# Genetic Epidemiological Pipeline Identifies Candidate Markers of Clozapine-Induced Metabolic Dysfunction Revealing Potential Avenues for Precision Clozapine Prescription

**DOI:** 10.64898/2026.03.17.26348301

**Authors:** Rory Shepherd, Vijayaprakash Suppiah, Anwar Mulugeta, Scott R. Clark, Elina Hyppönen, David Stacey

## Abstract

Clozapine is the gold-standard for treatment-resistant schizophrenia despite its severe metabolic complications, including metabolic syndrome (MetS) and type 2 diabetes (T2D) risk. A better understanding of the genetic factors influencing clozapine pharmacokinetics and their relationship to metabolic risk could help inform precision medicine approaches to clozapine prescribing. Using a series of genetic-epidemiological approaches, we aimed to identify candidate biomarkers associated with clozapine-induced metabolic dysfunction. We first used two-sample Mendelian randomization (MR) leveraging genome-wide association summary data to investigate evidence of causal relationships between clozapine metabolism and cardiometabolic traits. These analyses indicated that higher plasma clozapine levels and a higher clozapine-norclozapine ratio were both associated with a higher risk of T2D and higher blood pressure. We then applied a Phenome-scan-colocalization-MR pipeline to identify traits influenced by clozapine-metabolism loci that might serve as biomarkers of cardiometabolic risk. This pipeline identified 28 colocalizing candidate biomarkers associated with clozapine metabolising genetic loci. Subsequent MR highlighted associations for 16 of these 28 biomarker candidates with cardiometabolic outcomes, which included haematological markers and excretory traits (e.g. gamma-glutamyl transferase, red cell distribution width, and urea). These findings may inform the development of biomarker-guided monitoring approaches for risk stratification and early intervention, enabling a shift from reactive monitoring to predictive approaches in managing clozapine-induced metabolic dysfunction with appropriate clinical validation. These findings may also help to mitigate the risk of metabolic dysfunction associated with other antipsychotic medications.

## 1. Background

Clozapine is the gold-standard medication for treatment-resistant schizophrenia (TRS), which comprises ∼30% of all people with schizophrenia (SCZ). However, its therapeutic potential is severely constrained by adverse effects, particularly impacting the cardiometabolic system (1–5). Between 10 – 60% of all clozapine-treated patients develop metabolic syndrome (MetS), which is a cluster of interconnected cardiometabolic abnormalities including increased blood pressure and glucose levels, abnormal cholesterol, and excess abdominal fat (6, 7). These metabolic side effects are major contributors to the substantial mortality gap in SCZ, where patients experience a 15–20-year reduction in life expectancy compared to the general population (3, 8–12). Therefore, a precision medicine approach to safe clozapine prescribing has the potential to improve cardiometabolic outcomes, through early identification of high-risk patients and targeted co-administration of protective medications such as metformin or GLP-1 antagonists (13).

Both clozapine response and metabolism demonstrate substantial heritability, with twin studies indicating that approximately 73% of interindividual variation in *CYP1A2* activity, the primary enzyme metabolizing clozapine, is attributable to genetic factors (14). Several candidate gene studies have implicated serotonin receptor (*HTR2C*), leptin (*LEP*), and melanocortin 4 receptor (*MC4R*) genes in clozapine-induced weight gain and metabolic dysfunction (15–18), though well-powered GWAS of these metabolic outcomes have not yet been conducted due to modest sample sizes. However, GWAS of plasma clozapine and its primary metabolite norclozapine levels in people undergoing clozapine treatment have been undertaken (18–21). These studies have identified genome-wide significant and suggestive loci in key drug-metabolizing enzymes, refining our knowledge of clozapine pharmacokinetics and, with appropriate extension, other antipsychotic medications.

Clozapine is primarily metabolized in the liver by the cytochrome P450 enzyme complex, with *CYP1A2* playing a predominant role in converting clozapine to its major metabolite, N-desmethylclozapine (norclozapine) (18, 22–24). Secondary metabolic pathways involve *CYP2D6* and *CYP3A4* (*25*). Loci in *UGT1A* and *UGT2B* gene clusters that impact clozapine and norclozapine levels have also been identified (18). Overall, four key independent loci (*CYP1A2*, *CYP2D6*/*CYP2C19*, *UGT1A*, and *UGT2B* gene clusters) have been highlighted as associated with clozapine pharmacokinetics, providing robust genetic instruments for investigating relationships between drug exposure and metabolic outcomes (18, 20, 23, 26–28). Since individual differences in clozapine pharmacokinetics are a key regulator of treatment response and adverse effects (18–21), these pharmacokinetic GWAS data may proxy the genetic regulation of clozapine-induced metabolic dysfunction, thereby providing opportunities to inform safe clozapine prescribing.

In this study, we leveraged summary data from GWAS of plasma clozapine and norclozapine levels to determine whether individual differences in drug exposure alter the risk of metabolic disturbances. We then applied a series of genetic epidemiological methods (phenome-scan, statistical colocalization, and MR analyses) to identify biomarkers influenced by clozapine-metabolizing genetic loci that could inform precision medicine approaches to clozapine prescribing (29, 30). The key strength of this approach lies in its ability to overcome traditional confounding factors that limit observational studies in clozapine-treated populations, providing more reliable evidence for potential causal relationships while identifying measurable biomarkers for potential clinical use (31, 32).

## 2. Methods

### 2.1. Study Design and Overview

This study followed the STROBE-MR reporting guidelines for transparent MR studies and employed a multi-layered genomic approach to investigate potential causal relationships between plasma clozapine levels and cardiometabolic outcomes. This approach addresses key limitations of traditional observational studies in clozapine-treated patients, including small sample sizes, treatment-related confounding, and difficulty establishing temporal relationships between biomarker changes and metabolic dysfunction. By using inherited genetic variants as instrumental variables, our design has enabled the use of large population-based datasets to identify potential biomarkers and causal relationships limiting confounding seen in clinical studies.

The analytical strategy integrated direct genetic associations and potential biomarkers through a pipeline of genetic epidemiological methods, including PheWAS, colocalization, and MR (Figure 1). Our approach relied on three core MR assumptions: (**1**) genetic variants are strongly associated with clozapine plasma levels (relevance); (**2**) genetic variants are not associated with confounders (independence); (**3**) genetic variants affect outcomes only through clozapine exposure (exclusion restriction) (Supplementary Figures 1.1–1.4). Drug-metabolizing loci used for biomarker identification are highly pleiotropic, which may violate assumption 3, but this does not invalidate discovery as traits sharing genetic architecture with clozapine pharmacokinetics may be informative, regardless of causal pathway (33, 34).

**Figure 1:**
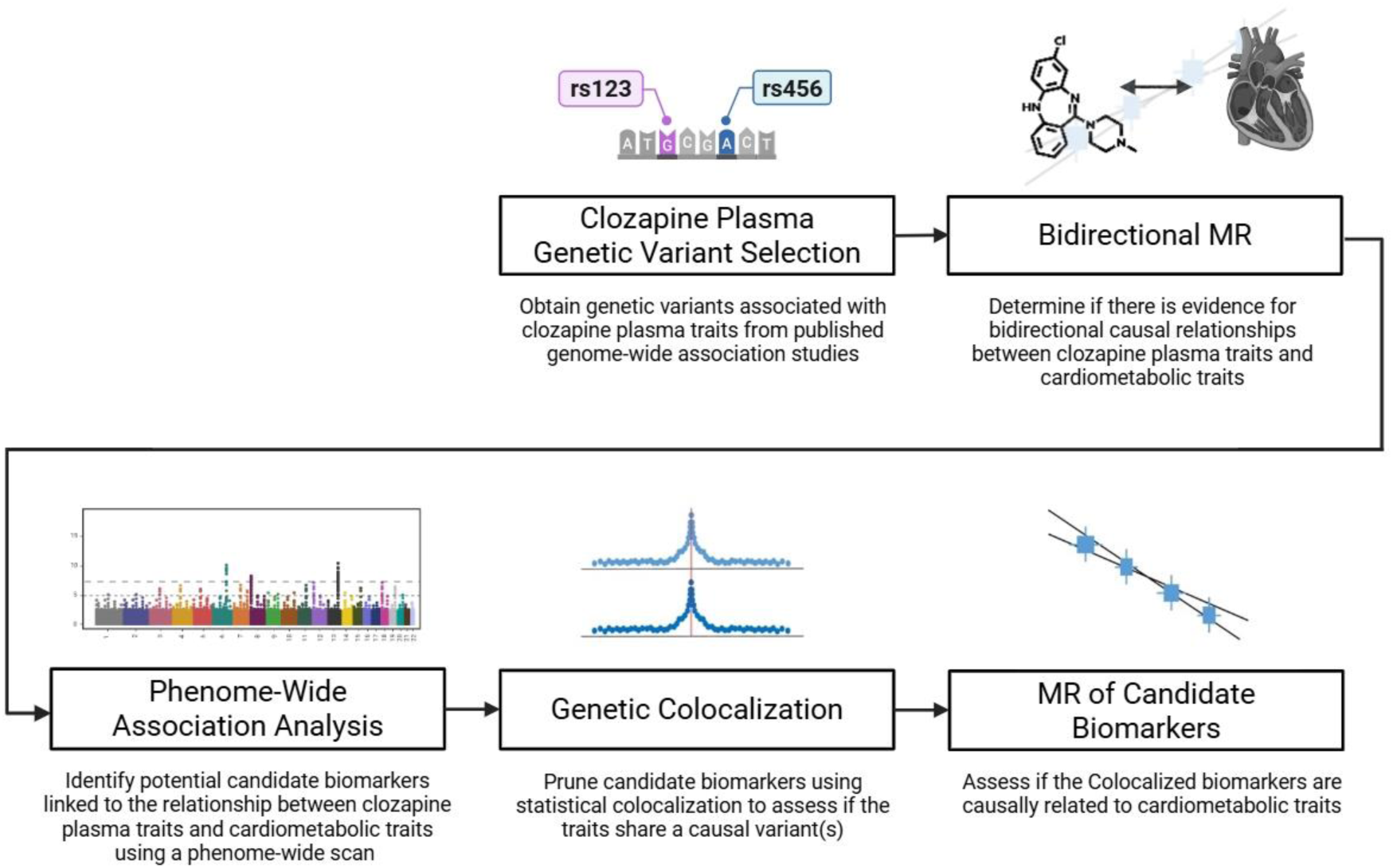
Multi-layered Analytical Framework for Investigating Clozapine-Induced Metabolic Dysfunction – Schematic representation of the four-stage analytical pipeline used in this study. The framework begins with bidirectional Mendelian randomization to examine direct causal relationships between clozapine plasma levels and cardiometabolic outcomes. This is followed by phenome-wide association study to identify candidate biomarkers, genetic colocalization to filter these candidates for shared genetic architecture, and finally targeted Mendelian randomization to assess potential causal effects of these biomarkers on metabolic outcomes.

### 2.2. Data Sources

Variant-level summary statistics for clozapine plasma levels were derived from the CLOZUK2 (Walters Group Data Repository, Centre for Neuropsychiatric Genetics and Genomics) GWAS (n=4,495), which provided statistics for plasma concentrations of clozapine, norclozapine, and the ratio of the two from a UK patient monitoring system (18). All datasets are European in ancestry to minimize population stratification bias.

Cardiometabolic GWAS represent population-based cohorts with minimal overlap with clozapine-treated populations, reducing sample overlap bias. Cardiometabolic outcomes were obtained from large-scale European GWAS studies through the OpenGWAS and GWAS Catalog platforms, including body mass index (BMI), blood pressure, HbA1c, lipid traits, T2D, and MetS (Table 1). Summary statistics for the identified candidate biomarkers were obtained based on PheWAS hits with the largest sample size and European ancestry.

**Table 1.**
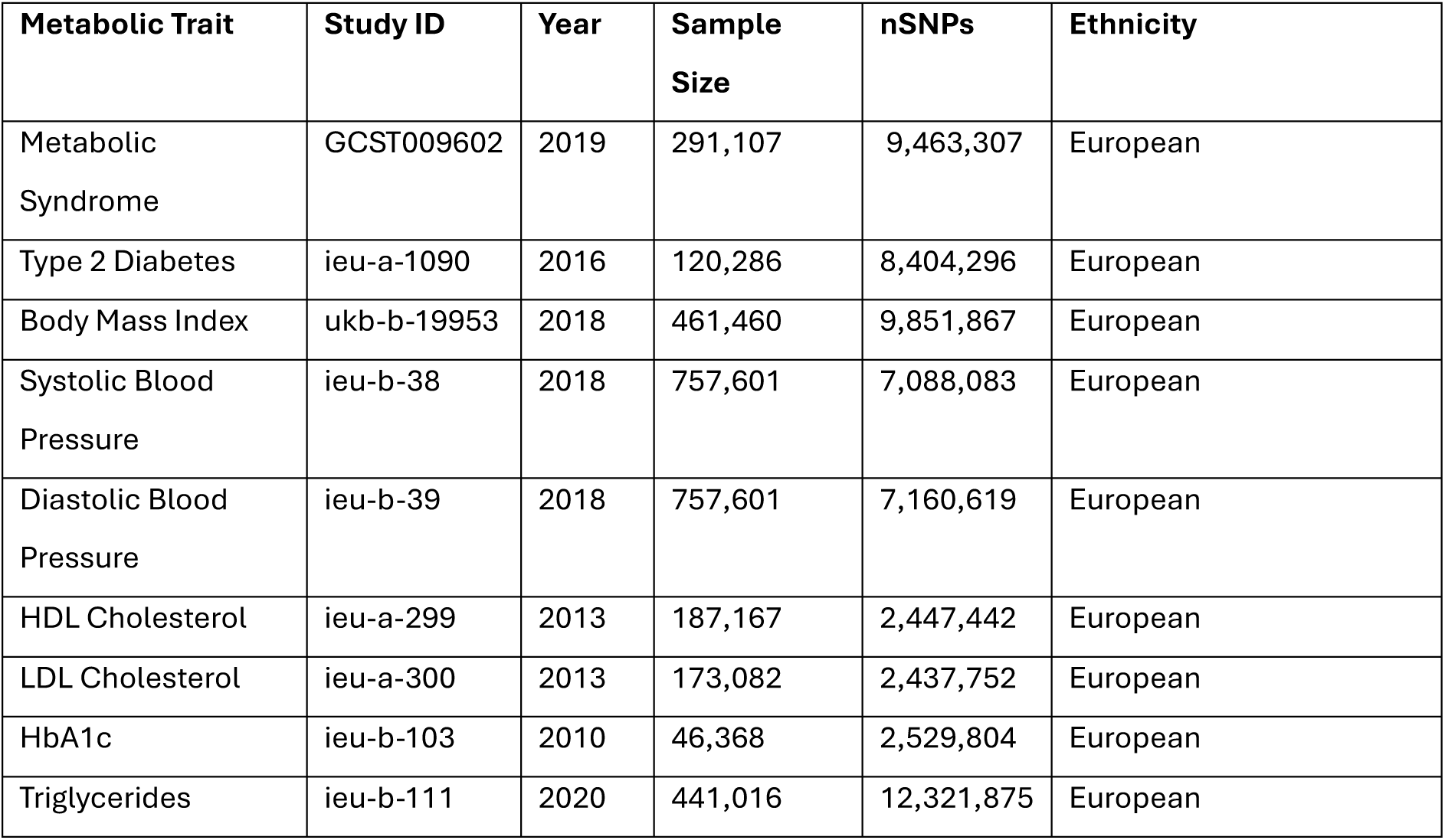
Metabolic outcome traits with their respective data sources obtained from OpenGWAS and GWAS Catalog.

### 2.3. Bidirectional Mendelian Randomization

To investigate genetic evidence for causal relationships between clozapine plasma levels and cardiometabolic outcomes, two-sample MR analyses were performed bidirectionally. Genetic instruments for clozapine pharmacokinetic traits were selected using a threshold of P<1.00×10⁻⁵ to ensure adequate instrument availability while maintaining biological relevance to drug metabolism pathways where genome-wide significant variants were insufficient. This threshold yielded 47 instrumental variables compared to only 9 at genome-wide significance, while maintaining robust instrument strength (Table 2; Supplementary Figures 1.1-1.2).

**Table 2.** Genetic instrumental variables for clozapine metabolism per clozapine exposure trait used in LD-Aware MR (clozapine metabolism effects on cardiometabolic outcomes). All instrumental variables were selected using P < 1.00×10⁻⁵ followed by linkage disequilibrium clumping (r² < 0.1, 10 Mb windows). F-statistics >10 indicate strong instruments with minimal weak instrument bias. Variants listed under multiple exposures serve as instruments for those respective phenotypes.

| SNP (GRCh37) | Chr | Position | EA | OA | EAF | Beta | SE | P-value | F-Statistic |
| --- | --- | --- | --- | --- | --- | --- | --- | --- | --- |
| **Clozapine Plasma** |  |  |  |  |  |  |  |  |  |
| rs3732218 | 2 | 234,627,304 | A | G | 0.1 | -0.217 | 0.031 | 3.30E-12 | 48.5 |
| rs6791266 | 3 | 1,446,510 | G | A | 0.21 | -0.114 | 0.024 | 2.10E-06 | 22.5 |
| rs6791077 | 3 | 140,176,199 | C | T | 0.23 | -0.101 | 0.022 | 3.90E-06 | 21.3 |
| rs75860583 | 3 | 165,859,297 | T | G | 8.00E-02 | -0.155 | 0.034 | 5.60E-06 | 20.6 |
| rs2100040 | 5 | 65,134,980 | A | G | 0.25 | 0.101 | 0.022 | 3.70E-06 | 21.4 |
| rs2389763 | 7 | 17,307,847 | C | T | 0.57 | -0.096 | 0.019 | 4.30E-07 | 25.6 |
| rs34872991 | 7 | 42,222,857 | A | G | 0.11 | 0.146 | 3.00E-02 | 1.10E-06 | 23.7 |
| rs1057868 | 7 | 75,615,006 | T | C | 0.27 | -0.094 | 0.021 | 9.00E-06 | 19.7 |
| rs56282157 | 7 | 37,411,377 | G | T | 8.00E-02 | -0.161 | 0.036 | 9.80E-06 | 19.5 |
| rs11020316 | 11 | 93,146,805 | C | T | 0.28 | -0.096 | 0.021 | 6.90E-06 | 20.2 |
| rs56133064 | 12 | 124,747,789 | G | A | 8.00E-02 | 0.155 | 0.034 | 3.50E-06 | 21.5 |
| rs73055220 | 12 | 17,655,750 | C | T | 0.14 | -0.126 | 0.027 | 4.60E-06 | 21 |
| rs2151870 | 13 | 72,813,751 | C | T | 0.21 | -0.108 | 0.023 | 3.80E-06 | 21.4 |
| rs1956171 | 14 | 94,677,145 | T | C | 0.28 | -0.101 | 0.021 | 1.90E-06 | 22.7 |
| rs854311 | 14 | 25,218,824 | A | C | 8.00E-02 | -0.161 | 0.036 | 5.70E-06 | 20.6 |
| rs2472297 | 15 | 75,027,880 | T | C | 0.24 | -0.15 | 0.023 | 4.40E-11 | 43.4 |
| rs62087495 | 18 | 9,398,708 | T | C | 8.00E-02 | 0.165 | 0.035 | 2.80E-06 | 22 |
| **Norclozapine Plasma** |  |  |  |  |  |  |  |  |  |
| rs4663325 | 2 | 234,616,357 | T | C | 0.12 | -0.203 | 0.028 | 8.10E-13 | 51.3 |
| rs11123265 | 2 | 115,736,492 | C | T | 0.27 | 0.101 | 0.021 | 1.30E-06 | 23.4 |
| rs77755140 | 3 | 165,859,270 | A | G | 8.00E-02 | -0.153 | 0.033 | 4.50E-06 | 21 |
| rs6791266 | 3 | 1,446,510 | G | A | 0.21 | -0.107 | 0.024 | 5.90E-06 | 20.5 |
| rs2926036 | 4 | 69,682,629 | G | A | 0.86 | 0.292 | 0.027 | 3.20E-28 | 121.4 |
| rs79870297 | 4 | 186,614,953 | T | C | 0.22 | -0.109 | 0.023 | 1.40E-06 | 23.2 |
| rs79204178 | 4 | 20,753,714 | C | T | 0.1 | 0.143 | 3.00E-02 | 2.50E-06 | 22.1 |
| rs112400791 | 5 | 175,991,431 | T | C | 6.00E-02 | -0.167 | 0.037 | 6.70E-06 | 20.3 |
| rs77634225 | 7 | 42,227,050 | C | T | 7.00E-02 | -0.164 | 0.036 | 5.80E-06 | 20.6 |
| rs12779195 | 10 | 77,631,585 | C | T | 0.49 | -0.083 | 0.019 | 7.40E-06 | 20.1 |
| rs34633304 | 11 | 11,419,070 | A | G | 0.13 | 0.133 | 0.028 | 2.70E-06 | 22 |
| rs12286285 | 11 | 107,399,326 | G | A | 0.31 | -0.088 | 0.019 | 5.80E-06 | 20.5 |
| rs854311 | 14 | 25,218,824 | A | C | 8.00E-02 | -0.178 | 0.034 | 1.50E-07 | 27.5 |
| rs1956168 | 14 | 94,680,291 | A | G | 0.28 | -0.093 | 2.00E-02 | 4.30E-06 | 21.1 |
| rs2472297 | 15 | 75,027,880 | T | C | 0.24 | -0.135 | 0.022 | 2.10E-09 | 35.9 |
| rs12918756 | 16 | 49,574,239 | T | G | 0.18 | -0.113 | 0.024 | 2.70E-06 | 22 |
| rs902589 | 18 | 63,842,961 | T | C | 0.87 | -0.134 | 0.029 | 3.70E-06 | 21.4 |
| rs4805573 | 19 | 30,995,755 | T | C | 0.1 | 0.141 | 0.032 | 8.20E-06 | 19.9 |
| **Clozapine-Norclozapine Plasma Ratio** |  |  |  |  |  |  |  |  |  |
| rs896809 | 2 | 19,976,506 | T | C | 0.28 | 0.094 | 0.021 | 7.30E-06 | 20.1 |
| rs835309 | 4 | 69,682,855 | G | T | 0.87 | -0.619 | 0.023 | 2.10E-153 | 696.1 |
| rs6830368 | 4 | 69,911,097 | G | A | 0.49 | 0.163 | 0.019 | 1.90E-18 | 76.8 |
| rs4272075 | 4 | 70,269,936 | A | G | 0.72 | -0.14 | 2.00E-02 | 3.90E-12 | 48.2 |
| rs115163219 | 4 | 70,366,972 | A | G | 5.00E-02 | 0.279 | 0.044 | 1.50E-10 | 41.1 |
| rs3777760 | 6 | 12,150,811 | A | G | 0.31 | 0.089 | 2.00E-02 | 8.70E-06 | 19.8 |
| rs41301394 | 7 | 75,612,803 | T | C | 0.27 | -0.115 | 0.021 | 4.80E-08 | 29.8 |
| rs62530100 | 9 | 7,652,916 | A | G | 0.1 | 0.143 | 3.00E-02 | 1.60E-06 | 23.1 |
| rs1926711 | 10 | 96,484,777 | A | G | 0.17 | 0.232 | 0.024 | 4.70E-22 | 93.2 |
| rs61832289 | 10 | 11,207,045 | A | G | 0.13 | -0.128 | 0.028 | 6.60E-06 | 20.3 |
| rs7103488 | 11 | 124,020,568 | T | C | 0.15 | -0.124 | 0.027 | 5.20E-06 | 20.8 |
| rs75929054 | 14 | 23,812,891 | C | T | 6.00E-02 | 0.177 | 0.039 | 5.10E-06 | 20.8 |

Instrumental variables were validated through LD clumping (r²<0.001) and strength assessment. All 47 variants exceeded the conventional F>10 threshold for strong instruments, with 91.5% demonstrating F>20, indicating minimal risk of weak instrument bias (35–38). Our genetic instruments captured steady-state plasma concentrations reflecting integrated pharmacokinetic processes rather than isolated metabolic activity. For reverse MR analyses, genetic instruments were selected at genome-wide significance for cardiometabolic traits (exposure) and tested against clozapine plasma traits as outcomes.

The primary analysis used inverse variance weighted (IVW) MR method, complemented by sensitivity analyses including MR-Egger and weighted median estimation which are robust to potential pleiotropy, and heterogeneity assessment using Q-statistics and I² values (39–42). Forest plots and scatterplots were generated to visualize the results (31, 43, 44). Leave-one-out plots used to test if the MR estimate was sensitive to a particular variant.

### 2.4. Phenome-wide Association Analysis

To identify candidate biomarkers for clozapine induced metabolic dysfunction, a PheWAS was conducted using the OpenGWAS platform (45). Ten lead variants at key loci previously identified at genome-wide significance as associated with clozapine pharmacokinetics in previous studies (Table 3) were used as the basis for this analysis (17, 18, 22, 46). The variants selected as leads are located on chromosomes 2, 4, 10, and 15 with a spread of mapped genes including *CYP1A, CYP2C, UGT1A,* and *UGT2B* genes. The PheWAS queried these variants against thousands of phenotypes available in OpenGWAS, with a genome-wide significance threshold applied to identify potentially relevant traits. The resulting associations were categorized into functional groups to identify key biological systems potentially involved in the clozapine-metabolic relationship (47–51).

**Table 3.** Lead variants used in the PheWAS-Colocalization-MR pipeline to identify potential intermediate phenotypes describing the biological pathways connecting clozapine metabolism to cardiometabolic abnormalities around key loci (*CYP* and *UGT* regions).

| SNP (GRCh37) | P-Value | Chr | Position | Mapped Gene(s) | Clozapine Associated Trait | Study Accession |
| --- | --- | --- | --- | --- | --- | --- |
| rs2011425 | 8.00E-9 | 2 | 233718962 | *UGT1A4* | Norclozapine | GCST007686 |
| rs3732218 | 3.00E-12 | 2 | 233718658 | *UGT1A9*,<br>*UGT1A6*,<br>*UGT1A8*,<br>*UGT1A5*,<br>*UGT1A7*,<br>*UGT1A10* | Clozapine,<br>Norclozapine | GCST90239836 |
| rs10000284 | 4.00E-20 | 4 | 68803465 | *UGT2B10* | Norclozapine | GCST010564 |
| rs10023464 | 2.00E-91 | 4 | 68794020 | *UGT2B10*,<br>*UGT2B15* | Ratio | GCST007684 |
| rs11725502 | 5.00E-15 | 4 | 68815224 | *UGT2B10* | Norclozapine | GCST007686 |
| rs7668556 | 4.00E-13 | 4 | 69164555 | *UGT2B11*,<br>*SPOPLP1* | Ratio | GCST007684 |
| rs835309 | 2.00E-153 | 4 | 68817137 | *UGT2B10* | Ratio | GCST90239838 |
| rs12767583 | 1.00E-16 | 10 | 94787706 | *CYP2C19* | Ratio | GCST010563 |
| rs1926711 | 5.00E-22 | 10 | 94725020 | *CYP2C18* | Ratio | GCST90239838 |
| rs2472297 | 4.00E-10 | 15 | 74735539 | *CYP1A1*,<br>*CYP1A2* | Clozapine | GCST007685 |

**Table 4.** Assessment of horizontal pleiotropy: Schizophrenia GWAS associations for clozapine pharmacokinetic lead variants. (*) Indicates that a proxy SNP was used due to missing lead variant in the schizophrenia dataset.

| Variant (GRCh37) | Chromosome : Position | Associated Trait | Clozapine Plasma Trait P-Value | Schizophrenia P-Value |
| --- | --- | --- | --- | --- |
| rs4663325 | 2 : 234,616,357 | Norclozapine | 8.10E-13 | 0.583 |
| rs3732218 | 2 : 234,627,304 | Clozapine | 3.30E-12 | 0.991 |
| rs2926036 | 4 : 69,682,629 | Norclozapine | 3.20E-28 | 0.841 |
| rs835309 | 4 : 69,682,855 | Ratio | 2.10E-153 | 0.358 |
| rs6830368 | 4 : 69,911,097 | Ratio | 1.90E-18 | 0.968 |
| rs13353587 | 4 : 70,312,793 | Ratio | 4.72E-40 | 0.137 |
| rs41301394 | 7 : 75,612,803 | Ratio | 4.80E-08 | 0.722 |
| rs12570829* | 10 : 96,474,651 | Ratio | 4.70E-22 | 0.135 |
| rs2472297 | 15 : 75,027,880 | Clozapine, Norclozapine | 4.40E-11 | 0.125 |

### 2.5. Statistical Colocalization

To distinguish true shared genetic effects from those attributable to LD, colocalization analyses were performed on the biomarkers identified through PheWAS. The Pairwise Conditional and Colocalization (PWCoCo) method was employed with the 1000 Genomes European reference panel (52). This method employs a Bayesian framework to calculate posterior probabilities for five hypotheses where the following hypotheses were implemented:

*H*_0_: No association with either trait

*H*_1_: Association with trait 1 only

*H*_2_: Association with trait 2 only

*H*_3_: Association with both traits, different causal variants

*H*_4_: Association with both traits, shared causal variant

For each identified phenotype, ±500Kb windows around lead variants for clozapine/norclozapine plasma levels were analysed, and a posterior probability threshold of H₄>0.80 was applied to signal robust colocalization indicating a shared causal variant between the traits (53–56).

### 2.6. Mendelian Randomization of Candidate Biomarkers

The phenotypes that demonstrated robust genetic colocalization with clozapine plasma traits were then used as exposures in a series of MR analyses against cardiometabolic outcomes bidirectionally. For these analyses, genetic instruments were selected at genome-wide significance. These analyses followed the same methodological approach as the primary MR analyses, including similar sensitivity analyses and visualization methods. This final analytical step aimed to quantify the potential causal effects of these candidates on metabolic outcomes, thereby identifying potential biological pathways that may be implicated in the relationship (Supplementary Figures 1.3-1.4) (31, 43, 44).

### 2.7. Genetic Correlation Analysis

To characterize the genetic architecture of candidate biomarkers and inform analytical strategy, we performed linkage disequilibrium score regression (LDSC) using the GenomicSEM R package (version 0.0.5). GWAS summary statistics for all 16 candidate biomarkers were first standardized using the munge function, which filtered SNPs to the HapMap3 reference panel, aligned alleles to the 1000 Genomes Phase 3 European population, and applied quality control filters (MAF > 0.01, INFO > 0.90). Genetic correlations (r_g_) were computed for all pairwise combinations using the ldsc function with pre-computed LD scores derived from the 1000 Genomes Phase 3 European reference panel (503 individuals, ∼1.2M HapMap3 SNPs). Hierarchical clustering using complete linkage was applied to the genetic correlation distance matrix (1 – |r_g_|). A height threshold of 0.85 was selected to define distinct biomarker clusters. Biomarkers with moderate/strong genetic correlations (|r_g_| > 0.15) clustering together were considered as sharing substantial genetic architecture, justifying joint analysis in multivariable frameworks.

### 2.8. Multivariable Mendelian Randomization

Based on genetic correlation results, we performed focused multivariable MR (MVMR) on biomarkers demonstrating moderate/strong genetic correlations. MVMR extends conventional univariable MR by simultaneously estimating the independent causal effects of multiple correlated exposures on an outcome. We used the MVMR R package to implement inverse-variance weighted (IVW) MVMR, selecting genetic instruments that were genome-wide significant. Instrument strength was assessed using conditional F-statistics, with F > 10 indicating strong instruments.

Additional statistical and procedural details are available in the Supplementary Methods.

## 3. Results

### 3.1. Clozapine Plasma Levels are Associated with Cardiometabolic Outcomes

To determine whether individual differences in clozapine plasma levels influence the risk of metabolic outcomes, we conducted two-sample MR analyses using plasma levels of clozapine, norclozapine, and their ratio as exposures against cardiometabolic outcomes. All genetic instruments demonstrated robust strength, indicating minimal weak instrument bias and supporting reliable causal inference (Table 2).

Using IVW MR as the main analysis with a Bonferroni adjusted p-value threshold of (P=0.05/27=1.85×10⁻³), we found that clozapine plasma levels were strongly associated with T2D (OR=1.62, SE=0.036, P=5.05×10⁻⁴¹) and systolic blood pressure (β=0.331, SE=0.054, P=7.63×10⁻¹⁰). We also found some evidence for a protective association with MetS (OR=0.99, SE=0.005, P=2.99×10⁻²), though this did not survive correction for multiple testing.

While norclozapine levels did not reveal any significant associations in primary IVW-MR analyses, the clozapine-norclozapine ratio was associated with diastolic blood pressure (β=0.210, SE=0.023, P=1.30×10⁻¹⁹) and T2D (OR=1.74, SE=0.165, P=7.92×10⁻⁴). A higher clozapine-norclozapine ratio was also associated with lower risk of MetS (β=-0.018, SE=0.005, P=1.48×10⁻⁴) (Figure 2).

**Figure 2:**
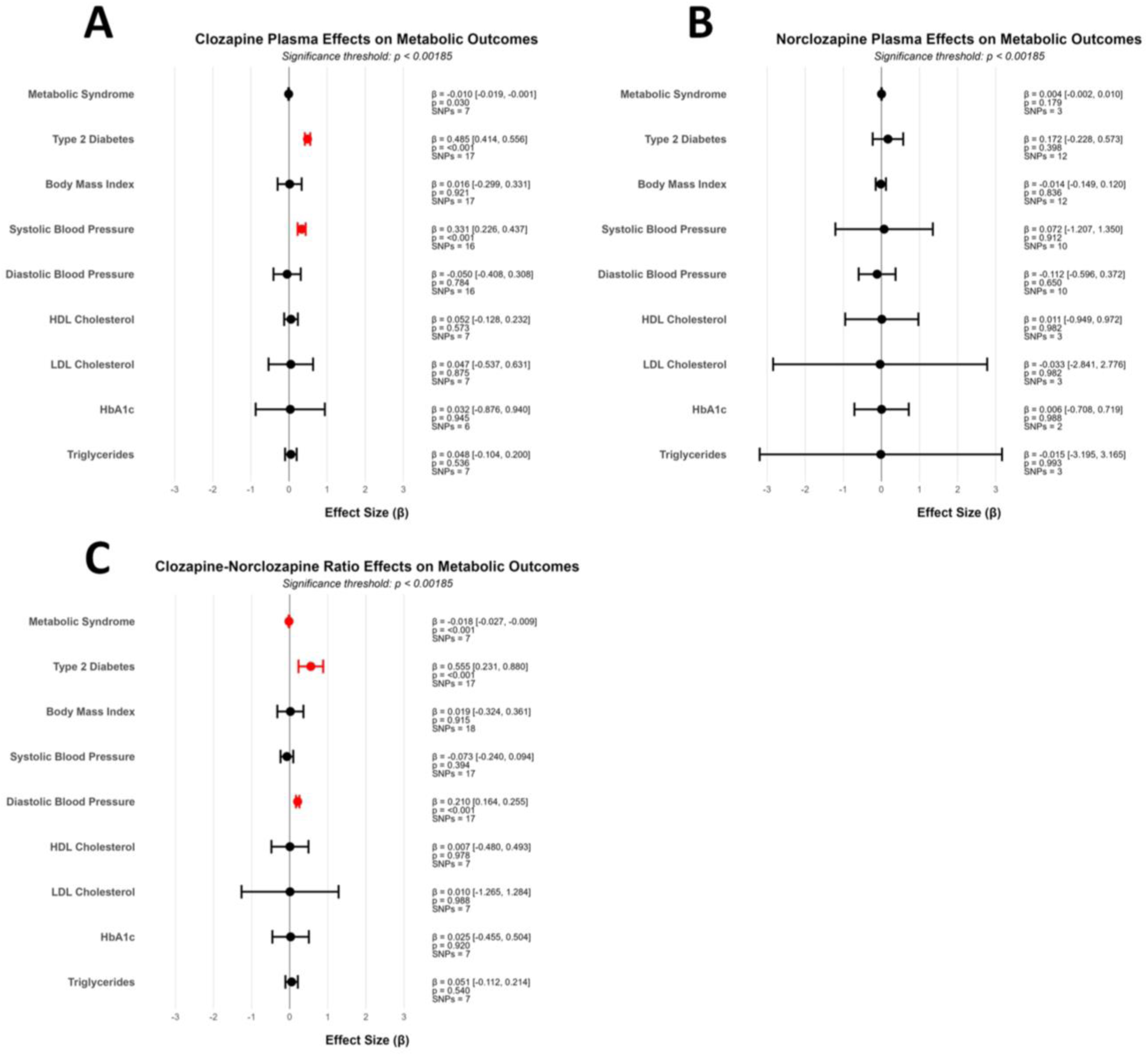
Direct Effects of Clozapine Metabolism on Cardiometabolic Outcomes – Forest plots showing causal effect estimates (β), 95% confidence intervals, number of instrumental variables, and P-values from inverse-variance weighted Mendelian randomization analyses. Results are presented for (A) Clozapine plasma levels, (B) Norclozapine plasma levels, and (C) Clozapine-Norclozapine ratio against multiple metabolic outcomes including metabolic syndrome, type 2 diabetes, body mass index, blood pressure, and lipid parameters. Red points indicate statistically significant associations after multiple testing correction (P < 1.85×10⁻³).

These findings remained consistent across multiple sensitivity analyses, with little evidence of directional pleiotropy for most analyses. MR-Egger regression detected directional pleiotropy in 5 of 27 analyses (18.5%). The remaining 22 analyses showed no evidence of directional pleiotropy (all Egger intercept p>0.05), suggesting instruments primarily influence metabolic outcomes through clozapine pharmacokinetic pathways rather than independent pleiotropic effects. For the clozapine plasma-T2D relationship (MR-Egger intercept P=3.10×10⁻³), the association remained significant and directionally consistent in pleiotropy-robust weighted median analysis (β=0.521, P=4.56×10⁻³). For MetS analyses of the clozapine-norclozapine ratio (MR-Egger intercept P=1.83×10⁻⁵), the association was attenuated and lost significance in weighted median analysis (β=-0.006, P=0.80), suggesting this relationship may be influenced by pleiotropy. (Supplementary Table 1; Supplementary Figures 2.1-2.3). We found limited evidence of reverse causation with the only exception being potential causal effects of T2D on blood plasma levels of both clozapine (β=0.098, SE=0.032, P=1.91×10⁻³) and norclozapine (β=0.090, SE=0.031, P=3.46×10⁻³). However, these associations did not withstand multiple testing correction (Supplementary Table 2; Supplementary Figures 2.4-2.6).

To ensure that observed associations between clozapine pharmacokinetic traits and metabolic outcomes reflect medication effects rather than disease liability, we verified that genetic variants associated with clozapine pharmacokinetics showed no association with SCZ risk. LD-clumping identified 9 independent variants across the three pharmacokinetic traits, spanning 6 genomic regions (chromosomes 2, 4, 7, 10, and 15). All 9 lead variants were present in the SCZ GWAS dataset (excluding one where a proxy SNP was used). None of the 9 variants showed even nominal significance in the SCZ dataset (all P>0.05), demonstrating strong evidence against horizontal pleiotropy through disease liability pathways. A proxy SNP rs12570829 was used for rs1926711 (10.1 Kb distant, same genomic region, r^2^=0.992) (Figure 3; Table 3).

**Figure 3:**
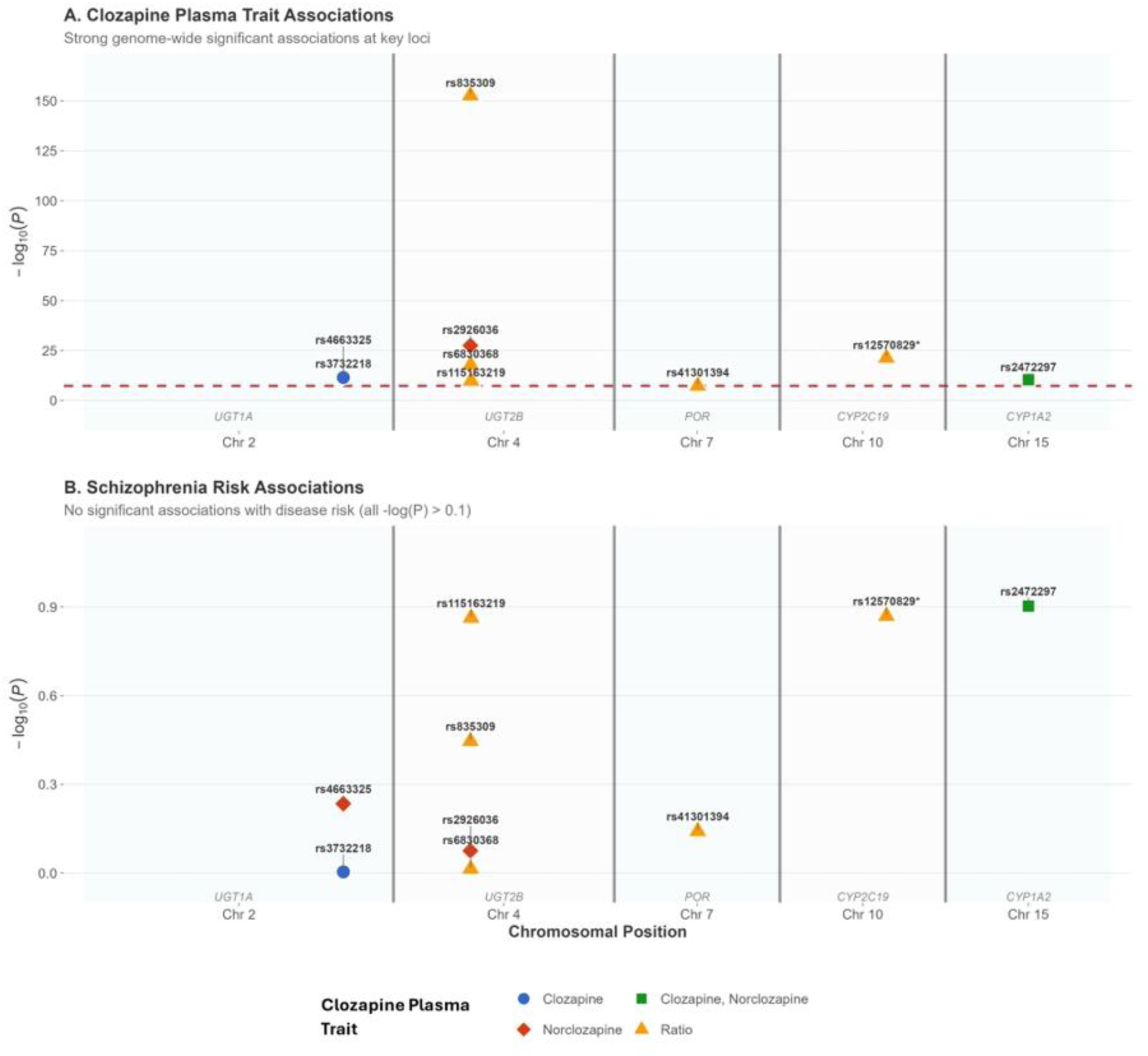
Assessment of Pleiotropy Between Schizophrenia and Clozapine Pharmacokinetics – Schizophrenia GWAS associations for clozapine pharmacokinetic lead variants. (A) Clozapine plasma trait associated –log(P) from the CLOZUK dataset, all variants significant at a genome-wide level. (B) Corresponding PGC schizophrenia dataset variant –log(P), no variants reach even nominal significance (P=0.05). Variant details: rs4663325 and rs3732218 (UGT1A cluster), rs2926036, rs835309, rs6830368 and rs115163219 (UGT2B cluster), rs41301394 (POR), rs12570829* (CYP2C19), rs2472297 (CYP1A2). Red line: genome-wide significance (P = 5.00×10⁻⁸). *rs12570829 used as a proxy SNP for rs1926711 which was not present in the PGC schizophrenia dataset.

### 3.2. Phenome-wide Association Identified Candidate Traits Influenced by Clozapine-Metabolising Genetic Loci

Given the evidence that individual differences in clozapine plasma levels influence cardiometabolic outcomes, we sought to identify measurable candidate traits influenced by clozapine-metabolizing genetic loci that could potentially be evaluated for risk stratification in people with SCZ. To this end, a previously established genetic epidemiological pipeline beginning with a PheWAS with a significance threshold of P<5×10⁻⁸ (Table 3) was employed (34). This approach yielded 391 significant associations across 203 unique studies, representing 98 distinct candidate biomarker traits. These hits clustered into several biological categories, including liver function markers, renal markers, body composition measurements, and dietary factors (Figure 4; Supplementary Table 3; Supplementary Figures 3).

**Figure 4:**
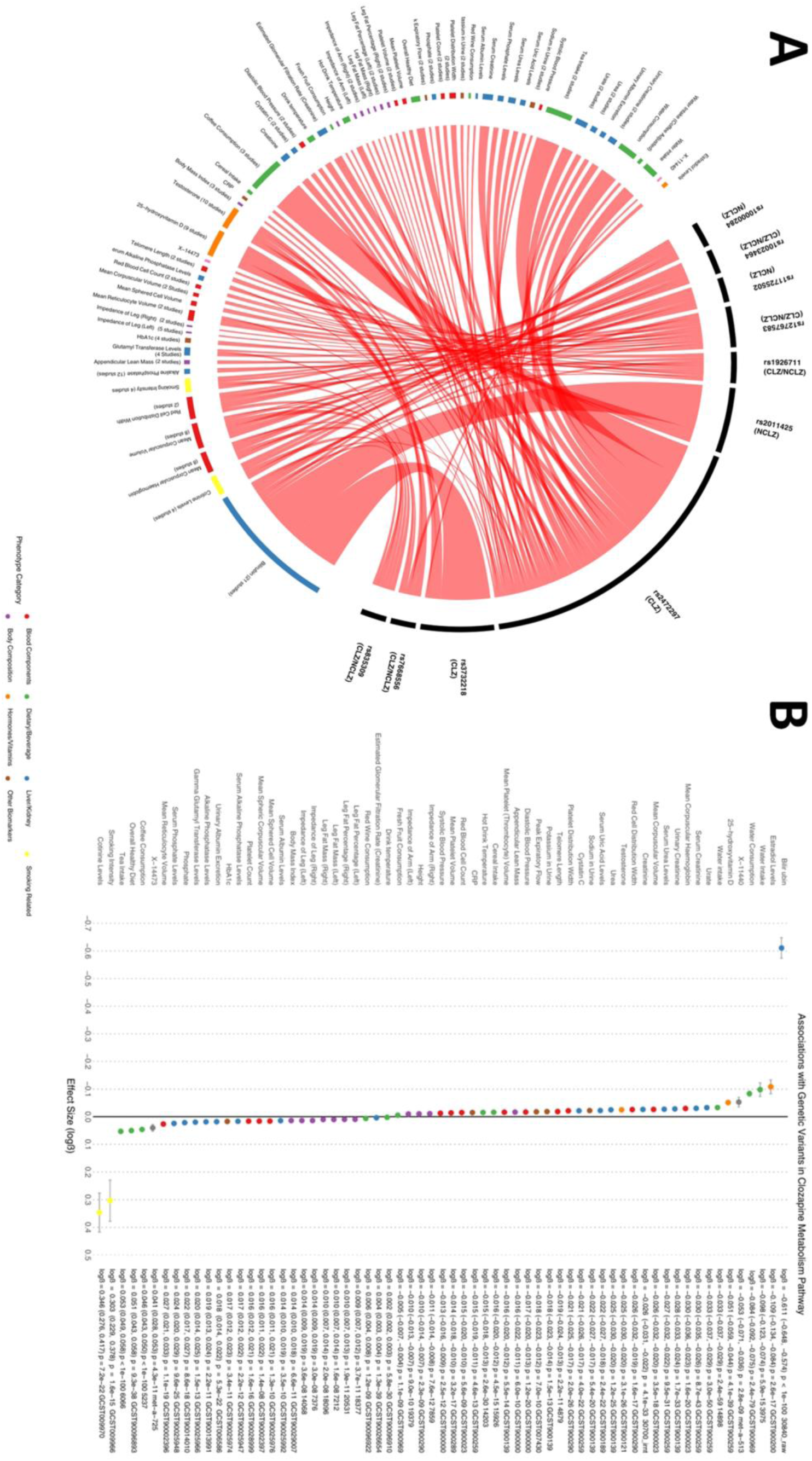
Phenome-Wide Analysis Results Identifying Candidate Biomarkers Modulating the Relationship Between Clozapine Metabolism and Cardiometabolic Outcomes – Graphical representation of the results from the PheWAS analysis run through OpenGWAS. (A) Circular plot of the associated phenotypes relating to a specific lead variant used for the analysis. When multiple studies reported the same trait, number of studies is displayed. Identified phenotypes are colour-coded by trait category. (B) Forest plot of the same identified phenotypes also colour-coded by trait category. Forest plot shows causal effect estimates (log(β)) with 95% confidence intervals, specific P-value, and associated study ID for each identified trait. When multiple studies reported the same trait, the most significant hit was used.

The lead variant rs2472297 near *CYP1A1*/*CYP1A2* emerged as the predominant driver of associations, demonstrating strong connections to caffeine metabolism and various metabolic traits. Similarly, rs3732218 (located within *UGT1A8*) demonstrated strong associations with bilirubin, while rs10000284 showed connections to smoking and nicotine metabolism.

### 3.3. Colocalization Confirms Shared Genetic Architecture Between Clozapine Pharmacokinetics and Candidate Biomarkers

The PheWAS results could potentially include traits that do not share the same genetic etiology with clozapine pharmacokinetic traits and are therefore confounded by LD. To identify and remove these spurious associations, we conducted pairwise colocalization analyses using PWCoCo, which combines conditional analyses with colocalization to address the single causal variant assumption limitation (Supplementary Methods).

From the 98 candidate biomarker traits identified in the PheWAS, we uncovered robust evidence of colocalization (H₄>0.80) with clozapine plasma traits for 28 phenotypes (28.6%) including renal, inflammatory, liver, and haematological markers (Supplementary Table 4; Supplementary Figures 4.1-4.2). The network visualization of colocalizing biomarker traits (Figure 5) illustrated a wide variety of traits impacted by the clozapine metabolising loci. The clustering of colocalization signals among functionally related traits (renal markers, body composition measures, liver function tests) suggested that these variants operate through coherent biological pathways. Notably, several phenotypes colocalized at multiple independent loci, providing particularly strong evidence for shared genetic architecture with clozapine pharmacokinetic traits. For example, bilirubin showed colocalization across the four independent loci with the clozapine-norclozapine ratio (H_4_ = 0.927-1.000), while total testosterone colocalized at two independent loci (H_4_ = 0.901-0.957), and 25-hydroxyvitamin D demonstrated both conditional and unconditional colocalization signals across multiple analyses (H_4_ = 0.939-1.000) (Supplementary Figures 4.3-4.5; Supplementary Table 4).

**Figure 5:**
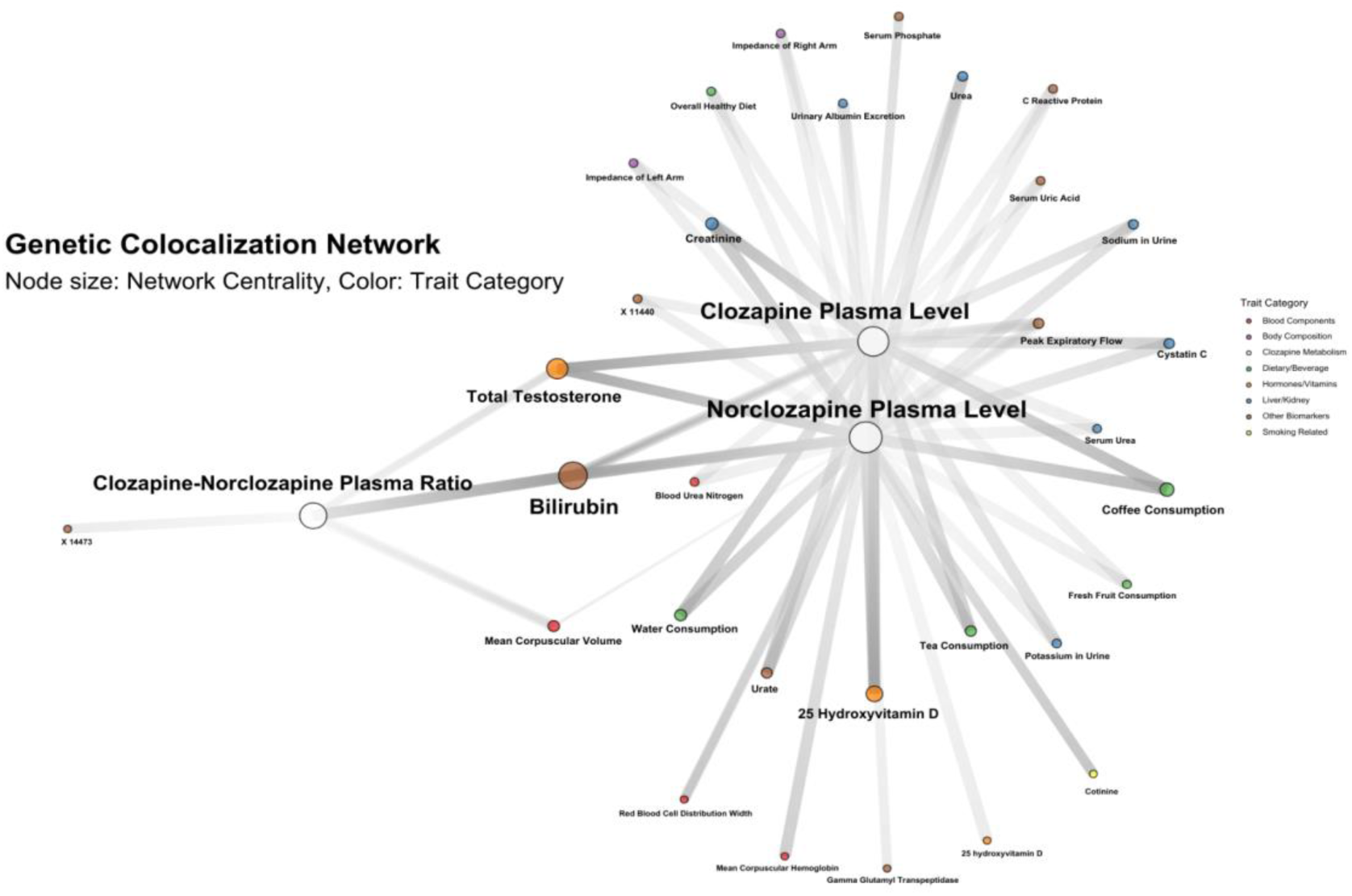
Genetic Colocalization Network of Clozapine Metabolism and Candidate Biomarkers-Network visualization of phenotypes demonstrating robust genetic colocalization with clozapine plasma traits. Nodes represent phenotypes color-coded by trait category as shown in the legend, with node size indicating network centrality. The three clozapine plasma traits (clozapine plasma level, norclozapine plasma level, and clozapine-norclozapine ratio) are positioned at the core of the network, with connecting lines representing robust genetic colocalization (H₄ > 0.80).

The colocalization analysis effectively prioritized the list of candidate biomarker traits to a set with strong evidence of shared genetic architecture with clozapine pharmacokinetics, establishing a foundation for subsequent MR.

### 3.4. Bidirectional Mendelian Randomization Identifies 16 Key Biomarkers with Evidence of Causality to Cardiometabolic Outcomes

To determine which of the colocalized phenotypes might causally influence metabolic outcomes, we conducted MR analyses using these phenotypes individually as exposures against 9 cardiometabolic outcomes. From the 28 colocalized phenotypes, 16 demonstrated evidence consistent with causal effects (P=0.05/9=5.56×10⁻³, correction for 9 cardiometabolic outcomes) on at least one cardiometabolic outcome, suggesting their potential utility as candidate clinical biomarkers.

Gamma glutamyl transferase (GGT) showed particularly strong effect on glucose homeostasis, with substantial impact on T2D risk (OR=0.072, SE=0.126, P=2.13×10⁻⁹⁷), along with effects on blood pressure and LDL cholesterol which were directionally consistent with T2D. Red blood cell distribution width (RDW) emerged as a potential biomarker of cardiovascular effects, with strong associations with both diastolic (β=0.653, SE=0.010, P<1.00×10⁻³⁰⁰) and systolic blood pressure (β=1.181, SE=0.031, P<1.00×10⁻³⁰⁰), as well as associations with LDL cholesterol and T2D. Additionally, 25-hydroxyvitamin D demonstrated effects on T2D and triglycerides (T2D: OR=1.680, SE=0.151, P<6.07×10⁻^4^; Triglycerides: β=0.507, SE=0.162, P<1.74×10⁻³) and total testosterone exhibited significant influence on BMI (β=0.047, SE=0.014, P<9.86×10⁻^4^) (Figure 6 (A); Supplementary Table 5).

**Figure 6:**
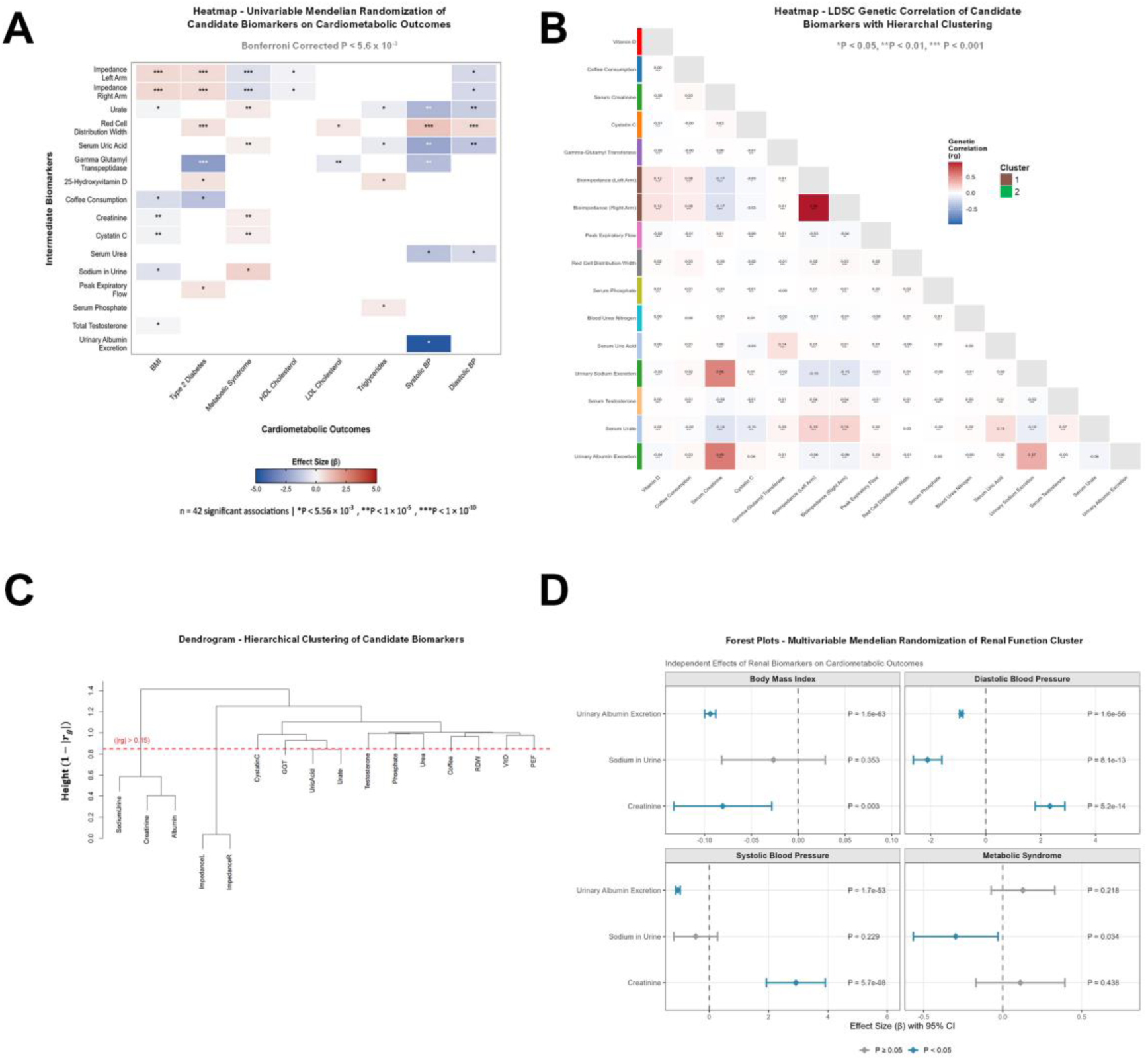
Genetic Architecture and Multivariable Causal Analysis of Biomarkers on Cardiometabolic Outcomes –. **(A)** Heatmap of univariable Mendelian randomization effect estimates for 16 candidate biomarkers across 8 cardiometabolic outcomes. Effect sizes (β) shown with significance indicated by asterisks (*P < 5.56 × 10^-3^, **P < 1 × 10^-5^, ***P < 1 × 10^-10^). Bonferroni-corrected significance threshold: P < 5.56 × 10^-3^. **(B)** Genetic correlations between candidate biomarkers estimated by linkage disequilibrium score regression (LDSC). Genetic correlation coefficients (r_g_) indicated by cell colour and significance levels indicated in each cell indicated by asterisks (***P < 0.001, **P < 0.01, *P < 0.05). Biomarkers ordered by hierarchical clustering (complete linkage, distance = 1 – |r_g_|, height threshold = 0.85) **(C)** Hierarchical clustering dendrogram of 16 candidate biomarkers based on genetic correlation distance (1 – |r_g_|). **(D)** Forest plots showing multivariable Mendelian randomization results for renal biomarkers against key cardiometabolic outcomes. Effect estimates (β) with 95% confidence intervals shown. Blue markers indicate P < 0.05 and grey markers indicate P > 0.05. Each biomarker shows independent causal effects despite genetic correlation.

Several other colocalizing phenotypes also demonstrated strong causal effects on metabolic outcomes. Body composition measures showed strong associations with MetS (left arm impedance: β=0.721, SE=0.031, P=1.41×10⁻¹¹⁸; right arm impedance: β=0.717, SE=0.032, P=1.02×10⁻¹¹⁰). Renal function markers, particularly blood urea nitrogen, showed significant associations with MetS and blood pressure. Coffee consumption demonstrated negative associations with both BMI (β=-0.754, SE=0.191, P=7.83×10⁻⁵) and T2D risk (OR=0.223 per SD increase in coffee intake, P=1.28×10⁻⁴), consistent with established protective effects of coffee consumption on metabolic outcomes. (Figure 6A; Supplementary Table 5; Supplementary Figures 5.1-5.3).

To assess bidirectionality, we conducted reverse MR analyses using cardiometabolic traits as exposures against the 28 colocalized phenotypes. After Bonferroni correction for multiple testing (P=0.05/28=1.79×10⁻³), reverse associations were identified across several key biomarker traits. Vitamin D and RDW both demonstrated reverse effects, suggesting complex feedback mechanisms rather than simple unidirectional pathways. Bioelectrical impedance also had noteworthy relationships with BMI and MetS (Supplementary Table 6; Supplementary Figures 5.4).

### 3.5. Genetic Correlation Analyses Highlight a Cluster of Renal System Markers

To identify opportunities for reducing redundancy among candidate biomarkers, we examined their genetic relationships using LD score regression and hierarchical clustering analyses. Overall, MR analyses prioritised 16 candidate biomarkers associated with metabolic dysfunction.

We therefore sought to determine which of these candidates are independent and whether there could be opportunities to remove redundancy. We first conducted LD score regression analyses, which revealed that most candidate biomarkers were genetically independent, with weak or negligible genetic correlations (|r_g_| < 0.15) (Figure 6B). Hierarchical clustering identified 2 distinct clusters, with 11 candidate markers failing to cluster. This is consistent with substantial genetic independence across most traits (Figure 6C).

Notably, three biomarkers formed a ‘renal function cluster’ (Cluster 2) consisting of serum creatinine, urinary sodium excretion, and urinary albumin excretion. These markers demonstrated moderate-to-strong genetic correlations, with the strongest correlation observed between creatinine and urinary albumin (r_g_ = 0.59, P = 8.2×10⁻¹¹⁷), followed by creatinine and urinary sodium (r_g_ = 0.56, P = 0.023), and finally urinary sodium and urinary albumin (r_g_ = 0.37, P = 0.035). The only other multi-biomarker cluster identified was Cluster 1, comprising bilateral bioimpedance measurements of the left and right arms, which showed near-perfect genetic correlation (r_g_ = 0.96, P = 5.0×10⁻²³⁹), as expected for measurements of the same physiological parameter (Supplementary Table 7 and 8).

### 3.6. Multivariable Mendelian Randomization Analysis Reveals Independent Causal Effects of Renal Biomarkers

To further explore potential relationships between the 16 candidate biomarkers, we conducted multivariable MR analyses (MVMR). We first conducted exploratory MVMR using all 16 candidates as exposures across 9 cardiometabolic outcomes, but this resulted in very weak instruments (F<10). The findings should therefore be treated with caution (Supplementary Table 9; Supplementary Figures 6).

We then conducted targeted MVMR analyses focused on the ‘renal function’ cluster we identified in LDSC analyses. This analysis wielded F-statistics (F ∼ 10) stronger than the exploratory MVMR but still below the generally accepted threshold for strong causal inference. Despite the substantial genetic correlations observed, all three renal markers showed independent associations with multiple cardiometabolic outcomes (Figure 6D; Supplementary Table 8). For body mass index, a genetic propensity to higher creatinine levels was associated with lower BMI (β = −0.26, P = 3.5×10⁻²³), while higher urinary sodium (β = 0.41, P = 1.4×10⁻¹⁵²) and urinary albumin (β = 0.40, P = 2.3×10⁻³⁹) were associated with higher BMI (Figure 6D; Supplementary Table 10).

Similar patterns of independent effects were observed for blood pressure, with all three markers showing associations with both DBP (creatinine: β = −0.05, P = 5.2×10⁻¹⁴; urinary sodium: β = 0.03, P = 9.1×10⁻¹³; urinary albumin: β = 0.03, P = 1.7×10⁻⁵¹) and SBP. For MetS, urinary albumin (β = 0.26, P = 0.04) and urinary sodium (β = 0.14, P = 0.03) were nominally associated. Notably, creatinine and urinary albumin/sodium demonstrated opposing directions of effect on BMI and blood pressure despite their positive genetic correlations, a pattern consistent with the clinical use of urinary albumin-to-creatinine ratio (UACR) as a marker of kidney damage (57–59). These results indicate that the renal function markers, despite their genetic correlation, each provide independent and complementary information about cardiometabolic risk, suggesting that composite measures such as ratios may be particularly informative for monitoring metabolic health in people undergoing clozapine treatment.

## 4. Discussion

This study presented a comprehensive genetic investigation of clozapine-induced metabolic dysfunction using established genetic epidemiological methods. We systematically investigated the complex relationships between clozapine plasma traits and cardiometabolic outcomes, identifying evidence of causal relationships and 16 candidate biomarkers potentially useful for identifying patients at risk of clozapine-induced metabolic dysfunction.

This study has addressed methodological challenges that limit research into clozapine-induced metabolic dysfunction. Traditional approaches to identifying biomarkers of cardiometabolic risk in clozapine-treated patients face significant constraints due to the relatively small population receiving this medication, polypharmacy, and inconsistent treatment timelines complicating longitudinal studies. Furthermore, existing studies have established only correlational relationships that cannot definitively determine whether changes in biomarkers precede metabolic disturbance or represent consequences of existing dysfunction. To circumvent these limitations, we leveraged genetic loci robustly associated with clozapine plasma levels, which enabled us to utilize GWAS summary data from the general population to establish potential causal relationships and identify candidate biomarkers. Our approach is inherently exploratory and hypothesis-generating, applying established methods to identify candidate biomarkers that require prospective validation in clinical cohorts before translation to practice.

### 4.1. Evidence of Causal Effects

Our analysis revealed strong evidence for causal effects of clozapine plasma levels on T2D (OR=1.62, P=5.05×10⁻⁴¹), while a nominal protective association with MetS (OR=0.99, P=0.030) did not survive multiple testing correction. This paradox requires careful interpretation. First, the MetS effect size is extremely small (OR=0.99) and unlikely to represent a clinically meaningful protective effect. Second, this pattern could reflect collider bias: MetS is diagnosed based on clustering of multiple risk factors including diabetes. Because clozapine has strong effects on T2D (one component of MetS) but potentially weaker or null effects on other components, the composite MetS outcome may produce spurious associations that don’t reflect the true causal effect on MetS as a whole. In statistical terms, analysing a composite outcome when the exposure has heterogeneous effects on its components can induce bias. Third, while MR-Egger intercept testing did not detect significant directional pleiotropy (P=0.250), the association was attenuated and lost significance in weighted median analysis (P=0.80), suggesting it may not represent a genuine causal effect. Therefore, the nominal MetS association should not be considered evidence of a true protective effect. The strong T2D association likely reflects the effect of clozapine through well-established diabetogenic mechanisms including insulin resistance and impaired glucose metabolism.

The analysis of plasma clozapine, norclozapine and their ratio revealed metabolite-specific effects on metabolic outcomes (60). While primary IVW analysis for norclozapine showed non-significant associations with BMI and metabolic outcomes, sensitivity analyses revealed significant negative associations with LDL cholesterol and triglycerides, contrasting with positive effects observed for clozapine plasma. These opposing directions between parent compound and metabolite align with pharmacological precedence where metabolites possess distinct biological activity. For instance, acetaldehyde rather than alcohol itself appears to drive cancer associations (61, 62). The clozapine-norclozapine ratio analyses revealed significant associations with diastolic blood pressure, T2D, and MetS, suggesting that relative concentrations of parent compound and metabolite have distinct cardiovascular implications.

These genetic findings partially align with clinical meta-analysis data showing positive correlations between clozapine serum levels and triglycerides and norclozapine levels with both triglycerides and total cholesterol (63). However, the opposing genetic effects between clozapine and norclozapine on lipid metabolism suggest complex regulatory mechanisms that may not be captured in observational clinical correlations (61, 64).

### 4.2. Downstream Effects of Clozapine Metabolising Loci

PheWAS-colocalization-MR analyses revealed a network of biological pathways that provided a detailed genetic picture regarding clozapine’s influences on metabolic outcomes. The identification of these candidate biomarkers is key, as it helps establish a hierarchical organization of metabolic effects, moving beyond simple correlations.

Body composition measures serve as positive controls that validate our primary findings rather than representing independent biomarkers. Bioelectrical impedance measurements showed consistent effects across both arms, which is strongly supported by reverse MR showing evidence that BMI and MetS causally influence impedance measurements. These findings confirm the expected relationships between clozapine exposure and body composition changes.

The strong association demonstrating higher RDW and increased blood pressure indicated that haematological changes may potentially play a role in clozapine’s cardiovascular effects (65). However, reverse MR analyses revealed significant reverse relationships with triglycerides and MetS, suggesting complex feedback loops rather than simple unidirectional relationships. Similarly, GGT’s impact on glucose homeostasis is clinically significant as GGT elevation indicates hepatic toxicity, which may be related to inflammatory processes (66).

The effects of 25-hydroxyvitamin D on T2D and triglycerides revealed vitamin D pathways’ role in energy regulation during clozapine treatment, though bidirectional analyses showed some reverse causation, indicating complex vitamin D-metabolic feedback mechanisms. Higher total testosterone associated with lower BMI demonstrated that endocrine disruption represents a potential mechanism, with reverse MR confirming bidirectional testosterone-metabolic relationships. This finding has important clinical implications as males have been reported to be more likely to develop metabolic abnormalities such as high cholesterol or triglycerides, hypertension, and cardiovascular risk (67). This suggests sex-specific biomarker profiles may be clinically useful for risk stratification.

Notably, many of the excretory traits identified in the analyses demonstrated significant associations with blood pressure parameters. Markers of renal function such as urinary albumin excretion, serum urea, and creatinine all showed robust associations with both systolic and diastolic blood pressure. This pattern is biologically plausible given the fundamental role of excretory systems, particularly the kidneys, in blood pressure homeostasis through regulation of fluid volume and electrolyte balance (68). However, these findings warrant further investigation as they contradict the typical understanding derived from observational studies that increased urate, uric acid, and urea, lead to increased blood pressure (69–71). This apparent contradiction potentially reflects a distinction between observational correlations and MR. While observational studies consistently demonstrate that people with high blood pressure tend to have elevated uric acid and urea levels, our MR analyses suggest these associations may not reflect causal relationships (69–71). Instead, this could represent compensatory physiological responses to existing blood pressure dysfunction rather than causal drivers of hypertension. This pattern of contradictory findings is well-documented in psychiatric genetic epidemiology and elsewhere, exemplified by studies of C-reactive protein and SCZ where observational associations do not align with estimates from genetic instruments (72, 73). These discrepancies underscore the value of MR in distinguishing candidate causal estimates from confounded observational associations (43).

LD score regression demonstrated that most biomarkers were genetically independent, with only three forming a coherent renal function cluster characterized by moderate-to-strong genetic correlations. This genetic clustering provided strong justification for focused multivariable MR analysis of these markers, as their shared genetic architecture necessitates joint modelling to obtain valid estimates of independent causal effects. Importantly, this data-driven clustering approach addresses potential concerns about selective biomarker inclusion. The naive MVMR sensitivity analysis, while expected to suffer from weak instrument bias when including all 16 biomarkers simultaneously, provides potential validation: the renal markers retained very small *p*-values, while other biomarkers showed substantial attenuation. This differential behaviour strongly suggests that the Cluster 2 findings reflect true causal effects rather than statistical artifacts or selective reporting. Moreover, the genetic architecture revealed by LDSC analysis justified our focused approach over exploratory inclusion of all biomarkers.

By identifying these specific biological pathways, our analysis has provided clarity regarding the potential mechanisms of clozapine-induced metabolic dysfunction. This disentanglement not only advances the theoretical understanding but has potential implications for clinical monitoring and intervention strategies, allowing for more targeted approaches in managing cardiometabolic risks in clozapine-treated patients.

### 4.3. Clinical Implications and Future Directions

The identified candidate biomarkers offer potential tools for risk stratification and early intervention, but translation to clinical practice requires extensive validation. Prospective studies in cohorts of TRS patients initiating clozapine treatment are essential to determine whether these candidate biomarkers can accurately predict metabolic disturbances and, crucially, whether monitoring these biomarkers leads to improved clinical outcomes through early intervention.

The strong associations between liver function markers and metabolic outcomes suggest that enhanced monitoring protocols focusing on GGT and other hepatic markers could enable earlier intervention, though this hypothesis requires clinical validation. Similarly, endocrine and nutritional assessment, particularly vitamin D and testosterone levels, represents a candidate approach for targeted supplementation strategies that requires testing in clinical trials (74). The identification of excretory system markers as potential mediators of blood pressure effects suggests value in monitoring renal function parameters, though the clinical utility remains to be established.

The opposing causal effects observed for genetically correlated renal markers in MVMR analyses suggest that composite measures may provide more informative risk assessment than individual biomarkers considered in isolation. Specifically, the UACR, an established clinical marker of kidney damage in diabetes and chronic kidney disease (57–59), exemplifies how ratios can capture meaningful pathophysiological relationships that individual components cannot reveal independently. Our findings that creatinine associates with lower BMI while urinary albumin and sodium associate with higher BMI, despite their positive genetic correlations, parallel the clinical observation that UACR integrates information about both glomerular filtration and tubular damage. This pattern suggests that monitoring composite renal measures, rather than relying on any single marker, may enable more nuanced assessment of clozapine-induced cardiometabolic risk. Future validation studies should therefore evaluate whether ratios such as UACR, or novel composite measures derived from these genetically correlated markers, provide superior predictive performance compared to individual biomarkers for identifying patients at risk of metabolic dysfunction during clozapine treatment.

These findings also suggest potential for patient stratification prior to clozapine initiation that requires further investigation. Genetic testing for variants in the *CYP1A2* region, particularly rs2472297 which has demonstrated the strongest and most widespread associations with metabolic traits in our analyses, could potentially identify patients predisposed to altered clozapine metabolism and higher metabolic risk before treatment begins (27, 75, 76). However, the limited SNP-based heritability of several key biomarkers (h² range: 0.05–0.49; Supplementary Table 8) suggests that genetic prediction alone may have modest clinical utility, particularly for urinary markers like albumin excretion (h² = 0.08) and sodium (h² = 0.10) where direct biomarker measurement is likely more informative for risk stratification. The clinical value of genetic testing would need to be demonstrated through studies showing that genetic information leads to improved outcomes through more intensive monitoring or early co-administration of protective medications such as metformin or GLP-1 antagonists for high-risk patients (13). This pharmacogenomic approach could be supplemented by screening of key clozapine metabolizing genes to identify rare loss-of-function variants, though the clinical utility remains to be established (77).

Several high-priority research areas have emerged from these findings. Large-scale pharmacogenetic studies focusing specifically on clozapine metabolism are needed to better characterize metabolite-specific effects and strengthen genetic instruments. These studies should focus on characterizing the mechanisms underlying clozapine’s diabetogenic effects and their relationship to other metabolic disturbances.

Most critically, prospective clinical validation studies are needed to determine whether the identified genetic variants and candidate biomarkers can predict metabolic dysfunction in real-world clinical settings and whether this information can guide interventions that improve patient outcomes. Such studies should evaluate the incremental value of genetic and biomarker information beyond standard clinical risk factors.

By targeting specific biological pathways and leveraging candidate biomarkers, clinicians could potentially identify at-risk patients earlier and implement pathway-specific interventions before significant metabolic dysfunction occurs, though this approach requires clinical validation. This represents a potential advancement over current monitoring protocols, which primarily focus on detecting complications after they have developed, leading to potential safer prescribing of clozapine.

### 4.4. Strengths and Limitations

One of the difficulties in studying clozapine-induced metabolic effects is confounding by clinical variables like medication adherence, dosage variation, polypharmacy, and treatment duration. However, our MR approach minimizes these issues because the outcome datasets were derived from generally healthy populations, allowing isolation of genetic effects from treatment-related confounders (36, 78). While MR-Egger regression and other sensitivity analyses suggested minimal directional pleiotropy, residual horizontal pleiotropy remains a potential limitation, particularly for analyses with fewer genetic instruments (40). Additionally, the focus on European ancestry populations due to data availability limits generalizability across diverse ancestral groups. Genetic architecture and metabolic responses to clozapine may differ across populations (79).

These considerations should be viewed within the context of the study’s overall methodological robustness and the convergent evidence observed across multiple analytical approaches, which collectively strengthen confidence in the suggested possible causal pathways while acknowledging the inherent limitations of genetic epidemiological methods in fully characterizing complex pharmacological relationships.

### 4.5. Conclusions

This study employed a comprehensive genetic epidemiological approach to identify candidate biomarkers for clozapine-induced metabolic dysfunction, leveraging the strengths of MR to overcome confounding inherent in observational studies of clozapine-treated populations. Our findings indicate that genetically predicted higher plasma clozapine levels and clozapine-norclozapine ratio are associated with increased risk of T2D and elevated blood pressure, supporting a causal role for clozapine pharmacokinetics in metabolic dysfunction. Through phenome-wide scanning, colocalization, and subsequent MR analyses, we identified 16 candidate biomarkers spanning hepatic, renal, haematological, and endocrine systems that show evidence of associations with cardiometabolic outcomes. Notably, genetic correlation analyses revealed substantial independence among most candidate biomarkers, apart from a renal function cluster comprising creatinine, urinary sodium, and urinary albumin. MVMR demonstrated that these renal markers exert independent causal effects on metabolic outcomes despite their genetic correlations, suggesting that composite measures such as urinary albumin-to-creatinine ratio may provide superior risk assessment. While these findings require extensive clinical validation before implementation, they provide a rational foundation for developing biomarker-guided monitoring strategies that could enable earlier identification and intervention for patients at high risk of clozapine-induced metabolic complications, potentially improving the benefit-risk profile of this essential medication for TRS.

## Data Availability

All data produced in the present work are contained in the manuscript and/or supplementary files

https://opengwas.io/datasets/

https://walters.psycm.cf.ac.uk/

https://www.ebi.ac.uk/gwas/

## 5. Acknowledgements

All data used for this study is publicly available and accessible through source publications and/or genomics databases as disclosed in this manuscript.

## References

1. Yuen JWY, Kim DD, Procyshyn RM, Panenka WJ, Honer WG, Barr AM. A Focused Review of the Metabolic Side-Effects of Clozapine. Front Endocrinol (Lausanne). 2021;12:609240.

2. Pandey A, Kalita KN. Treatment-resistant schizophrenia: How far have we traveled? Front Psychiatry. 2022;13:994425.

3. Hurley K, O’Brien S, Halleran C, Byrne D, Foley E, Cunningham J, et al. Metabolic Syndrome in Adults Receiving Clozapine; The Need for Pharmacist Support. Pharmacy (Basel). 2023;11(1).

4. Myles N, Myles H, Xia S, Large M, Kisely S, Galletly C, et al. Meta-analysis examining the epidemiology of clozapine-associated neutropenia. Acta Psychiatr Scand. 2018;138(2):101–9.

5. Smith J, Griffiths LA, Band M, Horne D. Cardiometabolic Risk in First Episode Psychosis Patients. Front Endocrinol (Lausanne). 2020;11:564240.

6. Stancakova A, Laakso M. Genetics of metabolic syndrome. Rev Endocr Metab Disord. 2014;15(4):243–52.

7. Brown AE, Walker M. Genetics of Insulin Resistance and the Metabolic Syndrome. Curr Cardiol Rep. 2016;18(8):75.

8. Grover S, Hazari N, Chakrabarti S, Avasthi A. Metabolic Disturbances, Side Effect Profile and Effectiveness of Clozapine in Adolescents. Indian J Psychol Med. 2016;38(3):224–33.

9. Lamberti JS, Olson D, Crilly JF, Olivares T, Williams GC, Tu X, et al. Prevalence of the metabolic syndrome among patients receiving clozapine. Am J Psychiatry. 2006;163(7):1273–6.

10. Quek YF, See YM, Yee JY, Rekhi G, Ng BT, Tang C, et al. Metabolic syndrome and cardiovascular risk between clozapine and non-clozapine antipsychotic users with schizophrenia. Asian J Psychiatr. 2022;74:103192.

11. Walker AM, Lanza LL, Arellano F, Rothman KJ. Mortality in current and former users of clozapine. Epidemiology. 1997;8(6):671–7.

12. van der Weide K, Loovers H, Pondman K, Bogers J, van der Straaten T, Langemeijer E, et al. Genetic risk factors for clozapine-induced neutropenia and agranulocytosis in a Dutch psychiatric population. Pharmacogenomics J. 2017;17(5):471–8.

13. Per BL, Loeser S, Edwards S, Lee WS, Wilton LR, Clark SR. The Impact of Metformin on Weight and Waist Circumference in Patients Treated With Clozapine: A One-Year Retrospective Cohort Study. Acta Psychiatr Scand. 2025;151(6):719–30.

14. Rasmussen BB, Brix TH, Kyvik KO, Brosen K. The interindividual differences in the 3-demthylation of caffeine alias CYP1A2 is determined by both genetic and environmental factors. Pharmacogenetics. 2002;12(6):473–8.

15. Hernandez M, Cullell N, Cendros M, Serra-Llovich A, Arranz MJ. Clinical Utility and Implementation of Pharmacogenomics for the Personalisation of Antipsychotic Treatments. Pharmaceutics. 2024;16(2).

16. Gunes A, Melkersson KI, Scordo MG, Dahl ML. Association between HTR2C and HTR2A polymorphisms and metabolic abnormalities in patients treated with olanzapine or clozapine. J Clin Psychopharmacol. 2009;29(1):65–8.

17. Kang SH, Lee JI, Han HR, Soh M, Hong JP. Polymorphisms of the leptin and HTR2C genes and clozapine-induced weight change and baseline BMI in patients with chronic schizophrenia. Psychiatr Genet. 2014;24(6):249–56.

18. Pardinas AF, Kappel DB, Roberts M, Tipple F, Shitomi-Jones LM, King A, et al. Pharmacokinetics and pharmacogenomics of clozapine in an ancestrally diverse sample: a longitudinal analysis and genome-wide association study using UK clinical monitoring data. Lancet Psychiatry. 2023;10(3):209–19.

19. Lesche D, Mostafa S, Everall I, Pantelis C, Bousman CA. Impact of CYP1A2, CYP2C19, and CYP2D6 genotype– and phenoconversion-predicted enzyme activity on clozapine exposure and symptom severity. Pharmacogenomics J. 2020;20(2):192–201.

20. Menus A, Kiss A, Toth K, Sirok D, Deri M, Fekete F, et al. Association of clozapine-related metabolic disturbances with CYP3A4 expression in patients with schizophrenia. Sci Rep. 2020;10(1):21283.

21. Numata S, Umehara H, Ohmori T, Hashimoto R. Clozapine Pharmacogenetic Studies in Schizophrenia: Efficacy and Agranulocytosis. Front Pharmacol. 2018;9:1049.

22. Olsson E, Edman G, Bertilsson L, Hukic DS, Lavebratt C, Eriksson SV, et al. Genetic and Clinical Factors Affecting Plasma Clozapine Concentration. Prim Care Companion CNS Disord. 2015;17(1).

23. Erickson-Ridout KK, Sun D, Lazarus P. Glucuronidation of the second-generation antipsychotic clozapine and its active metabolite N-desmethylclozapine. Potential importance of the UGT1A1 A(TA)(7)TAA and UGT1A4 L48V polymorphisms. Pharmacogenet Genomics. 2012;22(8):561–76.

24. Bousman CA. Pharmacogenomics-informed clozapine therapy. The Lancet Psychiatry. 2023;10(3):160–2.

25. Spina E, Hiemke C, de Leon J. Assessing drug-drug interactions through therapeutic drug monitoring when administering oral second-generation antipsychotics. Expert Opin Drug Metab Toxicol. 2016;12(4):407–22.

26. Kohlrausch FB. Pharmacogenetics in schizophrenia: a review of clozapine studies. Braz J Psychiatry. 2013;35(3):305–17.

27. Doude van Troostwijk LJ, Koopmans RP, Vermeulen HD, Guchelaar HJ. CYP1A2 activity is an important determinant of clozapine dosage in schizophrenic patients. Eur J Pharm Sci. 2003;20(4-5):451–7.

28. Manu P, Dima L, Shulman M, Vancampfort D, De Hert M, Correll CU. Weight gain and obesity in schizophrenia: epidemiology, pathobiology, and management. Acta Psychiatr Scand. 2015;132(2):97–108.

29. Wu Y, Zeng J, Zhang F, Zhu Z, Qi T, Zheng Z, et al. Integrative analysis of omics summary data reveals putative mechanisms underlying complex traits. Nat Commun. 2018;9(1):918.

30. Richardson TG, Hemani G, Gaunt TR, Relton CL, Davey Smith G. A transcriptome-wide Mendelian randomization study to uncover tissue-dependent regulatory mechanisms across the human phenome. Nat Commun. 2020;11(1):185.

31. Emdin CA, Khera AV, Kathiresan S. Mendelian Randomization. JAMA. 2017;318(19):1925–6.

32. Huhn M, Nikolakopoulou A, Schneider-Thoma J, Krause M, Samara M, Peter N, et al. Comparative Efficacy and Tolerability of 32 Oral Antipsychotics for the Acute Treatment of Adults With Multi-Episode Schizophrenia: A Systematic Review and Network Meta-Analysis. Focus (Am Psychiatr Publ). 2020;18(4):443–55.

33. Burgess S, Davey Smith G, Davies NM, Dudbridge F, Gill D, Glymour MM, et al. Guidelines for performing Mendelian randomization investigations: update for summer 2023. Wellcome Open Res. 2019;4:186.

34. Stacey D, Chen L, Stanczyk PJ, Howson JMM, Mason AM, Burgess S, et al. Elucidating mechanisms of genetic cross-disease associations at the PROCR vascular disease locus. Nat Commun. 2022;13(1):1222.

35. Sanderson E, Spiller W, Bowden J. Testing and correcting for weak and pleiotropic instruments in two-sample multivariable Mendelian randomization. Stat Med. 2021;40(25):5434–52.

36. Pierce BL, Ahsan H, Vanderweele TJ. Power and instrument strength requirements for Mendelian randomization studies using multiple genetic variants. Int J Epidemiol. 2011;40(3):740–52.

37. Liu J, Cheng Y, Li M, Zhang Z, Li T, Luo XJ. Genome-wide Mendelian randomization identifies actionable novel drug targets for psychiatric disorders. Neuropsychopharmacology. 2023;48(2):270–80.

38. Burgess S, Small DS, Thompson SG. A review of instrumental variable estimators for Mendelian randomization. Stat Methods Med Res. 2017;26(5):2333–55.

39. Bowden J, Davey Smith G, Haycock PC, Burgess S. Consistent Estimation in Mendelian Randomization with Some Invalid Instruments Using a Weighted Median Estimator. Genet Epidemiol. 2016;40(4):304–14.

40. Verbanck M, Chen CY, Neale B, Do R. Detection of widespread horizontal pleiotropy in causal relationships inferred from Mendelian randomization between complex traits and diseases. Nat Genet. 2018;50(5):693–8.

41. Brion MJ, Shakhbazov K, Visscher PM. Calculating statistical power in Mendelian randomization studies. Int J Epidemiol. 2013;42(5):1497–501.

42. Winkler TW, Day FR, Croteau-Chonka DC, Wood AR, Locke AE, Magi R, et al. Quality control and conduct of genome-wide association meta-analyses. Nat Protoc. 2014;9(5):1192–212.

43. Sanderson E, Glymour MM, Holmes MV, Kang H, Morrison J, Munafo MR, et al. Mendelian randomization. Nat Rev Methods Primers. 2022;2.

44. Wickham H. ggplot2: Springer-Verlag New York; 2016.

45. Hemani G, Zheng J, Elsworth B, Wade KH, Haberland V, Baird D, et al. The MR-Base platform supports systematic causal inference across the human phenome. eLife. 2018;7.

46. Okhuijsen-Pfeifer C, van der Horst MZ, Bousman CA, Lin B, van Eijk KR, Ripke S, et al. Genome-wide association analyses of symptom severity among clozapine-treated patients with schizophrenia spectrum disorders. Transl Psychiatry. 2022;12(1):145.

47. Zheng J, Haberland V, Baird D, Walker V, Haycock PC, Hurle MR, et al. Phenome-wide Mendelian randomization mapping the influence of the plasma proteome on complex diseases. Nat Genet. 2020;52(10):1122–31.

48. Machiela MJ, Chanock SJ. LDlink: a web-based application for exploring population-specific haplotype structure and linking correlated alleles of possible functional variants. Bioinformatics. 2015;31(21):3555–7.

49. Dennis JK, Sealock JM, Straub P, Lee YH, Hucks D, Actkins K, et al. Clinical laboratory test-wide association scan of polygenic scores identifies biomarkers of complex disease. Genome Med. 2021;13(1):6.

50. Bastarache L, Denny JC, Roden DM. Phenome-Wide Association Studies. JAMA. 2022;327(1):75–6.

51. Denny JC, Bastarache L, Roden DM. Phenome-Wide Association Studies as a Tool to Advance Precision Medicine. Annu Rev Genomics Hum Genet. 2016;17:353–73.

52. Liu B, Gloudemans MJ, Rao AS, Ingelsson E, Montgomery SB. Abundant associations with gene expression complicate GWAS follow-up. Nat Genet. 2019;51(5):768–9.

53. Giambartolomei C, Vukcevic D, Schadt EE, Franke L, Hingorani AD, Wallace C, et al. Bayesian test for colocalisation between pairs of genetic association studies using summary statistics. PLoS Genet. 2014;10(5):e1004383.

54. Wallace C. Eliciting priors and relaxing the single causal variant assumption in colocalisation analyses. PLoS Genet. 2020;16(4):e1008720.

55. Foley CN, Staley JR, Breen PG, Sun BB, Kirk PDW, Burgess S, et al. A fast and efficient colocalization algorithm for identifying shared genetic risk factors across multiple traits. Nat Commun. 2021;12(1):764.

56. Robinson JW, Hemani G, Babaei MS, Huang Y, Baird DA, Tsai EA, et al. 2022.

57. American Diabetes Association Professional Practice C. 11. Chronic Kidney Disease and Risk Management: Standards of Care in Diabetes-2025. Diabetes Care. 2025;48(1 Suppl 1):S239–S51.

58. Tang WH, Hung WC, Wang CP, Wu CC, Hsuan CF, Yu TH, et al. The Lower Limit of Reference of Urinary Albumin/Creatinine Ratio and the Risk of Chronic Kidney Disease Progression in Patients With Type 2 Diabetes Mellitus. Front Endocrinol (Lausanne). 2022;13:858267.

59. Tangri N, Singh R, Chen Y, Betts KA, Farag YM, Beeman S, et al. Change in urine albumin-to-creatinine ratio and clinical outcomes in patients with chronic kidney disease and type 2 diabetes. BMJ Open Diabetes Res Care. 2025;13(5).

60. Newcomer JW. Second-generation (atypical) antipsychotics and metabolic effects: a comprehensive literature review. CNS Drugs. 2005;19 Suppl 1:1–93.

61. Seitz HK, Stickel F. Acetaldehyde as an underestimated risk factor for cancer development: role of genetics in ethanol metabolism. Genes Nutr. 2010;5(2):121–8.

62. Salaspuro M. Acetaldehyde and gastric cancer. J Dig Dis. 2011;12(2):51–9.

63. Tan MSA, Honarparvar F, Falconer JR, Parekh HS, Pandey P, Siskind DJ. A systematic review and meta-analysis of the association between clozapine and norclozapine serum levels and peripheral adverse drug reactions. Psychopharmacology (Berl). 2021;238(3):615–37.

64. Brooks PJ, Goldman D, Li TK. Alleles of alcohol and acetaldehyde metabolism genes modulate susceptibility to oesophageal cancer from alcohol consumption. Hum Genomics. 2009;3(2):103–5.

65. Leung JG, Zhang L, Markota M, Ellingrod VL, Gerberi DJ, Bishop JR. A systematic review of clozapine-associated inflammation and related monitoring. Pharmacotherapy. 2023;43(12):1364–96.

66. Thakur S, Kumar V, Das R, Sharma V, Mehta DK. Biomarkers of Hepatic Toxicity: An Overview. Curr Ther Res Clin Exp. 2024;100:100737.

67. Alberich S, Fernandez-Sevillano J, Gonzalez-Ortega I, Usall J, Saenz M, Gonzalez-Fraile E, et al. A systematic review of sex-based differences in effectiveness and adverse effects of clozapine. Psychiatry Res. 2019;280:112506.

68. Suzumoto Y, Zucaro L, Iervolino A, Capasso G. Kidney and blood pressure regulation-latest evidence for molecular mechanisms. Clin Kidney J. 2023;16(6):952–64.

69. Sedaghat S, Pazoki R, Uitterlinden AG, Hofman A, Stricker BH, Ikram MA, et al. Association of uric acid genetic risk score with blood pressure: the Rotterdam study. Hypertension. 2014;64(5):1061–6.

70. Lanaspa MA, Andres-Hernando A, Kuwabara M. Uric acid and hypertension. Hypertens Res. 2020;43(8):832–4.

71. Sanchez-Lozada LG, Rodriguez-Iturbe B, Kelley EE, Nakagawa T, Madero M, Feig DI, et al. Uric Acid and Hypertension: An Update With Recommendations. American Journal of Hypertension. 2020;33(7):583–94.

72. Wang Z, Li P, Chi D, Wu T, Mei Z, Cui G. Association between C-reactive protein and risk of schizophrenia: An updated meta-analysis. Oncotarget. 2017;8(43):75445–54.

73. Prins BP, Abbasi A, Wong A, Vaez A, Nolte I, Franceschini N, et al. Investigating the Causal Relationship of C-Reactive Protein with 32 Complex Somatic and Psychiatric Outcomes: A Large-Scale Cross-Consortium Mendelian Randomization Study. PLoS Med. 2016;13(6):e1001976.

74. Hypponen E, Boucher BJ. Adiposity, vitamin D requirements, and clinical implications for obesity-related metabolic abnormalities. Nutr Rev. 2018;76(9):678–92.

75. Viikki M, Kampman O, Seppala N, Mononen N, Lehtimaki T, Leinonen E. CYP1A2 polymorphism – 1545C > T (rs2470890) is associated with increased side effects to clozapine. BMC Psychiatry. 2014;14:50.

76. Mihara K, Suzuki A, Kondo T, Yasui N, Furukori H, Nagashima U, et al. Effect of a genetic polymorphism of CYP1A2 inducibility on the steady state plasma concentrations of haloperidol and reduced haloperidol in Japanese patients with schizophrenia. Ther Drug Monit. 2000;22(3):245–9.

77. Zhou Y, Lauschke VM. The genetic landscape of major drug metabolizing cytochrome P450 genes-an updated analysis of population-scale sequencing data. Pharmacogenomics J. 2022;22(5-6):284–93.

78. Khasawneh LQ, Al-Mahayri ZN, Ali BR. Mendelian randomization in pharmacogenomics: The unforeseen potentials. Biomed Pharmacother. 2022;150:112952.

79. Ju D, Hui D, Hammond DA, Wonkam A, Tishkoff SA. Importance of Including Non-European Populations in Large Human Genetic Studies to Enhance Precision Medicine. Annu Rev Biomed Data Sci. 2022;5:321–39.

