## Supplementary Figures for "Genetic Epidemiological Pipeline Identifies Candidate Markers of Clozapine-Induced Metabolic Dysfunction Revealing Potential Avenues for Precision Clozapine Prescription"

#### Supplementary Figures 1 – Directive Acyclic Graphs

Directive acyclic graphs (DAGs) representing the assumptions of valid causal inference in Mendelian randomization: the genetic variant must be strongly associated with the exposure (relevance assumption), it must be independent of the outcome except through the exposure (independence assumption), and it must affect the outcome only through the exposure (exclusion restriction).

*Supplementary Figures 1. 1 – Clozapine Plasma Traits Effects on Cardiometabolic Traits*

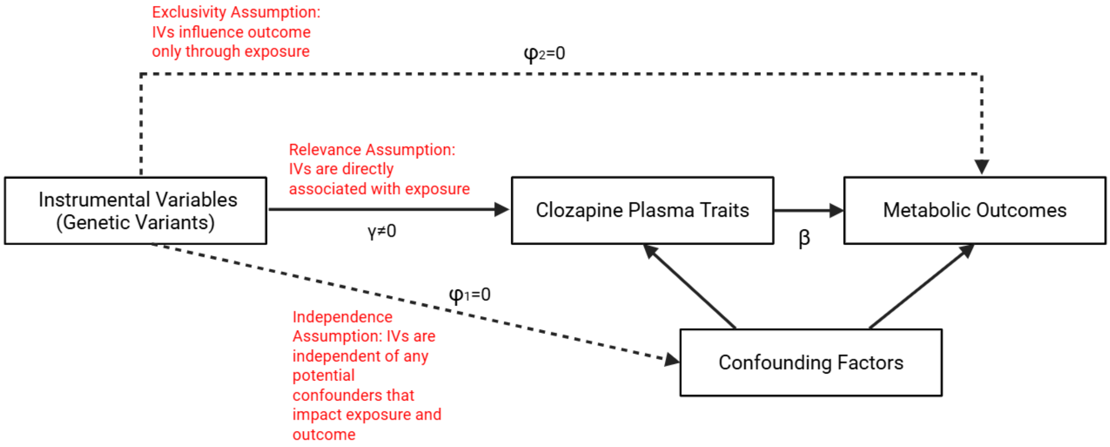

*Supplementary Figures 1. 2 – Cardiometabolic Effects on Clozapine Plasma Traits*

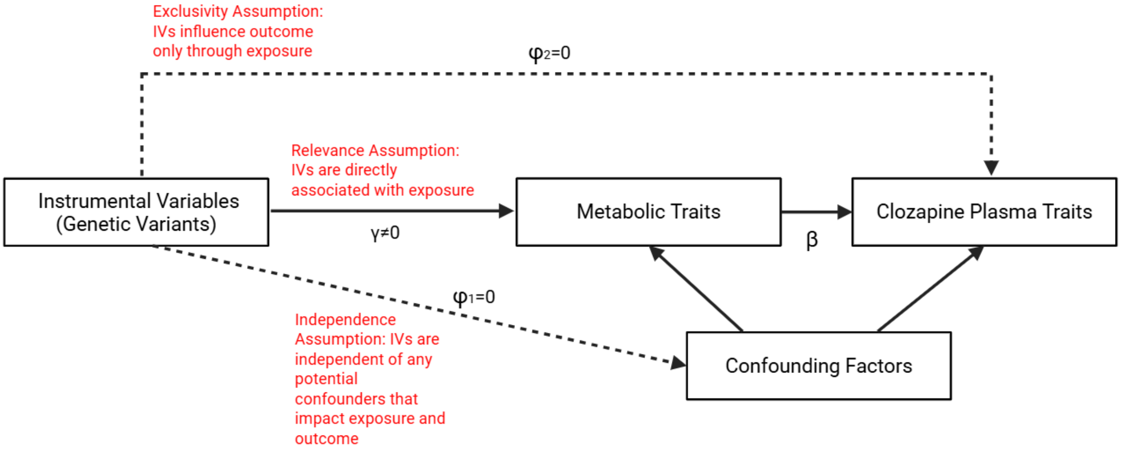

*Supplementary Figures 1. 3 – Candidate Biomarkers Effects on Cardiometabolic Traits*

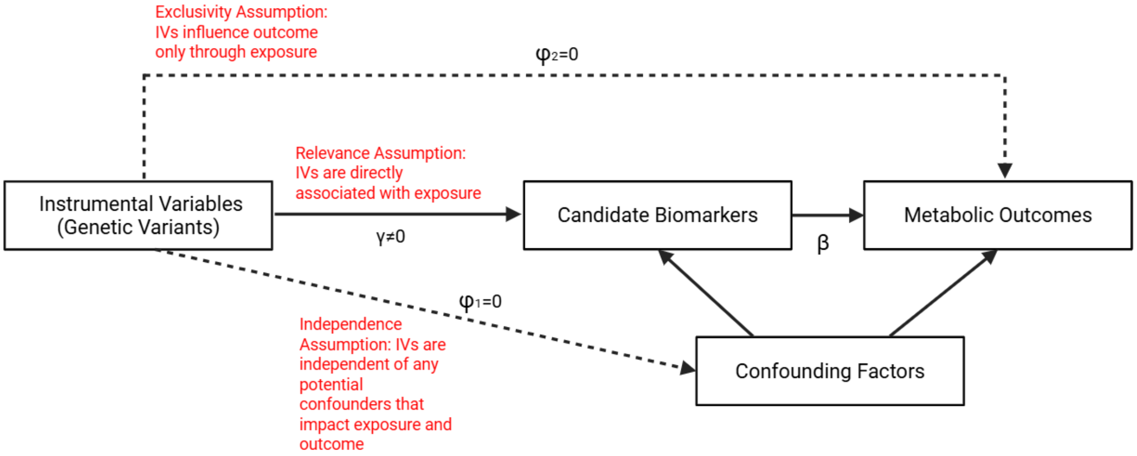

*Supplementary Figures 1. 4 – Cardiometabolic Effects on Candidate Biomarkers*

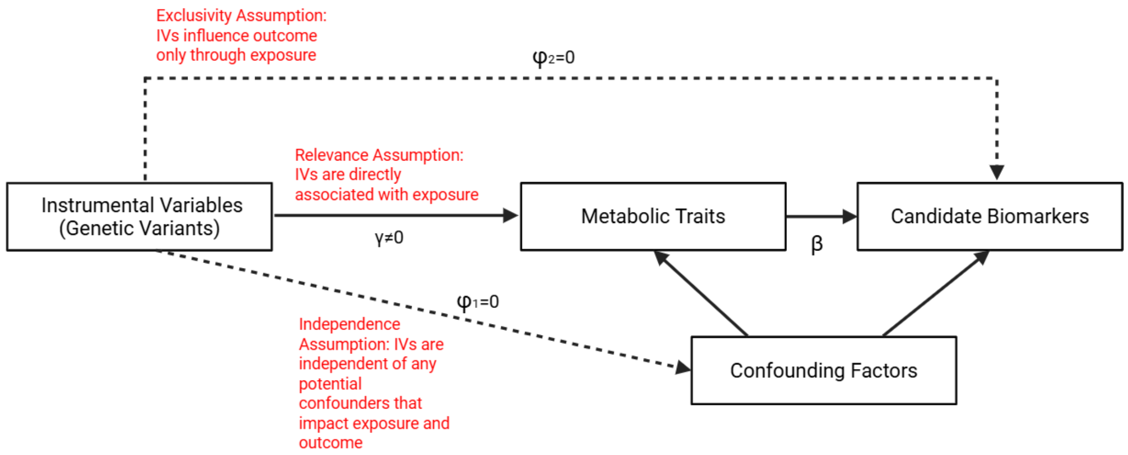

#### Supplementary Figures 2 – Bidirectional Effects of Clozapine Plasma Traits on Cardiometabolic Outcomes

Analysis results from linkage-disequilibrium-aware bidirectional Mendelian randomization of clozapine on cardiometabolic outcomes. Forest plots demonstrate the causal estimate of the exposure ($\boldsymbol{\beta}$) on $\boldsymbol{n}$ outcomes with $\mathbf{95\%}$ $\boldsymbol{CIs}$ and $\boldsymbol{Pvals}$. Significance threshold is adjusted for multiple testing (Bonferroni Correction) ($\boldsymbol{Pval}$ $(\boldsymbol{adj}) = \mathbf{0.05}/\boldsymbol{n}$). Scatterplots depict sensitivity analyses results including MR-Egger and weighted median to test for directional pleiotropy and causal estimate assessment under invalid instruments respectively. Leave-one-out plots demonstrate the robustness of causal inference by testing if one genetic variant is driving the observed association. Raw results for direct and inverse linkage-disequilibrium-aware bidirectional Mendelian randomization are available in Supplementary Table 1 and 2. Information about the Mendelian randomization analysis can be found in the Supplementary Methods.

*Supplementary Figures 2. 1 – Forest Plots – Clozapine Plasma Traits Effects on Cardiometabolic Outcomes*

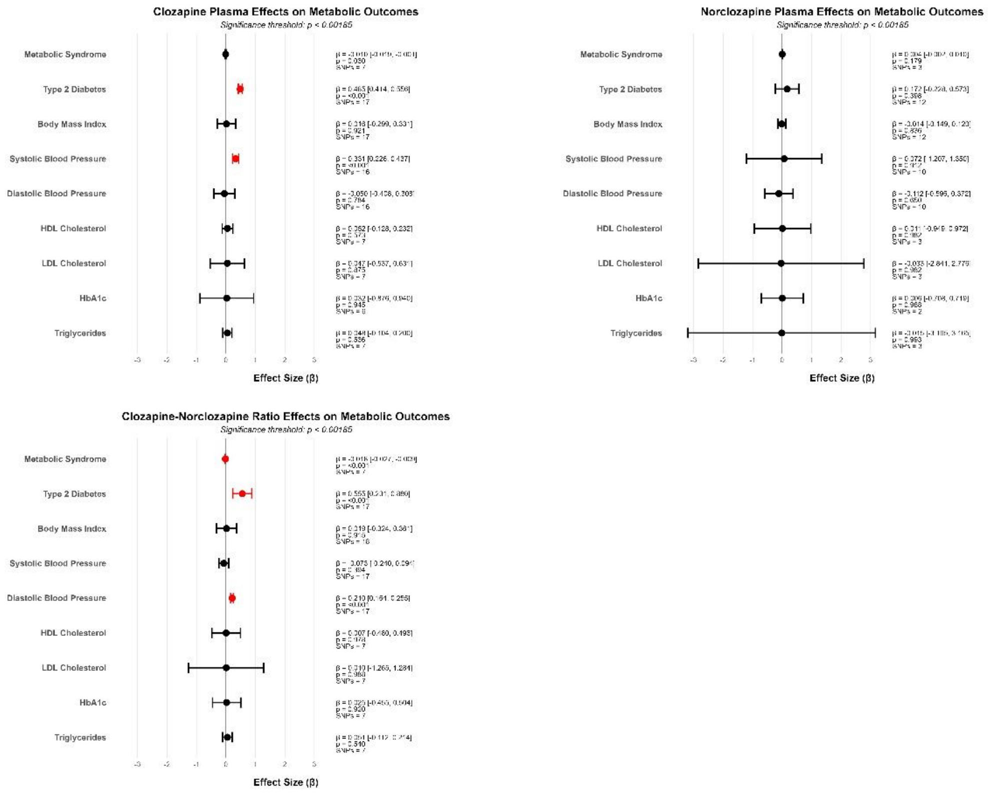

#### *Supplementary Figures 2. 2 – Scatterplots – Clozapine Plasma Traits Effects on Cardiometabolic Outcomes*

##### Clozapine Plasma

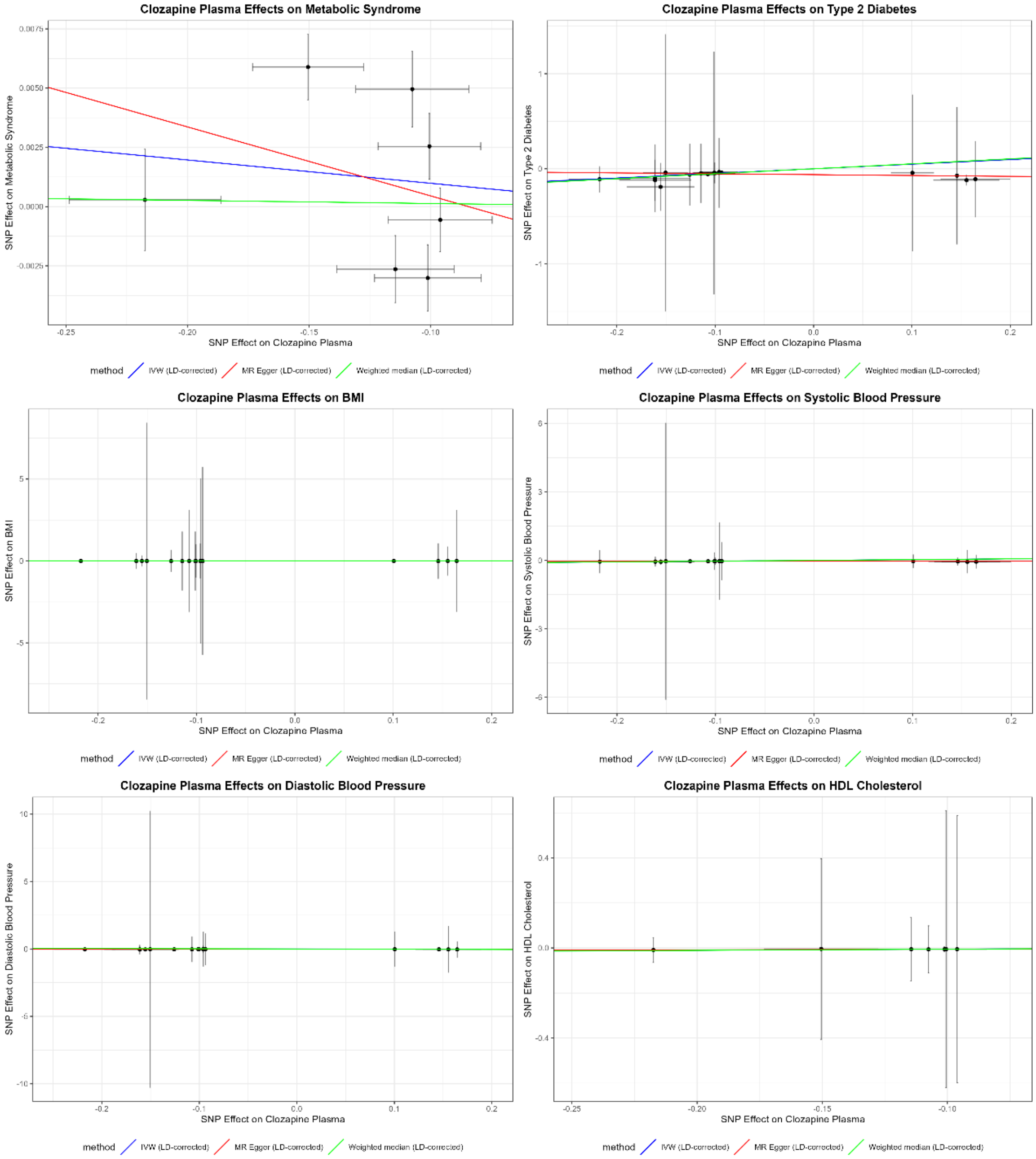

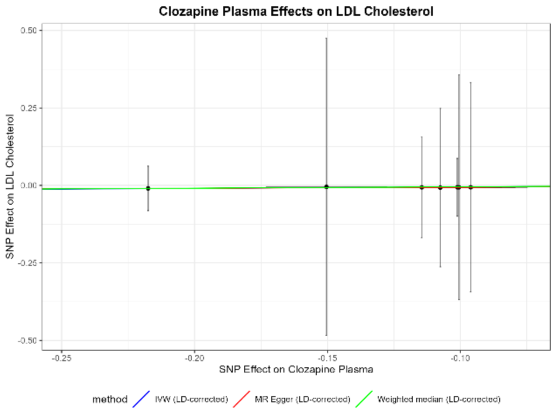

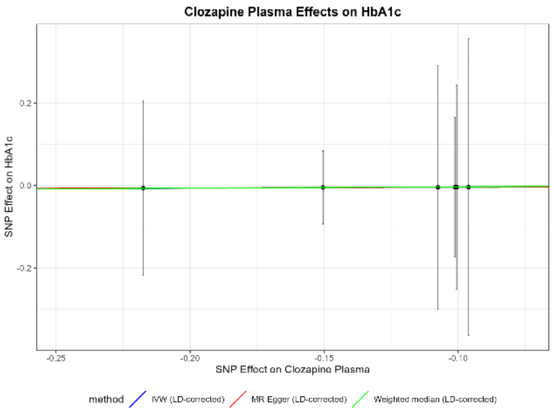

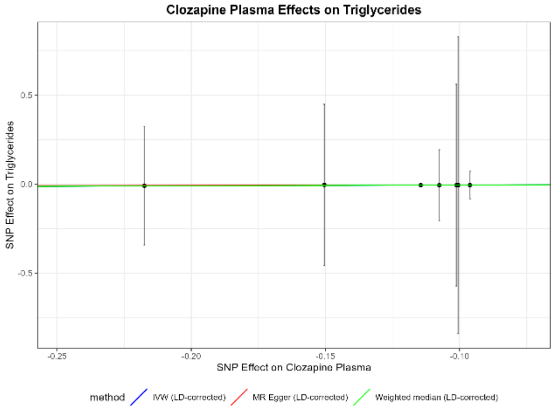

#### Norclozapine Plasma

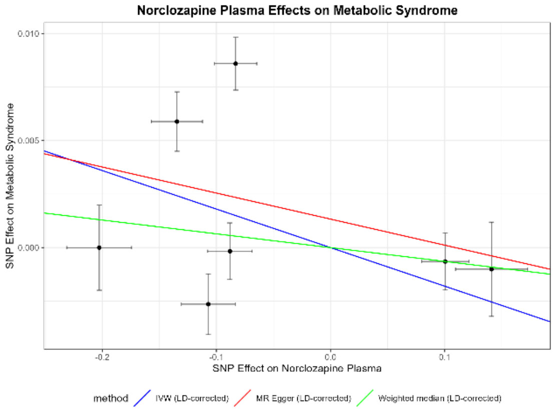

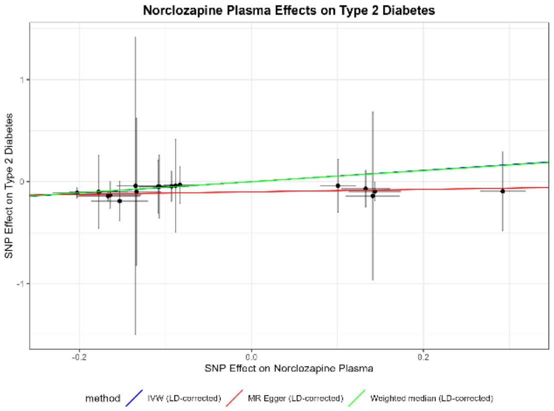

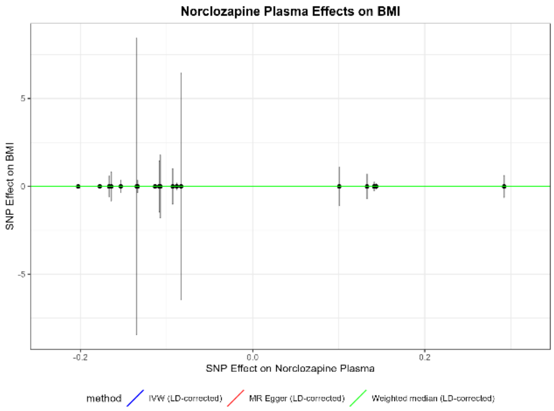

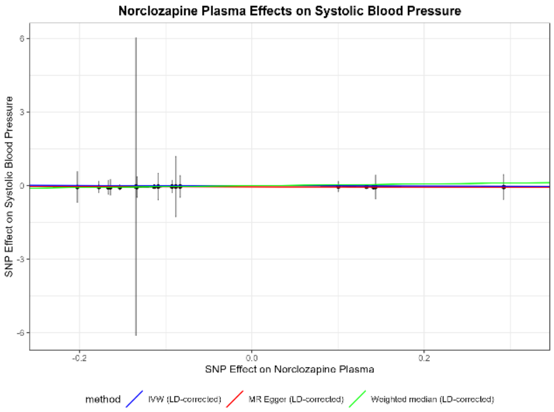

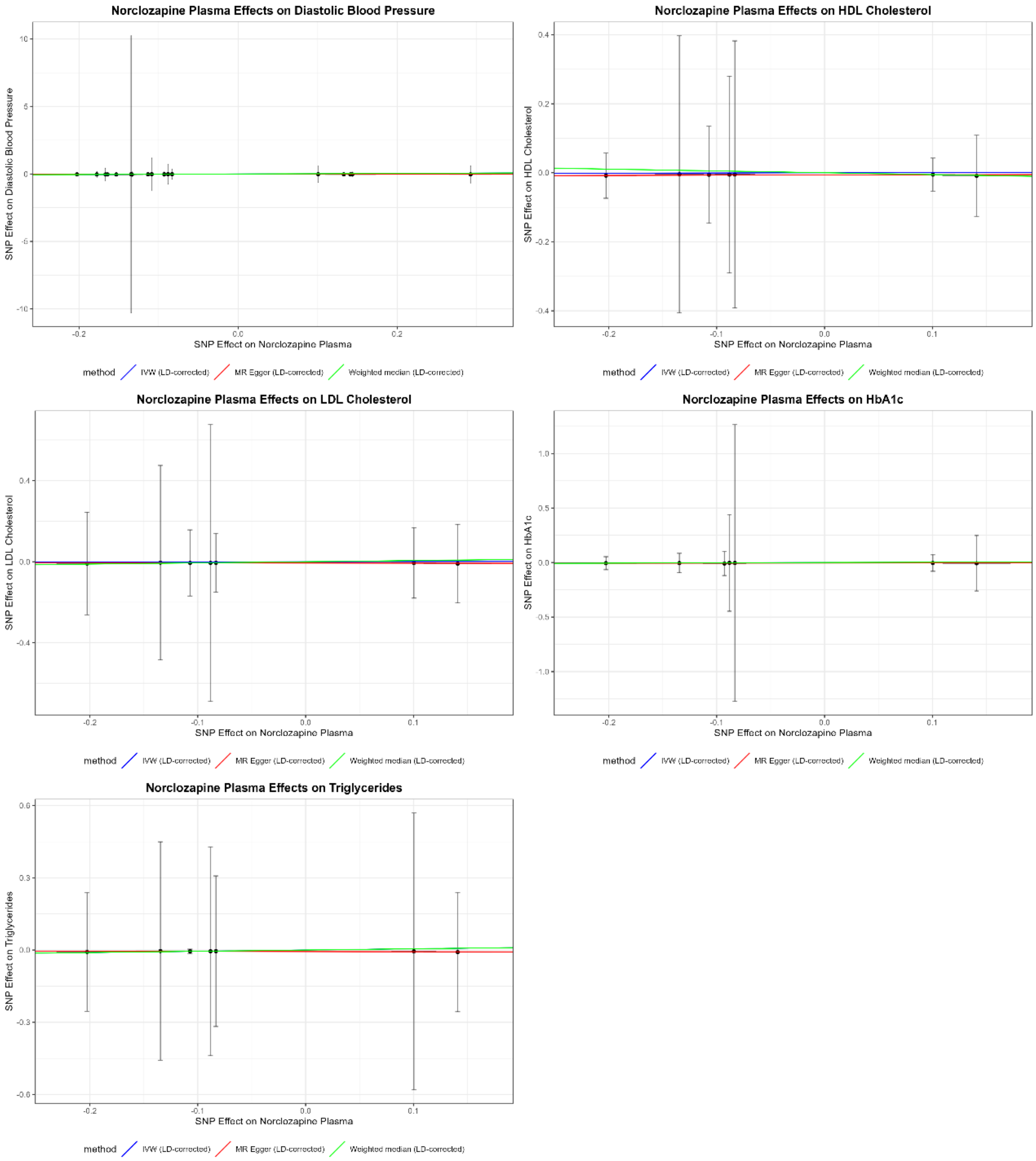
Norclozapine Plasma Effects on Diastolic Blood Pressure
SNP Effect on Diastolic Blood Pressure
SNP Effect on Norclozapine Plasma
method
IVW (LD-corrected)
MR Egger (LD-corrected)
Weighted median (LD-corrected)
Norclozapine Plasma Effects on HDL Cholesterol
SNP Effect on HDL Cholesterol
SNP Effect on Norclozapine Plasma
method
IVW (LD-corrected)
MR Egger (LD-corrected)
Weighted median (LD-corrected)
Norclozapine Plasma Effects on LDL Cholesterol
SNP Effect on LDL Cholesterol
SNP Effect on Norclozapine Plasma
method
IVW (LD-corrected)
MR Egger (LD-corrected)
Weighted median (LD-corrected)
Norclozapine Plasma Effects on HbA1c
SNP Effect on HbA1c
SNP Effect on Norclozapine Plasma
method
IVW (LD-corrected)
MR Egger (LD-corrected)
Weighted median (LD-corrected)
Norclozapine Plasma Effects on Triglycerides
SNP Effect on Triglycerides
SNP Effect on Norclozapine Plasma
method
IVW (LD-corrected)
MR Egger (LD-corrected)
Weighted median (LD-corrected)

### Clozapine-Norclozapine Plasma Ratio

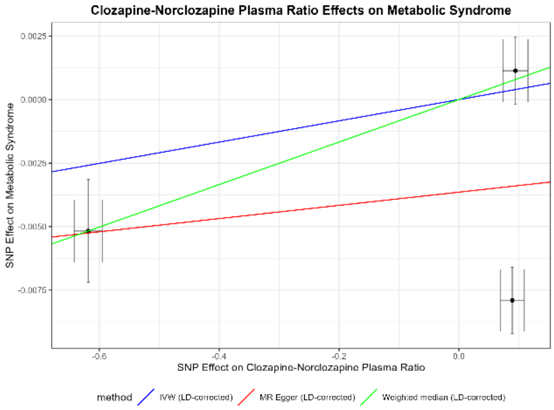

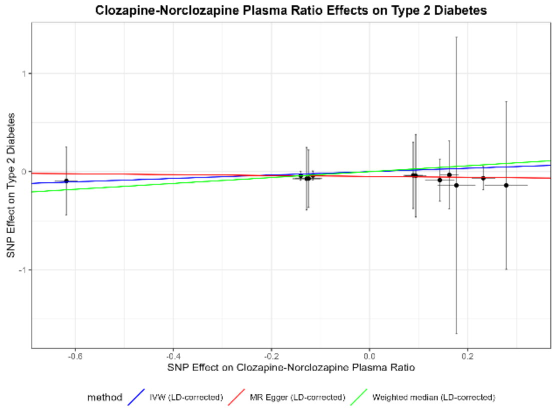

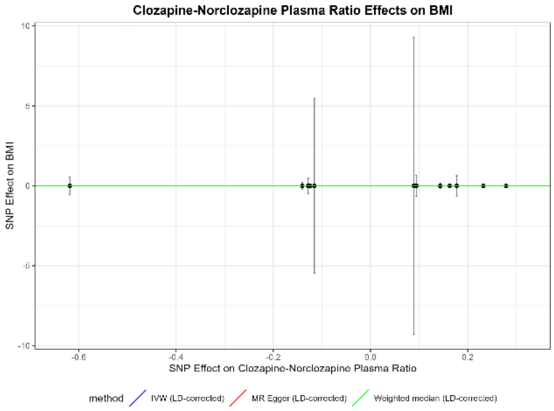

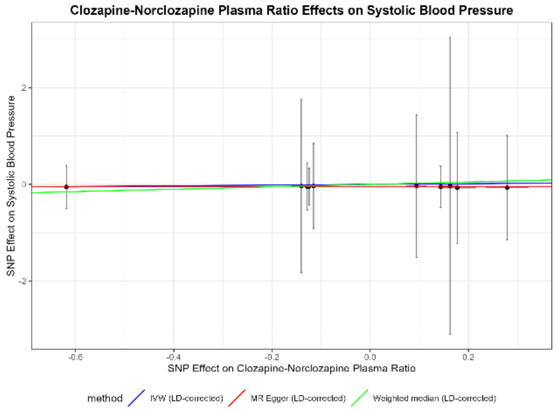

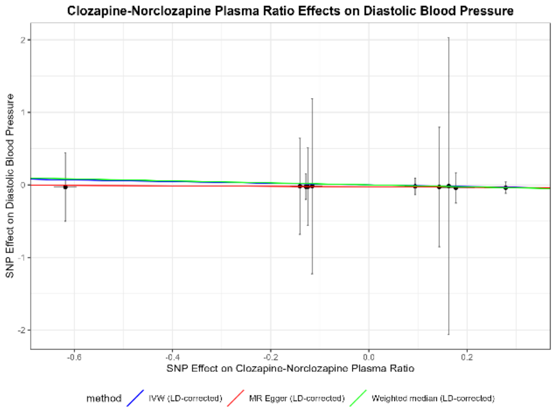

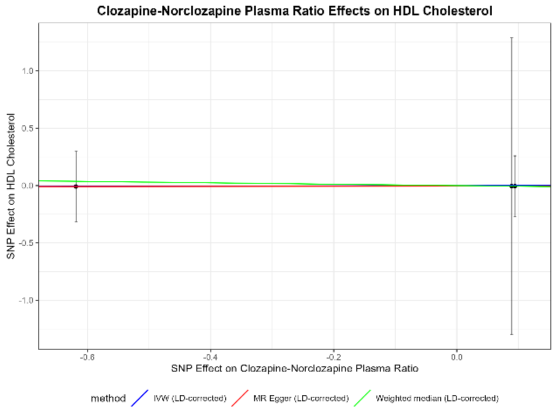

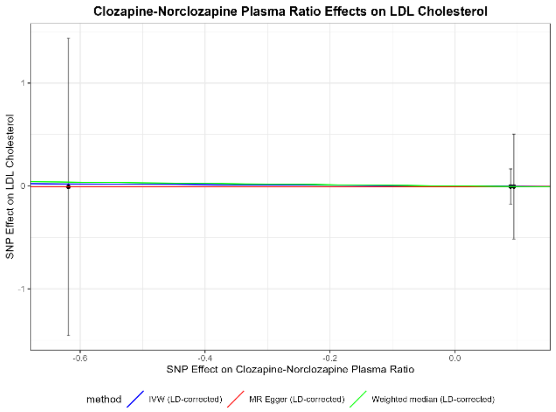
Clozapine-Norclozapine Plasma Ratio Effects on LDL Cholesterol
SNP Effect on LDL Cholesterol
1
0
-1
-0.6
-0.4
-0.2
0.0
SNP Effect on Clozapine-Norclozapine Plasma Ratio
method
IVW (LD-corrected)
MR Egger (LD-corrected)
Weighted median (LD-corrected)

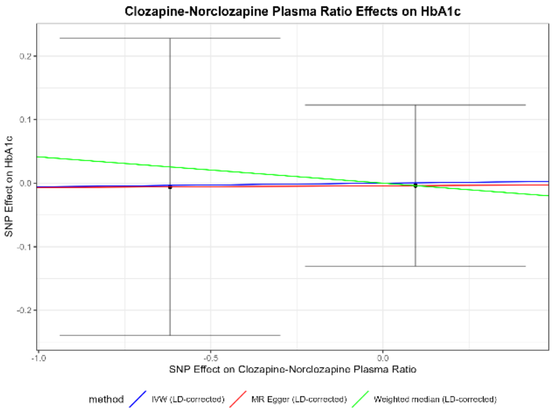
Clozapine-Norclozapine Plasma Ratio Effects on HbA1c
SNP Effect on HbA1c
0.2
0.1
0.0
-0.1
-0.2
-1.0
-0.5
0.0
SNP Effect on Clozapine-Norclozapine Plasma Ratio
method
IVW (LD-corrected)
MR Egger (LD-corrected)
Weighted median (LD-corrected)

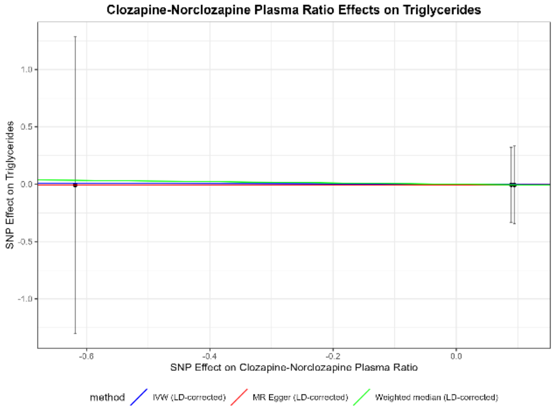
Clozapine-Norclozapine Plasma Ratio Effects on Triglycerides
SNP Effect on Triglycerides
1.0
0.5
0.0
-0.5
-1.0
-0.6
-0.4
-0.2
0.0
SNP Effect on Clozapine-Norclozapine Plasma Ratio
method
IVW (LD-corrected)
MR Egger (LD-corrected)
Weighted median (LD-corrected)

*Supplementary Figures 2. 3 – Scatterplots – Clozapine Plasma Traits Effects on Cardiometabolic Traits*

#### Clozapine Plasma

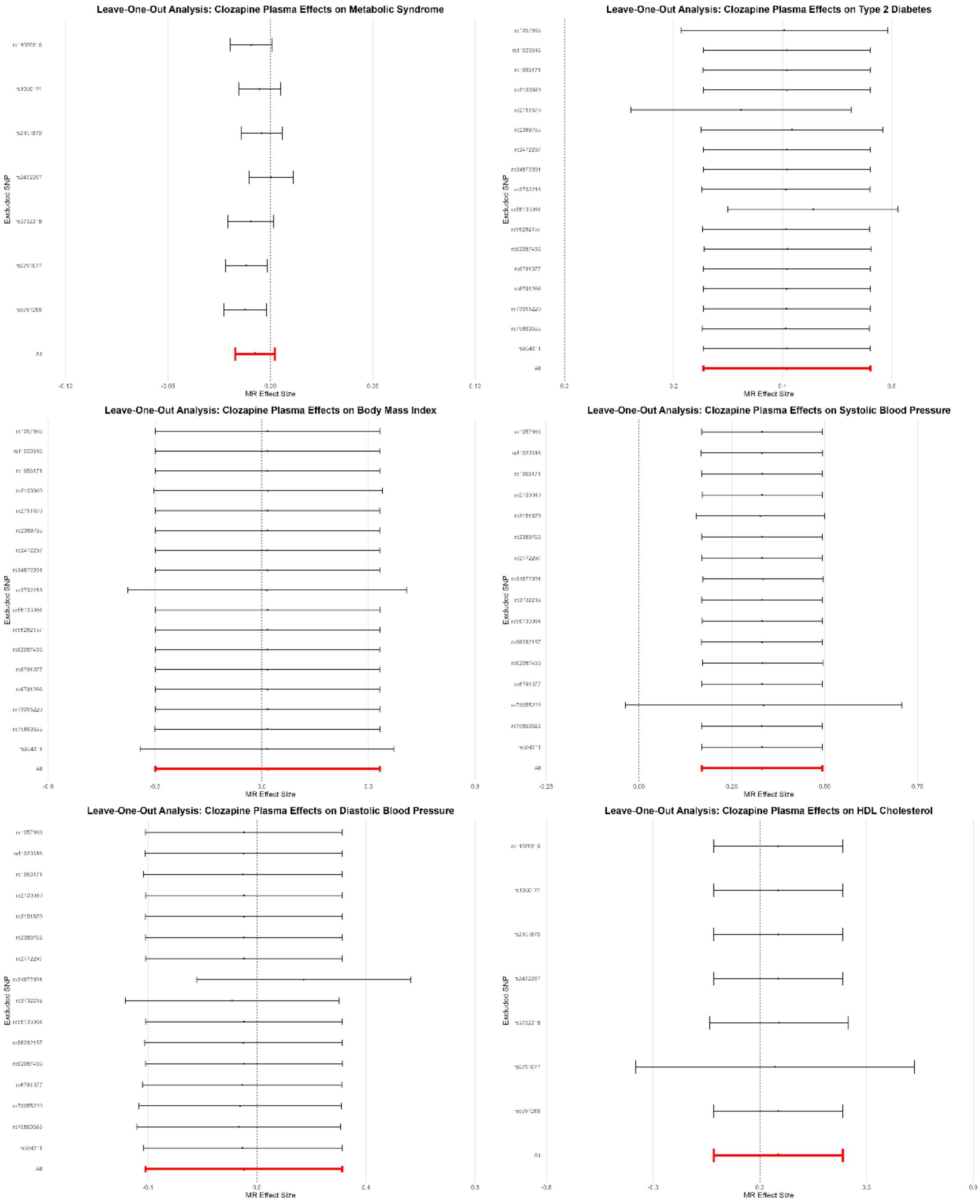

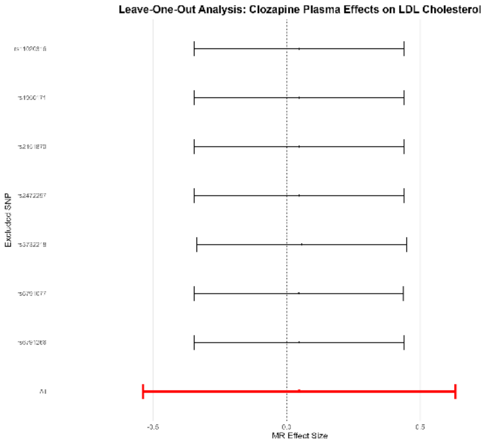

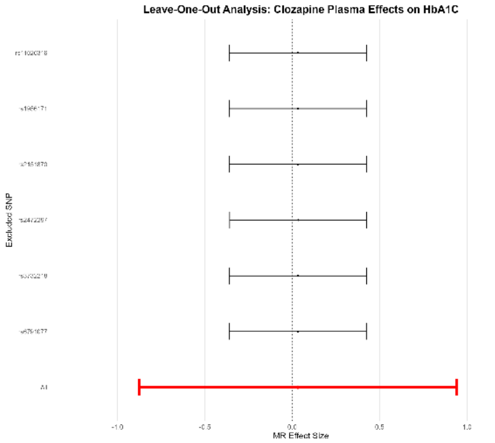

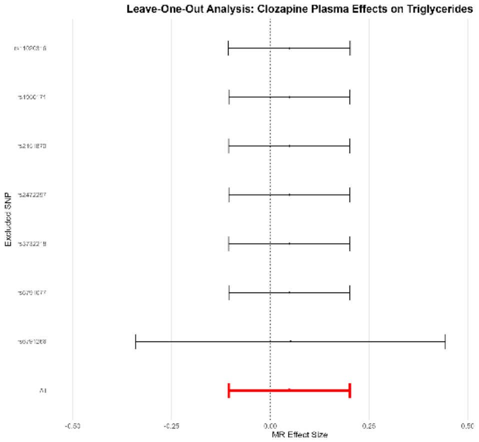

#### Norclozapine Plasma

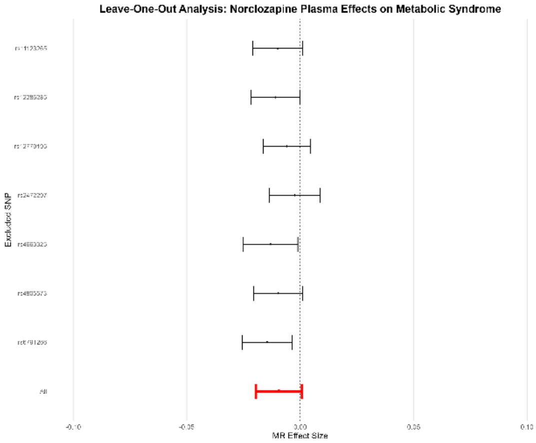

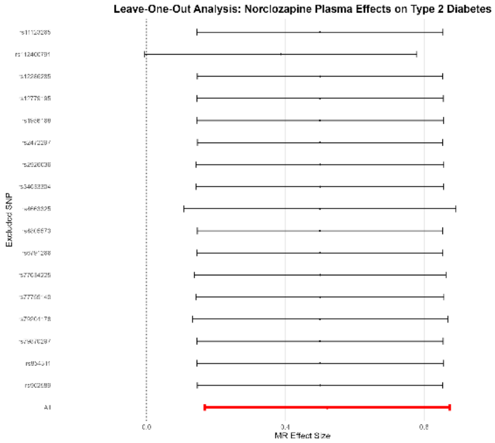

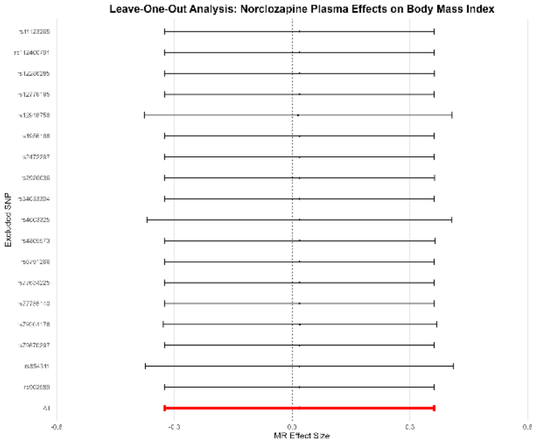

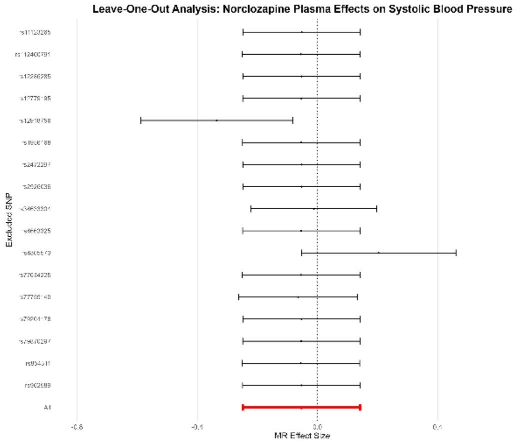

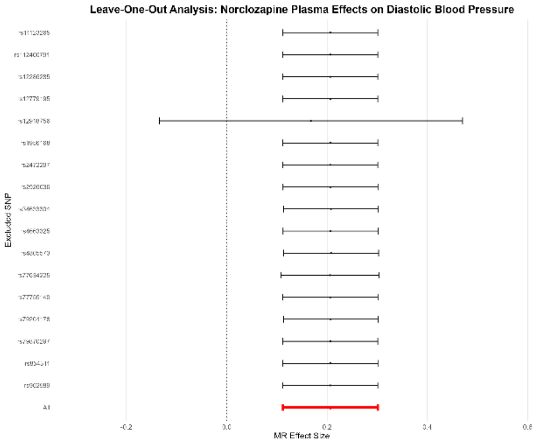
Leave-One-Out Analysis: Norclozapine Plasma Effects on Diastolic Blood Pressure
Excluded SNP
All
MR Effect Size

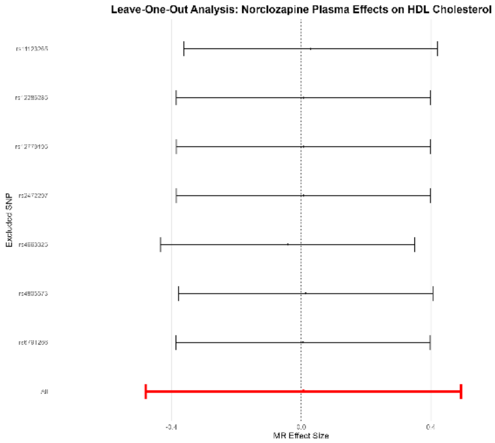
Leave-One-Out Analysis: Norclozapine Plasma Effects on HDL Cholesterol
Excluded SNP
All
MR Effect Size

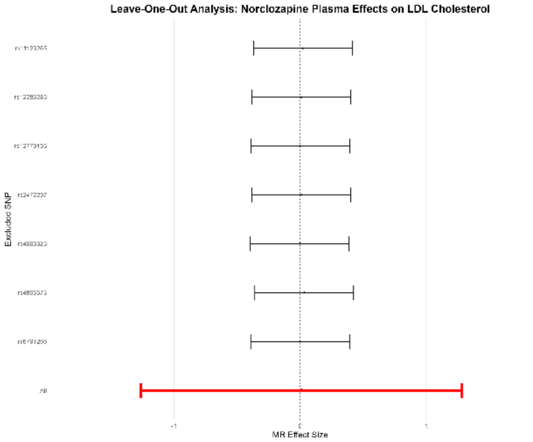
Leave-One-Out Analysis: Norclozapine Plasma Effects on LDL Cholesterol
Excluded SNP
All
MR Effect Size

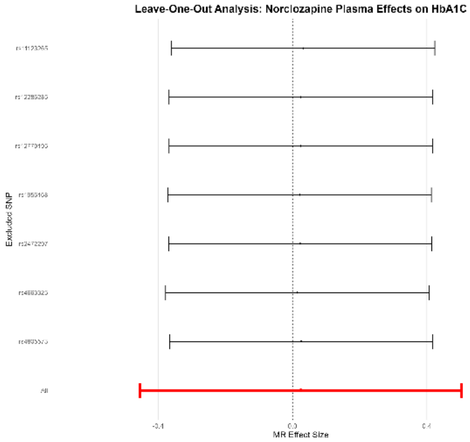
Leave-One-Out Analysis: Norclozapine Plasma Effects on HbA1C
Excluded SNP
All
MR Effect Size

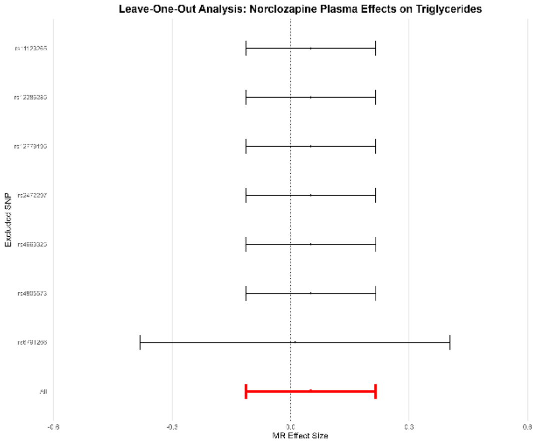
Leave-One-Out Analysis: Norclozapine Plasma Effects on Triglycerides
Excluded SNP
All
MR Effect Size

#### Clozapine-Norclozapine Plasma Ratio

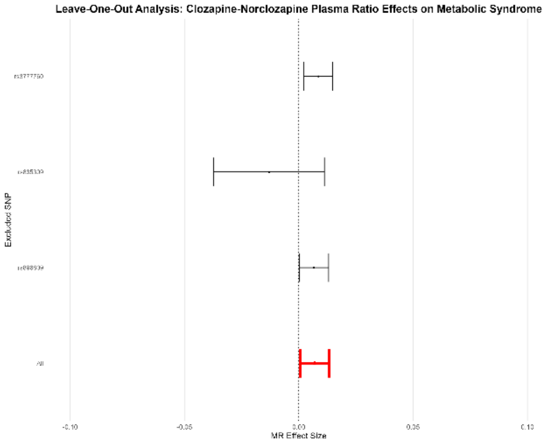

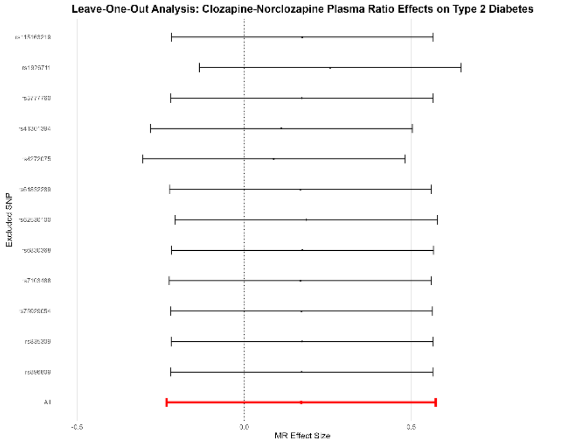

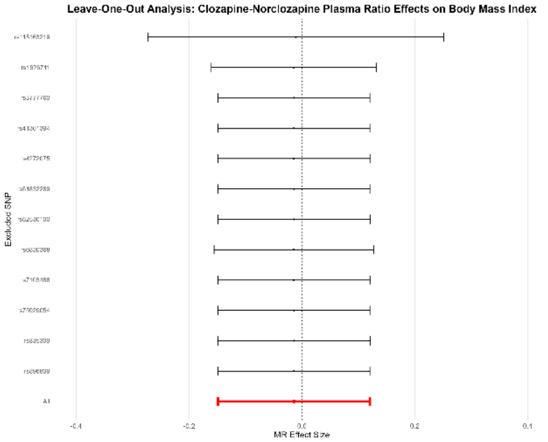

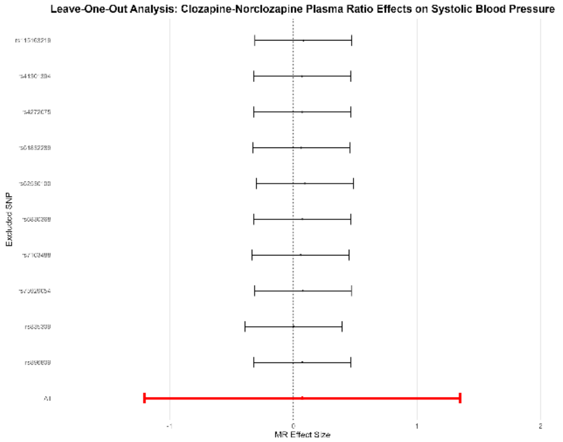

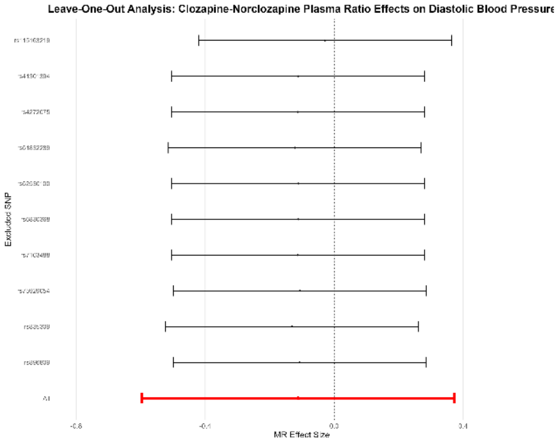

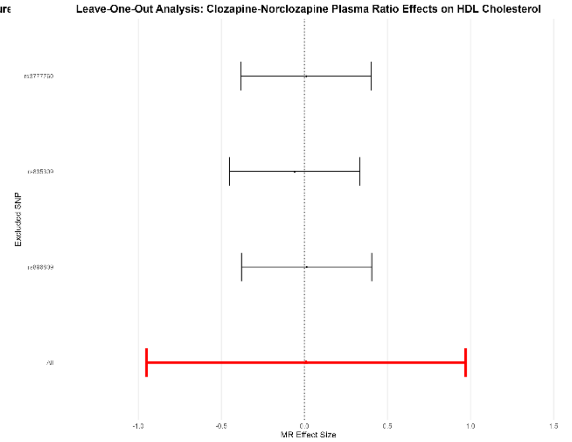

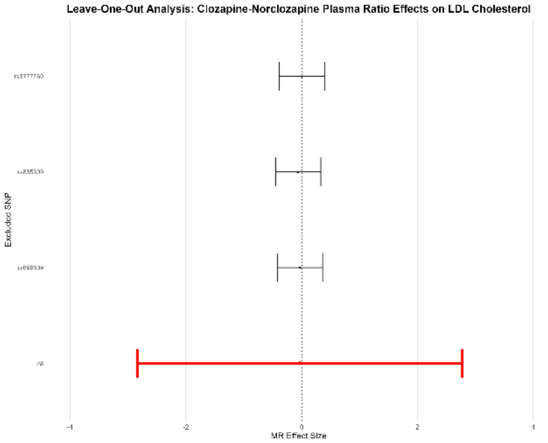

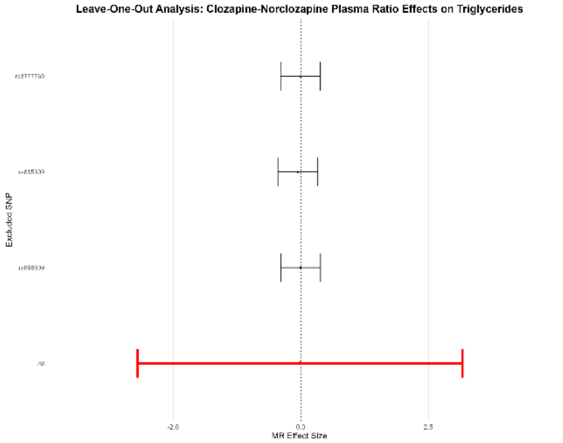

*Supplementary Figures 2. 4 – Forest Plots – Cardiometabolic Effects on Clozapine Plasma Traits*

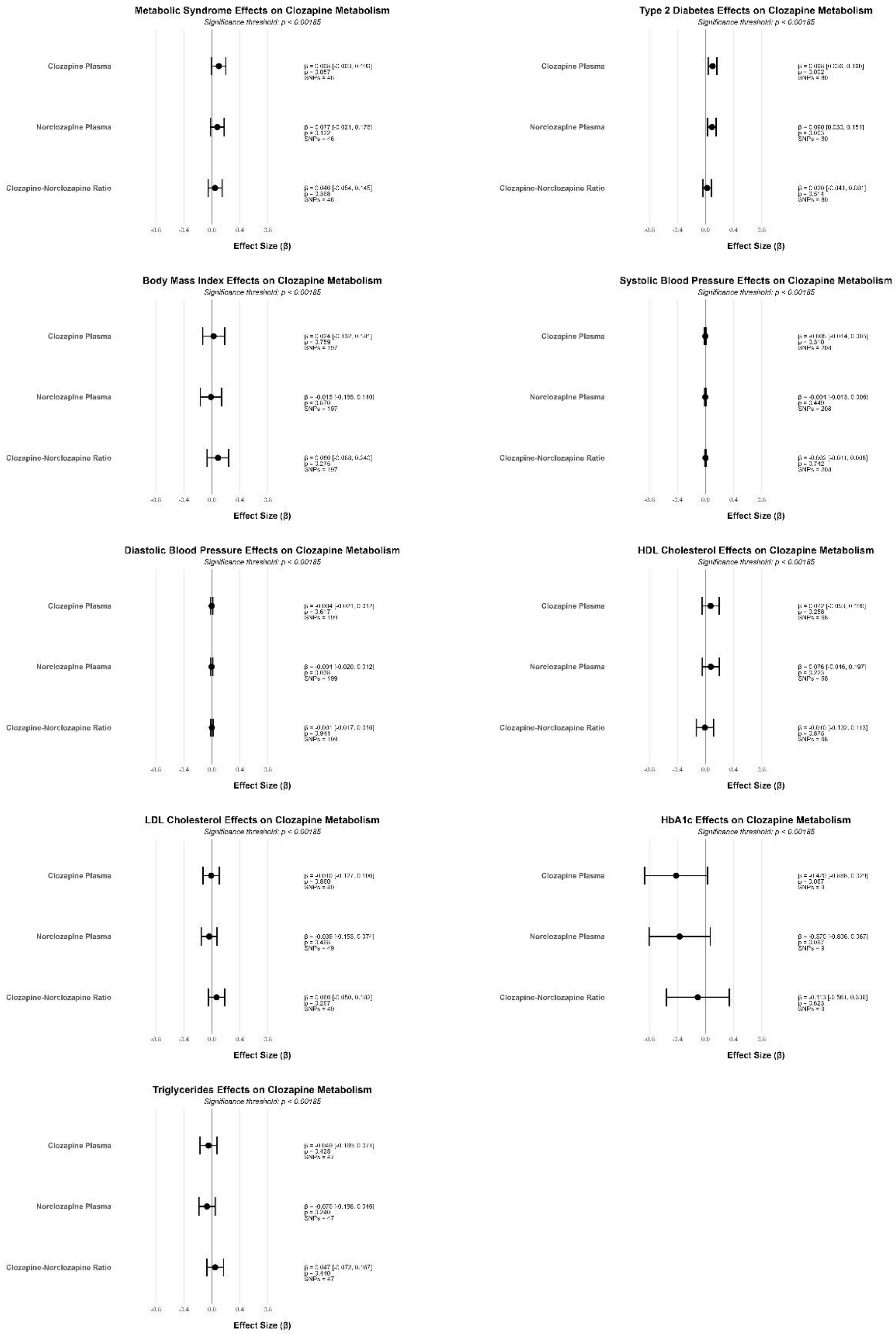

*Supplementary Figures 2. 5 – Scatterplots – Cardiometabolic Effects on Clozapine Plasma Traits*

#### Metabolic Syndrome

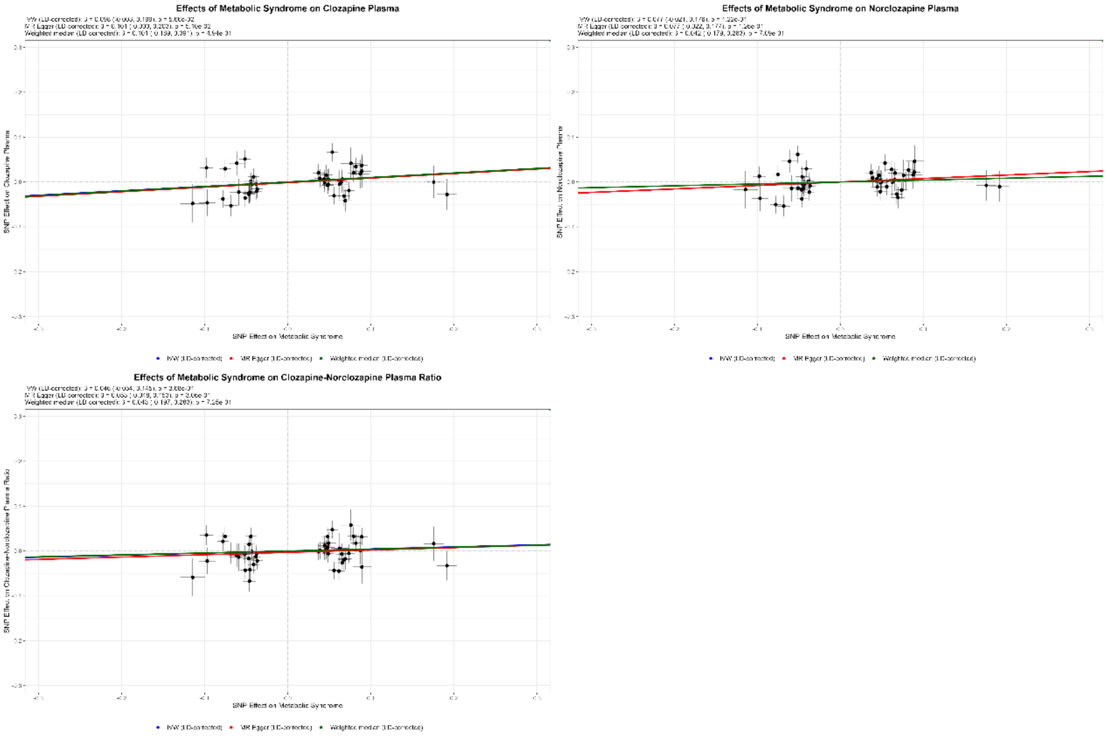

#### Type 2 Diabetes

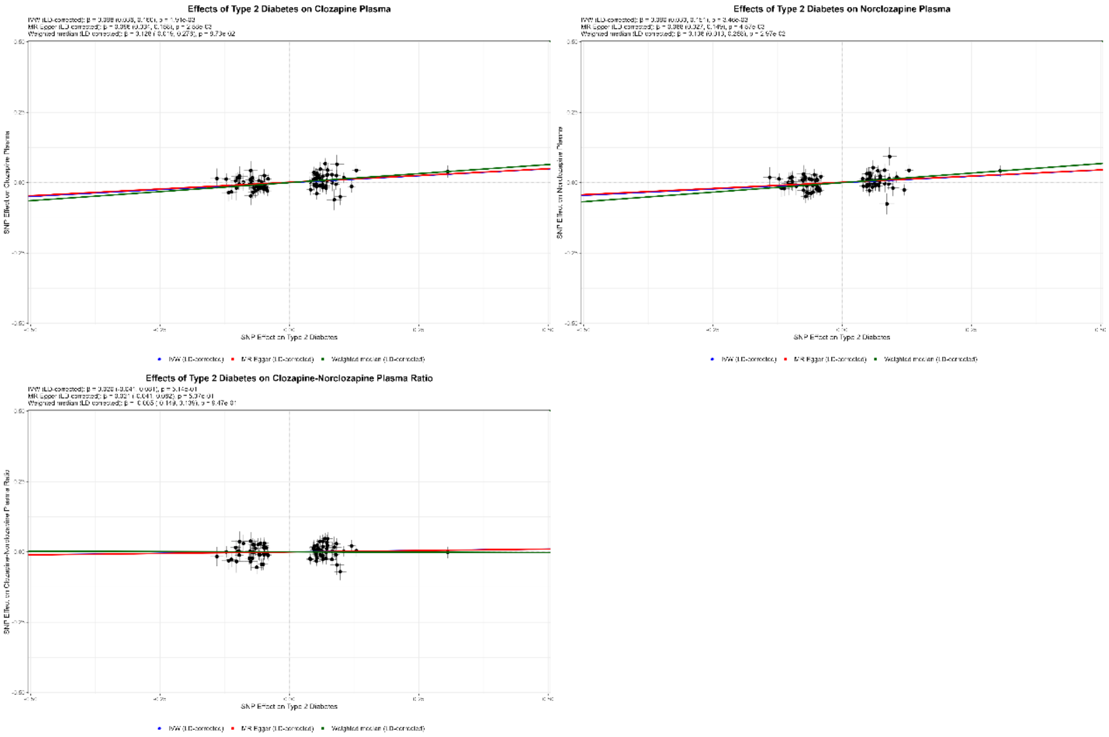

### Body Mass Index

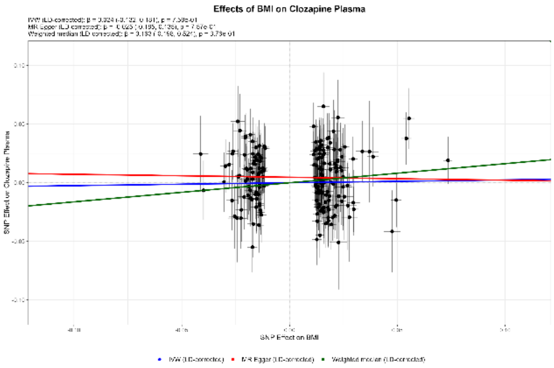

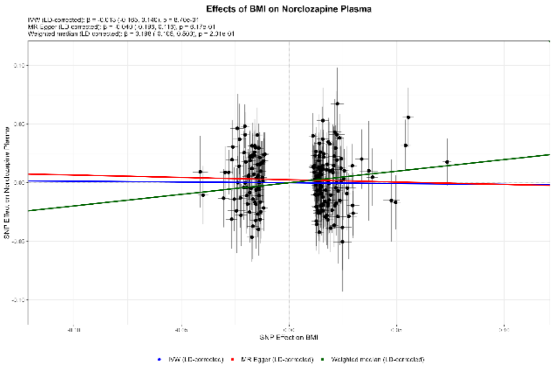

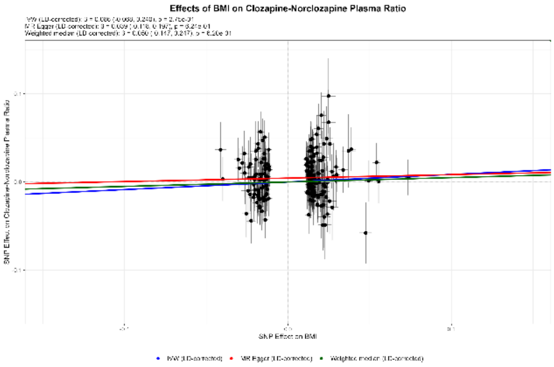

### Systolic Blood Pressure

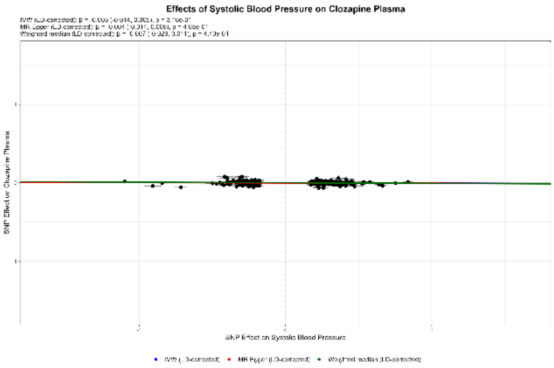

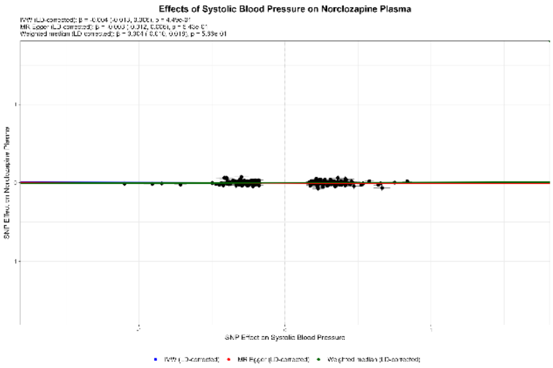

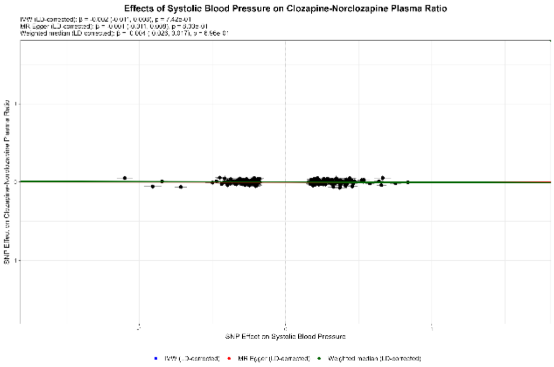

#### Diastolic Blood Pressure

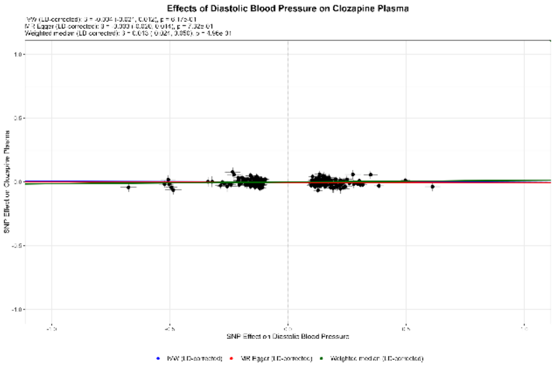

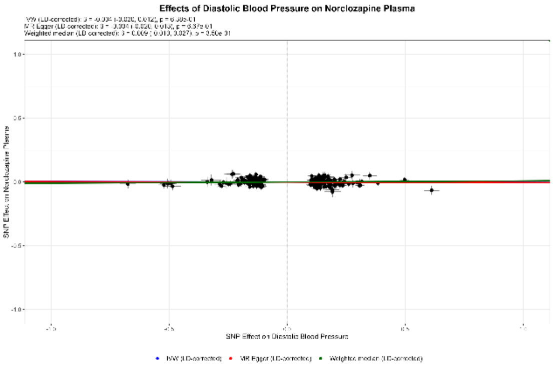

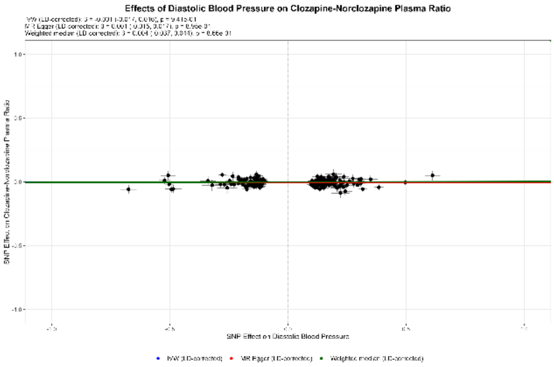

#### HDL

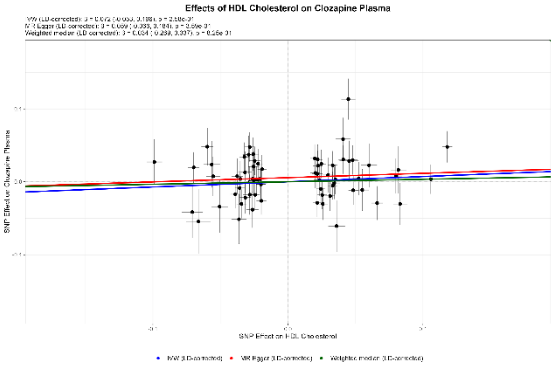

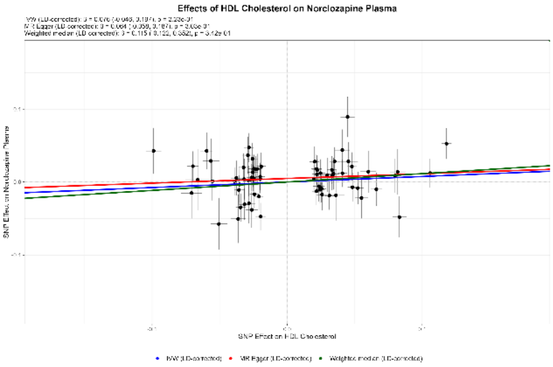

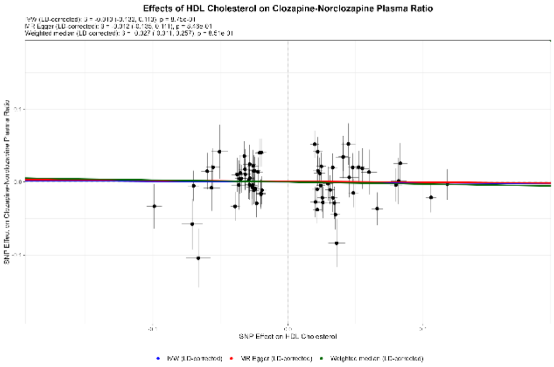

### LDL

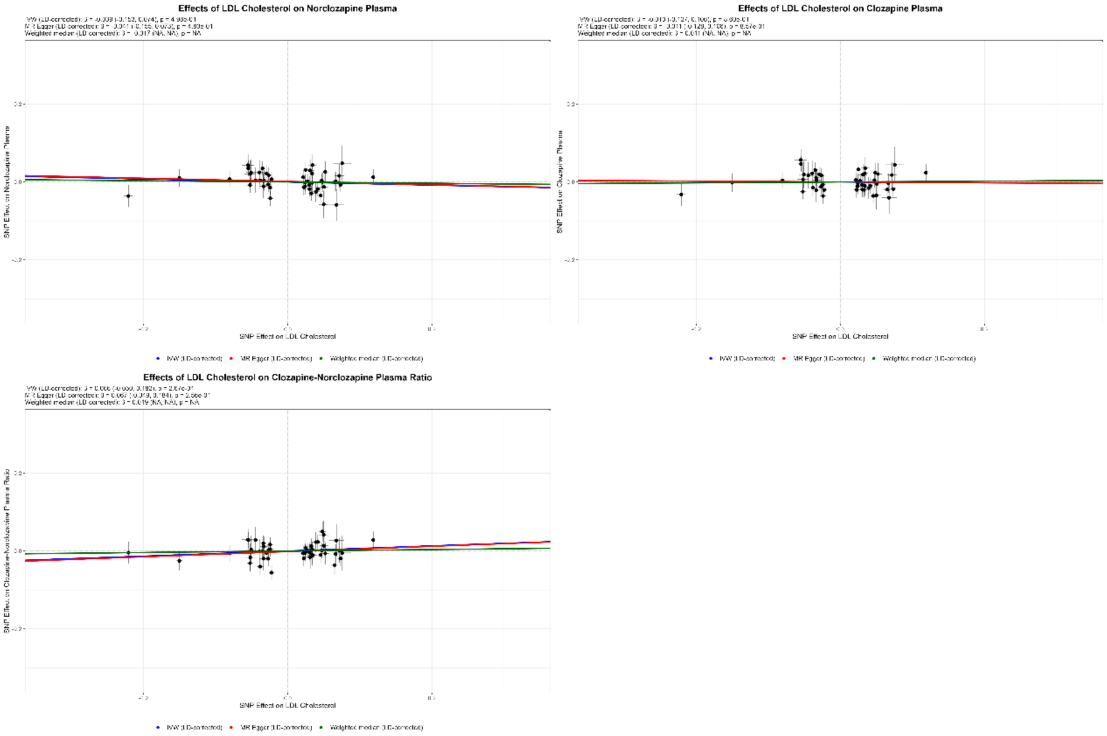
Effects of LDL Cholesterol on Norclozapine Plasma
Effects of LDL Cholesterol on Clozapine Plasma
Effects of LDL Cholesterol on Clozapine-Norclozapine Plasma Ratio
SNP Effect on LDL Cholesterol

### HbA1c

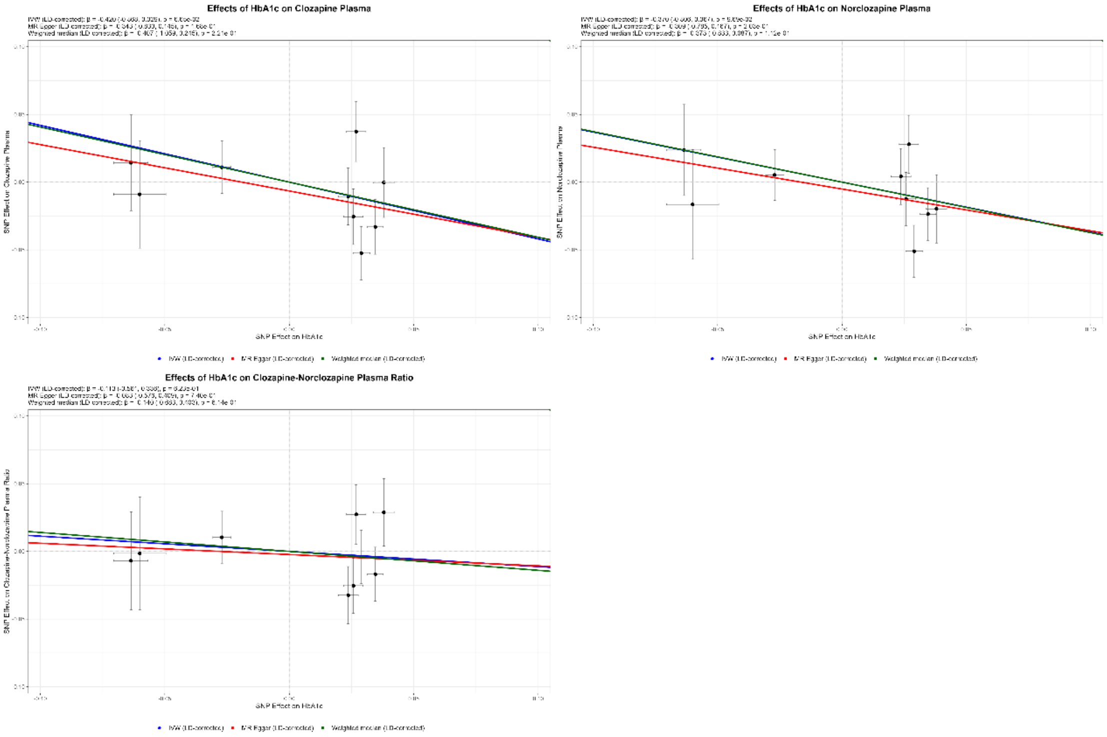
Effects of HbA1c on Clozapine Plasma
Effects of HbA1c on Norclozapine Plasma
Effects of HbA1c on Clozapine-Norclozapine Plasma Ratio
SNP Effect on HbA1c

#### Triglycerides

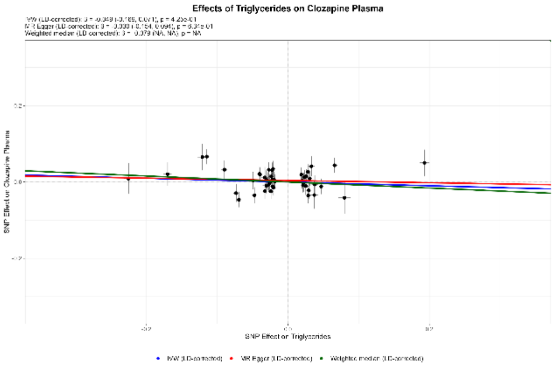

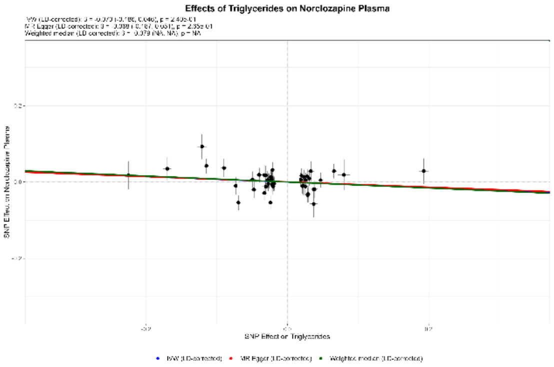

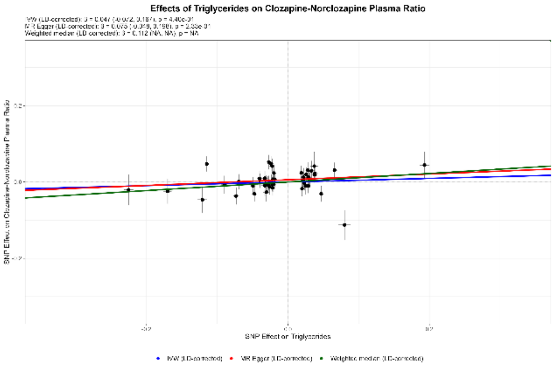

*Supplementary Figures 2. 6 – Leave-one-out Plots – Cardiometabolic Effects on Clozapine Plasma Traits*

#### Metabolic Syndrome

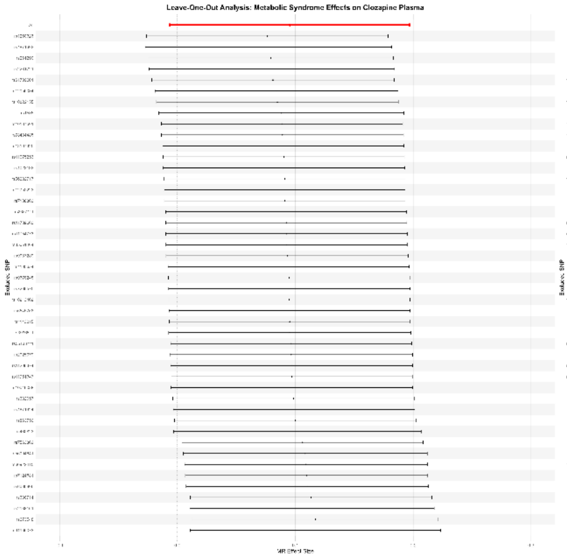

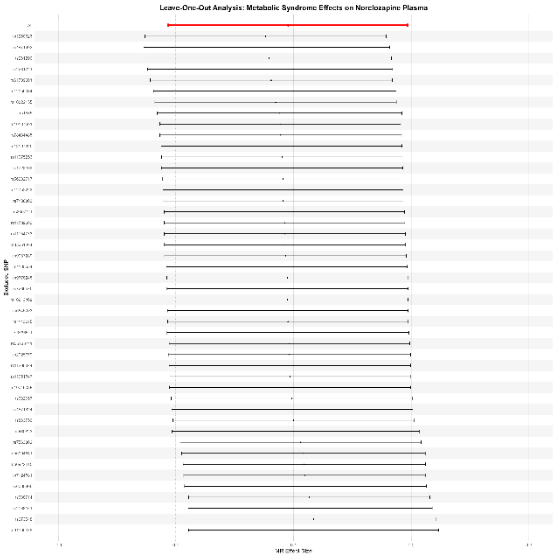

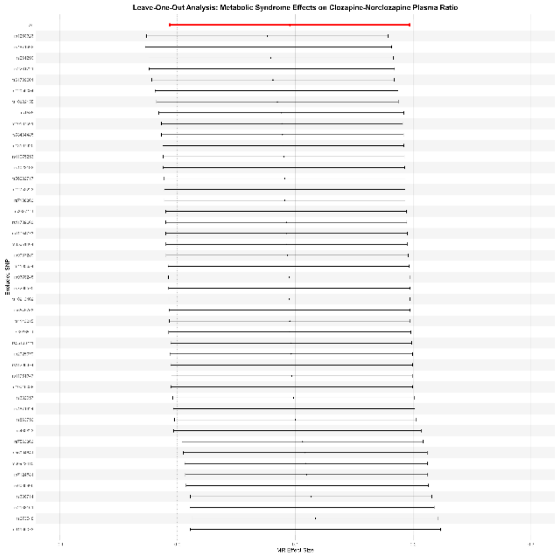

### Type 2 Diabetes

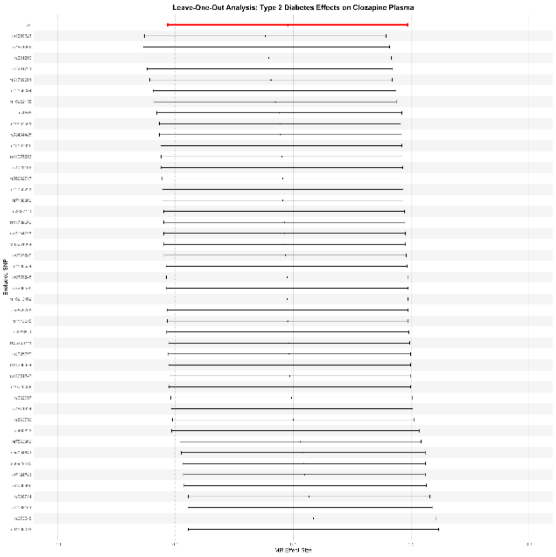

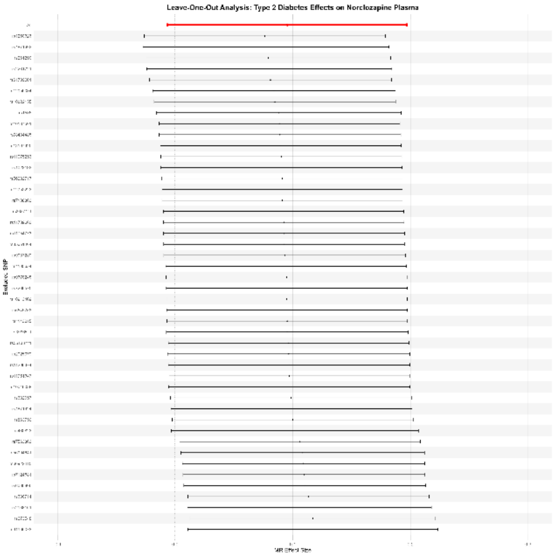

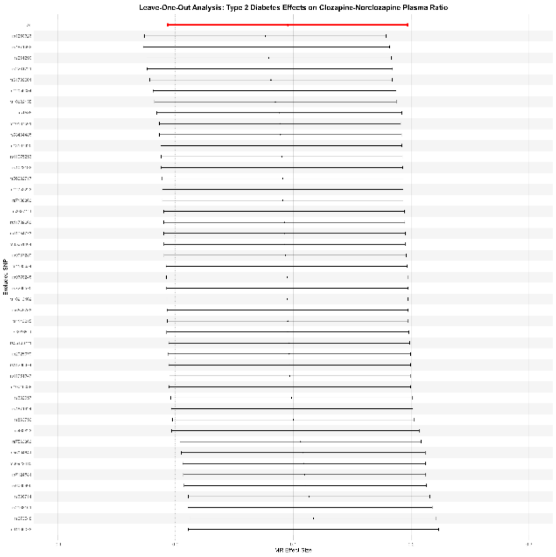

#### Body Mass Index

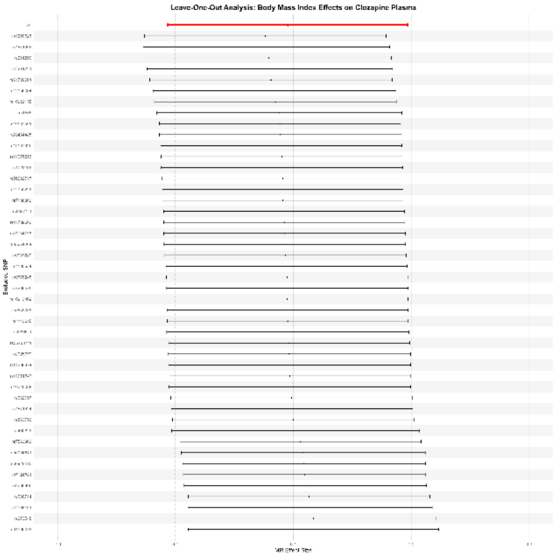

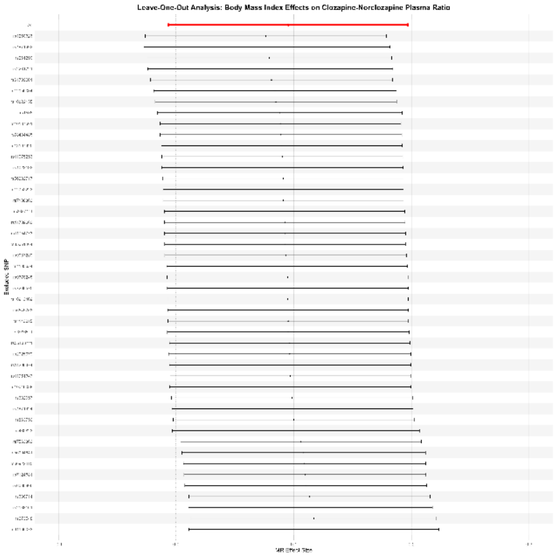

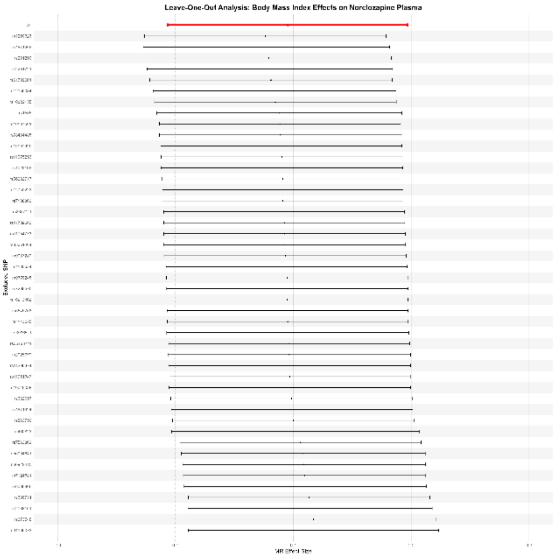

#### Systolic Blood Pressure

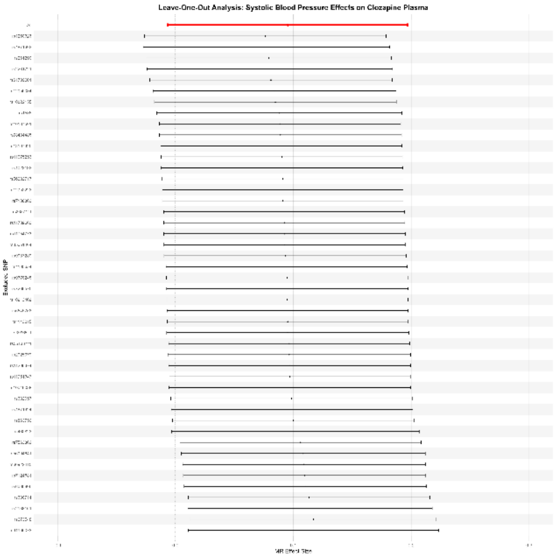

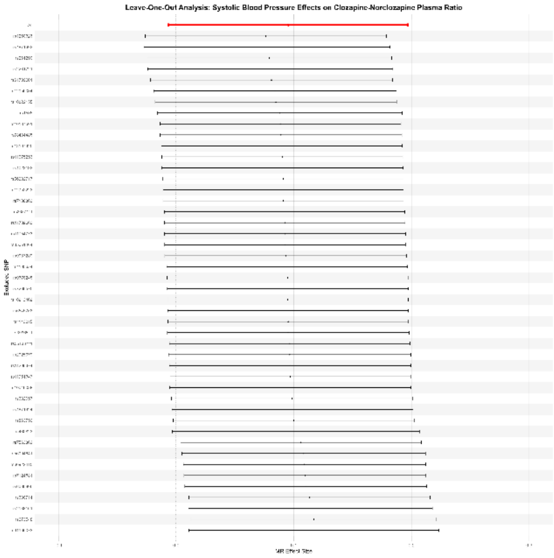

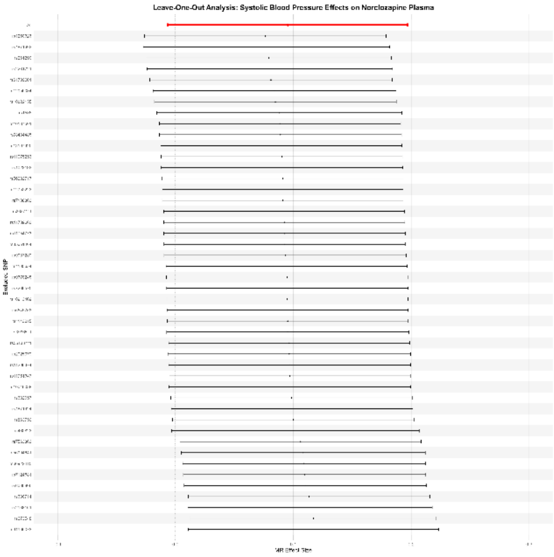

#### Diastolic Blood Pressure

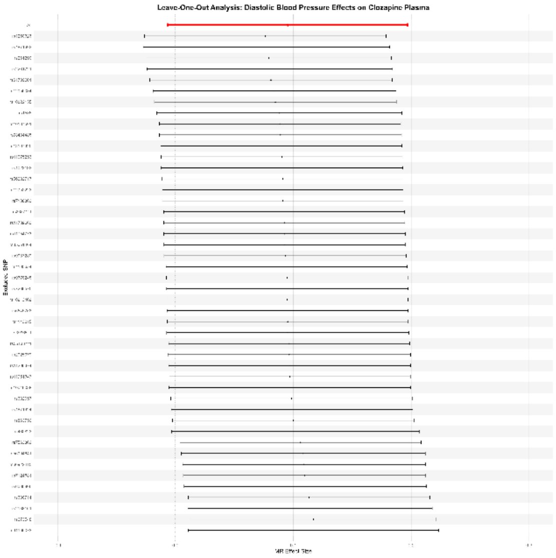

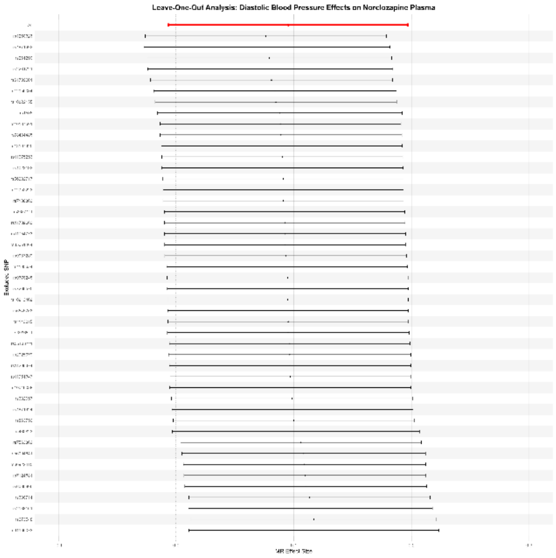

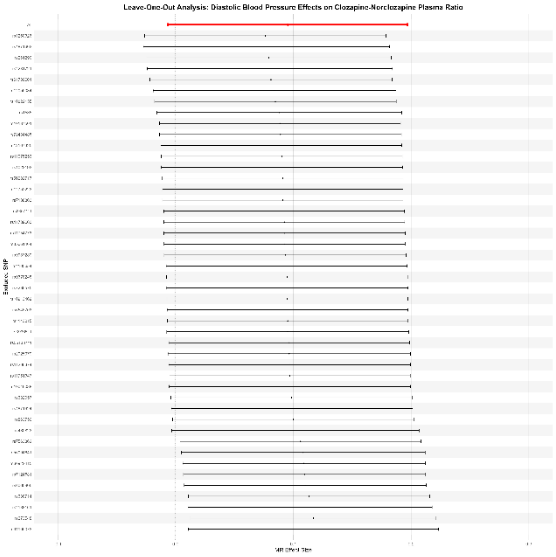

#### HDL

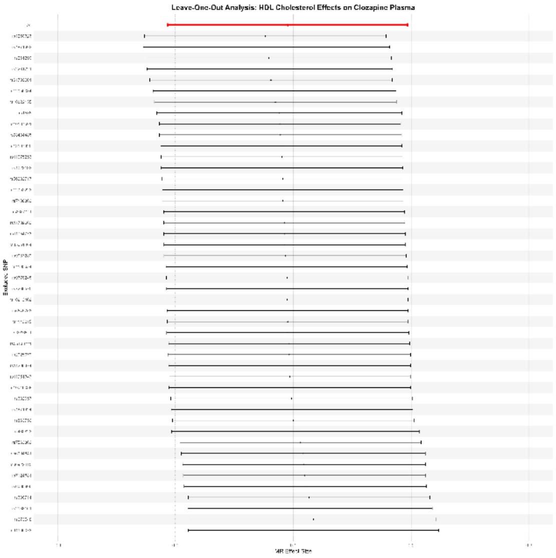
Leave-One-Out Analysis: HDL Cholesterol Effects on Clozapine Plasma
Excluded SNP
MR Effect Size

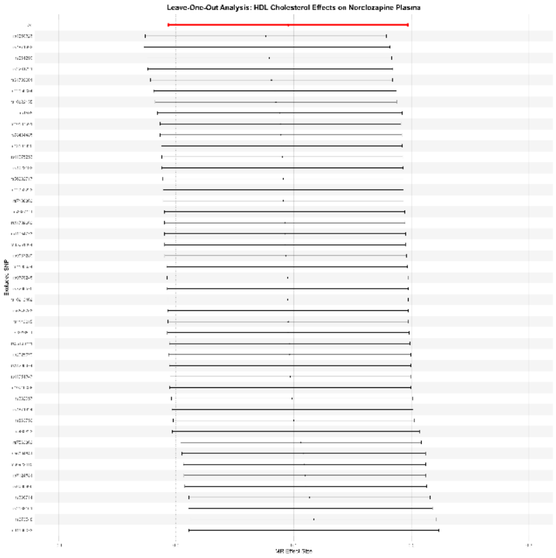
Leave-One-Out Analysis: HDL Cholesterol Effects on Norclozapine Plasma
Excluded SNP
MR Effect Size

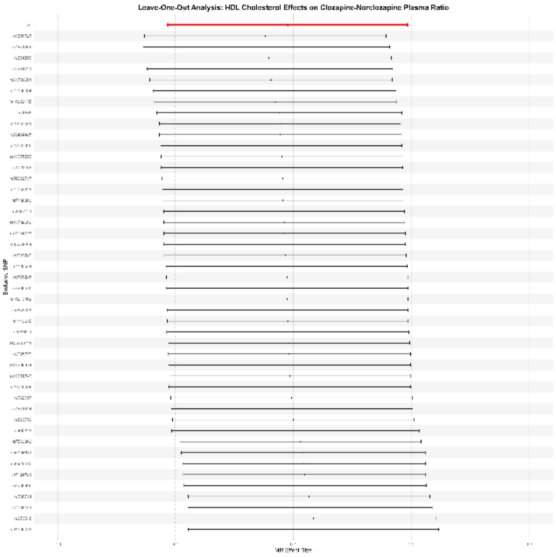
Leave-One-Out Analysis: HDL Cholesterol Effects on Clozapine-Norclozapine Plasma Ratio
Excluded SNP
MR Effect Size

#### LDL

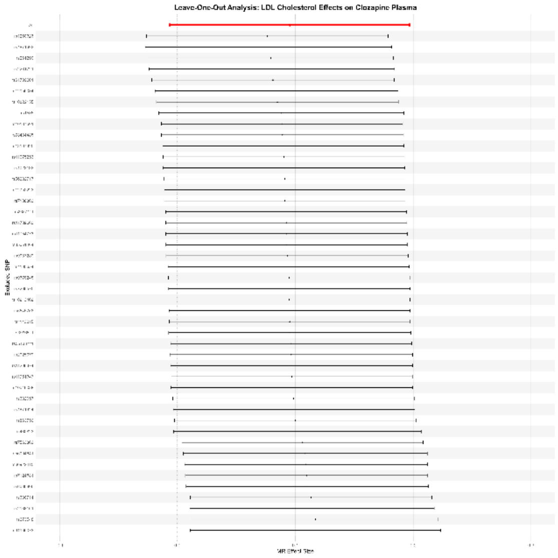
Leave-One-Out Analysis: LDL Cholesterol Effects on Clozapine Plasma
Excluded SNP
MR Effect Size

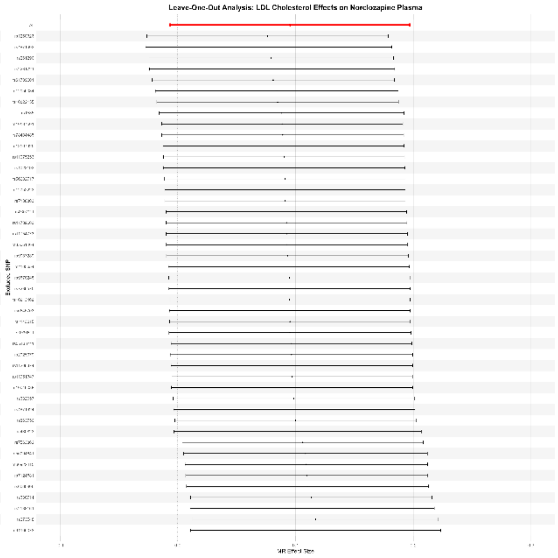
Leave-One-Out Analysis: LDL Cholesterol Effects on Norclozapine Plasma
Excluded SNP
MR Effect Size

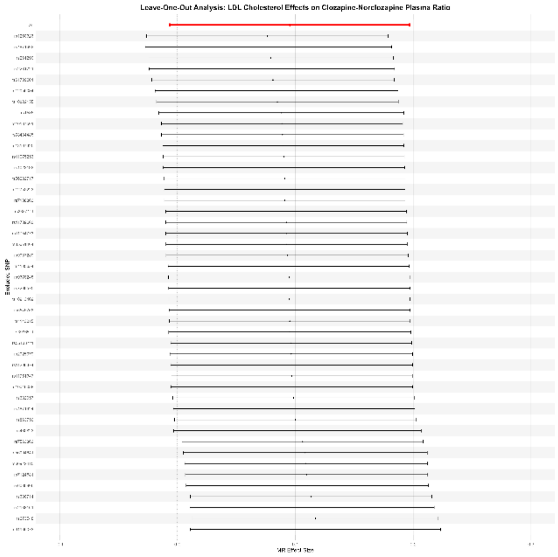
Leave-One-Out Analysis: LDL Cholesterol Effects on Clozapine-Norclozapine Plasma Ratio
Excluded SNP
MR Effect Size

#### HbA1c

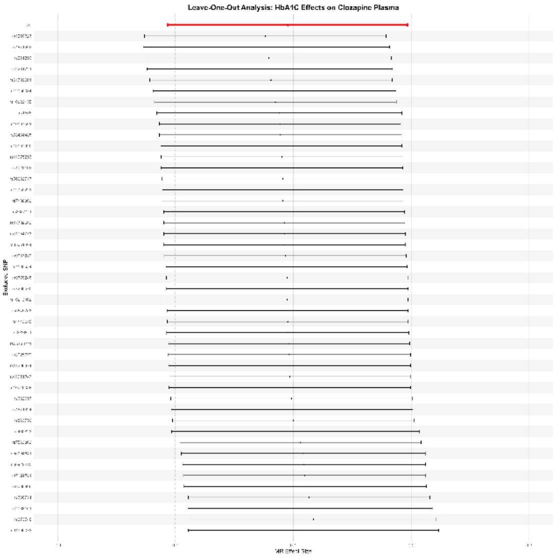
Leave-One-Out Analysis: HbA1C Effects on Clozapine Plasma
Exposure SNP
MR Effect Size

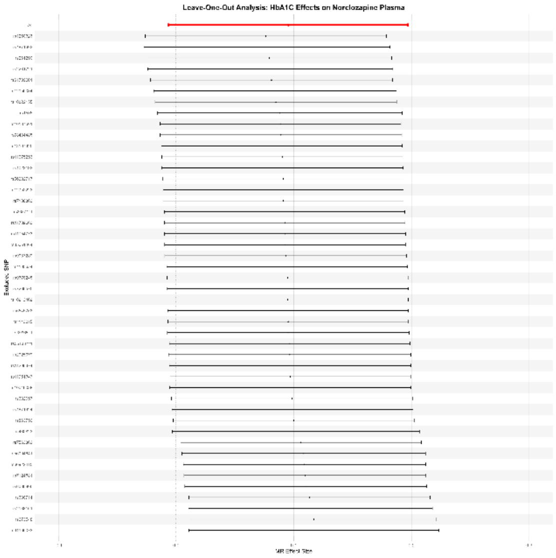
Leave-One-Out Analysis: HbA1C Effects on Norclozapine Plasma
Exposure SNP
MR Effect Size

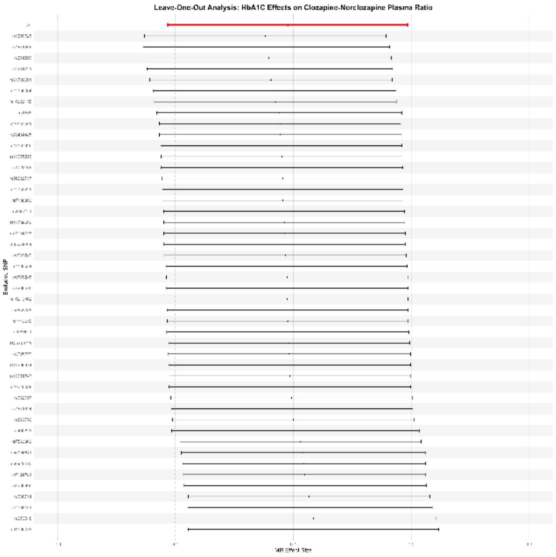
Leave-One-Out Analysis: HbA1C Effects on Clozapine-Norclozapine Plasma Ratio
Exposure SNP
MR Effect Size

#### Triglycerides

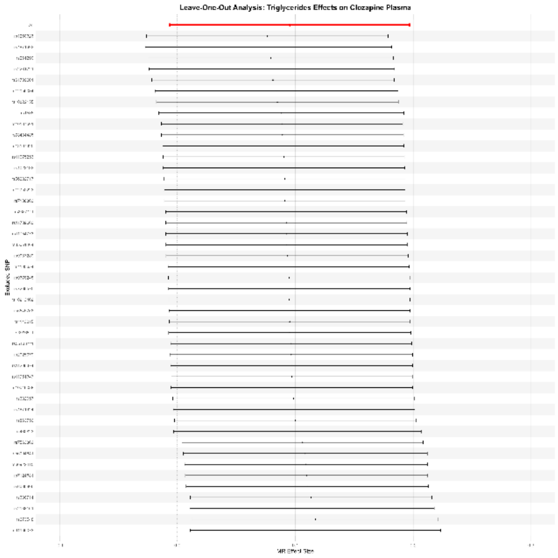

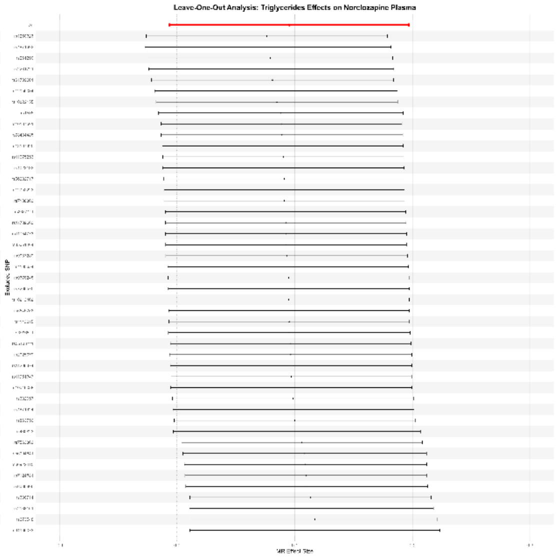

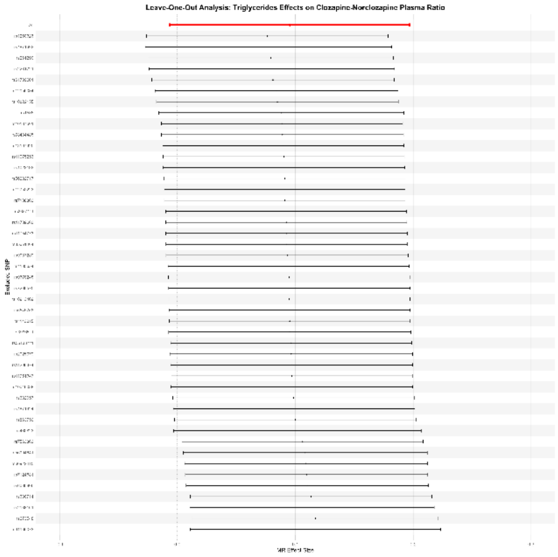

#### Supplementary Figures 3 – Phenome-wide Association Study Results

Extra plots depicting the output from the phenome-wide scan using clozapine variants (SNPs). The heatmap demonstrates the ($-\boldsymbol{log}\ (\boldsymbol{pval})$) of phenotypes identified through the PheWAS analysis with the highest p-value. The heatmap also demonstrates which traits are linked to which genetic variant used as a lead in phenome-scan. The individual variant phenotype hits further demonstrate this information whilst also demonstrating if a trait was identified through multiple GWAS studies and their relative ($-\boldsymbol{log}\ (\boldsymbol{pval})$). Phenome-wide raw results can be found in Supplementary Table 3 and information surrounding the PheWAS analysis itself can be found in the Supplementary Methods.

Supplementary Figure 3. 1 – PheWAS Results for Clozapine Variants, Heatmap ..........32
Supplementary Figure 3. 2 – PheWAS Individual Variant Phenotype Hits......................33

*Supplementary Figure 3. 1 – PheWAS Results for Clozapine Variants, Heatmap*

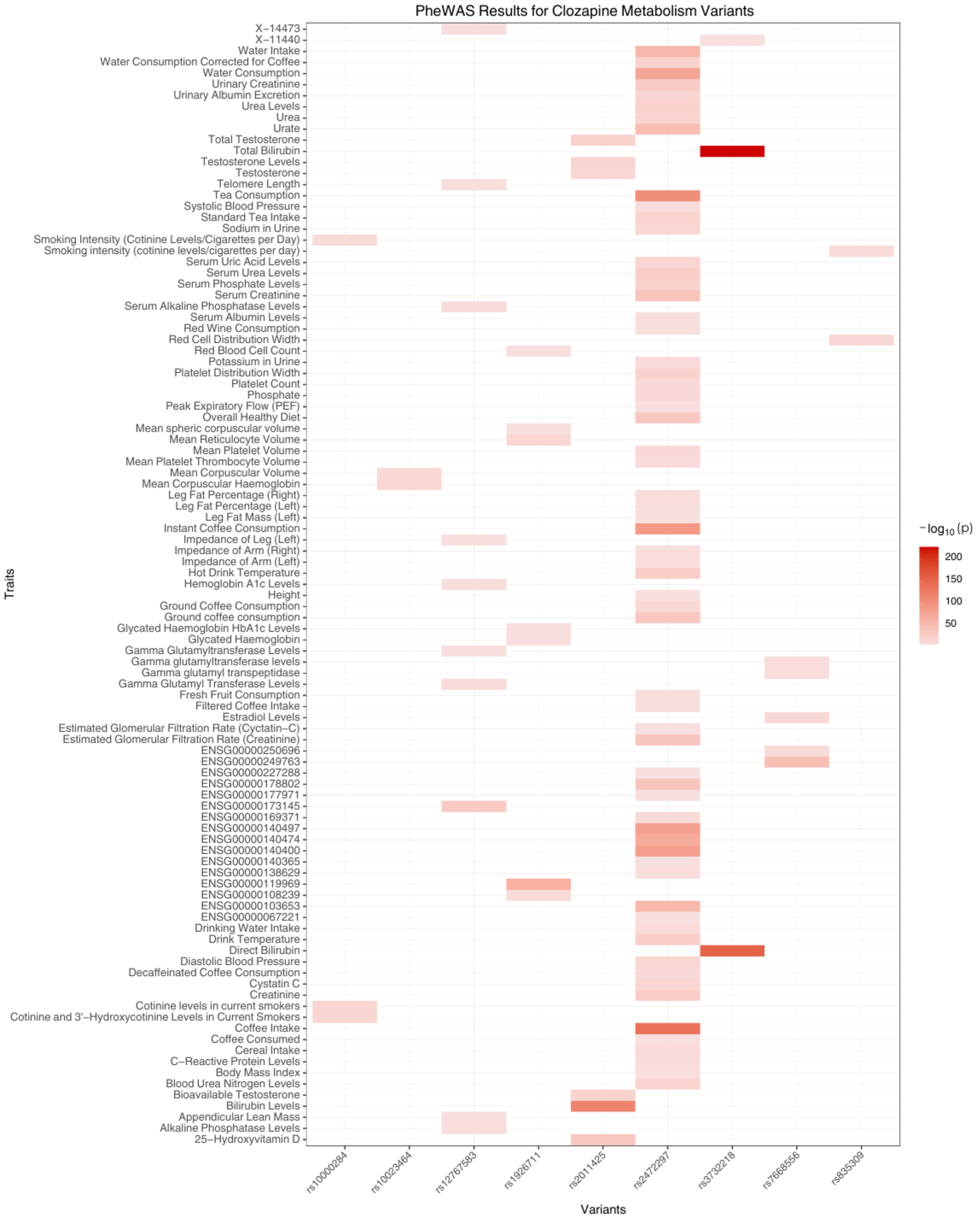

*Supplementary Figure 3. 2 – PheWAS Individual Variant Phenotype Hits*

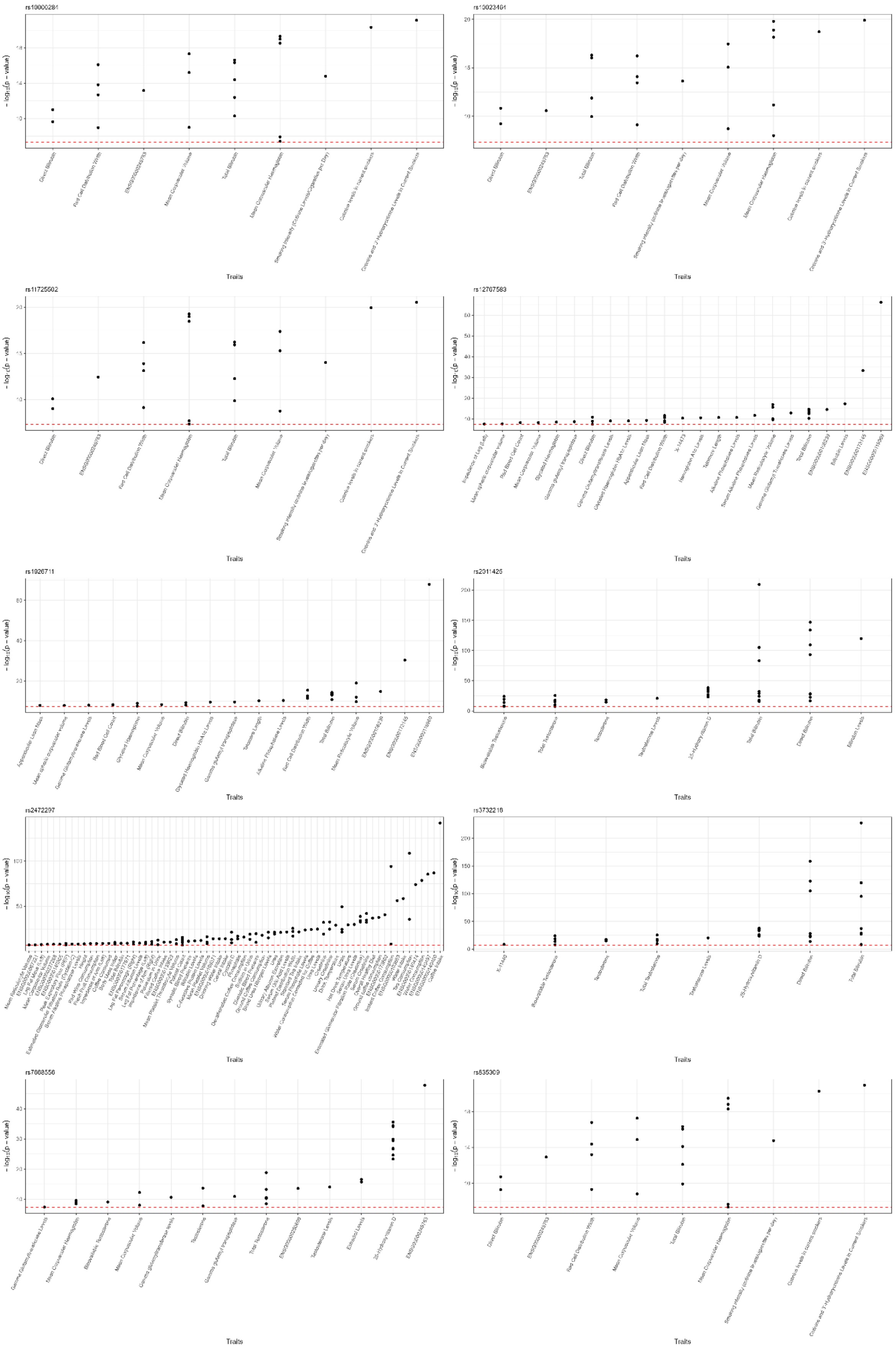

#### Supplementary Figures 4 – Colocalization Results

Extra plots depicting the PWCoCo colocalization results. All graphical representations show only results with a $H_4 > 0.80$, indicating robust colocalization. The bar graph demonstrates robust colocalization results split by clozapine metabolism exposure trait. Colocalized phenotypes are also split into trait categories. The circular plots demonstrate visually which intermediate phenotypes is being colocalized by which clozapine metabolism trait, even when the intermediate phenotype is colocalizing at multiple clozapine traits. Intermediate phenotypes are also categorised by colour depending upon their classification. The Stacked regional plots demonstrate the statistical strength of associations between genetic variants and a trait or disease within a specific genomic region. These plots demonstrate these relationships for trait that colocalized with a $H_4 > 0.80$. These plots demonstrate the genomic regions in chromosomes; 2, 4, 10, and 15 around the lead variants used for the PheWAS analysis . Raw results can be found in Supplementary Table 4.

Supplementary Figures 4. 1 – Bar Graph of Colocalization Results (H4 > 0.80) for Clozapine Plasma, Norclozapine Plasma and Clozapine-Norclozapine Plasma Ratio. .35

*Supplementary Figures 4. 1 – Bar Graph of Colocalization Results (H4 > 0.80) for Clozapine Plasma, Norclozapine Plasma and Clozapine-Norclozapine Plasma Ratio.*

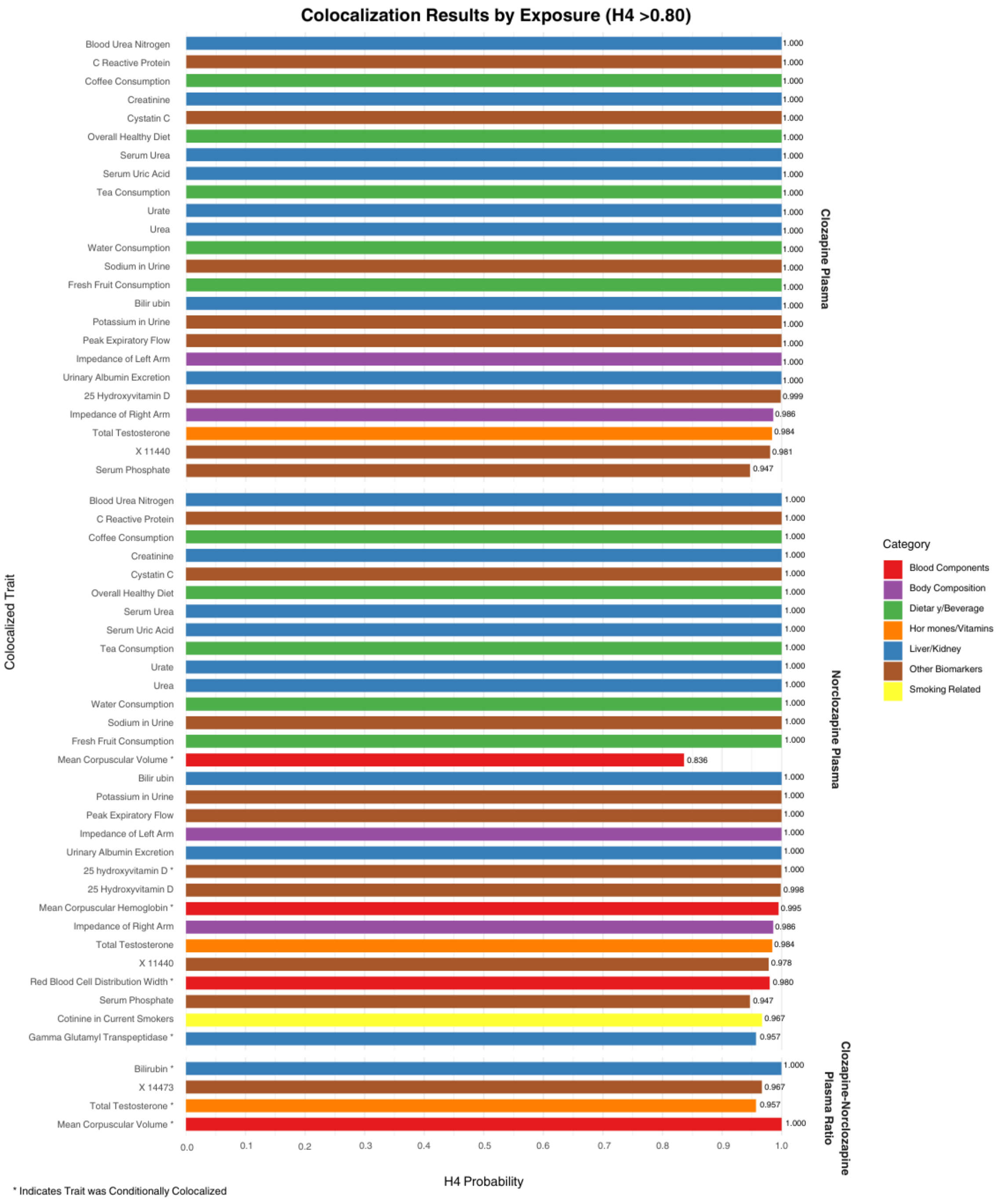

*Supplementary Figures 4. 2 – Circular Plots Demonstrating Clozapine Plasma Traits Colocalizing with PheWAS Phenotypes (H4 > 0.80).*

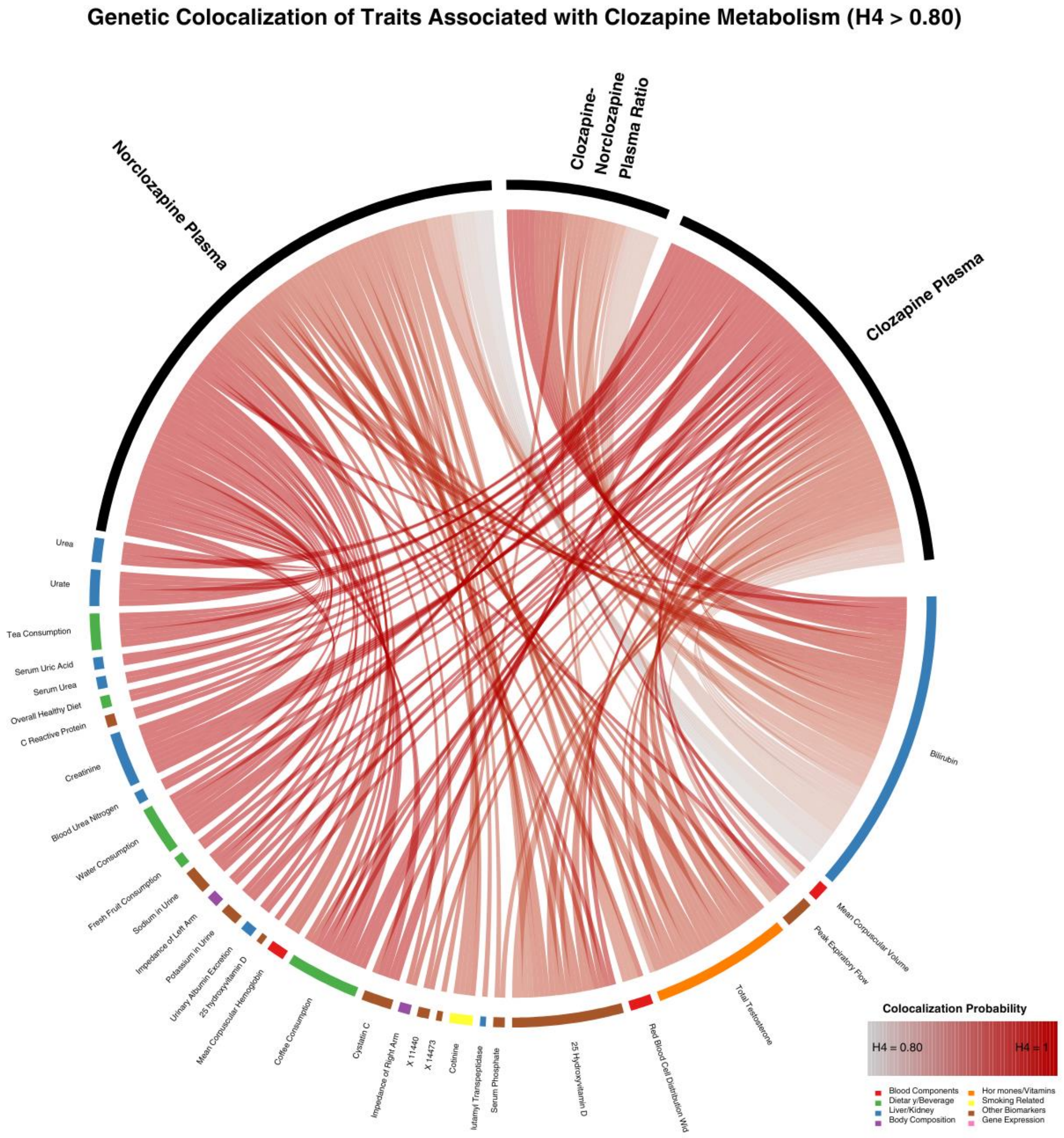

*Supplementary Figures 4. 3 – Stacked Regional Association Plots for Clozapine Colocalizing with PheWAS Identified Candidate Biomarkers Across 4 Lead Chromosomes.*

#### 25-Hydroxyvitamin D

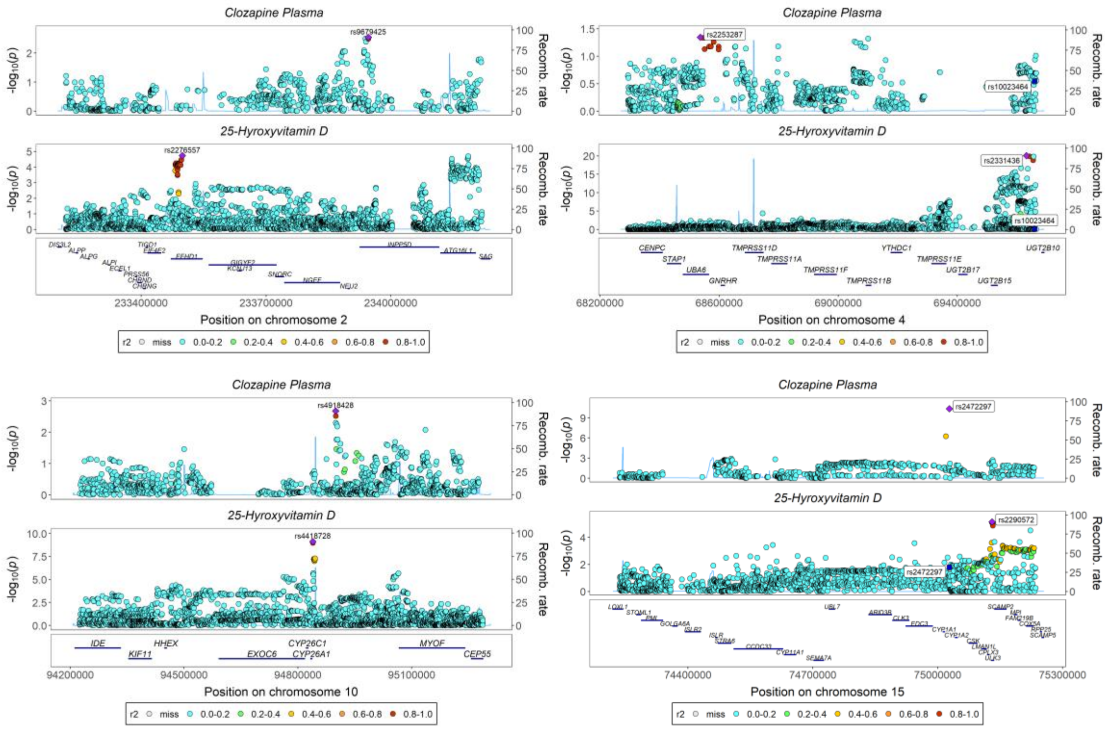

#### Bilirubin

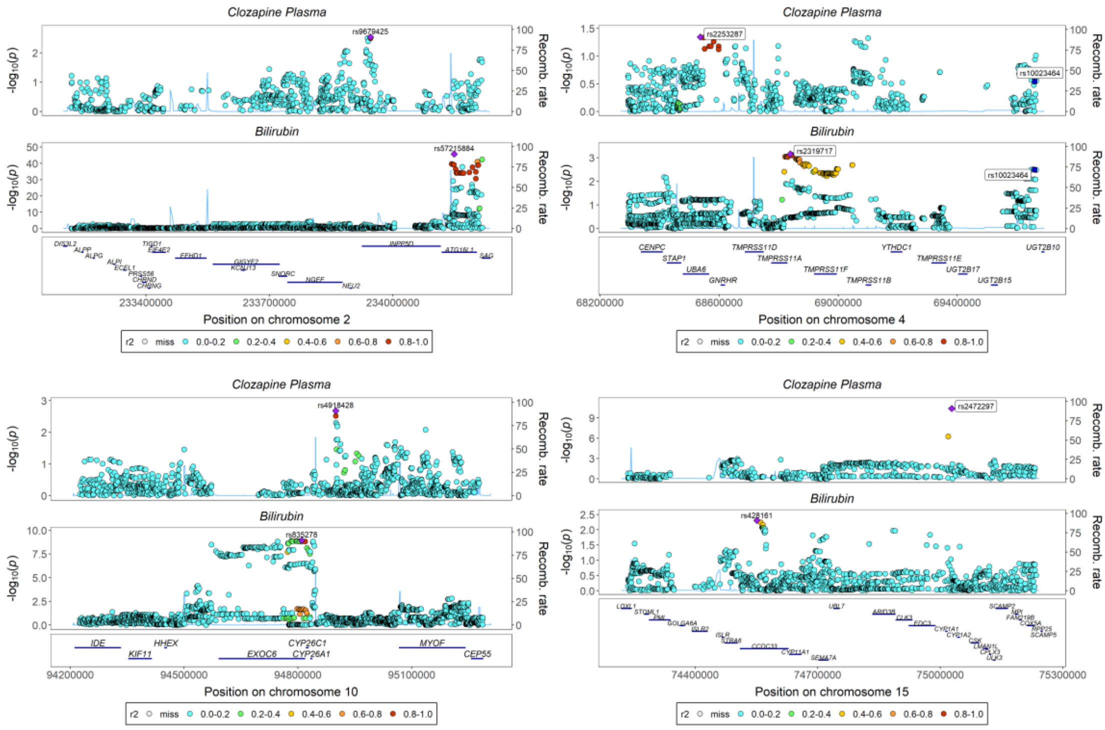

#### Blood Urea Nitrogen

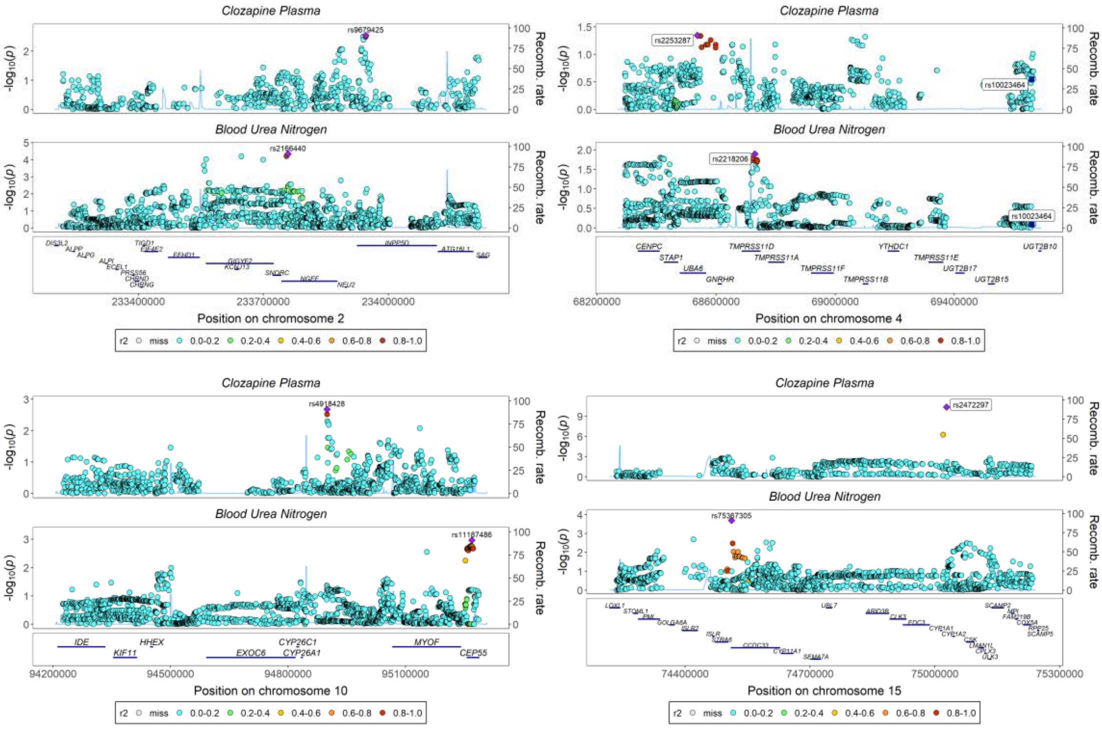

#### Coffee Consumption

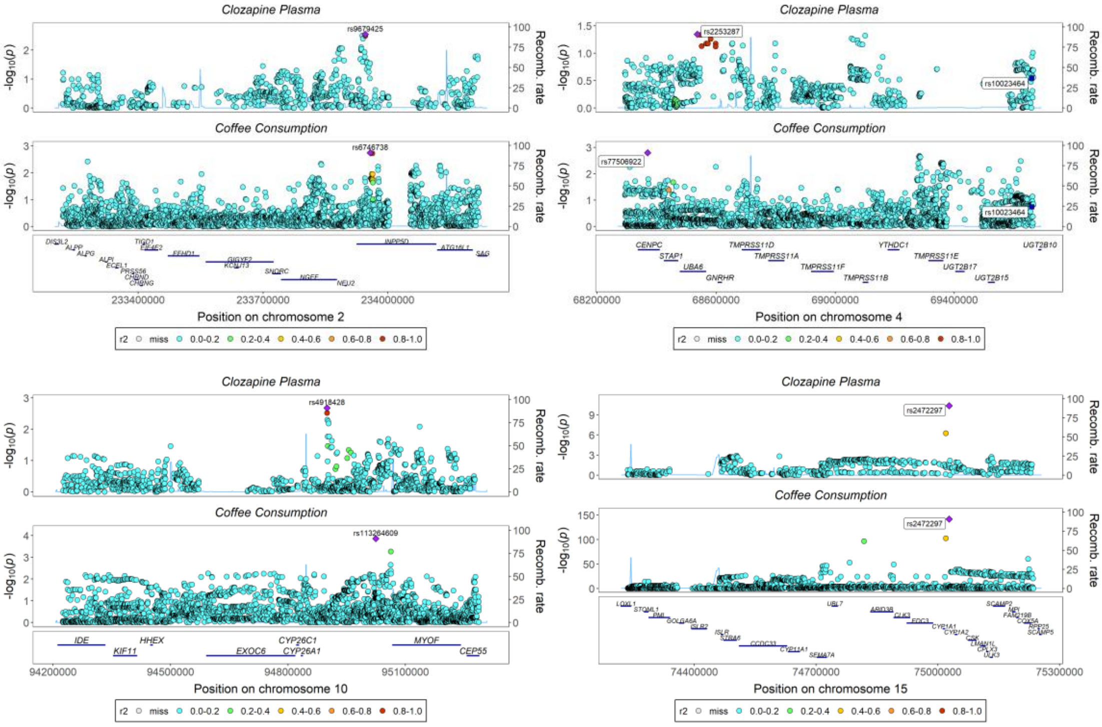

### Creatinine

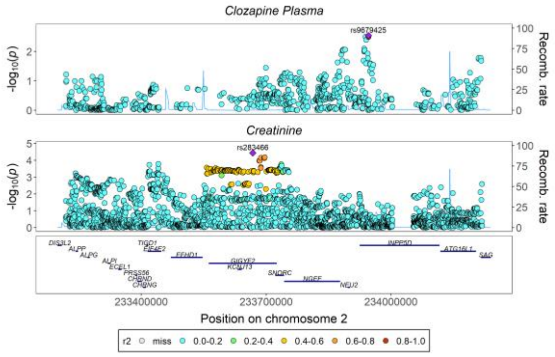

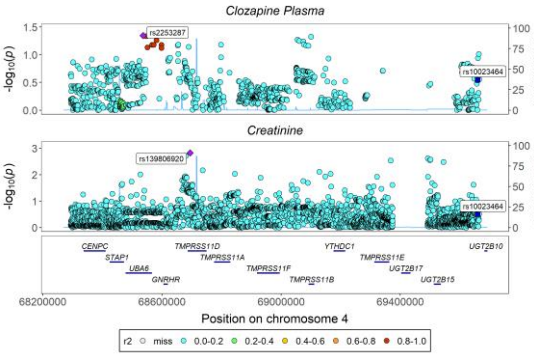

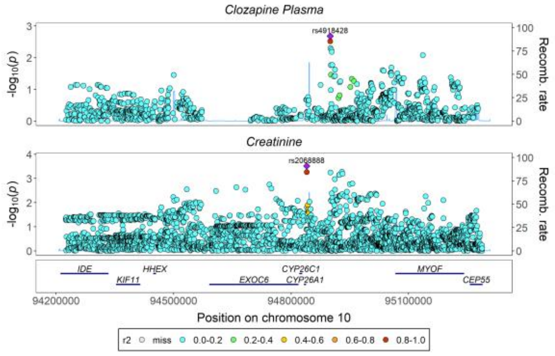

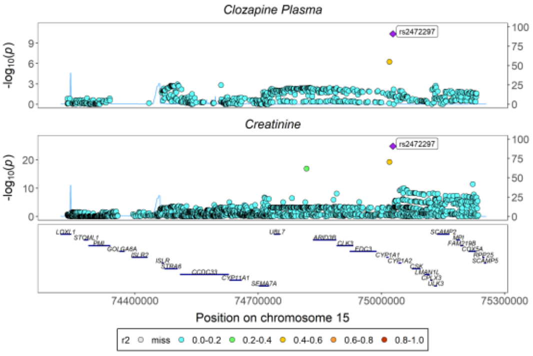

### C-Reactive Protein

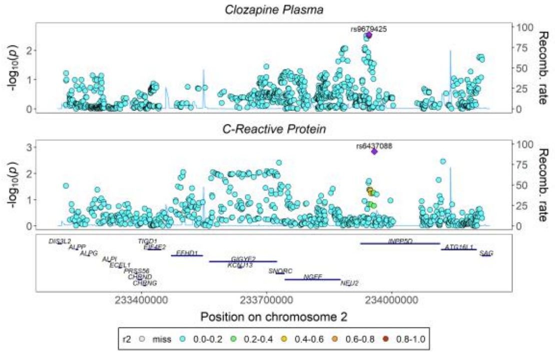

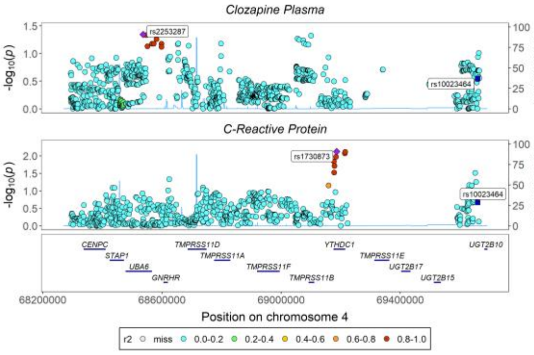

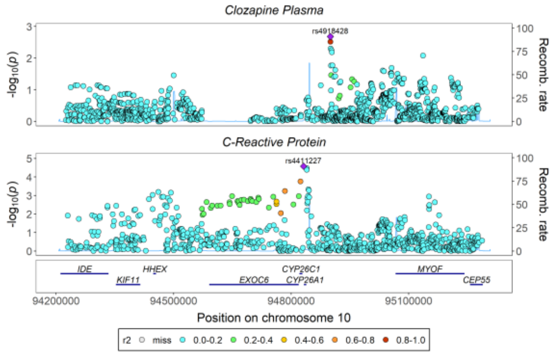

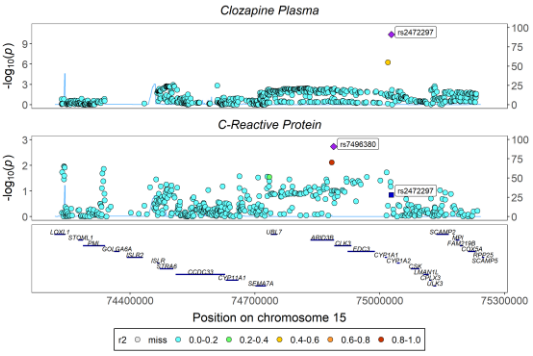

### Cystatin-C

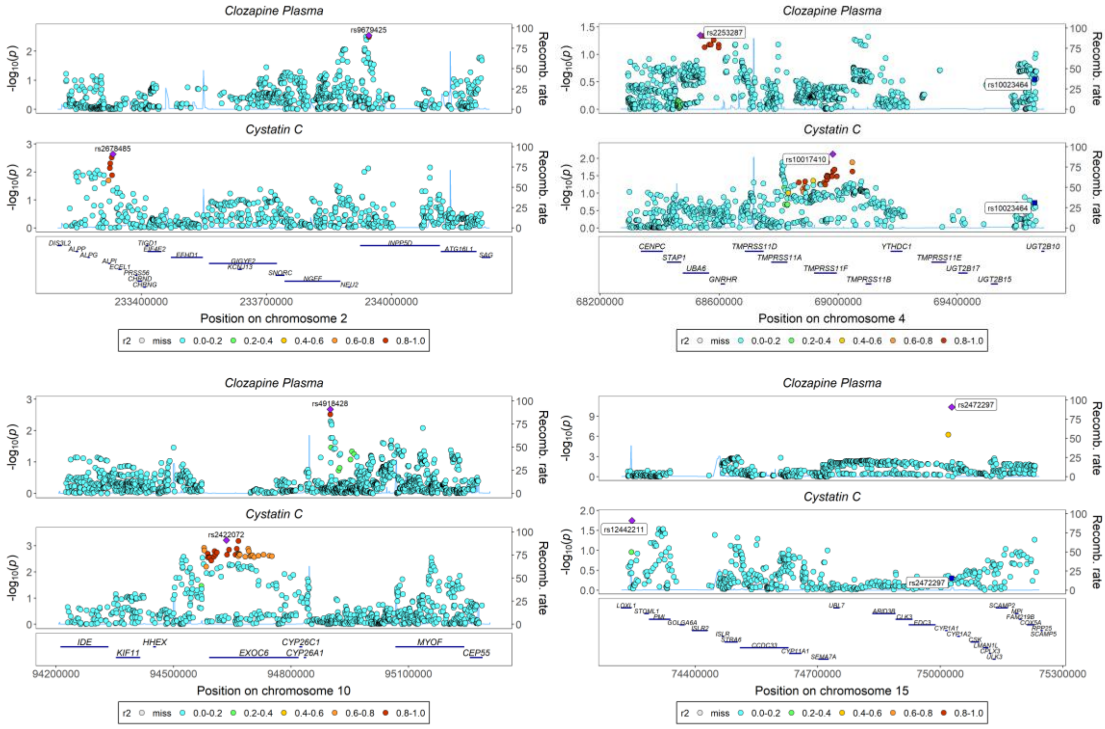

### Fresh Fruit Consumption

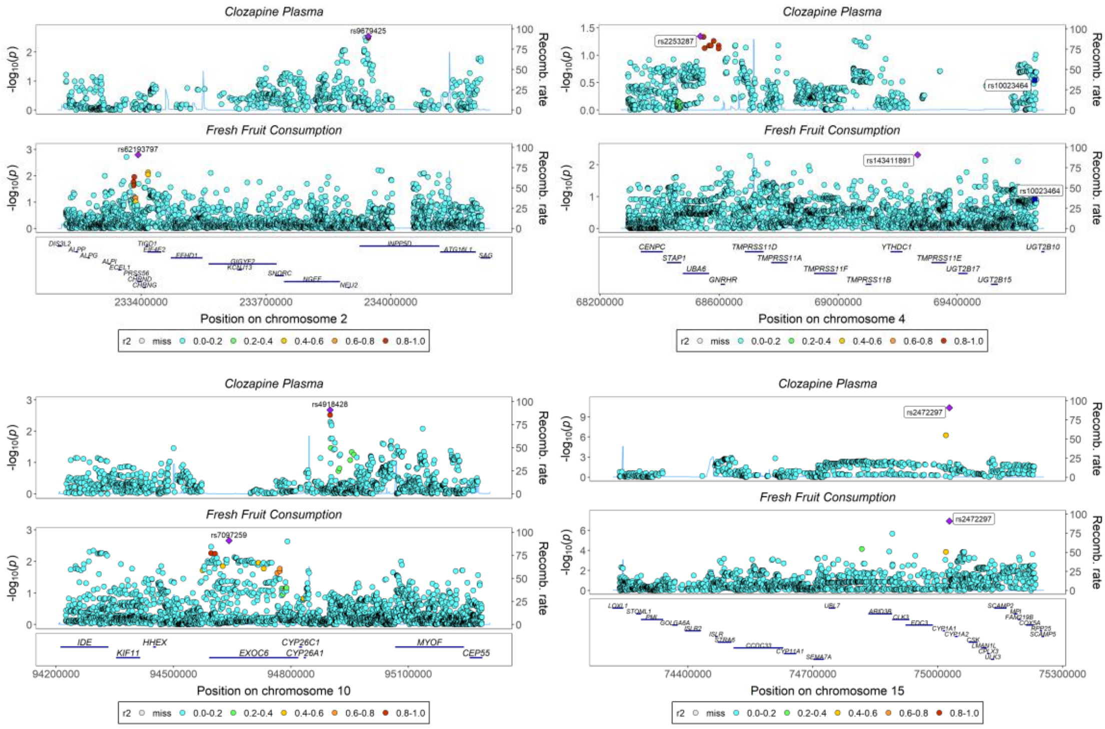

#### Impedance of Left Arm

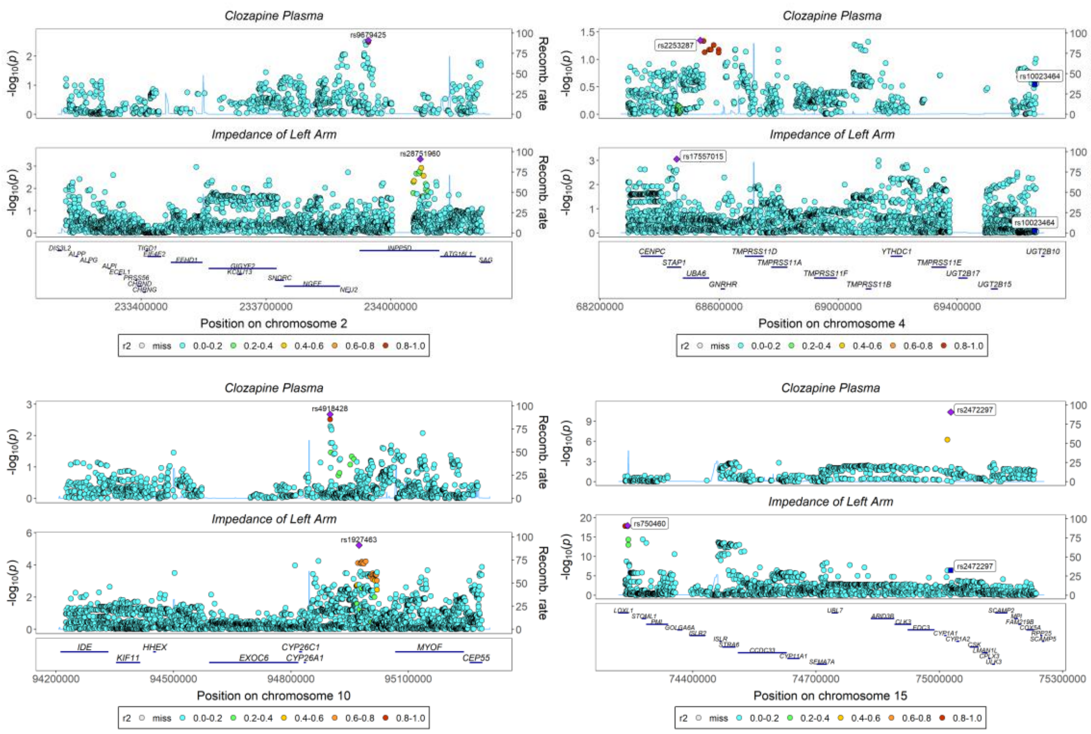

#### Impedance of Right Arm

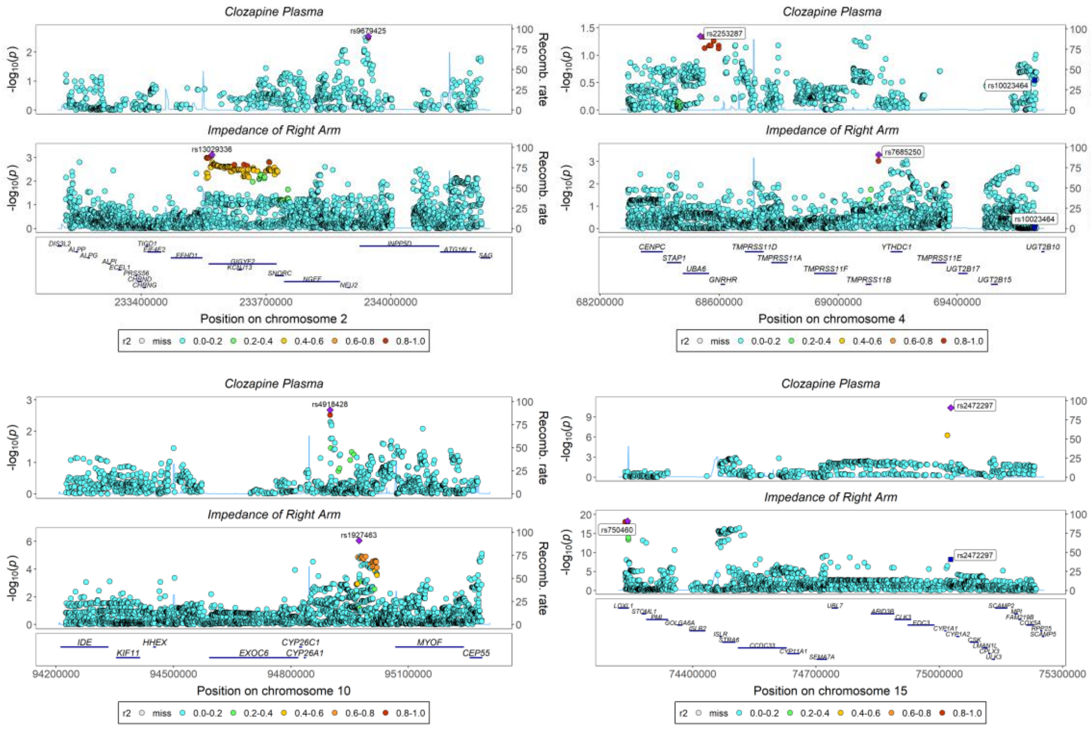

#### Overall Healthy Diet

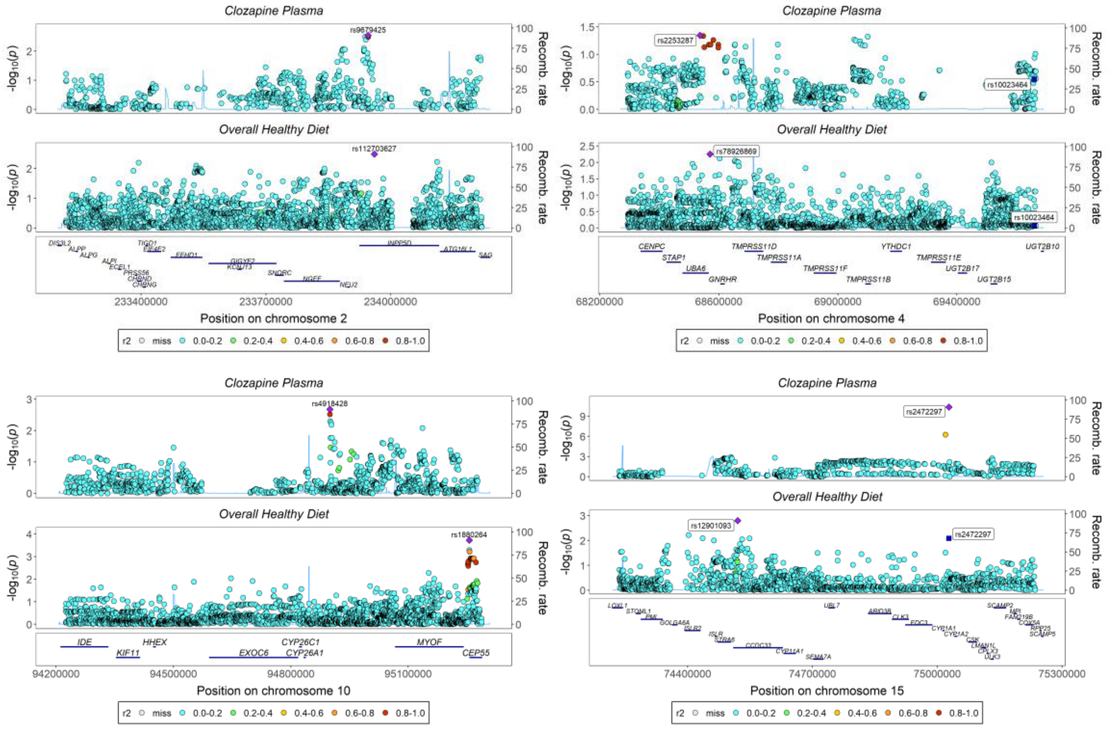

#### Peak Expiratory Flow

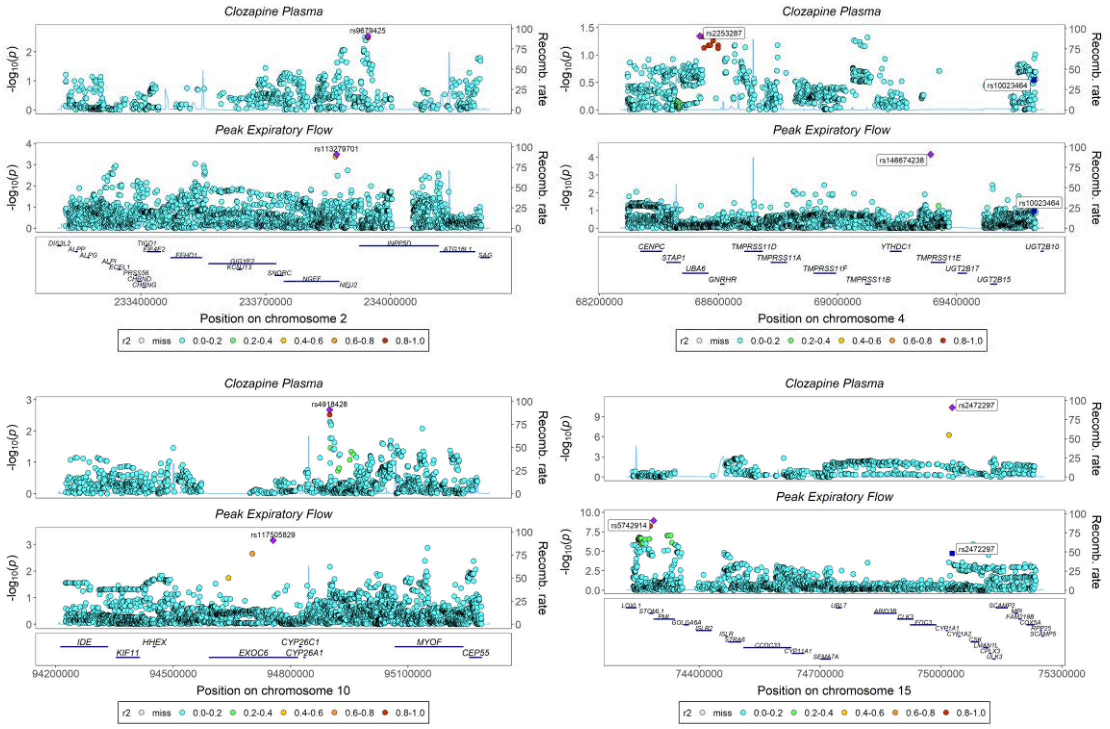

### Potassium in Urine

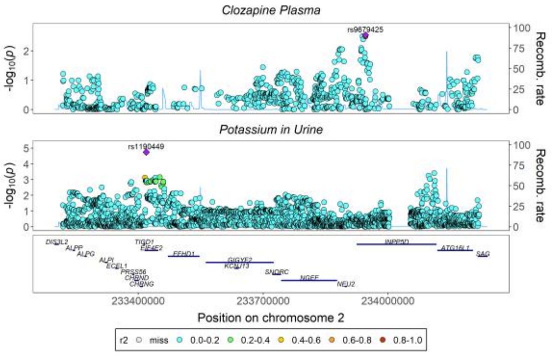

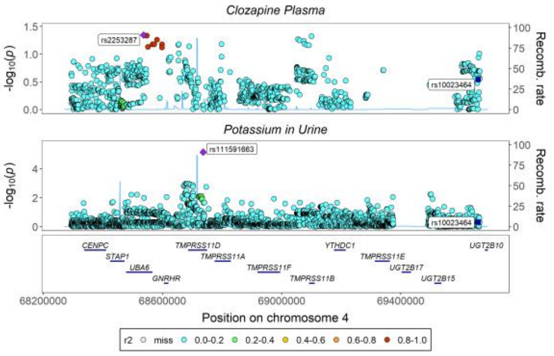

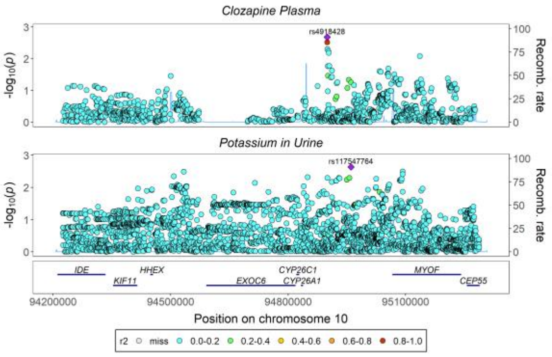

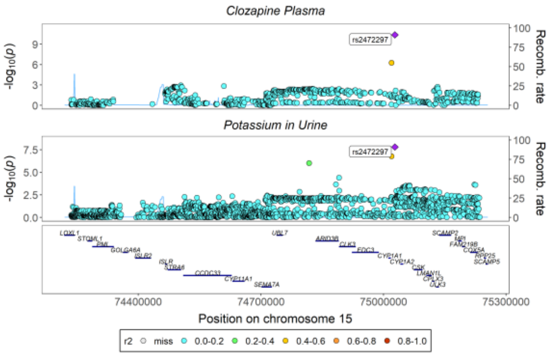

### Serum Phosphate

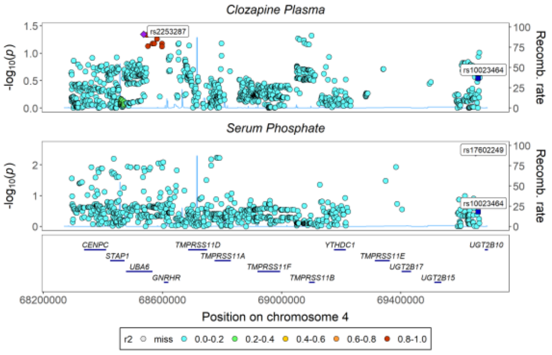

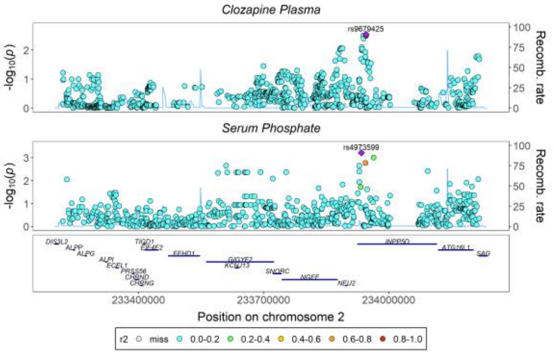

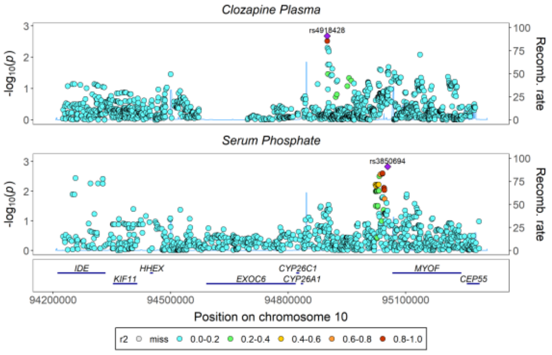

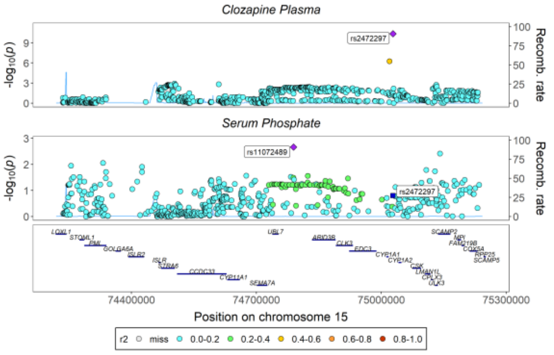

#### Serum Urea

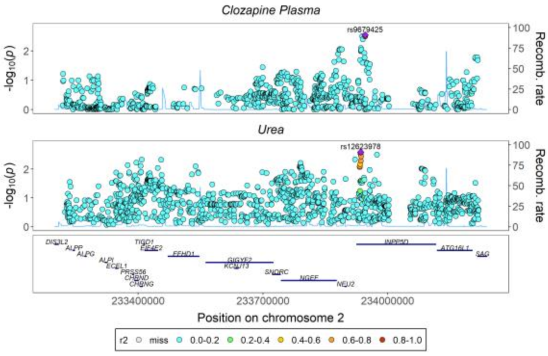

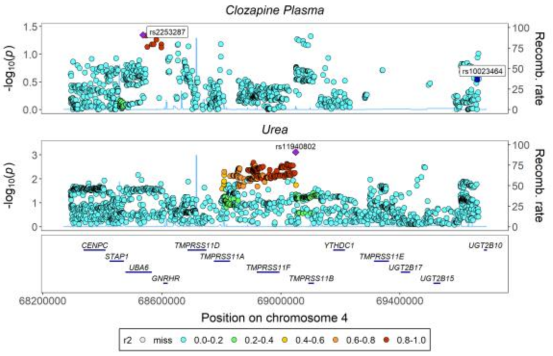

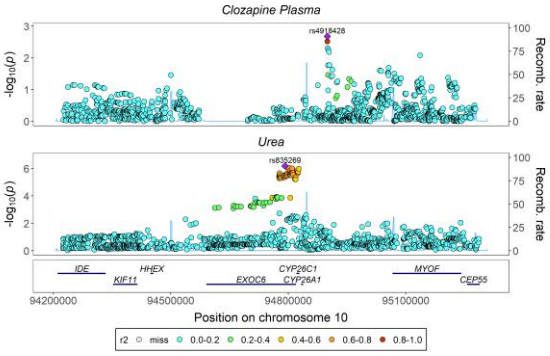

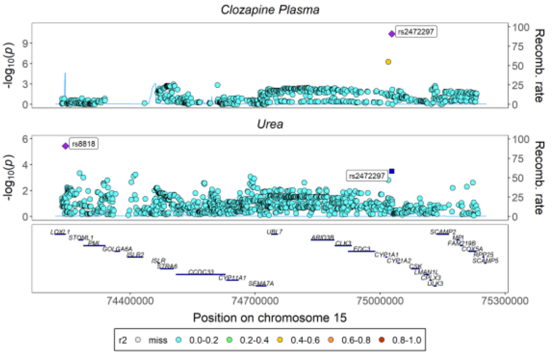

#### Serum Uric Acid

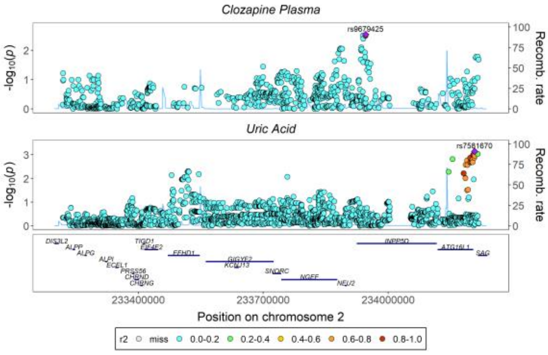

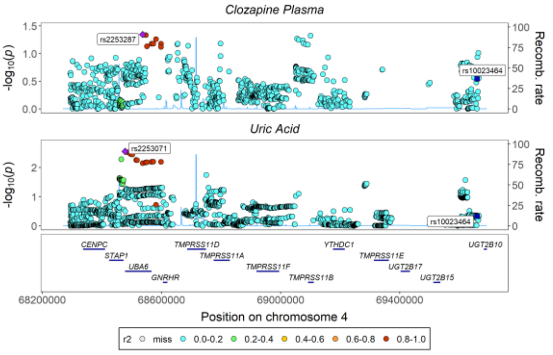

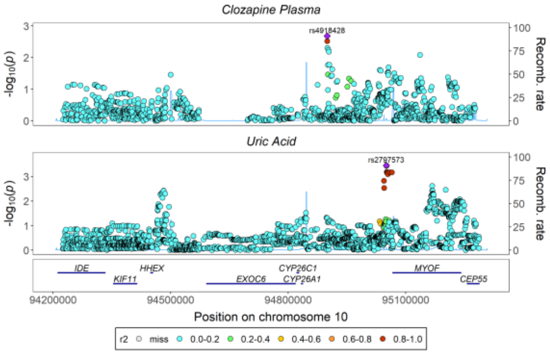

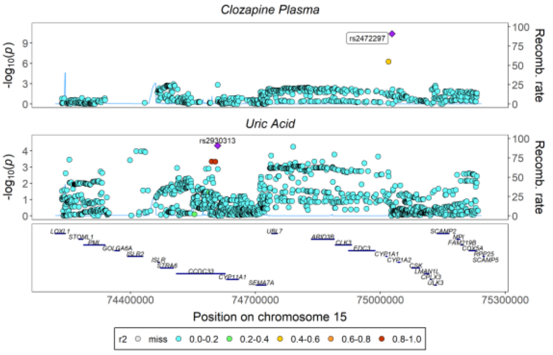

#### Sodium in Urine

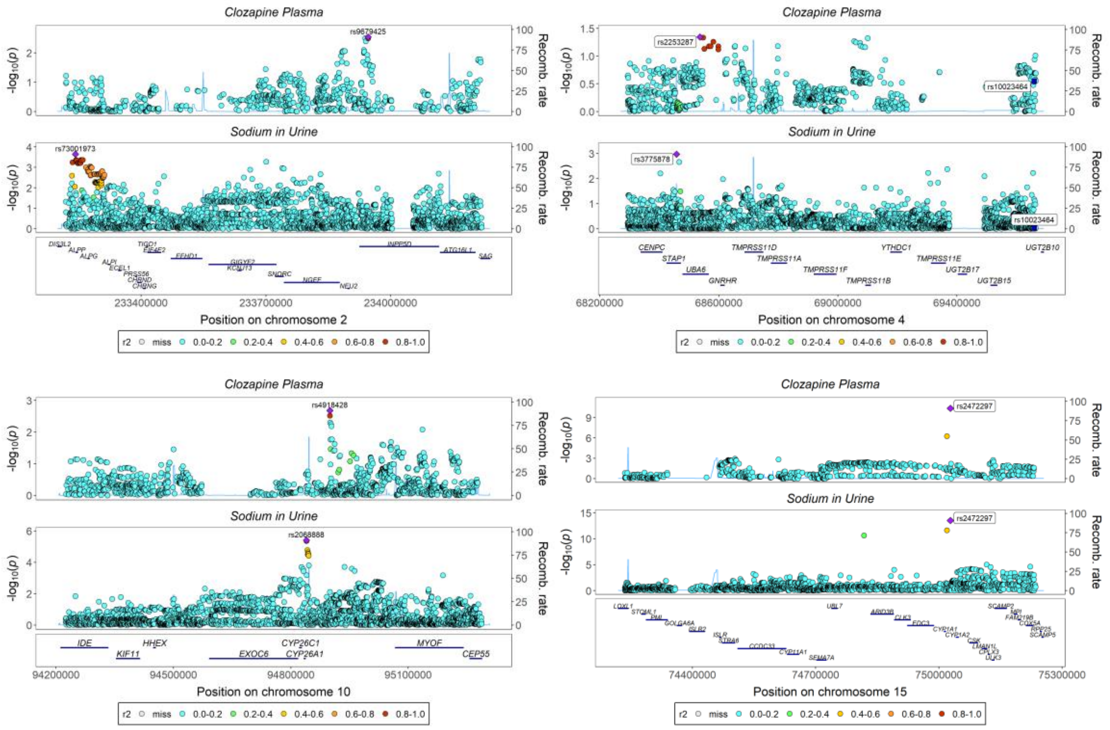

#### Tea Consumption

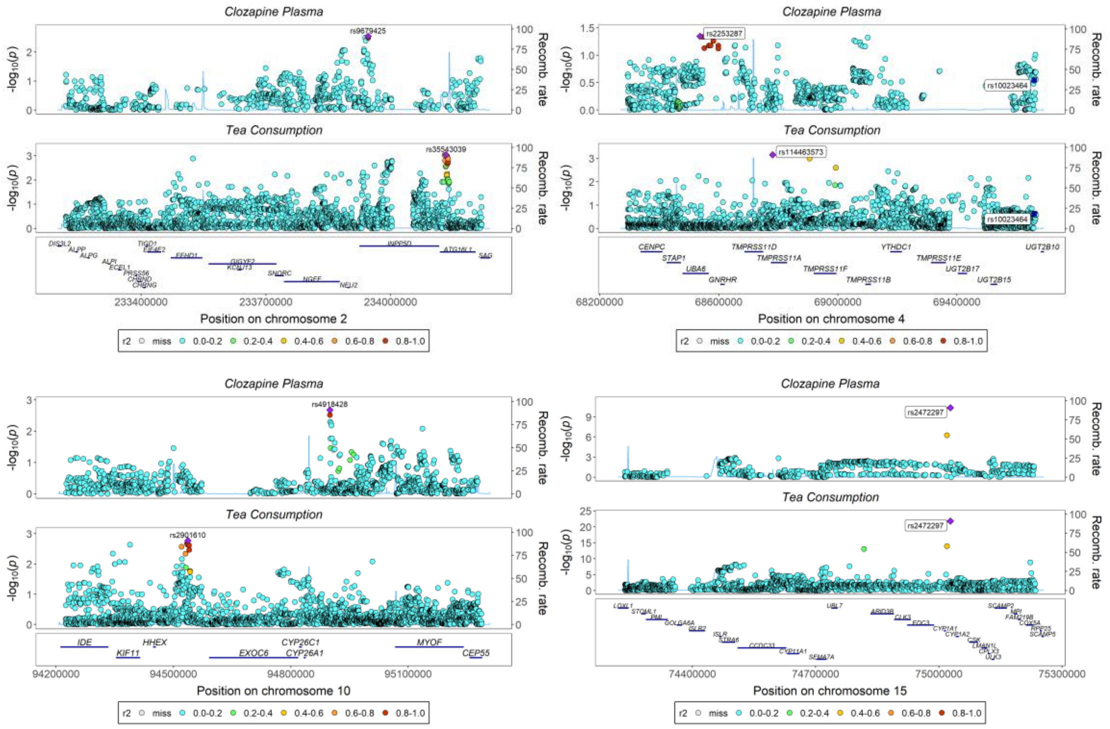

### Total Testosterone

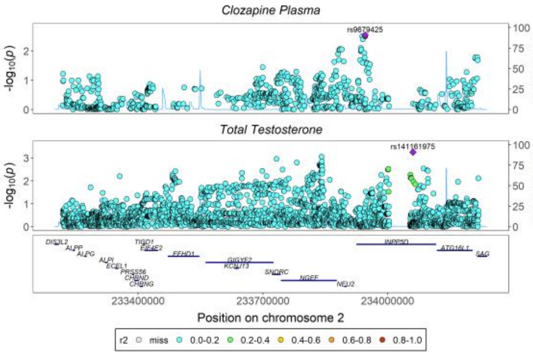

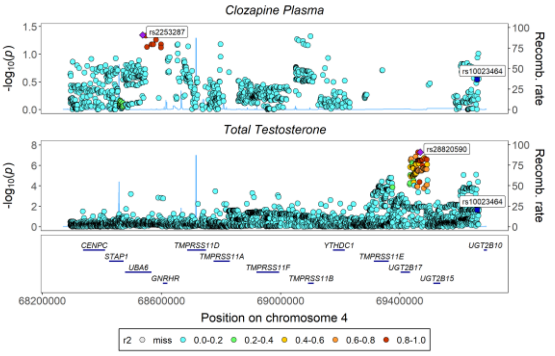

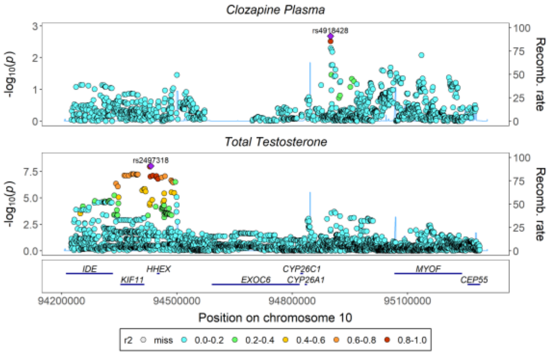

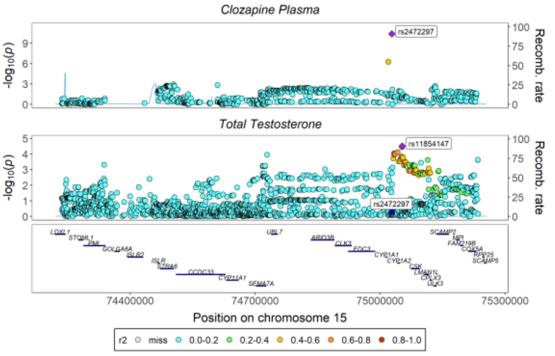

### Urinary Albumin Excretion

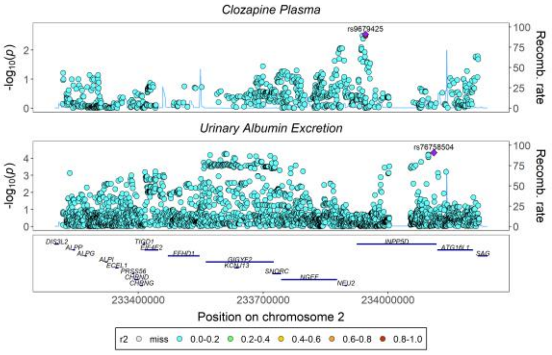

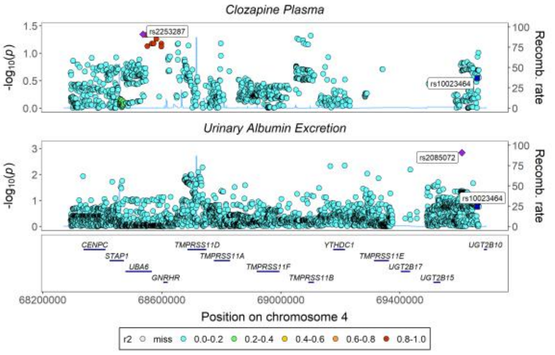

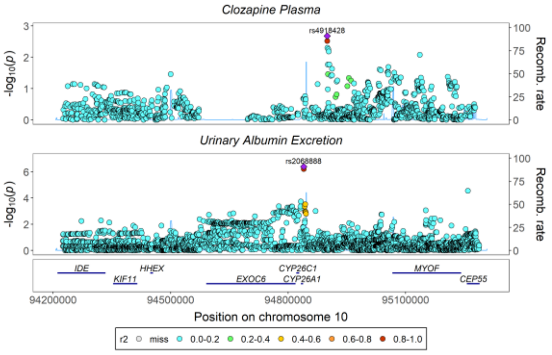

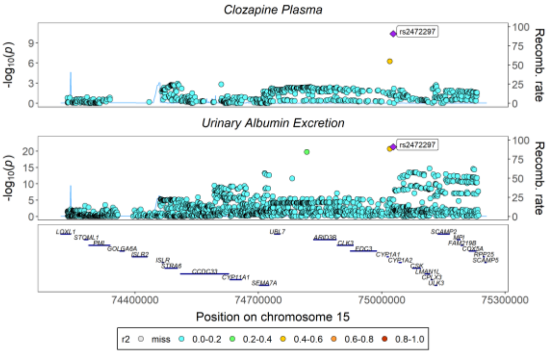

### Water Consumption

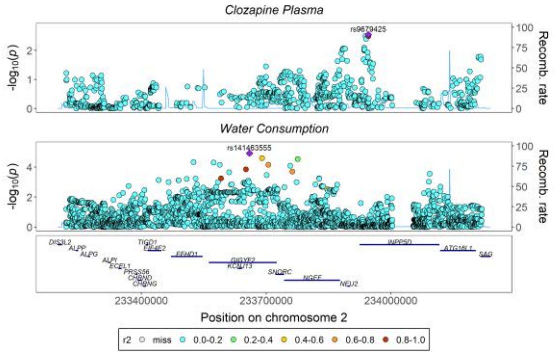

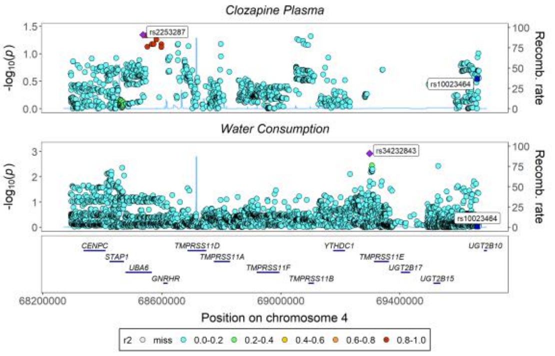

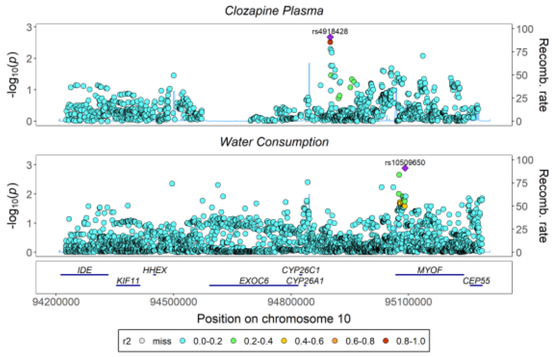

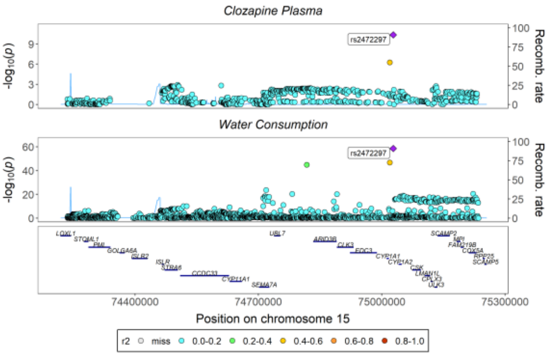

# X-11440

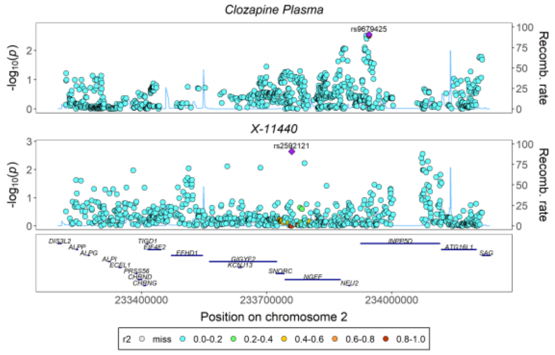

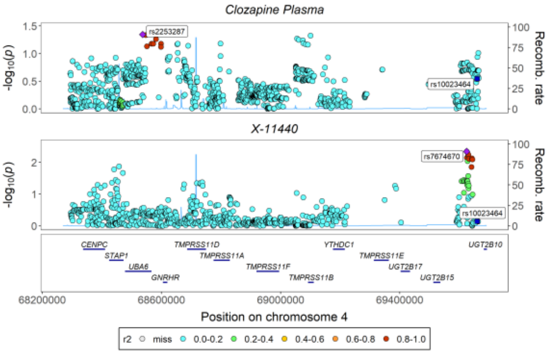

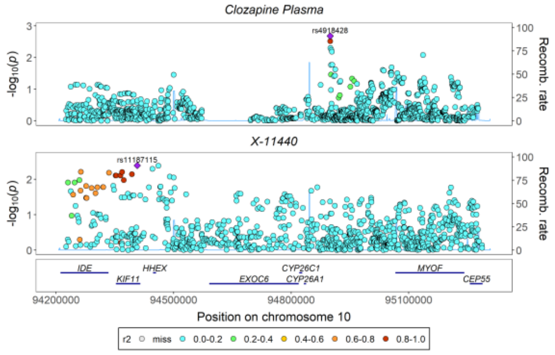

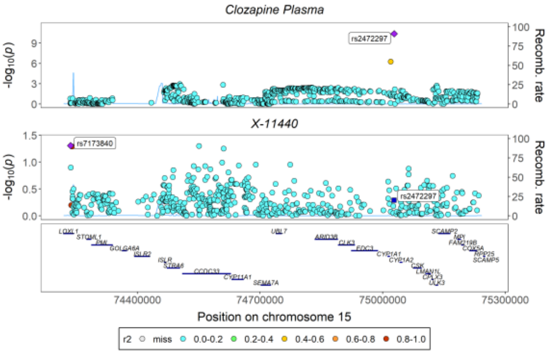

*Supplementary Figures 4. 4 – Stacked Regional Association Plots for Norclozapine Colocalizing with PheWAS Identified Candidate Biomarkers Across 4 Lead Chromosomes.*

#### 25-Hydroxyvitamin D

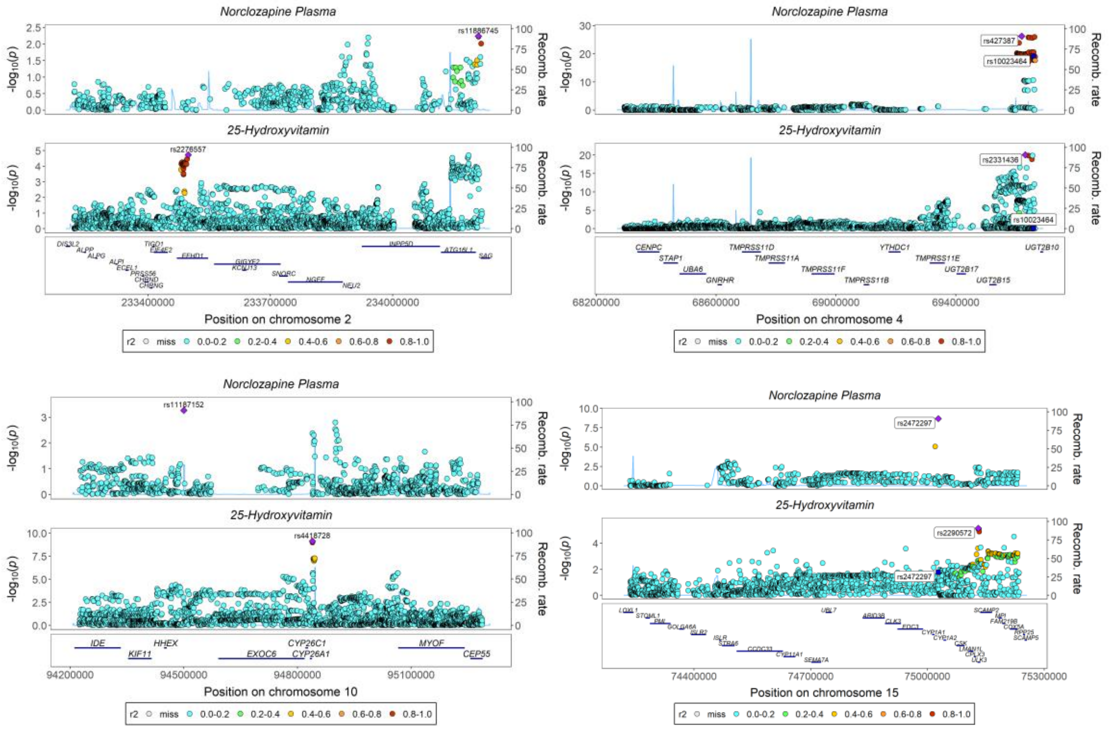

#### Bilirubin

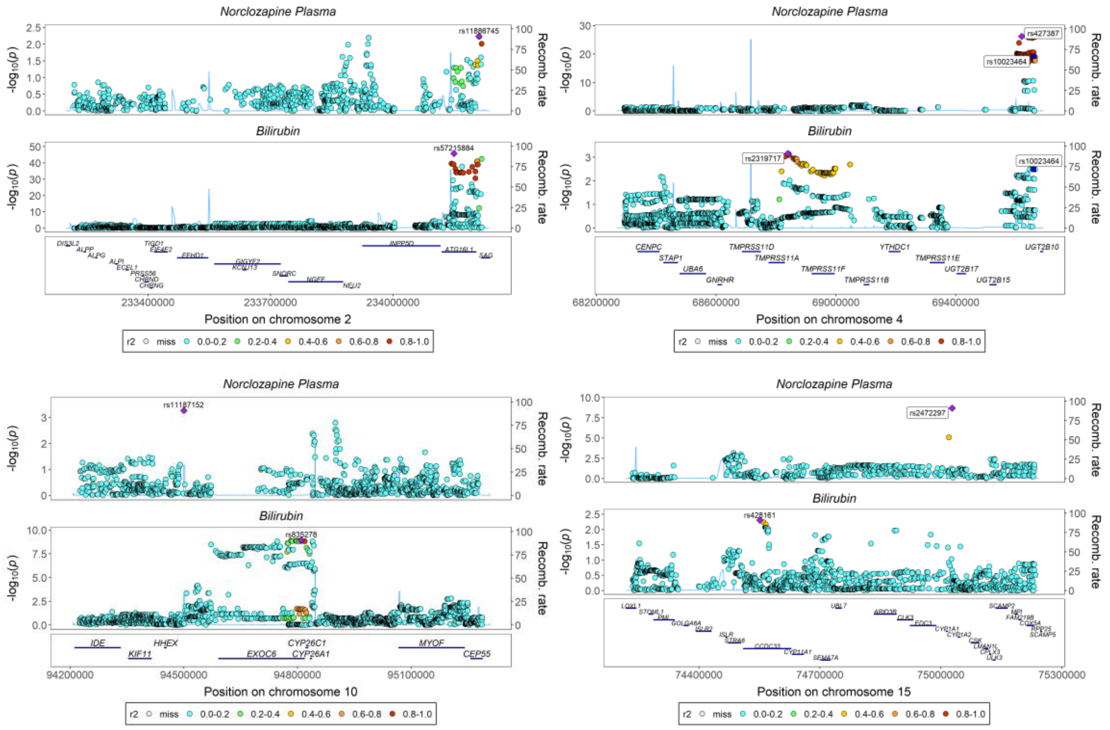

#### Blood Urea Nitrogen

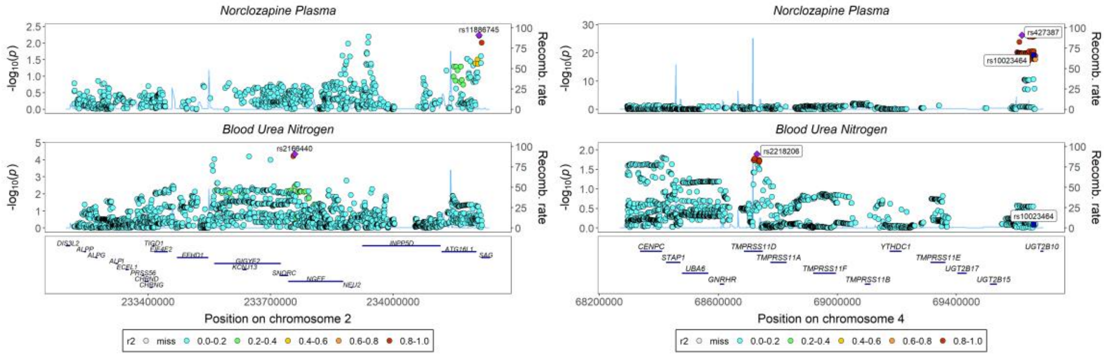

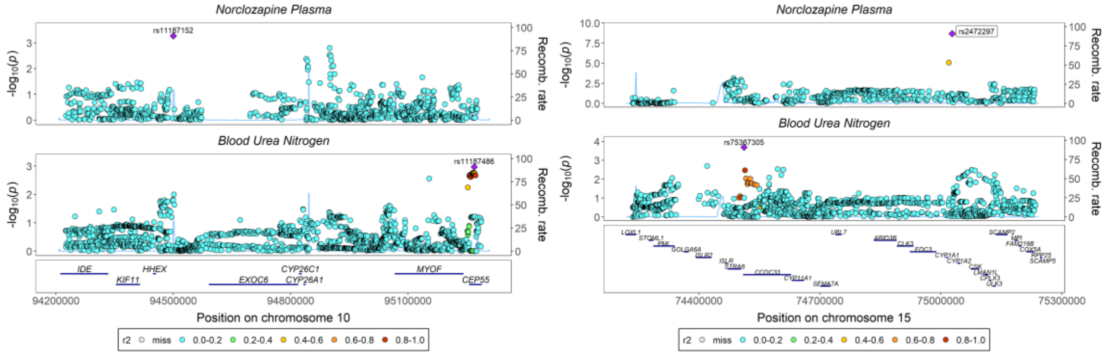

#### Coffee Consumption

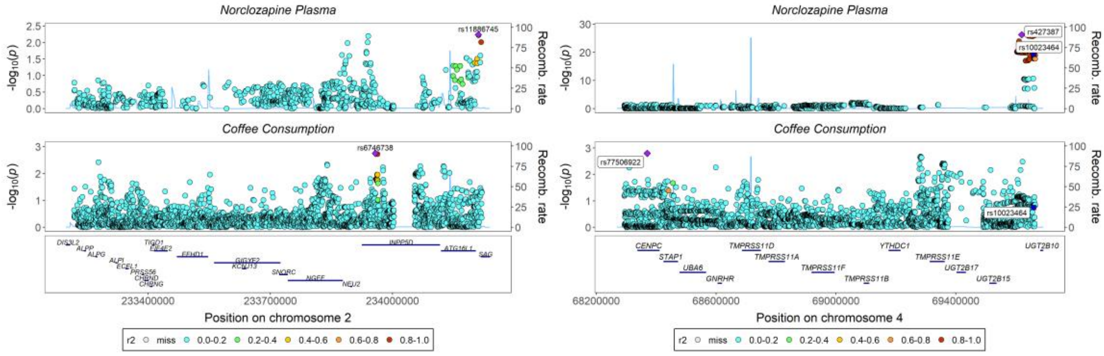

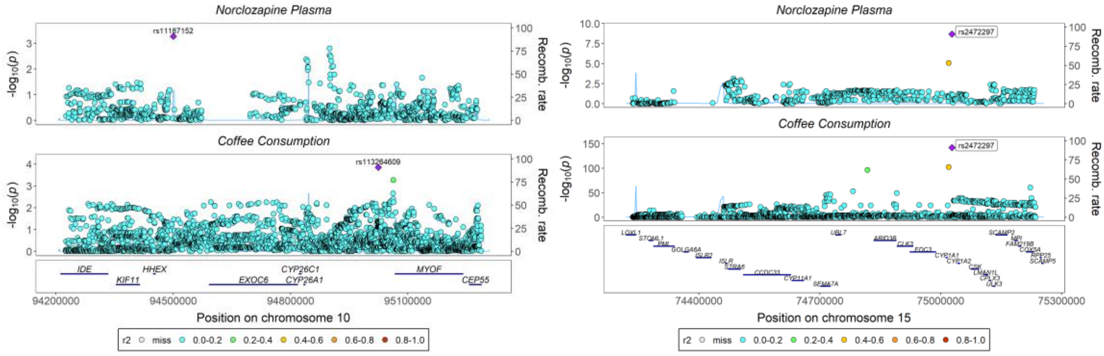

### Cotinine

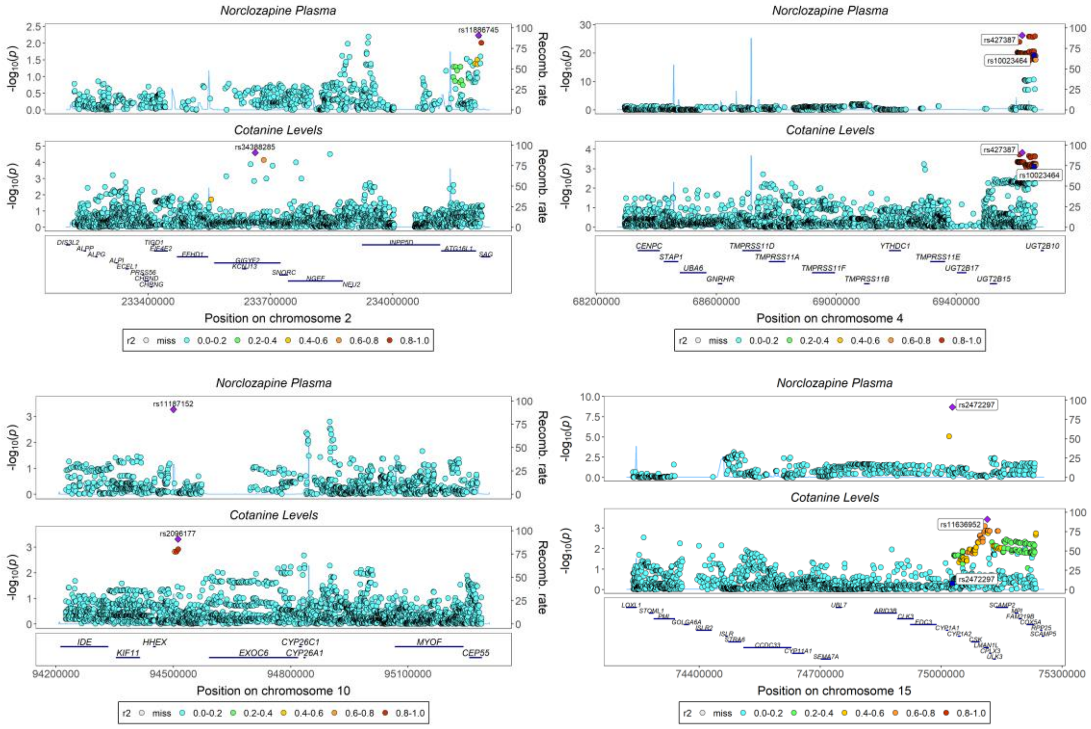

### Creatinine

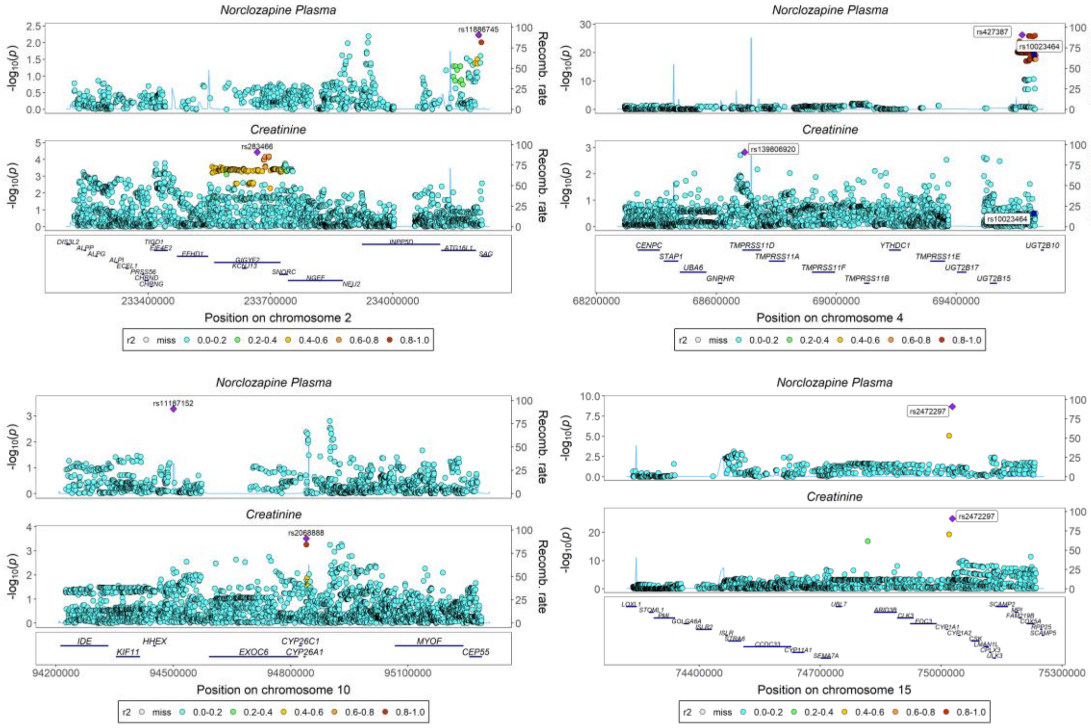

#### C-Reactive Protein

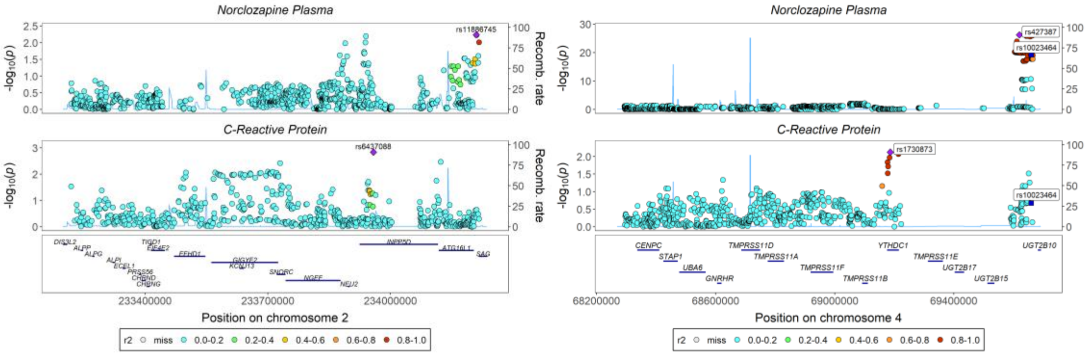

#### Cystatin C

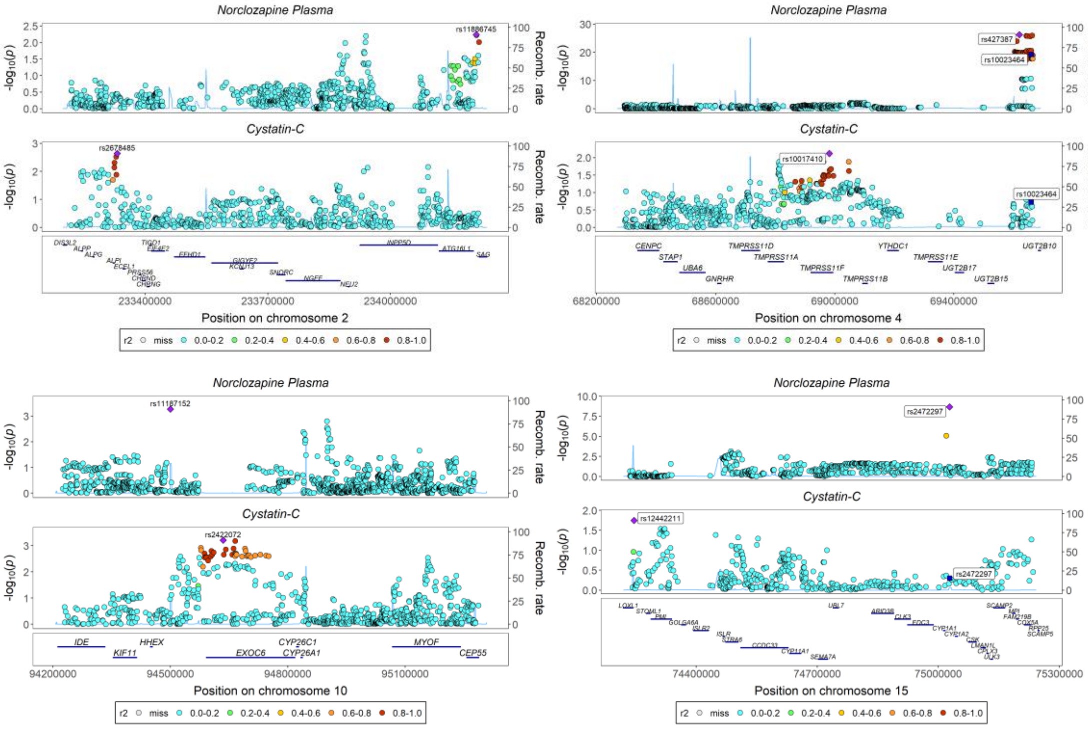

#### Fresh Fruit Consumption

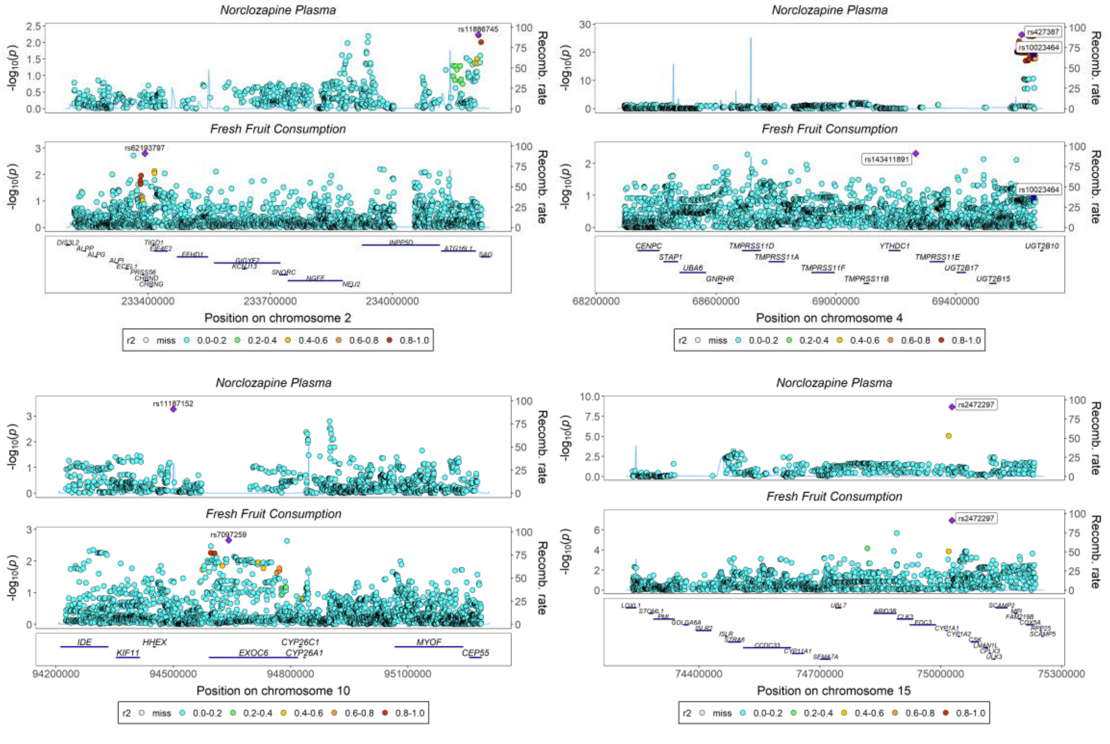

#### Gamma Glutamyl Transferase

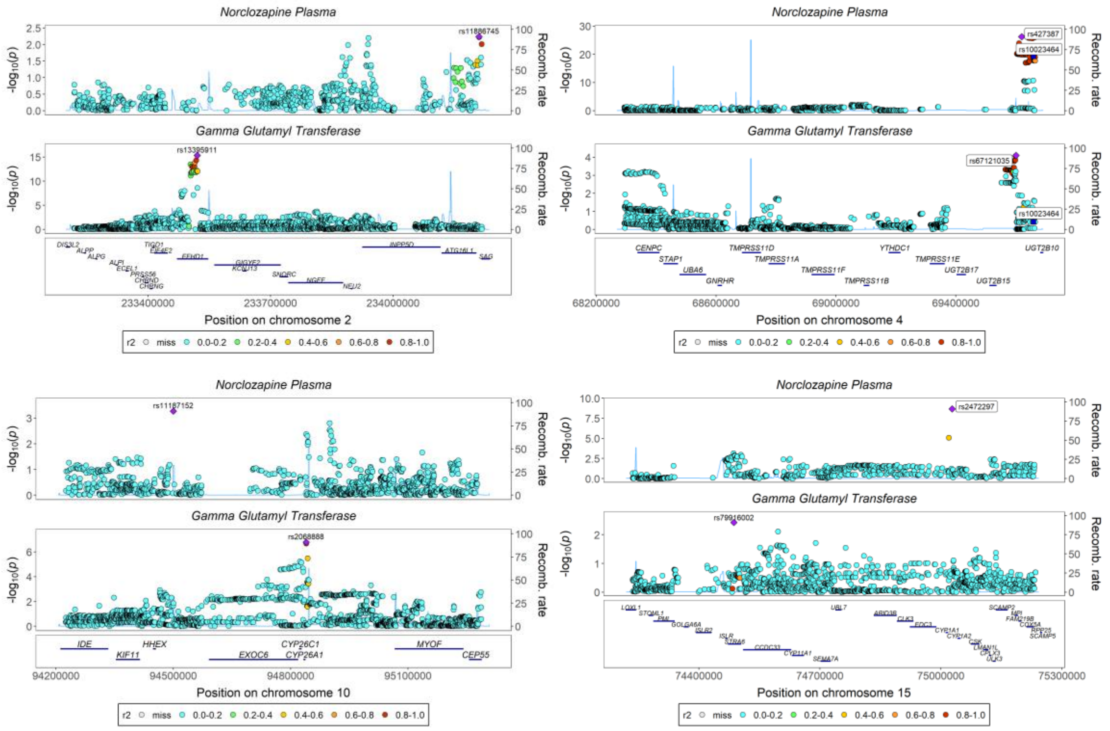

### Impedance of Left Arm

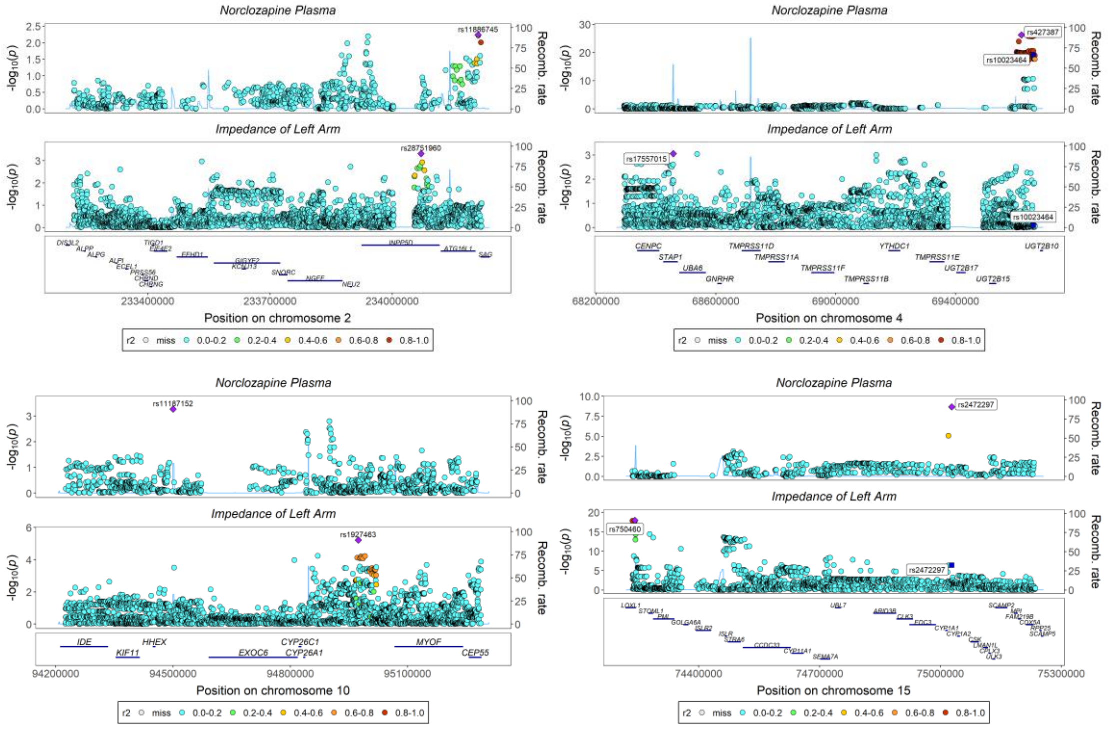

### Impedance of Right Arm

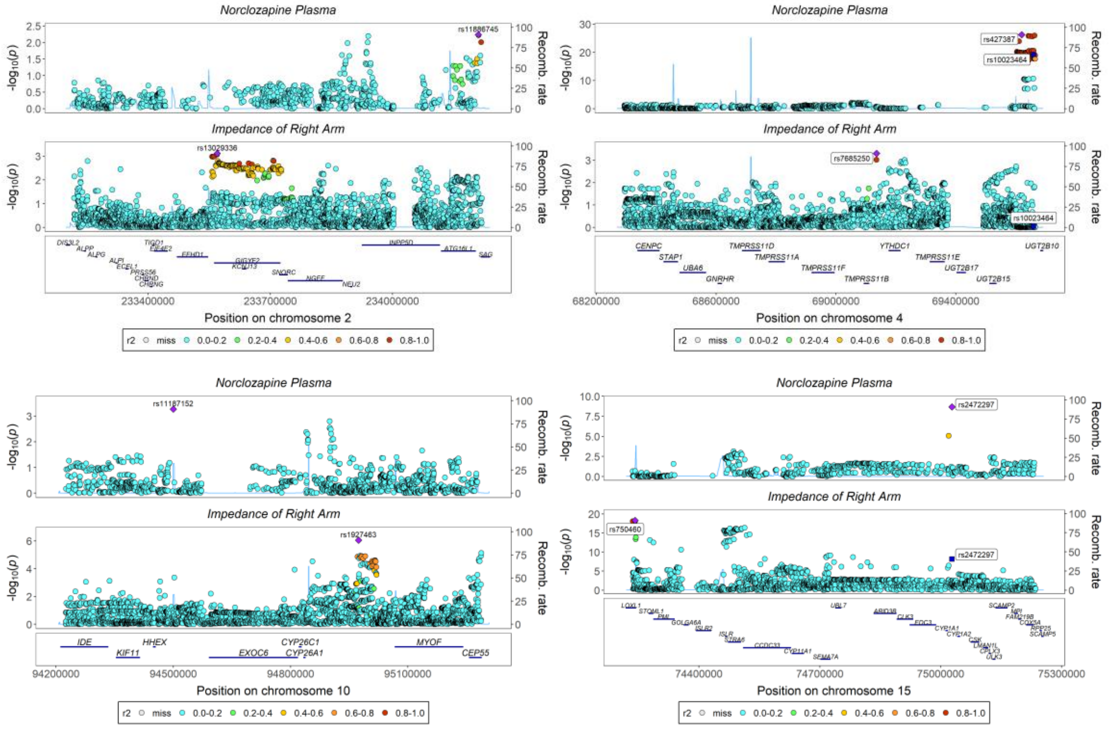

#### Overall Healthy Diet

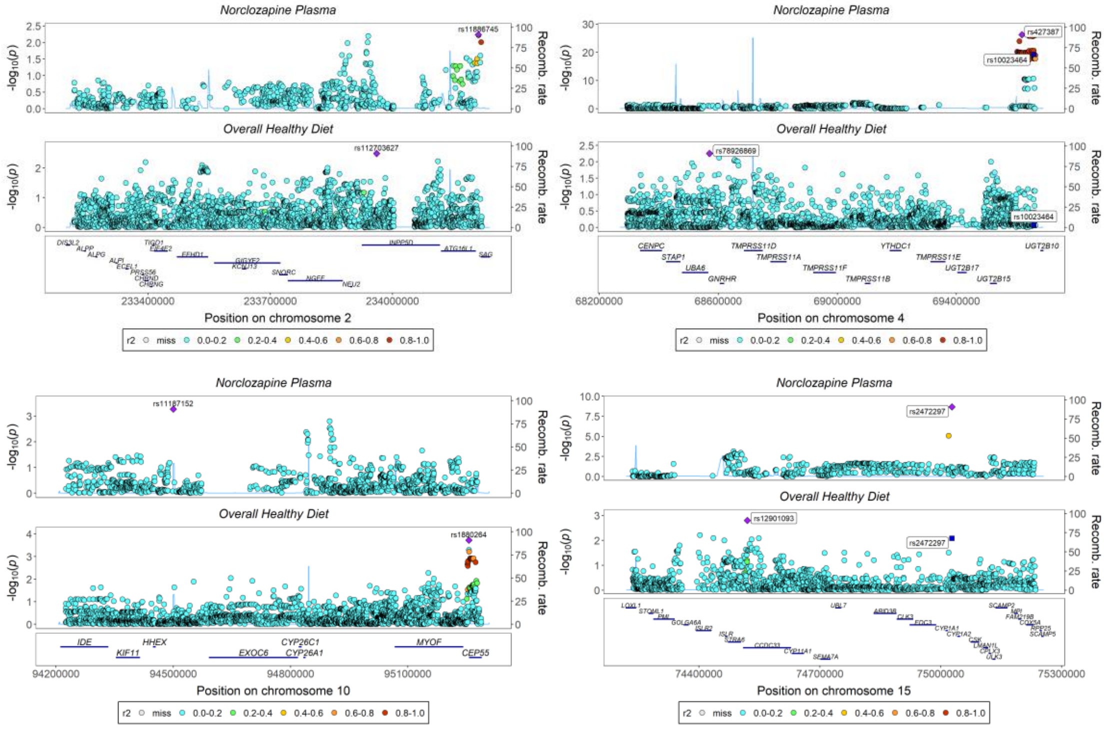

#### Peak Expiratory Flow

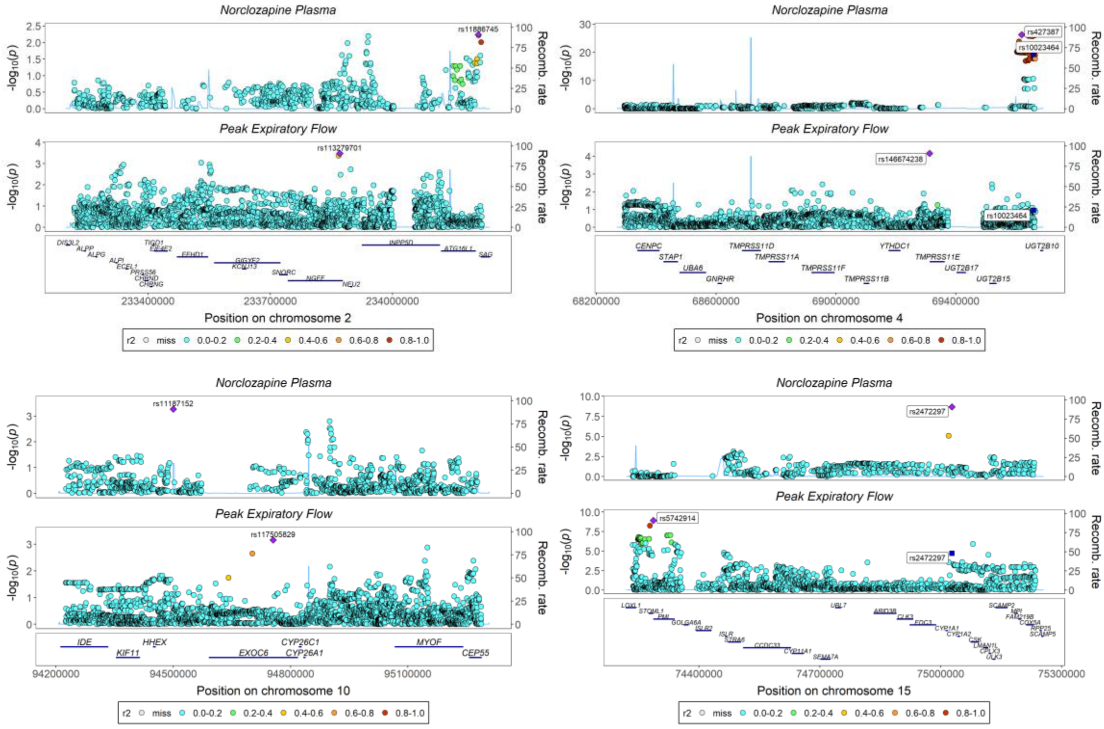

### Potassium in Urine

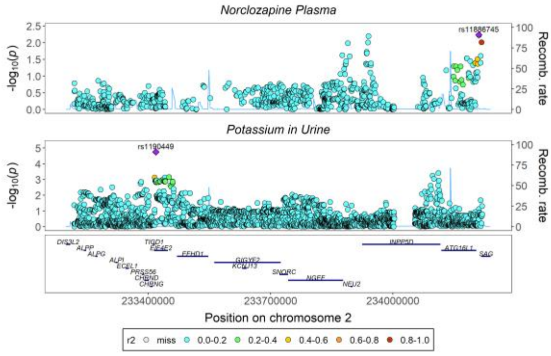

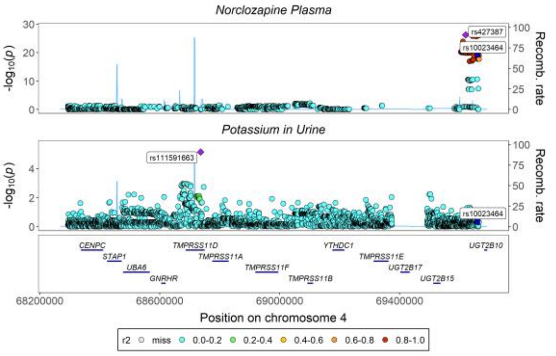

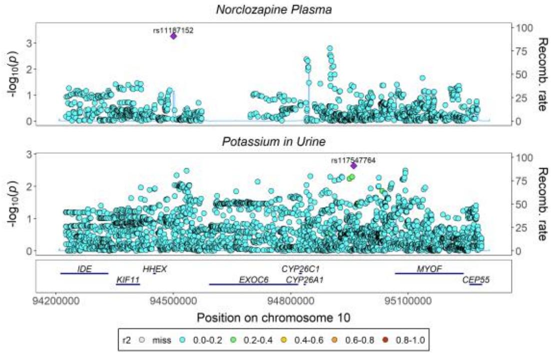

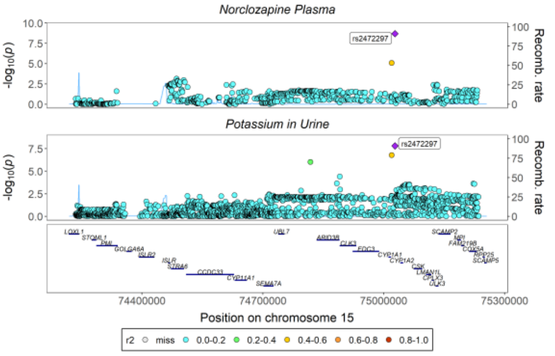

### Red Cell Distribution Width

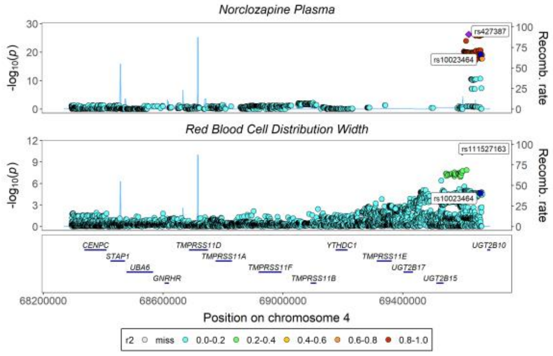

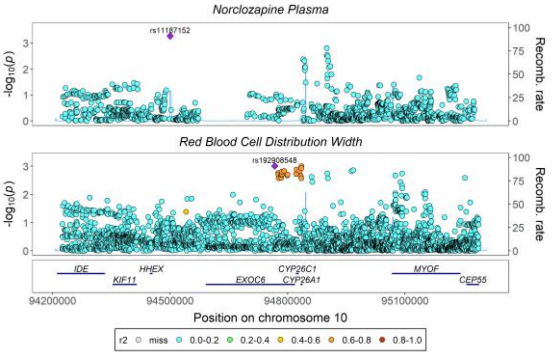

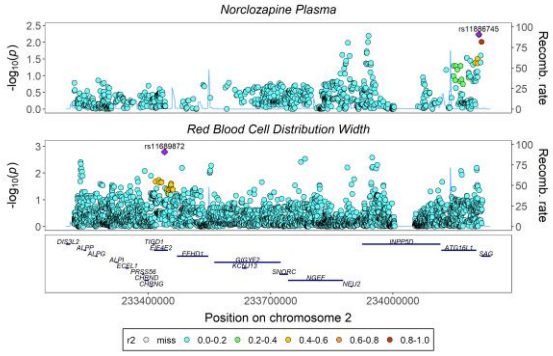

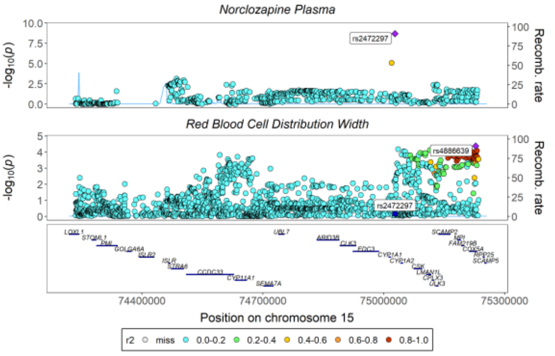

### Serum Phosphate

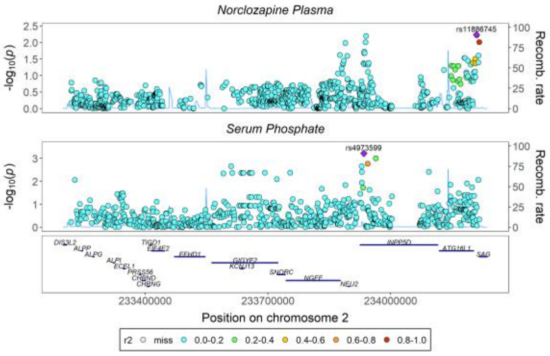

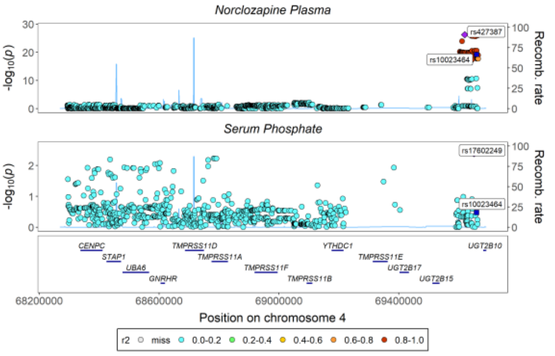

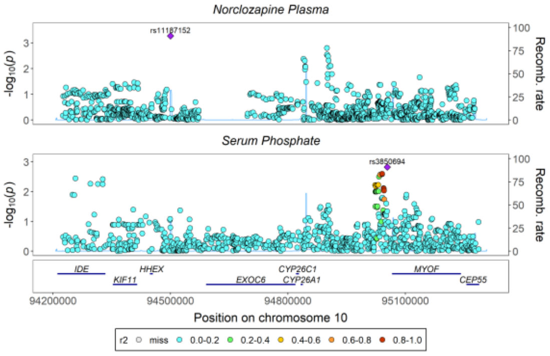

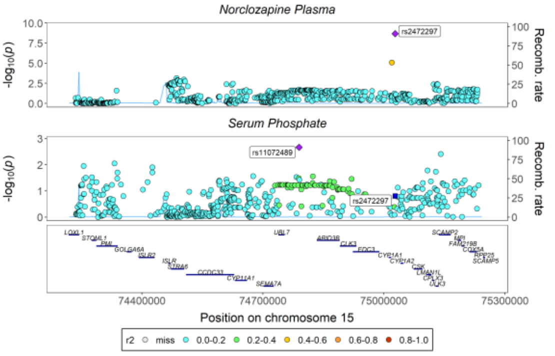

### Urea

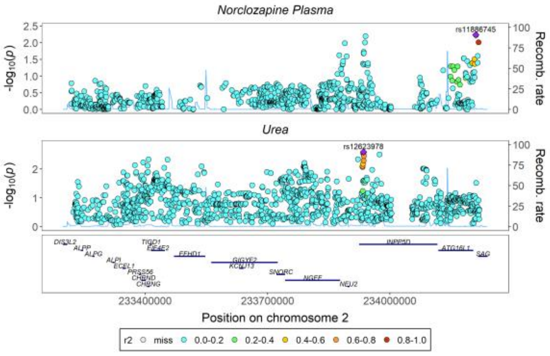

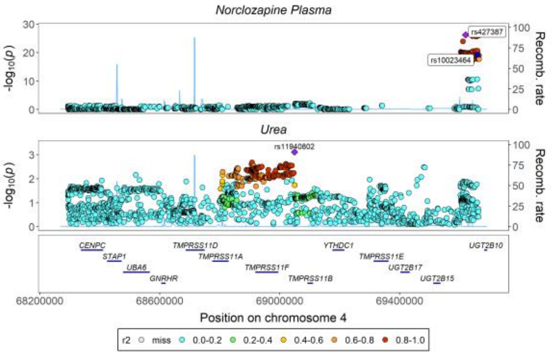

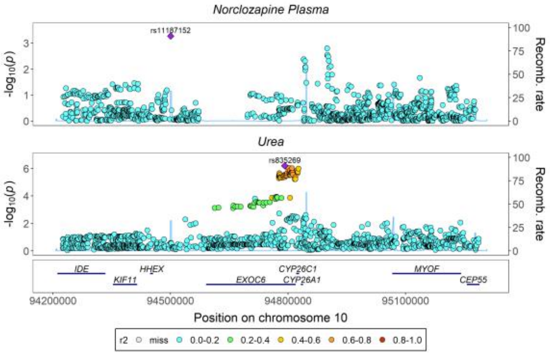

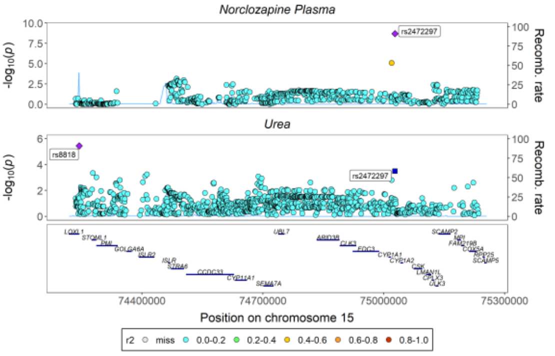

#### Uric Acid

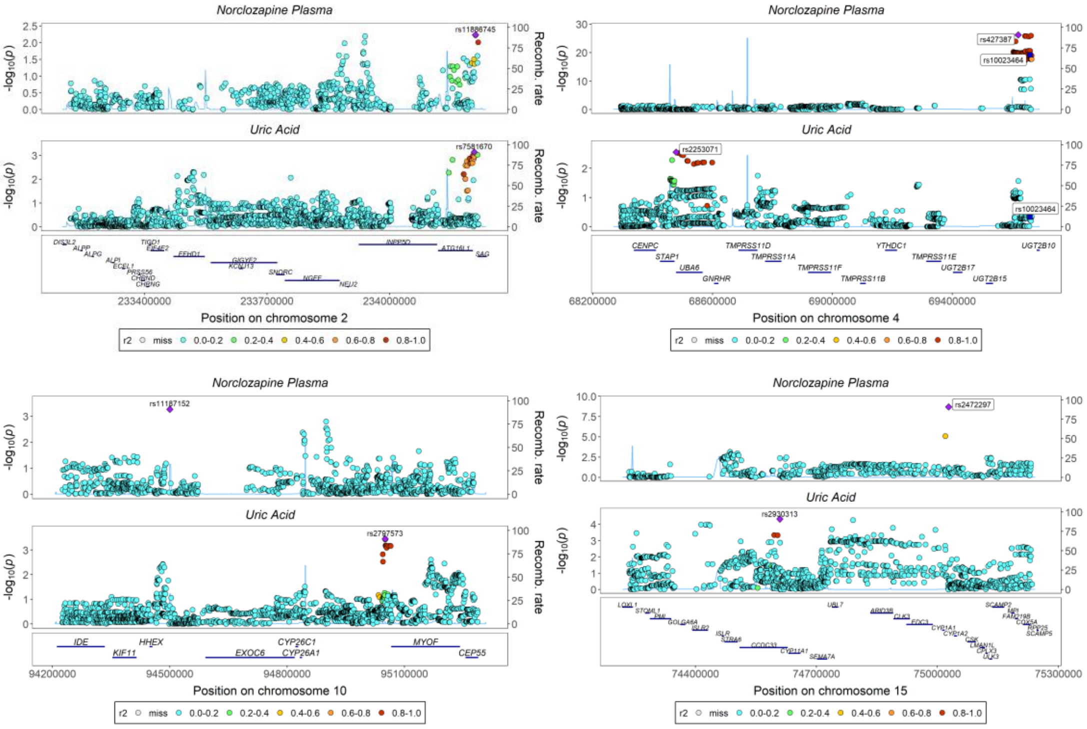

#### Sodium in Urine

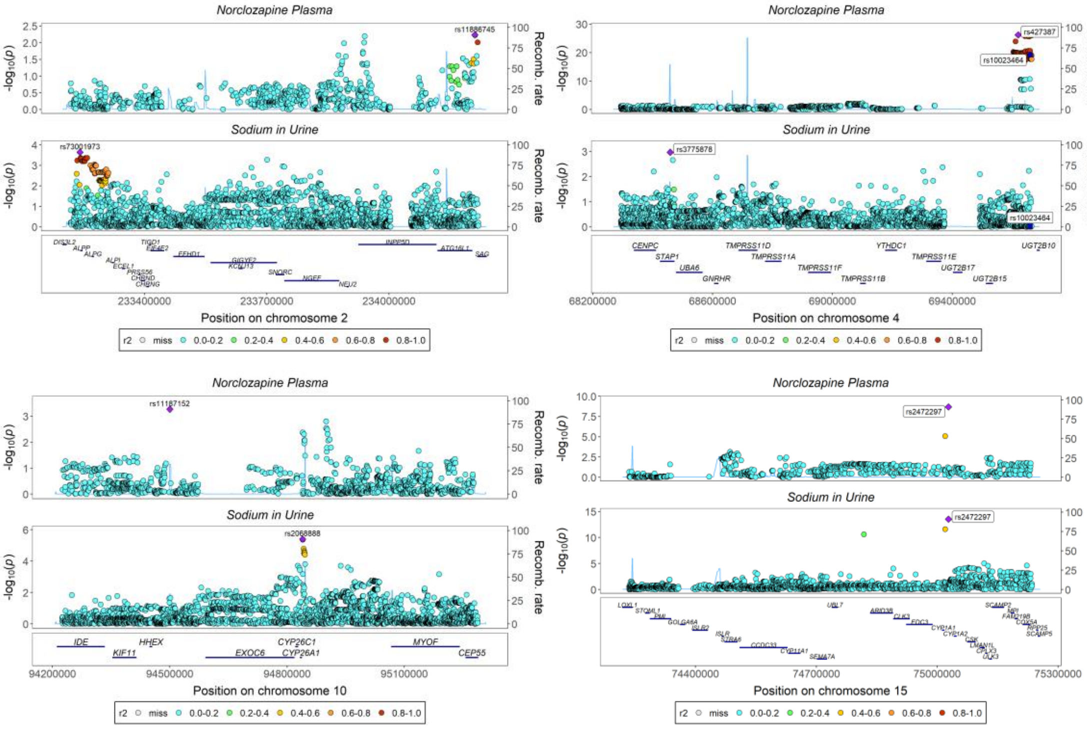

### Tea Consumption

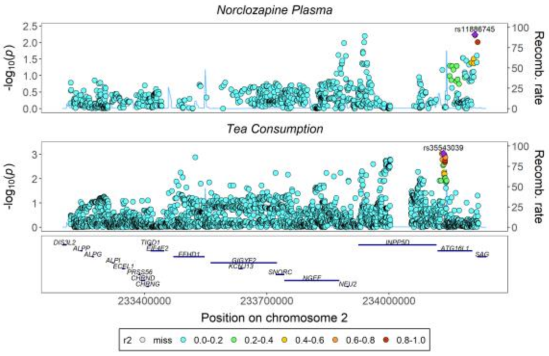

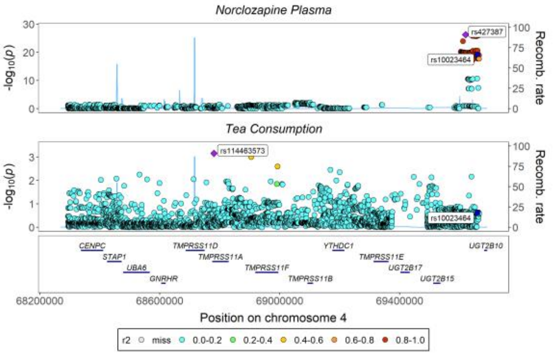

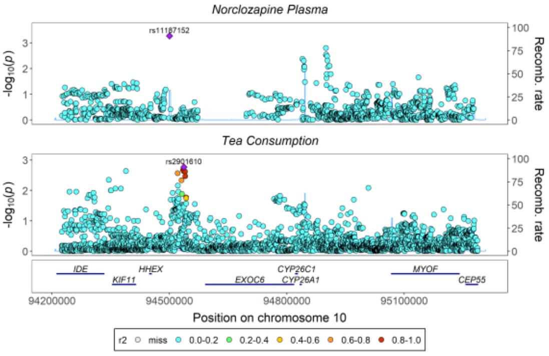

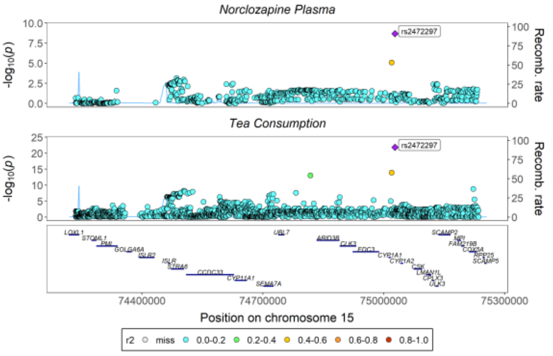

### Total Testosterone

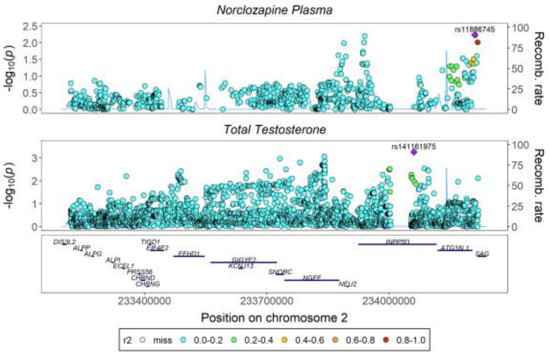

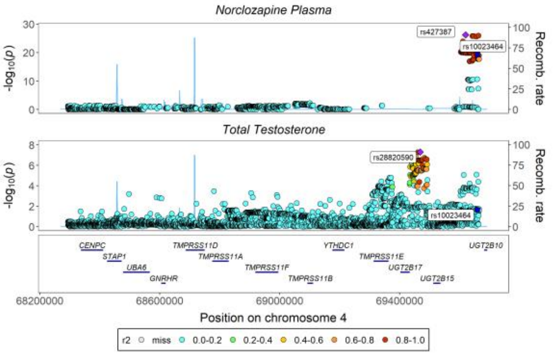

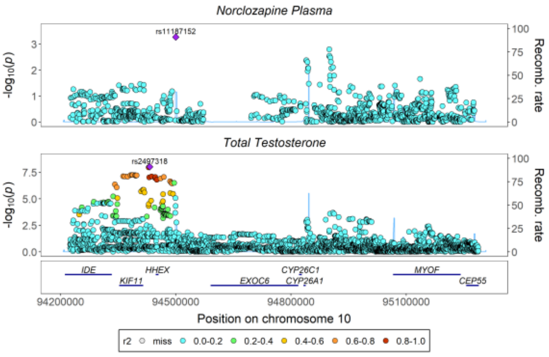

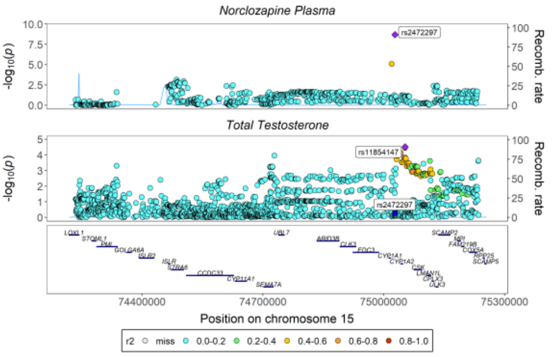

#### Urinary Albumin Excretion

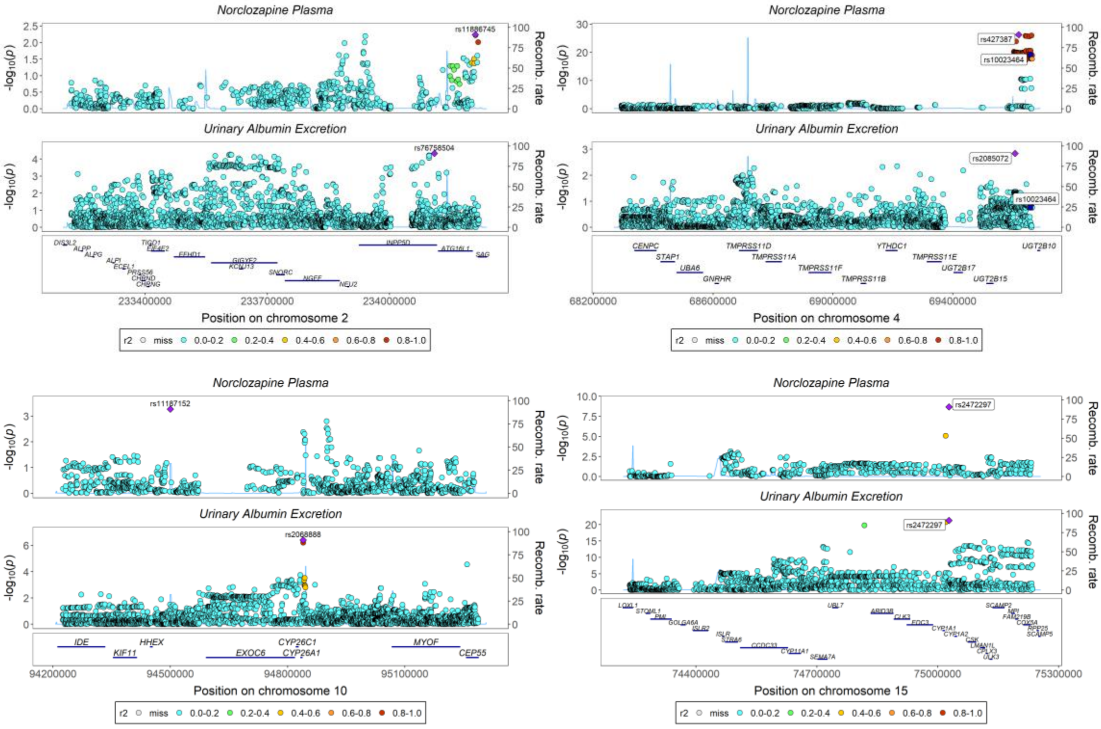

#### Water Consumption

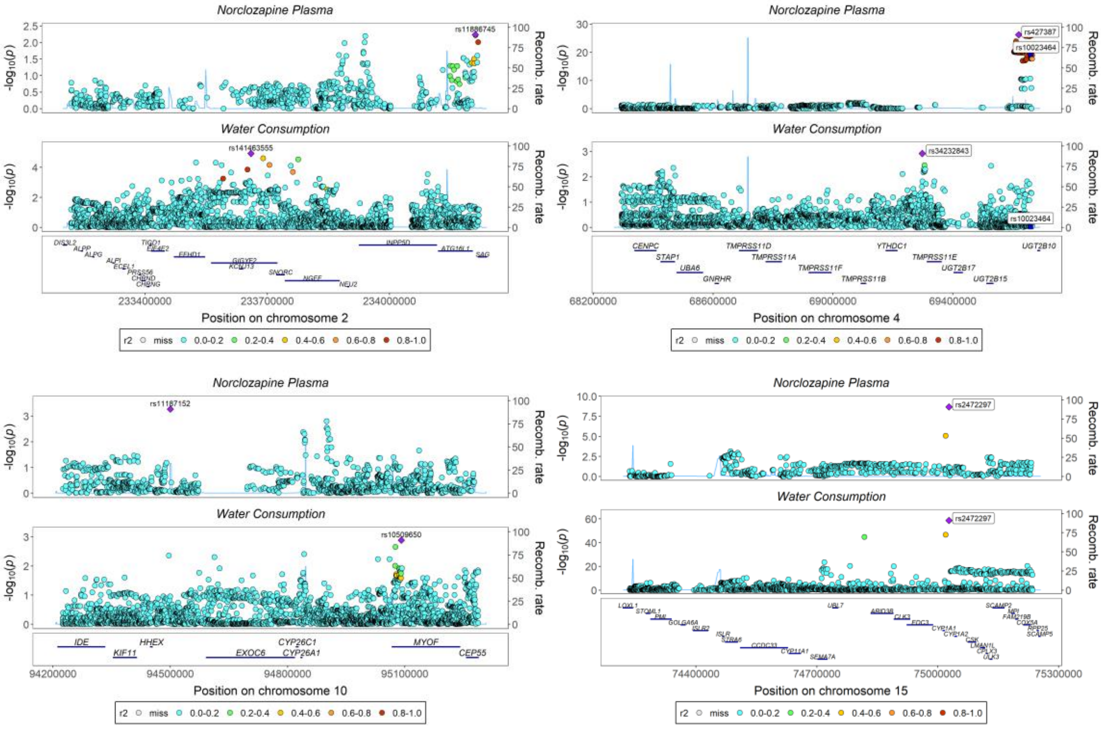

# X-11440

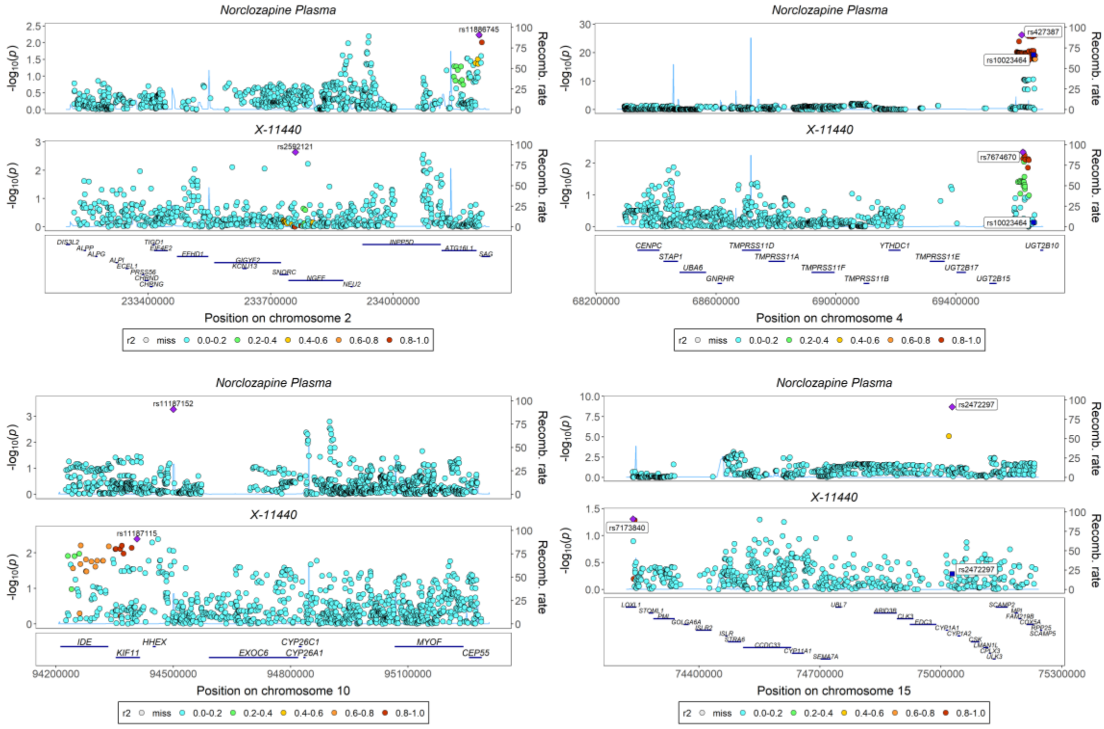

Norclozapine Plasma
X-11440
rs11886745
rs2592121
Position on chromosome 2
rs427387
rs10023464
rs7674670
Position on chromosome 4
rs11187152
rs11187115
Position on chromosome 10
rs2472297
rs7173840
Position on chromosome 15
-log10(p)
Recomb. rate
r2 miss 0.0-0.2 0.2-0.4 0.4-0.6 0.6-0.8 0.8-1.0

*Supplementary Figures 4. 5 - Stacked Regional Association Plots for Clozapine-Norclozapine Ratio Colocalizing with PheWAS Identified Candidate Biomarkers Across 4 Lead Chromosomes.*

#### Bilirubin

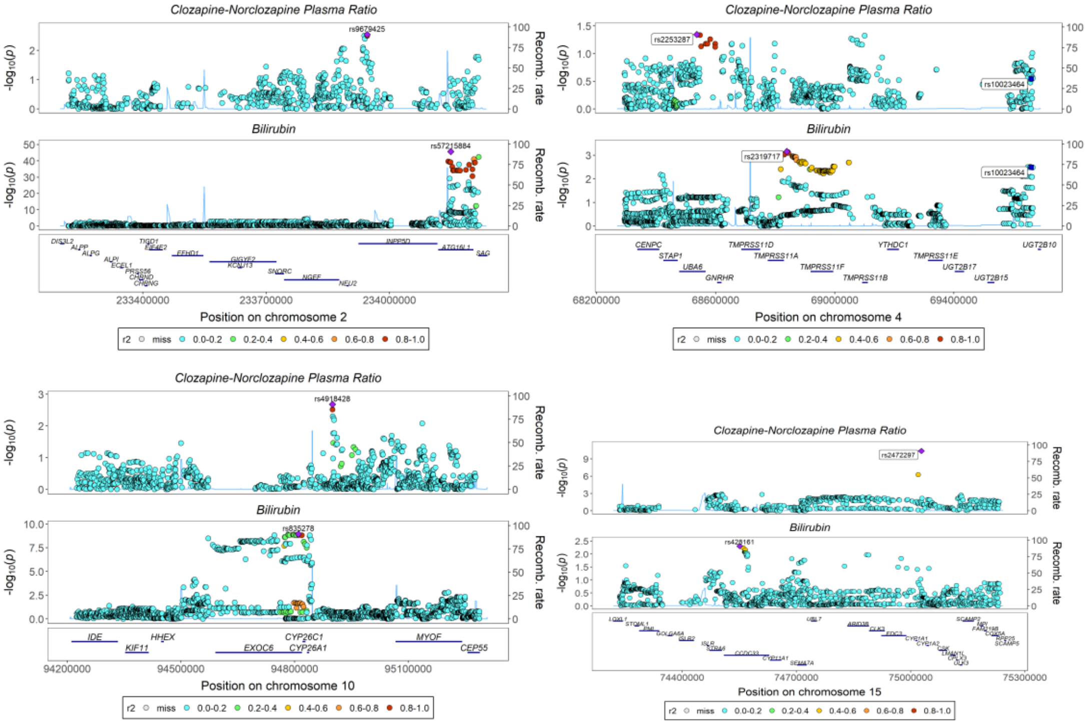

#### Mean Corpuscular Volume

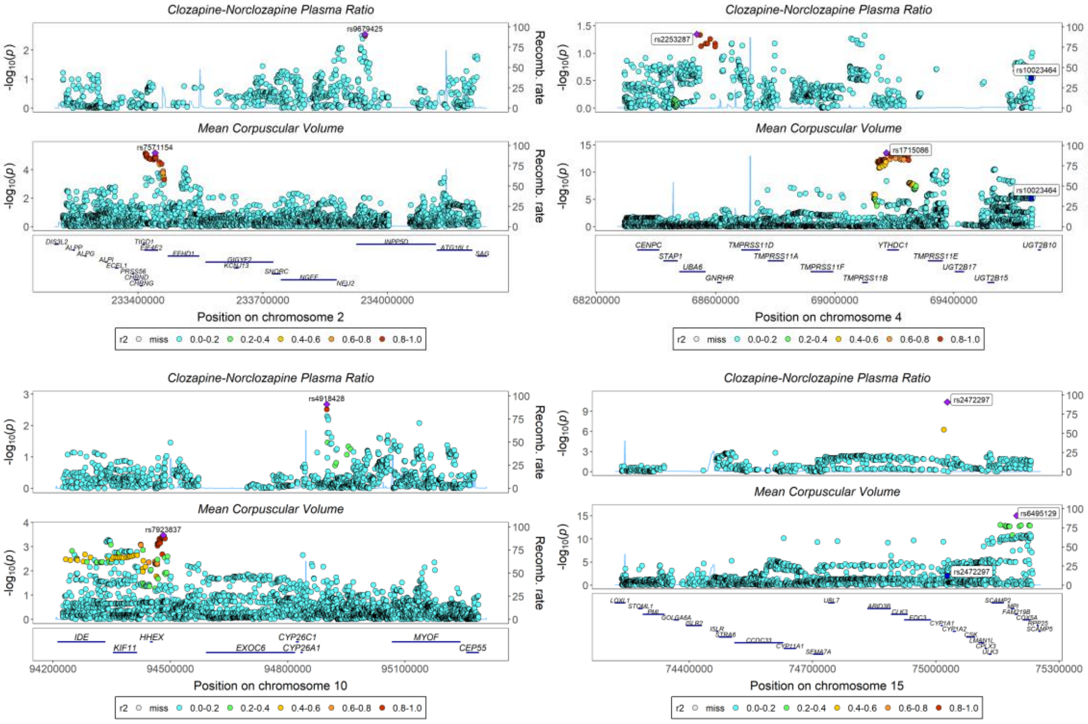

#### Total Testosterone

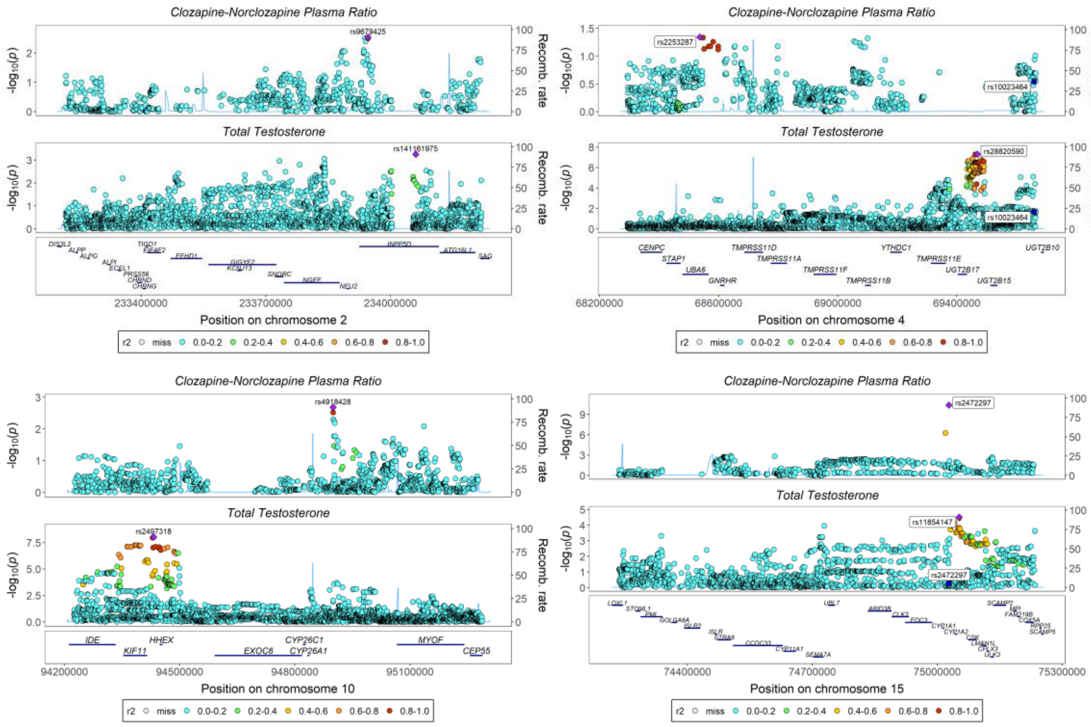

## X-14473

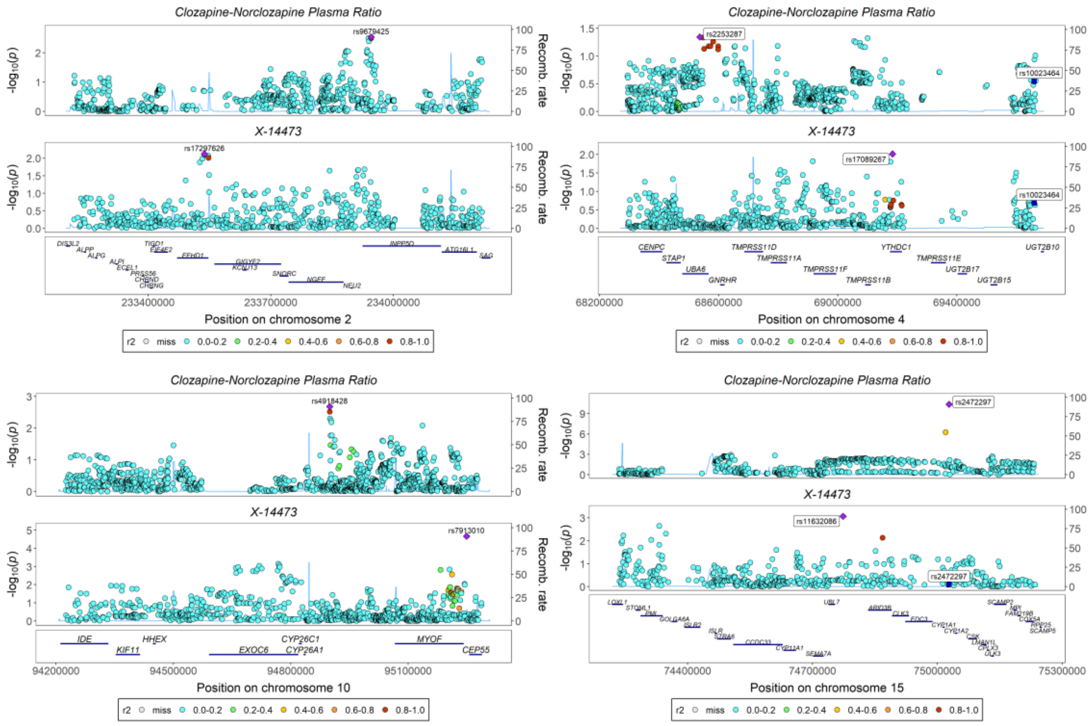

#### Supplementary Figures 5 – Candidate Biomarkers Effects on Cardiometabolic Traits

Analysis results from linkage-disequilibrium-aware bidirectional Mendelian randomization of clozapine on cardiometabolic outcomes where a colocalized phenotype ($\boldsymbol{H_4} > \mathbf{0.80}$) demonstrated significant causal effects on at least 1 of the 9 metabolic phenotypes after multiple testing correction (16/28 colocalized phenotypes). Forest plots demonstrate the causal estimate of the exposure ($\boldsymbol{\beta}$) on $\boldsymbol{n}$ outcomes with $\mathbf{95\%}\boldsymbol{CIs}$ and $\boldsymbol{Pvals}$. Significance threshold is adjusted for multiple testing (Bonferroni Correction) ($\boldsymbol{Pval}\ (\boldsymbol{adj}) = \mathbf{0.05}/\boldsymbol{n}$). Scatterplots depict sensitivity analyses results including MR-Egger and weighted median to test for directional pleiotropy and causal estimate assessment under invalid instruments respectively. Weighted mode demonstrates the causal estimate even if there are pleiotropic effects. Leave-one-out plots demonstrate the robustness of causal inference by testing if one genetic variant is driving the observed association. Raw results for bidirectional Mendelian randomization are available in Supplementary Table 5 and 6. Information about the Mendelian randomization analyses are available in the Supplementary Methods.

*Supplementary Figures 5. 1 – Forest Plots – Candidate Biomarkers on Cardiometabolic Traits*

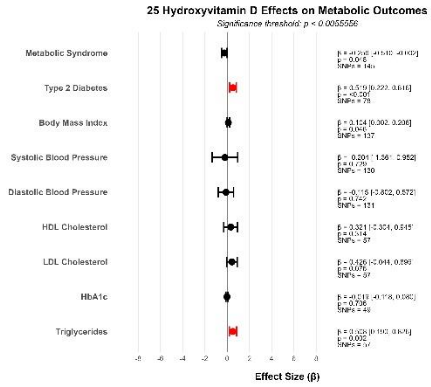

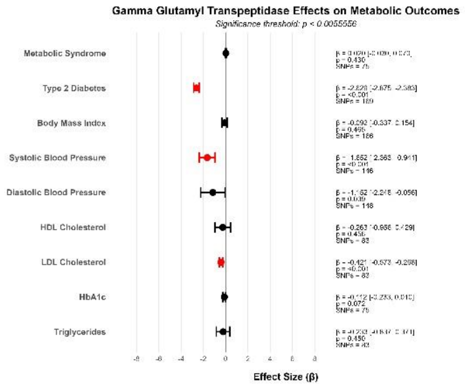

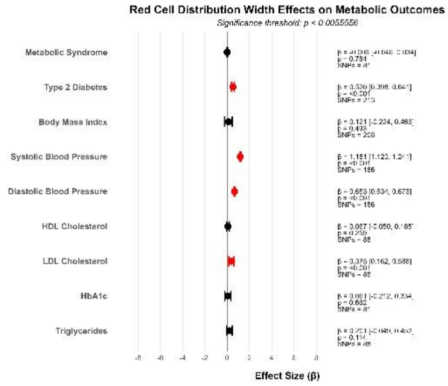

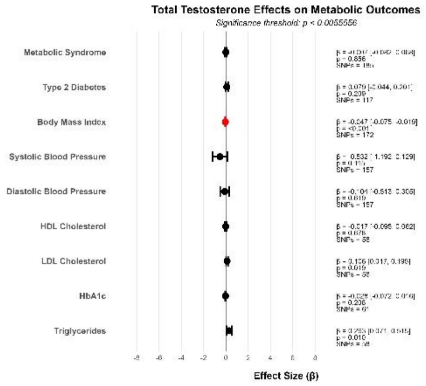

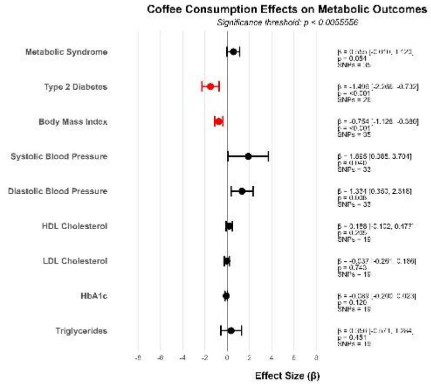

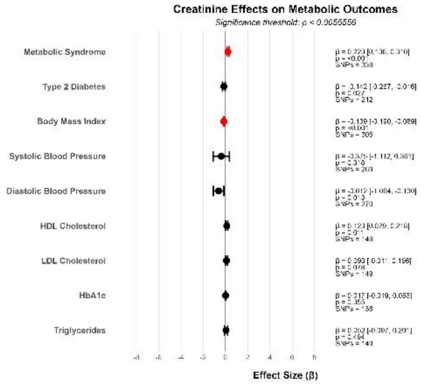

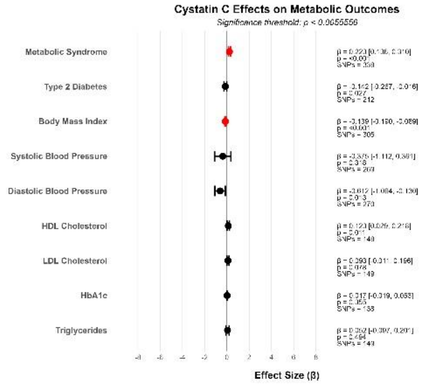

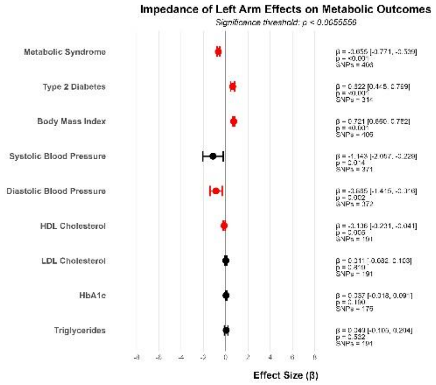

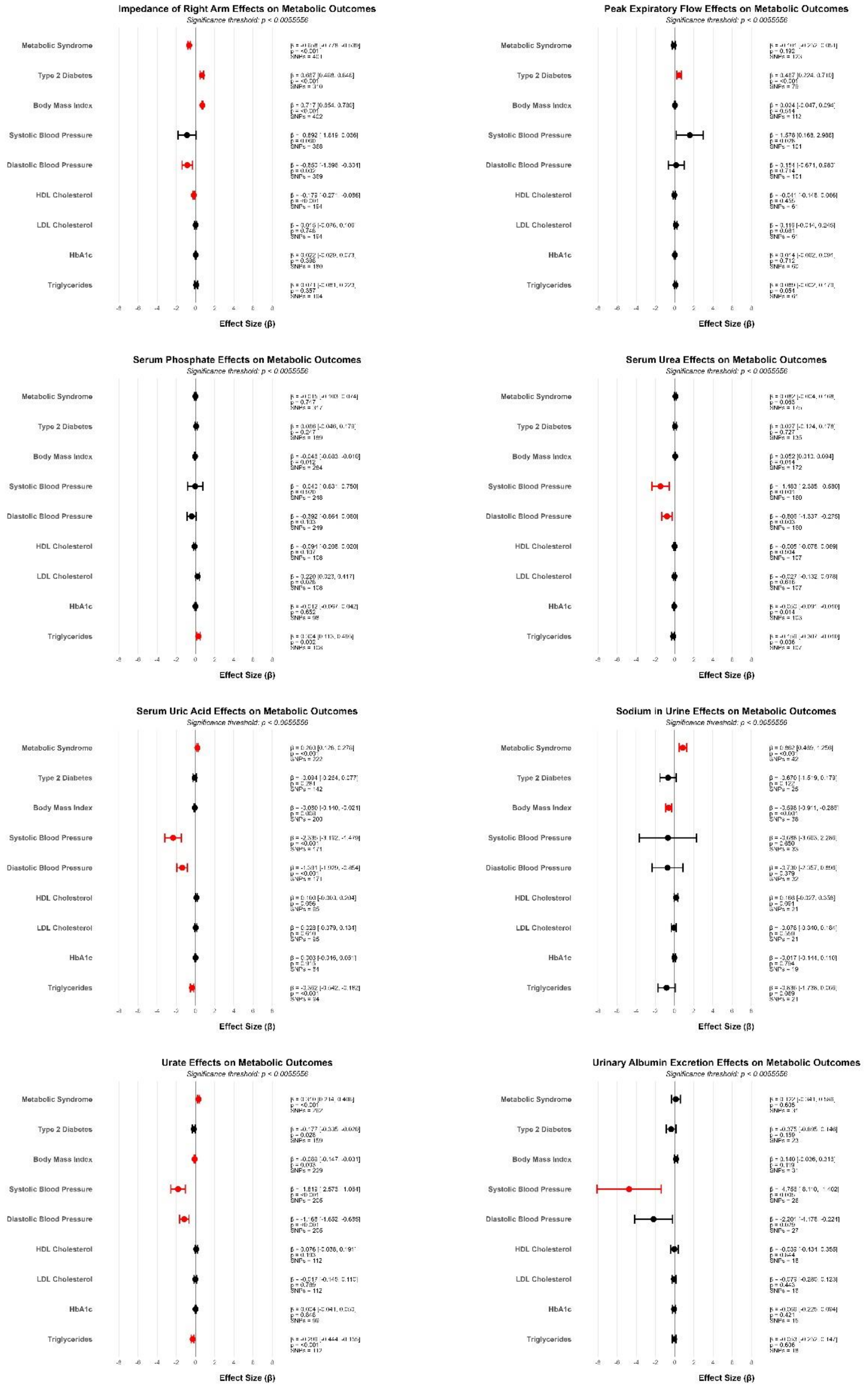
Impedance of Right Arm Effects on Metabolic Outcomes
Significance threshold: p < 0.0055556
Peak Expiratory Flow Effects on Metabolic Outcomes
Significance threshold: p < 0.0055556
Serum Phosphate Effects on Metabolic Outcomes
Significance threshold: p < 0.0055556
Serum Urea Effects on Metabolic Outcomes
Significance threshold: p < 0.0055556
Serum Uric Acid Effects on Metabolic Outcomes
Significance threshold: p < 0.0055556
Sodium in Urine Effects on Metabolic Outcomes
Significance threshold: p < 0.0055556
Urate Effects on Metabolic Outcomes
Significance threshold: p < 0.0055556
Urinary Albumin Excretion Effects on Metabolic Outcomes
Significance threshold: p < 0.0055556
Metabolic Syndrome
Type 2 Diabetes
Body Mass Index
Systolic Blood Pressure
Diastolic Blood Pressure
HDL Cholesterol
LDL Cholesterol
HbA1c
Triglycerides
Effect Size (β)

*Supplementary Figures 5. 2 – Scatterplots – Candidate Biomarkers on Cardiometabolic Traits*

#### 25-Hydroxyvitamin D

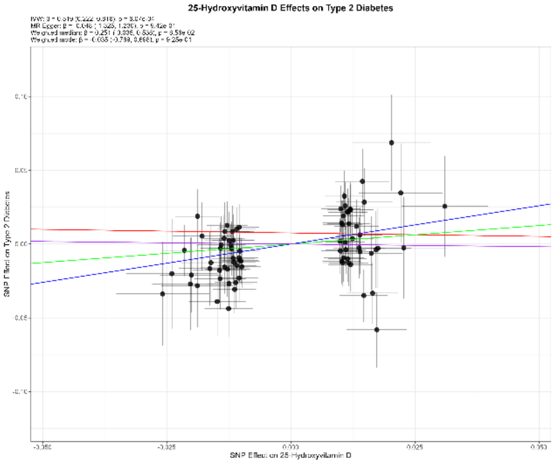

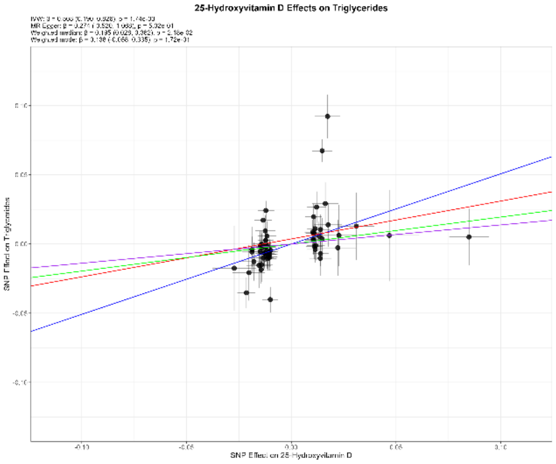

#### Gamma Glutamyl Transferase

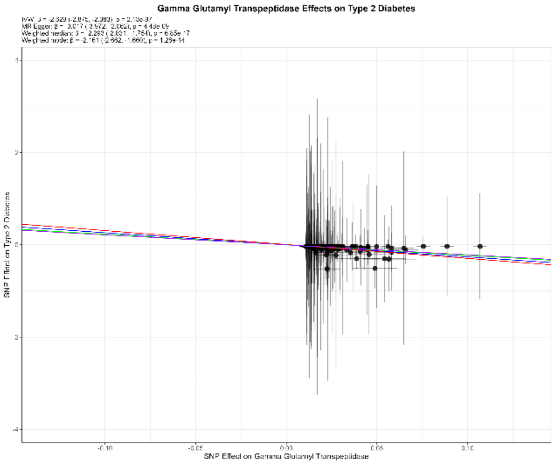

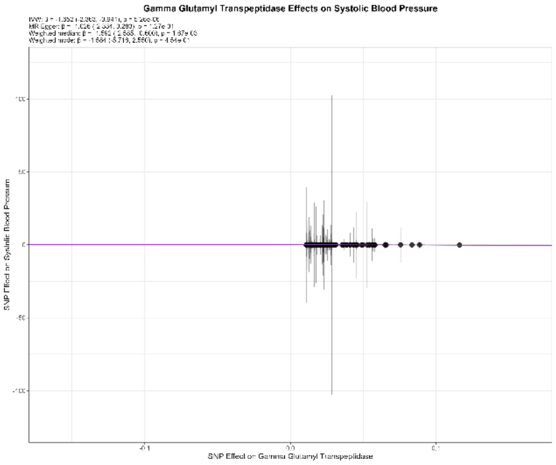

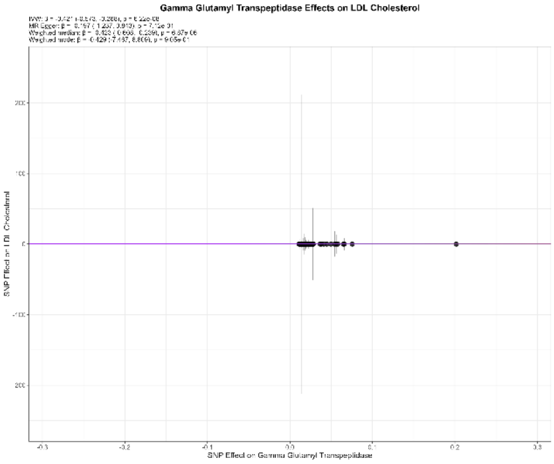

#### Red Cell Distribution Width

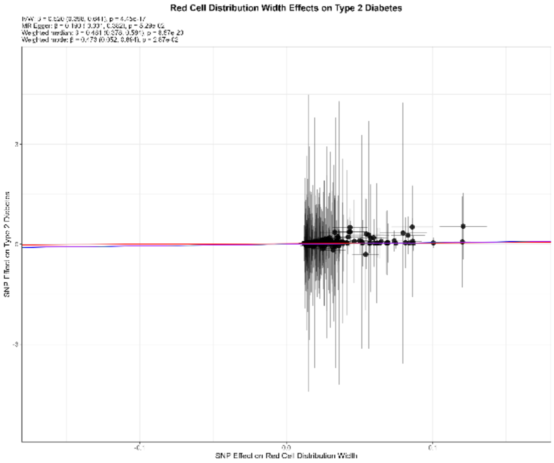

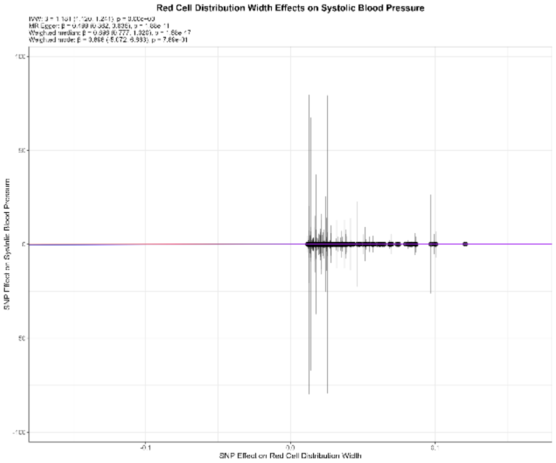

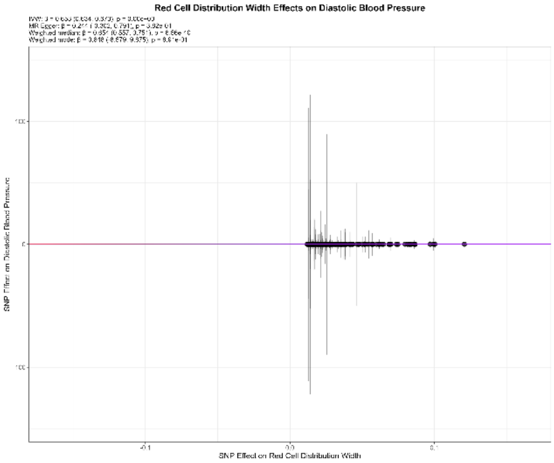

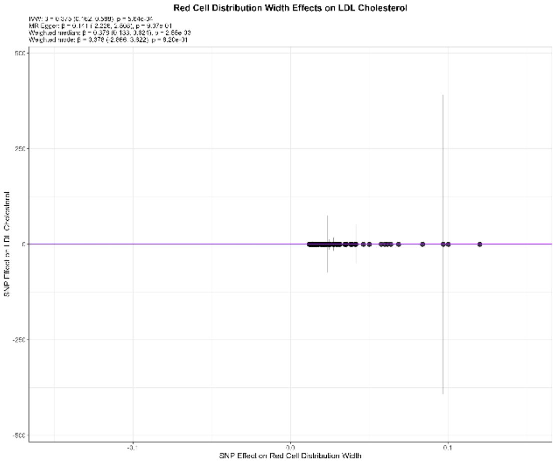

#### Total Testosterone

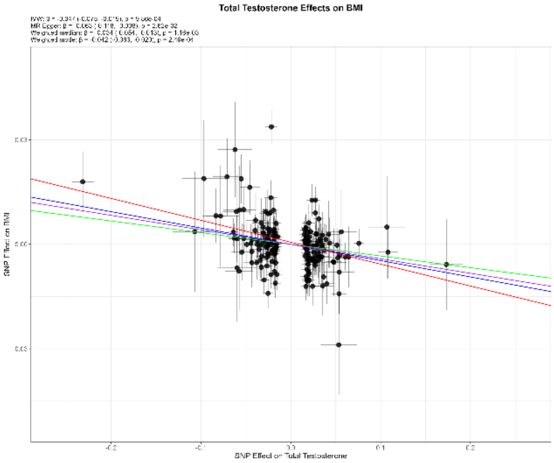

#### Coffee Consumption

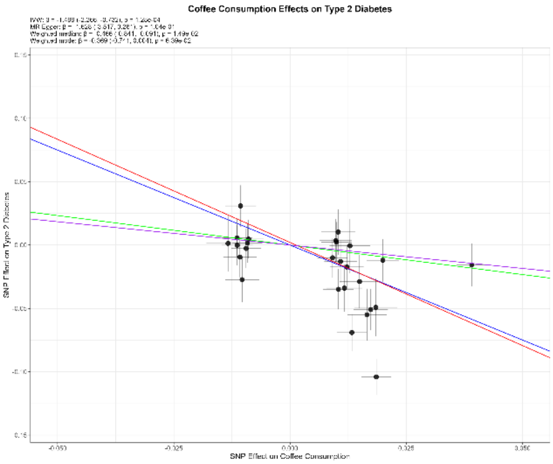

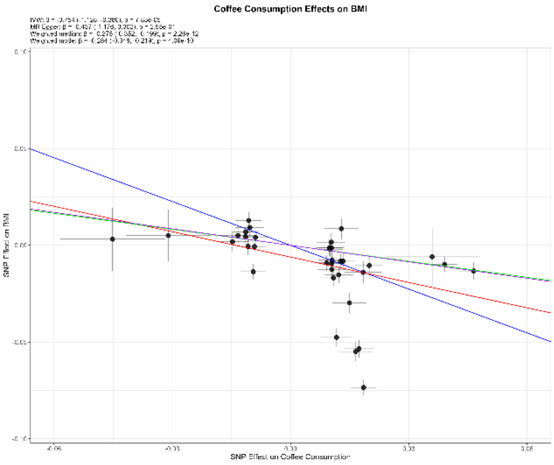

#### Creatinine

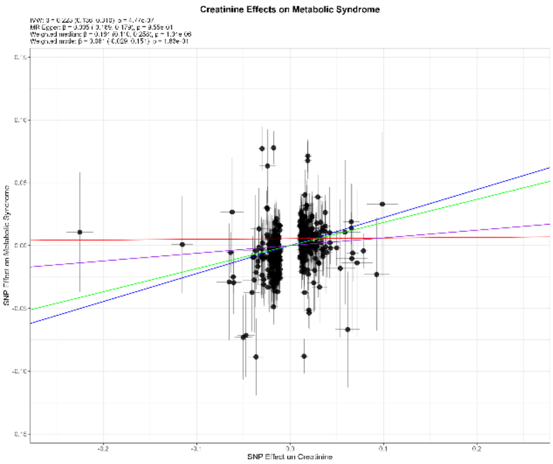

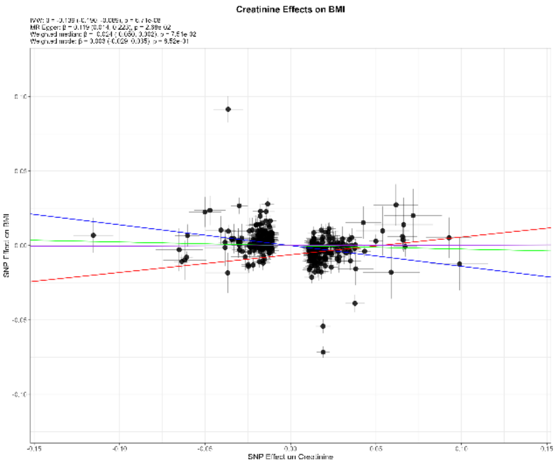

#### Cystatin C

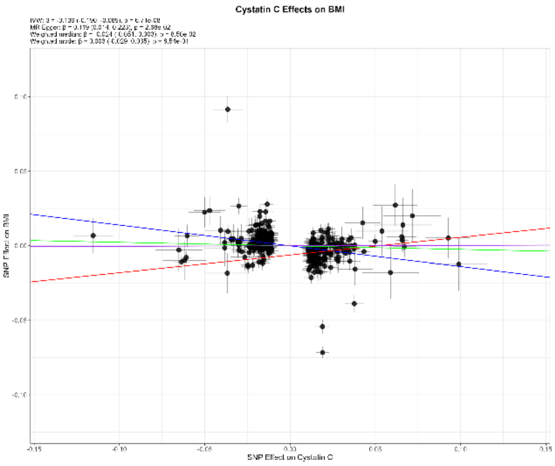

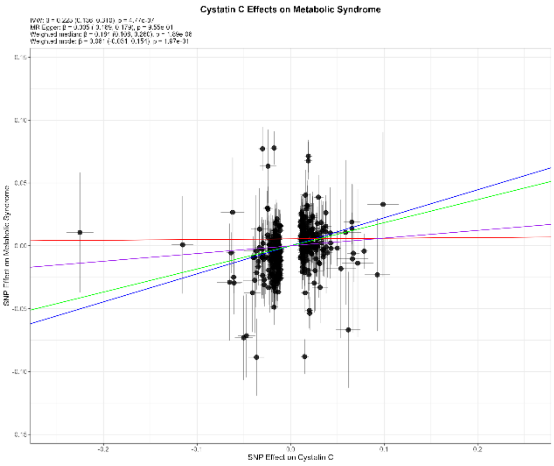

#### Impedance of Left Arm

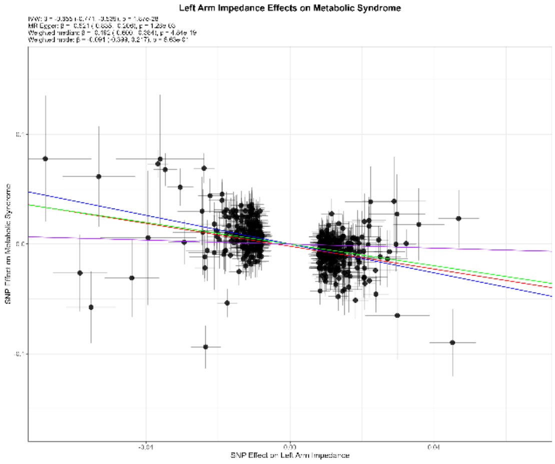

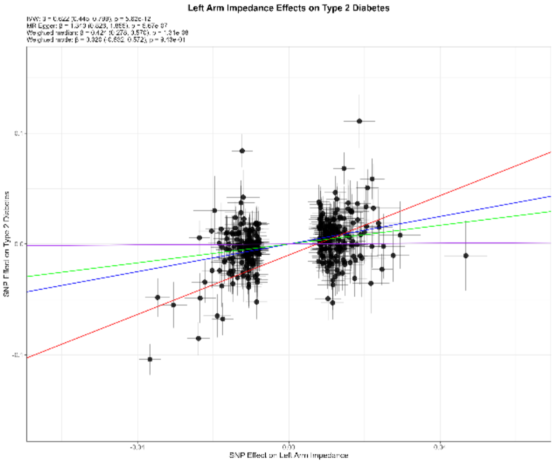

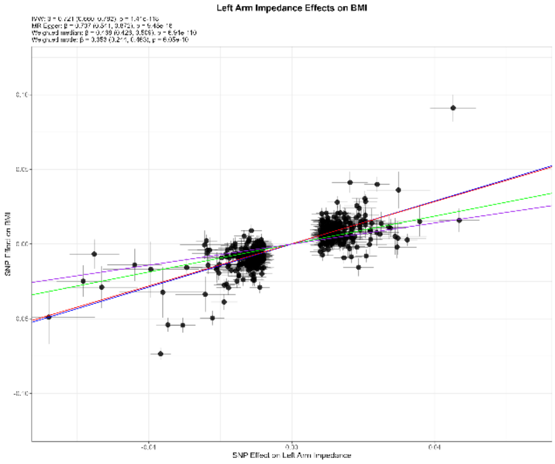

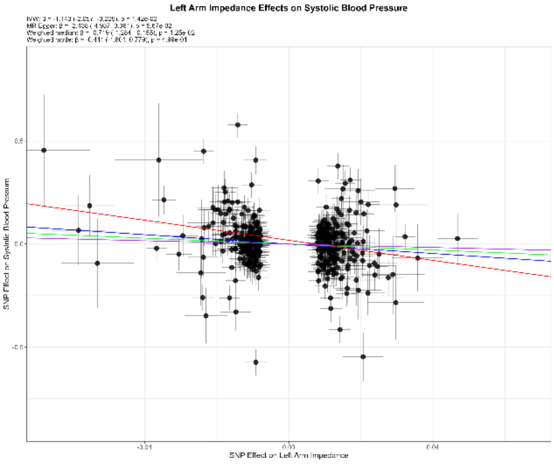

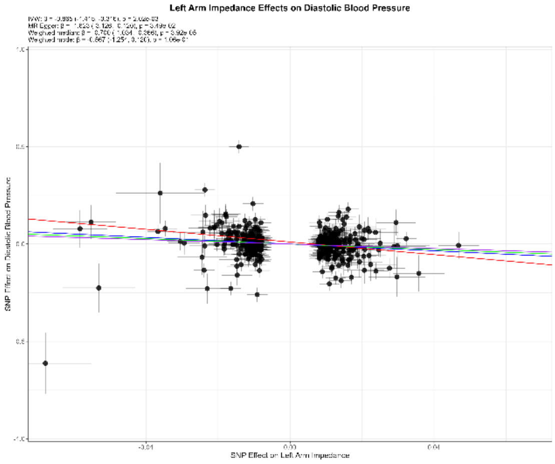

#### Impedance of Right Arm

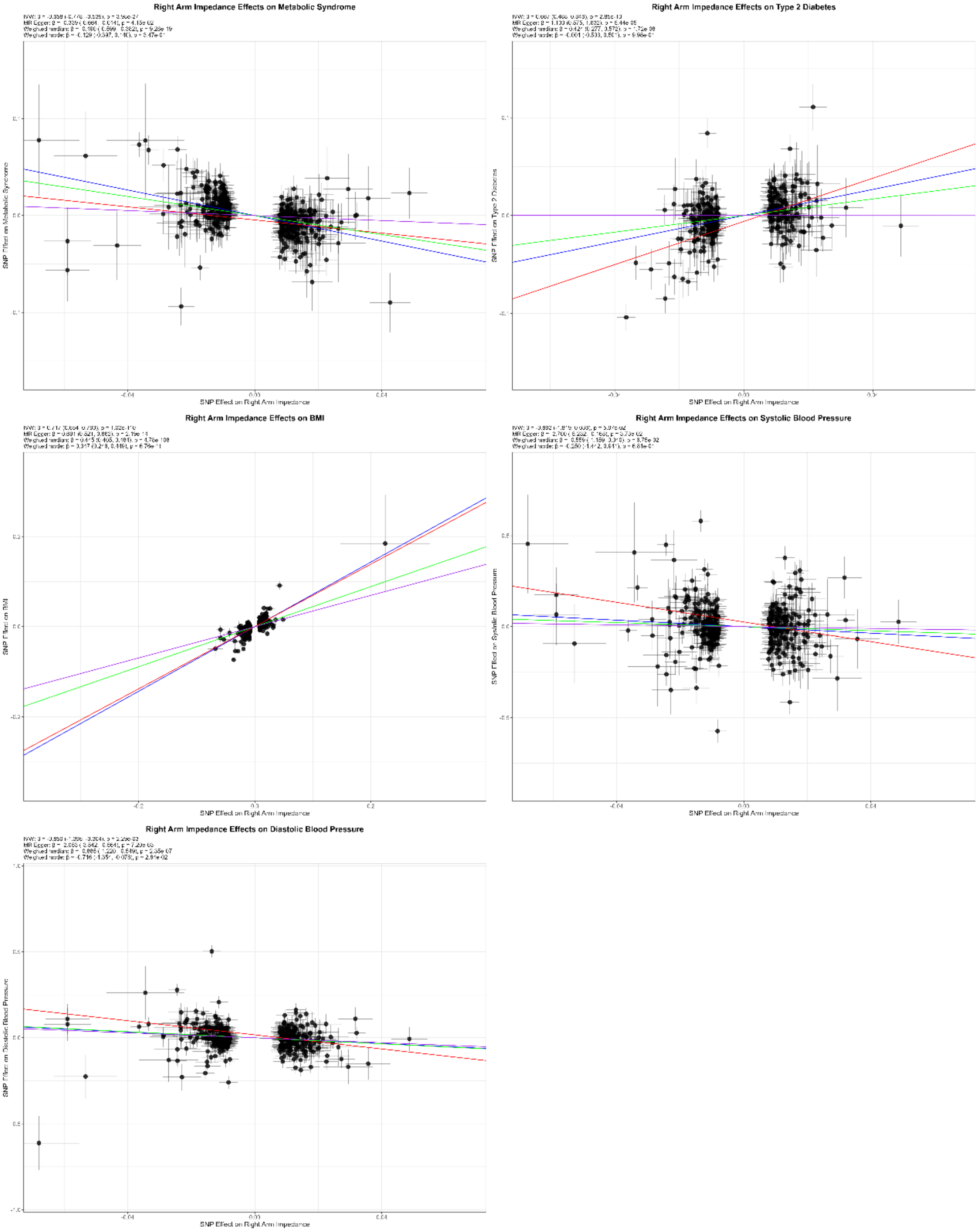

### Peak Expiratory Flow

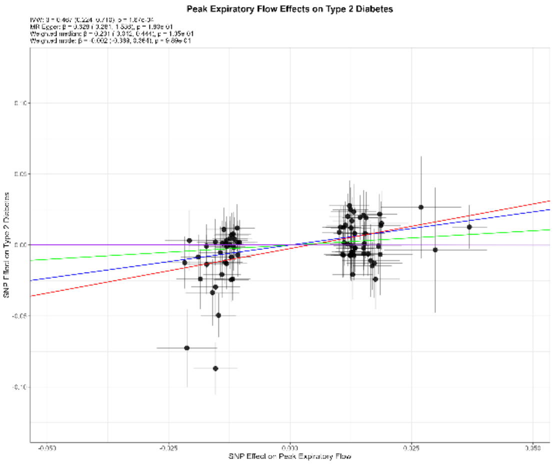

#### Phosphate

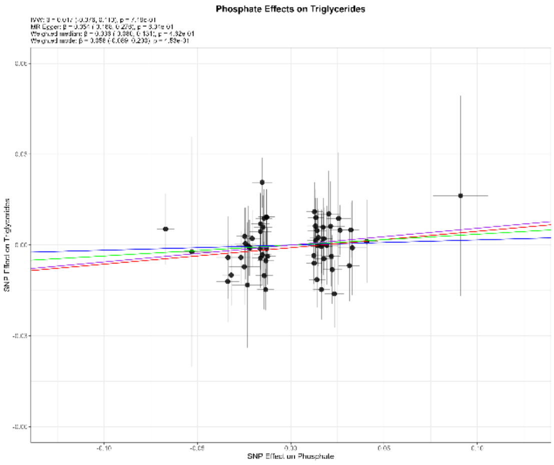

#### Urea

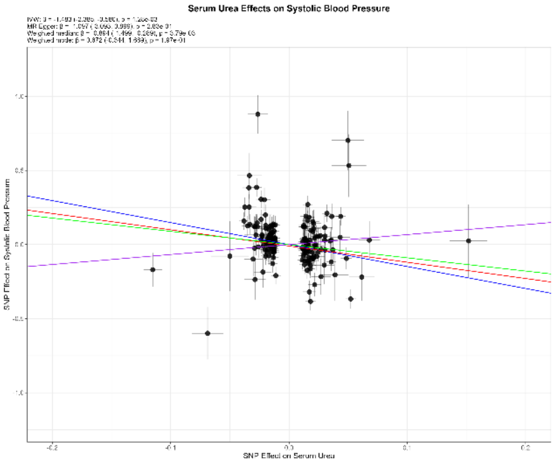

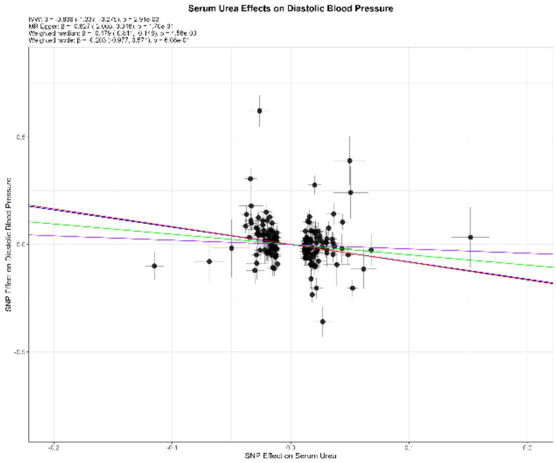

### Uric Acid

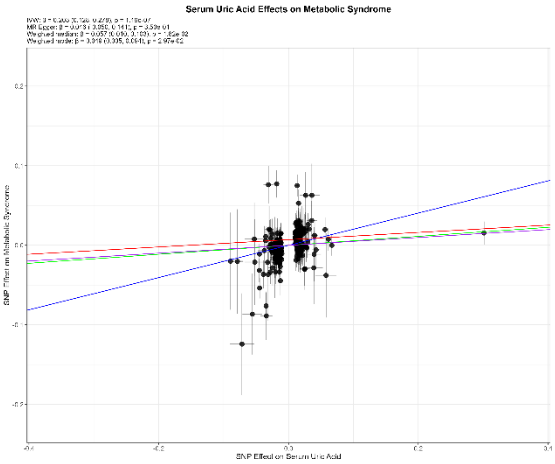

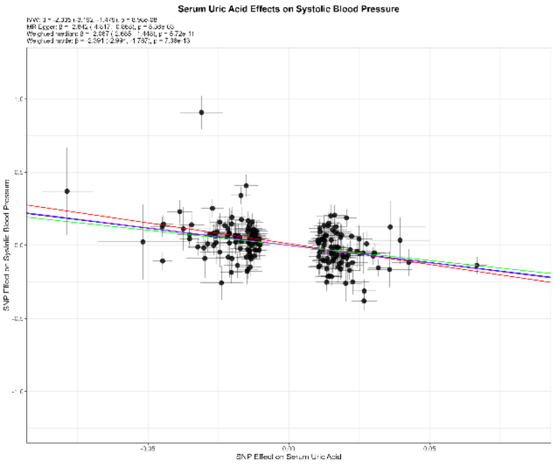

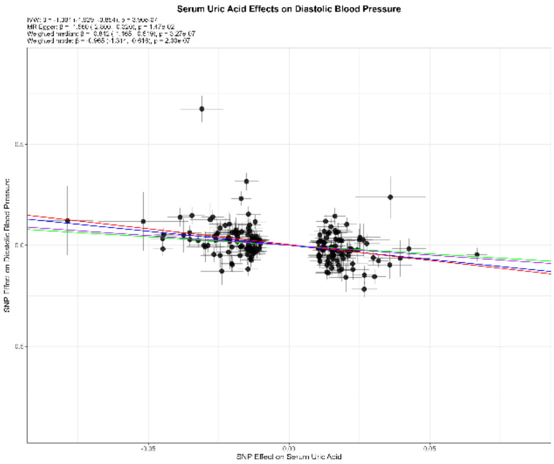

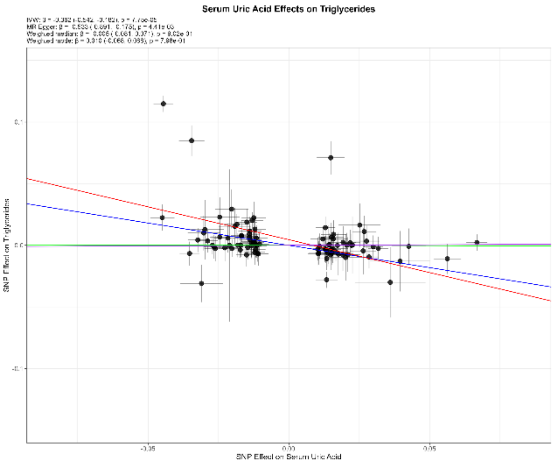

### Sodium in Urine

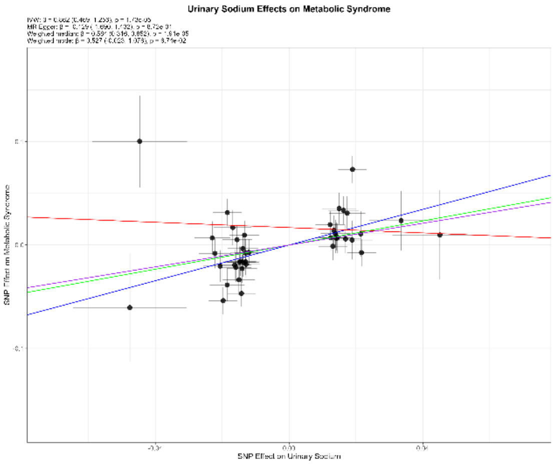

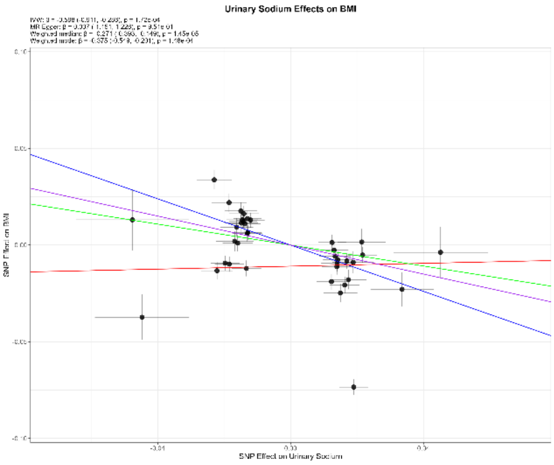

### Urate

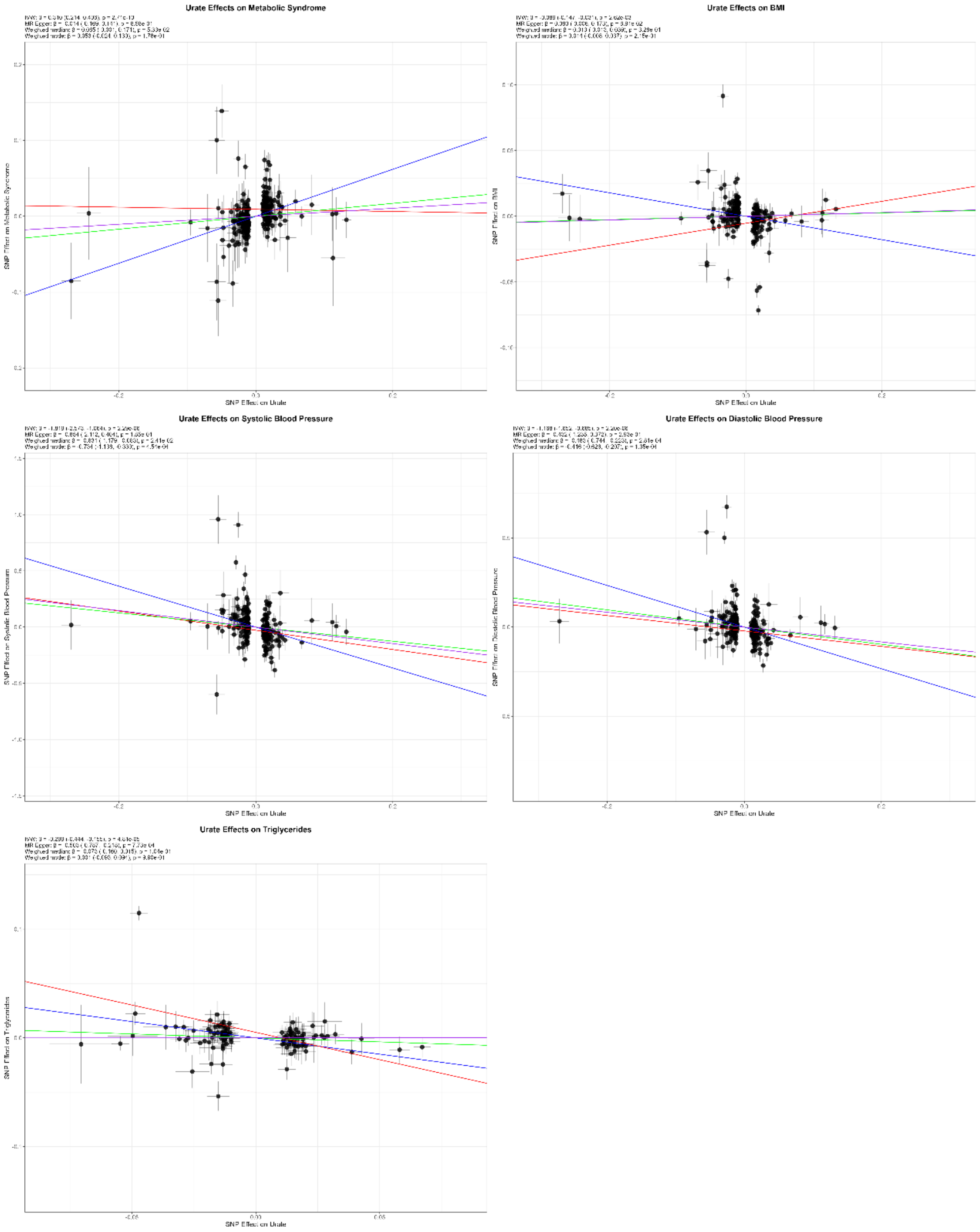
Urate Effects on Metabolic Syndrome
SNP Effect on Metabolic Syndrome
SNP Effect on Urate
Urate Effects on BMI
SNP Effect on BMI
SNP Effect on Urate
Urate Effects on Systolic Blood Pressure
SNP Effect on Systolic Blood Pressure
SNP Effect on Urate
Urate Effects on Diastolic Blood Pressure
SNP Effect on Diastolic Blood Pressure
SNP Effect on Urate
Urate Effects on Triglycerides
SNP Effect on Triglycerides
SNP Effect on Urate

### Urinary Albumin Excretion

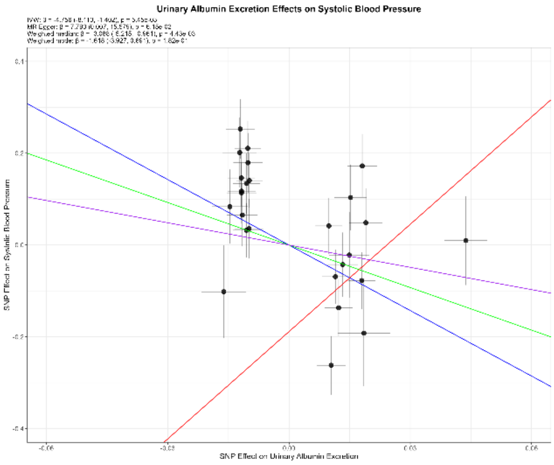

*Supplementary Figures 5. 3 – Leave-one-out Plots – Candidate Biomarkers Effect on Cardiometabolic Traits*

#### 25-Hydroxyvitamin D

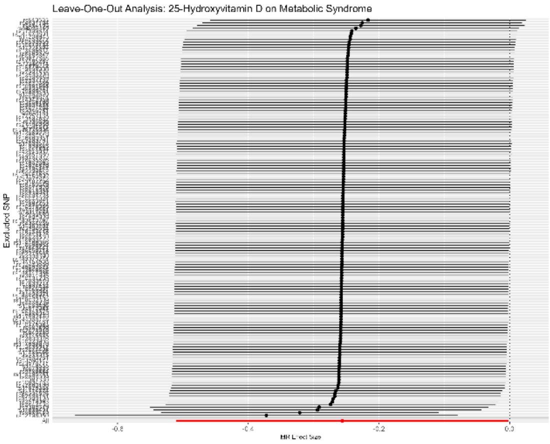

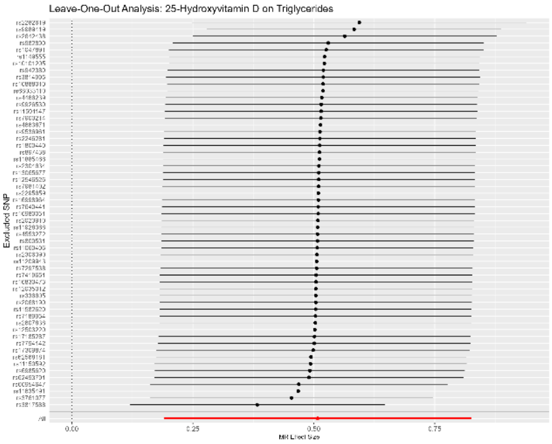

#### Gamma Glutamyl Transferase

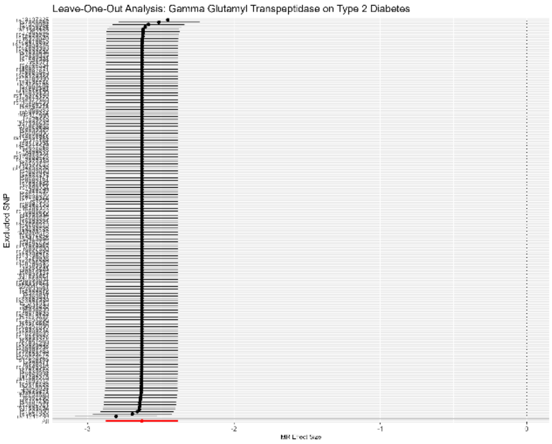

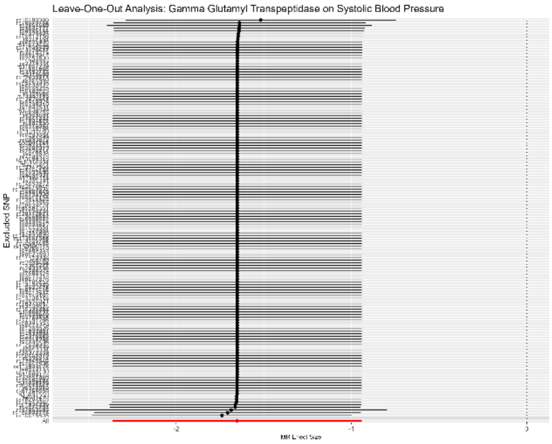

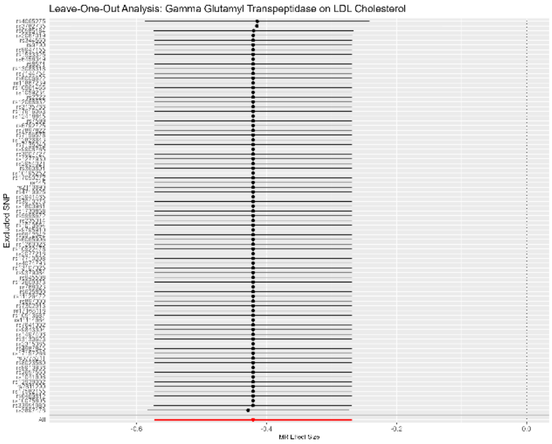

### Red Cell Distribution Width

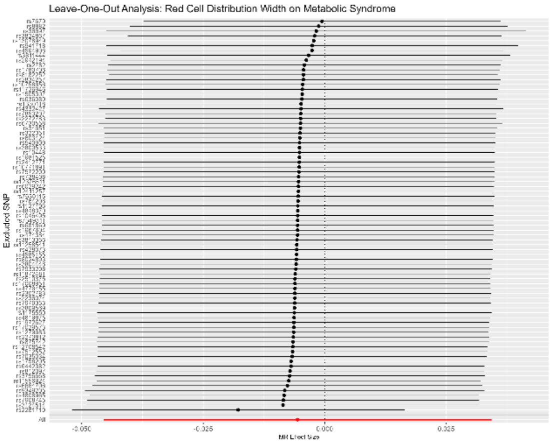

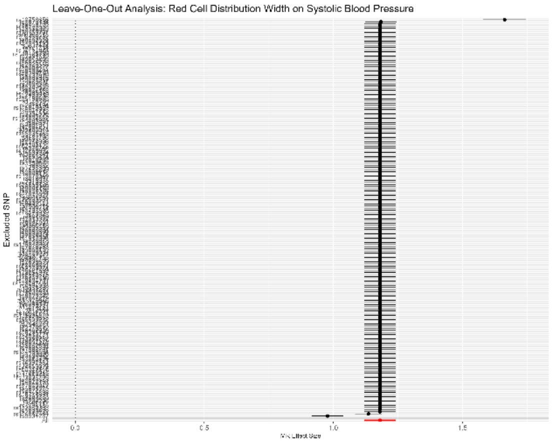

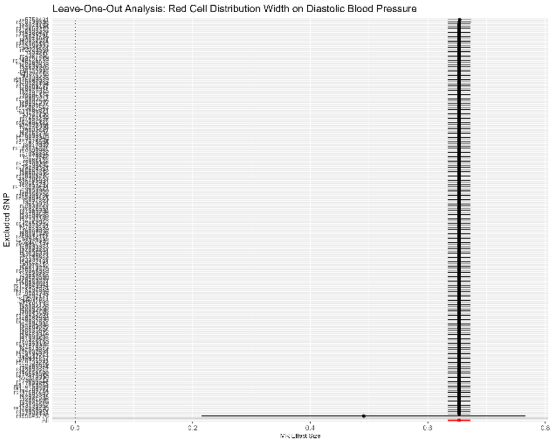

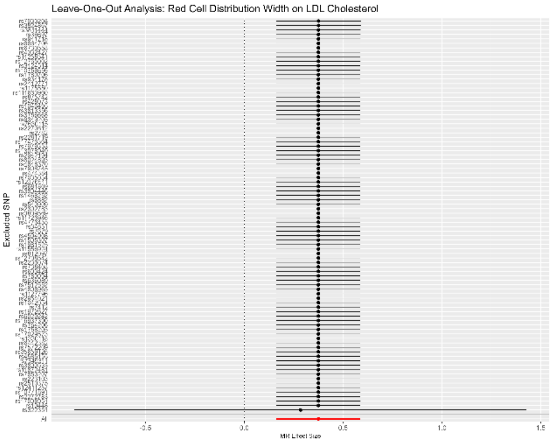

### Total Testosterone

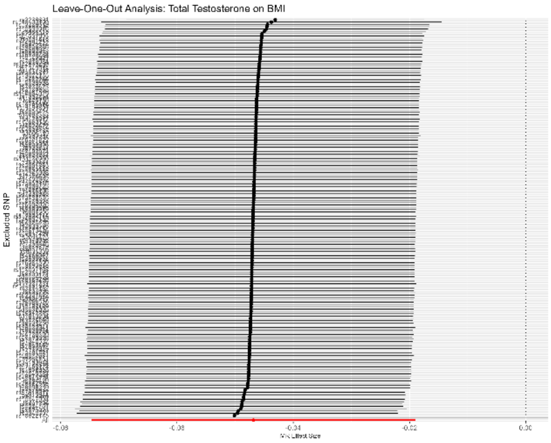

### Coffee Consumption

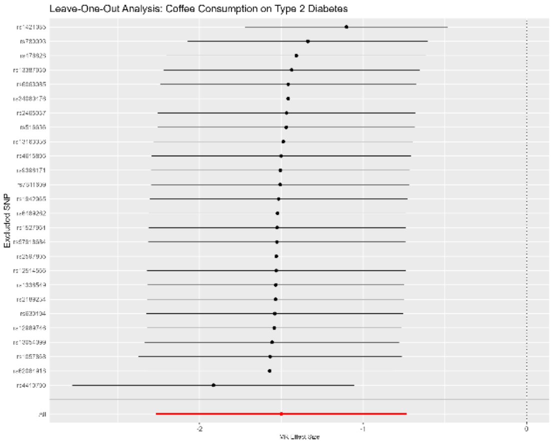

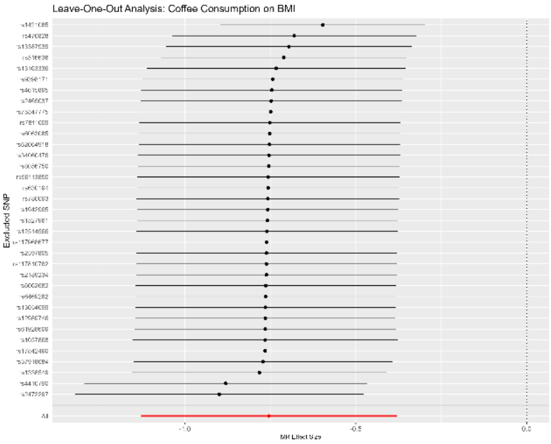

### Creatinine

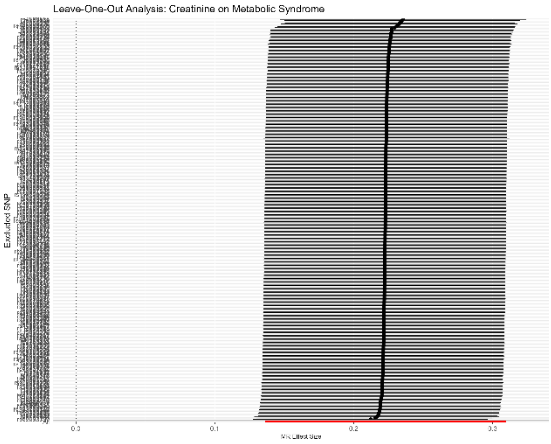

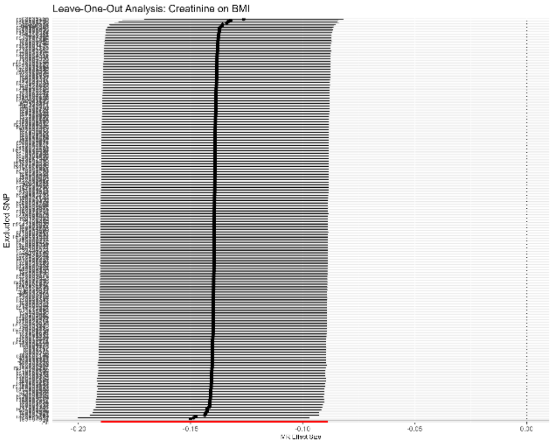

### Cystatin C

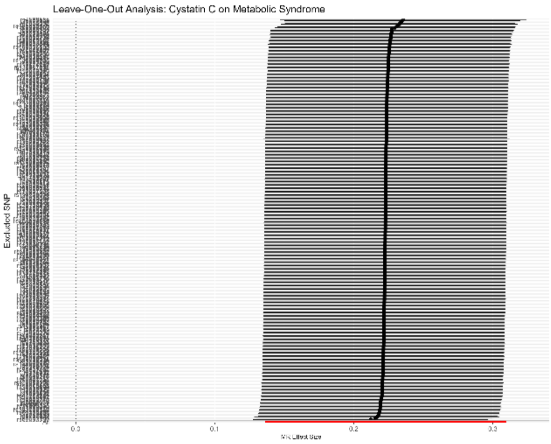

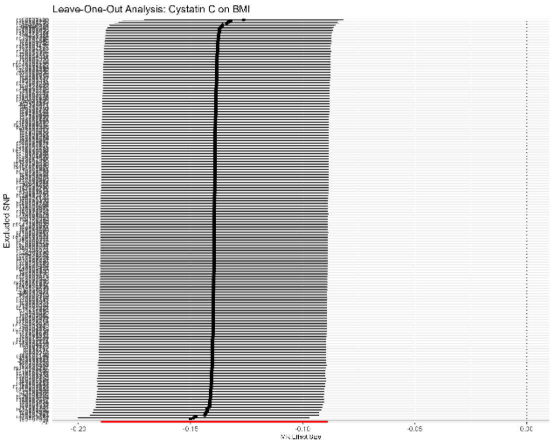

#### Impedance of Left Arm

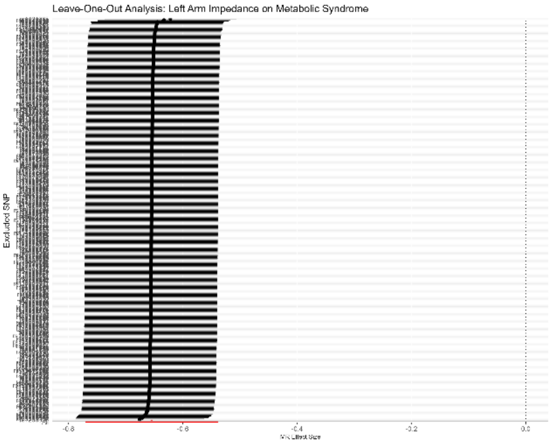

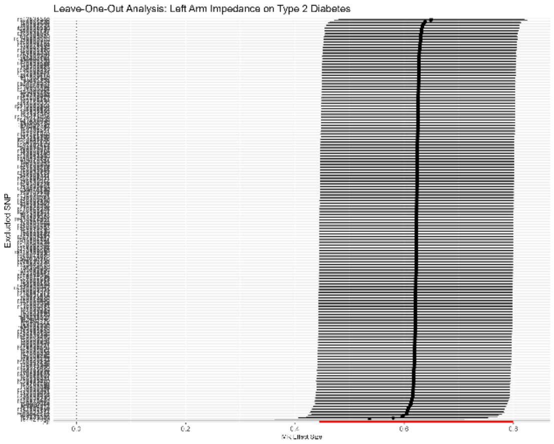

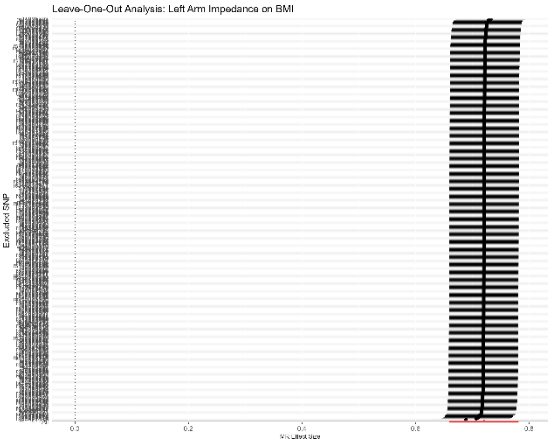

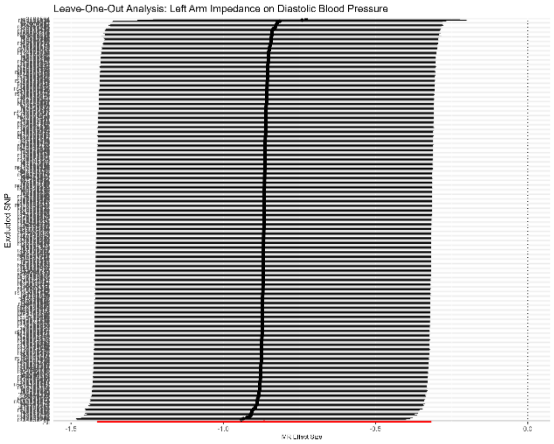

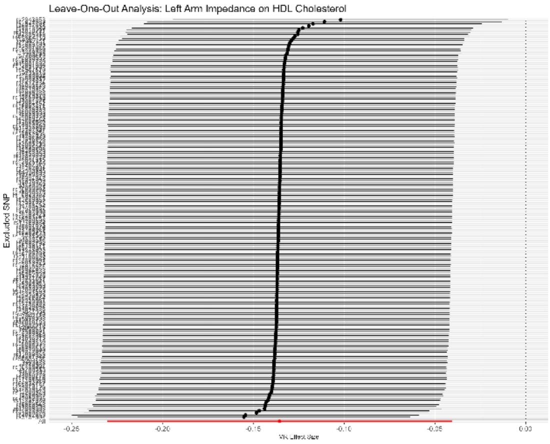

### Impedance of Right Arm

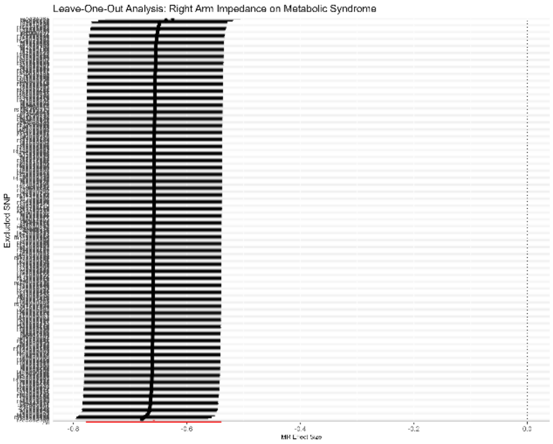

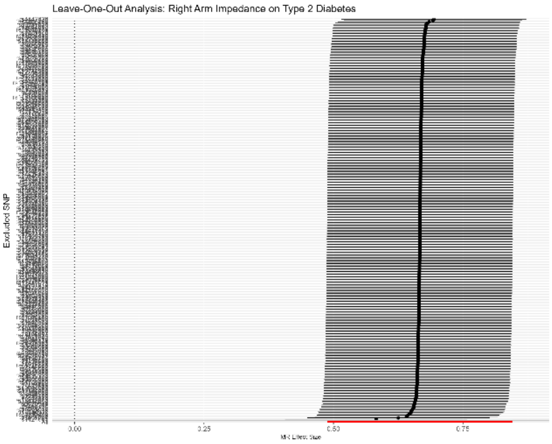

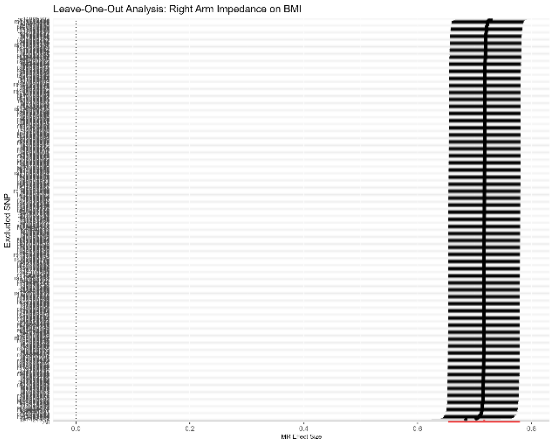

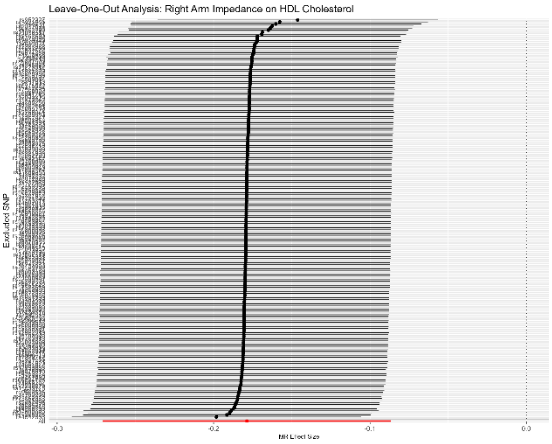

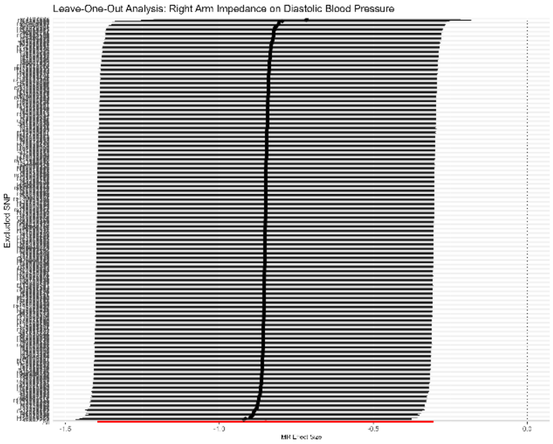

#### Peak Expiratory Flow

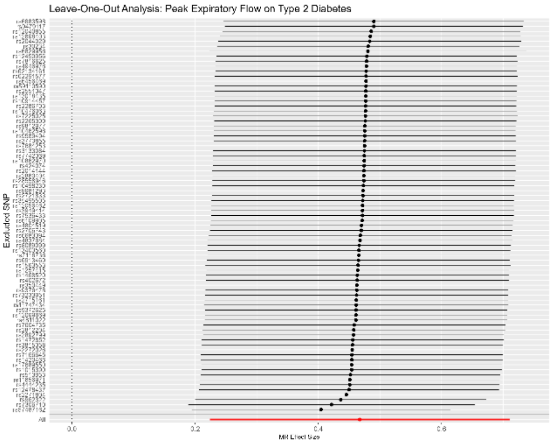

#### Phosphate

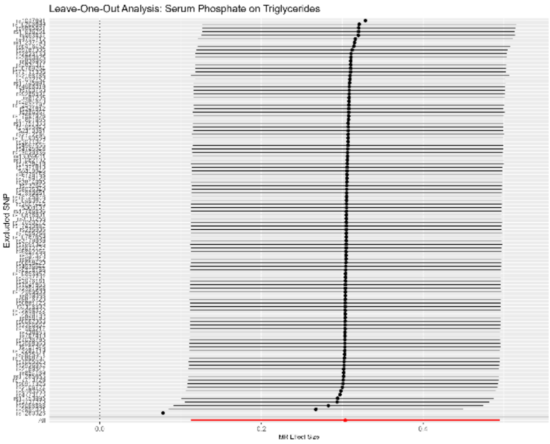

#### Urea

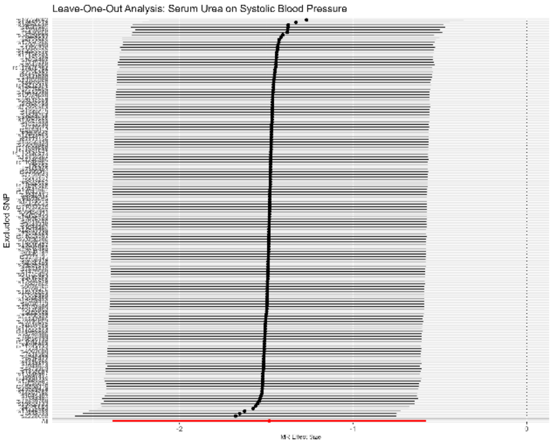

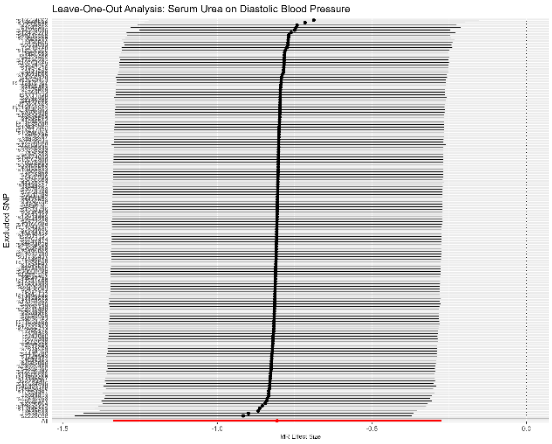

### Uric Acid

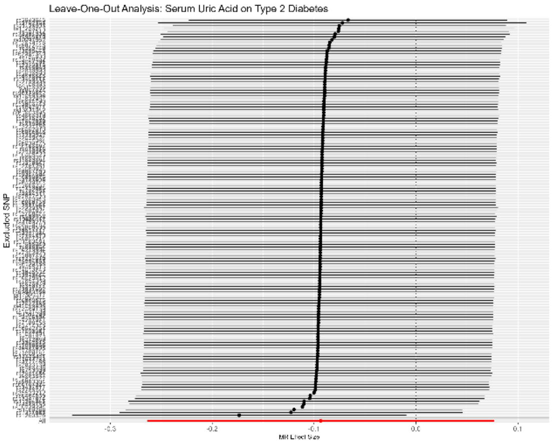

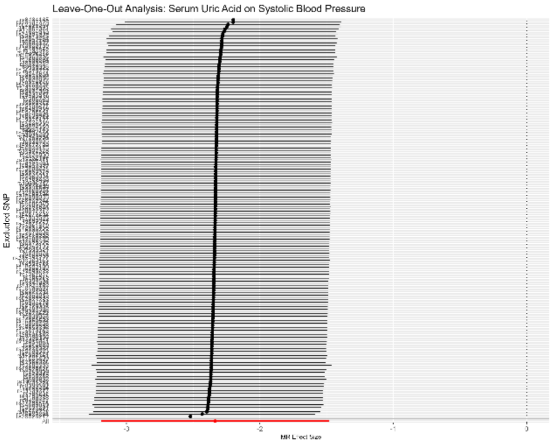

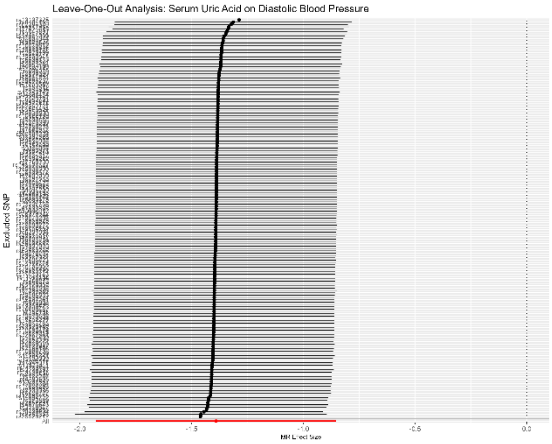

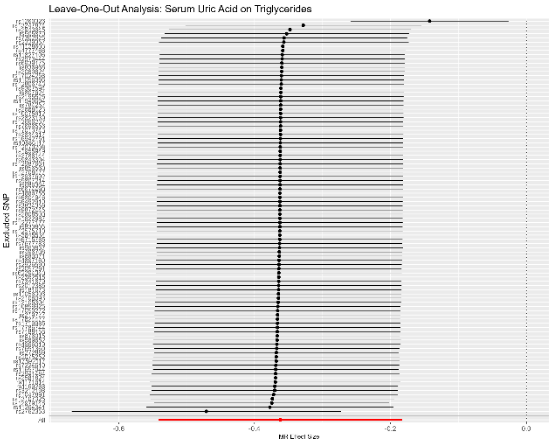

### Sodium in Urine

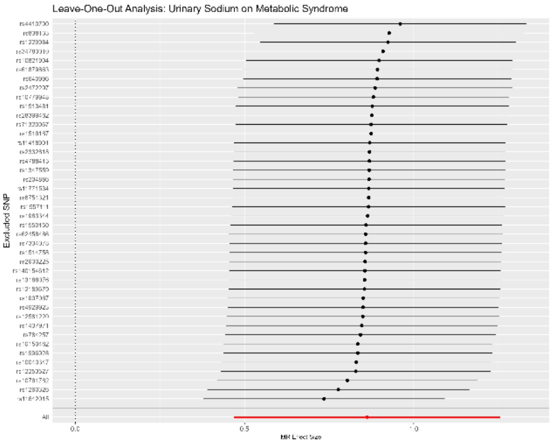

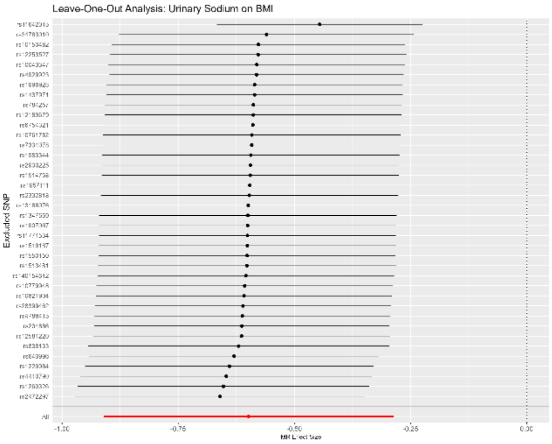

#### Urate

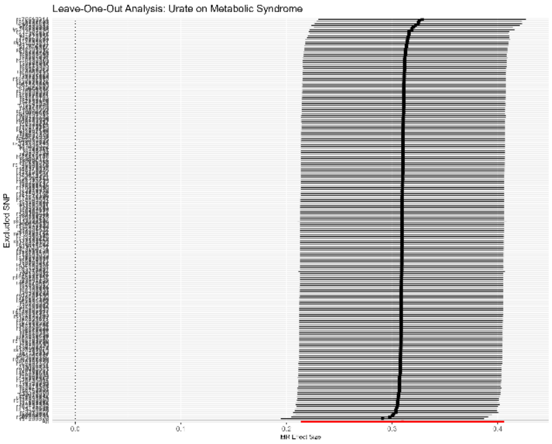
Leave-One-Out Analysis: Urate on Metabolic Syndrome
Excluded SNP
MR Effect Size

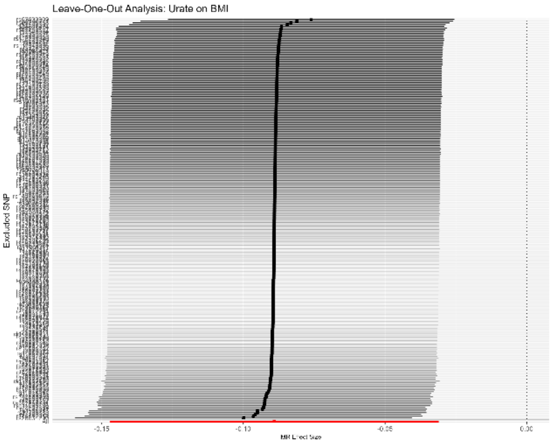
Leave-One-Out Analysis: Urate on BMI
Excluded SNP
MR Effect Size

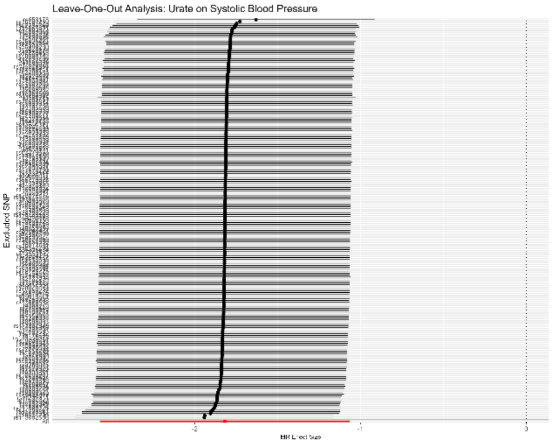
Leave-One-Out Analysis: Urate on Systolic Blood Pressure
Excluded SNP
MR Effect Size

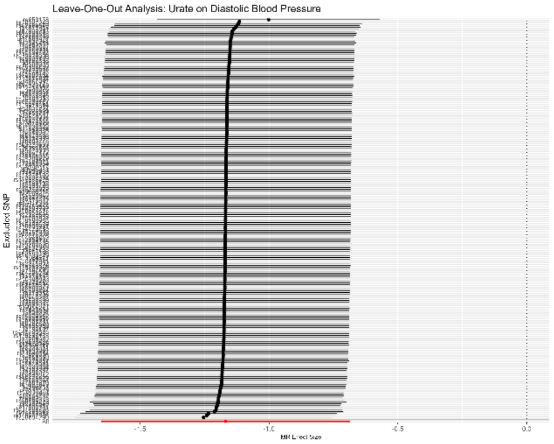
Leave-One-Out Analysis: Urate on Diastolic Blood Pressure
Excluded SNP
MR Effect Size

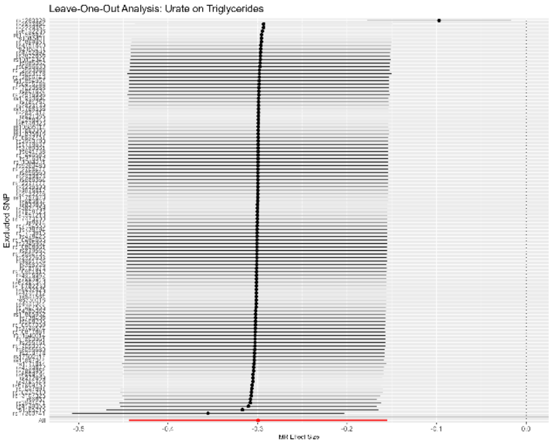
Leave-One-Out Analysis: Urate on Triglycerides
Excluded SNP
MR Effect Size

### Urinary Albumin Excretion

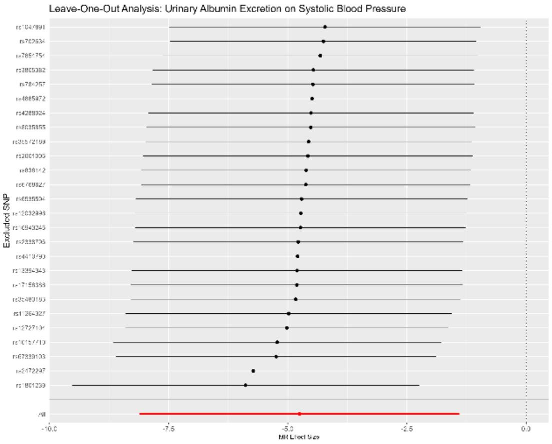

*Supplementary Figures 5. 4 – Forest Plots – Cardiometabolic Traits Effects on Candidate Biomarkers*

#### Metabolic Syndrome

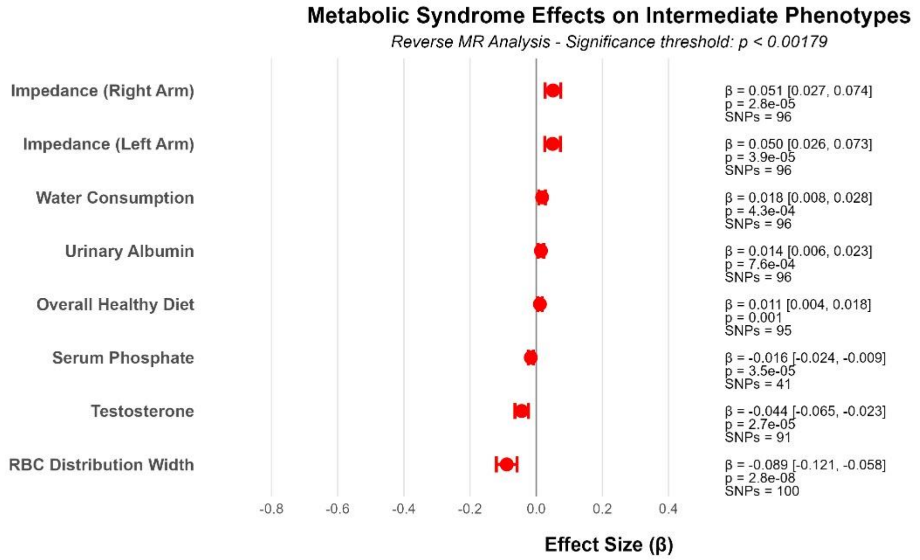

#### Type 2 Diabetes

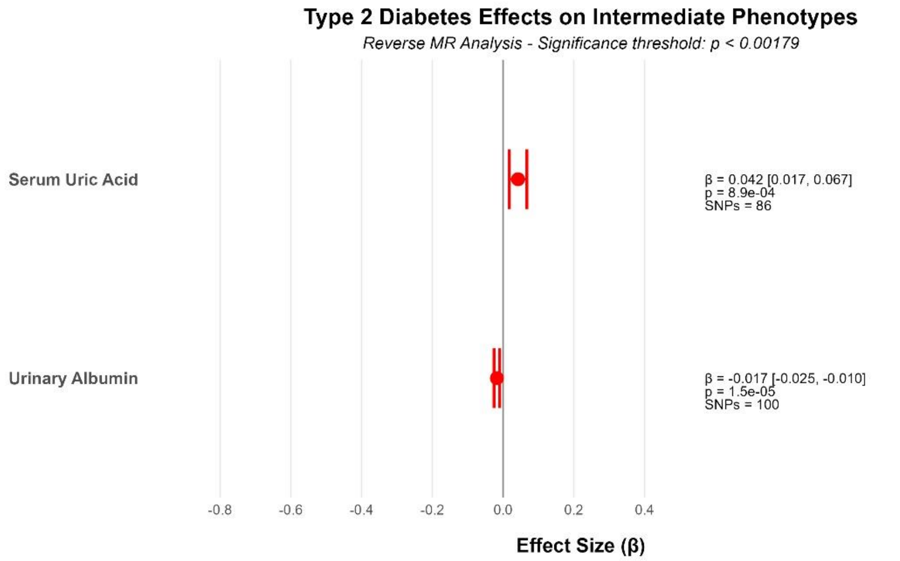

#### Body Mass Index

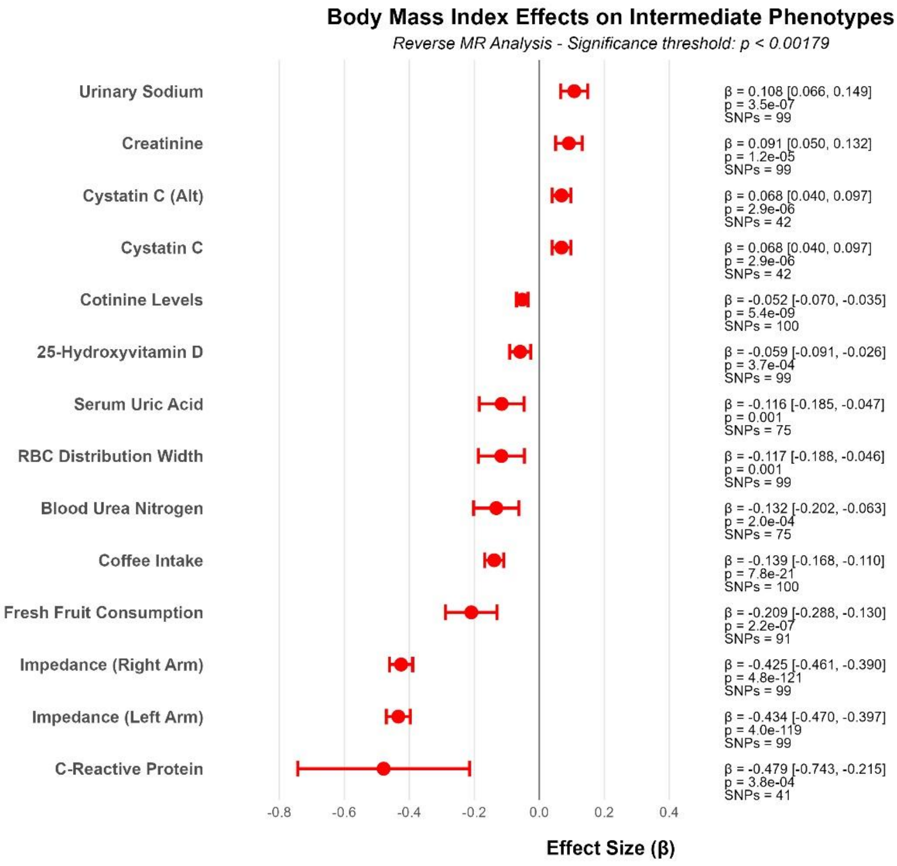

#### Systolic Blood Pressure

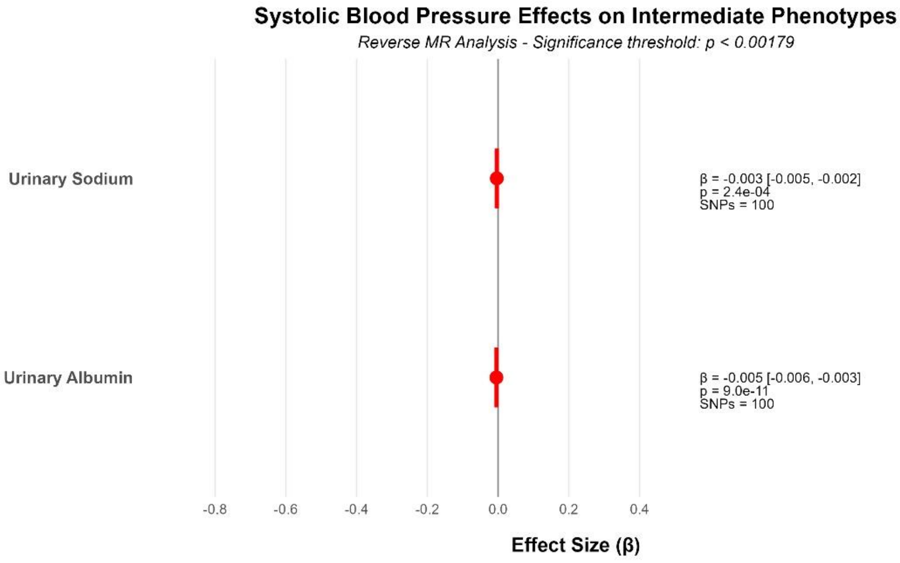

Diastolic Blood Pressure

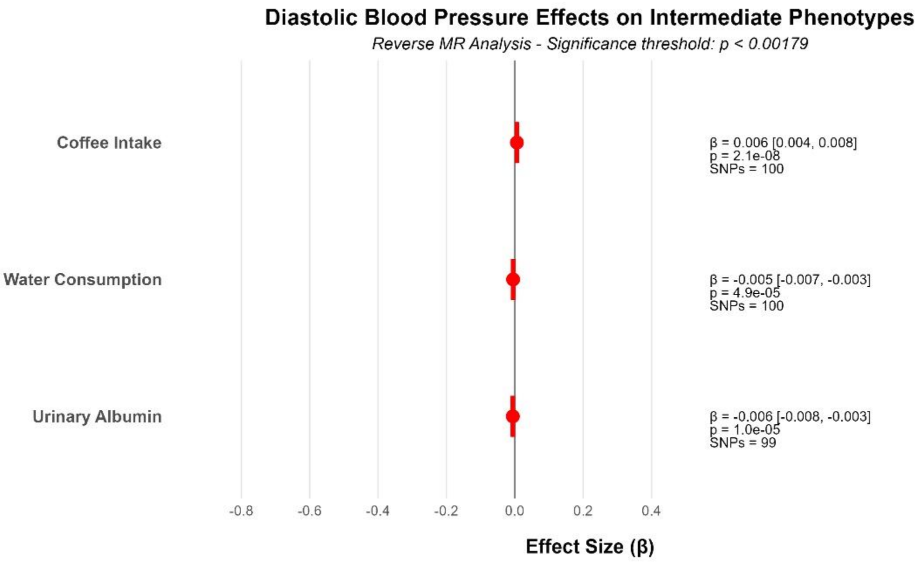

HDL

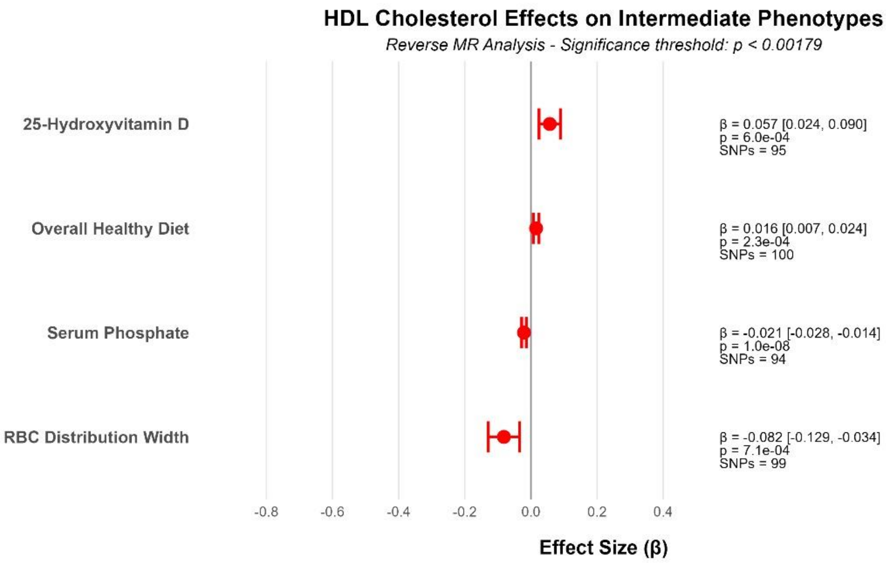

LDL

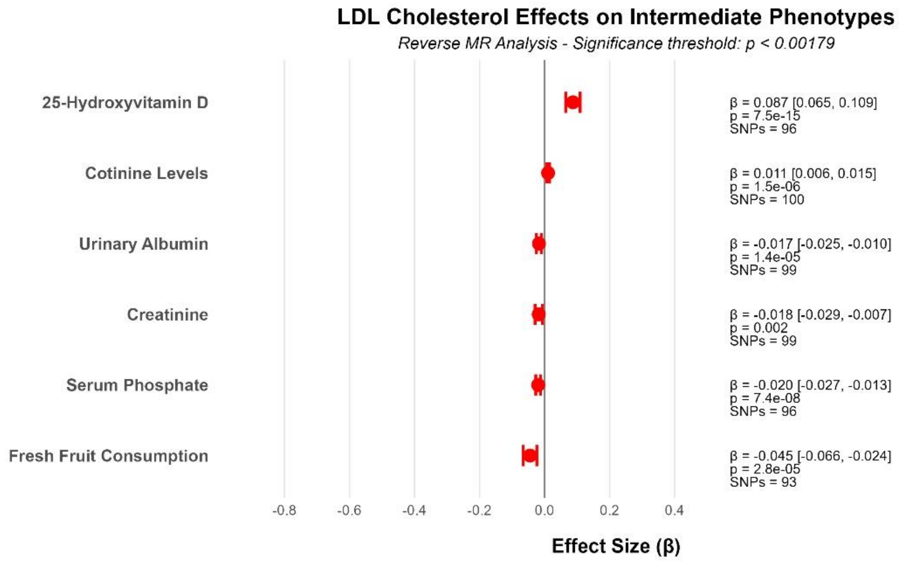

Triglycerides

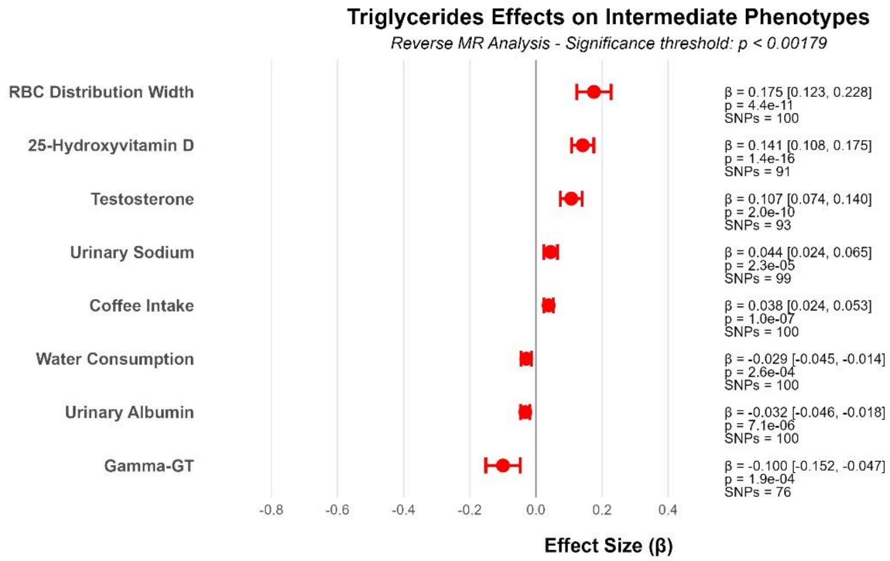

#### Supplementary Figures 6 – MVMR Analysis of 16 Candidate Biomarkers on Cardiometabolic Outcomes

*Forest plots showing multivariable Mendelian randomization results for 16 candidate biomarkers against cardiometabolic outcomes. Effect estimates (β) with 95% confidence intervals shown. Blue markers indicate $P < 0.05$ and grey markers indicate $P > 0.05$. Each biomarker shows independent causal effects despite genetic correlation.*

Supplementary Figures 6.1 – Forest Plots – MVMR of Candidate Biomarkers on Cardiometabolic Outcomes ........................................................................ 90

#### *Supplementary Figures 6. 1 – Forest Plots – MVMR of Candidate Biomarkers on Cardiometabolic Outcomes*

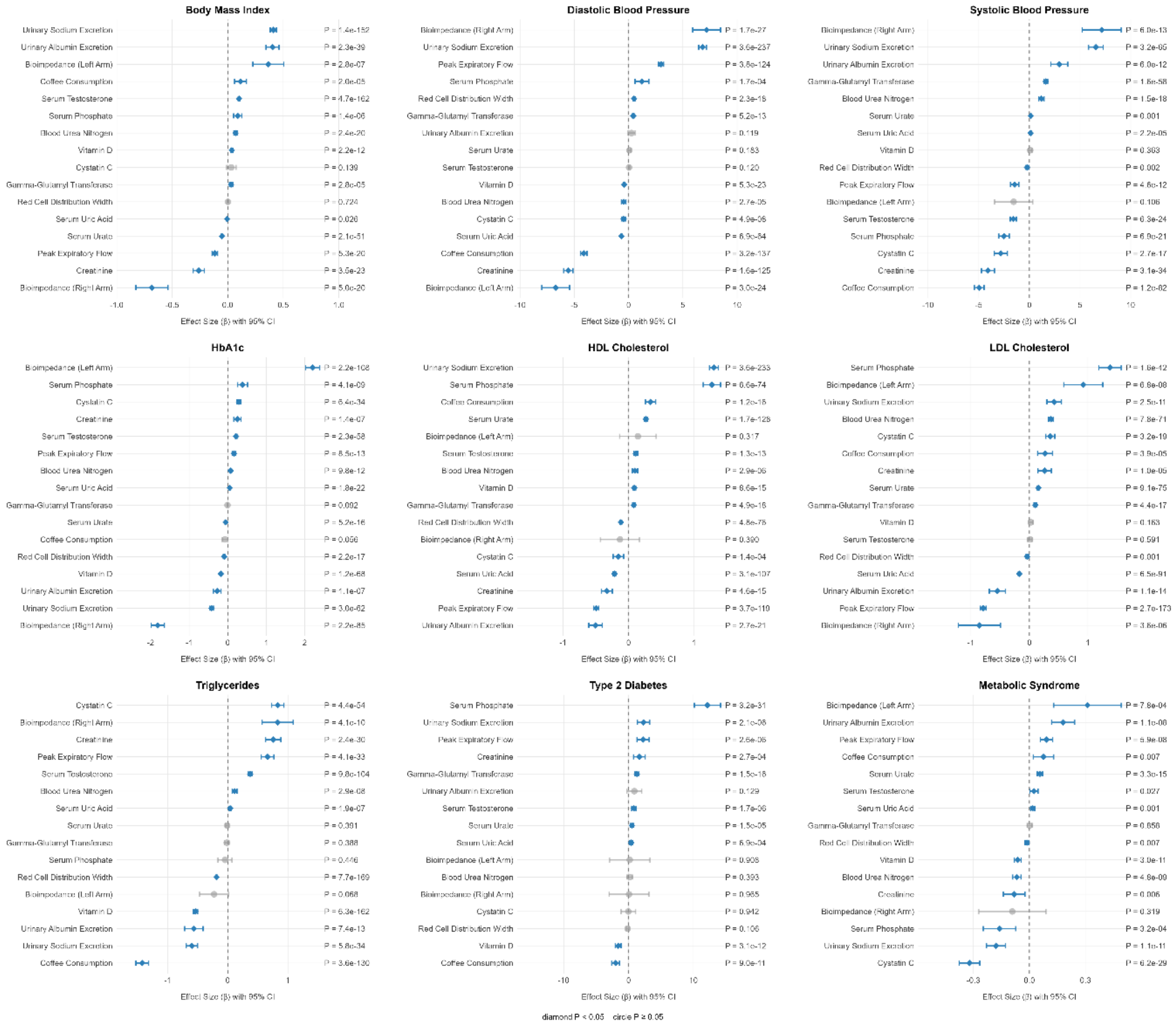
