## Supplementary Methods for "Genetic Epidemiological Pipeline Identifies Candidate Markers of Clozapine-Induced Metabolic Dysfunction Revealing Potential Avenues for Precision Clozapine Prescription"

### 1. Study Design and Analytical Framework

This study employed a multi-layered genomic approach, integrating Mendelian randomization (MR) with phenome-wide association studies (PheWAS) and colocalization techniques to investigate potential causal relationships between clozapine plasma levels and cardiometabolic outcomes. The analytical framework consisted of two main pathways of interconnected analysis; **(1)** Bidirectional Causal Investigation: Examining direct effects of clozapine plasma levels on metabolic outcomes; **(2.1)** Phenome-Wide Analysis: PheWAS approach to identify candidate biomarkers associated with clozapine-induced metabolic dysfunction based on a selection of lead variants; **(2.2)** Colocalization Analysis: Prune the list of candidate biomarkers using pairwise conditional colocalization (PWCoCo) to identify if they share causal variants with clozapine plasma levels; and **(2.3)** Causal Investigation of colocalized biomarkers: With the list of colocalized biomarkers, determine if these are causally related to cardiometabolic outcomes indicating potential biomarkers linked to metabolic risk as a result of clozapine use.

Figure 1 in the main text provides a visual representation of this pipeline, while the directed acyclic graphs (DAGs) in the supplementary figures depict the assumptions and relationships for each MR analysis component.

### 2. Data Sources and Instrument Selection

#### 2.1. Clozapine Plasma Traits Dataset

Genetic instruments for clozapine plasma levels were derived from summary statistics from the CLOZUK2 GWAS (Walters Group Data Repository, Centre for Neuropsychiatric Genetics and Genomics; n=4,495), which provided variant-level data for clozapine plasma concentration, N-desmethylclozapine (norclozapine) plasma concentration, and clozapine-norclozapine plasma ratio. Population from a UK study.

#### 2.2. Cardiometabolic Datasets

Cardiometabolic outcome data were obtained from large-scale European GWAS through the OpenGWAS platform (https://www.ebi.ac.uk/gwas/) and GWAS Catalog (https://gwas.mrcieu.ac.uk/) , including:

- Metabolic syndrome (GCST009602; n=291,107)
- Type 2 diabetes (ieu-a-1090; n=120,286)
- Body mass index (ukb-b-19953; n=461,460)
- Systolic blood pressure (ieu-b-38; n=757,601)
- Diastolic blood pressure (ieu-b-39; n=757,601)
- HDL cholesterol (ieu-a-299; n=187,167)
- LDL cholesterol (ieu-a-300; n=173,082)
- HbA1c (ieu-b-103; n=46,368)
- Triglycerides (ieu-b-111; n=441,016)

#### 2.3. Genetic Instrument Selection

For MR analyses using clozapine plasma instruments, clumping was done using the following parameters to ensure independence; clumping $r^2$ threshold: 0.001, clumping window: 10,000 kb, and significance threshold: $P < 1.00\times10^{-5}$. Standard quality control procedures were applied to the genetic data using PLINK v1.9. This included filtering variants based on minor allele frequency (MAF > 0.01), genotype missingness (< 5%), imputation quality scores (INFO > 0.8), and Hardy-Weinberg equilibrium ($P > 1.00\times10^{-6}$). These QC steps ensured that only high-quality genetic variants ($F \approx 20$) were used as instruments in the MR analyses (Table 2 main text).

A systematic approach was implemented to select genetic instruments for clozapine plasma traits to be used in the PheWAS-colocalization analysis: **(1)** Initial SNP (single nucleotide polymorphism or genetic variant) collection: 92 SNPs for clozapine metabolism were collected from Open Targets (https://genetics.opentargets.org/) and GWAS Catalog (https://www.ebi.ac.uk/gwas/); **(2)** Significance thresholding: Applied a P-value threshold of $5.00\times10^{-8}$, reducing the list to 28 SNPs; **(3)** Duplicate removal: After removing duplicates, 16 SNPs remained; **(4)** Linkage disequilibrium (LD) assessment: LD relationships between SNPs on the same chromosome were assessed using LDmatrix by the National Institutes of Health (NIH) (https://ldlink.nih.gov/?tab=ldmatrix); and **(5)** Final selection: After careful consideration of LD patterns, particularly for the complex locus on chromosome 4 with multiple partially independent signals and implicated gene clusters previously identified with clozapine plasma traits, 10 lead SNPs were retained as genetic instruments (Table 3 main text).

### 3. Primary Causality Analysis

#### 3.1. Direct MR Analysis

For the primary analysis examining direct effects of clozapine plasma traits on metabolic outcomes, we performed LD-aware inverse variance weighted (IVW) Mendelian randomization using the selected genetic instruments.

The analysis was conducted using the TwoSampleMR package (v0.5.6) in R (v4.3.3), with custom modifications to incorporate LD information. For each clozapine plasma trait (clozapine plasma, norclozapine plasma, and clozapine-norclozapine plasma ratio) against each metabolic outcome, we: **(1)** Harmonized exposure and outcome data to ensure consistent effect allele alignment; **(2)** Incorporated LD matrices derived from the 1000 Genomes European reference panel; **(3)** Computed LD-aware IVW estimates using the following formula:

$$\hat{\beta}_{IVW} = (X'\Omega^{-1}X)^{-1}X'\Omega^{-1}Y$$

Where, $X$ is the matrix of SNP-exposure effects ($\hat{\beta}_X$) with dimensions $n \times 1$ ($n$ SNPs), $Y$ is the vector of SNP-outcome effects ($\hat{\beta}_y$) with dimensions $n \times 1$, $\Omega$ is the LD-adjusted variance-covariance matrix with dimensions $n \times n$, where $\Omega = \sum Y \times R$ ($\sum Y$ is a diagonal matrix of outcome variances ($SE_Y^2$) and $R$ is the genetic correlation matrix); and (4) Calculated F-statistics to assess instrument strength using:

$$F_j = (\frac{\beta_{X_j}}{SE_{X_j}})^2$$

$$\bar{F} = (\frac{1}{n})\sum_{j=1}^{n} F_j$$

To assess the robustness of our findings, we conducted several sensitivity analyses. MR-Egger regression to evaluate directional pleiotropy. This method allows for the estimation of a non-zero intercept that can indicate the average pleiotropic effect:

$$\beta_Y = \beta_0 + \beta_{Egger} \times \beta_X + \varepsilon$$

Where $\beta_0$ is the intercept term, representing the average pleiotropic effect. $\beta_{Egger}$ is the slope coefficient, representing the causal effect adjusted for directional pleiotropy. The intercept term ($\beta_0$) provides a test for directional pleiotropy. Weighted median estimation to provide a robust causal estimate when some genetic variants used as instruments might be invalid.

$$\hat{\beta}_{WMed} = Median_{w_j}(\beta_j)$$

Where $\beta_j = \frac{(\beta_{Y_j})}{(\beta_{X_j})}$ which is the Wald ratio estimate for the $j^{th}$ SNP. $w_j \propto \frac{1}{Var(\frac{\beta_{Y_j}}{\beta_{X_j}})}$ are weights which are inversely proportional to the variance of the Wald ratio. This approach provides consistent estimates when up to 50% of the information comes from invalid instruments. Heterogeneity assessment using Q-statistics and $I^2$ values were calculated by:

$$Q = \sum_{j=1}^{n} w_j (\frac{(\beta_{Y_j})}{(\beta_{X_j})} - \hat{\beta}_{IVW})^2$$

$$I^2 = \max\,[0{,}100\% \times \frac{Q - df}{Q}]$$

where Q is Cochran's Q statistic and df is the degrees of freedom (1, 2). Leave-one-out analysis to identify influential outliers that might drive the overall effect was also run generating plots available in the supplementary figures for each individual analysis (3).

#### 3.2. Reverse Causation Analysis

To investigate potential bidirectional relationships, we conducted reverse MR analyses using metabolic traits as exposures and clozapine plasma phenotypes as outcomes. Metabolic trait instruments extracted using Plink1.9 at a genome-wide significance p-value threshold. The same analytical approach and sensitivity analyses were applied, with metabolic traits as the exposure and clozapine metabolism phenotypes as the outcomes.

### 4. Phenome-Wide Association Analysis

To identify potential biomarkers linked to the clozapine-metabolic relationship, we conducted a phenome-wide association study (PheWAS) using the R integrated API for OpenGWAS. For each of the 10 lead SNPs identified for clozapine plasma traits (Table 3 main text), we: **(1)** Queried the OpenGWAS database; and **(2)** Prioritized biomarkers based on significance, consistency across SNPs, and biological relevance to metabolic outcomes (4, 5).

### 5. Colocalization Analysis

To evaluate whether the genetic signals for clozapine plasma levels and identified biomarkers via the PheWAS analysis share causal variants, we performed colocalization analysis using PWCoCo (Pairwise Colocalization with Conditioning). PWCoCo implements colocalization analyses from coloc.abf (colocalization R package) for colocalization evidence. A limitation of the coloc.abf method is where the loci being assessed for colocalization evidence must contain only one causal variant (single variant assumption). PWCoCo allows for this assumption to hold as the conditional analyses will identify and then systematically condition upon each independent signal within a region. These regions are then assessed for colocalization evidence using each pairwise combination of conditionally independent signal (6). This was done using the following pipeline: **(1)** Data preparation: Summary statistics were extracted for each lead SNP and its associated traits within a ±500kb window. Allele frequencies were calculated using the appropriate chromosomal reference datasets from 1000 Genomes; **(2)** Reference dataset preparation: Chromosome-specific reference files were merged using PLINK v1.9; and **(3)** Colocalization implementation: PWCoCo analysis was performed using the merged reference dataset, applying a Bayesian framework to calculate posterior probabilities for five hypotheses in a high-performance computing suite where the following hypotheses were implemented:

$H_0$: No association with either trait
$H_1$: Association with trait 1 only
$H_2$: Association with trait 2 only
$H_3$: Association with both traits, different causal variants
$H_4$: Association with both traits, shared causal variant

Estimates were calculated by PWCoCo using the following model: Posterior probability of hypothesis $H_4$

$$PP(H_4) = \frac{P(D|H_4) \times prior_4}{\sum_{j=0}^{4} P(D|H_i) \times prior_i}$$

Where $P(D|H_i)$ is the likelihood of the data under hypothesis $i$, $prior_i$ is the prior probability assigned to hypothesis $i$. Default prior probabilities were used: $p_1 = p_2 = 1 \times 10^{-4}$ for single-trait association and $p_{12} = 1 \times 10^{-5}$ for shared causal variants, consistent with PWCoCo recommendations. The bayes factor calculation for comparing $H_4$ to $H_3$ is:

$$BF_{4,3} = \frac{(P(D|H_4))}{(P(D|H_3))}$$

Where a high bayes factor supports the hypothesis of a shared causal variant; **(4)** Conditional analysis: PWCoCo was further used to evaluate conditional relationships, allowing us to identify independent biomarkers by adjusting for other colocalized traits:

$$PP(H_4|Z) = \frac{P(D|H_4, Z) \times prior_4}{\sum_{j=0}^{4} P(D|H_i, Z) \times prior_i}$$

Where $Z$ represent the conditioning variable (another trait) and $P(D|H_i, Z)$ is the likelihood of the data under hypothesis $i$ after conditioning on $Z$. The likelihood calculation under conditioning is:

$$P(D|H_4, Z) = \int P(D|\beta, Z) \times P(\beta|H_4) d\beta$$

Where $\beta$ represents the effect sizes of variants, $P(D|\beta, Z)$ is the likelihood of the data given effect sizes $\beta$ after conditioning on $Z$, and $P(\beta|H_4)$ is the prior distribution of the effect sizes under $H_4$; and **(5)** Threshold application: We applied a $H_4 > 0.8$ threshold for robust colocalization, indicating strong evidence for a shared causal variant.

To visualise the colocalization results, we generated stacked regional association plots for each exposure-outcome relationship. These plots display the regional association patterns for clozapine plasma traits, association patterns for colocalized biomarkers, LD structure, and position of genes and variants within the region. Stacked association plots were generated using geni.plots (v0.1.2) in R and provide a visualisation of the colocalization patterns seen by this analysis. These are available in the supplementary figures.

#### 6. Causality Assessment of Candidate Biomarkers on Cardiometabolic Outcomes

For biomarkers identified through the PheWAS-colocalization pipeline ($H_4$ > 0.8), we conducted MR analyses to estimate their causal effects on metabolic outcomes: **(1)** Instrument Selection: For each unconditional phenotype, genetic instruments were selected using the R integrated OpenGWAS API, using the study with the highest sample size where the data was of European ancestry at genome-wide significance; **(2)** Data harmonization and quality control: Instruments were harmonized with outcome data and screened for duplicate variants and missing values; **(3)** MR estimation: IVW analysis (non-LD-aware) was performed using the TwoSampleMR package:

$$\hat{\beta}_{IVW} = \frac{\sum_{j=1}^{n} \beta_{Y_j} \times \beta_{X_j} \times w_j}{\sum_{j=1}^{n} \beta_{X_j}^2 \times w_j}$$

Where $\beta_Y, j$ is the effect of SNP $j$ on the outcome, $\beta_X, j$ is the effect of SNP $j$ on the exposure, and $w_j \propto \frac{1}{Var(\frac{\beta_{Y},j}{\beta_{X},j})}$ are the weights (inverse variance of the Wald ratio). Sensitivity analyses (MR-Egger, weighted median) were conducted as described in section 3.1. Weighted mode was also conducted as an additional sensitivity analysis:

$$\hat{\beta}_{WMode} = argmax_{\beta}[\sum_{j=1}^{n} K_h(\beta - \beta_j) \times w_j]$$

Where $\beta_j = \frac{(\beta_{Y_j})}{(\beta_{X_j})}$ which is the Wald ratio estimate for the $j^{th}$ SNP, $K_h$ is the kernel function with bandwidth $h$, $w_j \propto \frac{1}{Var(\frac{\beta_{Y_j}}{\beta_{X_j}})}$ are the weights inversely proportional to the squared standard error of the Wald ratio. This estimator is robust to horizontal pleiotropy when up to 50% of the genetic variants are invalid instruments, as it identifies the most common (modal) causal effect estimate among all genetic variants; and **(4)** Result visualization: Forest plots, scatter plots, and leave-one-out plots were generated to visualize the effect estimates (1-3).

The same analysis was conducted with cardiometabolic traits as exposures with the biomarkers as outcomes to assess potential bidirectionality.

#### 7. Linkage Disequilibrium Score Regression Analysis

To characterize the genetic architecture of candidate biomarkers and inform multivariable analysis strategy, we performed linkage disequilibrium score regression (LDSC) using the GenomicSEM R package (v0.0.5). LDSC estimates genetic correlation ($r_g$) between trait pairs by regressing the product of z-scores on LD scores, providing estimates of shared genetic architecture that are robust to confounding from population stratification.

GWAS summary statistics for all 16 candidate biomarkers were standardized using the munge function, which filtered SNPs to the HapMap3 reference panel (~1.2M SNPs), aligned alleles to the 1000 Genomes Phase 3 European reference, and applied quality control filters (MAF > 0.01, INFO > 0.90 when available). Duplicate SNPs and variants in the MHC region (chr6:25–34 Mb) were excluded. Palindromic SNPs (A/T, G/C) were removed to prevent strand mismatches.

Genetic correlations were estimated for all 120 pairwise combinations using the ldsc function with pre-computed LD scores from the 1000 Genomes Phase 3 European reference panel (503 individuals, distributed with GenomicSEM package). Standard errors were computed using block jackknife with 200 genome-wide blocks. For binary traits, liability-scale genetic correlations were estimated by providing sample and population prevalence. The LDSC intercept was examined for each trait to assess potential confounding from population stratification.

Hierarchical clustering was performed using the hclust function in R with complete linkage on the genetic correlation distance matrix ($D = 1 - |r_g|$). A height threshold of 0.85 was selected based on dendrogram structure and biological interpretability, corresponding to a minimum absolute genetic correlation of $|r_g| = 0.15$ for cluster membership. This threshold was validated by varying between 0.75 and 0.95.

#### 8. Multivariate Mendelian Randomization

Multivariable Mendelian randomization (MVMR) extends standard MR by simultaneously estimating independent causal effects of multiple correlated exposures on an outcome. For genetic variant $j$ and exposures $X_1, X_2, \ldots, X_K$, the MVMR model assumes:

$$\Gamma Y_j = \beta_1 \Gamma X_{1j} + \beta_2 \Gamma X_{2j} + \ldots + \beta_K \Gamma X_{Kj} + \varepsilon_j,$$

where $\Gamma$ terms represent genetic associations and $\beta$ coefficients represent conditional causal effects. MVMR requires three assumptions: (1) genetic variants are associated with at least one exposure (relevance), (2) variants are independent of confounders (independence), and (3) variants affect the outcome only through the modeled exposures (exclusion restriction).

For the focused Cluster, genetic instruments were selected by identifying SNPs reaching genome-wide significance ($P < 5\times10^{-8}$) for at least one exposure, followed by LD clumping ($r^2 < 0.001$, 10,000 kb window) using 1000 Genomes European reference. Variants were harmonized across all GWAS, with palindromic SNPs excluded when MAF was 0.40–0.60. IVW MVMR was implemented using the MVMR R package (v0.4), which estimates causal effects by regressing variant-outcome associations on variant-exposure associations, weighted by inverse variances:

$$(\Gamma X' W \Gamma X) - 1 \Gamma X' W \Gamma Y$$

Instrument strength was assessed using conditional F-statistics for each exposure, computed as the squared t-statistic from regressing each exposure's GWAS effects on other exposures' GWAS effects, weighted by inverse variances. $F > 10$ indicates strong instruments. Cochran's Q statistic was computed to assess heterogeneity in variant-specific estimates, with significant Q potentially indicating pleiotropy violations.

As a sensitivity analysis, naive MVMR was performed including all biomarkers simultaneously across nine outcomes. Instruments were selected using the same approach (genome-wide significant for ≥1 biomarker, LD clumping $r^2 < 0.001$). As anticipated for high-dimensional MVMR, this resulted in weak instruments due to multicollinearity. However, this analysis served to validate that focused Cluster findings were robust and not artifacts of selective biomarker inclusion.

All analyses were conducted in R version 4.3.0 using GenomicSEM (v0.0.5) for genetic correlations and MVMR (v0.4) for multivariable MR. Hierarchical clustering used the hclust function (stats package) and visualization used ggplot2 (v3.4.2).

### 9. Software and Implementation Details

All analyses were conducted using R (v4.3.3) with the following key packages: TwoSampleMR (v0.6.8) for MR analysis and data harmonization, ieugwasr (v1.0.1) for accessing OpenGWAS data and PheWAS analysis, ggplot2 (v3.5.1) for visualization, data.table (v1.16.1) for data manipulation, dplyr (v1.1.4) for data transformation, circlize (v0.3.16) for circular representation of PheWAS and colocalization results, and geni.plots (v0.1.2) for colocalization visualization. PWCoCo colocalization analysis was performed on a high-performance computing system using custom scripts. PLINK v1.9 was used for handling genetic data, particularly for LD calculations, file formatting, and genetic instrument clumping. A 1000 Genomes Reference Data set was also used. Genetic correlation analyses were performed using the GenomicSEM R package (version 0.0.5) in R version 4.3.0. All MVMR analyses were conducted using the MVMR R package (version 0.4). Hierarchical clustering and visualization were performed in R using the hclust function (stats package) and ggplot2 (version 3.5.1) for figure generation.

### 10. Statistical Analysis and Interpretation

#### 10.1. Significance Thresholds and Bonferroni P-Value Adjustment

To account for multiple-testing (controlling for increased risk of type I error), Bonferroni adjustment was made on P-values by dividing the overall significance threshold by the number of tests being run. This adjustment results in a stricter significance level for each individual test, reducing the overall probability of a false positive.

- Forward clozapine plasma on cardiometabolic traits ($P=0.05/27=1.85\times10^{-3}$)
- Reverse clozapine plasma on cardiometabolic traits ($P=0.05/27=1.85\times10^{-3}$)
- Forward candidate biomarkers on cardiometabolic traits ($P=0.05/9=5.56\times10^{-3}$)
- Reverse candidate biomarkers on cardiometabolic traits ($P=0.05/28=1.79\times10^{-3}$)

#### 10.2. Causal Inference Framework

Interpretation of results follows the principles of Mendelian randomization as a causal inference method, requiring the following assumptions: **(1)** The genetic variants are robustly associated with the exposure; **(2)** The genetic variants affect the outcome only through the exposure; and **(3)** The genetic variants are not associated with confounders.

The multi-layered approach employed in this study was specifically designed to address potential violations of these assumptions through sensitivity analyses, colocalization methods, and adjusting for potential confounders.

#### 10.3. Interpretation Considerations and Limitations

Several methodological aspects require caution when interpreting results from this multi-layered analysis: **(1)** while our colocalization approach identified significant shared genetic signals between clozapine plasma traits and identified biomarkers, these statistical associations do not definitively prove biological causality or fully elucidate the molecular mechanisms involved. The identified colocalized biomarkers represent promising candidates for further experimental validation; **(2)** despite our comprehensive approach to addressing pleiotropy through sensitivity analyses and adjusting for confounding factors, residual horizontal pleiotropy may still influence some findings. While MR-Egger intercepts suggested minimal directional pleiotropy, this test has limited power, especially when using smaller numbers of genetic instruments; and **(3)** our analysis focused primarily on individuals of European ancestry due to data availability, limiting the generalizability of our findings to other populations. Genetic architecture and metabolic responses to clozapine may differ across ancestral groups, highlighting the need for future studies with diverse population representation.

### 11. References

1. Emdin CA, Khera AV, Kathiresan S. Mendelian Randomization. JAMA. 2017;318(19):1925-6.

2. Sanderson E, Glymour MM, Holmes MV, Kang H, Morrison J, Munafo MR, et al. Mendelian randomization. Nat Rev Methods Primers. 2022;2.

3. Wickham H. ggplot2: Springer-Verlag New York; 2016.

4. Bastarache L, Denny JC, Roden DM. Phenome-Wide Association Studies. JAMA. 2022;327(1):75-6.

5. Denny JC, Bastarache L, Roden DM. Phenome-Wide Association Studies as a Tool to Advance Precision Medicine. Annu Rev Genomics Hum Genet. 2016;17:353-73.

6. Robinson JW, Hemani G, Babaei MS, Huang Y, Baird DA, Tsai EA, et al. 2022.
