## Supplementary Tables for "Genetic Epidemiological Pipeline Identifies Candidate Markers of Clozapine-Induced Metabolic Dysfunction Revealing Potential Avenues for Precision Clozapine Prescription"

Supplementary Table 1 - Clozapine Traits' Effects on Cardiometabolic Outcomes (MR)

| **Exposure** | **Outcome** | **MR Method** | **nSNP** | **Beta** | **SE** | **P-Value** | **F Stat (Mean)** | **F Stat (Min)** | **Q Stat** | **Q P-Value** | **I^2** | **Egger Intercept** | **SE Egger Intercept** | **Egger Intercept P-Value** |
|---|---|---|---|---|---|---|---|---|---|---|---|---|---|---|
| Norclozapine Plasma | HbA1c | IVW (LD-corrected) | 2 | 0.005663635 | 0.363887411 | 0.987582026 | 358.120146 | 20.10514773 | NA | NA | NA | NA | NA | NA |
| Norclozapine Plasma | HbA1c | MR Egger (LD-corrected) | 2 | 0.002665231 | NA | NA | 358.120146 | 20.10514773 | 0.001318309 | 0.971036355 | 0 | -0.004151356 | 0.114335584 | 0.916004796 |
| Norclozapine Plasma | HbA1c | Weighted median (LD-corrected) | 2 | -0.041353291 | 0.021424171 | 0.053580009 | 358.120146 | 20.10514773 | NA | NA | NA | NA | NA | NA |
| Norclozapine Plasma | HDL | IVW (LD-corrected) | 3 | 0.011113608 | 0.490017785 | 0.981905521 | 245.3419288 | 19.7854944 | NA | NA | NA | NA | NA | NA |
| Norclozapine Plasma | HDL | MR Egger (LD-corrected) | 3 | 0.003821647 | NA | NA | 245.3419288 | 19.7854944 | 0.000675429 | 0.999662343 | 0 | -0.005935779 | 0.228432911 | 0.853545528 |
| Norclozapine Plasma | HDL | Weighted median (LD-corrected) | 3 | -0.059379084 | 0.034756546 | 0.087556806 | 245.3419288 | 19.7854944 | NA | NA | NA | NA | NA | NA |
| Norclozapine Plasma | LDL | IVW (LD-corrected) | 3 | -0.03262356 | 1.433046901 | 0.981837589 | 245.3419288 | 19.7854944 | NA | NA | NA | NA | NA | NA |
| Norclozapine Plasma | LDL | MR Egger (LD-corrected) | 3 | 0.004928147 | NA | NA | 245.3419288 | 19.7854944 | 0.000654138 | 0.999672984 | 0 | -0.00591299 | 0.231513069 | 0.93299973 |
| Norclozapine Plasma | LDL | Weighted median (LD-corrected) | 3 | -0.0606238 | 0.015737798 | 0.000117102 | 245.3419288 | 19.7854944 | NA | NA | NA | NA | NA | NA |
| Norclozapine Plasma | Triglycerides | IVW (LD-corrected) | 3 | -0.014706882 | 1.622377521 | 0.992767261 | 245.3419288 | 19.7854944 | NA | NA | NA | NA | NA | NA |

| | | | | | | | | | | | | | | |
|---|---|---|---|---|---|---|---|---|---|---|---|---|---|---|
| Norclozapine Plasma | Triglycerides | MR Egger (LD-corrected) | 3 | 0.004048142 | NA | NA | 245.3419288 | 19.7854944 | 0.000443975 | 0.999778037 | 0 | -0.005561429 | 0.264459116 | 0.947248169 |
| Norclozapine Plasma | Triglycerides | Weighted median (LD-corrected) | 3 | -0.055010485 | 0.012322127 | 8.03E-06 | 245.3419288 | 19.7854944 | NA | NA | NA | NA | NA | NA |
| Norclozapine Plasma | MetS | IVW (LD-corrected) | 3 | 0.004184411 | 0.003112573 | 0.17883274 | 245.3419288 | 19.7854944 | NA | NA | NA | NA | NA | NA |
| Norclozapine Plasma | MetS | MR Egger (LD-corrected) | 3 | 0.00259982 | NA | NA | 245.3419288 | 19.7854944 | 41.95910882 | 7.74E-10 | 95.23345453 | -0.003642965 | 0.000850198 | 0.038012163 |
| Norclozapine Plasma | MetS | Weighted median (LD-corrected) | 3 | 0.008361582 | 0.047264524 | 0.859578838 | 245.3419288 | 19.7854944 | NA | NA | NA | NA | NA | NA |
| Norclozapine Plasma | DBP | IVW (LD-corrected) | 10 | -0.111882181 | 0.246840666 | 0.650363922 | 99.70394317 | 20.10514773 | NA | NA | NA | NA | NA | NA |
| Norclozapine Plasma | DBP | MR Egger (LD-corrected) | 10 | -0.031648831 | NA | NA | 99.70394317 | 20.10514773 | 0.128975247 | 0.99999992 | 0 | -0.026842823 | 0.079009059 | 0.466526898 |
| Norclozapine Plasma | DBP | Weighted median (LD-corrected) | 10 | -0.136976649 | 0.105632138 | 0.19472317 | 99.70394317 | 20.10514773 | NA | NA | NA | NA | NA | NA |
| Norclozapine Plasma | SBP | IVW (LD-corrected) | 10 | 0.071693847 | 0.652322679 | 0.912484325 | 99.70394317 | 20.10514773 | NA | NA | NA | NA | NA | NA |
| Norclozapine Plasma | SBP | MR Egger (LD-corrected) | 10 | 0.000203746 | NA | NA | 99.70394317 | 20.10514773 | 0.047812973 | 0.999999999 | 0 | -0.04786301 | 0.222540679 | 1.21E-05 |
| Norclozapine Plasma | SBP | Weighted median (LD-corrected) | 10 | 0.248617239 | 0.238729024 | 0.29768055 | 99.70394317 | 20.10514773 | NA | NA | NA | NA | NA | NA |
| Norclozapine Plasma | T2D | IVW (LD-corrected) | 12 | 0.172450271 | 0.204148862 | 0.398262692 | 92.50154795 | 19.7854944 | NA | NA | NA | NA | NA | NA |

| | | | | | | | | | | | | | | |
|---|---|---|---|---|---|---|---|---|---|---|---|---|---|---|
| Norclozapine Plasma | T2D | MR Egger (LD-corrected) | 12 | -0.044859341 | NA | NA | 92.50154795 | 19.7854944 | 1.57853218 | 0.99951256 | 0 | -0.048530873 | 0.040139552 | 0.019914403 |
| Norclozapine Plasma | T2D | Weighted median (LD-corrected) | 12 | 0.299195531 | 0.19319662 | 0.121463906 | 92.50154795 | 19.7854944 | NA | NA | NA | NA | NA | NA |
| Norclozapine Plasma | BMI | IVW (LD-corrected) | 12 | -0.014171573 | 0.068597334 | 0.836329501 | 92.50154795 | 19.7854944 | NA | NA | NA | NA | NA | NA |
| Norclozapine Plasma | BMI | MR Egger (LD-corrected) | 12 | -0.00823656 | NA | NA | 92.50154795 | 19.7854944 | 0.002781623 | 1 | 0 | -0.001543774 | 0.052514678 | 0.956864267 |
| Norclozapine Plasma | BMI | Weighted median (LD-corrected) | 12 | -0.015341564 | 0.006003113 | 0.010600442 | 92.50154795 | 19.7854944 | NA | NA | NA | NA | NA | NA |
| Cloz-Norcloz Plasma Ratio | DBP | IVW (LD-corrected) | 17 | 0.209895676 | 0.023167129 | 1.30E-19 | 30.2245365 | 19.89666534 | NA | NA | NA | NA | NA | NA |
| Cloz-Norcloz Plasma Ratio | DBP | MR Egger (LD-corrected) | 17 | 0.030628848 | NA | NA | 30.2245365 | 19.89666534 | 0.678287536 | 0.999999997 | 0 | -0.020532052 | 0.028197883 | 0.734049618 |
| Cloz-Norcloz Plasma Ratio | DBP | Weighted median (LD-corrected) | 17 | 0.21086976 | 0.064991118 | 0.001176184 | 30.2245365 | 19.89666534 | NA | NA | NA | NA | NA | NA |
| Cloz-Norcloz Plasma Ratio | SBP | IVW (LD-corrected) | 17 | -0.072733882 | 0.085258125 | 0.393602711 | 30.2245365 | 19.89666534 | NA | NA | NA | NA | NA | NA |
| Cloz-Norcloz Plasma Ratio | SBP | MR Egger (LD-corrected) | 17 | -0.048317557 | NA | NA | 30.2245365 | 19.89666534 | 19.3061321 | 0.253108204 | 17.12477718 | -0.047688421 | 0.010899572 | 0.829708588 |
| Cloz-Norcloz Plasma Ratio | SBP | Weighted median (LD-corrected) | 17 | 0.366154603 | 0.376780965 | 0.331151559 | 30.2245365 | 19.89666534 | NA | NA | NA | NA | NA | NA |
| Cloz-Norcloz Plasma Ratio | T2D | IVW (LD-corrected) | 17 | 0.555227286 | 0.165464658 | 0.000792033 | 30.13758144 | 19.89666534 | NA | NA | NA | NA | NA | NA |

| | | | | | | | | | | | | | |
|---|---|---|---|---|---|---|---|---|---|---|---|---|---|
| Cloz-Norcloz Plasma Ratio | T2D | MR Egger (LD-corrected) | 17 | 0.121371711 | NA | NA | 30.13758144 | 19.89666534 | 6.42198589 | 0.982862674 | 0 | -0.09947558 | 0.042730855 | 0.226642222 |
| Cloz-Norcloz Plasma Ratio | T2D | Weighted median (LD-corrected) | 17 | 0.542614494 | 0.315478967 | 0.085437817 | 30.13758144 | 19.89666534 | NA | NA | NA | NA | NA | NA |
| Cloz-Norcloz Plasma Ratio | BMI | IVW (LD-corrected) | 18 | 0.018595983 | 0.174904762 | 0.915327971 | 29.68585626 | 19.89666534 | NA | NA | NA | NA | NA | NA |
| Cloz-Norcloz Plasma Ratio | BMI | MR Egger (LD-corrected) | 18 | 0.001273302 | NA | NA | 29.68585626 | 19.89666534 | 0.003168193 | 1 | 0 | -0.002794265 | 0.051660515 | 0.976547964 |
| Cloz-Norcloz Plasma Ratio | BMI | Weighted median (LD-corrected) | 18 | 0.023628265 | 0.00657974 | 0.000329331 | 29.68585626 | 19.89666534 | NA | NA | NA | NA | NA | NA |
| Clozapine Plasma | HbA1c | IVW (LD-corrected) | 6 | 0.032215382 | 0.463197658 | 0.944551861 | 29.59310964 | 20.20769488 | NA | NA | NA | NA | NA | NA |
| Clozapine Plasma | HbA1c | MR Egger (LD-corrected) | 6 | 0.020119763 | NA | NA | 29.59310964 | 20.20769488 | 4.73E-05 | 1 | 0 | -0.001810285 | 0.289051973 | 0.995003012 |
| Clozapine Plasma | HbA1c | Weighted median (LD-corrected) | 6 | 0.031248784 | 0.003569804 | 2.07E-18 | 29.59310964 | 20.20769488 | NA | NA | NA | NA | NA | NA |
| Clozapine Plasma | HDL | IVW (LD-corrected) | 7 | 0.051786991 | 0.091792518 | 0.572635434 | 28.58390925 | 20.20769488 | NA | NA | NA | NA | NA | NA |
| Clozapine Plasma | HDL | MR Egger (LD-corrected) | 7 | 0.03079577 | NA | NA | 28.58390925 | 20.20769488 | 0.002079172 | 1 | 0 | -0.002288648 | 0.051314906 | 0.964426086 |
| Clozapine Plasma | HDL | Weighted median (LD-corrected) | 7 | 0.053374952 | 0.005431462 | 8.62E-23 | 28.58390925 | 20.20769488 | NA | NA | NA | NA | NA | NA |
| Clozapine Plasma | LDL | IVW (LD-corrected) | 7 | 0.046831807 | 0.298085749 | 0.875159333 | 28.58390925 | 20.20769488 | NA | NA | NA | NA | NA | NA |
| Clozapine Plasma | LDL | MR Egger (LD-corrected) | 7 | 0.032472843 | NA | NA | 28.58390925 | 20.20769488 | 0.000330753 | 1 | 0 | -0.002608471 | 0.155301335 | 0.986599206 |

| | | | | | | | | | | | | | | |
|---|---|---|---|---|---|---|---|---|---|---|---|---|---|---|
| Clozapine Plasma | LDL | Weighted median (LD-corrected) | 7 | 0.044602785 | 0.006578338 | 1.20E-11 | 28.58390925 | 20.20769488 | NA | NA | NA | NA | NA | NA |
| Clozapine Plasma | Triglycerides | IVW (LD-corrected) | 7 | 0.048083776 | 0.077618243 | 0.535593182 | 28.58390925 | 20.20769488 | NA | NA | NA | NA | NA | NA |
| Clozapine Plasma | Triglycerides | MR Egger (LD-corrected) | 7 | 0.019682168 | NA | NA | 28.58390925 | 20.20769488 | 0.000181811 | 1 | 0 | -0.003250201 | 0.287063885 | 0.990966368 |
| Clozapine Plasma | Triglycerides | Weighted median (LD-corrected) | 7 | 0.04803685 | 0.003920202 | 1.61E-34 | 28.58390925 | 20.20769488 | NA | NA | NA | NA | NA | NA |
| Clozapine Plasma | MetS | IVW (LD-corrected) | 7 | -0.009837229 | 0.004530063 | 0.02989013 | 28.58390925 | 20.20769488 | NA | NA | NA | NA | NA | NA |
| Clozapine Plasma | MetS | MR Egger (LD-corrected) | 7 | -0.029117219 | NA | NA | 28.58390925 | 20.20769488 | 34.63794399 | 5.06E-06 | 82.67795571 | -0.002460813 | 0.002139193 | 0.250001272 |
| Clozapine Plasma | MetS | Weighted median (LD-corrected) | 7 | -0.00130527 | 0.019885487 | 0.947664995 | 28.58390925 | 20.20769488 | NA | NA | NA | NA | NA | NA |
| Cloz-Norcloz Plasma Ratio | HbA1c | IVW (LD-corrected) | 7 | 0.024560281 | 0.244487354 | 0.919982119 | 27.46053471 | 19.89666534 | NA | NA | NA | NA | NA | NA |
| Cloz-Norcloz Plasma Ratio | HbA1c | MR Egger (LD-corrected) | 7 | 0.005545249 | NA | NA | 27.46053471 | 19.89666534 | 0.012450472 | 0.99999996 | 0 | -0.005012107 | 0.047522723 | 0.971036355 |
| Cloz-Norcloz Plasma Ratio | HbA1c | Weighted median (LD-corrected) | 7 | 0.029103868 | 0.030405884 | 0.338476998 | 27.46053471 | 19.89666534 | NA | NA | NA | NA | NA | NA |
| Cloz-Norcloz Plasma Ratio | HDL | IVW (LD-corrected) | 7 | 0.006951815 | 0.248225862 | 0.977657362 | 27.37530277 | 19.89666534 | NA | NA | NA | NA | NA | NA |
| Cloz-Norcloz Plasma Ratio | HDL | MR Egger (LD-corrected) | 7 | 0.006465766 | NA | NA | 27.37530277 | 19.89666534 | 0.035265261 | 0.999999098 | 0 | -0.006465105 | 0.035022899 | 0.979269478 |

| | | | | | | | | | | | | | | |
|---|---|---|---|---|---|---|---|---|---|---|---|---|---|---|
| Cloz-Norcloz Plasma Ratio | HDL | Weighted median (LD-corrected) | 7 | -0.052730474 | 0.046521644 | 0.257020671 | 27.37530277 | 19.89666534 | NA | NA | NA | NA | NA | NA |
| Cloz-Norcloz Plasma Ratio | LDL | IVW (LD-corrected) | 7 | 0.009614707 | 0.650172269 | 0.988201363 | 27.37530277 | 19.89666534 | NA | NA | NA | NA | NA | NA |
| Cloz-Norcloz Plasma Ratio | LDL | MR Egger (LD-corrected) | 7 | -0.004078833 | NA | NA | 27.37530277 | 19.89666534 | 0.007475178 | 0.999999991 | 0 | -0.006700166 | 0.079696218 | 0.979623741 |
| Cloz-Norcloz Plasma Ratio | LDL | Weighted median (LD-corrected) | 7 | 0.056917023 | 0.050793979 | 0.262480887 | 27.37530277 | 19.89666534 | NA | NA | NA | NA | NA | NA |
| Cloz-Norcloz Plasma Ratio | Triglycerides | IVW (LD-corrected) | 7 | 0.050978759 | 0.083163708 | 0.539881064 | 27.37530277 | 19.89666534 | NA | NA | NA | NA | NA | NA |
| Cloz-Norcloz Plasma Ratio | Triglycerides | MR Egger (LD-corrected) | 7 | -0.006783066 | NA | NA | 27.37530277 | 19.89666534 | 0.004604597 | 0.999999998 | 0 | -0.006231228 | 0.094180159 | 0.983222166 |
| Cloz-Norcloz Plasma Ratio | Triglycerides | Weighted median (LD-corrected) | 7 | 0.051318627 | 0.028709611 | 0.073855602 | 27.37530277 | 19.89666534 | NA | NA | NA | NA | NA | NA |
| Cloz-Norcloz Plasma Ratio | MetS | IVW (LD-corrected) | 7 | -0.017993681 | 0.004743145 | 0.000148468 | 27.37530277 | 19.89666534 | NA | NA | NA | NA | NA | NA |
| Cloz-Norcloz Plasma Ratio | MetS | MR Egger (LD-corrected) | 7 | -0.012164309 | NA | NA | 27.37530277 | 19.89666534 | 56.25375939 | 2.59E-10 | 89.33404618 | 0.001331815 | 0.000641924 | 1.83E-05 |
| Cloz-Norcloz Plasma Ratio | MetS | Weighted median (LD-corrected) | 7 | -0.006439093 | 0.025688364 | 0.802075557 | 27.37530277 | 19.89666534 | NA | NA | NA | NA | NA | NA |
| Clozapine Plasma | DBP | IVW (LD-corrected) | 16 | -0.05003198 | 0.182494314 | 0.783964448 | 24.57485479 | 19.54228913 | NA | NA | NA | NA | NA | NA |
| Clozapine Plasma | DBP | MR Egger (LD-corrected) | 16 | 0.003690956 | NA | NA | 24.57485479 | 19.54228913 | 0.907051593 | 0.999999873 | 0 | -0.026913615 | 0.028485597 | 0.344753421 |

| | | | | | | | | | | | | | | |
|---|---|---|---|---|---|---|---|---|---|---|---|---|---|---|
| Clozapine Plasma | DBP | Weighted median (LD-corrected) | 16 | -0.183184565 | 0.180608128 | 0.310456157 | 24.57485479 | 19.54228913 | NA | NA | NA | NA | NA | NA |
| Clozapine Plasma | SBP | IVW (LD-corrected) | 16 | 0.331239776 | 0.05383875 | 7.63E-10 | 24.57485479 | 19.54228913 | NA | NA | NA | NA | NA | NA |
| Clozapine Plasma | SBP | MR Egger (LD-corrected) | 16 | 0.031877396 | NA | NA | 24.57485479 | 19.54228913 | 0.540915994 | 0.999999997 | 0 | -0.037510006 | 0.055144737 | 0.496371451 |
| Clozapine Plasma | SBP | Weighted median (LD-corrected) | 16 | 0.330551405 | 0.01666986 | 1.66E-87 | 24.57485479 | 19.54228913 | NA | NA | NA | NA | NA | NA |
| Clozapine Plasma | T2D | IVW (LD-corrected) | 17 | 0.485407043 | 0.036188229 | 5.05E-41 | 24.45449315 | 19.54228913 | NA | NA | NA | NA | NA | NA |
| Clozapine Plasma | T2D | MR Egger (LD-corrected) | 17 | -0.090308044 | NA | NA | 24.45449315 | 19.54228913 | 14.73386012 | 0.544210571 | 0 | -0.0615095 3 | 0.020794407 | 0.003096578 |
| Clozapine Plasma | T2D | Weighted median (LD-corrected) | 17 | 0.520664097 | 0.183545631 | 0.004558226 | 24.45449315 | 19.54228913 | NA | NA | NA | NA | NA | NA |
| Clozapine Plasma | BMI | IVW (LD-corrected) | 17 | 0.01604503 | 0.160931507 | 0.920581719 | 24.45449315 | 19.54228913 | NA | NA | NA | NA | NA | NA |
| Clozapine Plasma | BMI | MR Egger (LD-corrected) | 17 | 0.004042266 | NA | NA | 24.45449315 | 19.54228913 | 0.002684616 | 1 | 0 | -0.002743244 | 0.0530733 | 0.958777437 |
| Clozapine Plasma | BMI | Weighted median (LD-corrected) | 17 | 0.016491121 | 0.01259765 | 0.190512923 | 24.45449315 | 19.54228913 | NA | NA | NA | NA | NA | NA |

#### Supplementary Table 2 - Cardiometabolic Traits' Effects on Clozapine Traits (MR)

| Exposure | Outcome | MR Method | nSNP | Beta | SE | P-Value | F Stat (Mean) | F Stat (Min) | Q Stat | Q Stat P-Value | I^2 | Egger Intercept | Egger Intercept SE |
|---|---|---|---|---|---|---|---|---|---|---|---|---|---|
| BMI | Clozapine Plasma | IVW (LD-corrected) | 197 | 0.024476768 | 0.079719066 | 0.758814588 | 74.81586599 | 30.16352643 | 206.2459571 | 0.293811898 | 4.967834138 | NA | NA |
| BMI | Clozapine Plasma | MR Egger (LD-corrected) | 197 | -0.025207735 | 0.081513065 | 0.75713305 | 74.81586599 | 30.16352643 | NA | NA | NA | 0.004706978 | 0.001611232 |
| BMI | Clozapine Plasma | Weighted median (LD-corrected) | 197 | 0.162879535 | 0.184030357 | 0.376119504 | 74.81586599 | 30.16352643 | NA | NA | NA | NA | NA |
| BMI | Clozapine-Norclozapine Plasma Ratio | IVW (LD-corrected) | 197 | 0.085944907 | 0.078680319 | 0.274687888 | 74.81586599 | 30.16352643 | 204.2862018 | 0.327682007 | 4.056173008 | NA | NA |
| BMI | Clozapine-Norclozapine Plasma Ratio | MR Egger (LD-corrected) | 197 | 0.039380251 | 0.080435645 | 0.624426136 | 74.81586599 | 30.16352643 | NA | NA | NA | 0.004430439 | 0.001590106 |
| BMI | Clozapine-Norclozapine Plasma Ratio | Weighted median (LD-corrected) | 197 | 0.049854839 | 0.100622655 | 0.620273076 | 74.81586599 | 30.16352643 | NA | NA | NA | NA | NA |
| BMI | Norclozapine Plasma | IVW (LD-corrected) | 197 | -0.012751662 | 0.077874755 | 0.869931282 | 74.81586599 | 30.16352643 | 189.8364615 | 0.610591524 | 0 | NA | NA |
| BMI | Norclozapine Plasma | MR Egger (LD-corrected) | 197 | -0.039801325 | 0.079585369 | 0.616998629 | 74.81586599 | 30.16352643 | NA | NA | NA | 0.002589294 | 0.001571017 |
| BMI | Norclozapine Plasma | Weighted median (LD-corrected) | 197 | 0.197714804 | 0.154471808 | 0.200565877 | 74.81586599 | 30.16352643 | NA | NA | NA | NA | NA |
| DBP | Clozapine Plasma | IVW (LD-corrected) | 199 | -0.004221437 | 0.00844478 | 0.617154552 | 86.27207084 | 29.70012014 | 231.3126236 | 0.052459443 | 14.40155885 | NA | NA |
| DBP | Clozapine Plasma | MR Egger (LD-corrected) | 199 | -0.002904702 | 0.008483782 | 0.732062783 | 86.27207084 | 29.70012014 | NA | NA | NA | -0.002555204 | 0.001576822 |

| | | | | | | | | | | | | | |
|---|---|---|---|---|---|---|---|---|---|---|---|---|---|
| DBP | Clozapine Plasma | Weighted median (LD-corrected) | 199 | 0.012989361 | 0.019082695 | 0.496068943 | 86.27207084 | 29.70012014 | NA | NA | NA | NA | NA |
| DBP | Clozapine Plasma | IVW (LD-corrected) | 208 | -0.004932167 | 0.004858066 | 0.309985131 | 81.18791338 | 29.93981381 | 215.561611 | 0.327157379 | 3.971769833 | NA | NA |
| DBP | Clozapine Plasma | MR Egger (LD-corrected) | 208 | -0.004073127 | 0.004896037 | 0.40545223 | 81.18791338 | 29.93981381 | NA | NA | NA | -0.002162276 | 0.00153186 |
| DBP | Clozapine Plasma | Weighted median (LD-corrected) | 208 | -0.007414677 | 0.009599002 | 0.439852413 | 81.18791338 | 29.93981381 | NA | NA | NA | NA | NA |
| DBP | Clozapine-Norclozapine Plasma Ratio | IVW (LD-corrected) | 199 | -0.000615343 | 0.008326263 | 0.941086916 | 86.27207084 | 29.70012014 | 236.4494097 | 0.03197096 | 16.26115699 | NA | NA |
| DBP | Clozapine-Norclozapine Plasma Ratio | MR Egger (LD-corrected) | 199 | 0.001099926 | 0.00835972 | 0.895320844 | 86.27207084 | 29.70012014 | NA | NA | NA | -0.003566948 | 0.001553752 |
| DBP | Clozapine-Norclozapine Plasma Ratio | Weighted median (LD-corrected) | 199 | 0.003516155 | 0.020802666 | 0.865777556 | 86.27207084 | 29.70012014 | NA | NA | NA | NA | NA |
| DBP | Clozapine-Norclozapine Plasma Ratio | IVW (LD-corrected) | 208 | -0.00157859 | 0.004803617 | 0.742438924 | 81.18791338 | 29.93981381 | 260.3180112 | 0.007018692 | 20.48187558 | NA | NA |
| DBP | Clozapine-Norclozapine Plasma Ratio | MR Egger (LD-corrected) | 208 | -0.001206632 | 0.004839643 | 0.803111251 | 81.18791338 | 29.93981381 | NA | NA | NA | -0.000955196 | 0.001513637 |
| DBP | Clozapine-Norclozapine Plasma Ratio | Weighted median (LD-corrected) | 208 | -0.004269743 | 0.010928049 | 0.696008543 | 81.18791338 | 29.93981381 | NA | NA | NA | NA | NA |
| DBP | Norclozapine Plasma | IVW (LD-corrected) | 199 | -0.003886872 | 0.00825122 | 0.637593319 | 86.27207084 | 29.70012014 | 225.6189641 | 0.08670963 | 12.2414196 | NA | NA |

| | | | | | | | | | | | | | |
|---|---|---|---|---|---|---|---|---|---|---|---|---|---|
| DBP | Norclozapine Plasma | MR Egger (LD-corrected) | 199 | -0.003566705 | 0.008291287 | 0.667068328 | 86.27207084 | 29.70012014 | NA | NA | NA | -0.000604849 | 0.001538041 |
| DBP | Norclozapine Plasma | Weighted median (LD-corrected) | 199 | 0.008856539 | 0.009467339 | 0.349539185 | 86.27207084 | 29.70012014 | NA | NA | NA | NA | NA |
| DBP | Norclozapine Plasma | IVW (LD-corrected) | 208 | -0.003593263 | 0.004743547 | 0.448747012 | 81.18791338 | 29.93981381 | 212.4905544 | 0.38201339 | 2.583905167 | NA | NA |
| DBP | Norclozapine Plasma | MR Egger (LD-corrected) | 208 | -0.002910866 | 0.00478236 | 0.54274499 | 81.18791338 | 29.93981381 | NA | NA | NA | -0.00167587 | 0.001493296 |
| DBP | Norclozapine Plasma | Weighted median (LD-corrected) | 208 | 0.00418177 | 0.007293063 | 0.566380615 | 81.18791338 | 29.93981381 | NA | NA | NA | NA | NA |
| HbA1c | Clozapine Plasma | IVW (LD-corrected) | 9 | -0.419704452 | 0.228752949 | 0.066542754 | 64.04395192 | 32.76199546 | 11.65395398 | 0.167324195 | 31.35377046 | NA | NA |
| HbA1c | Clozapine Plasma | MR Egger (LD-corrected) | 9 | -0.342617742 | 0.248722329 | 0.168354319 | 64.04395192 | 32.76199546 | NA | NA | NA | -0.006519644 | 0.008258518 |
| HbA1c | Clozapine Plasma | Weighted median (LD-corrected) | 9 | -0.407141456 | 0.332597637 | 0.220904652 | 64.04395192 | 32.76199546 | NA | NA | NA | NA | NA |
| HbA1c | Clozapine-Norclozapine Plasma Ratio | IVW (LD-corrected) | 9 | -0.112500389 | 0.228981584 | 0.623208915 | 64.04395192 | 32.76199546 | 7.630542128 | 0.470365606 | 0 | NA | NA |
| HbA1c | Clozapine-Norclozapine Plasma Ratio | MR Egger (LD-corrected) | 9 | -0.083454618 | 0.251217258 | 0.739737509 | 64.04395192 | 32.76199546 | NA | NA | NA | -0.002329644 | 0.008287859 |
| HbA1c | Clozapine-Norclozapine Plasma Ratio | Weighted median (LD-corrected) | 9 | -0.139898491 | 0.276985812 | 0.613506459 | 64.04395192 | 32.76199546 | NA | NA | NA | NA | NA |

| | | | | | | | | | | | | | |
|---|---|---|---|---|---|---|---|---|---|---|---|---|---|
| HbA1c | Norclozapine Plasma | IVW (LD-corrected) | 9 | -0.369720949 | 0.2227133 | 0.096899214 | 64.04395192 | 32.76199546 | 9.297435923 | 0.317829332 | 13.95477133 | NA | NA |
| HbA1c | Norclozapine Plasma | MR Egger (LD-corrected) | 9 | -0.308928137 | 0.24271159 | 0.203081965 | 64.04395192 | 32.76199546 | NA | NA | NA | -0.005073462 | 0.008051437 |
| HbA1c | Norclozapine Plasma | Weighted median (LD-corrected) | 9 | -0.373077732 | 0.234686155 | 0.111905181 | 64.04395192 | 32.76199546 | NA | NA | NA | NA | NA |
| HDL | Clozapine Plasma | IVW (LD-corrected) | 68 | 0.071736942 | 0.063480092 | 0.258446763 | 109.135486 | 29.94521605 | 95.04062207 | 0.013745147 | 29.50382843 | NA | NA |
| HDL | Clozapine Plasma | MR Egger (LD-corrected) | 68 | 0.058567269 | 0.063788565 | 0.358541993 | 109.135486 | 29.94521605 | NA | NA | NA | 0.005684641 | 0.002704565 |
| HDL | Clozapine Plasma | Weighted median (LD-corrected) | 68 | 0.034167474 | 0.154565977 | 0.825050151 | 109.135486 | 29.94521605 | NA | NA | NA | NA | NA |
| HDL | Clozapine-Norclozapine Plasma Ratio | IVW (LD-corrected) | 68 | -0.009864556 | 0.062559545 | 0.874706853 | 109.135486 | 29.94521605 | 92.49276525 | 0.021271958 | 27.56190193 | NA | NA |
| HDL | Clozapine-Norclozapine Plasma Ratio | MR Egger (LD-corrected) | 68 | -0.012226812 | 0.062871793 | 0.845806216 | 109.135486 | 29.94521605 | NA | NA | NA | 0.001007767 | 0.002669846 |
| HDL | Clozapine-Norclozapine Plasma Ratio | Weighted median (LD-corrected) | 68 | -0.027294717 | 0.14481799 | 0.850503519 | 109.135486 | 29.94521605 | NA | NA | NA | NA | NA |
| HDL | Norclozapine Plasma | IVW (LD-corrected) | 68 | 0.075678976 | 0.062087008 | 0.222875292 | 109.135486 | 29.94521605 | 81.57155378 | 0.108552444 | 17.86352363 | NA | NA |
| HDL | Norclozapine Plasma | MR Egger (LD-corrected) | 68 | 0.064235652 | 0.06239686 | 0.3032592 | 109.135486 | 29.94521605 | NA | NA | NA | 0.00486521 | 0.002640481 |
| HDL | Norclozapine Plasma | Weighted median (LD-corrected) | 68 | 0.115013204 | 0.120934343 | 0.341584884 | 109.135486 | 29.94521605 | NA | NA | NA | NA | NA |

| | | | | | | | | | | | | | |
|---|---|---|---|---|---|---|---|---|---|---|---|---|---|
| LDL | Clozapine Plasma | IVW (LD-corrected) | 49 | -0.010499617 | 0.059620651 | 0.860209842 | 146.5344204 | 27.78540649 | 44.39233882 | 0.621458782 | 0 | NA | NA |
| LDL | Clozapine Plasma | MR Egger (LD-corrected) | 49 | -0.010751146 | 0.059739501 | 0.857178382 | 146.5344204 | 27.78540649 | NA | NA | NA | 0.000219475 | 0.003286443 |
| LDL | Clozapine Plasma | Weighted median (LD-corrected) | 49 | 0.011048154 | NA | NA | 146.5344204 | 27.78540649 | NA | NA | NA | NA | NA |
| LDL | Clozapine-Norclozapine Plasma Ratio | IVW (LD-corrected) | 49 | 0.065675377 | 0.059198918 | 0.267256956 | 146.5344204 | 27.78540649 | 36.70714739 | 0.882641803 | 0 | NA | NA |
| LDL | Clozapine-Norclozapine Plasma Ratio | MR Egger (LD-corrected) | 49 | 0.067449312 | 0.059330281 | 0.255603242 | 146.5344204 | 27.78540649 | NA | NA | NA | -0.001467843 | 0.003265063 |
| LDL | Clozapine-Norclozapine Plasma Ratio | Weighted median (LD-corrected) | 49 | 0.019316793 | NA | NA | 146.5344204 | 27.78540649 | NA | NA | NA | NA | NA |
| LDL | Norclozapine Plasma | IVW (LD-corrected) | 49 | -0.039405164 | 0.058083114 | 0.497500876 | 146.5344204 | 27.78540649 | 45.46696209 | 0.577228764 | 0 | NA | NA |
| LDL | Norclozapine Plasma | MR Egger (LD-corrected) | 49 | -0.041126112 | 0.058204908 | 0.479830852 | 146.5344204 | 27.78540649 | NA | NA | NA | 0.001466115 | 0.003206126 |
| LDL | Norclozapine Plasma | Weighted median (LD-corrected) | 49 | -0.0165451 | NA | NA | 146.5344204 | 27.78540649 | NA | NA | NA | NA | NA |
| MetS | Clozapine Plasma | IVW (LD-corrected) | 46 | 0.097940241 | 0.051366119 | 0.056558282 | 71.18298489 | 29.88623736 | 64.94620642 | 0.027324019 | 30.71188838 | NA | NA |
| MetS | Clozapine Plasma | MR Egger (LD-corrected) | 46 | 0.101283994 | 0.051909625 | 0.051037995 | 71.18298489 | 29.88623736 | NA | NA | NA | -0.001487207 | 0.003332259 |
| MetS | Clozapine Plasma | Weighted median (LD-corrected) | 46 | 0.101008316 | 0.147844833 | 0.494477372 | 71.18298489 | 29.88623736 | NA | NA | NA | NA | NA |

| | | | | | | | | | | | | | |
|---|---|---|---|---|---|---|---|---|---|---|---|---|---|
| MetS | Clozapine-Norclozapine Plasma Ratio | IVW (LD-corrected) | 46 | 0.045688361 | 0.050800652 | 0.36845794 | 71.18298489 | 29.88623736 | 66.69866703 | 0.019441642 | 32.53238482 | NA | NA |
| MetS | Clozapine-Norclozapine Plasma Ratio | MR Egger (LD-corrected) | 46 | 0.052547121 | 0.051347498 | 0.306136326 | 71.18298489 | 29.88623736 | NA | NA | NA | -0.003022468 | 0.003293545 |
| MetS | Clozapine-Norclozapine Plasma Ratio | Weighted median (LD-corrected) | 46 | 0.043074802 | 0.122641189 | 0.725418637 | 71.18298489 | 29.88623736 | NA | NA | NA | NA | NA |
| MetS | Norclozapine Plasma | IVW (LD-corrected) | 46 | 0.077472684 | 0.050088062 | 0.121928478 | 71.18298489 | 29.88623736 | 56.72625308 | 0.112856453 | 20.67165103 | NA | NA |
| MetS | Norclozapine Plasma | MR Egger (LD-corrected) | 46 | 0.077346182 | 0.050612754 | 0.126464012 | 71.18298489 | 29.88623736 | NA | NA | NA | 5.66E-05 | 0.003251502 |
| MetS | Norclozapine Plasma | Weighted median (LD-corrected) | 46 | 0.042081159 | 0.112748137 | 0.708976134 | 71.18298489 | 29.88623736 | NA | NA | NA | NA | NA |
| T2D | Clozapine Plasma | IVW (LD-corrected) | 80 | 0.098122918 | 0.031607847 | 0.001906752 | 87.28145992 | 29.94180028 | 80.20400051 | 0.441051016 | 1.501172639 | NA | NA |
| T2D | Clozapine Plasma | MR Egger (LD-corrected) | 80 | 0.09589911 | 0.031815926 | 0.0025767 | 87.28145992 | 29.94180028 | NA | NA | NA | 0.001536923 | 0.002510703 |
| T2D | Clozapine Plasma | Weighted median (LD-corrected) | 80 | 0.128374996 | 0.075087917 | 0.087328302 | 87.28145992 | 29.94180028 | NA | NA | NA | NA | NA |
| T2D | Clozapine-Norclozapine Plasma Ratio | IVW (LD-corrected) | 80 | 0.020265659 | 0.031079338 | 0.514361098 | 87.28145992 | 29.94180028 | 85.75190109 | 0.282553734 | 7.873762571 | NA | NA |
| T2D | Clozapine-Norclozapine Plasma Ratio | MR Egger (LD-corrected) | 80 | 0.020779582 | 0.031281391 | 0.506511535 | 87.28145992 | 29.94180028 | NA | NA | NA | -0.000358627 | 0.00247704 |
| T2D | Clozapine-Norclozapine Plasma Ratio | Weighted median (LD-corrected) | 80 | -0.004905679 | 0.073378274 | 0.946697415 | 87.28145992 | 29.94180028 | NA | NA | NA | NA | NA |
| T2D | Norclozapine Plasma | IVW (LD-corrected) | 80 | 0.090127116 | 0.030828393 | 0.003461091 | 87.28145992 | 29.94180028 | 81.91159334 | 0.389065155 | 3.55455586 | NA | NA |
| T2D | Norclozapine Plasma | MR Egger (LD-corrected) | 80 | 0.088001941 | 0.03103159 | 0.004569927 | 87.28145992 | 29.94180028 | NA | NA | NA | 0.001464794 | 0.002443687 |

| | | | | | | | | | | | | | |
|---|---|---|---|---|---|---|---|---|---|---|---|---|---|
| T2D | Norclozapine Plasma | Weighted median (LD-corrected) | 80 | 0.135579764 | 0.062379821 | 0.029746053 | 87.28145992 | 29.94180028 | NA | NA | NA | NA | NA |
| Triglycerides | Clozapine Plasma | IVW (LD-corrected) | 47 | -0.048908222 | 0.061265216 | 0.424694521 | 179.9328287 | 30.90051903 | 62.45076815 | 0.053409409 | 26.34197887 | NA | NA |
| Triglycerides | Clozapine Plasma | MR Egger (LD-corrected) | 47 | -0.030005904 | 0.063036582 | 0.634068829 | 179.9328287 | 30.90051903 | NA | NA | NA | 0.004118315 | 0.003232938 |
| Triglycerides | Clozapine Plasma | Weighted median (LD-corrected) | 47 | -0.078822422 | NA | NA | 179.9328287 | 30.90051903 | NA | NA | NA | NA | NA |
| Triglycerides | Clozapine-Norclozapine Plasma Ratio | IVW (LD-corrected) | 47 | 0.047090429 | 0.060933596 | 0.439631571 | 179.9328287 | 30.90051903 | 62.46858238 | 0.05324476 | 26.36298401 | NA | NA |
| Triglycerides | Clozapine-Norclozapine Plasma Ratio | MR Egger (LD-corrected) | 47 | 0.074805589 | 0.062723701 | 0.233017919 | 179.9328287 | 30.90051903 | NA | NA | NA | 0.005953173 | 0.003195804 |
| Triglycerides | Clozapine-Norclozapine Plasma Ratio | Weighted median (LD-corrected) | 47 | 0.112364813 | NA | NA | 179.9328287 | 30.90051903 | NA | NA | NA | NA | NA |
| Triglycerides | Norclozapine Plasma | IVW (LD-corrected) | 47 | -0.069637505 | 0.059238913 | 0.239780135 | 179.9328287 | 30.90051903 | 59.43858962 | 0.088181572 | 22.60920002 | NA | NA |
| Triglycerides | Norclozapine Plasma | MR Egger (LD-corrected) | 47 | -0.067978152 | 0.06095283 | 0.264739731 | 179.9328287 | 30.90051903 | NA | NA | NA | 0.000364841 | 0.003155712 |
| Triglycerides | Norclozapine Plasma | Weighted median (LD-corrected) | 47 | -0.078782624 | NA | NA | 179.9328287 | 30.90051903 | NA | NA | NA | NA | NA |

#### Supplementary Table 3 - PheWAS Results (OpenGWAS)

| Chr | Pos | Beta | SE | P | n | ID | rsID | EA | NEA | EAF | Trait | Query SNP | Ethnicity | Window Start | Window End |
|---|---|---|---|---|---|---|---|---|---|---|---|---|---|---|---|
| 2 | 2.35E+08 | -0.04636 | 0.00361 | 9.66E-38 | NA | ieu-b-4812 | rs1337113701 | G | T | 0.07939 | 25-Hydroxyvitamin D | rs2011425 | EUR | 233627608 | 235627608 |
| 2 | 2.35E+08 | -0.04674 | 0.00373 | 5.07E-36 | 417580 | ebi-a-GCST90000614 | rs2011425 | G | T | 0.077893 | 25-Hydroxyvitamin D | rs2011425 | EUR | 233627608 | 235627608 |
| 2 | 2.35E+08 | -0.04664 | 0.003672 | 5.80E-37 | 417580 | ebi-a-GCST90000615 | rs2011425 | G | T | 0.077923 | 25-Hydroxyvitamin D | rs2011425 | EUR | 233627608 | 235627608 |
| 2 | 2.35E+08 | -0.04656 | 0.003674 | 8.28E-37 | 417580 | ebi-a-GCST90000616 | rs2011425 | G | T | 0.077893 | 25-Hydroxyvitamin D | rs2011425 | EUR | 233627608 | 235627608 |
| 2 | 2.35E+08 | -0.04382 | 0.00368 | 1.09E-32 | 417580 | ebi-a-GCST90000617 | rs2011425 | G | T | 0.077923 | 25-Hydroxyvitamin D | rs2011425 | EUR | 233627608 | 235627608 |
| 2 | 2.35E+08 | -0.04783 | 0.003761 | 4.66E-37 | 496946 | ebi-a-GCST90000618 | rs2011425 | G | T | 0.079578 | 25-Hydroxyvitamin D | rs2011425 | EUR | 233627608 | 235627608 |
| 2 | 2.35E+08 | -0.04589 | 0.00456 | 8.13E-24 | 329247 | ukb-d-30890_irnt | rs2011425 | G | T | 0.079392 | 25-Hydroxyvitamin D | rs2011425 | EUR | 233627608 | 235627608 |
| 2 | 2.35E+08 | -0.99468 | 0.095686 | 2.63E-25 | 329247 | ukb-d-30890_raw | rs2011425 | G | T | 0.079392 | 25-Hydroxyvitamin D | rs2011425 | EUR | 233627608 | 235627608 |
| 2 | 2.35E+08 | -0.05149 | 0.003958 | 4.10E-39 | 418691 | ebi-a-GCST90025967 | rs2011425 | G | T | 0.079601 | 25-Hydroxyvitamin D | rs2011425 | EUR | 233627608 | 235627608 |
| 2 | 2.35E+08 | -0.0458 | 0.00419 | 8.16E-28 | 373045 | ebi-a-GCST90014016 | rs2011425 | G | T | NA | 25-Hydroxyvitamin D | rs2011425 | EUR | 233627608 | 235627608 |
| 2 | 2.35E+08 | -0.04404 | 0.003543 | 1.81E-35 | NA | ieu-b-4812 | rs3732218 | A | G | 0.081658 | 25-Hydroxyvitamin D | rs3732218 | EUR | 233627304 | 235627304 |
| 2 | 2.35E+08 | -0.04647 | 0.003639 | 2.37E-37 | 417580 | ebi-a-GCST90000614 | rs3732218 | A | G | 0.081652 | 25-Hydroxyvitamin D | rs3732218 | EUR | 233627304 | 235627304 |
| 2 | 2.35E+08 | -0.04647 | 0.003582 | 1.76E-38 | 417580 | ebi-a-GCST90000615 | rs3732218 | A | G | 0.081687 | 25-Hydroxyvitamin D | rs3732218 | EUR | 233627304 | 235627304 |

| | | | | | | | | | | | | | | | |
|---|---|---|---|---|---|---|---|---|---|---|---|---|---|---|---|
| 2 | 2.35E+08 | -0.04628 | 0.003583 | 3.67E-38 | 417580 | ebi-a-GCST90000616 | rs3732218 | A | G | 0.081652 | 25-Hydroxyvitamin D | rs3732218 | EUR | 233627304 | 235627304 |
| 2 | 2.35E+08 | -0.04374 | 0.00359 | 3.78E-34 | 417580 | ebi-a-GCST90000617 | rs3732218 | A | G | 0.081687 | 25-Hydroxyvitamin D | rs3732218 | EUR | 233627304 | 235627304 |
| 2 | 2.35E+08 | -0.04718 | 0.00368 | 1.26E-37 | 496946 | ebi-a-GCST90000618 | rs3732218 | A | G | 0.082928 | 25-Hydroxyvitamin D | rs3732218 | EUR | 233627304 | 235627304 |
| 2 | 2.35E+08 | -0.04527 | 0.004479 | 5.11E-24 | 329247 | ukb-d-30890_irnt | rs3732218 | A | G | 0.081597 | 25-Hydroxyvitamin D | rs3732218 | EUR | 233627304 | 235627304 |
| 2 | 2.35E+08 | -0.97746 | 0.093973 | 2.46E-25 | 329247 | ukb-d-30890_raw | rs3732218 | A | G | 0.081597 | 25-Hydroxyvitamin D | rs3732218 | EUR | 233627304 | 235627304 |
| 2 | 2.35E+08 | -0.04424 | 0.004114 | 5.61E-27 | 373045 | ebi-a-GCST90014016 | rs3732218 | A | G | NA | 25-Hydroxyvitamin D | rs3732218 | EUR | 233627304 | 235627304 |
| 2 | 2.35E+08 | -0.06692 | 0.002903 | 1.90E-120 | 436055 | ebi-a-GCST90025973 | rs2011425 | G | T | 0.079614 | Bilirubin Levels | rs2011425 | EUR | 233627608 | 235627608 |
| 2 | 2.35E+08 | -0.04655 | 0.006033 | 1.20E-14 | NA | ieu-b-4868 | rs1337113701 | G | T | 0.080217 | Bioavailable Testosterone | rs2011425 | EUR | 233627608 | 235627608 |
| 2 | 2.35E+08 | -0.03534 | 0.00611 | 7.30E-09 | NA | ieu-b-4869 | rs1337113701 | G | T | 0.079462 | Bioavailable Testosterone | rs2011425 | EUR | 233627608 | 235627608 |
| 2 | 2.35E+08 | -0.02569 | 0.004701 | 3.30E-08 | 188507 | ebi-a-GCST90012102 | rs2011425 | G | T | 0.07945 | Bioavailable Testosterone | rs2011425 | EUR | 233627608 | 235627608 |
| 2 | 2.35E+08 | -0.05025 | 0.005656 | 6.40E-20 | 178782 | ebi-a-GCST90012103 | rs2011425 | G | T | 0.079615 | Bioavailable Testosterone | rs2011425 | EUR | 233627608 | 235627608 |
| 2 | 2.35E+08 | -0.03766 | 0.003714 | 8.90E-25 | 382988 | ebi-a-GCST90012104 | rs2011425 | G | T | 0.079474 | Bioavailable Testosterone | rs2011425 | EUR | 233627608 | 235627608 |
| 2 | 2.35E+08 | -0.04533 | 0.005924 | 2.00E-14 | NA | ieu-b-4868 | rs3732218 | A | G | 0.08243 | Bioavailable Testosterone | rs3732218 | EUR | 233627304 | 235627304 |
| 2 | 2.35E+08 | -0.03414 | 0.006004 | 1.30E-08 | NA | ieu-b-4869 | rs3732218 | A | G | 0.081613 | Bioavailable Testosterone | rs3732218 | EUR | 233627304 | 235627304 |
| 2 | 2.35E+08 | -0.02521 | 0.004619 | 3.10E-08 | 188507 | ebi-a-GCST90012102 | rs3732218 | A | G | 0.081646 | Bioavailable Testosterone | rs3732218 | EUR | 233627304 | 235627304 |

| | | | | | | | | | | | | | | | |
|---|---|---|---|---|---|---|---|---|---|---|---|---|---|---|---|
| 2 | 2.35E+08 | -0.04819 | 0.005553 | 3.10E-19 | 178782 | ebi-a-GCST90012103 | rs3732218 | A | G | 0.081856 | Bioavailable Testosterone | rs3732218 | EUR | 233627304 | 235627304 |
| 2 | 2.35E+08 | -0.0368 | 0.003647 | 1.10E-24 | 382988 | ebi-a-GCST90012104 | rs3732218 | A | G | 0.081698 | Bioavailable Testosterone | rs3732218 | EUR | 233627304 | 235627304 |
| 2 | 2.35E+08 | -0.09581 | 0.004659 | 6.62E-94 | 292933 | ukb-d-30660_irnt | rs2011425 | G | T | 0.079128 | Direct Bilirubin | rs2011425 | EUR | 233627608 | 235627608 |
| 2 | 2.35E+08 | -0.10485 | 0.004051 | 1.48E-147 | 292933 | ukb-d-30660_raw | rs2011425 | G | T | 0.079128 | Direct Bilirubin | rs2011425 | EUR | 233627608 | 235627608 |
| 2 | 2.35E+08 | -0.2908 | 0.03446 | 3.23E-17 | NA | ukb-e-30660_AFR | rs2011425 | G | T | 0.90436 | Direct Bilirubin | rs2011425 | AFR | 233627608 | 235627608 |
| 2 | 2.35E+08 | -0.2136 | 0.02143 | 2.09E-23 | NA | ukb-e-30660_CSA | rs2011425 | G | T | 0.8081 | Direct Bilirubin | rs2011425 | Mixed | 233627608 | 235627608 |
| 2 | 2.35E+08 | -0.08554 | 0.003526 | 1.60E-134 | 372420 | ebi-a-GCST90025983 | rs2011425 | G | T | 0.079238 | Direct Bilirubin | rs2011425 | EUR | 233627608 | 235627608 |
| 2 | 2.35E+08 | -0.09642 | 0.004326 | 4.99E-110 | 332739 | ebi-a-GCST90013997 | rs2011425 | G | T | NA | Direct Bilirubin | rs2011425 | EUR | 233627608 | 235627608 |
| 2 | 2.35E+08 | -0.406 | 0.03625 | 4.07E-29 | NA | ukb-e-recode1_AFR | rs2011425 | G | T | 0.90444 | Direct Bilirubin | rs2011425 | AFR | 233627608 | 235627608 |
| 2 | 2.35E+08 | -0.252 | 0.02271 | 1.31E-28 | NA | ukb-e-recode1_CSA | rs2011425 | G | T | 0.8077 | Direct Bilirubin | rs2011425 | Mixed | 233627608 | 235627608 |
| 2 | 2.35E+08 | -0.09998 | 0.004577 | 1.05E-105 | 292933 | ukb-d-30660_irnt | rs3732218 | A | G | 0.081285 | Direct Bilirubin | rs3732218 | EUR | 233627304 | 235627304 |
| 2 | 2.35E+08 | -0.10708 | 0.003979 | 2.55E-159 | 292933 | ukb-d-30660_raw | rs3732218 | A | G | 0.081285 | Direct Bilirubin | rs3732218 | EUR | 233627304 | 235627304 |
| 2 | 2.35E+08 | -0.2583 | 0.03357 | 1.42E-14 | NA | ukb-e-30660_AFR | rs3732218 | A | G | 0.8997 | Direct Bilirubin | rs3732218 | AFR | 233627304 | 235627304 |

| | | | | | | | | | | | | | | | |
|---|---|---|---|---|---|---|---|---|---|---|---|---|---|---|---|
| 2 | 2.35E+08 | -0.2114 | 0.02135 | 3.97E-23 | NA | ukb-e-30660_CSA | rs3732218 | A | G | 0.8082 | Direct Bilirubin | rs3732218 | Mixed | 233627304 | 235627304 |
| 2 | 2.35E+08 | -0.10042 | 0.004249 | 1.69E-123 | 332739 | ebi-a-GCST90013997 | rs3732218 | A | G | NA | Direct Bilirubin | rs3732218 | EUR | 233627304 | 235627304 |
| 2 | 2.35E+08 | -0.3614 | 0.03531 | 1.37E-24 | NA | ukb-e-recode1_AFR | rs3732218 | A | G | 0.8998 | Direct Bilirubin | rs3732218 | AFR | 233627304 | 235627304 |
| 2 | 2.35E+08 | -0.2516 | 0.02263 | 1.02E-28 | NA | ukb-e-recode1_CSA | rs3732218 | A | G | 0.8079 | Direct Bilirubin | rs3732218 | Mixed | 233627304 | 235627304 |
| 2 | 2.35E+08 | -0.0253 | 0.002844 | 5.73E-19 | 312102 | ukb-d-30850_irnt | rs2011425 | G | T | 0.079206 | Testosterone | rs2011425 | EUR | 233627608 | 235627608 |
| 2 | 2.35E+08 | -0.09949 | 0.012601 | 2.91E-15 | 312102 | ukb-d-30850_raw | rs2011425 | G | T | 0.079206 | Testosterone | rs2011425 | EUR | 233627608 | 235627608 |
| 2 | 2.35E+08 | -0.02433 | 0.002793 | 3.02E-18 | 312102 | ukb-d-30850_irnt | rs3732218 | A | G | 0.081417 | Testosterone | rs3732218 | EUR | 233627304 | 235627304 |
| 2 | 2.35E+08 | -0.09791 | 0.012377 | 2.57E-15 | 312102 | ukb-d-30850_raw | rs3732218 | A | G | 0.081417 | Testosterone | rs3732218 | EUR | 233627304 | 235627304 |
| 2 | 2.35E+08 | -0.02576 | 0.002705 | 1.67E-21 | 353805 | ebi-a-GCST90014013 | rs2011425 | G | T | NA | Testosterone Levels | rs2011425 | EUR | 233627608 | 235627608 |
| 2 | 2.35E+08 | -0.02491 | 0.002655 | 6.56E-21 | 353805 | ebi-a-GCST90014013 | rs3732218 | A | G | NA | Testosterone Levels | rs3732218 | EUR | 233627304 | 235627304 |
| 2 | 2.35E+08 | 0.05749 | 0.006589 | 2.63E-18 | NA | bbj-a-53 | rs2011425 | G | T | 0.1331 | Total Bilirubin | rs2011425 | EAS | 233627608 | 235627608 |
| 2 | 2.35E+08 | -0.08307 | 0.004277 | 5.83E-84 | 342829 | ukb-d-30840_irnt | rs2011425 | G | T | 0.079476 | Total Bilirubin | rs2011425 | EUR | 233627608 | 235627608 |
| 2 | 2.35E+08 | -0.59643 | 0.019265 | 3.66E-210 | 342829 | ukb-d-30840_raw | rs2011425 | G | T | 0.079476 | Total Bilirubin | rs2011425 | EUR | 233627608 | 235627608 |
| 2 | 2.35E+08 | -0.3826 | 0.03204 | 7.32E-33 | NA | ukb-e-30840_AFR | rs2011425 | G | T | 0.8972 | Total Bilirubin | rs2011425 | AFR | 233627608 | 235627608 |

| | | | | | | | | | | | | | | | |
|---|---|---|---|---|---|---|---|---|---|---|---|---|---|---|---|
| 2 | 2.35E+08 | -0.2218 | 0.01952 | 6.63E-30 | NA | ukb-e-30840_CSA | rs2011425 | G | T | 0.802 | Total Bilirubin | rs2011425 | Mixed | 233627608 | 235627608 |
| 2 | 2.35E+08 | 0.044991 | 0.005492 | 2.60E-16 | 124341 | ebi-a-GCST90018753 | rs2011425 | G | T | 0.132742 | Total Bilirubin | rs2011425 | EUR | 233627608 | 235627608 |
| 2 | 2.35E+08 | -0.0347 | 0.0034 | 8.07E-25 | 342829 | ebi-a-GCST90018973 | rs2011425 | G | T | 0.093653 | Total Bilirubin | rs2011425 | EUR | 233627608 | 235627608 |
| 2 | 2.35E+08 | -0.08572 | 0.003929 | 1.68E-105 | 388303 | ebi-a-GCST90013872 | rs2011425 | G | T | NA | Total Bilirubin | rs2011425 | EUR | 233627608 | 235627608 |
| 2 | 2.35E+08 | -0.08603 | 0.003936 | 6.44E-106 | 388303 | ebi-a-GCST90014012 | rs2011425 | G | T | NA | Total Bilirubin | rs2011425 | EUR | 233627608 | 235627608 |
| 2 | 2.35E+08 | 0.0355 | 0.006279 | 1.58E-08 | NA | bbj-a-53 | rs3732218 | A | G | 0.1325 | Total Bilirubin | rs3732218 | EAS | 233627304 | 235627304 |
| 2 | 2.35E+08 | -0.08738 | 0.004201 | 4.74E-96 | 342829 | ukb-d-30840_irnt | rs3732218 | A | G | 0.08168 | Total Bilirubin | rs3732218 | EUR | 233627304 | 235627304 |
| 2 | 2.35E+08 | -0.61094 | 0.018918 | 1.86E-228 | 342829 | ukb-d-30840_raw | rs3732218 | A | G | 0.08168 | Total Bilirubin | rs3732218 | EUR | 233627304 | 235627304 |
| 2 | 2.35E+08 | -0.3424 | 0.03137 | 9.91E-28 | NA | ukb-e-30840_AFR | rs3732218 | A | G | 0.8932 | Total Bilirubin | rs3732218 | AFR | 233627304 | 235627304 |
| 2 | 2.35E+08 | -0.2193 | 0.01947 | 1.98E-29 | NA | ukb-e-30840_CSA | rs3732218 | A | G | 0.8026 | Total Bilirubin | rs3732218 | Mixed | 233627304 | 235627304 |
| 2 | 2.35E+08 | 0.031648 | 0.005413 | 5.00E-09 | 124341 | ebi-a-GCST90018753 | rs3732218 | A | G | 0.131559 | Total Bilirubin | rs3732218 | EUR | 233627304 | 235627304 |
| 2 | 2.35E+08 | -0.0426 | 0.0033 | 8.64E-38 | 342829 | ebi-a-GCST90018973 | rs3732218 | A | G | 0.094956 | Total Bilirubin | rs3732218 | EUR | 233627304 | 235627304 |
| 2 | 2.35E+08 | -0.08998 | 0.003858 | 2.56E-120 | 388303 | ebi-a-GCST90013872 | rs3732218 | A | G | NA | Total Bilirubin | rs3732218 | EUR | 233627304 | 235627304 |

| | | | | | | | | | | | | | | | |
|---|---|---|---|---|---|---|---|---|---|---|---|---|---|---|---|
| 2 | 2.35E+08 | -0.09024 | 0.003864 | 1.21E-120 | 388303 | ebi-a-GCST90014012 | rs3732218 | A | G | NA | Total Bilirubin | rs3732218 | EUR | 233627304 | 235627304 |
| 2 | 2.35E+08 | -0.03206 | 0.005844 | 4.10E-08 | NA | ieu-b-4864 | rs1337113701 | G | T | 0.07939 | Total Testosterone | rs2011425 | EUR | 233627608 | 235627608 |
| 2 | 2.35E+08 | -0.046 | 0.005735 | 1.00E-15 | NA | ieu-b-4865 | rs1337113701 | G | T | 0.080366 | Total Testosterone | rs2011425 | EUR | 233627608 | 235627608 |
| 2 | 2.35E+08 | -0.03349 | 0.005199 | 6.60E-11 | 230454 | ebi-a-GCST90012112 | rs2011425 | G | T | 0.079396 | Total Testosterone | rs2011425 | EUR | 233627608 | 235627608 |
| 2 | 2.35E+08 | -0.04834 | 0.005631 | 1.90E-18 | 194453 | ebi-a-GCST90012113 | rs2011425 | G | T | 0.07973 | Total Testosterone | rs2011425 | EUR | 233627608 | 235627608 |
| 2 | 2.35E+08 | -0.02498 | 0.002389 | 3.10E-26 | 425097 | ebi-a-GCST90012114 | rs2011425 | G | T | 0.079563 | Total Testosterone | rs2011425 | EUR | 233627608 | 235627608 |
| 2 | 2.35E+08 | -0.04567 | 0.00563 | 5.00E-16 | NA | ieu-b-4865 | rs3732218 | A | G | 0.082622 | Total Testosterone | rs3732218 | EUR | 233627304 | 235627304 |
| 2 | 2.35E+08 | -0.03203 | 0.005108 | 1.40E-10 | 230454 | ebi-a-GCST90012112 | rs3732218 | A | G | 0.081552 | Total Testosterone | rs3732218 | EUR | 233627304 | 235627304 |
| 2 | 2.35E+08 | -0.04713 | 0.005527 | 2.50E-18 | 194453 | ebi-a-GCST90012113 | rs3732218 | A | G | 0.082015 | Total Testosterone | rs3732218 | EUR | 233627304 | 235627304 |
| 2 | 2.35E+08 | -0.02446 | 0.002346 | 3.80E-26 | 425097 | ebi-a-GCST90012114 | rs3732218 | A | G | 0.081777 | Total Testosterone | rs3732218 | EUR | 233627304 | 235627304 |
| 2 | 2.35E+08 | -0.0532 | 0.0089 | 2.77E-09 | 7250 | met-a-513 | rs3732218 | A | G | 0.0805 | X-11440 | rs3732218 | EUR | 233627304 | 235627304 |
| 4 | 70030273 | -0.02102 | 0.001939 | 2.18E-27 | NA | ieu-b-4812 | rs7668556 | G | A | 0.529883 | 25-Hydroxyvitamin D | rs7668556 | EUR | 69030273 | 71030273 |
| 4 | 70030273 | -0.02462 | 0.002 | 8.04E-35 | 417580 | ebi-a-GCST90000614 | rs7668556 | G | A | 0.531216 | 25-Hydroxyvitamin D | rs7668556 | EUR | 69030273 | 71030273 |
| 4 | 70030273 | -0.02436 | 0.001969 | 3.92E-35 | 417580 | ebi-a-GCST90000615 | rs7668556 | G | A | 0.531156 | 25-Hydroxyvitamin D | rs7668556 | EUR | 69030273 | 71030273 |
| 4 | 70030273 | -0.02479 | 0.00197 | 2.56E-36 | 417580 | ebi-a-GCST90000616 | rs7668556 | G | A | 0.531216 | 25-Hydroxyvitamin D | rs7668556 | EUR | 69030273 | 71030273 |

| | | | | | | | | | | | | | | | |
|---|---|---|---|---|---|---|---|---|---|---|---|---|---|---|---|
| 4 | 70030273 | -0.02251 | 0.001974 | 3.88E-30 | 417580 | ebi-a-GCST90000617 | rs7668556 | G | A | 0.531156 | 25-Hydroxyvitamin D | rs7668556 | EUR | 69030273 | 71030273 |
| 4 | 70030273 | -0.02349 | 0.002042 | 1.26E-30 | 496946 | ebi-a-GCST90000618 | rs7668556 | G | A | 0.531408 | 25-Hydroxyvitamin D | rs7668556 | EUR | 69030273 | 71030273 |
| 4 | 70030273 | -0.02481 | 0.002452 | 4.61E-24 | 329247 | ukb-d-30890_irnt | rs7668556 | G | A | 0.52896 | 25-Hydroxyvitamin D | rs7668556 | EUR | 69030273 | 71030273 |
| 4 | 70030273 | -0.53574 | 0.051451 | 2.19E-25 | 329247 | ukb-d-30890_raw | rs7668556 | G | A | 0.52896 | 25-Hydroxyvitamin D | rs7668556 | EUR | 69030273 | 71030273 |
| 4 | 70030273 | -0.02466 | 0.002267 | 1.44E-27 | 373045 | ebi-a-GCST90014016 | rs7668556 | G | A | NA | 25-Hydroxyvitamin D | rs7668556 | EUR | 69030273 | 71030273 |
| 4 | 70030273 | -0.01211 | 0.002 | 8.50E-10 | 382988 | ebi-a-GCST90012104 | rs7668556 | G | A | 0.530497 | Bioavailable Testosterone | rs7668556 | EUR | 69030273 | 71030273 |
| 4 | 69669183 | 0.346195 | 0.035995 | 7.17E-22 | 5185 | ebi-a-GCST009970 | rs10000284 | C | T | 0.079997 | Cotinine and 3'-Hydroxycotinine Levels in Current Smokers | rs10000284 | EUR | 68669183 | 70669183 |
| 4 | 69659738 | 0.324022 | 0.034764 | 1.23E-20 | 5185 | ebi-a-GCST009970 | rs10023464 | T | C | 0.085986 | Cotinine and 3'-Hydroxycotinine Levels in Current Smokers | rs10023464 | EUR | 68659738 | 70659738 |
| 4 | 69680942 | 0.348836 | 0.036823 | 2.89E-21 | 5185 | ebi-a-GCST009970 | rs11725502 | T | A | 0.077045 | Cotinine and 3'-Hydroxycotinine Levels in Current Smokers | rs11725502 | EUR | 68680942 | 70680942 |
| 4 | 69682855 | -0.34433 | 0.035959 | 1.08E-21 | 5185 | ebi-a-GCST009970 | rs835309 | G | T | 0.920031 | Cotinine and 3'-Hydroxycotinine Levels in Current Smokers | rs835309 | EUR | 68682855 | 70682855 |
| 4 | 69669183 | 0.345179 | 0.03663 | 4.65E-21 | 5185 | ebi-a-GCST009965 | rs10000284 | C | T | 0.079997 | Cotinine levels in current smokers | rs10000284 | EUR | 68669183 | 70669183 |
| 4 | 69659738 | 0.319984 | 0.035451 | 1.89E-19 | 5185 | ebi-a-GCST009965 | rs10023464 | T | C | 0.085986 | Cotinine levels in current smokers | rs10023464 | EUR | 68659738 | 70659738 |
| 4 | 69680942 | 0.349734 | 0.03748 | 1.11E-20 | 5185 | ebi-a-GCST009965 | rs11725502 | T | A | 0.077045 | Cotinine levels in current smokers | rs11725502 | EUR | 68680942 | 70680942 |

| | | | | | | | | | | | | | | | |
|---|---|---|---|---|---|---|---|---|---|---|---|---|---|---|---|
| 4 | 69682855 | -0.34445 | 0.036591 | 5.11E-21 | 5185 | ebi-a-GCST009965 | rs835309 | G | T | 0.920031 | Cotinine levels in current smokers | rs835309 | EUR | 68682855 | 70682855 |
| 4 | 69669183 | -0.0261 | 0.004118 | 2.34E-10 | 292933 | ukb-d-30660_irnt | rs10000284 | C | T | 0.10247 | Direct Bilirubin | rs10000284 | EUR | 68669183 | 70669183 |
| 4 | 69669183 | -0.02332 | 0.003427 | 1.01E-11 | 332739 | ebi-a-GCST90013997 | rs10000284 | C | T | NA | Direct Bilirubin | rs10000284 | EUR | 68669183 | 70669183 |
| 4 | 69659738 | -0.02543 | 0.00411 | 6.10E-10 | 292933 | ukb-d-30660_irnt | rs10023464 | T | C | 0.10312 | Direct Bilirubin | rs10023464 | EUR | 68659738 | 70659738 |
| 4 | 69659738 | -0.02307 | 0.00342 | 1.54E-11 | 332739 | ebi-a-GCST90013997 | rs10023464 | T | C | NA | Direct Bilirubin | rs10023464 | EUR | 68659738 | 70659738 |
| 4 | 69680942 | -0.02561 | 0.004189 | 9.79E-10 | 292933 | ukb-d-30660_irnt | rs11725502 | T | A | 0.10009 | Direct Bilirubin | rs11725502 | EUR | 68680942 | 70680942 |
| 4 | 69680942 | -0.02263 | 0.003485 | 8.45E-11 | 332739 | ebi-a-GCST90013997 | rs11725502 | T | A | NA | Direct Bilirubin | rs11725502 | EUR | 68680942 | 70680942 |
| 4 | 69682855 | 0.025585 | 0.004118 | 5.23E-10 | 292933 | ukb-d-30660_irnt | rs835309 | G | T | 0.89765 | Direct Bilirubin | rs835309 | EUR | 68682855 | 70682855 |
| 4 | 69682855 | 0.022998 | 0.003427 | 1.93E-11 | 332739 | ebi-a-GCST90013997 | rs835309 | G | T | NA | Direct Bilirubin | rs835309 | EUR | 68682855 | 70682855 |
| 4 | 69669183 | 0.153862 | 0.020525 | 6.58E-14 | 3956 | eqtl-a-ENSG00000249763 | rs10000284 | C | T | 0.092262 | ENSG00000249763 | rs10000284 | EUR | 68669183 | 70669183 |
| 4 | 69659738 | 0.136709 | 0.020519 | 2.69E-11 | 3691 | eqtl-a-ENSG00000249763 | rs10023464 | T | C | 0.092413 | ENSG00000249763 | rs10023464 | EUR | 68659738 | 70659738 |
| 4 | 69680942 | 0.158225 | 0.021782 | 3.75E-13 | 3871 | eqtl-a-ENSG00000249763 | rs11725502 | T | A | 0.080937 | ENSG00000249763 | rs11725502 | EUR | 68680942 | 70680942 |
| 4 | 70030273 | -0.17388 | 0.011876 | 1.53E-48 | 10239 | eqtl-a-ENSG00000249763 | rs7668556 | G | A | 0.551021 | ENSG00000249763 | rs7668556 | EUR | 69030273 | 71030273 |

| | | | | | | | | | | | | | | | |
|---|---|---|---|---|---|---|---|---|---|---|---|---|---|---|---|
| 4 | 69682855 | -0.15593 | 0.021015 | 1.17E-13 | 3956 | eqtl-a-ENSG00000249763 | rs835309 | G | T | 0.912433 | ENSG00000249763 | rs835309 | EUR | 68682855 | 70682855 |
| 4 | 70030273 | -0.09092 | 0.011942 | 2.67E-14 | 5493 | eqtl-a-ENSG00000250696 | rs7668556 | G | A | 0.551021 | ENSG00000250696 | rs7668556 | EUR | 69030273 | 71030273 |
| 4 | 70030273 | -0.00695 | 0.000847 | 1.90E-16 | 206927 | ebi-a-GCST90012105 | rs7668556 | G | A | 0.530887 | Estradiol Levels | rs7668556 | EUR | 69030273 | 71030273 |
| 4 | 70030273 | -0.10877 | 0.012853 | 2.62E-17 | 147690 | ebi-a-GCST90020091 | rs7668556 | G | A | 0.530151 | Estradiol Levels | rs7668556 | EUR | 69030273 | 71030273 |
| 4 | 70030273 | -0.0133 | 0.002 | 1.21E-11 | 344104 | ebi-a-GCST90018954 | rs7668556 | G | A | 0.599685 | Gamma glutamyl transpeptidase | rs7668556 | EUR | 69030273 | 71030273 |
| 4 | 70030273 | -0.01216 | 0.002223 | 4.47E-08 | 344104 | ukb-d-30730_irnt | rs7668556 | G | A | 0.52935 | Gamma Glutamyltransferase Levels | rs7668556 | EUR | 69030273 | 71030273 |
| 4 | 70030273 | -0.01305 | 0.001953 | 2.37E-11 | 389672 | ebi-a-GCST90014004 | rs7668556 | G | A | NA | Gamma glutamyltransferase levels | rs7668556 | EUR | 69030273 | 71030273 |
| 4 | 69669183 | -0.02136 | 0.003885 | 3.81E-08 | 350472 | ukb-d-30050_irnt | rs10000284 | C | T | 0.102974 | Mean Corpuscular Haemoglobin | rs10000284 | EUR | 68669183 | 70669183 |
| 4 | 69680942 | -0.0216 | 0.003951 | 4.54E-08 | 350472 | ukb-d-30050_irnt | rs11725502 | T | A | 0.100603 | Mean Corpuscular Haemoglobin | rs11725502 | EUR | 68680942 | 70680942 |
| 4 | 69682855 | 0.021215 | 0.003885 | 4.73E-08 | 350472 | ukb-d-30050_irnt | rs835309 | G | T | 0.897145 | Mean Corpuscular Haemoglobin | rs835309 | EUR | 68682855 | 70682855 |
| 4 | 69669183 | -0.03341 | 0.005865 | 1.22E-08 | 172332 | ebi-a-GCST004630 | rs10000284 | C | T | 0.1032 | Mean Corpuscular Haemoglobin | rs10000284 | EUR | 68669183 | 70669183 |
| 4 | 69669183 | -0.02932 | 0.003196 | 4.76E-20 | 486823 | ebi-a-GCST90002322 | rs10000284 | C | T | 0.101608 | Mean Corpuscular Haemoglobin | rs10000284 | EUR | 68669183 | 70669183 |
| 4 | 69669183 | -0.0312 | 0.003432 | 9.80E-20 | 408112 | ebi-a-GCST90002390 | rs10000284 | C | T | 0.102406 | Mean Corpuscular Haemoglobin | rs10000284 | EUR | 68669183 | 70669183 |
| 4 | 69669183 | -0.02628 | 0.003082 | 2.90E-19 | 572863 | ebi-a-GCST90028995 | rs10000284 | C | T | 0.10228 | Mean Corpuscular Haemoglobin | rs10000284 | EUR | 68669183 | 70669183 |

| | | | | | | | | | | | | | | | |
|---|---|---|---|---|---|---|---|---|---|---|---|---|---|---|---|
| 4 | 69659738 | -0.0336 | 0.005866 | 1.02E-08 | 172332 | ebi-a-GCST004630 | rs10023464 | T | C | 0.1017 | Mean Corpuscular Haemoglobin | rs10023464 | EUR | 68659738 | 70659738 |
| 4 | 69659738 | -0.02969 | 0.003195 | 1.63E-20 | 486823 | ebi-a-GCST90002322 | rs10023464 | T | C | 0.102344 | Mean Corpuscular Haemoglobin | rs10023464 | EUR | 68659738 | 70659738 |
| 4 | 69659738 | -0.03101 | 0.003423 | 1.30E-19 | 408112 | ebi-a-GCST90002390 | rs10023464 | T | C | 0.103188 | Mean Corpuscular Haemoglobin | rs10023464 | EUR | 68659738 | 70659738 |
| 4 | 69659738 | -0.0189 | 0.0028 | 7.05E-12 | 350472 | ebi-a-GCST90018964 | rs10023464 | T | C | 0.158403 | Mean Corpuscular Haemoglobin | rs10023464 | EUR | 68659738 | 70659738 |
| 4 | 69659738 | -0.02582 | 0.003075 | 7.00E-19 | 572863 | ebi-a-GCST90028995 | rs10023464 | T | C | 0.103017 | Mean Corpuscular Haemoglobin | rs10023464 | EUR | 68659738 | 70659738 |
| 4 | 69680942 | -0.0329 | 0.005864 | 2.01E-08 | 172332 | ebi-a-GCST004630 | rs11725502 | T | A | 0.1009 | Mean Corpuscular Haemoglobin | rs11725502 | EUR | 68680942 | 70680942 |
| 4 | 69680942 | -0.02977 | 0.003248 | 5.22E-20 | 486823 | ebi-a-GCST90002322 | rs11725502 | T | A | 0.099109 | Mean Corpuscular Haemoglobin | rs11725502 | EUR | 68680942 | 70680942 |
| 4 | 69680942 | -0.03173 | 0.003492 | 1.00E-19 | 408112 | ebi-a-GCST90002390 | rs11725502 | T | A | 0.099867 | Mean Corpuscular Haemoglobin | rs11725502 | EUR | 68680942 | 70680942 |
| 4 | 69680942 | -0.0216 | 0.004 | 4.54E-08 | 350472 | ebi-a-GCST90018964 | rs11725502 | T | A | 0.100603 | Mean Corpuscular Haemoglobin | rs11725502 | EUR | 68680942 | 70680942 |
| 4 | 69680942 | -0.02658 | 0.003137 | 3.30E-19 | 572863 | ebi-a-GCST90028995 | rs11725502 | T | A | 0.099726 | Mean Corpuscular Haemoglobin | rs11725502 | EUR | 68680942 | 70680942 |
| 4 | 70030273 | 0.012219 | 0.001932 | 2.61E-10 | 486823 | ebi-a-GCST90002322 | rs7668556 | G | A | 0.534333 | Mean Corpuscular Haemoglobin | rs7668556 | EUR | 69030273 | 71030273 |
| 4 | 70030273 | 0.012318 | 0.002086 | 3.50E-09 | 408112 | ebi-a-GCST90002390 | rs7668556 | G | A | 0.531746 | Mean Corpuscular Haemoglobin | rs7668556 | EUR | 69030273 | 71030273 |
| 4 | 70030273 | 0.011565 | 0.001873 | 1.20E-09 | 572863 | ebi-a-GCST90028995 | rs7668556 | G | A | 0.53231 | Mean Corpuscular Haemoglobin | rs7668556 | EUR | 69030273 | 71030273 |
| 4 | 69682855 | 0.032665 | 0.005845 | 2.29E-08 | 172332 | ebi-a-GCST004630 | rs835309 | G | T | 0.898 | Mean Corpuscular Haemoglobin | rs835309 | EUR | 68682855 | 70682855 |
| 4 | 69682855 | 0.029438 | 0.003192 | 3.12E-20 | 486823 | ebi-a-GCST90002322 | rs835309 | G | T | 0.89842 | Mean Corpuscular Haemoglobin | rs835309 | EUR | 68682855 | 70682855 |

| | | | | | | | | | | | | | | | |
|---|---|---|---|---|---|---|---|---|---|---|---|---|---|---|---|
| 4 | 69682855 | 0.031033 | 0.00343 | 1.50E-19 | 408112 | ebi-a-GCST90002390 | rs835309 | G | T | 0.897602 | Mean Corpuscular Haemoglobin | rs835309 | EUR | 68682855 | 70682855 |
| 4 | 69682855 | 0.025957 | 0.003079 | 4.90E-19 | 572863 | ebi-a-GCST90028995 | rs835309 | G | T | 0.89764 | Mean Corpuscular Haemoglobin | rs835309 | EUR | 68682855 | 70682855 |
| 4 | 69669183 | -0.02619 | 0.003021 | 4.53E-18 | 544127 | ebi-a-GCST90002334 | rs10000284 | C | T | 0.101832 | Mean Corpuscular Volume | rs10000284 | EUR | 68669183 | 70669183 |
| 4 | 69669183 | -0.02746 | 0.003396 | 6.10E-16 | 408112 | ebi-a-GCST90002392 | rs10000284 | C | T | 0.102408 | Mean Corpuscular Volume | rs10000284 | EUR | 68669183 | 70669183 |
| 4 | 69669183 | -0.02048 | 0.003352 | 1.00E-09 | 396624 | ebi-a-GCST90013979 | rs10000284 | C | T | NA | Mean Corpuscular Volume | rs10000284 | EUR | 68669183 | 70669183 |
| 4 | 69659738 | -0.02628 | 0.003021 | 3.52E-18 | 544127 | ebi-a-GCST90002334 | rs10023464 | T | C | 0.101803 | Mean Corpuscular Volume | rs10023464 | EUR | 68659738 | 70659738 |
| 4 | 69659738 | -0.02725 | 0.003388 | 8.70E-16 | 408112 | ebi-a-GCST90002392 | rs10023464 | T | C | 0.10319 | Mean Corpuscular Volume | rs10023464 | EUR | 68659738 | 70659738 |
| 4 | 69659738 | -0.02007 | 0.003345 | 1.98E-09 | 396624 | ebi-a-GCST90013979 | rs10023464 | T | C | NA | Mean Corpuscular Volume | rs10023464 | EUR | 68659738 | 70659738 |
| 4 | 69680942 | -0.02663 | 0.003068 | 4.15E-18 | 544127 | ebi-a-GCST90002334 | rs11725502 | T | A | 0.098463 | Mean Corpuscular Volume | rs11725502 | EUR | 68680942 | 70680942 |
| 4 | 69680942 | -0.02802 | 0.003456 | 5.20E-16 | 408112 | ebi-a-GCST90002392 | rs11725502 | T | A | 0.099869 | Mean Corpuscular Volume | rs11725502 | EUR | 68680942 | 70680942 |
| 4 | 69680942 | -0.0205 | 0.003408 | 1.80E-09 | 396624 | ebi-a-GCST90013979 | rs11725502 | T | A | NA | Mean Corpuscular Volume | rs11725502 | EUR | 68680942 | 70680942 |
| 4 | 70030273 | 0.013142 | 0.001822 | 5.71E-13 | 544127 | ebi-a-GCST90002334 | rs7668556 | G | A | 0.53683 | Mean Corpuscular Volume | rs7668556 | EUR | 69030273 | 71030273 |
| 4 | 70030273 | 0.011838 | 0.002064 | 9.80E-09 | 408112 | ebi-a-GCST90002392 | rs7668556 | G | A | 0.531735 | Mean Corpuscular Volume | rs7668556 | EUR | 69030273 | 71030273 |
| 4 | 69682855 | 0.026109 | 0.003017 | 5.22E-18 | 544127 | ebi-a-GCST90002334 | rs835309 | G | T | 0.898518 | Mean Corpuscular Volume | rs835309 | EUR | 68682855 | 70682855 |
| 4 | 69682855 | 0.027144 | 0.003394 | 1.30E-15 | 408112 | ebi-a-GCST90002392 | rs835309 | G | T | 0.897599 | Mean Corpuscular Volume | rs835309 | EUR | 68682855 | 70682855 |

| | | | | | | | | | | | | | | | |
|---|---|---|---|---|---|---|---|---|---|---|---|---|---|---|---|
| 4 | 69682855 | 0.020236 | 0.003352 | 1.57E-09 | 396624 | ebi-a-GCST90013979 | rs835309 | G | T | NA | Mean Corpuscular Volume | rs835309 | EUR | 68682855 | 70682855 |
| 4 | 69669183 | 0.023776 | 0.003901 | 1.10E-09 | 350473 | ukb-d-30070_irnt | rs10000284 | C | T | 0.102973 | Red Cell Distribution Width | rs10000284 | EUR | 68669183 | 70669183 |
| 4 | 69669183 | 0.022991 | 0.003129 | 2.11E-13 | 531774 | ebi-a-GCST90002369 | rs10000284 | C | T | 0.102247 | Red Cell Distribution Width | rs10000284 | EUR | 68669183 | 70669183 |
| 4 | 69669183 | 0.026987 | 0.003512 | 1.50E-14 | 408112 | ebi-a-GCST90002404 | rs10000284 | C | T | 0.102439 | Red Cell Distribution Width | rs10000284 | EUR | 68669183 | 70669183 |
| 4 | 69669183 | 0.025041 | 0.003247 | 7.90E-17 | 514929 | ebi-a-GCST90029001 | rs10000284 | C | T | 0.102311 | Red Cell Distribution Width | rs10000284 | EUR | 68669183 | 70669183 |
| 4 | 69659738 | 0.023944 | 0.003894 | 7.79E-10 | 350473 | ukb-d-30070_irnt | rs10023464 | T | C | 0.103608 | Red Cell Distribution Width | rs10023464 | EUR | 68659738 | 70659738 |
| 4 | 69659738 | 0.023714 | 0.003129 | 3.61E-14 | 531774 | ebi-a-GCST90002369 | rs10023464 | T | C | 0.102114 | Red Cell Distribution Width | rs10023464 | EUR | 68659738 | 70659738 |
| 4 | 69659738 | 0.027197 | 0.003504 | 8.30E-15 | 408112 | ebi-a-GCST90002404 | rs10023464 | T | C | 0.103223 | Red Cell Distribution Width | rs10023464 | EUR | 68659738 | 70659738 |
| 4 | 69659738 | 0.025113 | 0.003239 | 6.00E-17 | 514929 | ebi-a-GCST90029001 | rs10023464 | T | C | 0.103047 | Red Cell Distribution Width | rs10023464 | EUR | 68659738 | 70659738 |
| 4 | 69680942 | 0.024419 | 0.003967 | 7.51E-10 | 350473 | ukb-d-30070_irnt | rs11725502 | T | A | 0.100602 | Red Cell Distribution Width | rs11725502 | EUR | 68680942 | 70680942 |
| 4 | 69680942 | 0.023786 | 0.003178 | 7.44E-14 | 531774 | ebi-a-GCST90002369 | rs11725502 | T | A | 0.098866 | Red Cell Distribution Width | rs11725502 | EUR | 68680942 | 70680942 |
| 4 | 69680942 | 0.027556 | 0.003574 | 1.30E-14 | 408112 | ebi-a-GCST90002404 | rs11725502 | T | A | 0.099898 | Red Cell Distribution Width | rs11725502 | EUR | 68680942 | 70680942 |
| 4 | 69680942 | 0.025718 | 0.003304 | 6.60E-17 | 514929 | ebi-a-GCST90029001 | rs11725502 | T | A | 0.099754 | Red Cell Distribution Width | rs11725502 | EUR | 68680942 | 70680942 |
| 4 | 69682855 | -0.02426 | 0.003901 | 5.02E-10 | 350473 | ukb-d-30070_irnt | rs835309 | G | T | 0.897145 | Red Cell Distribution Width | rs835309 | EUR | 68682855 | 70682855 |
| 4 | 69682855 | -0.02345 | 0.003125 | 6.45E-14 | 531774 | ebi-a-GCST90002369 | rs835309 | G | T | 0.898116 | Red Cell Distribution Width | rs835309 | EUR | 68682855 | 70682855 |

| | | | | | | | | | | | | | | | |
|---|---|---|---|---|---|---|---|---|---|---|---|---|---|---|---|
| 4 | 69682855 | -0.02755 | 0.00351 | 4.20E-15 | 408112 | ebi-a-GCST90002404 | rs835309 | G | T | 0.897568 | Red Cell Distribution Width | rs835309 | EUR | 68682855 | 70682855 |
| 4 | 69682855 | -0.02569 | 0.003244 | 1.60E-17 | 514929 | ebi-a-GCST90029001 | rs835309 | G | T | 0.897611 | Red Cell Distribution Width | rs835309 | EUR | 68682855 | 70682855 |
| 4 | 69669183 | 0.30326 | 0.038037 | 1.63E-15 | 4772 | ebi-a-GCST009966 | rs10000284 | C | T | 0.079481 | Smoking Intensity (Cotinine Levels/Cigarettes per Day) | rs10000284 | EUR | 68669183 | 70669183 |
| 4 | 69659738 | 0.280343 | 0.036714 | 2.35E-14 | 4772 | ebi-a-GCST009966 | rs10023464 | T | C | 0.085454 | Smoking intensity (cotinine levels/cigarettes per day) | rs10023464 | EUR | 68659738 | 70659738 |
| 4 | 69680942 | 0.301806 | 0.038948 | 9.71E-15 | 4772 | ebi-a-GCST009966 | rs11725502 | T | A | 0.076431 | Smoking intensity (cotinine levels/cigarettes per day) | rs11725502 | EUR | 68680942 | 70680942 |
| 4 | 69682855 | -0.30292 | 0.038032 | 1.74E-15 | 4772 | ebi-a-GCST009966 | rs835309 | G | T | 0.920698 | Smoking intensity (cotinine levels/cigarettes per day) | rs835309 | EUR | 68682855 | 70682855 |
| 4 | 70030273 | -0.01168 | 0.001528 | 2.10E-14 | 312102 | ukb-d-30850_irnt | rs7668556 | G | A | 0.52886 | Testosterone | rs7668556 | EUR | 69030273 | 71030273 |
| 4 | 70030273 | -0.03816 | 0.00677 | 1.75E-08 | 312102 | ukb-d-30850_raw | rs7668556 | G | A | 0.52886 | Testosterone | rs7668556 | EUR | 69030273 | 71030273 |
| 4 | 70030273 | -0.01124 | 0.00145 | 9.13E-15 | 353805 | ebi-a-GCST90014013 | rs7668556 | G | A | NA | Testosterone Levels | rs7668556 | EUR | 69030273 | 71030273 |
| 4 | 69669183 | -0.02741 | 0.00378 | 4.13E-13 | 342829 | ukb-d-30840_irnt | rs10000284 | C | T | 0.10301 | Total Bilirubin | rs10000284 | EUR | 68669183 | 70669183 |
| 4 | 69669183 | -0.11192 | 0.017039 | 5.09E-11 | 342829 | ukb-d-30840_raw | rs10000284 | C | T | 0.10301 | Total Bilirubin | rs10000284 | EUR | 68669183 | 70669183 |
| 4 | 69669183 | -0.0263 | 0.0033 | 3.98E-15 | 342829 | ebi-a-GCST90018973 | rs10000284 | C | T | 0.095971 | Total Bilirubin | rs10000284 | EUR | 68669183 | 70669183 |
| 4 | 69669183 | -0.02484 | 0.002933 | 2.41E-17 | 388303 | ebi-a-GCST90013872 | rs10000284 | C | T | NA | Total Bilirubin | rs10000284 | EUR | 68669183 | 70669183 |
| 4 | 69669183 | -0.0248 | 0.002953 | 4.59E-17 | 388303 | ebi-a-GCST90014012 | rs10000284 | C | T | NA | Total Bilirubin | rs10000284 | EUR | 68669183 | 70669183 |
| 4 | 69659738 | -0.02673 | 0.003773 | 1.39E-12 | 342829 | ukb-d-30840_irnt | rs10023464 | T | C | 0.10364 | Total Bilirubin | rs10023464 | EUR | 68659738 | 70659738 |

| | | | | | | | | | | | | | | | |
|---|---|---|---|---|---|---|---|---|---|---|---|---|---|---|---|
| 4 | 69659738 | -0.10976 | 0.017007 | 1.09E-10 | 342829 | ukb-d-30840_raw | rs10023464 | T | C | 0.10364 | Total Bilirubin | rs10023464 | EUR | 68659738 | 70659738 |
| 4 | 69659738 | -0.0197 | 0.0028 | 1.32E-12 | 342829 | ebi-a-GCST90018973 | rs10023464 | T | C | 0.158256 | Total Bilirubin | rs10023464 | EUR | 68659738 | 70659738 |
| 4 | 69659738 | -0.02455 | 0.002927 | 4.96E-17 | 388303 | ebi-a-GCST90013872 | rs10023464 | T | C | NA | Total Bilirubin | rs10023464 | EUR | 68659738 | 70659738 |
| 4 | 69659738 | -0.0245 | 0.002947 | 9.49E-17 | 388303 | ebi-a-GCST90014012 | rs10023464 | T | C | NA | Total Bilirubin | rs10023464 | EUR | 68659738 | 70659738 |
| 4 | 69680942 | -0.02773 | 0.003844 | 5.44E-13 | 342829 | ukb-d-30840_irnt | rs11725502 | T | A | 0.10064 | Total Bilirubin | rs11725502 | EUR | 68680942 | 70680942 |
| 4 | 69680942 | -0.11124 | 0.017328 | 1.37E-10 | 342829 | ukb-d-30840_raw | rs11725502 | T | A | 0.10064 | Total Bilirubin | rs11725502 | EUR | 68680942 | 70680942 |
| 4 | 69680942 | -0.0277 | 0.0038 | 5.43E-13 | 342829 | ebi-a-GCST90018973 | rs11725502 | T | A | 0.10064 | Total Bilirubin | rs11725502 | EUR | 68680942 | 70680942 |
| 4 | 69680942 | -0.02495 | 0.002982 | 5.96E-17 | 388303 | ebi-a-GCST90013872 | rs11725502 | T | A | NA | Total Bilirubin | rs11725502 | EUR | 68680942 | 70680942 |
| 4 | 69680942 | -0.02488 | 0.003003 | 1.17E-16 | 388303 | ebi-a-GCST90014012 | rs11725502 | T | A | NA | Total Bilirubin | rs11725502 | EUR | 68680942 | 70680942 |
| 4 | 69682855 | 0.02709 | 0.00378 | 7.69E-13 | 342829 | ukb-d-30840_irnt | rs835309 | G | T | 0.8971 | Total Bilirubin | rs835309 | EUR | 68682855 | 70682855 |
| 4 | 69682855 | 0.10966 | 0.017039 | 1.23E-10 | 342829 | ukb-d-30840_raw | rs835309 | G | T | 0.8971 | Total Bilirubin | rs835309 | EUR | 68682855 | 70682855 |
| 4 | 69682855 | 0.0259 | 0.0033 | 7.85E-15 | 342829 | ebi-a-GCST90018973 | rs835309 | G | T | 0.90422 | Total Bilirubin | rs835309 | EUR | 68682855 | 70682855 |
| 4 | 69682855 | 0.024619 | 0.002933 | 4.68E-17 | 388303 | ebi-a-GCST90013872 | rs835309 | G | T | NA | Total Bilirubin | rs835309 | EUR | 68682855 | 70682855 |
| 4 | 69682855 | 0.02457 | 0.002953 | 8.81E-17 | 388303 | ebi-a-GCST90014012 | rs835309 | G | T | NA | Total Bilirubin | rs835309 | EUR | 68682855 | 70682855 |
| 4 | 70030273 | -0.02097 | 0.003148 | 2.70E-11 | NA | ieu-b-4864 | rs7668556 | G | A | 0.530847 | Total Testosterone | rs7668556 | EUR | 69030273 | 71030273 |

| | | | | | | | | | | | | | | | |
|---|---|---|---|---|---|---|---|---|---|---|---|---|---|---|---|
| 4 | 70030273 | -0.0183 | 0.003099 | 3.50E-09 | NA | ieu-b-4865 | rs7668556 | G | A | 0.533614 | Total Testosterone | rs7668556 | EUR | 69030273 | 71030273 |
| 4 | 70030273 | -0.02134 | 0.002808 | 5.40E-14 | 230454 | ebi-a-GCST90012112 | rs7668556 | G | A | 0.530193 | Total Testosterone | rs7668556 | EUR | 69030273 | 71030273 |
| 4 | 70030273 | -0.01981 | 0.003037 | 6.00E-11 | 194453 | ebi-a-GCST90012113 | rs7668556 | G | A | 0.531311 | Total Testosterone | rs7668556 | EUR | 69030273 | 71030273 |
| 4 | 70030273 | -0.01176 | 0.001288 | 1.50E-19 | 425097 | ebi-a-GCST90012114 | rs7668556 | G | A | 0.530689 | Total Testosterone | rs7668556 | EUR | 69030273 | 71030273 |
| 10 | 96547463 | 0.018622 | 0.002782 | 2.18E-11 | 389883 | ebi-a-GCST90013991 | rs12767583 | T | C | NA | Alkaline Phosphatase Levels | rs12767583 | EUR | 95547463 | 97547463 |
| 10 | 96484777 | 0.018062 | 0.00275 | 5.10E-11 | 389883 | ebi-a-GCST90013991 | rs1926711 | A | G | NA | Alkaline Phosphatase Levels | rs1926711 | EUR | 95484777 | 97484777 |
| 10 | 96547463 | -0.0164 | 0.0027 | 5.96E-10 | 450243 | ebi-a-GCST90000025 | rs12767583 | T | C | 0.1492 | Appendicular Lean Mass | rs12767583 | EUR | 95547463 | 97547463 |
| 10 | 96484777 | -0.0149 | 0.0026 | 1.41E-08 | 450243 | ebi-a-GCST90000025 | rs1926711 | A | G | 0.1534 | Appendicular Lean Mass | rs1926711 | EUR | 95484777 | 97484777 |
| 10 | 96547463 | -0.0214 | 0.002556 | 5.20E-18 | 436055 | ebi-a-GCST90025973 | rs12767583 | T | C | 0.14912 | Bilirubin Levels | rs12767583 | EUR | 95547463 | 97547463 |
| 10 | 96547463 | -0.01931 | 0.003512 | 3.83E-08 | 292933 | ukb-d-30660_irnt | rs12767583 | T | C | 0.14876 | Direct Bilirubin | rs12767583 | EUR | 95547463 | 97547463 |
| 10 | 96547463 | -0.01808 | 0.002949 | 1.70E-11 | 372420 | ebi-a-GCST90025983 | rs12767583 | T | C | 0.148524 | Direct Bilirubin | rs12767583 | EUR | 95547463 | 97547463 |
| 10 | 96547463 | -0.0177 | 0.002925 | 1.45E-09 | 332739 | ebi-a-GCST90013997 | rs12767583 | T | C | NA | Direct Bilirubin | rs12767583 | EUR | 95547463 | 97547463 |
| 10 | 96484777 | -0.01989 | 0.003471 | 1.01E-08 | 292933 | ukb-d-30660_irnt | rs1926711 | A | G | 0.15305 | Direct Bilirubin | rs1926711 | EUR | 95484777 | 97484777 |
| 10 | 96484777 | -0.01791 | 0.002891 | 5.87E-10 | 332739 | ebi-a-GCST90013997 | rs1926711 | A | G | NA | Direct Bilirubin | rs1926711 | EUR | 95484777 | 97484777 |
| 10 | 96547463 | -0.12693 | 0.016114 | 3.35E-15 | 9188 | eqtl-a-ENSG00000108239 | rs12767583 | T | C | 0.162097 | ENSG00000108239 | rs12767583 | EUR | 95547463 | 97547463 |

| | | | | | | | | | | | | | | | |
|---|---|---|---|---|---|---|---|---|---|---|---|---|---|---|---|
| 10 | 96484777 | -0.12689 | 0.015937 | 1.70E-15 | 9188 | eqtl-a-ENSG00000108239 | rs1926711 | A | G | 0.166598 | ENSG00000108239 | rs1926711 | EUR | 95484777 | 97484777 |
| 10 | 96547463 | 0.276319 | 0.015981 | 5.58E-67 | 31086 | eqtl-a-ENSG00000119969 | rs12767583 | T | C | 0.162097 | ENSG00000119969 | rs12767583 | EUR | 95547463 | 97547463 |
| 10 | 96484777 | 0.27673 | 0.015802 | 1.16E-68 | 31470 | eqtl-a-ENSG00000119969 | rs1926711 | A | G | 0.166598 | ENSG00000119969 | rs1926711 | EUR | 95484777 | 97484777 |
| 10 | 96547463 | -0.19527 | 0.016066 | 5.42E-34 | 31086 | eqtl-a-ENSG00000173145 | rs12767583 | T | C | 0.162097 | ENSG00000173145 | rs12767583 | EUR | 95547463 | 97547463 |
| 10 | 96484777 | -0.18419 | 0.015897 | 4.85E-31 | 31470 | eqtl-a-ENSG00000173145 | rs1926711 | A | G | 0.166598 | ENSG00000173145 | rs1926711 | EUR | 95484777 | 97484777 |
| 10 | 96547463 | 0.019709 | 0.002755 | 1.50E-13 | 437651 | ebi-a-GCST90025966 | rs12767583 | T | C | 0.149132 | Gamma Glutamyl Transferase Levels | rs12767583 | EUR | 95547463 | 97547463 |
| 10 | 96547463 | 0.0143 | 0.0024 | 2.39E-09 | 344104 | ebi-a-GCST90018954 | rs12767583 | T | C | 0.192329 | Gamma glutamyl transpeptidase | rs12767583 | EUR | 95547463 | 97547463 |
| 10 | 96484777 | 0.0145 | 0.0023 | 3.08E-10 | 344104 | ebi-a-GCST90018954 | rs1926711 | A | G | 0.228646 | Gamma glutamyl transpeptidase | rs1926711 | EUR | 95484777 | 97484777 |
| 10 | 96547463 | 0.016736 | 0.002737 | 9.65E-10 | 389672 | ebi-a-GCST90014004 | rs12767583 | T | C | NA | Gamma Glutamyltransferase Levels | rs12767583 | EUR | 95547463 | 97547463 |
| 10 | 96484777 | 0.015472 | 0.002705 | 1.07E-08 | 389672 | ebi-a-GCST90014004 | rs1926711 | A | G | NA | Gamma Glutamyltransferase Levels | rs1926711 | EUR | 95484777 | 97484777 |
| 10 | 96547463 | 0.018993 | 0.003223 | 3.78E-09 | 344182 | ukb-d-30750_irnt | rs12767583 | T | C | 0.14941 | Glycated Haemoglobin | rs12767583 | EUR | 95547463 | 97547463 |
| 10 | 96484777 | 0.01937 | 0.003186 | 1.21E-09 | 344182 | ukb-d-30750_irnt | rs1926711 | A | G | 0.15362 | Glycated Haemoglobin | rs1926711 | EUR | 95484777 | 97484777 |

| | | | | | | | | | | | | | | | |
|---|---|---|---|---|---|---|---|---|---|---|---|---|---|---|---|
| 10 | 96484777 | 0.11688 | 0.021257 | 3.83E-08 | 344182 | ukb-d-30750_raw | rs1926711 | A | G | 0.15362 | Glycated Haemoglobin | rs1926711 | EUR | 95484777 | 97484777 |
| 10 | 96547463 | 0.017493 | 0.002858 | 9.34E-10 | 389889 | ebi-a-GCST90014006 | rs12767583 | T | C | NA | Glycated Haemoglobin HbA1c Levels | rs12767583 | EUR | 95547463 | 97547463 |
| 10 | 96484777 | 0.017766 | 0.002825 | 3.20E-10 | 389889 | ebi-a-GCST90014006 | rs1926711 | A | G | NA | Glycated Haemoglobin HbA1c Levels | rs1926711 | EUR | 95484777 | 97484777 |
| 10 | 96547463 | 0.017312 | 0.002719 | 3.40E-11 | 437749 | ebi-a-GCST90025974 | rs12767583 | T | C | 0.149057 | Hemoglobin A1c Levels | rs12767583 | EUR | 95547463 | 97547463 |
| 10 | 96547463 | 0.014044 | 0.002549 | 3.60E-08 | 454857 | ukb-b-14068 | rs12767583 | T | C | 0.149125 | Impedance of Leg (Left) | rs12767583 | EUR | 95547463 | 97547463 |
| 10 | 96547463 | 0.014012 | 0.002528 | 3.00E-08 | 454863 | ukb-b-7376 | rs12767583 | T | C | 0.149124 | Impedance of Leg (Left) | rs12767583 | EUR | 95547463 | 97547463 |
| 10 | 96547463 | 0.014696 | 0.00254 | 7.44E-09 | 544127 | ebi-a-GCST90002334 | rs12767583 | T | C | 0.149218 | Mean Corpuscular Volume | rs12767583 | EUR | 95547463 | 97547463 |
| 10 | 96547463 | 0.013862 | 0.002595 | 6.30E-09 | 444035 | ebi-a-GCST90025963 | rs12767583 | T | C | 0.149074 | Mean Corpuscular Volume | rs12767583 | EUR | 95547463 | 97547463 |
| 10 | 96484777 | 0.01464 | 0.002515 | 6.02E-09 | 544127 | ebi-a-GCST90002334 | rs1926711 | A | G | 0.153611 | Mean Corpuscular Volume | rs1926711 | EUR | 95484777 | 97484777 |
| 10 | 96547463 | 0.024477 | 0.002989 | 2.60E-16 | 408112 | ebi-a-GCST90002396 | rs12767583 | T | C | 0.149413 | Mean Reticulocyte Volume | rs12767583 | EUR | 95547463 | 97547463 |
| 10 | 96547463 | 0.022653 | 0.002742 | 1.30E-17 | 436944 | ebi-a-GCST90025975 | rs12767583 | T | C | 0.14897 | Mean Reticulocyte Volume | rs12767583 | EUR | 95547463 | 97547463 |
| 10 | 96547463 | 0.021717 | 0.003355 | 9.66E-11 | 344728 | ukb-d-30260_irnt | rs12767583 | T | C | 0.149202 | Mean Reticulocyte Volume | rs12767583 | EUR | 95547463 | 97547463 |
| 10 | 96484777 | 0.026828 | 0.002955 | 1.10E-19 | 408112 | ebi-a-GCST90002396 | rs1926711 | A | G | 0.153669 | Mean Reticulocyte Volume | rs1926711 | EUR | 95484777 | 97484777 |
| 10 | 96484777 | 0.023564 | 0.003317 | 1.21E-12 | 344728 | ukb-d-30260_irnt | rs1926711 | A | G | 0.153446 | Mean Reticulocyte Volume | rs1926711 | EUR | 95484777 | 97484777 |
| 10 | 96547463 | 0.016185 | 0.002653 | 1.30E-10 | 437736 | ebi-a-GCST90025976 | rs12767583 | T | C | 0.148929 | Mean Reticulocyte Volume | rs12767583 | EUR | 95547463 | 97547463 |

| | | | | | | | | | | | | | | | |
|---|---|---|---|---|---|---|---|---|---|---|---|---|---|---|---|
| 10 | 96547463 | 0.016445 | 0.002749 | 2.70E-10 | 535537 | ebi-a-GCST90028997 | rs12767583 | T | C | 0.149023 | Mean Reticulocyte Volume | rs12767583 | EUR | 95547463 | 97547463 |
| 10 | 96484777 | 0.016344 | 0.002717 | 2.00E-10 | 535537 | ebi-a-GCST90028997 | rs1926711 | A | G | 0.153285 | Mean Reticulocyte Volume | rs1926711 | EUR | 95484777 | 97484777 |
| 10 | 96547463 | 0.016208 | 0.002912 | 2.60E-08 | 408112 | ebi-a-GCST90002397 | rs12767583 | T | C | 0.149354 | Mean spheric corpuscular volume | rs12767583 | EUR | 95547463 | 97547463 |
| 10 | 96484777 | 0.016322 | 0.002878 | 1.40E-08 | 408112 | ebi-a-GCST90002397 | rs1926711 | A | G | 0.153625 | Mean spheric corpuscular volume | rs1926711 | EUR | 95484777 | 97484777 |
| 10 | 96547463 | -0.01501 | 0.002588 | 6.85E-09 | 545203 | ebi-a-GCST90002363 | rs12767583 | T | C | 0.149199 | Red Blood Cell Count | rs12767583 | EUR | 95547463 | 97547463 |
| 10 | 96547463 | -0.01417 | 0.002682 | 6.30E-09 | 445305 | ebi-a-GCST90025964 | rs12767583 | T | C | 0.149097 | Red Blood Cell Count | rs12767583 | EUR | 95547463 | 97547463 |
| 10 | 96547463 | -0.01417 | 0.002704 | 7.60E-09 | 526540 | ebi-a-GCST90029002 | rs12767583 | T | C | 0.14917 | Red Blood Cell Count | rs12767583 | EUR | 95547463 | 97547463 |
| 10 | 96484777 | -0.01495 | 0.002562 | 5.58E-09 | 545203 | ebi-a-GCST90002363 | rs1926711 | A | G | 0.153583 | Red Blood Cell Count | rs1926711 | EUR | 95484777 | 97484777 |
| 10 | 96484777 | -0.01401 | 0.002673 | 8.90E-09 | 526540 | ebi-a-GCST90029002 | rs1926711 | A | G | 0.153435 | Red Blood Cell Count | rs1926711 | EUR | 95484777 | 97484777 |
| 10 | 96547463 | 0.019774 | 0.003326 | 2.77E-09 | 350473 | ukb-d-30070_irnt | rs12767583 | T | C | 0.149357 | Red Cell Distribution Width | rs12767583 | EUR | 95547463 | 97547463 |
| 10 | 96547463 | 0.016147 | 0.002636 | 9.26E-10 | 531774 | ebi-a-GCST90002369 | rs12767583 | T | C | 0.149166 | Red Cell Distribution Width | rs12767583 | EUR | 95547463 | 97547463 |
| 10 | 96547463 | 0.017473 | 0.002984 | 4.80E-09 | 408112 | ebi-a-GCST90002404 | rs12767583 | T | C | 0.149504 | Red Cell Distribution Width | rs12767583 | EUR | 95547463 | 97547463 |
| 10 | 96547463 | 0.017817 | 0.002762 | 2.50E-11 | 441263 | ebi-a-GCST90025988 | rs12767583 | T | C | 0.149079 | Red Cell Distribution Width | rs12767583 | EUR | 95547463 | 97547463 |
| 10 | 96547463 | 0.018651 | 0.002758 | 2.50E-12 | 514929 | ebi-a-GCST90029001 | rs12767583 | T | C | 0.149189 | Red Cell Distribution Width | rs12767583 | EUR | 95547463 | 97547463 |
| 10 | 96484777 | 0.0232 | 0.003288 | 1.72E-12 | 350473 | ukb-d-30070_irnt | rs1926711 | A | G | 0.153608 | Red Cell Distribution Width | rs1926711 | EUR | 95484777 | 97484777 |

| | | | | | | | | | | | | | | | |
|---|---|---|---|---|---|---|---|---|---|---|---|---|---|---|---|
| 10 | 96484777 | 0.019072 | 0.002607 | 2.69E-13 | 531774 | ebi-a-GCST90002369 | rs1926711 | A | G | 0.153544 | Red Cell Distribution Width | rs1926711 | EUR | 95484777 | 97484777 |
| 10 | 96484777 | 0.02041 | 0.00295 | 4.60E-12 | 408112 | ebi-a-GCST90002404 | rs1926711 | A | G | 0.153769 | Red Cell Distribution Width | rs1926711 | EUR | 95484777 | 97484777 |
| 10 | 96484777 | 0.021299 | 0.002726 | 3.90E-16 | 514929 | ebi-a-GCST90029001 | rs1926711 | A | G | 0.153449 | Red Cell Distribution Width | rs1926711 | EUR | 95484777 | 97484777 |
| 10 | 96547463 | 0.016849 | 0.002657 | 2.20E-12 | 437896 | ebi-a-GCST90025947 | rs12767583 | T | C | 0.149152 | Serum Alkaline Phosphatase Levels | rs12767583 | EUR | 95547463 | 97547463 |
| 10 | 96547463 | -0.01854 | 0.002769 | 2.20E-11 | 472174 | ieu-b-4879 | rs12767583 | T | C | 0.154478 | Telomere Length | rs12767583 | EUR | 95547463 | 97547463 |
| 10 | 96484777 | -0.0178 | 0.002726 | 6.60E-11 | 472174 | ieu-b-4879 | rs758646836 | A | G | 0.161331 | Telomere Length | rs1926711 | EUR | 95484777 | 97484777 |
| 10 | 96547463 | -0.02424 | 0.003222 | 5.31E-14 | 342829 | ukb-d-30840_irnt | rs12767583 | T | C | 0.14945 | Total Bilirubin | rs12767583 | EUR | 95547463 | 97547463 |
| 10 | 96547463 | -0.09508 | 0.014523 | 5.89E-11 | 342829 | ukb-d-30840_raw | rs12767583 | T | C | 0.14945 | Total Bilirubin | rs12767583 | EUR | 95547463 | 97547463 |
| 10 | 96547463 | -0.0185 | 0.0025 | 3.19E-13 | 342829 | ebi-a-GCST90018973 | rs12767583 | T | C | 0.190246 | Total Bilirubin | rs12767583 | EUR | 95547463 | 97547463 |
| 10 | 96547463 | -0.01986 | 0.002505 | 2.23E-15 | 388303 | ebi-a-GCST90013872 | rs12767583 | T | C | NA | Total Bilirubin | rs12767583 | EUR | 95547463 | 97547463 |
| 10 | 96547463 | -0.01964 | 0.002522 | 6.96E-15 | 388303 | ebi-a-GCST90014012 | rs12767583 | T | C | NA | Total Bilirubin | rs12767583 | EUR | 95547463 | 97547463 |
| 10 | 96484777 | -0.02366 | 0.003185 | 1.12E-13 | 342829 | ukb-d-30840_irnt | rs1926711 | A | G | 0.15368 | Total Bilirubin | rs1926711 | EUR | 95484777 | 97484777 |
| 10 | 96484777 | -0.09627 | 0.014358 | 2.02E-11 | 342829 | ukb-d-30840_raw | rs1926711 | A | G | 0.15368 | Total Bilirubin | rs1926711 | EUR | 95484777 | 97484777 |
| 10 | 96484777 | -0.0185 | 0.0024 | 4.35E-14 | 342829 | ebi-a-GCST90018973 | rs1926711 | A | G | 0.225109 | Total Bilirubin | rs1926711 | EUR | 95484777 | 97484777 |
| 10 | 96484777 | -0.01937 | 0.002476 | 5.17E-15 | 388303 | ebi-a-GCST90013872 | rs1926711 | A | G | NA | Total Bilirubin | rs1926711 | EUR | 95484777 | 97484777 |

| | | | | | | | | | | | | | | | |
|---|---|---|---|---|---|---|---|---|---|---|---|---|---|---|---|
| 10 | 96484777 | -0.01913 | 0.002493 | 1.66E-14 | 388303 | ebi-a-GCST90014012 | rs1926711 | A | G | NA | Total Bilirubin | rs1926711 | EUR | 95484777 | 97484777 |
| 10 | 96547463 | 0.0406 | 0.0062 | 4.34E-11 | 6486 | met-a-725 | rs12767583 | T | C | 0.1543 | X-14473 | rs12767583 | EUR | 95547463 | 97547463 |
| 15 | 75027880 | 0.013988 | 0.00208 | 1.40E-12 | 436055 | ebi-a-GCST90025973 | rs2472297 | T | C | 0.262586 | Bilirubin Levels | rs2472297 | EUR | 74027880 | 76027880 |
| 15 | 75027880 | -0.0251 | 0.0026 | 3.97E-22 | 344052 | ebi-a-GCST90018948 | rs2472297 | T | C | 0.26649 | Blood Urea Nitrogen Levels | rs2472297 | EUR | 74027880 | 76027880 |
| 15 | 75027880 | -0.005 | 6.00E-04 | 5.78E-16 | 173149 | ebi-a-GCST90103632 | rs2472297 | T | C | 0.2591 | Blood Urea Nitrogen Levels | rs2472297 | EUR | 74027880 | 76027880 |
| 15 | 75027880 | 0.009221 | 0.001557 | 3.20E-09 | NA | ieu-b-5118 | rs2472297 | T | C | 0.26172 | Body Mass Index | rs2472297 | EUR | 74027880 | 76027880 |
| 15 | 75027880 | 0.014059 | 0.002188 | 8.40E-11 | 457756 | ebi-a-GCST90025994 | rs2472297 | T | C | 0.262581 | Body Mass Index | rs2472297 | EUR | 74027880 | 76027880 |
| 15 | 75027880 | 0.014183 | 0.002188 | 6.60E-11 | 532396 | ebi-a-GCST90029007 | rs2472297 | T | C | 0.262425 | Body Mass Index | rs2472297 | EUR | 74027880 | 76027880 |
| 15 | 75027880 | 0.0122 | 0.0019 | 1.40E-10 | 786480 | ieu-b-40 | rs2472297 | T | C | 0.2496 | Body Mass Index | rs2472297 | EUR | 74027880 | 76027880 |
| 15 | 75027880 | 0.013257 | 0.002246 | 3.60E-09 | 461460 | ukb-b-19953 | rs2472297 | T | C | 0.261612 | Body Mass Index | rs2472297 | EUR | 74027880 | 76027880 |
| 15 | 75027880 | 0.01339 | 0.002263 | 3.30E-09 | 454884 | ukb-b-2303 | rs2472297 | T | C | 0.26163 | Body Mass Index | rs2472297 | EUR | 74027880 | 76027880 |
| 15 | 75027880 | 0.014287 | 0.002329 | 8.62E-10 | 407609 | ebi-a-GCST90013870 | rs2472297 | T | C | NA | Body Mass Index | rs2472297 | EUR | 74027880 | 76027880 |
| 15 | 75027880 | 0.014179 | 0.00233 | 1.16E-09 | 407609 | ebi-a-GCST90013974 | rs2472297 | T | C | NA | Body Mass Index | rs2472297 | EUR | 74027880 | 76027880 |
| 15 | 75027880 | -0.01585 | 0.002022 | 4.50E-15 | 441640 | ukb-b-15926 | rs2472297 | T | C | 0.261458 | Cereal Intake | rs2472297 | EUR | 74027880 | 76027880 |
| 15 | 75027880 | 0.017741 | 0.00288 | 7.30E-10 | 64949 | ukb-b-9508 | rs2472297 | T | C | 0.25771 | Coffee Consumed | rs2472297 | EUR | 74027880 | 76027880 |

| | | | | | | | | | | | | | | | |
|---|---|---|---|---|---|---|---|---|---|---|---|---|---|---|---|
| 15 | 75027880 | 0.046471 | 0.001827 | 1.10E-142 | 428860 | ukb-b-5237 | rs2472297 | T | C | 0.262883 | Coffee Intake | rs2472297 | EUR | 74027880 | 76027880 |
| 15 | 75027880 | -0.01501 | 0.002277 | 4.60E-13 | 436939 | ebi-a-GCST90025959 | rs2472297 | T | C | 0.262546 | C-Reactive Protein Levels | rs2472297 | EUR | 74027880 | 76027880 |
| 15 | 75027880 | -0.02623 | 0.002188 | 4.15E-33 | 344104 | ukb-d-30700_irnt | rs2472297 | T | C | 0.26651 | Creatinine | rs2472297 | EUR | 74027880 | 76027880 |
| 15 | 75027880 | -0.38844 | 0.042336 | 4.53E-20 | 344104 | ukb-d-30700_raw | rs2472297 | T | C | 0.26651 | Creatinine | rs2472297 | EUR | 74027880 | 76027880 |
| 15 | 75027880 | -0.01866 | 0.002459 | 3.26E-14 | 344264 | ukb-d-30720_irnt | rs2472297 | T | C | 0.2665 | Cystatin C | rs2472297 | EUR | 74027880 | 76027880 |
| 15 | 75027880 | -0.0028 | 0.000439 | 1.69E-10 | 344264 | ukb-d-30720_raw | rs2472297 | T | C | 0.2665 | Cystatin C | rs2472297 | EUR | 74027880 | 76027880 |
| 15 | 75027880 | -0.02138 | 0.0022 | 4.00E-22 | 437846 | ebi-a-GCST90025945 | rs2472297 | T | C | 0.262579 | Cystatin C | rs2472297 | EUR | 74027880 | 76027880 |
| 15 | 75027880 | -0.01594 | 0.002121 | 5.77E-14 | 389834 | ebi-a-GCST90014003 | rs2472297 | T | C | NA | Cystatin C | rs2472297 | EUR | 74027880 | 76027880 |
| 15 | 75027880 | 0.01402 | 0.001685 | 8.62E-17 | 62072 | ebi-a-GCST90096908 | rs2472297 | T | C | NA | Decaffeinated Coffee Consumption | rs2472297 | EUR | 74027880 | 76027880 |
| 15 | 75027880 | -0.01673 | 0.001803 | 1.69E-20 | 810865 | ebi-a-GCST90000059 | rs2472297 | T | C | 0.2604 | Diastolic Blood Pressure | rs2472297 | EUR | 74027880 | 76027880 |
| 15 | 75027880 | -0.01666 | 0.001788 | 1.21E-20 | 810865 | ebi-a-GCST90000063 | rs2472297 | T | C | 0.257 | Diastolic Blood Pressure | rs2472297 | EUR | 74027880 | 76027880 |
| 15 | 75027880 | -0.1321 | 0.0201 | 5.39E-11 | 724537 | ieu-b-39 | rs2472297 | T | C | 0.261 | Diastolic Blood Pressure | rs2472297 | EUR | 74027880 | 76027880 |
| 15 | 75027880 | 0.002353 | 0.000207 | 5.84E-30 | 444097 | ebi-a-GCST90096910 | rs2472297 | T | C | NA | Drink Temperature | rs2472297 | EUR | 74027880 | 76027880 |
| 15 | 75027880 | -0.09836 | 0.0126 | 5.90E-15 | 64949 | ukb-b-3975 | rs2472297 | T | C | 0.25771 | Drinking Water Intake | rs2472297 | EUR | 74027880 | 76027880 |

| | | | | | | | | | | | | | | | |
|---|---|---|---|---|---|---|---|---|---|---|---|---|---|---|---|
| 15 | 75027880 | -0.07735 | 0.014003 | 3.32E-08 | 31185 | eqtl-a-ENSG00000067221 | rs2472297 | T | C | 0.235925 | ENSG00000067221 | rs2472297 | EUR | 74027880 | 76027880 |
| 15 | 75027880 | -0.22109 | 0.013894 | 5.17E-57 | 31185 | eqtl-a-ENSG00000103653 | rs2472297 | T | C | 0.235925 | ENSG00000103653 | rs2472297 | EUR | 74027880 | 76027880 |
| 15 | 75027880 | 0.093351 | 0.013996 | 2.57E-11 | 31185 | eqtl-a-ENSG00000138629 | rs2472297 | T | C | 0.235925 | ENSG00000138629 | rs2472297 | EUR | 74027880 | 76027880 |
| 15 | 75027880 | 0.0812 | 0.014001 | 6.66E-09 | 30971 | eqtl-a-ENSG00000140365 | rs2472297 | T | C | 0.235925 | ENSG00000140365 | rs2472297 | EUR | 74027880 | 76027880 |
| 15 | 75027880 | -0.27429 | 0.013827 | 1.41E-87 | 30971 | eqtl-a-ENSG00000140400 | rs2472297 | T | C | 0.235925 | ENSG00000140400 | rs2472297 | EUR | 74027880 | 76027880 |
| 15 | 75027880 | -0.25348 | 0.013855 | 9.00E-75 | 31185 | eqtl-a-ENSG00000140474 | rs2472297 | T | C | 0.235925 | ENSG00000140474 | rs2472297 | EUR | 74027880 | 76027880 |
| 15 | 75027880 | 0.272187 | 0.01383 | 3.11E-86 | 31185 | eqtl-a-ENSG00000140497 | rs2472297 | T | C | 0.235925 | ENSG00000140497 | rs2472297 | EUR | 74027880 | 76027880 |
| 15 | 75027880 | -0.10915 | 0.013988 | 6.04E-15 | 26110 | eqtl-a-ENSG00000169371 | rs2472297 | T | C | 0.235925 | ENSG00000169371 | rs2472297 | EUR | 74027880 | 76027880 |
| 15 | 75027880 | 0.087079 | 0.013999 | 4.96E-10 | 31185 | eqtl-a-ENSG00000177971 | rs2472297 | T | C | 0.235925 | ENSG00000177971 | rs2472297 | EUR | 74027880 | 76027880 |
| 15 | 75027880 | 0.187867 | 0.013929 | 1.85E-41 | 31185 | eqtl-a-ENSG00000178802 | rs2472297 | T | C | 0.235925 | ENSG00000178802 | rs2472297 | EUR | 74027880 | 76027880 |
| 15 | 75027880 | -0.08065 | 0.014002 | 8.42E-09 | 4868 | eqtl-a-ENSG00000227288 | rs2472297 | T | C | 0.235925 | ENSG00000227288 | rs2472297 | EUR | 74027880 | 76027880 |

| | | | | | | | | | | | | | | | |
|---|---|---|---|---|---|---|---|---|---|---|---|---|---|---|---|
| 15 | 75027880 | 0.003762 | 0.000286 | 1.32E-39 | 1159871 | ebi-a-GCST90026654 | rs2472297 | T | C | 0.2552 | Estimated Glomerular Filtration Rate (Creatinine) | rs2472297 | EUR | 74027880 | 76027880 |
| 15 | 75027880 | 0.0039 | 3.00E-04 | 6.17E-35 | 165726 | ebi-a-GCST90103633 | rs2472297 | T | C | 0.2613 | Estimated Glomerular Filtration Rate (Creatinine) | rs2472297 | EUR | 74027880 | 76027880 |
| 15 | 75027880 | 0.0038 | 3.00E-04 | 1.94E-33 | 1004040 | ebi-a-GCST90103634 | rs2472297 | T | C | 0.2614 | Estimated Glomerular Filtration Rate (Creatinine) | rs2472297 | EUR | 74027880 | 76027880 |
| 15 | 75027880 | 0.0031 | 5.00E-04 | 4.51E-09 | 460826 | ebi-a-GCST90103635 | rs2472297 | T | C | 0.2572 | Estimated Glomerular Filtration Rate (Cyctatin-C) | rs2472297 | EUR | 74027880 | 76027880 |
| 15 | 75027880 | 0.050494 | 0.007604 | 3.10E-11 | 64949 | ukb-b-748 | rs2472297 | T | C | 0.25771 | Filtered Coffee Intake | rs2472297 | EUR | 74027880 | 76027880 |
| 15 | 75027880 | -0.00535 | 0.000877 | 1.09E-09 | 433186 | ebi-a-GCST90096912 | rs2472297 | T | C | NA | Fresh Fruit Consumption | rs2472297 | EUR | 74027880 | 76027880 |
| 15 | 75027880 | 0.011104 | 0.001252 | 7.38E-19 | 283449 | ukb-d-1508_3 | rs2472297 | T | C | 0.269937 | Ground Coffee Consumption | rs2472297 | EUR | 74027880 | 76027880 |
| 15 | 75027880 | 0.018267 | 0.001407 | 1.51E-38 | 72276 | ebi-a-GCST90096913 | rs2472297 | T | C | NA | Ground coffee consumption | rs2472297 | EUR | 74027880 | 76027880 |
| 15 | 75027880 | -0.01028 | 0.001784 | 3.10E-09 | 458235 | ebi-a-GCST90025949 | rs2472297 | T | C | 0.262577 | Height | rs2472297 | EUR | 74027880 | 76027880 |
| 15 | 75027880 | -0.01032 | 0.001783 | 2.30E-09 | 673878 | ebi-a-GCST90029008 | rs2472297 | T | C | 0.26242 | Height | rs2472297 | EUR | 74027880 | 76027880 |
| 15 | 75027880 | -0.01539 | 0.001345 | 2.60E-30 | 457873 | ukb-b-14203 | rs2472297 | T | C | 0.261771 | Hot Drink Temperature | rs2472297 | EUR | 74027880 | 76027880 |
| 15 | 75027880 | -0.00981 | 0.001601 | 9.00E-10 | 454850 | ukb-b-19379 | rs2472297 | T | C | 0.261628 | Impedance of Arm (Left) | rs2472297 | EUR | 74027880 | 76027880 |
| 15 | 75027880 | -0.01118 | 0.001928 | 6.78E-09 | 331279 | ukb-a-272 | rs2472297 | T | C | 0.266644 | Impedance of Arm (Right) | rs2472297 | EUR | 74027880 | 76027880 |
| 15 | 75027880 | -0.01092 | 0.001595 | 7.60E-12 | 454826 | ukb-b-7859 | rs2472297 | T | C | 0.261628 | Impedance of Arm (Right) | rs2472297 | EUR | 74027880 | 76027880 |
| 15 | 75027880 | 0.020452 | 0.000991 | 1.08E-94 | 180764 | ebi-a-GCST90096914 | rs2472297 | T | C | NA | Instant Coffee Consumption | rs2472297 | EUR | 74027880 | 76027880 |

| | | | | | | | | | | | | | | | |
|---|---|---|---|---|---|---|---|---|---|---|---|---|---|---|---|
| 15 | 75027880 | 0.073119 | 0.012472 | 4.60 E-09 | 64949 | ukb-b-930 | rs2472297 | T | C | 0.25771 | Instant Coffee Consumption | rs2472297 | EUR | 74027880 | 76027880 |
| 15 | 75027880 | 0.010169 | 0.001799 | 1.60 E-08 | 454823 | ukb-b-7212 | rs2472297 | T | C | 0.261624 | Leg Fat Mass (Left) | rs2472297 | EUR | 74027880 | 76027880 |
| 15 | 75027880 | 0.010216 | 0.00182 | 2.00 E-08 | 454846 | ukb-b-18096 | rs2472297 | T | C | 0.261629 | Leg Fat Mass (Left) | rs2472297 | EUR | 74027880 | 76027880 |
| 15 | 75027880 | 0.010148 | 0.001715 | 3.25 E-09 | 331278 | ukb-a-278 | rs2472297 | T | C | 0.266644 | Leg Fat Percentage (Left) | rs2472297 | EUR | 74027880 | 76027880 |
| 15 | 75027880 | 0.009465 | 0.001431 | 3.70 E-11 | 454826 | ukb-b-18377 | rs2472297 | T | C | 0.261627 | Leg Fat Percentage (Left) | rs2472297 | EUR | 74027880 | 76027880 |
| 15 | 75027880 | 0.010068 | 0.001737 | 6.74 E-09 | 331296 | ukb-a-274 | rs2472297 | T | C | 0.266644 | Leg Fat Percentage (Right) | rs2472297 | EUR | 74027880 | 76027880 |
| 15 | 75027880 | 0.009736 | 0.001449 | 1.90 E-11 | 454854 | ukb-b-20531 | rs2472297 | T | C | 0.261628 | Leg Fat Percentage (Right) | rs2472297 | EUR | 74027880 | 76027880 |
| 15 | 75027880 | 0.01223 | 0.002121 | 8.27 E-09 | 544127 | ebi-a-GCST90002334 | rs2472297 | T | C | 0.258502 | Mean Corpuscular Volume | rs2472297 | EUR | 74027880 | 76027880 |
| 15 | 75027880 | 0.009746 | 0.002109 | 8.90 E-09 | 444035 | ebi-a-GCST90025963 | rs2472297 | T | C | 0.262678 | Mean Corpuscular Volume | rs2472297 | EUR | 74027880 | 76027880 |
| 15 | 75027880 | -0.01632 | 0.002695 | 1.41 E-09 | 350470 | ukb-d-30100_irnt | rs2472297 | T | C | 0.266596 | Mean Platelet Thrombocyte Volume | rs2472297 | EUR | 74027880 | 76027880 |
| 15 | 75027880 | -0.01608 | 0.002138 | 5.48 E-14 | 396616 | ebi-a-GCST90013981 | rs2472297 | T | C | NA | Mean Platelet Thrombocyte Volume | rs2472297 | EUR | 74027880 | 76027880 |
| 15 | 75027880 | -0.01465 | 0.002196 | 2.58 E-11 | 460935 | ebi-a-GCST90002346 | rs2472297 | T | C | 0.262905 | Mean Platelet Volume | rs2472297 | EUR | 74027880 | 76027880 |
| 15 | 75027880 | -0.01413 | 0.002335 | 1.50 E-09 | 408112 | ebi-a-GCST90002395 | rs2472297 | T | C | 0.263703 | Mean Platelet Volume | rs2472297 | EUR | 74027880 | 76027880 |
| 15 | 75027880 | -0.0136 | 0.002011 | 4.00 E-17 | 445364 | ebi-a-GCST90025960 | rs2472297 | T | C | 0.262668 | Mean Platelet Volume | rs2472297 | EUR | 74027880 | 76027880 |
| 15 | 75027880 | -0.01391 | 0.002012 | 3.20 E-17 | 694866 | ebi-a-GCST90028996 | rs2472297 | T | C | 0.262515 | Mean Platelet Volume | rs2472297 | EUR | 74027880 | 76027880 |

| | | | | | | | | | | | | | | | |
|---|---|---|---|---|---|---|---|---|---|---|---|---|---|---|---|
| 15 | 75027880 | 0.010778 | 0.002226 | 3.90E-08 | 436944 | ebi-a-GCST90025975 | rs2472297 | T | C | 0.262698 | Mean Reticulocyte Volume | rs2472297 | EUR | 74027880 | 76027880 |
| 15 | 75027880 | 0.050613 | 0.003941 | 9.26E-38 | 258758 | ebi-a-GCST90096893 | rs2472297 | T | C | NA | Overall Healthy Diet | rs2472297 | EUR | 74027880 | 76027880 |
| 15 | 75027880 | -0.0176 | 0.0029 | 7.00E-10 | 321047 | ebi-a-GCST007430 | rs2472297 | T | C | 0.264 | Peak Expiratory Flow (PEF) | rs2472297 | EUR | 74027880 | 76027880 |
| 15 | 75027880 | -0.01205 | 0.002083 | 7.20E-09 | 421986 | ukb-b-12019 | rs2472297 | T | C | 0.261532 | Peak Expiratory Flow (PEF) | rs2472297 | EUR | 74027880 | 76027880 |
| 15 | 75027880 | -0.01139 | 0.002075 | 3.98E-08 | 374599 | ebi-a-GCST90013995 | rs2472297 | T | C | NA | Peak Expiratory Flow (PEF) | rs2472297 | EUR | 74027880 | 76027880 |
| 15 | 75027880 | 0.021282 | 0.002747 | 9.49E-15 | 314658 | ukb-d-30810_irnt | rs2472297 | T | C | 0.26642 | Phosphate | rs2472297 | EUR | 74027880 | 76027880 |
| 15 | 75027880 | 0.003423 | 0.000441 | 8.66E-15 | 314658 | ukb-d-30810_raw | rs2472297 | T | C | 0.26642 | Phosphate | rs2472297 | EUR | 74027880 | 76027880 |
| 15 | 75027880 | 0.021624 | 0.002517 | 8.56E-18 | 357302 | ebi-a-GCST90014010 | rs2472297 | T | C | NA | Phosphate | rs2472297 | EUR | 74027880 | 76027880 |
| 15 | 75027880 | 0.014749 | 0.002142 | 5.96E-12 | 542827 | ebi-a-GCST90002357 | rs2472297 | T | C | 0.258073 | Platelet Count | rs2472297 | EUR | 74027880 | 76027880 |
| 15 | 75027880 | 0.015701 | 0.002399 | 5.90E-11 | 408112 | ebi-a-GCST90002402 | rs2472297 | T | C | 0.26374 | Platelet Count | rs2472297 | EUR | 74027880 | 76027880 |
| 15 | 75027880 | 0.0143 | 0.0026 | 4.42E-08 | 350474 | ebi-a-GCST90018969 | rs2472297 | T | C | 0.266596 | Platelet Count | rs2472297 | EUR | 74027880 | 76027880 |
| 15 | 75027880 | 0.015031 | 0.002119 | 1.20E-14 | 444866 | ebi-a-GCST90025951 | rs2472297 | T | C | 0.262661 | Platelet Count | rs2472297 | EUR | 74027880 | 76027880 |
| 15 | 75027880 | 0.01639 | 0.002141 | 1.60E-16 | 600968 | ebi-a-GCST90028999 | rs2472297 | T | C | 0.262505 | Platelet Count | rs2472297 | EUR | 74027880 | 76027880 |
| 15 | 75027880 | 0.01428 | 0.002609 | 4.42E-08 | 350474 | ukb-d-30080_irnt | rs2472297 | T | C | 0.266596 | Platelet Count | rs2472297 | EUR | 74027880 | 76027880 |
| 15 | 75027880 | 0.016206 | 0.002224 | 3.18E-13 | 396621 | ebi-a-GCST90013980 | rs2472297 | T | C | NA | Platelet Count | rs2472297 | EUR | 74027880 | 76027880 |

| | | | | | | | | | | | | | | | |
|---|---|---|---|---|---|---|---|---|---|---|---|---|---|---|---|
| 15 | 75027880 | -0.02418 | 0.002448 | 5.20E-23 | 408112 | ebi-a-GCST90002401 | rs2472297 | T | C | 0.263713 | Platelet Distribution Width | rs2472297 | EUR | 74027880 | 76027880 |
| 15 | 75027880 | -0.02119 | 0.002192 | 2.00E-26 | 572201 | ebi-a-GCST90029000 | rs2472297 | T | C | 0.262487 | Platelet Distribution Width | rs2472297 | EUR | 74027880 | 76027880 |
| 15 | 75027880 | -0.02297 | 0.002668 | 7.36E-18 | 350470 | ukb-d-30110_irnt | rs2472297 | T | C | 0.266596 | Platelet Distribution Width | rs2472297 | EUR | 74027880 | 76027880 |
| 15 | 75027880 | -0.01556 | 0.002753 | 1.59E-08 | 326816 | ukb-a-334 | rs2472297 | T | C | 0.266644 | Potassium in Urine | rs2472297 | EUR | 74027880 | 76027880 |
| 15 | 75027880 | -0.01846 | 0.0025 | 1.51E-13 | 395995 | ebi-a-GCST90013988 | rs2472297 | T | C | NA | Potassium in Urine | rs2472297 | EUR | 74027880 | 76027880 |
| 15 | 75027880 | 0.006266 | 0.00103 | 1.18E-09 | 211628 | ebi-a-GCST90096922 | rs2472297 | T | C | NA | Red Wine Consumption | rs2472297 | EUR | 74027880 | 76027880 |
| 15 | 75027880 | 0.014479 | 0.002437 | 3.50E-10 | 400938 | ebi-a-GCST90025992 | rs2472297 | T | C | 0.262478 | Serum Albumin Levels | rs2472297 | EUR | 74027880 | 76027880 |
| 15 | 75027880 | -0.01203 | 0.002168 | 4.40E-09 | 437896 | ebi-a-GCST90025947 | rs2472297 | T | C | 0.262579 | Serum Alkaline Phosphatase Levels | rs2472297 | EUR | 74027880 | 76027880 |
| 15 | 75027880 | -0.03045 | 0.002261 | 8.70E-43 | 437660 | ebi-a-GCST90025946 | rs2472297 | T | C | 0.262585 | Serum Creatinine | rs2472297 | EUR | 74027880 | 76027880 |
| 15 | 75027880 | -0.02425 | 0.001951 | 1.81E-35 | 389678 | ebi-a-GCST90014001 | rs2472297 | T | C | NA | Serum Creatinine | rs2472297 | EUR | 74027880 | 76027880 |
| 15 | 75027880 | -0.0262 | 0.0022 | 4.08E-33 | 344104 | ebi-a-GCST90018979 | rs2472297 | T | C | 0.26651 | Serum Creatinine | rs2472297 | EUR | 74027880 | 76027880 |
| 15 | 75027880 | 0.02436 | 0.00244 | 9.60E-25 | 400159 | ebi-a-GCST90025948 | rs2472297 | T | C | 0.262471 | Serum Phosphate Levels | rs2472297 | EUR | 74027880 | 76027880 |
| 15 | 75027880 | -0.02706 | 0.00236 | 9.50E-31 | 437580 | ebi-a-GCST90025991 | rs2472297 | T | C | 0.262581 | Serum Urea Levels | rs2472297 | EUR | 74027880 | 76027880 |
| 15 | 75027880 | -0.0223 | 0.0023 | 2.42E-22 | 343836 | ebi-a-GCST90018977 | rs2472297 | T | C | 0.26649 | Serum Uric Acid Levels | rs2472297 | EUR | 74027880 | 76027880 |
| 15 | 75027880 | -0.02029 | 0.00267 | 2.98E-14 | 326831 | ukb-a-335 | rs2472297 | T | C | 0.266644 | Sodium in Urine | rs2472297 | EUR | 74027880 | 76027880 |

| | | | | | | | | | | | | | | | |
|---|---|---|---|---|---|---|---|---|---|---|---|---|---|---|---|
| 15 | 75027880 | -0.02215 | 0.002419 | 5.38E-20 | 396020 | ebi-a-GCST90013989 | rs2472297 | T | C | NA | Sodium in Urine | rs2472297 | EUR | 74027880 | 76027880 |
| 15 | 75027880 | 10.2075 | 1.04443 | 1.50E-22 | 64949 | ukb-b-3291 | rs2472297 | T | C | 0.25771 | Standard Tea Intake | rs2472297 | EUR | 74027880 | 76027880 |
| 15 | 75027880 | -0.01251 | 0.0018 | 3.63E-12 | 810865 | ebi-a-GCST90000062 | rs2472297 | T | C | 0.2604 | Systolic Blood Pressure | rs2472297 | EUR | 74027880 | 76027880 |
| 15 | 75027880 | -0.01251 | 0.001786 | 2.48E-12 | 810865 | ebi-a-GCST90000066 | rs2472297 | T | C | 0.2571 | Systolic Blood Pressure | rs2472297 | EUR | 74027880 | 76027880 |
| 15 | 75027880 | 0.017197 | 0.001363 | 1.65E-36 | 434171 | ebi-a-GCST90096926 | rs2472297 | T | C | NA | Tea Consumption | rs2472297 | EUR | 74027880 | 76027880 |
| 15 | 75027880 | 0.053345 | 0.002401 | 2.30E-109 | 447485 | ukb-b-6066 | rs2472297 | T | C | 0.262049 | Tea Consumption | rs2472297 | EUR | 74027880 | 76027880 |
| 15 | 75027880 | 0.012504 | 0.002012 | 5.17E-10 | 388303 | ebi-a-GCST90013872 | rs2472297 | T | C | NA | Total Bilirubin | rs2472297 | EUR | 74027880 | 76027880 |
| 15 | 75027880 | 0.012546 | 0.002026 | 5.95E-10 | 388303 | ebi-a-GCST90014012 | rs2472297 | T | C | NA | Total Bilirubin | rs2472297 | EUR | 74027880 | 76027880 |
| 15 | 75027880 | -0.02232 | 0.002295 | 2.44E-22 | 343836 | ukb-d-30880_irnt | rs2472297 | T | C | 0.26649 | Urate | rs2472297 | EUR | 74027880 | 76027880 |
| 15 | 75027880 | -1.8062 | 0.18521 | 1.82E-22 | 343836 | ukb-d-30880_raw | rs2472297 | T | C | 0.26649 | Urate | rs2472297 | EUR | 74027880 | 76027880 |
| 15 | 75027880 | -0.03295 | 0.00222 | 3.00E-50 | 437354 | ebi-a-GCST90025965 | rs2472297 | T | C | 0.262583 | Urate | rs2472297 | EUR | 74027880 | 76027880 |
| 15 | 75027880 | -0.0209 | 0.002006 | 2.10E-25 | 389404 | ebi-a-GCST90014015 | rs2472297 | T | C | NA | Urate | rs2472297 | EUR | 74027880 | 76027880 |
| 15 | 75027880 | -0.02506 | 0.002591 | 3.99E-22 | 344052 | ukb-d-30670_irnt | rs2472297 | T | C | 0.26649 | Urea | rs2472297 | EUR | 74027880 | 76027880 |
| 15 | 75027880 | -0.03348 | 0.003633 | 3.12E-20 | 344052 | ukb-d-30670_raw | rs2472297 | T | C | 0.26649 | Urea | rs2472297 | EUR | 74027880 | 76027880 |
| 15 | 75027880 | -0.0249 | 0.002379 | 1.23E-25 | 389608 | ebi-a-GCST90013998 | rs2472297 | T | C | NA | Urea Levels | rs2472297 | EUR | 74027880 | 76027880 |

| | | | | | | | | | | | | | | | |
|---|---|---|---|---|---|---|---|---|---|---|---|---|---|---|---|
| 15 | 75027880 | 0.018124 | 0.00188 | 5.31E-22 | 382500 | ebi-a-GCST006586 | rs2472297 | T | C | 0.267268 | Urinary Albumin Excretion | rs2472297 | EUR | 74027880 | 76027880 |
| 15 | 75027880 | -0.02715 | 0.002597 | 1.42E-25 | 327525 | ukb-a-333 | rs2472297 | T | C | 0.266644 | Urinary Creatinine | rs2472297 | EUR | 74027880 | 76027880 |
| 15 | 75027880 | -0.02842 | 0.002356 | 1.66E-33 | 396837 | ebi-a-GCST90013987 | rs2472297 | T | C | NA | Urinary Creatinine | rs2472297 | EUR | 74027880 | 76027880 |
| 15 | 75027880 | -0.08378 | 0.004442 | 2.45E-79 | 445799 | ebi-a-GCST90096929 | rs2472297 | T | C | NA | Water Consumption | rs2472297 | EUR | 74027880 | 76027880 |
| 15 | 75027880 | -0.04809 | 0.004639 | 3.50E-25 | 400642 | ebi-a-GCST90096928 | rs2472297 | T | C | NA | Water Consumption Corrected for Coffee | rs2472297 | EUR | 74027880 | 76027880 |
| 15 | 75027880 | -0.03342 | 0.002057 | 2.40E-59 | 427588 | ukb-b-14898 | rs2472297 | T | C | 0.26061 | Water Intake | rs2472297 | EUR | 74027880 | 76027880 |

#### Supplementary Table 4 - Genetic Colocalization Results (PWCoCo)

| Exposure | Outcome | SNP1 | SNP2 | H4 | Conditional Status | LogABF |
|---|---|---|---|---|---|---|
| Clozapine Plasma Level | 25 Hydroxyvitamin D | Unconditioned | Unconditioned | 0.964721 | Unconditional | 87.8816 |
| Clozapine Plasma Level | 25 Hydroxyvitamin D | Unconditioned | Unconditioned | 0.9906 | Unconditional | 86.8581 |
| Clozapine Plasma Level | 25 Hydroxyvitamin D | Unconditioned | Unconditioned | 0.939667 | Unconditional | 92.2257 |
| Clozapine Plasma Level | 25 Hydroxyvitamin D | Unconditioned | Unconditioned | 0.939667 | Unconditional | 92.2257 |
| Clozapine Plasma Level | 25 Hyroxyvitamin D | Unconditioned | Unconditioned | 0.962933 | Unconditional | 61.8826 |
| Clozapine Plasma Level | 25 Hyroxyvitamin D | Unconditioned | Unconditioned | 0.962933 | Unconditional | 61.8826 |
| Clozapine Plasma Level | 25 Hyroxyvitamin D | Unconditioned | Unconditioned | 0.998514 | Unconditional | 87.0637 |
| Clozapine Plasma Level | 25 Hyroxyvitamin D | Unconditioned | Unconditioned | 0.968536 | Unconditional | 68.7752 |
| Clozapine Plasma Level | 25 Hyroxyvitamin D | Unconditioned | Unconditioned | 0.968536 | Unconditional | 68.7752 |
| Clozapine Plasma Level | Bilirubin | rs3732218* | rs887829 | 0.969434 | Conditional Both | 1394.23 |
| Clozapine Plasma Level | Bilirubin | rs3732218* | rs10180090 | 0.852145 | Conditional Both | 1155.23 |
| Clozapine Plasma Level | Bilirubin | rs3732218* | rs10180090 | 0.854293 | Conditional Both | 1129.24 |
| Clozapine Plasma Level | Bilirubin | rs3732218* | rs887829 | 0.968581 | Conditional Both | 234.689 |
| Clozapine Plasma Level | Bilirubin | rs3732218* | rs7567229 | 0.854093 | Conditional Both | 12745 |
| Clozapine Plasma Level | Bilirubin | rs3732218* | rs7567229 | 0.854093 | Conditional Both | 12745 |
| Clozapine Plasma Level | Bilirubin | rs3732218* | rs2284294 | 0.889816 | Conditional Both | 33.021 |
| Clozapine Plasma Level | Bilirubin | rs3732218* | rs2284294 | 0.889816 | Conditional Both | 33.021 |
| Clozapine Plasma Level | Bilirubin | Unconditioned | Unconditioned | 0.999965 | Unconditional | 26.302 |
| Clozapine Plasma Level | Blood Urea Nitrogen | Unconditioned | Unconditioned | 1 | Unconditional | 50.7767 |
| Clozapine Plasma Level | Coffee Consumption | Unconditioned | Unconditioned | 0.999997 | Unconditional | 23.5822 |
| Clozapine Plasma Level | Coffee Consumption | Unconditioned | Unconditioned | 1 | Unconditional | 326.977 |

| | | | | | | |
|---|---|---|---|---|---|---|
| Clozapine Plasma Level | Coffee Consumption | Unconditioned | Unconditioned | 0.993918 | Unconditional | 26.6595 |
| Clozapine Plasma Level | Coffee Consumption | Unconditioned | Unconditioned | 1 | Unconditional | 88.7291 |
| Clozapine Plasma Level | Coffee Consumption | Unconditioned | Unconditioned | 1 | Unconditional | 217.095 |
| Clozapine Plasma Level | Coffee Consumption | Unconditioned | Unconditioned | 0.99999 | Unconditional | 21.7994 |
| Clozapine Plasma Level | Creatinine | Unconditioned | Unconditioned | 1 | Unconditional | 75.6414 |
| Clozapine Plasma Level | Creatinine | Unconditioned | Unconditioned | 1 | Unconditional | 58.4524 |
| Clozapine Plasma Level | Creatinine | Unconditioned | Unconditioned | 1 | Unconditional | 94.2892 |
| Clozapine Plasma Level | Creatinine | Unconditioned | Unconditioned | 1 | Unconditional | 80.9571 |
| Clozapine Plasma Level | Creatinine | Unconditioned | Unconditioned | 1 | Unconditional | 74.833 |
| Clozapine Plasma Level | C Reactive Protein | Unconditioned | Unconditioned | 1 | Unconditional | 25.4212 |
| Clozapine Plasma Level | Cystatin C | Unconditioned | Unconditioned | 1 | Unconditional | 32.5826 |
| Clozapine Plasma Level | Cystatin C | Unconditioned | Unconditioned | 1 | Unconditional | 50.9251 |
| Clozapine Plasma Level | Cystatin C | Unconditioned | Unconditioned | 0.990486 | Unconditional | 31.9614 |
| Clozapine Plasma Level | Fresh Fruit Consumption | Unconditioned | Unconditioned | 0.999989 | Unconditional | 22.258 |
| Clozapine Plasma Level | Impedance of Left Arm | Unconditioned | Unconditioned | 0.999856 | Unconditional | 22.4278 |
| Clozapine Plasma Level | Impedance of Right Arm | Unconditioned | Unconditioned | 0.986041 | Unconditional | 20.6286 |
| Clozapine Plasma Level | Overall Healthy Diet | Unconditioned | Unconditioned | 1 | Unconditional | 86.3939 |
| Clozapine Plasma Level | Peak Expiratory Flow | Unconditioned | Unconditioned | 0.999875 | Unconditional | 20.4378 |
| Clozapine Plasma Level | Peak Expiratory Flow | Unconditioned | Unconditioned | 0.999774 | Unconditional | 18.8317 |
| Clozapine Plasma Level | Peak Expiratory Flow | Unconditioned | Unconditioned | 0.893014 | Unconditional | 40.7914 |
| Clozapine Plasma Level | Potassium in Urine | Unconditioned | Unconditioned | 0.999836 | Unconditional | 19.7952 |
| Clozapine Plasma Level | Potassium in Urine | Unconditioned | Unconditioned | 0.999991 | Unconditional | 30.9988 |
| Clozapine Plasma Level | Serum Phosphate | Unconditioned | Unconditioned | 0.946901 | Unconditional | 53.6206 |
| Clozapine Plasma Level | Serum Urea | Unconditioned | Unconditioned | 1 | Unconditional | 69.383 |

| | | | | | | |
|---|---|---|---|---|---|---|
| Clozapine Plasma Level | Serum Uric Acid | Unconditioned | Unconditioned | 1 | Unconditional | 50.884 |
| Clozapine Plasma Level | Sodium in Urine | Unconditioned | Unconditioned | 0.999987 | Unconditional | 32.6943 |
| Clozapine Plasma Level | Sodium in Urine | Unconditioned | Unconditioned | 0.999999 | Unconditional | 45.6347 |
| Clozapine Plasma Level | Tea Consumption | Unconditioned | Unconditioned | 1 | Unconditional | 52.3182 |
| Clozapine Plasma Level | Tea Consumption | Unconditioned | Unconditioned | 1 | Unconditional | 83.289 |
| Clozapine Plasma Level | Tea Consumption | Unconditioned | Unconditioned | 1 | Unconditional | 250.435 |
| Clozapine Plasma Level | Total Testosterone | Unconditioned | Unconditioned | 0.969384 | Unconditional | 37.9852 |
| Clozapine Plasma Level | Total Testosterone | Unconditioned | Unconditioned | 0.983943 | Unconditional | 35.8764 |
| Clozapine Plasma Level | Total Testosterone | Unconditioned | Unconditioned | 0.957861 | Unconditional | 44.0121 |
| Clozapine Plasma Level | Total Testosterone | Unconditioned | Unconditioned | 0.970804 | Unconditional | 23.4109 |
| Clozapine Plasma Level | Total Testosterone | Unconditioned | Unconditioned | 0.960916 | Unconditional | 47.0756 |
| Clozapine Plasma Level | Total Testosterone | Unconditioned | Unconditioned | 0.960916 | Unconditional | 47.0756 |
| Clozapine Plasma Level | Total Testosterone | Unconditioned | Unconditioned | 0.967799 | Unconditional | 52.7835 |
| Clozapine Plasma Level | Total Testosterone | Unconditioned | Unconditioned | 0.967799 | Unconditional | 52.7835 |
| Clozapine Plasma Level | Total Testosterone | Unconditioned | Unconditioned | 0.967653 | Unconditional | 22.8486 |
| Clozapine Plasma Level | Total Testosterone | Unconditioned | Unconditioned | 0.979196 | Unconditional | 42.4074 |
| Clozapine Plasma Level | Total Testosterone | Unconditioned | Unconditioned | 0.967461 | Unconditional | 27.8788 |
| Clozapine Plasma Level | Total Testosterone | Unconditioned | Unconditioned | 0.972234 | Unconditional | 59.9247 |
| Clozapine Plasma Level | Urate | Unconditioned | Unconditioned | 1 | Unconditional | 51.0502 |
| Clozapine Plasma Level | Urate | Unconditioned | Unconditioned | 1 | Unconditional | 113.743 |
| Clozapine Plasma Level | Urate | Unconditioned | Unconditioned | 1 | Unconditional | 57.9671 |
| Clozapine Plasma Level | Urea | Unconditioned | Unconditioned | 1 | Unconditional | 50.5606 |
| Clozapine Plasma Level | Urea | Unconditioned | Unconditioned | 1 | Unconditional | 58.4965 |
| Clozapine Plasma Level | Urinary Albumin Excretion | Unconditioned | Unconditioned | 0.99972 | Unconditional | 50.2262 |

| | | | | | | |
|---|---|---|---|---|---|---|
| Clozapine Plasma Level | Water Consumption | Unconditioned | Unconditioned | 0.999991 | Unconditional | 35.0593 |
| Clozapine Plasma Level | Water Consumption | Unconditioned | Unconditioned | 1 | Unconditional | 181.468 |
| Clozapine Plasma Level | Water Consumption | Unconditioned | Unconditioned | 1 | Unconditional | 57.4477 |
| Clozapine Plasma Level | Water Consumption | Unconditioned | Unconditioned | 1 | Unconditional | 135.618 |
| Clozapine Plasma Level | X 11440 | Unconditioned | Unconditioned | 0.980717 | Unconditional | 26.9575 |
| Clozapine-Norclozapine Plasma Ratio | Bilirubin | rs11726899 | rs293435 | 0.92734 | Conditional Both | 123.232 |
| Clozapine-Norclozapine Plasma Ratio | Bilirubin | rs11726899 | rs293435 | 0.927342 | Conditional Both | 123.223 |
| Clozapine-Norclozapine Plasma Ratio | Bilirubin | rs11726899 | rs293435 | 0.927342 | Conditional Both | 123.223 |
| Clozapine-Norclozapine Plasma Ratio | Bilirubin | rs835309 | rs293435 | 0.957796 | Conditional Both | 13.7865 |
| Clozapine-Norclozapine Plasma Ratio | Bilirubin | rs835309 | rs293435 | 0.957801 | Conditional Both | 13.7889 |
| Clozapine-Norclozapine Plasma Ratio | Bilirubin | rs835309 | rs293435 | 0.957801 | Conditional Both | 13.789 |
| Clozapine-Norclozapine Plasma Ratio | Bilirubin | rs835309 | rs2670430 | 0.989571 | Conditional Both | 19.6749 |
| Clozapine-Norclozapine Plasma Ratio | Bilirubin | rs835309 | rs2670430 | 0.989571 | Conditional Both | 19.6751 |
| Clozapine-Norclozapine Plasma Ratio | Bilirubin | rs835309 | rs2670430 | 0.989571 | Conditional Both | 19.6727 |
| Clozapine-Norclozapine Plasma Ratio | Bilirubin | rs72846859 | rs2670430 | 0.999718 | Conditional Both | 31.5329 |
| Clozapine-Norclozapine Plasma Ratio | Bilirubin | rs72846859 | rs2670430 | 0.999718 | Conditional Both | 31.5331 |
| Clozapine-Norclozapine Plasma Ratio | Bilirubin | rs72846859 | rs2670430 | 0.999718 | Conditional Both | 31.5318 |
| Clozapine-Norclozapine Plasma Ratio | Bilirubin | Unconditioned | Unconditioned | 0.991493 | Unconditional | 62.3425 |
| Clozapine-Norclozapine Plasma Ratio | Bilirubin | Unconditioned | Unconditioned | 0.847452 | Unconditional | 46.1441 |
| Clozapine-Norclozapine Plasma Ratio | Bilirubin | Unconditioned | Unconditioned | 0.847464 | Unconditional | 46.1452 |
| Clozapine-Norclozapine Plasma Ratio | Bilirubin | Unconditioned | Unconditioned | 0.846334 | Unconditional | 347.183 |
| Clozapine-Norclozapine Plasma Ratio | Bilirubin | Unconditioned | Unconditioned | 0.846334 | Unconditional | 347.187 |
| Clozapine-Norclozapine Plasma Ratio | Bilirubin | Unconditioned | Unconditioned | 0.846334 | Unconditional | 347.175 |
| Clozapine-Norclozapine Plasma Ratio | Bilirubin | Unconditioned | Unconditioned | 0.846334 | Unconditional | 347.173 |

| | | | | | | |
|---|---|---|---|---|---|---|
| Clozapine-Norclozapine Plasma Ratio | Bilirubin | Unconditioned | Unconditioned | 0.862011 | Unconditional | 58.1654 |
| Clozapine-Norclozapine Plasma Ratio | Bilirubin | Unconditioned | Unconditioned | 0.918877 | Unconditional | 57.1685 |
| Clozapine-Norclozapine Plasma Ratio | Bilirubin | Unconditioned | Unconditioned | 0.945142 | Unconditional | 58.4516 |
| Clozapine-Norclozapine Plasma Ratio | Bilirubin | Unconditioned | Unconditioned | 0.992387 | Unconditional | 352.125 |
| Clozapine-Norclozapine Plasma Ratio | Bilirubin | Unconditioned | Unconditioned | 0.998647 | Unconditional | 350.946 |
| Clozapine-Norclozapine Plasma Ratio | Mean Corpuscular Volume | rs835309 | rs11726946 | 0.898953 | Conditional Both | 40.0267 |
| Clozapine-Norclozapine Plasma Ratio | Mean Corpuscular Volume | rs72846859 | rs72846859 | 0.999998 | Conditional Both | 61.7142 |
| Clozapine-Norclozapine Plasma Ratio | Total Testosterone | rs12646327 | rs7438942 | 0.901361 | Conditional Both | 56.5107 |
| Clozapine-Norclozapine Plasma Ratio | Total Testosterone | rs835309 | rs9994887 | 0.95699 | Conditional Both | 10.3865 |
| Clozapine-Norclozapine Plasma Ratio | X 14473 | Unconditioned | Unconditioned | 0.96687 | Unconditional | 52.7449 |
| Norclozapine Plasma Level | 25 hydroxyvitamin D | rs1824264 | rs4466107 | 0.999562 | Conditional Both | 28.4326 |
| Norclozapine Plasma Level | 25 Hydroxyvitamin D | rs1824264 | rs114024927 | 0.957364 | Conditional Both | 26.2375 |
| Norclozapine Plasma Level | 25 Hydroxyvitamin D | Unconditioned | Unconditioned | 0.964099 | Unconditional | 89.3327 |
| Norclozapine Plasma Level | 25 Hydroxyvitamin D | Unconditioned | Unconditioned | 0.990584 | Unconditional | 88.3373 |
| Norclozapine Plasma Level | 25 Hydroxyvitamin D | Unconditioned | Unconditioned | 0.942417 | Unconditional | 93.7584 |
| Norclozapine Plasma Level | 25 Hydroxyvitamin D | Unconditioned | Unconditioned | 0.942417 | Unconditional | 93.7584 |
| Norclozapine Plasma Level | 25 Hyroxyvitamin D | Unconditioned | Unconditioned | 0.961088 | Unconditional | 63.3093 |
| Norclozapine Plasma Level | 25 Hyroxyvitamin D | Unconditioned | Unconditioned | 0.961088 | Unconditional | 63.3093 |
| Norclozapine Plasma Level | 25 Hyroxyvitamin D | Unconditioned | Unconditioned | 0.998486 | Unconditional | 88.2741 |
| Norclozapine Plasma Level | 25 Hyroxyvitamin D | Unconditioned | Unconditioned | 0.967552 | Unconditional | 70.2313 |
| Norclozapine Plasma Level | 25 Hyroxyvitamin D | Unconditioned | Unconditioned | 0.967552 | Unconditional | 70.2313 |
| Norclozapine Plasma Level | Bilirubin | rs4663325* | rs17863787 | 0.808566 | Conditional Both | 666.838 |
| Norclozapine Plasma Level | Bilirubin | rs4663325* | rs17863787 | 0.809381 | Conditional Both | 707.713 |
| Norclozapine Plasma Level | Bilirubin | rs4663325* | rs10180090 | 0.895005 | Conditional Both | 1129.59 |

| | | | | | | |
|---|---|---|---|---|---|---|
| Norclozapine Plasma Level | Bilirubin | rs4663325* | rs10180090 | 0.895471 | Conditional Both | 1155.58 |
| Norclozapine Plasma Level | Bilirubin | rs2926036 | rs2670430 | 0.985766 | Conditional Both | 63.405 |
| Norclozapine Plasma Level | Bilirubin | rs2926036 | rs2670430 | 0.985766 | Conditional Both | 63.4053 |
| Norclozapine Plasma Level | Bilirubin | rs2926036 | rs2670430 | 0.985766 | Conditional Both | 63.4085 |
| Norclozapine Plasma Level | Bilirubin | rs4663325* | rs17863787 | 0.8105 | Conditional Both | 302.017 |
| Norclozapine Plasma Level | Bilirubin | rs4663325* | rs17863787 | 0.8105 | Conditional Both | 302.017 |
| Norclozapine Plasma Level | Bilirubin | rs4663325* | rs7567229 | 0.896151 | Conditional Both | 12745.3 |
| Norclozapine Plasma Level | Bilirubin | rs4663325* | rs7567229 | 0.896151 | Conditional Both | 12745.3 |
| Norclozapine Plasma Level | Bilirubin | rs4663325* | rs2284294 | 0.937328 | Conditional Both | 33.6031 |
| Norclozapine Plasma Level | Bilirubin | rs4663325* | rs2284294 | 0.937328 | Conditional Both | 33.6031 |
| Norclozapine Plasma Level | Bilirubin | Unconditioned | Unconditioned | 0.999963 | Unconditional | 22.6255 |
| Norclozapine Plasma Level | Bilirubin | Unconditioned | Unconditioned | 0.82273 | Unconditional | 68.8938 |
| Norclozapine Plasma Level | Bilirubin | Unconditioned | Unconditioned | 0.82273 | Unconditional | 68.8939 |
| Norclozapine Plasma Level | Bilirubin | Unconditioned | Unconditioned | 0.822731 | Unconditional | 68.8942 |
| Norclozapine Plasma Level | Bilirubin | Unconditioned | Unconditioned | 0.822731 | Unconditional | 68.8944 |
| Norclozapine Plasma Level | Bilirubin | Unconditioned | Unconditioned | 0.973565 | Unconditional | 73.1564 |
| Norclozapine Plasma Level | Bilirubin | Unconditioned | Unconditioned | 0.973565 | Unconditional | 73.1562 |
| Norclozapine Plasma Level | Bilirubin | Unconditioned | Unconditioned | 0.984126 | Unconditional | 72.8209 |
| Norclozapine Plasma Level | Bilirubin | Unconditioned | Unconditioned | 0.99165 | Unconditional | 71.8527 |
| Norclozapine Plasma Level | Blood Urea Nitrogen | Unconditioned | Unconditioned | 0.999998 | Unconditional | 47.1006 |
| Norclozapine Plasma Level | Coffee Consumption | Unconditioned | Unconditioned | 0.999995 | Unconditional | 19.9061 |
| Norclozapine Plasma Level | Coffee Consumption | Unconditioned | Unconditioned | 0.999998 | Unconditional | 323.301 |
| Norclozapine Plasma Level | Coffee Consumption | Unconditioned | Unconditioned | 0.993917 | Unconditional | 22.9852 |
| Norclozapine Plasma Level | Coffee Consumption | Unconditioned | Unconditioned | 0.999998 | Unconditional | 85.0531 |

| | | | | | | |
|---|---|---|---|---|---|---|
| Norclozapine Plasma Level | Coffee Consumption | Unconditioned | Unconditioned | 0.999998 | Unconditional | 213.419 |
| Norclozapine Plasma Level | Coffee Consumption | Unconditioned | Unconditioned | 0.999988 | Unconditional | 18.1233 |
| Norclozapine Plasma Level | Cotinine in Current Smokers | Unconditioned | Unconditioned | 0.957686 | Unconditional | 86.6263 |
| Norclozapine Plasma Level | Cotinine in Current Smokers | Unconditioned | Unconditioned | 0.957686 | Unconditional | 86.6263 |
| Norclozapine Plasma Level | Cotinine in Current Smokers | Unconditioned | Unconditioned | 0.957687 | Unconditional | 86.6268 |
| Norclozapine Plasma Level | Cotinine in Current Smokers | Unconditioned | Unconditioned | 0.967169 | Unconditional | 85.913 |
| Norclozapine Plasma Level | Creatinine | Unconditioned | Unconditioned | 0.999998 | Unconditional | 71.9653 |
| Norclozapine Plasma Level | Creatinine | Unconditioned | Unconditioned | 0.999998 | Unconditional | 54.7763 |
| Norclozapine Plasma Level | Creatinine | Unconditioned | Unconditioned | 0.999998 | Unconditional | 90.6127 |
| Norclozapine Plasma Level | Creatinine | Unconditioned | Unconditioned | 0.999998 | Unconditional | 71.1569 |
| Norclozapine Plasma Level | C Reactive Protein | Unconditioned | Unconditioned | 0.999998 | Unconditional | 21.7447 |
| Norclozapine Plasma Level | Cystatin C | Unconditioned | Unconditioned | 0.999998 | Unconditional | 28.9065 |
| Norclozapine Plasma Level | Cystatin C | Unconditioned | Unconditioned | 0.999998 | Unconditional | 47.2486 |
| Norclozapine Plasma Level | Fresh Fruit Consumption | Unconditioned | Unconditioned | 0.999987 | Unconditional | 18.5819 |
| Norclozapine Plasma Level | Gamma Glutamyl Transpeptidase | rs2926036 | rs113185623 | 0.95701 | Conditional Both | 55.5757 |
| Norclozapine Plasma Level | Impedance of Left Arm | Unconditioned | Unconditioned | 0.999854 | Unconditional | 18.7517 |
| Norclozapine Plasma Level | Impedance of Right Arm | Unconditioned | Unconditioned | 0.986011 | Unconditional | 16.9526 |
| Norclozapine Plasma Level | Mean Corpuscular Hemoglobin | rs2926036 | rs294776 | 0.994891 | Conditional Both | 62.8769 |
| Norclozapine Plasma Level | Mean Corpuscular Hemoglobin | rs2926036 | rs294776 | 0.994891 | Conditional Both | 62.8607 |
| Norclozapine Plasma Level | Mean Corpuscular Hemoglobin | rs2926036 | rs294776 | 0.994891 | Conditional Both | 62.8664 |
| Norclozapine Plasma Level | Mean Corpuscular Volume | rs1824264 | rs62298399 | 0.836111 | Conditional Both | 15.6027 |
| Norclozapine Plasma Level | Overall Healthy Diet | Unconditioned | Unconditioned | 0.999998 | Unconditional | 82.7179 |
| Norclozapine Plasma Level | Peak Expiratory Flow | Unconditioned | Unconditioned | 0.999873 | Unconditional | 16.7618 |
| Norclozapine Plasma Level | Peak Expiratory Flow | Unconditioned | Unconditioned | 0.892823 | Unconditional | 37.1155 |

| | | | | | | |
|---|---|---|---|---|---|---|
| Norclozapine Plasma Level | Potassium in Urine | Unconditioned | Unconditioned | 0.999834 | Unconditional | 16.1192 |
| Norclozapine Plasma Level | Red Blood Cell Distribution Width | rs1824264 | rs10020490 | 0.937493 | Conditional Both | 13.4812 |
| Norclozapine Plasma Level | Red Blood Cell Distribution Width | rs1824264 | rs10020490 | 0.937618 | Conditional Both | 13.482 |
| Norclozapine Plasma Level | Red Blood Cell Distribution Width | rs1824264 | rs10020490 | 0.937688 | Conditional Both | 13.4821 |
| Norclozapine Plasma Level | Red Blood Cell Distribution Width | rs2926036 | rs10020490 | 0.979931 | Conditional Both | 63.2371 |
| Norclozapine Plasma Level | Serum Phosphate | Unconditioned | Unconditioned | 0.946806 | Unconditional | 49.9442 |
| Norclozapine Plasma Level | Serum Urea | Unconditioned | Unconditioned | 0.999998 | Unconditional | 65.7065 |
| Norclozapine Plasma Level | Serum Uric Acid | Unconditioned | Unconditioned | 0.999998 | Unconditional | 47.2079 |
| Norclozapine Plasma Level | Sodium in Urine | Unconditioned | Unconditioned | 0.999985 | Unconditional | 29.0183 |
| Norclozapine Plasma Level | Sodium in Urine | Unconditioned | Unconditioned | 0.999997 | Unconditional | 41.9587 |
| Norclozapine Plasma Level | Tea Consumption | Unconditioned | Unconditioned | 0.999998 | Unconditional | 48.6422 |
| Norclozapine Plasma Level | Tea Consumption | Unconditioned | Unconditioned | 0.999998 | Unconditional | 79.613 |
| Norclozapine Plasma Level | Tea Consumption | Unconditioned | Unconditioned | 0.999998 | Unconditional | 246.759 |
| Norclozapine Plasma Level | Total Testosterone | Unconditioned | Unconditioned | 0.966643 | Unconditional | 39.3702 |
| Norclozapine Plasma Level | Total Testosterone | Unconditioned | Unconditioned | 0.984377 | Unconditional | 37.3405 |
| Norclozapine Plasma Level | Total Testosterone | Unconditioned | Unconditioned | 0.95724 | Unconditional | 45.4847 |
| Norclozapine Plasma Level | Total Testosterone | Unconditioned | Unconditioned | 0.9679 | Unconditional | 24.8059 |
| Norclozapine Plasma Level | Total Testosterone | Unconditioned | Unconditioned | 0.960195 | Unconditional | 48.5317 |
| Norclozapine Plasma Level | Total Testosterone | Unconditioned | Unconditioned | 0.960195 | Unconditional | 48.5317 |
| Norclozapine Plasma Level | Total Testosterone | Unconditioned | Unconditioned | 0.964927 | Unconditional | 24.2381 |
| Norclozapine Plasma Level | Total Testosterone | Unconditioned | Unconditioned | 0.976717 | Unconditional | 43.8151 |
| Norclozapine Plasma Level | Total Testosterone | Unconditioned | Unconditioned | 0.965848 | Unconditional | 29.3186 |

| | | | | | | |
|---|---|---|---|---|---|---|
| Norclozapine Plasma Level | Total Testosterone | Unconditioned | Unconditioned | 0.968346 | Unconditional | 61.2868 |
| Norclozapine Plasma Level | Urate | Unconditioned | Unconditioned | 0.999998 | Unconditional | 47.3742 |
| Norclozapine Plasma Level | Urate | Unconditioned | Unconditioned | 0.999998 | Unconditional | 110.067 |
| Norclozapine Plasma Level | Urate | Unconditioned | Unconditioned | 0.999998 | Unconditional | 54.291 |
| Norclozapine Plasma Level | Urea | Unconditioned | Unconditioned | 0.999998 | Unconditional | 46.8845 |
| Norclozapine Plasma Level | Urea | Unconditioned | Unconditioned | 0.999998 | Unconditional | 54.8205 |
| Norclozapine Plasma Level | Urinary Albumin Excretion | Unconditioned | Unconditioned | 0.999718 | Unconditional | 46.5503 |
| Norclozapine Plasma Level | Water Consumption | Unconditioned | Unconditioned | 0.999989 | Unconditional | 31.3833 |
| Norclozapine Plasma Level | Water Consumption | Unconditioned | Unconditioned | 0.999998 | Unconditional | 177.792 |
| Norclozapine Plasma Level | Water Consumption | Unconditioned | Unconditioned | 0.999998 | Unconditional | 53.7716 |
| Norclozapine Plasma Level | Water Consumption | Unconditioned | Unconditioned | 0.999998 | Unconditional | 131.942 |
| Norclozapine Plasma Level | X 11440 | Unconditioned | Unconditioned | 0.978314 | Unconditional | 28.3318 |

Supplementary Table 5 - Candidate Biomarkers' Effects on Cardiometabolic Outcomes (MR)

| Exposure | Outcome | Method | nSNP | Beta | SE | P-Value | F Stat (Mean) | F Stat (Min) | Q Stat | Q Stat P-Value | I^2 | Egger Intercept | Egger Intercept SE | Egger Intercept P-Value |
|---|---|---|---|---|---|---|---|---|---|---|---|---|---|---|
| 25 Hydroxyvitamin D | BMI | IVW | 137 | 1.0E-01 | 5.2E-02 | 4.6E-02 | 3.5E+01 | 2.5E+01 | 1.7E+03 | 2.3E-269 | 9.2E+01 | NA | NA | NA |
| 25 Hydroxyvitamin D | BMI | MR Egger | 137 | -1.6E-01 | 1.2E-01 | 1.7E-01 | 3.5E+01 | 2.5E+01 | NA | NA | NA | 4.5E-03 | 1.8E-03 | 1.2E-02 |
| 25 Hydroxyvitamin D | BMI | WMed | 137 | -3.2E-02 | 2.7E-02 | 2.4E-01 | 3.5E+01 | 2.5E+01 | NA | NA | NA | NA | NA | NA |
| 25 Hydroxyvitamin D | BMI | WMode | 137 | -6.1E-02 | 3.4E-02 | 7.8E-02 | 3.5E+01 | 2.5E+01 | NA | NA | NA | NA | NA | NA |
| 25 Hydroxyvitamin D | DBP | IVW | 131 | -1.2E-01 | 3.5E-01 | 7.4E-01 | 3.4E+01 | 2.5E+01 | 8.6E+02 | 8.2E-109 | 8.5E+01 | NA | NA | NA |
| 25 Hydroxyvitamin D | DBP | MR Egger | 131 | -6.0E-03 | 8.5E-01 | 9.9E-01 | 3.4E+01 | 2.5E+01 | NA | NA | NA | -1.8E-03 | 1.3E-02 | 8.9E-01 |
| 25 Hydroxyvitamin D | DBP | WMed | 131 | 1.9E-01 | 2.4E-01 | 4.4E-01 | 3.4E+01 | 2.5E+01 | NA | NA | NA | NA | NA | NA |
| 25 Hydroxyvitamin D | DBP | WMode | 131 | -2.8E-01 | 4.1E-01 | 5.0E-01 | 3.4E+01 | 2.5E+01 | NA | NA | NA | NA | NA | NA |
| 25 Hydroxyvitamin D | HbA1c | IVW | 49 | -1.9E-02 | 5.1E-02 | 7.1E-01 | 3.8E+01 | 2.5E+01 | 7.1E+01 | 1.7E-02 | 3.2E+01 | NA | NA | NA |
| 25 Hydroxyvitamin D | HbA1c | MR Egger | 49 | -8.5E-02 | 1.2E-01 | 4.8E-01 | 3.8E+01 | 2.5E+01 | NA | NA | NA | 1.1E-03 | 1.8E-03 | 5.5E-01 |
| 25 Hydroxyvitamin D | HbA1c | WMed | 49 | -2.7E-02 | 7.1E-02 | 7.0E-01 | 3.8E+01 | 2.5E+01 | NA | NA | NA | NA | NA | NA |
| 25 Hydroxyvitamin D | HbA1c | WMode | 49 | -2.8E-02 | 8.4E-02 | 7.4E-01 | 3.8E+01 | 2.5E+01 | NA | NA | NA | NA | NA | NA |
| 25 Hydroxyvitamin D | HDL | IVW | 57 | 3.2E-01 | 3.2E-01 | 3.1E-01 | 3.7E+01 | 2.5E+01 | 2.2E+03 | 0.0E+00 | 9.7E+01 | NA | NA | NA |

| | | | | | | | | | | | | | | |
|---|---|---|---|---|---|---|---|---|---|---|---|---|---|---|
| 25 Hydroxyvitamin D | HDL | MR Egger | 57 | -4.2E-02 | 8.0E-01 | 9.6E-01 | 3.7E+01 | 2.5E+01 | NA | NA | NA | 5.7E-03 | 1.2E-02 | 6.3E-01 |
| 25 Hydroxyvitamin D | HDL | WMed | 57 | -2.3E-01 | 9.4E-02 | 1.6E-02 | 3.7E+01 | 2.5E+01 | NA | NA | NA | NA | NA | NA |
| 25 Hydroxyvitamin D | HDL | WMode | 57 | -2.8E-01 | 1.2E-01 | 1.7E-02 | 3.7E+01 | 2.5E+01 | NA | NA | NA | NA | NA | NA |
| 25 Hydroxyvitamin D | LDL | IVW | 57 | 4.3E-01 | 2.4E-01 | 7.6E-02 | 3.7E+01 | 2.5E+01 | 1.1E+03 | 1.1E-187 | 9.5E+01 | NA | NA | NA |
| 25 Hydroxyvitamin D | LDL | MR Egger | 57 | 1.2E+00 | 6.0E-01 | 5.1E-02 | 3.7E+01 | 2.5E+01 | NA | NA | NA | -1.2E-02 | 8.6E-03 | 1.7E-01 |
| 25 Hydroxyvitamin D | LDL | WMed | 57 | 9.7E-02 | 1.1E-01 | 3.6E-01 | 3.7E+01 | 2.5E+01 | NA | NA | NA | NA | NA | NA |
| 25 Hydroxyvitamin D | LDL | WMode | 57 | 6.2E-02 | 1.2E-01 | 6.0E-01 | 3.7E+01 | 2.5E+01 | NA | NA | NA | NA | NA | NA |
| 25 Hydroxyvitamin D | MetS | IVW | 145 | -2.6E-01 | 1.3E-01 | 4.8E-02 | 4.6E+01 | 2.5E+01 | 1.4E+03 | 5.0E-211 | 9.0E+01 | NA | NA | NA |
| 25 Hydroxyvitamin D | MetS | MR Egger | 145 | 1.6E-01 | 2.2E-01 | 4.7E-01 | 4.6E+01 | 2.5E+01 | NA | NA | NA | -8.9E-03 | 3.9E-03 | 2.4E-02 |
| 25 Hydroxyvitamin D | MetS | WMed | 145 | 7.4E-02 | 7.8E-02 | 3.4E-01 | 4.6E+01 | 2.5E+01 | NA | NA | NA | NA | NA | NA |
| 25 Hydroxyvitamin D | MetS | WMode | 145 | 5.9E-02 | 7.6E-02 | 4.4E-01 | 4.6E+01 | 2.5E+01 | NA | NA | NA | NA | NA | NA |
| 25 Hydroxyvitamin D | SBP | IVW | 130 | -2.0E-01 | 5.9E-01 | 7.3E-01 | 3.4E+01 | 2.5E+01 | 8.0E+02 | 8.8E-97 | 8.4E+01 | NA | NA | NA |
| 25 Hydroxyvitamin D | SBP | MR Egger | 130 | 4.7E-01 | 1.4E+00 | 7.4E-01 | 3.4E+01 | 2.5E+01 | NA | NA | NA | -1.1E-02 | 2.1E-02 | 6.1E-01 |
| 25 Hydroxyvitamin D | SBP | WMed | 130 | -6.7E-01 | 4.7E-01 | 1.5E-01 | 3.4E+01 | 2.5E+01 | NA | NA | NA | NA | NA | NA |
| 25 Hydroxyvitamin D | SBP | WMode | 130 | -7.6E-01 | 7.1E-01 | 2.8E-01 | 3.4E+01 | 2.5E+01 | NA | NA | NA | NA | NA | NA |

| | | | | | | | | | | | | | | |
|---|---|---|---|---|---|---|---|---|---|---|---|---|---|---|
| 25 Hydroxyvitamin D | T2D | IVW | 78 | 5.2E-01 | 1.5E-01 | 6.1E-04 | 3.1E+01 | 2.5E+01 | 2.7E+02 | 8.5E-23 | 7.1E+01 | NA | NA | NA |
| 25 Hydroxyvitamin D | T2D | MR Egger | 78 | -4.8E-02 | 6.5E-01 | 9.4E-01 | 3.1E+01 | 2.5E+01 | NA | NA | NA | 7.6E-03 | 8.4E-03 | 3.7E-01 |
| 25 Hydroxyvitamin D | T2D | WMed | 78 | 2.5E-01 | 1.5E-01 | 8.6E-02 | 3.1E+01 | 2.5E+01 | NA | NA | NA | NA | NA | NA |
| 25 Hydroxyvitamin D | T2D | WMode | 78 | -3.5E-02 | 3.7E-01 | 9.3E-01 | 3.1E+01 | 2.5E+01 | NA | NA | NA | NA | NA | NA |
| 25 Hydroxyvitamin D | Triglycerides | IVW | 57 | 5.1E-01 | 1.6E-01 | 1.7E-03 | 3.7E+01 | 2.5E+01 | 6.1E+02 | 9.8E-94 | 9.1E+01 | NA | NA | NA |
| 25 Hydroxyvitamin D | Triglycerides | MR Egger | 57 | 2.7E-01 | 4.0E-01 | 5.0E-01 | 3.7E+01 | 2.5E+01 | NA | NA | NA | 3.7E-03 | 5.9E-03 | 5.3E-01 |
| 25 Hydroxyvitamin D | Triglycerides | WMed | 57 | 2.0E-01 | 8.5E-02 | 2.2E-02 | 3.7E+01 | 2.5E+01 | NA | NA | NA | NA | NA | NA |
| 25 Hydroxyvitamin D | Triglycerides | WMode | 57 | 1.4E-01 | 1.0E-01 | 1.7E-01 | 3.7E+01 | 2.5E+01 | NA | NA | NA | NA | NA | NA |
| Bilirubin | BMI | IVW | 178 | 3.7E-03 | 8.4E-03 | 6.6E-01 | 8.0E+02 | 2.0E+01 | 1.1E+03 | 4.4E-141 | 8.4E+01 | NA | NA | NA |
| Bilirubin | BMI | MR Egger | 178 | 4.8E-04 | 9.2E-03 | 9.6E-01 | 8.0E+02 | 2.0E+01 | NA | NA | NA | 4.1E-04 | 5.0E-04 | 4.0E-01 |
| Bilirubin | BMI | WMed | 178 | -2.4E-03 | 3.6E-03 | 5.0E-01 | 8.0E+02 | 2.0E+01 | NA | NA | NA | NA | NA | NA |
| Bilirubin | BMI | WMode | 178 | -3.9E-03 | 4.3E-03 | 3.7E-01 | 8.0E+02 | 2.0E+01 | NA | NA | NA | NA | NA | NA |
| Bilirubin | DBP | IVW | 162 | -8.4E-02 | 1.2E-01 | 4.9E-01 | 8.8E+02 | 2.0E+01 | 2.9E+03 | 0.0E+00 | 9.4E+01 | NA | NA | NA |
| Bilirubin | DBP | MR Egger | 162 | 2.8E-02 | 1.3E-01 | 8.4E-01 | 8.8E+02 | 2.0E+01 | NA | NA | NA | -1.5E-02 | 7.4E-03 | 4.2E-02 |
| Bilirubin | DBP | WMed | 162 | 1.3E-02 | 3.3E-02 | 6.9E-01 | 8.8E+02 | 2.0E+01 | NA | NA | NA | NA | NA | NA |
| Bilirubin | DBP | WMode | 162 | 3.5E-02 | 3.9E-02 | 3.6E-01 | 8.8E+02 | 2.0E+01 | NA | NA | NA | NA | NA | NA |
| Bilirubin | HbA1c | IVW | 108 | 3.1E-02 | 1.5E-02 | 3.3E-02 | 1.2E+03 | 2.5E+01 | 6.8E+02 | 8.1E-85 | 8.2E+01 | NA | NA | NA |
| Bilirubin | HbA1c | MR Egger | 108 | 7.5E-03 | 1.5E-02 | 6.1E-01 | 1.2E+03 | 2.5E+01 | NA | NA | NA | 4.2E-03 | 9.8E-04 | 4.2E-05 |
| Bilirubin | HbA1c | WMed | 108 | 3.7E-03 | 6.3E-03 | 5.6E-01 | 1.2E+03 | 2.5E+01 | NA | NA | NA | NA | NA | NA |

| | | | | | | | | | | | | | | |
|---|---|---|---|---|---|---|---|---|---|---|---|---|---|---|
| Bilirubin | HbA1c | WMode | 108 | 1.6E-03 | 6.3E-03 | 8.1E-01 | 1.2E+03 | 2.5E+01 | NA | NA | NA | NA | NA | NA |
| Bilirubin | HDL | IVW | 119 | 1.2E-03 | 2.0E-02 | 9.5E-01 | 1.1E+03 | 2.4E+01 | 1.3E+03 | 5.3E-203 | 9.1E+01 | NA | NA | NA |
| Bilirubin | HDL | MR Egger | 119 | -1.2E-03 | 2.2E-02 | 9.6E-01 | 1.1E+03 | 2.4E+01 | NA | NA | NA | 4.9E-04 | 1.7E-03 | 7.7E-01 |
| Bilirubin | HDL | WMed | 119 | 3.4E-04 | 6.2E-03 | 9.6E-01 | 1.1E+03 | 2.4E+01 | NA | NA | NA | NA | NA | NA |
| Bilirubin | HDL | WMode | 119 | -3.1E-03 | 7.6E-03 | 6.8E-01 | 1.1E+03 | 2.4E+01 | NA | NA | NA | NA | NA | NA |
| Bilirubin | LDL | IVW | 119 | 4.2E-02 | 2.3E-02 | 7.2E-02 | 1.1E+03 | 2.4E+01 | 1.5E+03 | 2.7E-244 | 9.2E+01 | NA | NA | NA |
| Bilirubin | LDL | MR Egger | 119 | 4.5E-02 | 2.5E-02 | 7.5E-02 | 1.1E+03 | 2.4E+01 | NA | NA | NA | -7.1E-04 | 1.9E-03 | 7.1E-01 |
| Bilirubin | LDL | WMed | 119 | 3.3E-02 | 6.7E-03 | 7.8E-07 | 1.1E+03 | 2.4E+01 | NA | NA | NA | NA | NA | NA |
| Bilirubin | LDL | WMode | 119 | 4.1E-02 | 8.8E-03 | 9.9E-06 | 1.1E+03 | 2.4E+01 | NA | NA | NA | NA | NA | NA |
| Bilirubin | MetS | IVW | 180 | 3.0E-03 | 3.1E-02 | 9.2E-01 | 7.9E+02 | 2.0E+01 | 1.4E+03 | 4.8E-191 | 8.7E+01 | NA | NA | NA |
| Bilirubin | MetS | MR Egger | 180 | 7.9E-03 | 3.4E-02 | 8.2E-01 | 7.9E+02 | 2.0E+01 | NA | NA | NA | -6.2E-04 | 1.8E-03 | 7.3E-01 |
| Bilirubin | MetS | WMed | 180 | 1.5E-03 | 1.2E-02 | 9.0E-01 | 7.9E+02 | 2.0E+01 | NA | NA | NA | NA | NA | NA |
| Bilirubin | MetS | WMode | 180 | 1.0E-02 | 1.4E-02 | 4.7E-01 | 7.9E+02 | 2.0E+01 | NA | NA | NA | NA | NA | NA |
| Bilirubin | SBP | IVW | 160 | -4.9E-03 | 1.7E-01 | 9.8E-01 | 8.9E+02 | 2.0E+01 | 1.7E+03 | 2.0E-264 | 9.1E+01 | NA | NA | NA |
| Bilirubin | SBP | MR Egger | 160 | 7.5E-02 | 1.8E-01 | 6.8E-01 | 8.9E+02 | 2.0E+01 | NA | NA | NA | -1.1E-02 | 1.0E-02 | 2.8E-01 |
| Bilirubin | SBP | WMed | 160 | 6.5E-02 | 5.5E-02 | 2.4E-01 | 8.9E+02 | 2.0E+01 | NA | NA | NA | NA | NA | NA |
| Bilirubin | SBP | WMode | 160 | 9.4E-02 | 7.0E-02 | 1.8E-01 | 8.9E+02 | 2.0E+01 | NA | NA | NA | NA | NA | NA |
| Bilirubin | T2D | IVW | 131 | -3.6E-02 | 2.6E-02 | 1.6E-01 | 1.0E+03 | 2.4E+01 | 5.8E+02 | 1.6E-58 | 7.8E+01 | NA | NA | NA |
| Bilirubin | T2D | MR Egger | 131 | -3.8E-02 | 2.8E-02 | 1.8E-01 | 1.0E+03 | 2.4E+01 | NA | NA | NA | 2.7E-04 | 1.8E-03 | 8.8E-01 |
| Bilirubin | T2D | WMed | 131 | -3.8E-02 | 1.3E-02 | 3.4E-03 | 1.0E+03 | 2.4E+01 | NA | NA | NA | NA | NA | NA |
| Bilirubin | T2D | WMode | 131 | -2.9E-02 | 1.5E-02 | 5.2E-02 | 1.0E+03 | 2.4E+01 | NA | NA | NA | NA | NA | NA |
| Bilirubin | Triglycerides | IVW | 119 | 2.1E-02 | 2.2E-02 | 3.5E-01 | 1.1E+03 | 2.4E+01 | 1.7E+03 | 2.6E-273 | 9.3E+01 | NA | NA | NA |
| Bilirubin | Triglycerides | MR Egger | 119 | 2.2E-02 | 2.4E-02 | 3.5E-01 | 1.1E+03 | 2.4E+01 | NA | NA | NA | -3.1E-04 | 1.8E-03 | 8.7E-01 |

| | | | | | | | | | | | | | | |
|---|---|---|---|---|---|---|---|---|---|---|---|---|---|---|
| Bilirubin | Triglycerides | WMed | 119 | 1.7E-02 | 6.2E-03 | 5.6E-03 | 1.1E+03 | 2.4E+01 | NA | NA | NA | NA | NA | NA |
| Bilirubin | Triglycerides | WMode | 119 | 5.2E-03 | 8.3E-03 | 5.3E-01 | 1.1E+03 | 2.4E+01 | NA | NA | NA | NA | NA | NA |
| Blood Urea Nitrogen | BMI | IVW | 147 | -1.3E-02 | 3.6E-02 | 7.2E-01 | 9.2E+01 | 2.9E+01 | 2.5E+03 | 0.0E+00 | 9.4E+01 | NA | NA | NA |
| Blood Urea Nitrogen | BMI | MR Egger | 147 | 1.4E-02 | 7.9E-02 | 8.6E-01 | 9.2E+01 | 2.9E+01 | NA | NA | NA | -6.7E-04 | 1.8E-03 | 7.0E-01 |
| Blood Urea Nitrogen | BMI | WMed | 147 | 3.3E-02 | 1.7E-02 | 6.1E-02 | 9.2E+01 | 2.9E+01 | NA | NA | NA | NA | NA | NA |
| Blood Urea Nitrogen | BMI | WMode | 147 | 3.4E-02 | 2.3E-02 | 1.4E-01 | 9.2E+01 | 2.9E+01 | NA | NA | NA | NA | NA | NA |
| Blood Urea Nitrogen | DBP | IVW | 133 | -6.0E-01 | 3.2E-01 | 5.9E-02 | 9.2E+01 | 2.9E+01 | 2.1E+03 | 0.0E+00 | 9.4E+01 | NA | NA | NA |
| Blood Urea Nitrogen | DBP | MR Egger | 133 | -4.2E-01 | 6.8E-01 | 5.3E-01 | 9.2E+01 | 2.9E+01 | NA | NA | NA | -4.4E-03 | 1.5E-02 | 7.7E-01 |
| Blood Urea Nitrogen | DBP | WMed | 133 | -3.9E-01 | 1.9E-01 | 3.8E-02 | 9.2E+01 | 2.9E+01 | NA | NA | NA | NA | NA | NA |
| Blood Urea Nitrogen | DBP | WMode | 133 | 4.6E-02 | 2.8E-01 | 8.7E-01 | 9.2E+01 | 2.9E+01 | NA | NA | NA | NA | NA | NA |
| Blood Urea Nitrogen | HbA1c | IVW | 53 | -4.1E-02 | 2.7E-02 | 1.3E-01 | 1.0E+02 | 2.9E+01 | 6.4E+01 | 1.2E-01 | 1.8E+01 | NA | NA | NA |
| Blood Urea Nitrogen | HbA1c | MR Egger | 53 | -1.0E-01 | 6.9E-02 | 1.6E-01 | 1.0E+02 | 2.9E+01 | NA | NA | NA | 1.4E-03 | 1.5E-03 | 3.6E-01 |
| Blood Urea Nitrogen | HbA1c | WMed | 53 | -6.5E-02 | 3.9E-02 | 9.2E-02 | 1.0E+02 | 2.9E+01 | NA | NA | NA | NA | NA | NA |
| Blood Urea Nitrogen | HbA1c | WMode | 53 | -1.2E-01 | 6.2E-02 | 6.2E-02 | 1.0E+02 | 2.9E+01 | NA | NA | NA | NA | NA | NA |
| Blood Urea Nitrogen | HDL | IVW | 60 | -8.0E-02 | 5.0E-02 | 1.1E-01 | 1.1E+02 | 2.9E+01 | 1.9E+02 | 1.7E-15 | 6.8E+01 | NA | NA | NA |

| | | | | | | | | | | | | | | |
|---|---|---|---|---|---|---|---|---|---|---|---|---|---|---|
| Blood Urea Nitrogen | HDL | MR Egger | 60 | -1.9E-01 | 1.3E-01 | 1.4E-01 | 1.1E+02 | 2.9E+01 | NA | NA | NA | 2.8E-03 | 2.9E-03 | 3.4E-01 |
| Blood Urea Nitrogen | HDL | WMed | 60 | -8.9E-02 | 4.9E-02 | 6.9E-02 | 1.1E+02 | 2.9E+01 | NA | NA | NA | NA | NA | NA |
| Blood Urea Nitrogen | HDL | WMode | 60 | -7.3E-02 | 6.0E-02 | 2.3E-01 | 1.1E+02 | 2.9E+01 | NA | NA | NA | NA | NA | NA |
| Blood Urea Nitrogen | LDL | IVW | 60 | -5.5E-02 | 6.6E-02 | 4.0E-01 | 1.1E+02 | 2.9E+01 | 2.8E+02 | 3.2E-30 | 7.9E+01 | NA | NA | NA |
| Blood Urea Nitrogen | LDL | MR Egger | 60 | -1.4E-01 | 1.7E-01 | 4.0E-01 | 1.1E+02 | 2.9E+01 | NA | NA | NA | 2.2E-03 | 3.9E-03 | 5.8E-01 |
| Blood Urea Nitrogen | LDL | WMed | 60 | -8.8E-02 | 5.5E-02 | 1.1E-01 | 1.1E+02 | 2.9E+01 | NA | NA | NA | NA | NA | NA |
| Blood Urea Nitrogen | LDL | WMode | 60 | -6.7E-02 | 5.7E-02 | 2.4E-01 | 1.1E+02 | 2.9E+01 | NA | NA | NA | NA | NA | NA |
| Blood Urea Nitrogen | MetS | IVW | 166 | 1.1E-01 | 5.4E-02 | 3.4E-02 | 8.8E+01 | 2.9E+01 | 6.4E+02 | 1.5E-57 | 7.4E+01 | NA | NA | NA |
| Blood Urea Nitrogen | MetS | MR Egger | 166 | 1.8E-02 | 1.2E-01 | 8.8E-01 | 8.8E+01 | 2.9E+01 | NA | NA | NA | 2.3E-03 | 2.6E-03 | 3.7E-01 |
| Blood Urea Nitrogen | MetS | WMed | 166 | 1.3E-02 | 4.8E-02 | 7.9E-01 | 8.8E+01 | 2.9E+01 | NA | NA | NA | NA | NA | NA |
| Blood Urea Nitrogen | MetS | WMode | 166 | -2.7E-02 | 6.4E-02 | 6.7E-01 | 8.8E+01 | 2.9E+01 | NA | NA | NA | NA | NA | NA |
| Blood Urea Nitrogen | SBP | IVW | 132 | -1.1E+00 | 5.2E-01 | 4.2E-02 | 9.2E+01 | 2.9E+01 | 1.9E+03 | 3.2E-303 | 9.3E+01 | NA | NA | NA |
| Blood Urea Nitrogen | SBP | MR Egger | 132 | -4.2E-01 | 1.1E+00 | 7.1E-01 | 9.2E+01 | 2.9E+01 | NA | NA | NA | -1.6E-02 | 2.5E-02 | 5.2E-01 |
| Blood Urea Nitrogen | SBP | WMed | 132 | -2.8E-01 | 3.4E-01 | 4.1E-01 | 9.2E+01 | 2.9E+01 | NA | NA | NA | NA | NA | NA |
| Blood Urea Nitrogen | SBP | WMode | 132 | 3.8E-01 | 4.0E-01 | 3.4E-01 | 9.2E+01 | 2.9E+01 | NA | NA | NA | NA | NA | NA |

| | | | | | | | | | | | | | | |
|---|---|---|---|---|---|---|---|---|---|---|---|---|---|---|
| Blood Urea Nitrogen | T2D | IVW | 105 | -7.4E-03 | 1.0E-01 | 9.4E-01 | 9.6E+01 | 2.9E+01 | 7.2E+02 | 3.6E-93 | 8.6E+01 | NA | NA | NA |
| Blood Urea Nitrogen | T2D | MR Egger | 105 | 8.0E-02 | 2.3E-01 | 7.3E-01 | 9.6E+01 | 2.9E+01 | NA | NA | NA | -2.2E-03 | 5.0E-03 | 6.7E-01 |
| Blood Urea Nitrogen | T2D | WMed | 105 | 1.5E-01 | 7.2E-02 | 3.6E-02 | 9.6E+01 | 2.9E+01 | NA | NA | NA | NA | NA | NA |
| Blood Urea Nitrogen | T2D | WMode | 105 | 1.4E-01 | 8.2E-02 | 8.6E-02 | 9.6E+01 | 2.9E+01 | NA | NA | NA | NA | NA | NA |
| Blood Urea Nitrogen | Triglycerides | IVW | 60 | -1.7E-01 | 1.3E-01 | 2.1E-01 | 1.1E+02 | 2.9E+01 | 1.4E+03 | 2.8E-258 | 9.6E+01 | NA | NA | NA |
| Blood Urea Nitrogen | Triglycerides | MR Egger | 60 | 2.1E-01 | 3.3E-01 | 5.3E-01 | 1.1E+02 | 2.9E+01 | NA | NA | NA | -9.7E-03 | 7.8E-03 | 2.2E-01 |
| Blood Urea Nitrogen | Triglycerides | WMed | 60 | 0.0E+00 | 4.8E-02 | 1.0E+00 | 1.1E+02 | 2.9E+01 | NA | NA | NA | NA | NA | NA |
| Blood Urea Nitrogen | Triglycerides | WMode | 60 | -1.9E-02 | 6.0E-02 | 7.5E-01 | 1.1E+02 | 2.9E+01 | NA | NA | NA | NA | NA | NA |
| Coffee Consumption | BMI | IVW | 35 | -7.5E-01 | 1.9E-01 | 7.8E-05 | 7.8E+01 | 3.0E+01 | 2.2E+03 | 0.0E+00 | 9.8E+01 | NA | NA | NA |
| Coffee Consumption | BMI | MR Egger | 35 | -4.4E-01 | 3.8E-01 | 2.5E-01 | 7.8E+01 | 3.0E+01 | NA | NA | NA | -6.1E-03 | 6.2E-03 | 3.4E-01 |
| Coffee Consumption | BMI | WMed | 35 | -2.8E-01 | 3.9E-02 | 2.3E-12 | 7.8E+01 | 3.0E+01 | NA | NA | NA | NA | NA | NA |
| Coffee Consumption | BMI | WMode | 35 | -2.8E-01 | 3.3E-02 | 4.1E-10 | 7.8E+01 | 3.0E+01 | NA | NA | NA | NA | NA | NA |
| Coffee Consumption | DBP | IVW | 33 | 1.3E+00 | 5.0E-01 | 7.9E-03 | 8.0E+01 | 3.0E+01 | 1.8E+02 | 7.6E-23 | 8.2E+01 | NA | NA | NA |
| Coffee Consumption | DBP | MR Egger | 33 | 1.4E+00 | 1.0E+00 | 1.8E-01 | 8.0E+01 | 3.0E+01 | NA | NA | NA | -7.4E-04 | 1.7E-02 | 9.6E-01 |
| Coffee Consumption | DBP | WMed | 33 | 1.4E+00 | 5.0E-01 | 3.7E-03 | 8.0E+01 | 3.0E+01 | NA | NA | NA | NA | NA | NA |

| | | | | | | | | | | | | | | |
|---|---|---|---|---|---|---|---|---|---|---|---|---|---|---|
| Coffee Consumption | DBP | WMode | 33 | 2.3E+00 | 8.0E-01 | 8.5E-03 | 8.0E+01 | 3.0E+01 | NA | NA | NA | NA | NA | NA |
| Coffee Consumption | HbA1c | IVW | 19 | -8.9E-02 | 5.7E-02 | 1.2E-01 | 1.0E+02 | 3.0E+01 | 2.1E+01 | 2.8E-01 | 8.4E+00 | NA | NA | NA |
| Coffee Consumption | HbA1c | MR Egger | 19 | 4.9E-03 | 1.1E-01 | 9.6E-01 | 1.0E+02 | 3.0E+01 | NA | NA | NA | -1.9E-03 | 1.8E-03 | 3.0E-01 |
| Coffee Consumption | HbA1c | WMed | 19 | -6.1E-02 | 6.8E-02 | 3.7E-01 | 1.0E+02 | 3.0E+01 | NA | NA | NA | NA | NA | NA |
| Coffee Consumption | HbA1c | WMode | 19 | -4.3E-02 | 7.2E-02 | 5.6E-01 | 1.0E+02 | 3.0E+01 | NA | NA | NA | NA | NA | NA |
| Coffee Consumption | HDL | IVW | 19 | 1.9E-01 | 1.5E-01 | 2.0E-01 | 1.1E+02 | 3.0E+01 | 1.2E+02 | 6.7E-17 | 8.5E+01 | NA | NA | NA |
| Coffee Consumption | HDL | MR Egger | 19 | 6.6E-02 | 2.8E-01 | 8.2E-01 | 1.1E+02 | 3.0E+01 | NA | NA | NA | 2.7E-03 | 5.3E-03 | 6.1E-01 |
| Coffee Consumption | HDL | WMed | 19 | -7.5E-02 | 1.1E-01 | 4.7E-01 | 1.1E+02 | 3.0E+01 | NA | NA | NA | NA | NA | NA |
| Coffee Consumption | HDL | WMode | 19 | 2.5E-02 | 9.3E-02 | 7.9E-01 | 1.1E+02 | 3.0E+01 | NA | NA | NA | NA | NA | NA |
| Coffee Consumption | LDL | IVW | 19 | -3.7E-02 | 1.1E-01 | 7.4E-01 | 1.1E+02 | 3.0E+01 | 6.1E+01 | 1.4E-06 | 7.0E+01 | NA | NA | NA |
| Coffee Consumption | LDL | MR Egger | 19 | -1.4E-01 | 2.2E-01 | 5.3E-01 | 1.1E+02 | 3.0E+01 | NA | NA | NA | 2.2E-03 | 4.1E-03 | 5.9E-01 |
| Coffee Consumption | LDL | WMed | 19 | -1.1E-01 | 8.4E-02 | 2.0E-01 | 1.1E+02 | 3.0E+01 | NA | NA | NA | NA | NA | NA |
| Coffee Consumption | LDL | WMode | 19 | -1.2E-01 | 7.3E-02 | 1.2E-01 | 1.1E+02 | 3.0E+01 | NA | NA | NA | NA | NA | NA |
| Coffee Consumption | MetS | IVW | 35 | 5.6E-01 | 2.9E-01 | 5.4E-02 | 7.8E+01 | 3.0E+01 | 4.7E+02 | 6.7E-78 | 9.3E+01 | NA | NA | NA |
| Coffee Consumption | MetS | MR Egger | 35 | 5.0E-01 | 5.8E-01 | 3.9E-01 | 7.8E+01 | 3.0E+01 | NA | NA | NA | 1.0E-03 | 9.5E-03 | 9.1E-01 |

| | | | | | | | | | | | | | | |
|---|---|---|---|---|---|---|---|---|---|---|---|---|---|---|
| Coffee Consumption | MetS | WMed | 35 | 5.0E-01 | 1.7E-01 | 3.9E-03 | 7.8E+01 | 3.0E+01 | NA | NA | NA | NA | NA | NA |
| Coffee Consumption | MetS | WMode | 35 | 4.1E-01 | 1.3E-01 | 3.4E-03 | 7.8E+01 | 3.0E+01 | NA | NA | NA | NA | NA | NA |
| Coffee Consumption | SBP | IVW | 33 | 1.9E+00 | 9.2E-01 | 4.0E-02 | 8.0E+01 | 3.0E+01 | 2.0E+02 | 1.3E-26 | 8.4E+01 | NA | NA | NA |
| Coffee Consumption | SBP | MR Egger | 33 | 3.0E-01 | 1.8E+00 | 8.7E-01 | 8.0E+01 | 3.0E+01 | NA | NA | NA | 3.0E-02 | 3.0E-02 | 3.2E-01 |
| Coffee Consumption | SBP | WMed | 33 | 2.3E+00 | 8.4E-01 | 7.3E-03 | 8.0E+01 | 3.0E+01 | NA | NA | NA | NA | NA | NA |
| Coffee Consumption | SBP | WMode | 33 | 8.2E-01 | 1.3E+00 | 5.4E-01 | 8.0E+01 | 3.0E+01 | NA | NA | NA | NA | NA | NA |
| Coffee Consumption | T2D | IVW | 26 | -1.5E+00 | 3.9E-01 | 1.3E-04 | 6.7E+01 | 3.0E+01 | 2.9E+02 | 1.9E-46 | 9.1E+01 | NA | NA | NA |
| Coffee Consumption | T2D | MR Egger | 26 | -1.6E+00 | 9.6E-01 | 1.0E-01 | 6.7E+01 | 3.0E+01 | NA | NA | NA | 2.1E-03 | 1.4E-02 | 8.8E-01 |
| Coffee Consumption | T2D | WMed | 26 | -4.7E-01 | 1.9E-01 | 1.5E-02 | 6.7E+01 | 3.0E+01 | NA | NA | NA | NA | NA | NA |
| Coffee Consumption | T2D | WMode | 26 | -3.7E-01 | 1.9E-01 | 6.4E-02 | 6.7E+01 | 3.0E+01 | NA | NA | NA | NA | NA | NA |
| Coffee Consumption | Triglycerides | IVW | 19 | 3.6E-01 | 4.7E-01 | 4.5E-01 | 1.1E+02 | 3.0E+01 | 1.3E+03 | 3.7E-267 | 9.9E+01 | NA | NA | NA |
| Coffee Consumption | Triglycerides | MR Egger | 19 | -2.5E-01 | 8.8E-01 | 7.8E-01 | 1.1E+02 | 3.0E+01 | NA | NA | NA | 1.4E-02 | 1.7E-02 | 4.2E-01 |
| Coffee Consumption | Triglycerides | WMed | 19 | 2.7E-03 | 1.1E-01 | 9.8E-01 | 1.1E+02 | 3.0E+01 | NA | NA | NA | NA | NA | NA |
| Coffee Consumption | Triglycerides | WMode | 19 | 4.3E-02 | 9.1E-02 | 6.4E-01 | 1.1E+02 | 3.0E+01 | NA | NA | NA | NA | NA | NA |
| Cotinine in Current Smokers | BMI | IVW | 3 | 1.4E-02 | 1.8E-02 | 4.5E-01 | 8.4E+01 | 4.6E+01 | 1.8E+01 | 1.2E-04 | 0.0E+00 | NA | NA | NA |

| | | | | | | | | | | | | | | |
|---|---|---|---|---|---|---|---|---|---|---|---|---|---|---|
| Cotinine in Current Smokers | BMI | MR Egger | 3 | -5.6E-02 | 1.7E-02 | 1.9E-01 | 8.4E+01 | 4.6E+01 | NA | NA | NA | 1.6E-02 | 3.8E-03 | 1.5E-01 |
| Cotinine in Current Smokers | BMI | WMed | 3 | 8.0E-03 | 7.5E-03 | 2.8E-01 | 8.4E+01 | 4.6E+01 | NA | NA | NA | NA | NA | NA |
| Cotinine in Current Smokers | BMI | WMode | 3 | 4.7E-03 | 8.2E-03 | 6.2E-01 | 8.4E+01 | 4.6E+01 | NA | NA | NA | NA | NA | NA |
| Cotinine in Current Smokers | DBP | IVW | 3 | -1.0E-01 | 1.2E-01 | 3.8E-01 | 8.4E+01 | 4.6E+01 | 9.0E+00 | 1.1E-02 | 0.0E+00 | NA | NA | NA |
| Cotinine in Current Smokers | DBP | MR Egger | 3 | 3.3E-01 | 1.7E-01 | 3.0E-01 | 8.4E+01 | 4.6E+01 | NA | NA | NA | -1.0E-01 | 3.7E-02 | 2.2E-01 |
| Cotinine in Current Smokers | DBP | WMed | 3 | -1.0E-01 | 7.5E-02 | 1.7E-01 | 8.4E+01 | 4.6E+01 | NA | NA | NA | NA | NA | NA |
| Cotinine in Current Smokers | DBP | WMode | 3 | -1.4E-01 | 1.1E-01 | 3.4E-01 | 8.4E+01 | 4.6E+01 | NA | NA | NA | NA | NA | NA |
| Cotinine in Current Smokers | MetS | IVW | 3 | 1.2E-02 | 2.0E-02 | 5.4E-01 | 8.4E+01 | 4.6E+01 | 6.6E-01 | 7.2E-01 | 0.0E+00 | NA | NA | NA |
| Cotinine in Current Smokers | MetS | MR Egger | 3 | 3.7E-02 | 5.7E-02 | 6.3E-01 | 8.4E+01 | 4.6E+01 | NA | NA | NA | -5.9E-03 | 1.3E-02 | 7.2E-01 |
| Cotinine in Current Smokers | MetS | WMed | 3 | 1.6E-02 | 2.2E-02 | 4.7E-01 | 8.4E+01 | 4.6E+01 | NA | NA | NA | NA | NA | NA |
| Cotinine in Current Smokers | MetS | WMode | 3 | 1.8E-02 | 2.6E-02 | 5.6E-01 | 8.4E+01 | 4.6E+01 | NA | NA | NA | NA | NA | NA |
| Cotinine in Current Smokers | SBP | IVW | 3 | -1.1E-01 | 1.4E-01 | 4.6E-01 | 8.4E+01 | 4.6E+01 | 4.5E+00 | 1.1E-01 | 0.0E+00 | NA | NA | NA |
| Cotinine in Current Smokers | SBP | MR Egger | 3 | 3.3E-01 | 3.9E-01 | 5.5E-01 | 8.4E+01 | 4.6E+01 | NA | NA | NA | -1.0E-01 | 8.5E-02 | 4.5E-01 |
| Cotinine in Current Smokers | SBP | WMed | 3 | -2.6E-01 | 1.2E-01 | 3.5E-02 | 8.4E+01 | 4.6E+01 | NA | NA | NA | NA | NA | NA |
| Cotinine in Current Smokers | SBP | WMode | 3 | -2.7E-01 | 2.0E-01 | 3.1E-01 | 8.4E+01 | 4.6E+01 | NA | NA | NA | NA | NA | NA |

| | | | | | | | | | | | | | | |
|---|---|---|---|---|---|---|---|---|---|---|---|---|---|---|
| Creatinine | BMI | IVW | 305 | -1.4E-01 | 2.6E-02 | 6.7E-08 | 8.9E+01 | 3.0E+01 | 5.0E+03 | 0.0E+00 | 9.3E+01 | NA | NA | NA |
| Creatinine | BMI | MR Egger | 305 | 1.2E-01 | 5.3E-02 | 2.7E-02 | 8.9E+01 | 3.0E+01 | NA | NA | NA | -6.2E-03 | 1.1E-03 | 1.0E-07 |
| Creatinine | BMI | WMed | 305 | -2.4E-02 | 1.3E-02 | 7.5E-02 | 8.9E+01 | 3.0E+01 | NA | NA | NA | NA | NA | NA |
| Creatinine | BMI | WMode | 305 | 3.0E-03 | 1.6E-02 | 8.5E-01 | 8.9E+01 | 3.0E+01 | NA | NA | NA | NA | NA | NA |
| Creatinine | DBP | IVW | 270 | -6.1E-01 | 2.5E-01 | 1.3E-02 | 9.2E+01 | 3.0E+01 | 4.7E+03 | 0.0E+00 | 9.4E+01 | NA | NA | NA |
| Creatinine | DBP | MR Egger | 270 | -1.5E+00 | 5.3E-01 | 5.0E-03 | 9.2E+01 | 3.0E+01 | NA | NA | NA | 2.1E-02 | 1.1E-02 | 6.0E-02 |
| Creatinine | DBP | WMed | 270 | -1.8E-01 | 1.4E-01 | 2.0E-01 | 9.2E+01 | 3.0E+01 | NA | NA | NA | NA | NA | NA |
| Creatinine | DBP | WMode | 270 | 2.8E-01 | 3.0E-01 | 3.5E-01 | 9.2E+01 | 3.0E+01 | NA | NA | NA | NA | NA | NA |
| Creatinine | HbA1c | IVW | 138 | 1.7E-02 | 1.9E-02 | 3.6E-01 | 1.0E+02 | 3.0E+01 | 1.9E+02 | 2.2E-03 | 2.7E+01 | NA | NA | NA |
| Creatinine | HbA1c | MR Egger | 138 | 4.1E-02 | 4.0E-02 | 3.0E-01 | 1.0E+02 | 3.0E+01 | NA | NA | NA | -6.0E-04 | 8.7E-04 | 4.9E-01 |
| Creatinine | HbA1c | WMed | 138 | 3.5E-03 | 2.6E-02 | 8.9E-01 | 1.0E+02 | 3.0E+01 | NA | NA | NA | NA | NA | NA |
| Creatinine | HbA1c | WMode | 138 | -3.9E-05 | 4.5E-02 | 1.0E+00 | 1.0E+02 | 3.0E+01 | NA | NA | NA | NA | NA | NA |
| Creatinine | HDL | IVW | 149 | 1.2E-01 | 4.8E-02 | 1.1E-02 | 9.8E+01 | 3.0E+01 | 8.9E+02 | 9.6E-106 | 8.3E+01 | NA | NA | NA |
| Creatinine | HDL | MR Egger | 149 | 1.8E-01 | 1.1E-01 | 9.1E-02 | 9.8E+01 | 3.0E+01 | NA | NA | NA | -1.5E-03 | 2.3E-03 | 5.3E-01 |
| Creatinine | HDL | WMed | 149 | 1.3E-01 | 3.8E-02 | 7.4E-04 | 9.8E+01 | 3.0E+01 | NA | NA | NA | NA | NA | NA |
| Creatinine | HDL | WMode | 149 | 8.2E-02 | 9.5E-02 | 3.9E-01 | 9.8E+01 | 3.0E+01 | NA | NA | NA | NA | NA | NA |
| Creatinine | LDL | IVW | 149 | 9.3E-02 | 5.3E-02 | 7.8E-02 | 9.8E+01 | 3.0E+01 | 9.0E+02 | 3.7E-108 | 8.4E+01 | NA | NA | NA |
| Creatinine | LDL | MR Egger | 149 | 3.1E-02 | 1.2E-01 | 7.9E-01 | 9.8E+01 | 3.0E+01 | NA | NA | NA | 1.5E-03 | 2.6E-03 | 5.6E-01 |
| Creatinine | LDL | WMed | 149 | 1.9E-02 | 4.1E-02 | 6.4E-01 | 9.8E+01 | 3.0E+01 | NA | NA | NA | NA | NA | NA |
| Creatinine | LDL | WMode | 149 | 1.9E-03 | 8.4E-02 | 9.8E-01 | 9.8E+01 | 3.0E+01 | NA | NA | NA | NA | NA | NA |
| Creatinine | MetS | IVW | 339 | 2.2E-01 | 4.4E-02 | 4.8E-07 | 9.6E+01 | 3.0E+01 | 1.8E+03 | 2.5E-196 | 8.1E+01 | NA | NA | NA |
| Creatinine | MetS | MR Egger | 339 | 5.0E-03 | 8.9E-02 | 9.6E-01 | 9.6E+01 | 3.0E+01 | NA | NA | NA | 5.5E-03 | 1.9E-03 | 5.0E-03 |
| Creatinine | MetS | WMed | 339 | 1.8E-01 | 3.8E-02 | 1.0E-06 | 9.6E+01 | 3.0E+01 | NA | NA | NA | NA | NA | NA |
| Creatinine | MetS | WMode | 339 | 6.1E-02 | 4.6E-02 | 1.8E-01 | 9.6E+01 | 3.0E+01 | NA | NA | NA | NA | NA | NA |

| | | | | | | | | | | | | | |
|---|---|---|---|---|---|---|---|---|---|---|---|---|---|
| Creatinine | SBP | IVW | 269 | -3.8E-01 | 3.8E-01 | 3.2E-01 | 9.2E+01 | 3.0E+01 | 3.6E+03 | 0.0E+00 | 9.2E+01 | NA | NA | NA |
| Creatinine | SBP | MR Egger | 269 | -1.5E+00 | 8.2E-01 | 7.7E-02 | 9.2E+01 | 3.0E+01 | NA | NA | NA | 2.6E-02 | 1.7E-02 | 1.4E-01 |
| Creatinine | SBP | WMed | 269 | 1.2E-01 | 2.5E-01 | 6.3E-01 | 9.2E+01 | 3.0E+01 | NA | NA | NA | NA | NA | NA |
| Creatinine | SBP | WMode | 269 | 1.1E+00 | 5.9E-01 | 6.1E-02 | 9.2E+01 | 3.0E+01 | NA | NA | NA | NA | NA | NA |
| Creatinine | T2D | IVW | 212 | -1.4E-01 | 6.4E-02 | 2.7E-02 | 9.5E+01 | 3.0E+01 | 1.0E+03 | 4.4E-107 | 7.9E+01 | NA | NA | NA |
| Creatinine | T2D | MR Egger | 212 | 2.1E-01 | 1.5E-01 | 1.5E-01 | 9.5E+01 | 3.0E+01 | NA | NA | NA | -8.2E-03 | 3.0E-03 | 7.7E-03 |
| Creatinine | T2D | WMed | 212 | 1.9E-02 | 5.8E-02 | 7.4E-01 | 9.5E+01 | 3.0E+01 | NA | NA | NA | NA | NA | NA |
| Creatinine | T2D | WMode | 212 | 4.8E-02 | 8.9E-02 | 5.9E-01 | 9.5E+01 | 3.0E+01 | NA | NA | NA | NA | NA | NA |
| Creatinine | Triglycerides | IVW | 149 | 5.2E-02 | 7.6E-02 | 4.9E-01 | 9.8E+01 | 3.0E+01 | 2.3E+03 | 0.0E+00 | 9.4E+01 | NA | NA | NA |
| Creatinine | Triglycerides | MR Egger | 149 | -1.3E-01 | 1.7E-01 | 4.4E-01 | 9.8E+01 | 3.0E+01 | NA | NA | NA | 4.4E-03 | 3.7E-03 | 2.3E-01 |
| Creatinine | Triglycerides | WMed | 149 | -1.4E-01 | 3.6E-02 | 1.3E-04 | 9.8E+01 | 3.0E+01 | NA | NA | NA | NA | NA | NA |
| Creatinine | Triglycerides | WMode | 149 | -1.2E-01 | 3.6E-02 | 6.2E-04 | 9.8E+01 | 3.0E+01 | NA | NA | NA | NA | NA | NA |
| CRP | BMI | IVW | 224 | -1.3E-03 | 1.2E-02 | 9.1E-01 | 2.5E+02 | 2.4E+01 | 1.9E+03 | 2.6E-257 | 8.8E+01 | NA | NA | NA |
| CRP | BMI | MR Egger | 224 | -3.1E-02 | 1.6E-02 | 4.8E-02 | 2.5E+02 | 2.4E+01 | NA | NA | NA | 1.8E-03 | 6.0E-04 | 3.5E-03 |
| CRP | BMI | WMed | 224 | -5.0E-03 | 8.5E-03 | 5.5E-01 | 2.5E+02 | 2.4E+01 | NA | NA | NA | NA | NA | NA |
| CRP | BMI | WMode | 224 | -2.2E-02 | 6.0E-03 | 2.4E-04 | 2.5E+02 | 2.4E+01 | NA | NA | NA | NA | NA | NA |
| CRP | DBP | IVW | 201 | -5.2E-02 | 1.2E-01 | 6.7E-01 | 2.7E+02 | 2.4E+01 | 2.0E+03 | 7.4E-292 | 9.0E+01 | NA | NA | NA |
| CRP | DBP | MR Egger | 201 | 2.0E-02 | 1.6E-01 | 9.0E-01 | 2.7E+02 | 2.4E+01 | NA | NA | NA | -4.3E-03 | 6.0E-03 | 4.7E-01 |
| CRP | DBP | WMed | 201 | -5.8E-02 | 7.5E-02 | 4.4E-01 | 2.7E+02 | 2.4E+01 | NA | NA | NA | NA | NA | NA |
| CRP | DBP | WMode | 201 | 1.2E-02 | 5.2E-02 | 8.2E-01 | 2.7E+02 | 2.4E+01 | NA | NA | NA | NA | NA | NA |
| CRP | HbA1c | IVW | 121 | 1.1E-02 | 1.3E-02 | 4.1E-01 | 2.8E+02 | 2.4E+01 | 2.3E+02 | 2.1E-08 | 4.6E+01 | NA | NA | NA |
| CRP | HbA1c | MR Egger | 121 | -1.4E-03 | 1.7E-02 | 9.4E-01 | 2.8E+02 | 2.4E+01 | NA | NA | NA | 7.3E-04 | 6.7E-04 | 2.8E-01 |

| | | | | | | | | | | | | | | |
|---|---|---|---|---|---|---|---|---|---|---|---|---|---|---|
| CRP | HbA1c | WMed | 121 | -9.9E-03 | 1.5E-02 | 5.1E-01 | 2.8E+02 | 2.4E+01 | NA | NA | NA | NA | NA | NA |
| CRP | HbA1c | WMode | 121 | -2.2E-03 | 1.2E-02 | 8.5E-01 | 2.8E+02 | 2.4E+01 | NA | NA | NA | NA | NA | NA |
| CRP | HDL | IVW | 130 | -3.4E-02 | 4.0E-02 | 3.9E-01 | 2.7E+02 | 2.4E+01 | 1.8E+03 | 1.9E-298 | 9.3E+01 | NA | NA | NA |
| CRP | HDL | MR Egger | 130 | 1.7E-02 | 5.1E-02 | 7.4E-01 | 2.7E+02 | 2.4E+01 | NA | NA | NA | -3.4E-03 | 2.2E-03 | 1.2E-01 |
| CRP | HDL | WMed | 130 | 5.3E-02 | 2.2E-02 | 1.6E-02 | 2.7E+02 | 2.4E+01 | NA | NA | NA | NA | NA | NA |
| CRP | HDL | WMode | 130 | 4.5E-02 | 1.4E-02 | 1.3E-03 | 2.7E+02 | 2.4E+01 | NA | NA | NA | NA | NA | NA |
| CRP | LDL | IVW | 130 | 2.9E-02 | 5.0E-02 | 5.6E-01 | 2.7E+02 | 2.4E+01 | 2.4E+03 | 0.0E+00 | 9.5E+01 | NA | NA | NA |
| CRP | LDL | MR Egger | 130 | -1.5E-02 | 6.4E-02 | 8.1E-01 | 2.7E+02 | 2.4E+01 | NA | NA | NA | 3.0E-03 | 2.8E-03 | 2.8E-01 |
| CRP | LDL | WMed | 130 | -7.4E-03 | 2.0E-02 | 7.1E-01 | 2.7E+02 | 2.4E+01 | NA | NA | NA | NA | NA | NA |
| CRP | LDL | WMode | 130 | 4.7E-02 | 1.8E-02 | 1.0E-02 | 2.7E+02 | 2.4E+01 | NA | NA | NA | NA | NA | NA |
| CRP | MetS | IVW | 224 | -7.9E-02 | 3.4E-02 | 2.2E-02 | 2.5E+02 | 2.4E+01 | 1.4E+03 | 2.6E-162 | 8.4E+01 | NA | NA | NA |
| CRP | MetS | MR Egger | 224 | -8.9E-02 | 4.5E-02 | 5.0E-02 | 2.5E+02 | 2.4E+01 | NA | NA | NA | 5.9E-04 | 1.7E-03 | 7.3E-01 |
| CRP | MetS | WMed | 224 | -6.1E-02 | 3.3E-02 | 6.4E-02 | 2.5E+02 | 2.4E+01 | NA | NA | NA | NA | NA | NA |
| CRP | MetS | WMode | 224 | -6.9E-02 | 2.1E-02 | 1.0E-03 | 2.5E+02 | 2.4E+01 | NA | NA | NA | NA | NA | NA |
| CRP | SBP | IVW | 199 | 5.0E-04 | 1.9E-01 | 1.0E+00 | 2.6E+02 | 2.4E+01 | 1.6E+03 | 2.0E-213 | 8.7E+01 | NA | NA | NA |
| CRP | SBP | MR Egger | 199 | 1.3E-01 | 2.5E-01 | 6.1E-01 | 2.6E+02 | 2.4E+01 | NA | NA | NA | -7.5E-03 | 9.4E-03 | 4.3E-01 |
| CRP | SBP | WMed | 199 | 1.8E-01 | 1.3E-01 | 1.7E-01 | 2.6E+02 | 2.4E+01 | NA | NA | NA | NA | NA | NA |
| CRP | SBP | WMode | 199 | 1.3E-01 | 9.5E-02 | 1.7E-01 | 2.6E+02 | 2.4E+01 | NA | NA | NA | NA | NA | NA |
| CRP | T2D | IVW | 155 | 5.1E-02 | 3.9E-02 | 1.9E-01 | 2.4E+02 | 2.4E+01 | 6.1E+02 | 4.6E-55 | 7.4E+01 | NA | NA | NA |
| CRP | T2D | MR Egger | 155 | 1.2E-01 | 5.1E-02 | 2.4E-02 | 2.4E+02 | 2.4E+01 | NA | NA | NA | -3.7E-03 | 1.9E-03 | 5.2E-02 |
| CRP | T2D | WMed | 155 | 5.7E-03 | 3.5E-02 | 8.7E-01 | 2.4E+02 | 2.4E+01 | NA | NA | NA | NA | NA | NA |
| CRP | T2D | WMode | 155 | 3.2E-02 | 2.8E-02 | 2.5E-01 | 2.4E+02 | 2.4E+01 | NA | NA | NA | NA | NA | NA |
| CRP | Triglycerides | IVW | 130 | -5.8E-02 | 4.2E-02 | 1.6E-01 | 2.7E+02 | 2.4E+01 | 2.1E+03 | 0.0E+00 | 9.4E+01 | NA | NA | NA |

| | | | | | | | | | | | | | | |
|---|---|---|---|---|---|---|---|---|---|---|---|---|---|---|
| CRP | Triglycerides | MR Egger | 130 | -9.0E-02 | 5.4E-02 | 9.5E-02 | 2.7E+02 | 2.4E+01 | NA | NA | NA | 2.2E-03 | 2.3E-03 | 3.5E-01 |
| CRP | Triglycerides | WMed | 130 | 1.9E-03 | 1.5E-02 | 9.0E-01 | 2.7E+02 | 2.4E+01 | NA | NA | NA | NA | NA | NA |
| CRP | Triglycerides | WMode | 130 | 1.4E-02 | 1.2E-02 | 2.5E-01 | 2.7E+02 | 2.4E+01 | NA | NA | NA | NA | NA | NA |
| Cystatin C | BMI | IVW | 305 | -1.4E-01 | 2.6E-02 | 6.7E-08 | 8.9E+01 | 3.0E+01 | 5.0E+03 | 0.0E+00 | 9.3E+01 | NA | NA | NA |
| Cystatin C | BMI | MR Egger | 305 | 1.2E-01 | 5.3E-02 | 2.7E-02 | 8.9E+01 | 3.0E+01 | NA | NA | NA | -6.2E-03 | 1.1E-03 | 1.0E-07 |
| Cystatin C | BMI | WMed | 305 | -2.4E-02 | 1.4E-02 | 8.5E-02 | 8.9E+01 | 3.0E+01 | NA | NA | NA | NA | NA | NA |
| Cystatin C | BMI | WMode | 305 | 3.0E-03 | 1.6E-02 | 8.5E-01 | 8.9E+01 | 3.0E+01 | NA | NA | NA | NA | NA | NA |
| Cystatin C | DBP | IVW | 270 | -6.1E-01 | 2.5E-01 | 1.3E-02 | 9.2E+01 | 3.0E+01 | 4.7E+03 | 0.0E+00 | 9.4E+01 | NA | NA | NA |
| Cystatin C | DBP | MR Egger | 270 | -1.5E+00 | 5.3E-01 | 5.0E-03 | 9.2E+01 | 3.0E+01 | NA | NA | NA | 2.1E-02 | 1.1E-02 | 6.0E-02 |
| Cystatin C | DBP | WMed | 270 | -1.8E-01 | 1.4E-01 | 1.9E-01 | 9.2E+01 | 3.0E+01 | NA | NA | NA | NA | NA | NA |
| Cystatin C | DBP | WMode | 270 | 2.8E-01 | 3.0E-01 | 3.5E-01 | 9.2E+01 | 3.0E+01 | NA | NA | NA | NA | NA | NA |
| Cystatin C | HbA1c | IVW | 138 | 1.7E-02 | 1.9E-02 | 3.6E-01 | 1.0E+02 | 3.0E+01 | 1.9E+02 | 2.2E-03 | 2.7E+01 | NA | NA | NA |
| Cystatin C | HbA1c | MR Egger | 138 | 4.1E-02 | 4.0E-02 | 3.0E-01 | 1.0E+02 | 3.0E+01 | NA | NA | NA | -6.0E-04 | 8.7E-04 | 4.9E-01 |
| Cystatin C | HbA1c | WMed | 138 | 3.5E-03 | 2.7E-02 | 9.0E-01 | 1.0E+02 | 3.0E+01 | NA | NA | NA | NA | NA | NA |
| Cystatin C | HbA1c | WMode | 138 | -3.9E-05 | 4.6E-02 | 1.0E+00 | 1.0E+02 | 3.0E+01 | NA | NA | NA | NA | NA | NA |
| Cystatin C | HDL | IVW | 149 | 1.2E-01 | 4.8E-02 | 1.1E-02 | 9.8E+01 | 3.0E+01 | 8.9E+02 | 9.6E-106 | 8.3E+01 | NA | NA | NA |
| Cystatin C | HDL | MR Egger | 149 | 1.8E-01 | 1.1E-01 | 9.1E-02 | 9.8E+01 | 3.0E+01 | NA | NA | NA | -1.5E-03 | 2.3E-03 | 5.3E-01 |
| Cystatin C | HDL | WMed | 149 | 1.3E-01 | 4.0E-02 | 1.2E-03 | 9.8E+01 | 3.0E+01 | NA | NA | NA | NA | NA | NA |
| Cystatin C | HDL | WMode | 149 | 8.2E-02 | 1.0E-01 | 4.1E-01 | 9.8E+01 | 3.0E+01 | NA | NA | NA | NA | NA | NA |
| Cystatin C | LDL | IVW | 149 | 9.3E-02 | 5.3E-02 | 7.8E-02 | 9.8E+01 | 3.0E+01 | 9.0E+02 | 3.7E-108 | 8.4E+01 | NA | NA | NA |
| Cystatin C | LDL | MR Egger | 149 | 3.1E-02 | 1.2E-01 | 7.9E-01 | 9.8E+01 | 3.0E+01 | NA | NA | NA | 1.5E-03 | 2.6E-03 | 5.6E-01 |
| Cystatin C | LDL | WMed | 149 | 1.9E-02 | 4.3E-02 | 6.6E-01 | 9.8E+01 | 3.0E+01 | NA | NA | NA | NA | NA | NA |

| | | | | | | | | | | | | | | |
|---|---|---|---|---|---|---|---|---|---|---|---|---|---|---|
| Cystatin C | LDL | WMode | 149 | 1.9E-03 | 8.7E-02 | 9.8E-01 | 9.8E+01 | 3.0E+01 | NA | NA | NA | NA | NA | NA |
| Cystatin C | MetS | IVW | 339 | 2.2E-01 | 4.4E-02 | 4.8E-07 | 9.6E+01 | 3.0E+01 | 1.8E+03 | 2.5E-196 | 8.1E+01 | NA | NA | NA |
| Cystatin C | MetS | MR Egger | 339 | 5.0E-03 | 8.9E-02 | 9.6E-01 | 9.6E+01 | 3.0E+01 | NA | NA | NA | 5.5E-03 | 1.9E-03 | 5.0E-03 |
| Cystatin C | MetS | WMed | 339 | 1.8E-01 | 3.9E-02 | 1.9E-06 | 9.6E+01 | 3.0E+01 | NA | NA | NA | NA | NA | NA |
| Cystatin C | MetS | WMode | 339 | 6.1E-02 | 4.7E-02 | 2.0E-01 | 9.6E+01 | 3.0E+01 | NA | NA | NA | NA | NA | NA |
| Cystatin C | SBP | IVW | 269 | -3.8E-01 | 3.8E-01 | 3.2E-01 | 9.2E+01 | 3.0E+01 | 3.6E+03 | 0.0E+00 | 9.2E+01 | NA | NA | NA |
| Cystatin C | SBP | MR Egger | 269 | -1.5E+00 | 8.2E-01 | 7.7E-02 | 9.2E+01 | 3.0E+01 | NA | NA | NA | 2.6E-02 | 1.7E-02 | 1.4E-01 |
| Cystatin C | SBP | WMed | 269 | 1.2E-01 | 2.5E-01 | 6.2E-01 | 9.2E+01 | 3.0E+01 | NA | NA | NA | NA | NA | NA |
| Cystatin C | SBP | WMode | 269 | 1.1E+00 | 5.1E-01 | 3.0E-02 | 9.2E+01 | 3.0E+01 | NA | NA | NA | NA | NA | NA |
| Cystatin C | T2D | IVW | 212 | -1.4E-01 | 6.4E-02 | 2.7E-02 | 9.5E+01 | 3.0E+01 | 1.0E+03 | 4.4E-107 | 7.9E+01 | NA | NA | NA |
| Cystatin C | T2D | MR Egger | 212 | 2.1E-01 | 1.5E-01 | 1.5E-01 | 9.5E+01 | 3.0E+01 | NA | NA | NA | -8.2E-03 | 3.0E-03 | 7.7E-03 |
| Cystatin C | T2D | WMed | 212 | 1.9E-02 | 5.8E-02 | 7.4E-01 | 9.5E+01 | 3.0E+01 | NA | NA | NA | NA | NA | NA |
| Cystatin C | T2D | WMode | 212 | 4.8E-02 | 8.8E-02 | 5.9E-01 | 9.5E+01 | 3.0E+01 | NA | NA | NA | NA | NA | NA |
| Cystatin C | Triglycerides | IVW | 149 | 5.2E-02 | 7.6E-02 | 4.9E-01 | 9.8E+01 | 3.0E+01 | 2.3E+03 | 0.0E+00 | 9.4E+01 | NA | NA | NA |
| Cystatin C | Triglycerides | MR Egger | 149 | -1.3E-01 | 1.7E-01 | 4.4E-01 | 9.8E+01 | 3.0E+01 | NA | NA | NA | 4.4E-03 | 3.7E-03 | 2.3E-01 |
| Cystatin C | Triglycerides | WMed | 149 | -1.4E-01 | 3.6E-02 | 1.1E-04 | 9.8E+01 | 3.0E+01 | NA | NA | NA | NA | NA | NA |
| Cystatin C | Triglycerides | WMode | 149 | -1.2E-01 | 3.6E-02 | 6.4E-04 | 9.8E+01 | 3.0E+01 | NA | NA | NA | NA | NA | NA |
| Fresh Fruit Consumption | BMI | IVW | 47 | -2.2E-02 | 2.8E-01 | 9.4E-01 | 4.9E+01 | 3.0E+01 | 1.3E+03 | 1.6E-237 | 9.6E+01 | NA | NA | NA |
| Fresh Fruit Consumption | BMI | MR Egger | 47 | -3.1E-01 | 1.0E+00 | 7.7E-01 | 4.9E+01 | 3.0E+01 | NA | NA | NA | 1.8E-03 | 6.2E-03 | 7.8E-01 |
| Fresh Fruit Consumption | BMI | WMed | 47 | 1.2E-01 | 1.3E-01 | 3.4E-01 | 4.9E+01 | 3.0E+01 | NA | NA | NA | NA | NA | NA |

| | | | | | | | | | | | | | | |
|---|---|---|---|---|---|---|---|---|---|---|---|---|---|---|
| Fresh Fruit Consumption | BMI | WMode | 47 | 2.8E-01 | 1.4E-01 | 4.6E-02 | 4.9E+01 | 3.0E+01 | NA | NA | NA | NA | NA | NA |
| Fresh Fruit Consumption | DBP | IVW | 41 | 7.0E-01 | 1.5E+0 0 | 6.4E-01 | 5.1E+01 | 3.0E+01 | 3.8E+02 | 1.8E-56 | 8.9E+01 | NA | NA | NA |
| Fresh Fruit Consumption | DBP | MR Egger | 41 | 3.6E+00 | 5.5E+0 0 | 5.2E-01 | 5.1E+01 | 3.0E+01 | NA | NA | NA | -1.8E-02 | 3.3E-02 | 5.9E-01 |
| Fresh Fruit Consumption | DBP | WMed | 41 | 1.8E+00 | 9.2E-01 | 4.9E-02 | 5.1E+01 | 3.0E+01 | NA | NA | NA | NA | NA | NA |
| Fresh Fruit Consumption | DBP | WMode | 41 | 2.6E+00 | 1.1E+0 0 | 2.2E-02 | 5.1E+01 | 3.0E+01 | NA | NA | NA | NA | NA | NA |
| Fresh Fruit Consumption | HbA1c | IVW | 23 | -1.7E-01 | 1.3E-01 | 1.8E-01 | 5.5E+01 | 3.0E+01 | 1.4E+01 | 8.9E-01 | 0.0E+00 | NA | NA | NA |
| Fresh Fruit Consumption | HbA1c | MR Egger | 23 | 2.4E-01 | 4.3E-01 | 5.9E-01 | 5.5E+01 | 3.0E+01 | NA | NA | NA | -2.7E-03 | 2.7E-03 | 3.3E-01 |
| Fresh Fruit Consumption | HbA1c | WMed | 23 | -1.8E-01 | 1.7E-01 | 3.0E-01 | 5.5E+01 | 3.0E+01 | NA | NA | NA | NA | NA | NA |
| Fresh Fruit Consumption | HbA1c | WMode | 23 | -1.1E-01 | 2.5E-01 | 6.8E-01 | 5.5E+01 | 3.0E+01 | NA | NA | NA | NA | NA | NA |
| Fresh Fruit Consumption | HDL | IVW | 24 | 1.4E-01 | 2.2E-01 | 5.3E-01 | 5.7E+01 | 3.0E+01 | 4.6E+01 | 2.9E-03 | 5.0E+01 | NA | NA | NA |
| Fresh Fruit Consumption | HDL | MR Egger | 24 | 1.9E-01 | 8.2E-01 | 8.2E-01 | 5.7E+01 | 3.0E+01 | NA | NA | NA | -3.5E-04 | 5.1E-03 | 9.5E-01 |
| Fresh Fruit Consumption | HDL | WMed | 24 | 3.2E-01 | 2.4E-01 | 1.8E-01 | 5.7E+01 | 3.0E+01 | NA | NA | NA | NA | NA | NA |
| Fresh Fruit Consumption | HDL | WMode | 24 | 3.0E-01 | 2.9E-01 | 3.1E-01 | 5.7E+01 | 3.0E+01 | NA | NA | NA | NA | NA | NA |
| Fresh Fruit Consumption | LDL | IVW | 24 | -3.2E-01 | 3.5E-01 | 3.6E-01 | 5.7E+01 | 3.0E+01 | 9.6E+01 | 6.8E-11 | 7.5E+01 | NA | NA | NA |
| Fresh Fruit Consumption | LDL | MR Egger | 24 | 9.5E-01 | 1.3E+0 0 | 4.6E-01 | 5.7E+01 | 3.0E+01 | NA | NA | NA | -8.2E-03 | 7.9E-03 | 3.1E-01 |

| | | | | | | | | | | | | | | |
|---|---|---|---|---|---|---|---|---|---|---|---|---|---|---|
| Fresh Fruit Consumption | LDL | WMed | 24 | 4.6E-02 | 2.4E-01 | 8.5E-01 | 5.7E+01 | 3.0E+01 | NA | NA | NA | NA | NA | NA |
| Fresh Fruit Consumption | LDL | WMode | 24 | 2.6E-02 | 3.0E-01 | 9.3E-01 | 5.7E+01 | 3.0E+01 | NA | NA | NA | NA | NA | NA |
| Fresh Fruit Consumption | MetS | IVW | 47 | 1.6E-01 | 4.5E-01 | 7.2E-01 | 4.9E+01 | 3.0E+01 | 3.1E+02 | 5.8E-40 | 8.5E+01 | NA | NA | NA |
| Fresh Fruit Consumption | MetS | MR Egger | 47 | 6.5E-01 | 1.7E+00 | 7.0E-01 | 4.9E+01 | 3.0E+01 | NA | NA | NA | -3.0E-03 | 9.9E-03 | 7.6E-01 |
| Fresh Fruit Consumption | MetS | WMed | 47 | -3.6E-02 | 3.4E-01 | 9.2E-01 | 4.9E+01 | 3.0E+01 | NA | NA | NA | NA | NA | NA |
| Fresh Fruit Consumption | MetS | WMode | 47 | -2.0E-01 | 4.7E-01 | 6.7E-01 | 4.9E+01 | 3.0E+01 | NA | NA | NA | NA | NA | NA |
| Fresh Fruit Consumption | SBP | IVW | 41 | -5.4E-02 | 2.6E+00 | 9.8E-01 | 5.1E+01 | 3.0E+01 | 3.6E+02 | 1.3E-53 | 8.8E+01 | NA | NA | NA |
| Fresh Fruit Consumption | SBP | MR Egger | 41 | 1.3E+01 | 9.1E+00 | 1.6E-01 | 5.1E+01 | 3.0E+01 | NA | NA | NA | -8.1E-02 | 5.5E-02 | 1.5E-01 |
| Fresh Fruit Consumption | SBP | WMed | 41 | 4.3E+00 | 1.5E+00 | 4.6E-03 | 5.1E+01 | 3.0E+01 | NA | NA | NA | NA | NA | NA |
| Fresh Fruit Consumption | SBP | WMode | 41 | 4.8E+00 | 1.7E+00 | 8.0E-03 | 5.1E+01 | 3.0E+01 | NA | NA | NA | NA | NA | NA |
| Fresh Fruit Consumption | T2D | IVW | 41 | -6.0E-01 | 4.2E-01 | 1.6E-01 | 4.9E+01 | 3.0E+01 | 1.4E+02 | 1.9E-13 | 7.1E+01 | NA | NA | NA |
| Fresh Fruit Consumption | T2D | MR Egger | 41 | -2.3E+00 | 1.5E+00 | 1.3E-01 | 4.9E+01 | 3.0E+01 | NA | NA | NA | 1.1E-02 | 8.8E-03 | 2.4E-01 |
| Fresh Fruit Consumption | T2D | WMed | 41 | -9.9E-01 | 4.0E-01 | 1.3E-02 | 4.9E+01 | 3.0E+01 | NA | NA | NA | NA | NA | NA |
| Fresh Fruit Consumption | T2D | WMode | 41 | -5.8E-01 | 6.3E-01 | 3.6E-01 | 4.9E+01 | 3.0E+01 | NA | NA | NA | NA | NA | NA |
| Fresh Fruit Consumption | Triglycerides | IVW | 24 | -4.1E-01 | 2.3E-01 | 7.3E-02 | 5.7E+01 | 3.0E+01 | 5.2E+01 | 5.5E-04 | 5.2E+01 | NA | NA | NA |

| | | | | | | | | | | | | | | |
|---|---|---|---|---|---|---|---|---|---|---|---|---|---|---|
| Fresh Fruit Consumption | Triglycerides | MR Egger | 24 | 6.6E-01 | 8.1E-01 | 4.3E-01 | 5.7E+01 | 3.0E+01 | NA | NA | NA | -7.0E-03 | 5.1E-03 | 1.9E-01 |
| Fresh Fruit Consumption | Triglycerides | WMed | 24 | -4.9E-01 | 2.7E-01 | 7.0E-02 | 5.7E+01 | 3.0E+01 | NA | NA | NA | NA | NA | NA |
| Fresh Fruit Consumption | Triglycerides | WMode | 24 | -8.8E-01 | 5.4E-01 | 1.2E-01 | 5.7E+01 | 3.0E+01 | NA | NA | NA | NA | NA | NA |
| Gamma Glutamyl Transpeptidase | BMI | IVW | 166 | -9.2E-02 | 1.3E-01 | 4.7E-01 | 1.3E+02 | 3.0E+01 | 1.8E-02 | 1.0E+00 | 0.0E+00 | NA | NA | NA |
| Gamma Glutamyl Transpeptidase | BMI | MR Egger | 166 | -7.2E-02 | 2.1E-01 | 7.3E-01 | 1.3E+02 | 3.0E+01 | NA | NA | NA | -1.0E-03 | 9.1E-03 | 9.1E-01 |
| Gamma Glutamyl Transpeptidase | BMI | WMed | 166 | -9.1E-02 | 1.4E-01 | 5.3E-01 | 1.3E+02 | 3.0E+01 | NA | NA | NA | NA | NA | NA |
| Gamma Glutamyl Transpeptidase | BMI | WMode | 166 | -9.0E-02 | 2.3E+00 | 9.7E-01 | 1.3E+02 | 3.0E+01 | NA | NA | NA | NA | NA | NA |
| Gamma Glutamyl Transpeptidase | DBP | IVW | 148 | -1.2E+00 | 5.6E-01 | 3.9E-02 | 1.2E+02 | 3.0E+01 | 5.3E-01 | 1.0E+00 | 0.0E+00 | NA | NA | NA |
| Gamma Glutamyl Transpeptidase | DBP | MR Egger | 148 | -3.2E-01 | 1.4E+00 | 8.2E-01 | 1.2E+02 | 3.0E+01 | NA | NA | NA | -1.5E-02 | 2.3E-02 | 5.1E-01 |
| Gamma Glutamyl Transpeptidase | DBP | WMed | 148 | -1.1E+00 | 7.4E-01 | 1.2E-01 | 1.2E+02 | 3.0E+01 | NA | NA | NA | NA | NA | NA |
| Gamma Glutamyl Transpeptidase | DBP | WMode | 148 | -7.7E-01 | 2.7E+00 | 7.7E-01 | 1.2E+02 | 3.0E+01 | NA | NA | NA | NA | NA | NA |
| Gamma Glutamyl Transpeptidase | HbA1C | IVW | 75 | -1.1E-01 | 6.2E-02 | 7.2E-02 | 1.1E+02 | 3.0E+01 | 2.2E-01 | 1.0E+00 | 0.0E+00 | NA | NA | NA |
| Gamma Glutamyl Transpeptidase | HbA1C | MR Egger | 75 | 5.3E-02 | 3.9E-01 | 8.9E-01 | 1.1E+02 | 3.0E+01 | NA | NA | NA | -6.7E-03 | 1.6E-02 | 6.7E-01 |
| Gamma Glutamyl Transpeptidase | HbA1C | WMed | 75 | -1.1E-01 | 6.4E-02 | 8.5E-02 | 1.1E+02 | 3.0E+01 | NA | NA | NA | NA | NA | NA |
| Gamma Glutamyl Transpeptidase | HbA1C | WMode | 75 | -1.1E-01 | 2.8E-01 | 7.0E-01 | 1.1E+02 | 3.0E+01 | NA | NA | NA | NA | NA | NA |

| | | | | | | | | | | | | | | |
|---|---|---|---|---|---|---|---|---|---|---|---|---|---|---|
| Gamma Glutamyl Transpeptidase | HDL | IVW | 83 | -2.6E-01 | 3.5E-01 | 4.6E-01 | 1.2E+02 | 3.0E+01 | 1.1E-01 | 1.0E+00 | 0.0E+00 | NA | NA | NA |
| Gamma Glutamyl Transpeptidase | HDL | MR Egger | 83 | -8.8E-02 | 7.5E-01 | 9.1E-01 | 1.2E+02 | 3.0E+01 | NA | NA | NA | -4.4E-03 | 1.7E-02 | 7.9E-01 |
| Gamma Glutamyl Transpeptidase | HDL | WMed | 83 | -3.4E-01 | 4.6E-01 | 4.6E-01 | 1.2E+02 | 3.0E+01 | NA | NA | NA | NA | NA | NA |
| Gamma Glutamyl Transpeptidase | HDL | WMode | 83 | -3.4E-01 | 1.8E+00 | 8.5E-01 | 1.2E+02 | 3.0E+01 | NA | NA | NA | NA | NA | NA |
| Gamma Glutamyl Transpeptidase | LDL | IVW | 83 | -4.2E-01 | 7.8E-02 | 6.2E-08 | 1.2E+02 | 3.0E+01 | 3.1E-01 | 1.0E+00 | 0.0E+00 | NA | NA | NA |
| Gamma Glutamyl Transpeptidase | LDL | MR Egger | 83 | -2.0E-01 | 5.3E-01 | 7.1E-01 | 1.2E+02 | 3.0E+01 | NA | NA | NA | -3.3E-03 | 7.8E-03 | 6.7E-01 |
| Gamma Glutamyl Transpeptidase | LDL | WMed | 83 | -4.2E-01 | 9.4E-02 | 6.9E-06 | 1.2E+02 | 3.0E+01 | NA | NA | NA | NA | NA | NA |
| Gamma Glutamyl Transpeptidase | LDL | WMode | 83 | -4.3E-01 | 3.6E+00 | 9.1E-01 | 1.2E+02 | 3.0E+01 | NA | NA | NA | NA | NA | NA |
| Gamma Glutamyl Transpeptidase | MetS | IVW | 75 | 2.0E-02 | 2.6E-02 | 4.3E-01 | 1.4E+02 | 3.0E+01 | 1.3E+03 | 3.4E-226 | 9.4E+01 | NA | NA | NA |
| Gamma Glutamyl Transpeptidase | MetS | MR Egger | 75 | 1.9E-02 | 4.2E-02 | 6.5E-01 | 1.4E+02 | 3.0E+01 | NA | NA | NA | 3.4E-05 | 1.1E-03 | 9.8E-01 |
| Gamma Glutamyl Transpeptidase | MetS | WMed | 75 | 2.4E-03 | 1.2E-02 | 8.3E-01 | 1.4E+02 | 3.0E+01 | NA | NA | NA | NA | NA | NA |
| Gamma Glutamyl Transpeptidase | MetS | WMode | 75 | 1.0E-02 | 9.0E-03 | 2.6E-01 | 1.4E+02 | 3.0E+01 | NA | NA | NA | NA | NA | NA |
| Gamma Glutamyl Transpeptidase | SBP | IVW | 146 | -1.7E+00 | 3.6E-01 | 5.3E-06 | 1.2E+02 | 3.0E+01 | 2.0E+00 | 1.0E+00 | 0.0E+00 | NA | NA | NA |
| Gamma Glutamyl Transpeptidase | SBP | MR Egger | 146 | -1.0E+00 | 6.7E-01 | 1.3E-01 | 1.2E+02 | 3.0E+01 | NA | NA | NA | -1.7E-02 | 1.6E-02 | 2.7E-01 |
| Gamma Glutamyl Transpeptidase | SBP | WMed | 146 | -1.6E+00 | 5.1E-01 | 1.7E-03 | 1.2E+02 | 3.0E+01 | NA | NA | NA | NA | NA | NA |

| | | | | | | | | | | | | | | |
|---|---|---|---|---|---|---|---|---|---|---|---|---|---|---|
| Gamma Glutamyl Transpeptidase | SBP | WMode | 146 | -1.6E+00 | 2.1E+00 | 4.5E-01 | 1.2E+02 | 3.0E+01 | NA | NA | NA | NA | NA | NA |
| Gamma Glutamyl Transpeptidase | T2D | IVW | 169 | -2.6E+00 | 1.3E-01 | 2.1E-97 | 1.3E+02 | 3.0E+01 | 1.1E+02 | 1.0E+00 | 0.0E+00 | NA | NA | NA |
| Gamma Glutamyl Transpeptidase | T2D | MR Egger | 169 | -3.0E+00 | 4.9E-01 | 4.5E-09 | 1.3E+02 | 3.0E+01 | NA | NA | NA | 6.9E-03 | 8.4E-03 | 4.1E-01 |
| Gamma Glutamyl Transpeptidase | T2D | WMed | 169 | -2.3E+00 | 2.7E-01 | 6.9E-17 | 1.3E+02 | 3.0E+01 | NA | NA | NA | NA | NA | NA |
| Gamma Glutamyl Transpeptidase | T2D | WMode | 169 | -2.2E+00 | 2.6E-01 | 1.3E-14 | 1.3E+02 | 3.0E+01 | NA | NA | NA | NA | NA | NA |
| Gamma Glutamyl Transpeptidase | Triglycerides | IVW | 83 | -2.3E-01 | 3.1E-01 | 4.5E-01 | 1.2E+02 | 3.0E+01 | 1.3E-01 | 1.0E+00 | 0.0E+00 | NA | NA | NA |
| Gamma Glutamyl Transpeptidase | Triglycerides | MR Egger | 83 | -1.5E-01 | 4.7E-01 | 7.5E-01 | 1.2E+02 | 3.0E+01 | NA | NA | NA | -3.9E-03 | 1.7E-02 | 8.2E-01 |
| Gamma Glutamyl Transpeptidase | Triglycerides | WMed | 83 | -2.1E-01 | 3.9E-01 | 5.9E-01 | 1.2E+02 | 3.0E+01 | NA | NA | NA | NA | NA | NA |
| Gamma Glutamyl Transpeptidase | Triglycerides | WMode | 83 | -2.1E-01 | 2.8E+00 | 9.4E-01 | 1.2E+02 | 3.0E+01 | NA | NA | NA | NA | NA | NA |
| Impedance of Left Arm | BMI | IVW | 406 | 7.2E-01 | 3.1E-02 | 1.4E-118 | 6.7E+01 | 3.0E+01 | 5.4E+03 | 0.0E+00 | 9.3E+01 | NA | NA | NA |
| Impedance of Left Arm | BMI | MR Egger | 406 | 7.1E-01 | 8.4E-02 | 9.4E-16 | 6.7E+01 | 3.0E+01 | NA | NA | NA | 2.0E-04 | 1.1E-03 | 8.6E-01 |
| Impedance of Left Arm | BMI | WMed | 406 | 4.7E-01 | 2.1E-02 | 6.9E-110 | 6.7E+01 | 3.0E+01 | NA | NA | NA | NA | NA | NA |
| Impedance of Left Arm | BMI | WMode | 406 | 3.5E-01 | 5.6E-02 | 6.1E-10 | 6.7E+01 | 3.0E+01 | NA | NA | NA | NA | NA | NA |
| Impedance of Left Arm | DBP | IVW | 372 | -8.7E-01 | 2.8E-01 | 2.0E-03 | 6.7E+01 | 3.0E+01 | 4.8E+03 | 0.0E+00 | 9.2E+01 | NA | NA | NA |
| Impedance of Left Arm | DBP | MR Egger | 372 | -1.6E+00 | 7.7E-01 | 3.5E-02 | 6.7E+01 | 3.0E+01 | NA | NA | NA | 1.1E-02 | 1.0E-02 | 2.9E-01 |

| | | | | | | | | | | | | | | |
|---|---|---|---|---|---|---|---|---|---|---|---|---|---|---|
| Impedance of Left Arm | DBP | WMed | 372 | -7.0E-01 | 1.7E-01 | 3.9E-05 | 6.7E+01 | 3.0E+01 | NA | NA | NA | NA | NA | NA |
| Impedance of Left Arm | DBP | WMode | 372 | -5.7E-01 | 3.5E-01 | 1.1E-01 | 6.7E+01 | 3.0E+01 | NA | NA | NA | NA | NA | NA |
| Impedance of Left Arm | HbA1c | IVW | 176 | 3.7E-02 | 2.8E-02 | 1.9E-01 | 7.2E+01 | 3.0E+01 | 2.7E+02 | 2.3E-06 | 3.6E+01 | NA | NA | NA |
| Impedance of Left Arm | HbA1c | MR Egger | 176 | 3.1E-02 | 8.2E-02 | 7.0E-01 | 7.2E+01 | 3.0E+01 | NA | NA | NA | 7.9E-05 | 1.1E-03 | 9.4E-01 |
| Impedance of Left Arm | HbA1c | WMed | 176 | 1.8E-02 | 3.6E-02 | 6.2E-01 | 7.2E+01 | 3.0E+01 | NA | NA | NA | NA | NA | NA |
| Impedance of Left Arm | HbA1c | WMode | 176 | 5.7E-02 | 7.1E-02 | 4.3E-01 | 7.2E+01 | 3.0E+01 | NA | NA | NA | NA | NA | NA |
| Impedance of Left Arm | HDL | IVW | 191 | -1.4E-01 | 4.8E-02 | 5.2E-03 | 7.5E+01 | 3.0E+01 | 7.0E+02 | 5.1E-59 | 7.3E+01 | NA | NA | NA |
| Impedance of Left Arm | HDL | MR Egger | 191 | -2.9E-01 | 1.3E-01 | 2.7E-02 | 7.5E+01 | 3.0E+01 | NA | NA | NA | 2.3E-03 | 1.8E-03 | 2.0E-01 |
| Impedance of Left Arm | HDL | WMed | 191 | -3.8E-02 | 5.2E-02 | 4.6E-01 | 7.5E+01 | 3.0E+01 | NA | NA | NA | NA | NA | NA |
| Impedance of Left Arm | HDL | WMode | 191 | 1.3E-01 | 2.4E-01 | 5.9E-01 | 7.5E+01 | 3.0E+01 | NA | NA | NA | NA | NA | NA |
| Impedance of Left Arm | LDL | IVW | 191 | 1.1E-02 | 4.7E-02 | 8.2E-01 | 7.5E+01 | 3.0E+01 | 5.7E+02 | 2.7E-39 | 6.6E+01 | NA | NA | NA |
| Impedance of Left Arm | LDL | MR Egger | 191 | 1.0E-01 | 1.3E-01 | 4.2E-01 | 7.5E+01 | 3.0E+01 | NA | NA | NA | -1.4E-03 | 1.8E-03 | 4.3E-01 |
| Impedance of Left Arm | LDL | WMed | 191 | 2.6E-02 | 5.0E-02 | 6.0E-01 | 7.5E+01 | 3.0E+01 | NA | NA | NA | NA | NA | NA |
| Impedance of Left Arm | LDL | WMode | 191 | 6.0E-02 | 6.5E-02 | 3.6E-01 | 7.5E+01 | 3.0E+01 | NA | NA | NA | NA | NA | NA |
| Impedance of Left Arm | MetS | IVW | 406 | -6.5E-01 | 5.9E-02 | 1.9E-28 | 6.7E+01 | 3.0E+01 | 1.8E+03 | 4.9E-173 | 7.7E+01 | NA | NA | NA |

| | | | | | | | | | | | | | | |
|---|---|---|---|---|---|---|---|---|---|---|---|---|---|---|
| Impedance of Left Arm | MetS | MR Egger | 406 | -5.2E-01 | 1.6E-01 | 1.3E-03 | 6.7E+01 | 3.0E+01 | NA | NA | NA | -1.9E-03 | 2.1E-03 | 3.7E-01 |
| Impedance of Left Arm | MetS | WMed | 406 | -4.9E-01 | 5.5E-02 | 4.8E-19 | 6.7E+01 | 3.0E+01 | NA | NA | NA | NA | NA | NA |
| Impedance of Left Arm | MetS | WMode | 406 | -9.1E-02 | 1.6E-01 | 5.6E-01 | 6.7E+01 | 3.0E+01 | NA | NA | NA | NA | NA | NA |
| Impedance of Left Arm | SBP | IVW | 371 | -1.1E+00 | 4.7E-01 | 1.4E-02 | 6.7E+01 | 3.0E+01 | 4.3E+03 | 0.0E+00 | 9.1E+01 | NA | NA | NA |
| Impedance of Left Arm | SBP | MR Egger | 371 | -2.4E+00 | 1.3E+00 | 5.7E-02 | 6.7E+01 | 3.0E+01 | NA | NA | NA | 1.8E-02 | 1.7E-02 | 2.8E-01 |
| Impedance of Left Arm | SBP | WMed | 371 | -7.2E-01 | 2.9E-01 | 1.3E-02 | 6.7E+01 | 3.0E+01 | NA | NA | NA | NA | NA | NA |
| Impedance of Left Arm | SBP | WMode | 371 | -4.1E-01 | 6.1E-01 | 5.0E-01 | 6.7E+01 | 3.0E+01 | NA | NA | NA | NA | NA | NA |
| Impedance of Left Arm | T2D | IVW | 314 | 6.2E-01 | 9.0E-02 | 5.6E-12 | 6.6E+01 | 3.0E+01 | 1.8E+03 | 2.4E-203 | 8.2E+01 | NA | NA | NA |
| Impedance of Left Arm | T2D | MR Egger | 314 | 1.3E+00 | 2.6E-01 | 5.7E-07 | 6.6E+01 | 3.0E+01 | NA | NA | NA | -9.9E-03 | 3.4E-03 | 3.9E-03 |
| Impedance of Left Arm | T2D | WMed | 314 | 4.2E-01 | 7.5E-02 | 1.3E-08 | 6.6E+01 | 3.0E+01 | NA | NA | NA | NA | NA | NA |
| Impedance of Left Arm | T2D | WMode | 314 | 2.0E-02 | 2.8E-01 | 9.4E-01 | 6.6E+01 | 3.0E+01 | NA | NA | NA | NA | NA | NA |
| Impedance of Left Arm | Triglycerides | IVW | 191 | 4.9E-02 | 7.9E-02 | 5.3E-01 | 7.5E+01 | 3.0E+01 | 1.9E+03 | 2.2E-286 | 9.0E+01 | NA | NA | NA |
| Impedance of Left Arm | Triglycerides | MR Egger | 191 | 6.6E-02 | 2.1E-01 | 7.5E-01 | 7.5E+01 | 3.0E+01 | NA | NA | NA | -2.6E-04 | 3.0E-03 | 9.3E-01 |
| Impedance of Left Arm | Triglycerides | WMed | 191 | 1.2E-01 | 4.9E-02 | 1.6E-02 | 7.5E+01 | 3.0E+01 | NA | NA | NA | NA | NA | NA |
| Impedance of Left Arm | Triglycerides | WMode | 191 | 1.2E-01 | 1.3E-01 | 3.8E-01 | 7.5E+01 | 3.0E+01 | NA | NA | NA | NA | NA | NA |

| | | | | | | | | | | | | | | |
|---|---|---|---|---|---|---|---|---|---|---|---|---|---|---|
| Impedance of Right Arm | BMI | IVW | 402 | 7.2E-01 | 3.2E-02 | 1.0E-110 | 6.7E+01 | 3.0E+01 | 5.6E+03 | 0.0E+00 | 9.3E+01 | NA | NA | NA |
| Impedance of Right Arm | BMI | MR Egger | 402 | 6.9E-01 | 8.7E-02 | 2.2E-14 | 6.7E+01 | 3.0E+01 | NA | NA | NA | 3.7E-04 | 1.2E-03 | 7.5E-01 |
| Impedance of Right Arm | BMI | WMed | 402 | 4.4E-01 | 2.0E-02 | 4.8E-108 | 6.7E+01 | 3.0E+01 | NA | NA | NA | NA | NA | NA |
| Impedance of Right Arm | BMI | WMode | 402 | 3.5E-01 | 5.2E-02 | 6.8E-11 | 6.7E+01 | 3.0E+01 | NA | NA | NA | NA | NA | NA |
| Impedance of Right Arm | DBP | IVW | 369 | -8.5E-01 | 2.8E-01 | 2.3E-03 | 6.7E+01 | 3.0E+01 | 4.6E+03 | 0.0E+00 | 9.2E+01 | NA | NA | NA |
| Impedance of Right Arm | DBP | MR Egger | 369 | -2.1E+00 | 7.6E-01 | 7.2E-03 | 6.7E+01 | 3.0E+01 | NA | NA | NA | 1.7E-02 | 1.0E-02 | 9.0E-02 |
| Impedance of Right Arm | DBP | WMed | 369 | -8.8E-01 | 1.7E-01 | 2.4E-07 | 6.7E+01 | 3.0E+01 | NA | NA | NA | NA | NA | NA |
| Impedance of Right Arm | DBP | WMode | 369 | -7.2E-01 | 3.3E-01 | 2.8E-02 | 6.7E+01 | 3.0E+01 | NA | NA | NA | NA | NA | NA |
| Impedance of Right Arm | HbA1c | IVW | 180 | 2.2E-02 | 2.6E-02 | 4.0E-01 | 7.3E+01 | 3.0E+01 | 2.5E+02 | 1.8E-04 | 3.0E+01 | NA | NA | NA |
| Impedance of Right Arm | HbA1c | MR Egger | 180 | 4.5E-02 | 7.4E-02 | 5.5E-01 | 7.3E+01 | 3.0E+01 | NA | NA | NA | -3.2E-04 | 9.9E-04 | 7.4E-01 |
| Impedance of Right Arm | HbA1c | WMed | 180 | 3.0E-02 | 3.8E-02 | 4.2E-01 | 7.3E+01 | 3.0E+01 | NA | NA | NA | NA | NA | NA |
| Impedance of Right Arm | HbA1c | WMode | 180 | 1.0E-01 | 7.4E-02 | 1.7E-01 | 7.3E+01 | 3.0E+01 | NA | NA | NA | NA | NA | NA |
| Impedance of Right Arm | HDL | IVW | 194 | -1.8E-01 | 4.7E-02 | 1.5E-04 | 7.4E+01 | 3.0E+01 | 6.8E+02 | 2.9E-56 | 7.2E+01 | NA | NA | NA |
| Impedance of Right Arm | HDL | MR Egger | 194 | -3.4E-01 | 1.3E-01 | 8.9E-03 | 7.4E+01 | 3.0E+01 | NA | NA | NA | 2.4E-03 | 1.8E-03 | 1.8E-01 |
| Impedance of Right Arm | HDL | WMed | 194 | -5.2E-02 | 5.1E-02 | 3.0E-01 | 7.4E+01 | 3.0E+01 | NA | NA | NA | NA | NA | NA |

| | | | | | | | | | | | | | | |
|---|---|---|---|---|---|---|---|---|---|---|---|---|---|---|
| Impedance of Right Arm | HDL | WMode | 194 | 5.3E-02 | 2.5E-01 | 8.3E-01 | 7.4E+01 | 3.0E+01 | NA | NA | NA | NA | NA | NA |
| Impedance of Right Arm | LDL | IVW | 194 | 1.5E-02 | 4.7E-02 | 7.5E-01 | 7.4E+01 | 3.0E+01 | 5.8E+02 | 1.0E-39 | 6.6E+01 | NA | NA | NA |
| Impedance of Right Arm | LDL | MR Egger | 194 | 6.3E-03 | 1.3E-01 | 9.6E-01 | 7.4E+01 | 3.0E+01 | NA | NA | NA | 1.3E-04 | 1.8E-03 | 9.4E-01 |
| Impedance of Right Arm | LDL | WMed | 194 | 2.8E-02 | 5.1E-02 | 5.9E-01 | 7.4E+01 | 3.0E+01 | NA | NA | NA | NA | NA | NA |
| Impedance of Right Arm | LDL | WMode | 194 | 7.2E-02 | 6.3E-02 | 2.6E-01 | 7.4E+01 | 3.0E+01 | NA | NA | NA | NA | NA | NA |
| Impedance of Right Arm | MetS | IVW | 401 | -6.6E-01 | 6.1E-02 | 4.0E-27 | 6.7E+01 | 3.0E+01 | 1.8E+03 | 5.2E-184 | 7.8E+01 | NA | NA | NA |
| Impedance of Right Arm | MetS | MR Egger | 401 | -3.4E-01 | 1.7E-01 | 4.2E-02 | 6.7E+01 | 3.0E+01 | NA | NA | NA | -4.6E-03 | 2.2E-03 | 3.9E-02 |
| Impedance of Right Arm | MetS | WMed | 401 | -4.9E-01 | 5.5E-02 | 9.3E-19 | 6.7E+01 | 3.0E+01 | NA | NA | NA | NA | NA | NA |
| Impedance of Right Arm | MetS | WMode | 401 | -1.3E-01 | 1.4E-01 | 3.5E-01 | 6.7E+01 | 3.0E+01 | NA | NA | NA | NA | NA | NA |
| Impedance of Right Arm | SBP | IVW | 368 | -8.9E-01 | 4.7E-01 | 6.0E-02 | 6.7E+01 | 3.0E+01 | 4.3E+03 | 0.0E+00 | 9.1E+01 | NA | NA | NA |
| Impedance of Right Arm | SBP | MR Egger | 368 | -2.7E+00 | 1.3E+0 0 | 3.7E-02 | 6.7E+01 | 3.0E+01 | NA | NA | NA | 2.6E-02 | 1.7E-02 | 1.3E-01 |
| Impedance of Right Arm | SBP | WMed | 368 | -5.6E-01 | 3.1E-01 | 6.8E-02 | 6.7E+01 | 3.0E+01 | NA | NA | NA | NA | NA | NA |
| Impedance of Right Arm | SBP | WMode | 368 | -2.5E-01 | 6.1E-01 | 6.8E-01 | 6.7E+01 | 3.0E+01 | NA | NA | NA | NA | NA | NA |
| Impedance of Right Arm | T2D | IVW | 310 | 6.7E-01 | 9.1E-02 | 2.9E-13 | 6.7E+01 | 3.0E+01 | 1.8E+03 | 3.7E-204 | 8.2E+01 | NA | NA | NA |
| Impedance of Right Arm | T2D | MR Egger | 310 | 1.1E+00 | 2.7E-01 | 5.4E-05 | 6.7E+01 | 3.0E+01 | NA | NA | NA | -6.0E-03 | 3.5E-03 | 8.6E-02 |

| | | | | | | | | | | | | | | |
|---|---|---|---|---|---|---|---|---|---|---|---|---|---|---|
| Impedance of Right Arm | T2D | WMed | 310 | 4.2E-01 | 7.5E-02 | 1.7E-08 | 6.7E+01 | 3.0E+01 | NA | NA | NA | NA | NA | NA |
| Impedance of Right Arm | T2D | WMode | 310 | -7.2E-04 | 2.6E-01 | 1.0E+0 0 | 6.7E+01 | 3.0E+01 | NA | NA | NA | NA | NA | NA |
| Impedance of Right Arm | Triglycerides | IVW | 194 | 7.1E-02 | 7.8E-02 | 3.6E-01 | 7.4E+01 | 3.0E+01 | 2.0E+03 | 1.3E-288 | 9.0E+01 | NA | NA | NA |
| Impedance of Right Arm | Triglycerides | MR Egger | 194 | 1.0E-01 | 2.1E-01 | 6.3E-01 | 7.4E+01 | 3.0E+01 | NA | NA | NA | -4.9E-04 | 3.0E-03 | 8.7E-01 |
| Impedance of Right Arm | Triglycerides | WMed | 194 | 1.4E-01 | 4.6E-02 | 2.8E-03 | 7.4E+01 | 3.0E+01 | NA | NA | NA | NA | NA | NA |
| Impedance of Right Arm | Triglycerides | WMode | 194 | 1.5E-01 | 1.1E-01 | 1.9E-01 | 7.4E+01 | 3.0E+01 | NA | NA | NA | NA | NA | NA |
| Overall Healthy Diet | BMI | IVW | 15 | -1.7E-01 | 1.1E-01 | 1.3E-01 | 5.4E+01 | 3.1E+01 | 4.4E+02 | 3.7E-85 | 9.7E+01 | NA | NA | NA |
| Overall Healthy Diet | BMI | MR Egger | 15 | -3.9E-01 | 4.0E-01 | 3.4E-01 | 5.4E+01 | 3.1E+01 | NA | NA | NA | 6.2E-03 | 1.1E-02 | 5.7E-01 |
| Overall Healthy Diet | BMI | WMed | 15 | -2.6E-01 | 3.4E-02 | 1.3E-13 | 5.4E+01 | 3.1E+01 | NA | NA | NA | NA | NA | NA |
| Overall Healthy Diet | BMI | WMode | 15 | -2.5E-01 | 3.4E-02 | 4.1E-06 | 5.4E+01 | 3.1E+01 | NA | NA | NA | NA | NA | NA |
| Overall Healthy Diet | DBP | IVW | 13 | 2.4E-01 | 6.5E-01 | 7.1E-01 | 5.7E+01 | 3.1E+01 | 1.5E+02 | 3.8E-26 | 9.1E+01 | NA | NA | NA |
| Overall Healthy Diet | DBP | MR Egger | 13 | 2.5E+00 | 2.2E+0 0 | 2.9E-01 | 5.7E+01 | 3.1E+01 | NA | NA | NA | -6.4E-02 | 6.1E-02 | 3.2E-01 |
| Overall Healthy Diet | DBP | WMed | 13 | -7.1E-01 | 3.3E-01 | 3.2E-02 | 5.7E+01 | 3.1E+01 | NA | NA | NA | NA | NA | NA |
| Overall Healthy Diet | DBP | WMode | 13 | -8.3E-01 | 3.8E-01 | 4.8E-02 | 5.7E+01 | 3.1E+01 | NA | NA | NA | NA | NA | NA |
| Overall Healthy Diet | HbA1c | IVW | 8 | -1.3E-02 | 4.7E-02 | 7.9E-01 | 6.4E+01 | 3.1E+01 | 3.2E+00 | 8.7E-01 | 0.0E+00 | NA | NA | NA |

| | | | | | | | | | | | | | | |
|---|---|---|---|---|---|---|---|---|---|---|---|---|---|---|
| Overall Healthy Diet | HbA1c | MR Egger | 8 | -2.0E-02 | 1.4E-01 | 9.0E-01 | 6.4E+01 | 3.1E+01 | NA | NA | NA | 2.0E-04 | 4.0E-03 | 9.6E-01 |
| Overall Healthy Diet | HbA1c | WMed | 8 | -2.9E-02 | 6.2E-02 | 6.4E-01 | 6.4E+01 | 3.1E+01 | NA | NA | NA | NA | NA | NA |
| Overall Healthy Diet | HbA1c | WMode | 8 | -4.3E-02 | 7.7E-02 | 6.0E-01 | 6.4E+01 | 3.1E+01 | NA | NA | NA | NA | NA | NA |
| Overall Healthy Diet | HDL | IVW | 10 | -5.7E-02 | 1.1E-01 | 6.0E-01 | 6.0E+01 | 3.1E+01 | 4.0E+01 | 6.5E-06 | 7.3E+01 | NA | NA | NA |
| Overall Healthy Diet | HDL | MR Egger | 10 | 3.0E-01 | 3.1E-01 | 3.6E-01 | 6.0E+01 | 3.1E+01 | NA | NA | NA | -1.1E-02 | 9.0E-03 | 2.5E-01 |
| Overall Healthy Diet | HDL | WMed | 10 | -6.6E-02 | 7.9E-02 | 4.0E-01 | 6.0E+01 | 3.1E+01 | NA | NA | NA | NA | NA | NA |
| Overall Healthy Diet | HDL | WMode | 10 | -6.5E-02 | 9.9E-02 | 5.3E-01 | 6.0E+01 | 3.1E+01 | NA | NA | NA | NA | NA | NA |
| Overall Healthy Diet | LDL | IVW | 10 | -5.0E-02 | 1.3E-01 | 6.9E-01 | 6.0E+01 | 3.1E+01 | 4.7E+01 | 3.3E-07 | 8.1E+01 | NA | NA | NA |
| Overall Healthy Diet | LDL | MR Egger | 10 | -2.1E-01 | 3.9E-01 | 6.0E-01 | 6.0E+01 | 3.1E+01 | NA | NA | NA | 5.0E-03 | 1.1E-02 | 6.7E-01 |
| Overall Healthy Diet | LDL | WMed | 10 | -1.1E-01 | 7.4E-02 | 1.6E-01 | 6.0E+01 | 3.1E+01 | NA | NA | NA | NA | NA | NA |
| Overall Healthy Diet | LDL | WMode | 10 | -1.1E-01 | 7.5E-02 | 1.6E-01 | 6.0E+01 | 3.1E+01 | NA | NA | NA | NA | NA | NA |
| Overall Healthy Diet | MetS | IVW | 16 | 8.6E-02 | 2.4E-01 | 7.2E-01 | 5.3E+01 | 3.1E+01 | 2.1E+02 | 1.6E-36 | 9.3E+01 | NA | NA | NA |
| Overall Healthy Diet | MetS | MR Egger | 16 | 2.2E-01 | 8.8E-01 | 8.1E-01 | 5.3E+01 | 3.1E+01 | NA | NA | NA | -3.8E-03 | 2.4E-02 | 8.8E-01 |
| Overall Healthy Diet | MetS | WMed | 16 | 8.1E-02 | 1.4E-01 | 5.6E-01 | 5.3E+01 | 3.1E+01 | NA | NA | NA | NA | NA | NA |
| Overall Healthy Diet | MetS | WMode | 16 | 1.2E-01 | 2.2E-01 | 5.7E-01 | 5.3E+01 | 3.1E+01 | NA | NA | NA | NA | NA | NA |

| | | | | | | | | | | | | | | |
|---|---|---|---|---|---|---|---|---|---|---|---|---|---|---|
| Overall Healthy Diet | SBP | IVW | 13 | 1.3E-01 | 1.1E+00 | 9.1E-01 | 5.7E+01 | 3.1E+01 | 1.5E+02 | 1.2E-25 | 9.2E+01 | NA | NA | NA |
| Overall Healthy Diet | SBP | MR Egger | 13 | 2.3E+00 | 3.9E+00 | 5.7E-01 | 5.7E+01 | 3.1E+01 | NA | NA | NA | -6.2E-02 | 1.1E-01 | 5.8E-01 |
| Overall Healthy Diet | SBP | WMed | 13 | -1.4E+00 | 5.4E-01 | 8.2E-03 | 5.7E+01 | 3.1E+01 | NA | NA | NA | NA | NA | NA |
| Overall Healthy Diet | SBP | WMode | 13 | -1.7E+00 | 6.1E-01 | 1.4E-02 | 5.7E+01 | 3.1E+01 | NA | NA | NA | NA | NA | NA |
| Overall Healthy Diet | T2D | IVW | 11 | -4.9E-01 | 3.2E-01 | 1.2E-01 | 4.8E+01 | 3.3E+01 | 1.0E+02 | 6.6E-18 | 9.0E+01 | NA | NA | NA |
| Overall Healthy Diet | T2D | MR Egger | 11 | 6.3E-02 | 1.6E+00 | 9.7E-01 | 4.8E+01 | 3.3E+01 | NA | NA | NA | -1.4E-02 | 4.1E-02 | 7.4E-01 |
| Overall Healthy Diet | T2D | WMed | 11 | -3.9E-01 | 1.5E-01 | 9.8E-03 | 4.8E+01 | 3.3E+01 | NA | NA | NA | NA | NA | NA |
| Overall Healthy Diet | T2D | WMode | 11 | -4.2E-01 | 1.7E-01 | 3.5E-02 | 4.8E+01 | 3.3E+01 | NA | NA | NA | NA | NA | NA |
| Overall Healthy Diet | Triglycerides | IVW | 10 | 5.6E-01 | 5.3E-01 | 2.9E-01 | 6.0E+01 | 3.1E+01 | 1.0E+03 | 3.2E-217 | 9.9E+01 | NA | NA | NA |
| Overall Healthy Diet | Triglycerides | MR Egger | 10 | -1.1E+00 | 1.5E+00 | 4.9E-01 | 6.0E+01 | 3.1E+01 | NA | NA | NA | 5.2E-02 | 4.4E-02 | 2.8E-01 |
| Overall Healthy Diet | Triglycerides | WMed | 10 | 2.5E-02 | 7.6E-02 | 7.4E-01 | 6.0E+01 | 3.1E+01 | NA | NA | NA | NA | NA | NA |
| Overall Healthy Diet | Triglycerides | WMode | 10 | 6.7E-02 | 8.2E-02 | 4.3E-01 | 6.0E+01 | 3.1E+01 | NA | NA | NA | NA | NA | NA |
| Peak Expiratory Flow | BMI | IVW | 112 | 2.4E-02 | 3.6E-02 | 5.1E-01 | 5.2E+01 | 3.0E+01 | 7.3E+02 | 5.1E-91 | 8.5E+01 | NA | NA | NA |
| Peak Expiratory Flow | BMI | MR Egger | 112 | -6.4E-02 | 1.2E-01 | 5.8E-01 | 5.2E+01 | 3.0E+01 | NA | NA | NA | 1.4E-03 | 1.8E-03 | 4.3E-01 |
| Peak Expiratory Flow | BMI | WMed | 112 | -8.4E-03 | 2.6E-02 | 7.5E-01 | 5.2E+01 | 3.0E+01 | NA | NA | NA | NA | NA | NA |

| | | | | | | | | | | | | | | |
|---|---|---|---|---|---|---|---|---|---|---|---|---|---|---|
| Peak Expiratory Flow | BMI | WMode | 112 | -4.8E-02 | 4.6E-02 | 2.9E-01 | 5.2E+01 | 3.0E+01 | NA | NA | NA | NA | NA | NA |
| Peak Expiratory Flow | DBP | IVW | 101 | 1.5E-01 | 4.2E-01 | 7.1E-01 | 5.3E+01 | 3.0E+01 | 1.1E+03 | 2.9E-158 | 9.0E+01 | NA | NA | NA |
| Peak Expiratory Flow | DBP | MR Egger | 101 | 9.0E-01 | 1.4E+00 | 5.2E-01 | 5.3E+01 | 3.0E+01 | NA | NA | NA | -1.2E-02 | 2.2E-02 | 5.8E-01 |
| Peak Expiratory Flow | DBP | WMed | 101 | 1.7E-01 | 2.9E-01 | 5.7E-01 | 5.3E+01 | 3.0E+01 | NA | NA | NA | NA | NA | NA |
| Peak Expiratory Flow | DBP | WMode | 101 | 5.3E-01 | 3.3E-01 | 1.1E-01 | 5.3E+01 | 3.0E+01 | NA | NA | NA | NA | NA | NA |
| Peak Expiratory Flow | HbA1c | IVW | 60 | 1.4E-02 | 3.9E-02 | 7.1E-01 | 5.7E+01 | 3.0E+01 | 8.1E+01 | 3.3E-02 | 2.7E+01 | NA | NA | NA |
| Peak Expiratory Flow | HbA1c | MR Egger | 60 | -2.9E-02 | 1.3E-01 | 8.2E-01 | 5.7E+01 | 3.0E+01 | NA | NA | NA | 7.1E-04 | 2.0E-03 | 7.2E-01 |
| Peak Expiratory Flow | HbA1c | WMed | 60 | -5.3E-03 | 5.4E-02 | 9.2E-01 | 5.7E+01 | 3.0E+01 | NA | NA | NA | NA | NA | NA |
| Peak Expiratory Flow | HbA1c | WMode | 60 | -3.9E-02 | 9.7E-02 | 6.9E-01 | 5.7E+01 | 3.0E+01 | NA | NA | NA | NA | NA | NA |
| Peak Expiratory Flow | HDL | IVW | 61 | -4.1E-02 | 5.5E-02 | 4.5E-01 | 5.6E+01 | 3.0E+01 | 9.3E+01 | 4.3E-03 | 3.4E+01 | NA | NA | NA |
| Peak Expiratory Flow | HDL | MR Egger | 61 | 1.3E-01 | 1.8E-01 | 4.7E-01 | 5.6E+01 | 3.0E+01 | NA | NA | NA | -2.8E-03 | 2.8E-03 | 3.2E-01 |
| Peak Expiratory Flow | HDL | WMed | 61 | 6.2E-02 | 8.0E-02 | 4.3E-01 | 5.6E+01 | 3.0E+01 | NA | NA | NA | NA | NA | NA |
| Peak Expiratory Flow | HDL | WMode | 61 | 6.6E-02 | 1.1E-01 | 5.7E-01 | 5.6E+01 | 3.0E+01 | NA | NA | NA | NA | NA | NA |
| Peak Expiratory Flow | LDL | IVW | 61 | 1.2E-01 | 6.6E-02 | 8.1E-02 | 5.6E+01 | 3.0E+01 | 1.1E+02 | 2.6E-05 | 4.8E+01 | NA | NA | NA |
| Peak Expiratory Flow | LDL | MR Egger | 61 | 2.0E-02 | 2.2E-01 | 9.3E-01 | 5.6E+01 | 3.0E+01 | NA | NA | NA | 1.6E-03 | 3.5E-03 | 6.5E-01 |

| | | | | | | | | | | | | | | |
|---|---|---|---|---|---|---|---|---|---|---|---|---|---|---|
| Peak Expiratory Flow | LDL | WMed | 61 | 9.5E-03 | 8.0E-02 | 9.1E-01 | 5.6E+01 | 3.0E+01 | NA | NA | NA | NA | NA | NA |
| Peak Expiratory Flow | LDL | WMode | 61 | -8.4E-02 | 1.1E-01 | 4.5E-01 | 5.6E+01 | 3.0E+01 | NA | NA | NA | NA | NA | NA |
| Peak Expiratory Flow | MetS | IVW | 123 | -1.0E-01 | 7.7E-02 | 1.9E-01 | 5.1E+01 | 3.0E+01 | 3.6E+02 | 3.1E-25 | 6.6E+01 | NA | NA | NA |
| Peak Expiratory Flow | MetS | MR Egger | 123 | 3.5E-02 | 2.6E-01 | 8.9E-01 | 5.1E+01 | 3.0E+01 | NA | NA | NA | -2.2E-03 | 3.9E-03 | 5.8E-01 |
| Peak Expiratory Flow | MetS | WMed | 123 | -5.3E-02 | 8.4E-02 | 5.3E-01 | 5.1E+01 | 3.0E+01 | NA | NA | NA | NA | NA | NA |
| Peak Expiratory Flow | MetS | WMode | 123 | -1.9E-02 | 1.4E-01 | 8.9E-01 | 5.1E+01 | 3.0E+01 | NA | NA | NA | NA | NA | NA |
| Peak Expiratory Flow | SBP | IVW | 101 | 1.6E+00 | 7.2E-01 | 2.8E-02 | 5.3E+01 | 3.0E+01 | 1.0E+03 | 1.9E-150 | 9.0E+01 | NA | NA | NA |
| Peak Expiratory Flow | SBP | MR Egger | 101 | 2.7E-01 | 2.4E+00 | 9.1E-01 | 5.3E+01 | 3.0E+01 | NA | NA | NA | 2.1E-02 | 3.7E-02 | 5.7E-01 |
| Peak Expiratory Flow | SBP | WMed | 101 | 8.9E-01 | 5.2E-01 | 8.5E-02 | 5.3E+01 | 3.0E+01 | NA | NA | NA | NA | NA | NA |
| Peak Expiratory Flow | SBP | WMode | 101 | 3.5E-01 | 6.1E-01 | 5.6E-01 | 5.3E+01 | 3.0E+01 | NA | NA | NA | NA | NA | NA |
| Peak Expiratory Flow | T2D | IVW | 79 | 4.7E-01 | 1.2E-01 | 1.7E-04 | 5.1E+01 | 3.0E+01 | 2.7E+02 | 1.1E-22 | 7.1E+01 | NA | NA | NA |
| Peak Expiratory Flow | T2D | MR Egger | 79 | 6.3E-01 | 4.6E-01 | 1.8E-01 | 5.1E+01 | 3.0E+01 | NA | NA | NA | -2.5E-03 | 6.8E-03 | 7.2E-01 |
| Peak Expiratory Flow | T2D | WMed | 79 | 2.0E-01 | 1.2E-01 | 1.0E-01 | 5.1E+01 | 3.0E+01 | NA | NA | NA | NA | NA | NA |
| Peak Expiratory Flow | T2D | WMode | 79 | -2.5E-03 | 1.9E-01 | 9.9E-01 | 5.1E+01 | 3.0E+01 | NA | NA | NA | NA | NA | NA |
| Peak Expiratory Flow | Triglycerides | IVW | 61 | 8.9E-02 | 4.6E-02 | 5.4E-02 | 5.6E+01 | 3.0E+01 | 6.9E+01 | 2.0E-01 | 1.3E+01 | NA | NA | NA |

| | | | | | | | | | | | | | | |
|---|---|---|---|---|---|---|---|---|---|---|---|---|---|---|
| Peak Expiratory Flow | Triglycerides | MR Egger | 61 | 3.1E-02 | 1.5E-01 | 8.4E-01 | 5.6E+01 | 3.0E+01 | NA | NA | NA | 9.5E-04 | 2.4E-03 | 6.9E-01 |
| Peak Expiratory Flow | Triglycerides | WMed | 61 | 7.3E-02 | 6.9E-02 | 2.9E-01 | 5.6E+01 | 3.0E+01 | NA | NA | NA | NA | NA | NA |
| Peak Expiratory Flow | Triglycerides | WMode | 61 | 8.9E-02 | 9.4E-02 | 3.5E-01 | 5.6E+01 | 3.0E+01 | NA | NA | NA | NA | NA | NA |
| Phosphate | BMI | IVW | 148 | -6.7E-03 | 1.7E-02 | 6.9E-01 | 9.7E+01 | 3.0E+01 | 7.7E+02 | 2.9E-85 | 8.1E+01 | NA | NA | NA |
| Phosphate | BMI | MR Egger | 148 | 5.7E-03 | 3.0E-02 | 8.5E-01 | 9.7E+01 | 3.0E+01 | NA | NA | NA | -4.0E-04 | 8.0E-04 | 6.2E-01 |
| Phosphate | BMI | WMed | 148 | 6.8E-03 | 1.4E-02 | 6.1E-01 | 9.7E+01 | 3.0E+01 | NA | NA | NA | NA | NA | NA |
| Phosphate | BMI | WMode | 148 | -3.6E-05 | 1.2E-02 | 1.0E+00 | 9.7E+01 | 3.0E+01 | NA | NA | NA | NA | NA | NA |
| Phosphate | DBP | IVW | 133 | -1.0E-02 | 2.1E-01 | 9.6E-01 | 1.0E+02 | 3.0E+01 | 1.3E+03 | 2.0E-184 | 8.9E+01 | NA | NA | NA |
| Phosphate | DBP | MR Egger | 133 | 4.0E-01 | 3.7E-01 | 2.8E-01 | 1.0E+02 | 3.0E+01 | NA | NA | NA | -1.3E-02 | 1.0E-02 | 1.8E-01 |
| Phosphate | DBP | WMed | 133 | 1.5E-02 | 1.3E-01 | 9.1E-01 | 1.0E+02 | 3.0E+01 | NA | NA | NA | NA | NA | NA |
| Phosphate | DBP | WMode | 133 | -4.5E-02 | 1.2E-01 | 7.2E-01 | 1.0E+02 | 3.0E+01 | NA | NA | NA | NA | NA | NA |
| Phosphate | HbA1c | IVW | 62 | -5.6E-02 | 2.4E-02 | 2.0E-02 | 9.7E+01 | 3.0E+01 | 4.9E+01 | 8.7E-01 | 0.0E+00 | NA | NA | NA |
| Phosphate | HbA1c | MR Egger | 62 | -3.7E-02 | 5.1E-02 | 4.7E-01 | 9.7E+01 | 3.0E+01 | NA | NA | NA | -4.8E-04 | 1.1E-03 | 6.7E-01 |
| Phosphate | HbA1c | WMed | 62 | -3.6E-02 | 3.6E-02 | 3.2E-01 | 9.7E+01 | 3.0E+01 | NA | NA | NA | NA | NA | NA |
| Phosphate | HbA1c | WMode | 62 | -2.9E-02 | 5.0E-02 | 5.6E-01 | 9.7E+01 | 3.0E+01 | NA | NA | NA | NA | NA | NA |
| Phosphate | HDL | IVW | 59 | 2.4E-02 | 4.6E-02 | 6.0E-01 | 7.5E+01 | 3.0E+01 | 1.1E+02 | 2.5E-05 | 4.7E+01 | NA | NA | NA |
| Phosphate | HDL | MR Egger | 59 | -1.0E-01 | 1.1E-01 | 3.7E-01 | 7.5E+01 | 3.0E+01 | NA | NA | NA | 2.9E-03 | 2.3E-03 | 2.2E-01 |
| Phosphate | HDL | WMed | 59 | -3.1E-02 | 5.8E-02 | 5.9E-01 | 7.5E+01 | 3.0E+01 | NA | NA | NA | NA | NA | NA |
| Phosphate | HDL | WMode | 59 | -7.2E-03 | 6.8E-02 | 9.2E-01 | 7.5E+01 | 3.0E+01 | NA | NA | NA | NA | NA | NA |
| Phosphate | LDL | IVW | 59 | -8.6E-02 | 5.4E-02 | 1.1E-01 | 7.5E+01 | 3.0E+01 | 1.3E+02 | 1.9E-07 | 5.5E+01 | NA | NA | NA |
| Phosphate | LDL | MR Egger | 59 | -4.8E-02 | 1.3E-01 | 7.1E-01 | 7.5E+01 | 3.0E+01 | NA | NA | NA | -8.9E-04 | 2.8E-03 | 7.5E-01 |
| Phosphate | LDL | WMed | 59 | -8.5E-02 | 5.7E-02 | 1.4E-01 | 7.5E+01 | 3.0E+01 | NA | NA | NA | NA | NA | NA |

| | | | | | | | | | | | | | | |
|---|---|---|---|---|---|---|---|---|---|---|---|---|---|---|
| Phosphate | LDL | WMode | 59 | -7.8E-02 | 6.6E-02 | 2.4E-01 | 7.5E+01 | 3.0E+01 | NA | NA | NA | NA | NA | NA |
| Phosphate | MetS | IVW | 168 | 4.4E-03 | 4.0E-02 | 9.1E-01 | 9.4E+01 | 3.0E+01 | 4.8E+02 | 1.3E-32 | 6.5E+01 | NA | NA | NA |
| Phosphate | MetS | MR Egger | 168 | -9.0E-03 | 7.2E-02 | 9.0E-01 | 9.4E+01 | 3.0E+01 | NA | NA | NA | 4.2E-04 | 1.9E-03 | 8.2E-01 |
| Phosphate | MetS | WMed | 168 | 3.8E-02 | 3.9E-02 | 3.3E-01 | 9.4E+01 | 3.0E+01 | NA | NA | NA | NA | NA | NA |
| Phosphate | MetS | WMode | 168 | 9.6E-03 | 3.8E-02 | 8.0E-01 | 9.4E+01 | 3.0E+01 | NA | NA | NA | NA | NA | NA |
| Phosphate | SBP | IVW | 133 | -1.0E-03 | 3.4E-01 | 1.0E+00 | 1.0E+02 | 3.0E+01 | 1.1E+03 | 1.8E-148 | 8.8E+01 | NA | NA | NA |
| Phosphate | SBP | MR Egger | 133 | 6.0E-01 | 5.9E-01 | 3.2E-01 | 1.0E+02 | 3.0E+01 | NA | NA | NA | -2.0E-02 | 1.6E-02 | 2.2E-01 |
| Phosphate | SBP | WMed | 133 | 1.7E-01 | 2.4E-01 | 4.7E-01 | 1.0E+02 | 3.0E+01 | NA | NA | NA | NA | NA | NA |
| Phosphate | SBP | WMode | 133 | 1.2E-01 | 2.2E-01 | 5.9E-01 | 1.0E+02 | 3.0E+01 | NA | NA | NA | NA | NA | NA |
| Phosphate | T2D | IVW | 107 | 4.1E-02 | 6.1E-02 | 5.0E-01 | 9.4E+01 | 3.0E+01 | 3.2E+02 | 1.9E-23 | 6.7E+01 | NA | NA | NA |
| Phosphate | T2D | MR Egger | 107 | 1.2E-01 | 1.1E-01 | 2.8E-01 | 9.4E+01 | 3.0E+01 | NA | NA | NA | -2.4E-03 | 2.7E-03 | 3.9E-01 |
| Phosphate | T2D | WMed | 107 | 9.7E-02 | 5.6E-02 | 8.4E-02 | 9.4E+01 | 3.0E+01 | NA | NA | NA | NA | NA | NA |
| Phosphate | T2D | WMode | 107 | 1.1E-01 | 5.6E-02 | 6.4E-02 | 9.4E+01 | 3.0E+01 | NA | NA | NA | NA | NA | NA |
| Phosphate | Triglycerides | IVW | 59 | 1.7E-02 | 4.7E-02 | 7.2E-01 | 7.5E+01 | 3.0E+01 | 1.2E+02 | 1.3E-06 | 5.3E+01 | NA | NA | NA |
| Phosphate | Triglycerides | MR Egger | 59 | 5.4E-02 | 1.1E-01 | 6.3E-01 | 7.5E+01 | 3.0E+01 | NA | NA | NA | -8.7E-04 | 2.4E-03 | 7.2E-01 |
| Phosphate | Triglycerides | WMed | 59 | 3.6E-02 | 4.9E-02 | 4.6E-01 | 7.5E+01 | 3.0E+01 | NA | NA | NA | NA | NA | NA |
| Phosphate | Triglycerides | WMode | 59 | 5.6E-02 | 7.4E-02 | 4.5E-01 | 7.5E+01 | 3.0E+01 | NA | NA | NA | NA | NA | NA |
| Potassium in Urine | BMI | IVW | 13 | 5.2E-02 | 1.6E-01 | 7.5E-01 | 4.2E+01 | 3.0E+01 | 2.2E+02 | 6.9E-40 | 9.4E+01 | NA | NA | NA |
| Potassium in Urine | BMI | MR Egger | 13 | -6.2E-02 | 1.0E+00 | 9.5E-01 | 4.2E+01 | 3.0E+01 | NA | NA | NA | 1.9E-03 | 1.7E-02 | 9.1E-01 |

| | | | | | | | | | | | | | | |
|---|---|---|---|---|---|---|---|---|---|---|---|---|---|---|
| Potassium in Urine | BMI | WMed | 13 | 1.6E-01 | 8.3E-02 | 4.9E-02 | 4.2E+01 | 3.0E+01 | NA | NA | NA | NA | NA | NA |
| Potassium in Urine | BMI | WMode | 13 | 2.6E-01 | 1.3E-01 | 6.8E-02 | 4.2E+01 | 3.0E+01 | NA | NA | NA | NA | NA | NA |
| Potassium in Urine | DBP | IVW | 12 | 1.5E-01 | 1.3E+0 0 | 9.1E-01 | 4.3E+01 | 3.0E+01 | 1.6E+02 | 1.8E-28 | 9.2E+01 | NA | NA | NA |
| Potassium in Urine | DBP | MR Egger | 12 | -9.6E+00 | 7.9E+0 0 | 2.5E-01 | 4.3E+01 | 3.0E+01 | NA | NA | NA | 1.6E-01 | 1.3E-01 | 2.4E-01 |
| Potassium in Urine | DBP | WMed | 12 | 1.1E+00 | 6.7E-01 | 9.3E-02 | 4.3E+01 | 3.0E+01 | NA | NA | NA | NA | NA | NA |
| Potassium in Urine | DBP | WMode | 12 | 1.8E+00 | 9.5E-01 | 8.5E-02 | 4.3E+01 | 3.0E+01 | NA | NA | NA | NA | NA | NA |
| Potassium in Urine | HbA1c | IVW | 9 | 1.9E-01 | 9.0E-02 | 3.9E-02 | 4.1E+01 | 3.0E+01 | 8.7E+00 | 3.7E-01 | 0.0E+00 | NA | NA | NA |
| Potassium in Urine | HbA1c | MR Egger | 9 | 9.7E-01 | 4.8E-01 | 8.3E-02 | 4.1E+01 | 3.0E+01 | NA | NA | NA | -1.3E-02 | 7.7E-03 | 1.4E-01 |
| Potassium in Urine | HbA1c | WMed | 9 | 1.5E-01 | 1.1E-01 | 1.7E-01 | 4.1E+01 | 3.0E+01 | NA | NA | NA | NA | NA | NA |
| Potassium in Urine | HbA1c | WMode | 9 | 1.3E-01 | 1.6E-01 | 4.4E-01 | 4.1E+01 | 3.0E+01 | NA | NA | NA | NA | NA | NA |
| Potassium in Urine | HDL | IVW | 9 | -2.4E-01 | 1.9E-01 | 2.0E-01 | 4.1E+01 | 3.0E+01 | 2.5E+01 | 1.6E-03 | 6.8E+01 | NA | NA | NA |
| Potassium in Urine | HDL | MR Egger | 9 | -4.7E-02 | 1.2E+0 0 | 9.7E-01 | 4.1E+01 | 3.0E+01 | NA | NA | NA | -3.2E-03 | 1.9E-02 | 8.7E-01 |
| Potassium in Urine | HDL | WMed | 9 | -1.6E-01 | 1.7E-01 | 3.3E-01 | 4.1E+01 | 3.0E+01 | NA | NA | NA | NA | NA | NA |
| Potassium in Urine | HDL | WMode | 9 | -6.7E-02 | 2.1E-01 | 7.6E-01 | 4.1E+01 | 3.0E+01 | NA | NA | NA | NA | NA | NA |
| Potassium in Urine | LDL | IVW | 9 | -1.4E-01 | 2.4E-01 | 5.6E-01 | 4.1E+01 | 3.0E+01 | 3.3E+01 | 5.6E-05 | 7.6E+01 | NA | NA | NA |

| | | | | | | | | | | | | | | |
|---|---|---|---|---|---|---|---|---|---|---|---|---|---|---|
| Potassium in Urine | LDL | MR Egger | 9 | -1.4E-01 | 1.5E+00 | 9.3E-01 | 4.1E+01 | 3.0E+01 | NA | NA | NA | -5.2E-05 | 2.3E-02 | 1.0E+00 |
| Potassium in Urine | LDL | WMed | 9 | 1.8E-01 | 1.8E-01 | 3.3E-01 | 4.1E+01 | 3.0E+01 | NA | NA | NA | NA | NA | NA |
| Potassium in Urine | LDL | WMode | 9 | 1.9E-01 | 2.2E-01 | 4.2E-01 | 4.1E+01 | 3.0E+01 | NA | NA | NA | NA | NA | NA |
| Potassium in Urine | MetS | IVW | 16 | -4.3E-04 | 2.6E-01 | 1.0E+00 | 4.1E+01 | 3.0E+01 | 7.4E+01 | 9.4E-10 | 8.0E+01 | NA | NA | NA |
| Potassium in Urine | MetS | MR Egger | 16 | 4.4E-01 | 1.6E+00 | 7.8E-01 | 4.1E+01 | 3.0E+01 | NA | NA | NA | -7.0E-03 | 2.5E-02 | 7.8E-01 |
| Potassium in Urine | MetS | WMed | 16 | -2.9E-02 | 1.9E-01 | 8.8E-01 | 4.1E+01 | 3.0E+01 | NA | NA | NA | NA | NA | NA |
| Potassium in Urine | MetS | WMode | 16 | -1.5E-01 | 2.7E-01 | 5.8E-01 | 4.1E+01 | 3.0E+01 | NA | NA | NA | NA | NA | NA |
| Potassium in Urine | SBP | IVW | 12 | 3.4E+00 | 1.9E+00 | 7.6E-02 | 4.3E+01 | 3.0E+01 | 1.1E+02 | 1.6E-18 | 8.9E+01 | NA | NA | NA |
| Potassium in Urine | SBP | MR Egger | 12 | -8.8E+00 | 1.2E+01 | 4.6E-01 | 4.3E+01 | 3.0E+01 | NA | NA | NA | 2.0E-01 | 1.9E-01 | 3.1E-01 |
| Potassium in Urine | SBP | WMed | 12 | 3.0E+00 | 1.2E+00 | 1.0E-02 | 4.3E+01 | 3.0E+01 | NA | NA | NA | NA | NA | NA |
| Potassium in Urine | SBP | WMode | 12 | 2.6E+00 | 1.9E+00 | 2.0E-01 | 4.3E+01 | 3.0E+01 | NA | NA | NA | NA | NA | NA |
| Potassium in Urine | T2D | IVW | 8 | 7.9E-02 | 3.0E-01 | 7.9E-01 | 4.1E+01 | 3.0E+01 | 1.6E+01 | 2.1E-02 | 5.7E+01 | NA | NA | NA |
| Potassium in Urine | T2D | MR Egger | 8 | -3.7E-01 | 1.8E+00 | 8.5E-01 | 4.1E+01 | 3.0E+01 | NA | NA | NA | 7.2E-03 | 2.9E-02 | 8.1E-01 |
| Potassium in Urine | T2D | WMed | 8 | 3.6E-01 | 2.9E-01 | 2.1E-01 | 4.1E+01 | 3.0E+01 | NA | NA | NA | NA | NA | NA |
| Potassium in Urine | T2D | WMode | 8 | 6.5E-01 | 4.8E-01 | 2.2E-01 | 4.1E+01 | 3.0E+01 | NA | NA | NA | NA | NA | NA |

| | | | | | | | | | | | | | |
|---|---|---|---|---|---|---|---|---|---|---|---|---|---|
| Potassium in Urine | Triglycerides | IVW | 9 | -3.7E-01 | 2.9E-01 | 2.0E-01 | 4.1E+01 | 3.0E+01 | 6.0E+01 | 4.0E-10 | 8.4E+01 | NA | NA | NA |
| Potassium in Urine | Triglycerides | MR Egger | 9 | -2.2E+00 | 1.6E+00 | 2.2E-01 | 4.1E+01 | 3.0E+01 | NA | NA | NA | 3.0E-02 | 2.6E-02 | 2.9E-01 |
| Potassium in Urine | Triglycerides | WMed | 9 | -2.8E-01 | 1.5E-01 | 6.6E-02 | 4.1E+01 | 3.0E+01 | NA | NA | NA | NA | NA | NA |
| Potassium in Urine | Triglycerides | WMode | 9 | -2.4E-01 | 1.6E-01 | 1.8E-01 | 4.1E+01 | 3.0E+01 | NA | NA | NA | NA | NA | NA |
| Red Cell Distribution Width | BMI | IVW | 208 | 1.2E-01 | 1.8E-01 | 4.9E-01 | 2.2E+02 | 3.9E+01 | 4.8E-02 | 1.0E+00 | 0.0E+00 | NA | NA | NA |
| Red Cell Distribution Width | BMI | MR Egger | 208 | 5.0E-02 | 4.4E-01 | 9.1E-01 | 2.2E+02 | 3.9E+01 | NA | NA | NA | 1.5E-03 | 8.6E-03 | 8.6E-01 |
| Red Cell Distribution Width | BMI | WMed | 208 | 1.4E-01 | 2.5E-01 | 5.6E-01 | 2.2E+02 | 3.9E+01 | NA | NA | NA | NA | NA | NA |
| Red Cell Distribution Width | BMI | WMode | 208 | 1.5E-01 | 7.7E+00 | 9.8E-01 | 2.2E+02 | 3.9E+01 | NA | NA | NA | NA | NA | NA |
| Red Cell Distribution Width | DBP | IVW | 186 | 6.5E-01 | 1.0E-02 | 0.0E+00 | 2.2E+02 | 3.9E+01 | 5.1E+00 | 1.0E+00 | 0.0E+00 | NA | NA | NA |
| Red Cell Distribution Width | DBP | MR Egger | 186 | 2.4E-01 | 2.8E-01 | 3.8E-01 | 2.2E+02 | 3.9E+01 | NA | NA | NA | 2.0E-02 | 1.3E-02 | 1.4E-01 |
| Red Cell Distribution Width | DBP | WMed | 186 | 6.5E-01 | 4.9E-02 | 6.6E-40 | 2.2E+02 | 3.9E+01 | NA | NA | NA | NA | NA | NA |
| Red Cell Distribution Width | DBP | WMode | 186 | 6.5E-01 | 4.7E+00 | 8.9E-01 | 2.2E+02 | 3.9E+01 | NA | NA | NA | NA | NA | NA |
| Red Cell Distribution Width | HbA1C | IVW | 81 | 6.1E-02 | 1.4E-01 | 6.6E-01 | 2.0E+02 | 3.9E+01 | 1.4E-01 | 1.0E+00 | 0.0E+00 | NA | NA | NA |
| Red Cell Distribution Width | HbA1C | MR Egger | 81 | 8.0E-03 | 2.1E-01 | 9.7E-01 | 2.0E+02 | 3.9E+01 | NA | NA | NA | 3.8E-03 | 1.1E-02 | 7.3E-01 |
| Red Cell Distribution Width | HbA1C | WMed | 81 | 4.3E-02 | 1.5E-01 | 7.7E-01 | 2.0E+02 | 3.9E+01 | NA | NA | NA | NA | NA | NA |

| | | | | | | | | | | | | | | |
|---|---|---|---|---|---|---|---|---|---|---|---|---|---|---|
| Red Cell Distribution Width | HbA1C | WMode | 81 | 4.0E-02 | 2.8E-01 | 8.8E-01 | 2.0E+02 | 3.9E+01 | NA | NA | NA | NA | NA | NA |
| Red Cell Distribution Width | HDL | IVW | 88 | 6.7E-02 | 6.0E-02 | 2.6E-01 | 2.1E+02 | 3.9E+01 | 1.4E-01 | 1.0E+00 | 0.0E+00 | NA | NA | NA |
| Red Cell Distribution Width | HDL | MR Egger | 88 | 2.4E-02 | 1.5E-01 | 8.7E-01 | 2.1E+02 | 3.9E+01 | NA | NA | NA | 4.8E-03 | 1.5E-02 | 7.4E-01 |
| Red Cell Distribution Width | HDL | WMed | 88 | 6.5E-02 | 6.2E-02 | 3.0E-01 | 2.1E+02 | 3.9E+01 | NA | NA | NA | NA | NA | NA |
| Red Cell Distribution Width | HDL | WMode | 88 | 6.5E-02 | 2.4E+00 | 9.8E-01 | 2.1E+02 | 3.9E+01 | NA | NA | NA | NA | NA | NA |
| Red Cell Distribution Width | LDL | IVW | 88 | 3.7E-01 | 1.1E-01 | 5.6E-04 | 2.1E+02 | 3.9E+01 | 6.2E-02 | 1.0E+00 | 0.0E+00 | NA | NA | NA |
| Red Cell Distribution Width | LDL | MR Egger | 88 | 1.4E-01 | 1.2E+00 | 9.1E-01 | 2.1E+02 | 3.9E+01 | NA | NA | NA | 3.6E-03 | 1.9E-02 | 8.5E-01 |
| Red Cell Distribution Width | LDL | WMed | 88 | 3.8E-01 | 1.3E-01 | 2.5E-03 | 2.1E+02 | 3.9E+01 | NA | NA | NA | NA | NA | NA |
| Red Cell Distribution Width | LDL | WMode | 88 | 3.8E-01 | 1.7E+00 | 8.2E-01 | 2.1E+02 | 3.9E+01 | NA | NA | NA | NA | NA | NA |
| Red Cell Distribution Width | MetS | IVW | 81 | -5.6E-03 | 2.0E-02 | 7.8E-01 | 1.9E+02 | 3.9E+01 | 1.3E+03 | 4.4E-214 | 9.4E+01 | NA | NA | NA |
| Red Cell Distribution Width | MetS | MR Egger | 81 | -1.6E-02 | 3.7E-02 | 6.6E-01 | 1.9E+02 | 3.9E+01 | NA | NA | NA | 3.8E-04 | 1.1E-03 | 7.3E-01 |
| Red Cell Distribution Width | MetS | WMed | 81 | -2.5E-02 | 8.9E-03 | 5.5E-03 | 1.9E+02 | 3.9E+01 | NA | NA | NA | NA | NA | NA |
| Red Cell Distribution Width | MetS | WMode | 81 | -2.2E-02 | 7.9E-03 | 7.7E-03 | 1.9E+02 | 3.9E+01 | NA | NA | NA | NA | NA | NA |
| Red Cell Distribution Width | SBP | IVW | 186 | 1.2E+00 | 3.1E-02 | 0.0E+00 | 2.2E+02 | 3.9E+01 | 2.6E+02 | 2.0E-04 | 0.0E+00 | NA | NA | NA |
| Red Cell Distribution Width | SBP | MR Egger | 186 | 5.0E-01 | 7.0E-02 | 1.9E-11 | 2.2E+02 | 3.9E+01 | NA | NA | NA | 2.0E-02 | 1.9E-03 | 1.2E-20 |

| | | | | | | | | | | | | | | |
|---|---|---|---|---|---|---|---|---|---|---|---|---|---|---|
| Red Cell Distribution Width | SBP | WMed | 186 | 9.0E-01 | 6.2E-02 | 1.6E-47 | 2.2E+02 | 3.9E+01 | NA | NA | NA | NA | NA | NA |
| Red Cell Distribution Width | SBP | WMode | 186 | 9.0E-01 | 3.0E+00 | 7.7E-01 | 2.2E+02 | 3.9E+01 | NA | NA | NA | NA | NA | NA |
| Red Cell Distribution Width | T2D | IVW | 213 | 5.2E-01 | 6.2E-02 | 4.5E-17 | 2.0E+02 | 3.9E+01 | 3.3E+02 | 1.6E-07 | 3.1E+01 | NA | NA | NA |
| Red Cell Distribution Width | T2D | MR Egger | 213 | 1.9E-01 | 9.8E-02 | 5.3E-02 | 2.0E+02 | 3.9E+01 | NA | NA | NA | 2.2E-02 | 5.1E-03 | 3.3E-05 |
| Red Cell Distribution Width | T2D | WMed | 213 | 4.8E-01 | 5.3E-02 | 6.6E-20 | 2.0E+02 | 3.9E+01 | NA | NA | NA | NA | NA | NA |
| Red Cell Distribution Width | T2D | WMode | 213 | 4.7E-01 | 2.1E-01 | 2.9E-02 | 2.0E+02 | 3.9E+01 | NA | NA | NA | NA | NA | NA |
| Red Cell Distribution Width | Triglycerides | IVW | 88 | 2.0E-01 | 1.3E-01 | 1.1E-01 | 2.1E+02 | 3.9E+01 | 3.5E-01 | 1.0E+00 | 0.0E+00 | NA | NA | NA |
| Red Cell Distribution Width | Triglycerides | MR Egger | 88 | 2.0E-02 | 3.4E-01 | 9.5E-01 | 2.1E+02 | 3.9E+01 | NA | NA | NA | 4.5E-03 | 7.7E-03 | 5.7E-01 |
| Red Cell Distribution Width | Triglycerides | WMed | 88 | 2.4E-01 | 1.5E-01 | 1.1E-01 | 2.1E+02 | 3.9E+01 | NA | NA | NA | NA | NA | NA |
| Red Cell Distribution Width | Triglycerides | WMode | 88 | 2.4E-01 | 5.4E+00 | 9.6E-01 | 2.1E+02 | 3.9E+01 | NA | NA | NA | NA | NA | NA |
| Serum Phosphate | BMI | IVW | 284 | -4.6E-02 | 1.8E-02 | 1.2E-02 | 8.8E+01 | 2.9E+01 | 1.7E+03 | 1.7E-208 | 8.4E+01 | NA | NA | NA |
| Serum Phosphate | BMI | MR Egger | 284 | -9.4E-02 | 4.1E-02 | 2.3E-02 | 8.8E+01 | 2.9E+01 | NA | NA | NA | 9.9E-04 | 7.7E-04 | 2.0E-01 |
| Serum Phosphate | BMI | WMed | 284 | -6.0E-02 | 1.5E-02 | 1.1E-04 | 8.8E+01 | 2.9E+01 | NA | NA | NA | NA | NA | NA |
| Serum Phosphate | BMI | WMode | 284 | -7.4E-02 | 3.5E-02 | 3.4E-02 | 8.8E+01 | 2.9E+01 | NA | NA | NA | NA | NA | NA |
| Serum Phosphate | DBP | IVW | 249 | -3.9E-01 | 2.4E-01 | 1.0E-01 | 9.1E+01 | 2.9E+01 | 3.0E+03 | 0.0E+00 | 9.2E+01 | NA | NA | NA |
| Serum Phosphate | DBP | MR Egger | 249 | -4.1E-02 | 5.4E-01 | 9.4E-01 | 9.1E+01 | 2.9E+01 | NA | NA | NA | -7.4E-03 | 1.0E-02 | 4.7E-01 |
| Serum Phosphate | DBP | WMed | 249 | -2.0E-01 | 1.9E-01 | 2.8E-01 | 9.1E+01 | 2.9E+01 | NA | NA | NA | NA | NA | NA |
| Serum Phosphate | DBP | WMode | 249 | 3.9E-01 | 6.6E-01 | 5.6E-01 | 9.1E+01 | 2.9E+01 | NA | NA | NA | NA | NA | NA |

| | | | | | | | | | | | | | | |
|---|---|---|---|---|---|---|---|---|---|---|---|---|---|---|
| Serum Phosphate | HbA1c | IVW | 98 | -1.2E-02 | 2.8E-02 | 6.5E-01 | 9.2E+01 | 2.9E+01 | 1.4E+02 | 1.2E-03 | 3.3E+01 | NA | NA | NA |
| Serum Phosphate | HbA1c | MR Egger | 98 | -3.0E-02 | 7.1E-02 | 6.7E-01 | 9.2E+01 | 2.9E+01 | NA | NA | NA | 3.4E-04 | 1.2E-03 | 7.9E-01 |
| Serum Phosphate | HbA1c | WMed | 98 | -1.2E-02 | 3.7E-02 | 7.5E-01 | 9.2E+01 | 2.9E+01 | NA | NA | NA | NA | NA | NA |
| Serum Phosphate | HbA1c | WMode | 98 | -4.6E-03 | 5.1E-02 | 9.3E-01 | 9.2E+01 | 2.9E+01 | NA | NA | NA | NA | NA | NA |
| Serum Phosphate | HDL | IVW | 108 | -9.4E-02 | 5.8E-02 | 1.1E-01 | 9.3E+01 | 2.9E+01 | 5.1E+02 | 5.4E-53 | 7.9E+01 | NA | NA | NA |
| Serum Phosphate | HDL | MR Egger | 108 | -4.3E-02 | 1.4E-01 | 7.7E-01 | 9.3E+01 | 2.9E+01 | NA | NA | NA | -1.0E-03 | 2.7E-03 | 7.0E-01 |
| Serum Phosphate | HDL | WMed | 108 | 1.4E-02 | 4.9E-02 | 7.7E-01 | 9.3E+01 | 2.9E+01 | NA | NA | NA | NA | NA | NA |
| Serum Phosphate | HDL | WMode | 108 | 1.8E-01 | 1.5E-01 | 2.3E-01 | 9.3E+01 | 2.9E+01 | NA | NA | NA | NA | NA | NA |
| Serum Phosphate | LDL | IVW | 108 | 2.2E-01 | 1.0E-01 | 2.8E-02 | 9.3E+01 | 2.9E+01 | 1.3E+03 | 2.5E-202 | 9.2E+01 | NA | NA | NA |
| Serum Phosphate | LDL | MR Egger | 108 | 3.3E-01 | 2.5E-01 | 1.9E-01 | 9.3E+01 | 2.9E+01 | NA | NA | NA | -2.1E-03 | 4.6E-03 | 6.4E-01 |
| Serum Phosphate | LDL | WMed | 108 | -2.4E-02 | 5.2E-02 | 6.4E-01 | 9.3E+01 | 2.9E+01 | NA | NA | NA | NA | NA | NA |
| Serum Phosphate | LDL | WMode | 108 | -7.7E-02 | 5.8E-02 | 1.9E-01 | 9.3E+01 | 2.9E+01 | NA | NA | NA | NA | NA | NA |
| Serum Phosphate | MetS | IVW | 317 | -1.5E-02 | 4.5E-02 | 7.5E-01 | 8.7E+01 | 2.9E+01 | 1.2E+03 | 8.0E-103 | 7.4E+01 | NA | NA | NA |
| Serum Phosphate | MetS | MR Egger | 317 | 7.9E-02 | 1.0E-01 | 4.4E-01 | 8.7E+01 | 2.9E+01 | NA | NA | NA | -2.0E-03 | 1.9E-03 | 3.1E-01 |
| Serum Phosphate | MetS | WMed | 317 | 9.1E-02 | 4.3E-02 | 3.5E-02 | 8.7E+01 | 2.9E+01 | NA | NA | NA | NA | NA | NA |
| Serum Phosphate | MetS | WMode | 317 | 1.5E-01 | 6.2E-02 | 1.9E-02 | 8.7E+01 | 2.9E+01 | NA | NA | NA | NA | NA | NA |
| Serum Phosphate | SBP | IVW | 248 | -4.0E-02 | 4.0E-01 | 9.2E-01 | 9.1E+01 | 2.9E+01 | 2.8E+03 | 0.0E+00 | 9.1E+01 | NA | NA | NA |
| Serum Phosphate | SBP | MR Egger | 248 | 4.9E-01 | 9.0E-01 | 5.9E-01 | 9.1E+01 | 2.9E+01 | NA | NA | NA | -1.1E-02 | 1.7E-02 | 5.1E-01 |
| Serum Phosphate | SBP | WMed | 248 | 4.1E-01 | 3.5E-01 | 2.4E-01 | 9.1E+01 | 2.9E+01 | NA | NA | NA | NA | NA | NA |
| Serum Phosphate | SBP | WMode | 248 | 1.4E+00 | 1.2E+0 0 | 2.3E-01 | 9.1E+01 | 2.9E+01 | NA | NA | NA | NA | NA | NA |
| Serum Phosphate | T2D | IVW | 189 | 6.6E-02 | 5.7E-02 | 2.5E-01 | 9.5E+01 | 2.9E+01 | 5.2E+02 | 4.8E-33 | 6.4E+01 | NA | NA | NA |
| Serum Phosphate | T2D | MR Egger | 189 | -1.4E-02 | 1.3E-01 | 9.2E-01 | 9.5E+01 | 2.9E+01 | NA | NA | NA | 1.7E-03 | 2.5E-03 | 5.0E-01 |
| Serum Phosphate | T2D | WMed | 189 | 1.0E-01 | 6.4E-02 | 1.1E-01 | 9.5E+01 | 2.9E+01 | NA | NA | NA | NA | NA | NA |
| Serum Phosphate | T2D | WMode | 189 | 1.2E-01 | 9.4E-02 | 2.2E-01 | 9.5E+01 | 2.9E+01 | NA | NA | NA | NA | NA | NA |

| | | | | | | | | | | | | | | |
|---|---|---|---|---|---|---|---|---|---|---|---|---|---|---|
| Serum Phosphate | Triglycerides | IVW | 108 | 3.0E-01 | 9.7E-02 | 1.8E-03 | 9.3E+01 | 2.9E+01 | 1.5E+03 | 6.0E-243 | 9.3E+01 | NA | NA | NA |
| Serum Phosphate | Triglycerides | MR Egger | 108 | 6.6E-01 | 2.4E-01 | 6.8E-03 | 9.3E+01 | 2.9E+01 | NA | NA | NA | -7.2E-03 | 4.4E-03 | 1.1E-01 |
| Serum Phosphate | Triglycerides | WMed | 108 | -1.8E-02 | 4.7E-02 | 7.1E-01 | 9.3E+01 | 2.9E+01 | NA | NA | NA | NA | NA | NA |
| Serum Phosphate | Triglycerides | WMode | 108 | -7.6E-02 | 6.2E-02 | 2.3E-01 | 9.3E+01 | 2.9E+01 | NA | NA | NA | NA | NA | NA |
| Serum Urea | BMI | IVW | 172 | 5.2E-02 | 2.1E-02 | 1.4E-02 | 8.5E+01 | 2.7E+01 | 1.3E+03 | 1.5E-167 | 8.7E+01 | NA | NA | NA |
| Serum Urea | BMI | MR Egger | 172 | 6.9E-02 | 4.8E-02 | 1.5E-01 | 8.5E+01 | 2.7E+01 | NA | NA | NA | -4.1E-04 | 1.1E-03 | 7.0E-01 |
| Serum Urea | BMI | WMed | 172 | 2.8E-02 | 1.6E-02 | 8.0E-02 | 8.5E+01 | 2.7E+01 | NA | NA | NA | NA | NA | NA |
| Serum Urea | BMI | WMode | 172 | 1.3E-02 | 2.2E-02 | 5.6E-01 | 8.5E+01 | 2.7E+01 | NA | NA | NA | NA | NA | NA |
| Serum Urea | DBP | IVW | 160 | -8.1E-01 | 2.7E-01 | 2.9E-03 | 8.6E+01 | 2.8E+01 | 2.3E+03 | 0.0E+00 | 9.3E+01 | NA | NA | NA |
| Serum Urea | DBP | MR Egger | 160 | -8.3E-01 | 6.0E-01 | 1.7E-01 | 8.6E+01 | 2.8E+01 | NA | NA | NA | 5.4E-04 | 1.4E-02 | 9.7E-01 |
| Serum Urea | DBP | WMed | 160 | -4.8E-01 | 1.7E-01 | 4.6E-03 | 8.6E+01 | 2.8E+01 | NA | NA | NA | NA | NA | NA |
| Serum Urea | DBP | WMode | 160 | -2.0E-01 | 3.9E-01 | 6.1E-01 | 8.6E+01 | 2.8E+01 | NA | NA | NA | NA | NA | NA |
| Serum Urea | HbA1c | IVW | 103 | -5.0E-02 | 2.1E-02 | 1.4E-02 | 8.9E+01 | 2.8E+01 | 1.4E+02 | 1.2E-02 | 2.5E+01 | NA | NA | NA |
| Serum Urea | HbA1c | MR Egger | 103 | -2.0E-02 | 4.6E-02 | 6.7E-01 | 8.9E+01 | 2.8E+01 | NA | NA | NA | -7.4E-04 | 1.0E-03 | 4.7E-01 |
| Serum Urea | HbA1c | WMed | 103 | -9.7E-03 | 2.9E-02 | 7.4E-01 | 8.9E+01 | 2.8E+01 | NA | NA | NA | NA | NA | NA |
| Serum Urea | HbA1c | WMode | 103 | -1.3E-02 | 4.6E-02 | 7.8E-01 | 8.9E+01 | 2.8E+01 | NA | NA | NA | NA | NA | NA |
| Serum Urea | HDL | IVW | 107 | -4.5E-03 | 3.8E-02 | 9.0E-01 | 9.0E+01 | 2.8E+01 | 3.1E+02 | 1.6E-21 | 6.6E+01 | NA | NA | NA |
| Serum Urea | HDL | MR Egger | 107 | -2.4E-02 | 8.8E-02 | 7.8E-01 | 9.0E+01 | 2.8E+01 | NA | NA | NA | 4.9E-04 | 1.9E-03 | 8.0E-01 |
| Serum Urea | HDL | WMed | 107 | 9.6E-03 | 4.0E-02 | 8.1E-01 | 9.0E+01 | 2.8E+01 | NA | NA | NA | NA | NA | NA |
| Serum Urea | HDL | WMode | 107 | 4.3E-02 | 4.8E-02 | 3.7E-01 | 9.0E+01 | 2.8E+01 | NA | NA | NA | NA | NA | NA |
| Serum Urea | LDL | IVW | 107 | -2.7E-02 | 5.4E-02 | 6.2E-01 | 9.0E+01 | 2.8E+01 | 5.4E+02 | 9.7E-59 | 8.0E+01 | NA | NA | NA |
| Serum Urea | LDL | MR Egger | 107 | 8.6E-02 | 1.2E-01 | 4.9E-01 | 9.0E+01 | 2.8E+01 | NA | NA | NA | -2.8E-03 | 2.8E-03 | 3.2E-01 |

| | | | | | | | | | | | | | | |
|---|---|---|---|---|---|---|---|---|---|---|---|---|---|---|
| Serum Urea | LDL | WMed | 107 | -1.9E-02 | 4.1E-02 | 6.3E-01 | 9.0E+01 | 2.8E+01 | NA | NA | NA | NA | NA | NA |
| Serum Urea | LDL | WMode | 107 | 2.0E-03 | 5.9E-02 | 9.7E-01 | 9.0E+01 | 2.8E+01 | NA | NA | NA | NA | NA | NA |
| Serum Urea | MetS | IVW | 175 | 8.2E-02 | 4.4E-02 | 6.3E-02 | 8.6E+01 | 2.8E+01 | 5.1E+02 | 3.0E-35 | 6.6E+01 | NA | NA | NA |
| Serum Urea | MetS | MR Egger | 175 | 6.3E-02 | 9.9E-02 | 5.2E-01 | 8.6E+01 | 2.8E+01 | NA | NA | NA | 4.8E-04 | 2.2E-03 | 8.3E-01 |
| Serum Urea | MetS | WMed | 175 | 5.4E-02 | 4.5E-02 | 2.3E-01 | 8.6E+01 | 2.8E+01 | NA | NA | NA | NA | NA | NA |
| Serum Urea | MetS | WMode | 175 | 2.4E-02 | 6.7E-02 | 7.2E-01 | 8.6E+01 | 2.8E+01 | NA | NA | NA | NA | NA | NA |
| Serum Urea | SBP | IVW | 160 | -1.5E+00 | 4.6E-01 | 1.3E-03 | 8.6E+01 | 2.8E+01 | 2.2E+03 | 0.0E+00 | 9.3E+01 | NA | NA | NA |
| Serum Urea | SBP | MR Egger | 160 | -1.1E+00 | 1.0E+00 | 2.8E-01 | 8.6E+01 | 2.8E+01 | NA | NA | NA | -9.8E-03 | 2.3E-02 | 6.7E-01 |
| Serum Urea | SBP | WMed | 160 | -8.9E-01 | 3.1E-01 | 3.8E-03 | 8.6E+01 | 2.8E+01 | NA | NA | NA | NA | NA | NA |
| Serum Urea | SBP | WMode | 160 | 6.7E-01 | 5.2E-01 | 2.0E-01 | 8.6E+01 | 2.8E+01 | NA | NA | NA | NA | NA | NA |
| Serum Urea | T2D | IVW | 135 | 2.7E-02 | 7.7E-02 | 7.3E-01 | 8.5E+01 | 2.8E+01 | 6.6E+02 | 1.4E-70 | 8.0E+01 | NA | NA | NA |
| Serum Urea | T2D | MR Egger | 135 | 1.1E-01 | 1.8E-01 | 5.3E-01 | 8.5E+01 | 2.8E+01 | NA | NA | NA | -2.1E-03 | 3.9E-03 | 5.9E-01 |
| Serum Urea | T2D | WMed | 135 | 5.3E-02 | 6.7E-02 | 4.3E-01 | 8.5E+01 | 2.8E+01 | NA | NA | NA | NA | NA | NA |
| Serum Urea | T2D | WMode | 135 | 5.3E-02 | 9.8E-02 | 5.9E-01 | 8.5E+01 | 2.8E+01 | NA | NA | NA | NA | NA | NA |
| Serum Urea | Triglycerides | IVW | 107 | -1.6E-01 | 7.6E-02 | 3.6E-02 | 9.0E+01 | 2.8E+01 | 1.3E+03 | 3.1E-206 | 9.2E+01 | NA | NA | NA |
| Serum Urea | Triglycerides | MR Egger | 107 | -1.9E-01 | 1.8E-01 | 2.8E-01 | 9.0E+01 | 2.8E+01 | NA | NA | NA | 7.6E-04 | 3.9E-03 | 8.5E-01 |
| Serum Urea | Triglycerides | WMed | 107 | -3.4E-02 | 3.8E-02 | 3.7E-01 | 9.0E+01 | 2.8E+01 | NA | NA | NA | NA | NA | NA |
| Serum Urea | Triglycerides | WMode | 107 | -1.9E-02 | 4.7E-02 | 6.8E-01 | 9.0E+01 | 2.8E+01 | NA | NA | NA | NA | NA | NA |
| Serum Uric Acid | BMI | IVW | 200 | -8.0E-02 | 3.0E-02 | 8.1E-03 | 8.8E+01 | 3.0E+01 | 2.7E+03 | 0.0E+00 | 9.2E+01 | NA | NA | NA |
| Serum Uric Acid | BMI | MR Egger | 200 | 9.4E-02 | 6.6E-02 | 1.6E-01 | 8.8E+01 | 3.0E+01 | NA | NA | NA | -4.0E-03 | 1.4E-03 | 3.6E-03 |
| Serum Uric Acid | BMI | WMed | 200 | -1.1E-02 | 1.7E-02 | 5.1E-01 | 8.8E+01 | 3.0E+01 | NA | NA | NA | NA | NA | NA |

| | | | | | | | | | | | | | | |
|---|---|---|---|---|---|---|---|---|---|---|---|---|---|---|
| Serum Uric Acid | BMI | WMode | 200 | -1.2E-03 | 2.2E-02 | 9.6E-01 | 8.8E+01 | 3.0E+01 | NA | NA | NA | NA | NA | NA |
| Serum Uric Acid | DBP | IVW | 171 | -1.4E+00 | 2.7E-01 | 3.9E-07 | 9.0E+01 | 3.0E+01 | 2.1E+03 | 0.0E+00 | 9.2E+01 | NA | NA | NA |
| Serum Uric Acid | DBP | MR Egger | 171 | -1.6E+00 | 6.3E-01 | 1.5E-02 | 9.0E+01 | 3.0E+01 | NA | NA | NA | 3.8E-03 | 1.3E-02 | 7.7E-01 |
| Serum Uric Acid | DBP | WMed | 171 | -8.4E-01 | 1.6E-01 | 3.3E-07 | 9.0E+01 | 3.0E+01 | NA | NA | NA | NA | NA | NA |
| Serum Uric Acid | DBP | WMode | 171 | -9.6E-01 | 1.8E-01 | 2.0E-07 | 9.0E+01 | 3.0E+01 | NA | NA | NA | NA | NA | NA |
| Serum Uric Acid | HbA1c | IVW | 84 | 2.6E-03 | 2.5E-02 | 9.2E-01 | 1.0E+02 | 3.0E+01 | 1.1E+02 | 1.4E-02 | 2.4E+01 | NA | NA | NA |
| Serum Uric Acid | HbA1c | MR Egger | 84 | 8.7E-02 | 5.3E-02 | 1.0E-01 | 1.0E+02 | 3.0E+01 | NA | NA | NA | -2.0E-03 | 1.1E-03 | 7.6E-02 |
| Serum Uric Acid | HbA1c | WMed | 84 | 7.6E-02 | 3.4E-02 | 2.4E-02 | 1.0E+02 | 3.0E+01 | NA | NA | NA | NA | NA | NA |
| Serum Uric Acid | HbA1c | WMode | 84 | 8.6E-02 | 4.0E-02 | 3.4E-02 | 1.0E+02 | 3.0E+01 | NA | NA | NA | NA | NA | NA |
| Serum Uric Acid | HDL | IVW | 95 | 1.0E-01 | 5.3E-02 | 5.6E-02 | 1.1E+02 | 3.0E+01 | 4.7E+02 | 4.6E-51 | 8.0E+01 | NA | NA | NA |
| Serum Uric Acid | HDL | MR Egger | 95 | 3.9E-02 | 1.1E-01 | 7.1E-01 | 1.1E+02 | 3.0E+01 | NA | NA | NA | 1.6E-03 | 2.4E-03 | 5.0E-01 |
| Serum Uric Acid | HDL | WMed | 95 | 5.4E-02 | 4.5E-02 | 2.2E-01 | 1.1E+02 | 3.0E+01 | NA | NA | NA | NA | NA | NA |
| Serum Uric Acid | HDL | WMode | 95 | 8.0E-02 | 3.9E-02 | 4.4E-02 | 1.1E+02 | 3.0E+01 | NA | NA | NA | NA | NA | NA |
| Serum Uric Acid | LDL | IVW | 95 | 2.8E-02 | 5.4E-02 | 6.1E-01 | 1.1E+02 | 3.0E+01 | 4.3E+02 | 7.2E-44 | 7.8E+01 | NA | NA | NA |
| Serum Uric Acid | LDL | MR Egger | 95 | -2.1E-02 | 1.1E-01 | 8.5E-01 | 1.1E+02 | 3.0E+01 | NA | NA | NA | 1.3E-03 | 2.5E-03 | 6.1E-01 |
| Serum Uric Acid | LDL | WMed | 95 | 6.5E-02 | 4.5E-02 | 1.5E-01 | 1.1E+02 | 3.0E+01 | NA | NA | NA | NA | NA | NA |
| Serum Uric Acid | LDL | WMode | 95 | 4.2E-02 | 6.1E-02 | 5.0E-01 | 1.1E+02 | 3.0E+01 | NA | NA | NA | NA | NA | NA |
| Serum Uric Acid | MetS | IVW | 222 | 2.0E-01 | 3.8E-02 | 1.2E-07 | 1.7E+02 | 3.0E+01 | 1.0E+03 | 2.6E-105 | 7.6E+01 | NA | NA | NA |
| Serum Uric Acid | MetS | MR Egger | 222 | 4.6E-02 | 4.9E-02 | 3.5E-01 | 1.7E+02 | 3.0E+01 | NA | NA | NA | 7.0E-03 | 1.5E-03 | 2.6E-06 |
| Serum Uric Acid | MetS | WMed | 222 | 5.7E-02 | 2.4E-02 | 1.8E-02 | 1.7E+02 | 3.0E+01 | NA | NA | NA | NA | NA | NA |
| Serum Uric Acid | MetS | WMode | 222 | 4.9E-02 | 2.3E-02 | 3.0E-02 | 1.7E+02 | 3.0E+01 | NA | NA | NA | NA | NA | NA |
| Serum Uric Acid | SBP | IVW | 171 | -2.3E+00 | 4.4E-01 | 9.0E-08 | 9.0E+01 | 3.0E+01 | 1.8E+03 | 3.0E-265 | 9.0E+01 | NA | NA | NA |
| Serum Uric Acid | SBP | MR Egger | 171 | -2.8E+00 | 1.0E+00 | 5.4E-03 | 9.0E+01 | 3.0E+01 | NA | NA | NA | 1.1E-02 | 2.0E-02 | 5.8E-01 |
| Serum Uric Acid | SBP | WMed | 171 | -2.1E+00 | 3.2E-01 | 5.7E-11 | 9.0E+01 | 3.0E+01 | NA | NA | NA | NA | NA | NA |

| | | | | | | | | | | | | | | |
|---|---|---|---|---|---|---|---|---|---|---|---|---|---|---|
| Serum Uric Acid | SBP | WMode | 171 | -2.4E+00 | 3.1E-01 | 7.4E-13 | 9.0E+01 | 3.0E+01 | NA | NA | NA | NA | NA | NA |
| Serum Uric Acid | T2D | IVW | 142 | -9.4E-02 | 8.7E-02 | 2.8E-01 | 9.7E+01 | 3.0E+01 | 8.2E+02 | 2.2E-95 | 8.2E+01 | NA | NA | NA |
| Serum Uric Acid | T2D | MR Egger | 142 | 4.3E-01 | 1.9E-01 | 2.7E-02 | 9.7E+01 | 3.0E+01 | NA | NA | NA | -1.2E-02 | 3.9E-03 | 2.7E-03 |
| Serum Uric Acid | T2D | WMed | 142 | -1.1E-01 | 6.6E-02 | 1.0E-01 | 9.7E+01 | 3.0E+01 | NA | NA | NA | NA | NA | NA |
| Serum Uric Acid | T2D | WMode | 142 | -1.1E-01 | 7.6E-02 | 1.4E-01 | 9.7E+01 | 3.0E+01 | NA | NA | NA | NA | NA | NA |
| Serum Uric Acid | Triglycerides | IVW | 94 | -3.6E-01 | 9.2E-02 | 7.8E-05 | 1.1E+02 | 3.0E+01 | 1.5E+03 | 5.3E-249 | 9.4E+01 | NA | NA | NA |
| Serum Uric Acid | Triglycerides | MR Egger | 94 | -5.3E-01 | 1.8E-01 | 4.4E-03 | 1.1E+02 | 3.0E+01 | NA | NA | NA | 4.6E-03 | 4.2E-03 | 2.8E-01 |
| Serum Uric Acid | Triglycerides | WMed | 94 | -4.8E-03 | 3.9E-02 | 9.0E-01 | 1.1E+02 | 3.0E+01 | NA | NA | NA | NA | NA | NA |
| Serum Uric Acid | Triglycerides | WMode | 94 | 1.0E-02 | 4.0E-02 | 8.0E-01 | 1.1E+02 | 3.0E+01 | NA | NA | NA | NA | NA | NA |
| Sodium in Urine | BMI | IVW | 38 | -6.0E-01 | 1.6E-01 | 1.7E-04 | 4.4E+01 | 3.0E+01 | 1.9E+03 | 0.0E+00 | 9.8E+01 | NA | NA | NA |
| Sodium in Urine | BMI | MR Egger | 38 | 3.7E-02 | 6.1E-01 | 9.5E-01 | 4.4E+01 | 3.0E+01 | NA | NA | NA | -1.1E-02 | 1.0E-02 | 2.8E-01 |
| Sodium in Urine | BMI | WMed | 38 | -2.7E-01 | 6.2E-02 | 1.4E-05 | 4.4E+01 | 3.0E+01 | NA | NA | NA | NA | NA | NA |
| Sodium in Urine | BMI | WMode | 38 | -3.8E-01 | 8.9E-02 | 1.5E-04 | 4.4E+01 | 3.0E+01 | NA | NA | NA | NA | NA | NA |
| Sodium in Urine | DBP | IVW | 32 | -7.3E-01 | 8.3E-01 | 3.8E-01 | 4.6E+01 | 3.0E+01 | 4.8E+02 | 4.0E-81 | 9.3E+01 | NA | NA | NA |
| Sodium in Urine | DBP | MR Egger | 32 | 3.8E+00 | 3.5E+00 | 2.8E-01 | 4.6E+01 | 3.0E+01 | NA | NA | NA | -7.8E-02 | 5.8E-02 | 1.9E-01 |
| Sodium in Urine | DBP | WMed | 32 | 1.3E-01 | 4.6E-01 | 7.8E-01 | 4.6E+01 | 3.0E+01 | NA | NA | NA | NA | NA | NA |
| Sodium in Urine | DBP | WMode | 32 | 6.2E-01 | 9.0E-01 | 5.0E-01 | 4.6E+01 | 3.0E+01 | NA | NA | NA | NA | NA | NA |
| Sodium in Urine | HbA1c | IVW | 19 | -1.7E-02 | 6.5E-02 | 7.9E-01 | 4.7E+01 | 3.0E+01 | 2.4E+01 | 1.5E-01 | 2.0E+01 | NA | NA | NA |
| Sodium in Urine | HbA1c | MR Egger | 19 | -4.2E-01 | 3.5E-01 | 2.5E-01 | 4.7E+01 | 3.0E+01 | NA | NA | NA | 6.6E-03 | 5.6E-03 | 2.6E-01 |
| Sodium in Urine | HbA1c | WMed | 19 | 1.4E-02 | 8.3E-02 | 8.6E-01 | 4.7E+01 | 3.0E+01 | NA | NA | NA | NA | NA | NA |
| Sodium in Urine | HbA1c | WMode | 19 | 8.6E-02 | 1.4E-01 | 5.5E-01 | 4.7E+01 | 3.0E+01 | NA | NA | NA | NA | NA | NA |

| | | | | | | | | | | | | | |
|---|---|---|---|---|---|---|---|---|---|---|---|---|---|
| Sodium in Urine | HDL | IVW | 21 | 1.7E-01 | 9.8E-02 | 9.1E-02 | 4.7E+01 | 3.0E+01 | 4.4E+01 | 1.5E-03 | 5.2E+01 | NA | NA | NA |
| Sodium in Urine | HDL | MR Egger | 21 | 6.8E-01 | 5.1E-01 | 2.0E-01 | 4.7E+01 | 3.0E+01 | NA | NA | NA | -8.5E-03 | 8.4E-03 | 3.2E-01 |
| Sodium in Urine | HDL | WMed | 21 | 1.7E-01 | 1.1E-01 | 1.0E-01 | 4.7E+01 | 3.0E+01 | NA | NA | NA | NA | NA | NA |
| Sodium in Urine | HDL | WMode | 21 | 1.8E-01 | 1.8E-01 | 3.4E-01 | 4.7E+01 | 3.0E+01 | NA | NA | NA | NA | NA | NA |
| Sodium in Urine | LDL | IVW | 21 | -7.8E-02 | 1.3E-01 | 5.6E-01 | 4.7E+01 | 3.0E+01 | 7.1E+01 | 1.5E-07 | 7.1E+01 | NA | NA | NA |
| Sodium in Urine | LDL | MR Egger | 21 | 4.8E-01 | 7.0E-01 | 5.1E-01 | 4.7E+01 | 3.0E+01 | NA | NA | NA | -9.3E-03 | 1.2E-02 | 4.3E-01 |
| Sodium in Urine | LDL | WMed | 21 | 4.8E-02 | 1.1E-01 | 6.6E-01 | 4.7E+01 | 3.0E+01 | NA | NA | NA | NA | NA | NA |
| Sodium in Urine | LDL | WMode | 21 | 9.7E-02 | 1.5E-01 | 5.3E-01 | 4.7E+01 | 3.0E+01 | NA | NA | NA | NA | NA | NA |
| Sodium in Urine | MetS | IVW | 42 | 8.6E-01 | 2.0E-01 | 1.7E-05 | 4.4E+01 | 3.0E+01 | 3.3E+02 | 5.9E-47 | 8.7E+01 | NA | NA | NA |
| Sodium in Urine | MetS | MR Egger | 42 | -1.3E-01 | 8.0E-01 | 8.7E-01 | 4.4E+01 | 3.0E+01 | NA | NA | NA | 1.7E-02 | 1.3E-02 | 2.1E-01 |
| Sodium in Urine | MetS | WMed | 42 | 5.8E-01 | 1.4E-01 | 1.9E-05 | 4.4E+01 | 3.0E+01 | NA | NA | NA | NA | NA | NA |
| Sodium in Urine | MetS | WMode | 42 | 5.3E-01 | 2.8E-01 | 6.7E-02 | 4.4E+01 | 3.0E+01 | NA | NA | NA | NA | NA | NA |
| Sodium in Urine | SBP | IVW | 33 | -6.9E-01 | 1.5E+0<br>0 | 6.5E-01 | 4.5E+01 | 3.0E+01 | 5.6E+02 | 6.8E-97 | 9.3E+01 | NA | NA | NA |
| Sodium in Urine | SBP | MR Egger | 33 | 1.2E+01 | 6.1E+0<br>0 | 6.8E-02 | 4.5E+01 | 3.0E+01 | NA | NA | NA | -2.1E-01 | 1.0E-01 | 4.8E-02 |
| Sodium in Urine | SBP | WMed | 33 | -7.6E-01 | 8.7E-01 | 3.8E-01 | 4.5E+01 | 3.0E+01 | NA | NA | NA | NA | NA | NA |
| Sodium in Urine | SBP | WMode | 33 | -2.5E+00 | 1.9E+0<br>0 | 2.1E-01 | 4.5E+01 | 3.0E+01 | NA | NA | NA | NA | NA | NA |
| Sodium in Urine | T2D | IVW | 25 | -6.7E-01 | 4.3E-01 | 1.2E-01 | 4.3E+01 | 3.0E+01 | 3.6E+02 | 1.5E-62 | 9.3E+01 | NA | NA | NA |
| Sodium in Urine | T2D | MR Egger | 25 | 1.1E-01 | 2.9E+0<br>0 | 9.7E-01 | 4.3E+01 | 3.0E+01 | NA | NA | NA | -1.2E-02 | 4.5E-02 | 7.9E-01 |
| Sodium in Urine | T2D | WMed | 25 | -7.7E-01 | 2.0E-01 | 1.0E-04 | 4.3E+01 | 3.0E+01 | NA | NA | NA | NA | NA | NA |
| Sodium in Urine | T2D | WMode | 25 | -9.2E-01 | 3.7E-01 | 1.9E-02 | 4.3E+01 | 3.0E+01 | NA | NA | NA | NA | NA | NA |
| Sodium in Urine | Triglyceri<br>des | IVW | 21 | -8.4E-01 | 4.6E-01 | 6.9E-02 | 4.7E+01 | 3.0E+01 | 1.0E+03 | 1.1E-206 | 9.8E+01 | NA | NA | NA |

| | | | | | | | | | | | | | | |
|---|---|---|---|---|---|---|---|---|---|---|---|---|---|---|
| Sodium in Urine | Triglycerides | MR Egger | 21 | -4.2E+00 | 2.3E+00 | 9.1E-02 | 4.7E+01 | 3.0E+01 | NA | NA | NA | 5.6E-02 | 3.8E-02 | 1.6E-01 |
| Sodium in Urine | Triglycerides | WMed | 21 | -1.5E-01 | 1.2E-01 | 2.1E-01 | 4.7E+01 | 3.0E+01 | NA | NA | NA | NA | NA | NA |
| Sodium in Urine | Triglycerides | WMode | 21 | -3.1E-01 | 2.0E-01 | 1.3E-01 | 4.7E+01 | 3.0E+01 | NA | NA | NA | NA | NA | NA |
| Tea Consumption | BMI | IVW | 33 | -4.5E-04 | 1.4E-01 | 1.0E+00 | 6.7E+01 | 3.0E+01 | 1.6E+03 | 3.3E-308 | 9.8E+01 | NA | NA | NA |
| Tea Consumption | BMI | MR Egger | 33 | -4.8E-01 | 2.9E-01 | 1.1E-01 | 6.7E+01 | 3.0E+01 | NA | NA | NA | 1.1E-02 | 5.7E-03 | 7.2E-02 |
| Tea Consumption | BMI | WMed | 33 | -2.4E-01 | 3.5E-02 | 5.7E-12 | 6.7E+01 | 3.0E+01 | NA | NA | NA | NA | NA | NA |
| Tea Consumption | BMI | WMode | 33 | -2.2E-01 | 3.2E-02 | 8.0E-08 | 6.7E+01 | 3.0E+01 | NA | NA | NA | NA | NA | NA |
| Tea Consumption | DBP | IVW | 29 | 7.3E-01 | 4.9E-01 | 1.3E-01 | 7.1E+01 | 3.0E+01 | 2.0E+02 | 1.3E-27 | 8.6E+01 | NA | NA | NA |
| Tea Consumption | DBP | MR Egger | 29 | 1.0E+00 | 1.1E+00 | 3.6E-01 | 7.1E+01 | 3.0E+01 | NA | NA | NA | -6.3E-03 | 2.1E-02 | 7.7E-01 |
| Tea Consumption | DBP | WMed | 29 | 3.7E-01 | 4.4E-01 | 4.0E-01 | 7.1E+01 | 3.0E+01 | NA | NA | NA | NA | NA | NA |
| Tea Consumption | DBP | WMode | 29 | 2.0E+00 | 1.7E+00 | 2.4E-01 | 7.1E+01 | 3.0E+01 | NA | NA | NA | NA | NA | NA |
| Tea Consumption | HbA1c | IVW | 12 | -1.3E-03 | 5.4E-02 | 9.8E-01 | 1.1E+02 | 3.0E+01 | 6.9E+00 | 8.1E-01 | 0.0E+00 | NA | NA | NA |
| Tea Consumption | HbA1c | MR Egger | 12 | -3.3E-02 | 1.0E-01 | 7.5E-01 | 1.1E+02 | 3.0E+01 | NA | NA | NA | 8.8E-04 | 2.4E-03 | 7.2E-01 |
| Tea Consumption | HbA1c | WMed | 12 | -3.1E-02 | 6.8E-02 | 6.5E-01 | 1.1E+02 | 3.0E+01 | NA | NA | NA | NA | NA | NA |
| Tea Consumption | HbA1c | WMode | 12 | -3.4E-02 | 6.6E-02 | 6.1E-01 | 1.1E+02 | 3.0E+01 | NA | NA | NA | NA | NA | NA |
| Tea Consumption | HDL | IVW | 11 | -4.1E-02 | 1.1E-01 | 7.0E-01 | 1.1E+02 | 3.0E+01 | 3.3E+01 | 2.3E-04 | 6.6E+01 | NA | NA | NA |
| Tea Consumption | HDL | MR Egger | 11 | 1.3E-01 | 1.9E-01 | 5.0E-01 | 1.1E+02 | 3.0E+01 | NA | NA | NA | -5.3E-03 | 4.8E-03 | 3.0E-01 |
| Tea Consumption | HDL | WMed | 11 | -6.1E-02 | 8.0E-02 | 4.5E-01 | 1.1E+02 | 3.0E+01 | NA | NA | NA | NA | NA | NA |
| Tea Consumption | HDL | WMode | 11 | -4.3E-02 | 9.3E-02 | 6.5E-01 | 1.1E+02 | 3.0E+01 | NA | NA | NA | NA | NA | NA |
| Tea Consumption | LDL | IVW | 11 | -1.3E-02 | 6.9E-02 | 8.5E-01 | 1.1E+02 | 3.0E+01 | 1.2E+01 | 2.6E-01 | 0.0E+00 | NA | NA | NA |
| Tea Consumption | LDL | MR Egger | 11 | -2.4E-01 | 1.1E-01 | 6.1E-02 | 1.1E+02 | 3.0E+01 | NA | NA | NA | 6.9E-03 | 2.8E-03 | 3.7E-02 |

| | | | | | | | | | | | | | | |
|---|---|---|---|---|---|---|---|---|---|---|---|---|---|---|
| Tea Consumption | LDL | WMed | 11 | -9.0E-02 | 7.6E-02 | 2.3E-01 | 1.1E+02 | 3.0E+01 | NA | NA | NA | NA | NA | NA |
| Tea Consumption | LDL | WMode | 11 | -1.1E-01 | 7.7E-02 | 1.8E-01 | 1.1E+02 | 3.0E+01 | NA | NA | NA | NA | NA | NA |
| Tea Consumption | MetS | IVW | 32 | 1.8E-02 | 1.8E-01 | 9.2E-01 | 6.8E+01 | 3.0E+01 | 2.3E+02 | 7.5E-33 | 8.6E+01 | NA | NA | NA |
| Tea Consumption | MetS | MR Egger | 32 | 3.9E-01 | 4.0E-01 | 3.4E-01 | 6.8E+01 | 3.0E+01 | NA | NA | NA | -8.1E-03 | 7.8E-03 | 3.1E-01 |
| Tea Consumption | MetS | WMed | 32 | -9.2E-02 | 1.1E-01 | 4.2E-01 | 6.8E+01 | 3.0E+01 | NA | NA | NA | NA | NA | NA |
| Tea Consumption | MetS | WMode | 32 | -5.9E-02 | 1.5E-01 | 6.9E-01 | 6.8E+01 | 3.0E+01 | NA | NA | NA | NA | NA | NA |
| Tea Consumption | SBP | IVW | 29 | 1.1E+00 | 8.5E-01 | 1.8E-01 | 7.1E+01 | 3.0E+01 | 2.0E+02 | 3.2E-28 | 8.6E+01 | NA | NA | NA |
| Tea Consumption | SBP | MR Egger | 29 | 2.5E-01 | 1.9E+00 | 9.0E-01 | 7.1E+01 | 3.0E+01 | NA | NA | NA | 2.0E-02 | 3.7E-02 | 6.0E-01 |
| Tea Consumption | SBP | WMed | 29 | 1.2E+00 | 7.5E-01 | 1.1E-01 | 7.1E+01 | 3.0E+01 | NA | NA | NA | NA | NA | NA |
| Tea Consumption | SBP | WMode | 29 | 2.3E+00 | 1.5E+00 | 1.3E-01 | 7.1E+01 | 3.0E+01 | NA | NA | NA | NA | NA | NA |
| Tea Consumption | T2D | IVW | 23 | 1.6E-01 | 3.5E-01 | 6.4E-01 | 5.4E+01 | 3.0E+01 | 2.4E+02 | 2.9E-39 | 9.0E+01 | NA | NA | NA |
| Tea Consumption | T2D | MR Egger | 23 | -1.0E+00 | 9.5E-01 | 3.1E-01 | 5.4E+01 | 3.0E+01 | NA | NA | NA | 2.1E-02 | 1.6E-02 | 2.1E-01 |
| Tea Consumption | T2D | WMed | 23 | -3.6E-01 | 1.8E-01 | 4.3E-02 | 5.4E+01 | 3.0E+01 | NA | NA | NA | NA | NA | NA |
| Tea Consumption | T2D | WMode | 23 | -3.3E-01 | 1.8E-01 | 7.8E-02 | 5.4E+01 | 3.0E+01 | NA | NA | NA | NA | NA | NA |
| Tea Consumption | Triglycerides | IVW | 11 | 8.6E-02 | 9.9E-02 | 3.9E-01 | 1.1E+02 | 3.0E+01 | 3.1E+01 | 5.4E-04 | 6.3E+01 | NA | NA | NA |
| Tea Consumption | Triglycerides | MR Egger | 11 | -8.6E-02 | 1.7E-01 | 6.3E-01 | 1.1E+02 | 3.0E+01 | NA | NA | NA | 5.3E-03 | 4.4E-03 | 2.6E-01 |
| Tea Consumption | Triglycerides | WMed | 11 | 1.2E-01 | 7.5E-02 | 1.2E-01 | 1.1E+02 | 3.0E+01 | NA | NA | NA | NA | NA | NA |
| Tea Consumption | Triglycerides | WMode | 11 | 1.3E-01 | 9.0E-02 | 1.9E-01 | 1.1E+02 | 3.0E+01 | NA | NA | NA | NA | NA | NA |
| Total Testosterone | BMI | IVW | 172 | -4.7E-02 | 1.4E-02 | 9.9E-04 | 8.7E+01 | 2.4E+01 | 1.0E+03 | 3.3E-125 | 8.4E+01 | NA | NA | NA |

| | | | | | | | | | | | | | | |
|---|---|---|---|---|---|---|---|---|---|---|---|---|---|---|
| Total Testosterone | BMI | MR Egger | 172 | -6.3E-02 | 2.8E-02 | 2.6E-02 | 8.7E+01 | 2.4E+01 | NA | NA | NA | 6.0E-04 | 9.1E-04 | 5.1E-01 |
| Total Testosterone | BMI | WMed | 172 | -3.4E-02 | 1.0E-02 | 1.2E-03 | 8.7E+01 | 2.4E+01 | NA | NA | NA | NA | NA | NA |
| Total Testosterone | BMI | WMode | 172 | -4.2E-02 | 1.1E-02 | 2.5E-04 | 8.7E+01 | 2.4E+01 | NA | NA | NA | NA | NA | NA |
| Total Testosterone | DBP | IVW | 157 | -1.0E-01 | 2.1E-01 | 6.2E-01 | 8.9E+01 | 2.4E+01 | 2.4E+03 | 0.0E+00 | 9.3E+01 | NA | NA | NA |
| Total Testosterone | DBP | MR Egger | 157 | -4.9E-01 | 4.1E-01 | 2.4E-01 | 8.9E+01 | 2.4E+01 | NA | NA | NA | 1.4E-02 | 1.3E-02 | 2.8E-01 |
| Total Testosterone | DBP | WMed | 157 | -1.4E-01 | 1.2E-01 | 2.6E-01 | 8.9E+01 | 2.4E+01 | NA | NA | NA | NA | NA | NA |
| Total Testosterone | DBP | WMode | 157 | -8.5E-02 | 1.3E-01 | 5.1E-01 | 8.9E+01 | 2.4E+01 | NA | NA | NA | NA | NA | NA |
| Total Testosterone | HbA1c | IVW | 61 | -2.8E-02 | 2.2E-02 | 2.1E-01 | 7.3E+01 | 2.8E+01 | 7.6E+01 | 7.7E-02 | 2.0E+01 | NA | NA | NA |
| Total Testosterone | HbA1c | MR Egger | 61 | -8.4E-02 | 6.0E-02 | 1.7E-01 | 7.3E+01 | 2.8E+01 | NA | NA | NA | 1.7E-03 | 1.7E-03 | 3.2E-01 |
| Total Testosterone | HbA1c | WMed | 61 | -5.6E-02 | 3.0E-02 | 6.1E-02 | 7.3E+01 | 2.8E+01 | NA | NA | NA | NA | NA | NA |
| Total Testosterone | HbA1c | WMode | 61 | -5.8E-02 | 4.5E-02 | 2.0E-01 | 7.3E+01 | 2.8E+01 | NA | NA | NA | NA | NA | NA |
| Total Testosterone | HDL | IVW | 58 | -1.7E-02 | 4.0E-02 | 6.8E-01 | 7.3E+01 | 2.8E+01 | 1.4E+02 | 5.3E-09 | 5.9E+01 | NA | NA | NA |
| Total Testosterone | HDL | MR Egger | 58 | -9.4E-03 | 1.1E-01 | 9.3E-01 | 7.3E+01 | 2.8E+01 | NA | NA | NA | -2.2E-04 | 3.1E-03 | 9.4E-01 |
| Total Testosterone | HDL | WMed | 58 | 1.2E-02 | 4.1E-02 | 7.7E-01 | 7.3E+01 | 2.8E+01 | NA | NA | NA | NA | NA | NA |
| Total Testosterone | HDL | WMode | 58 | 2.5E-02 | 6.4E-02 | 7.0E-01 | 7.3E+01 | 2.8E+01 | NA | NA | NA | NA | NA | NA |

| | | | | | | | | | | | | | | |
|---|---|---|---|---|---|---|---|---|---|---|---|---|---|---|
| Total Testosterone | LDL | IVW | 58 | 1.1E-01 | 4.5E-02 | 1.9E-02 | 7.3E+01 | 2.8E+01 | 1.6E+02 | 5.2E-11 | 6.3E+01 | NA | NA | NA |
| Total Testosterone | LDL | MR Egger | 58 | 2.2E-01 | 1.3E-01 | 9.2E-02 | 7.3E+01 | 2.8E+01 | NA | NA | NA | -3.3E-03 | 3.6E-03 | 3.5E-01 |
| Total Testosterone | LDL | WMed | 58 | 1.2E-01 | 4.5E-02 | 5.9E-03 | 7.3E+01 | 2.8E+01 | NA | NA | NA | NA | NA | NA |
| Total Testosterone | LDL | WMode | 58 | 1.5E-01 | 8.9E-02 | 1.0E-01 | 7.3E+01 | 2.8E+01 | NA | NA | NA | NA | NA | NA |
| Total Testosterone | MetS | IVW | 185 | -7.0E-03 | 3.9E-02 | 8.6E-01 | 8.5E+01 | 2.4E+01 | 7.9E+02 | 1.0E-75 | 7.6E+01 | NA | NA | NA |
| Total Testosterone | MetS | MR Egger | 185 | 1.1E-01 | 7.6E-02 | 1.4E-01 | 8.5E+01 | 2.4E+01 | NA | NA | NA | -4.4E-03 | 2.4E-03 | 6.8E-02 |
| Total Testosterone | MetS | WMed | 185 | 7.2E-02 | 3.8E-02 | 5.6E-02 | 8.5E+01 | 2.4E+01 | NA | NA | NA | NA | NA | NA |
| Total Testosterone | MetS | WMode | 185 | 7.3E-02 | 3.9E-02 | 6.1E-02 | 8.5E+01 | 2.4E+01 | NA | NA | NA | NA | NA | NA |
| Total Testosterone | SBP | IVW | 157 | -5.3E-01 | 3.4E-01 | 1.1E-01 | 8.9E+01 | 2.4E+01 | 2.1E+03 | 0.0E+00 | 9.2E+01 | NA | NA | NA |
| Total Testosterone | SBP | MR Egger | 157 | -1.5E+00 | 6.7E-01 | 2.7E-02 | 8.9E+01 | 2.4E+01 | NA | NA | NA | 3.6E-02 | 2.1E-02 | 9.9E-02 |
| Total Testosterone | SBP | WMed | 157 | -4.3E-01 | 2.0E-01 | 3.3E-02 | 8.9E+01 | 2.4E+01 | NA | NA | NA | NA | NA | NA |
| Total Testosterone | SBP | WMode | 157 | -6.7E-01 | 2.1E-01 | 2.1E-03 | 8.9E+01 | 2.4E+01 | NA | NA | NA | NA | NA | NA |
| Total Testosterone | T2D | IVW | 117 | 7.9E-02 | 6.3E-02 | 2.1E-01 | 8.3E+01 | 2.4E+01 | 5.7E+02 | 4.4E-61 | 7.9E+01 | NA | NA | NA |
| Total Testosterone | T2D | MR Egger | 117 | -3.9E-02 | 1.4E-01 | 7.9E-01 | 8.3E+01 | 2.4E+01 | NA | NA | NA | 3.9E-03 | 4.2E-03 | 3.6E-01 |
| Total Testosterone | T2D | WMed | 117 | 2.5E-02 | 5.4E-02 | 6.5E-01 | 8.3E+01 | 2.4E+01 | NA | NA | NA | NA | NA | NA |

| | | | | | | | | | | | | | | |
|---|---|---|---|---|---|---|---|---|---|---|---|---|---|---|
| Total Testosterone | T2D | WMode | 117 | -3.4E-05 | 8.1E-02 | 1.0E+00 | 8.3E+01 | 2.4E+01 | NA | NA | NA | NA | NA | NA |
| Total Testosterone | Triglycerides | IVW | 58 | 2.9E-01 | 1.1E-01 | 9.8E-03 | 7.3E+01 | 2.8E+01 | 1.2E+03 | 1.9E-212 | 9.5E+01 | NA | NA | NA |
| Total Testosterone | Triglycerides | MR Egger | 58 | 5.1E-01 | 3.1E-01 | 1.1E-01 | 7.3E+01 | 2.8E+01 | NA | NA | NA | -6.5E-03 | 8.9E-03 | 4.7E-01 |
| Total Testosterone | Triglycerides | WMed | 58 | 5.0E-02 | 4.1E-02 | 2.3E-01 | 7.3E+01 | 2.8E+01 | NA | NA | NA | NA | NA | NA |
| Total Testosterone | Triglycerides | WMode | 58 | 4.8E-02 | 6.2E-02 | 4.4E-01 | 7.3E+01 | 2.8E+01 | NA | NA | NA | NA | NA | NA |
| Urate | BMI | IVW | 229 | -8.9E-02 | 3.0E-02 | 2.6E-03 | 1.3E+02 | 3.0E+01 | 4.8E+03 | 0.0E+00 | 9.5E+01 | NA | NA | NA |
| Urate | BMI | MR Egger | 229 | 8.3E-02 | 4.6E-02 | 6.9E-02 | 1.3E+02 | 3.0E+01 | NA | NA | NA | -5.3E-03 | 1.1E-03 | 2.8E-06 |
| Urate | BMI | WMed | 229 | 1.3E-02 | 1.3E-02 | 3.3E-01 | 1.3E+02 | 3.0E+01 | NA | NA | NA | NA | NA | NA |
| Urate | BMI | WMode | 229 | 1.4E-02 | 1.1E-02 | 2.1E-01 | 1.3E+02 | 3.0E+01 | NA | NA | NA | NA | NA | NA |
| Urate | DBP | IVW | 205 | -1.2E+00 | 2.5E-01 | 2.2E-06 | 1.2E+02 | 3.0E+01 | 3.3E+03 | 0.0E+00 | 9.4E+01 | NA | NA | NA |
| Urate | DBP | MR Egger | 205 | -4.3E-01 | 4.1E-01 | 2.9E-01 | 1.2E+02 | 3.0E+01 | NA | NA | NA | -2.2E-02 | 9.6E-03 | 2.6E-02 |
| Urate | DBP | WMed | 205 | -4.8E-01 | 1.3E-01 | 2.8E-04 | 1.2E+02 | 3.0E+01 | NA | NA | NA | NA | NA | NA |
| Urate | DBP | WMode | 205 | -4.2E-01 | 1.1E-01 | 1.4E-04 | 1.2E+02 | 3.0E+01 | NA | NA | NA | NA | NA | NA |
| Urate | HbA1c | IVW | 99 | 4.5E-03 | 2.3E-02 | 8.5E-01 | 1.1E+02 | 3.0E+01 | 1.4E+02 | 1.6E-03 | 2.9E+01 | NA | NA | NA |
| Urate | HbA1c | MR Egger | 99 | 8.8E-02 | 4.7E-02 | 6.2E-02 | 1.1E+02 | 3.0E+01 | NA | NA | NA | -2.0E-03 | 9.8E-04 | 4.3E-02 |
| Urate | HbA1c | WMed | 99 | 7.1E-02 | 3.2E-02 | 2.9E-02 | 1.1E+02 | 3.0E+01 | NA | NA | NA | NA | NA | NA |
| Urate | HbA1c | WMode | 99 | 5.7E-02 | 3.2E-02 | 7.9E-02 | 1.1E+02 | 3.0E+01 | NA | NA | NA | NA | NA | NA |
| Urate | HDL | IVW | 112 | 7.6E-02 | 5.8E-02 | 1.9E-01 | 1.1E+02 | 3.0E+01 | 8.0E+02 | 9.5E-105 | 8.6E+01 | NA | NA | NA |
| Urate | HDL | MR Egger | 112 | -2.9E-02 | 1.2E-01 | 8.0E-01 | 1.1E+02 | 3.0E+01 | NA | NA | NA | 2.6E-03 | 2.5E-03 | 3.0E-01 |
| Urate | HDL | WMed | 112 | 3.7E-02 | 4.0E-02 | 3.5E-01 | 1.1E+02 | 3.0E+01 | NA | NA | NA | NA | NA | NA |
| Urate | HDL | WMode | 112 | 7.5E-02 | 4.2E-02 | 7.9E-02 | 1.1E+02 | 3.0E+01 | NA | NA | NA | NA | NA | NA |

| | | | | | | | | | | | | | | |
|---|---|---|---|---|---|---|---|---|---|---|---|---|---|---|
| Urate | LDL | IVW | 112 | -1.7E-02 | 6.5E-02 | 7.9E-01 | 1.1E+02 | 3.0E+01 | 8.5E+02 | 2.4E-114 | 8.7E+01 | NA | NA | NA |
| Urate | LDL | MR Egger | 112 | -6.9E-02 | 1.3E-01 | 5.9E-01 | 1.1E+02 | 3.0E+01 | NA | NA | NA | 1.3E-03 | 2.8E-03 | 6.4E-01 |
| Urate | LDL | WMed | 112 | -1.7E-02 | 4.9E-02 | 7.4E-01 | 1.1E+02 | 3.0E+01 | NA | NA | NA | NA | NA | NA |
| Urate | LDL | WMode | 112 | -2.7E-02 | 6.0E-02 | 6.5E-01 | 1.1E+02 | 3.0E+01 | NA | NA | NA | NA | NA | NA |
| Urate | MetS | IVW | 262 | 3.1E-01 | 4.9E-02 | 2.7E-10 | 1.1E+02 | 3.0E+01 | 1.4E+03 | 9.8E-159 | 8.0E+01 | NA | NA | NA |
| Urate | MetS | MR Egger | 262 | -1.4E-02 | 7.9E-02 | 8.6E-01 | 1.1E+02 | 3.0E+01 | NA | NA | NA | 9.2E-03 | 1.8E-03 | 7.1E-07 |
| Urate | MetS | WMed | 262 | 8.5E-02 | 4.4E-02 | 5.3E-02 | 1.1E+02 | 3.0E+01 | NA | NA | NA | NA | NA | NA |
| Urate | MetS | WMode | 262 | 5.3E-02 | 3.9E-02 | 1.8E-01 | 1.1E+02 | 3.0E+01 | NA | NA | NA | NA | NA | NA |
| Urate | SBP | IVW | 205 | -1.8E+00 | 3.8E-01 | 2.3E-06 | 1.2E+02 | 3.0E+01 | 2.7E+03 | 0.0E+00 | 9.2E+01 | NA | NA | NA |
| Urate | SBP | MR Egger | 205 | -8.5E-01 | 6.4E-01 | 1.8E-01 | 1.2E+02 | 3.0E+01 | NA | NA | NA | -2.8E-02 | 1.5E-02 | 6.3E-02 |
| Urate | SBP | WMed | 205 | -6.3E-01 | 2.8E-01 | 2.4E-02 | 1.2E+02 | 3.0E+01 | NA | NA | NA | NA | NA | NA |
| Urate | SBP | WMode | 205 | -7.3E-01 | 2.1E-01 | 4.5E-04 | 1.2E+02 | 3.0E+01 | NA | NA | NA | NA | NA | NA |
| Urate | T2D | IVW | 159 | -1.8E-01 | 8.0E-02 | 2.8E-02 | 1.2E+02 | 3.0E+01 | 1.0E+03 | 5.6E-127 | 8.4E+01 | NA | NA | NA |
| Urate | T2D | MR Egger | 159 | 2.2E-01 | 1.5E-01 | 1.4E-01 | 1.2E+02 | 3.0E+01 | NA | NA | NA | -1.0E-02 | 3.2E-03 | 1.8E-03 |
| Urate | T2D | WMed | 159 | -8.8E-02 | 5.7E-02 | 1.2E-01 | 1.2E+02 | 3.0E+01 | NA | NA | NA | NA | NA | NA |
| Urate | T2D | WMode | 159 | -1.2E-01 | 5.0E-02 | 2.1E-02 | 1.2E+02 | 3.0E+01 | NA | NA | NA | NA | NA | NA |
| Urate | Triglycerides | IVW | 112 | -3.0E-01 | 7.4E-02 | 4.8E-05 | 1.1E+02 | 3.0E+01 | 1.3E+03 | 1.2E-209 | 9.2E+01 | NA | NA | NA |
| Urate | Triglycerides | MR Egger | 112 | -5.0E-01 | 1.5E-01 | 7.7E-04 | 1.1E+02 | 3.0E+01 | NA | NA | NA | 5.1E-03 | 3.2E-03 | 1.1E-01 |
| Urate | Triglycerides | WMed | 112 | -7.3E-02 | 4.5E-02 | 1.0E-01 | 1.1E+02 | 3.0E+01 | NA | NA | NA | NA | NA | NA |
| Urate | Triglycerides | WMode | 112 | 5.7E-04 | 4.8E-02 | 9.9E-01 | 1.1E+02 | 3.0E+01 | NA | NA | NA | NA | NA | NA |
| Urea | BMI | IVW | 157 | 3.8E-03 | 3.4E-02 | 9.1E-01 | 7.8E+01 | 3.0E+01 | 2.5E+03 | 0.0E+00 | 9.4E+01 | NA | NA | NA |
| Urea | BMI | MR Egger | 157 | 1.4E-01 | 8.6E-02 | 9.6E-02 | 7.8E+01 | 3.0E+01 | NA | NA | NA | -3.3E-03 | 1.9E-03 | 7.8E-02 |

| | | | | | | | | | | | | | | |
|---|---|---|---|---|---|---|---|---|---|---|---|---|---|---|
| Urea | BMI | WMed | 157 | 4.1E-03 | 1.7E-02 | 8.1E-01 | 7.8E+01 | 3.0E+01 | NA | NA | NA | NA | NA | NA |
| Urea | BMI | WMode | 157 | -1.2E-02 | 2.3E-02 | 6.0E-01 | 7.8E+01 | 3.0E+01 | NA | NA | NA | NA | NA | NA |
| Urea | DBP | IVW | 133 | -7.3E-01 | 3.2E-01 | 2.1E-02 | 7.5E+01 | 3.0E+01 | 1.9E+03 | 1.611488915 21861e-313 | 9.3E+01 | NA | NA | NA |
| Urea | DBP | MR Egger | 133 | -1.0E+00 | 7.9E-01 | 2.1E-01 | 7.5E+01 | 3.0E+01 | NA | NA | NA | 6.3E-03 | 1.7E-02 | 7.1E-01 |
| Urea | DBP | WMed | 133 | -4.3E-01 | 1.8E-01 | 1.9E-02 | 7.5E+01 | 3.0E+01 | NA | NA | NA | NA | NA | NA |
| Urea | DBP | WMode | 133 | -3.3E-01 | 4.2E-01 | 4.4E-01 | 7.5E+01 | 3.0E+01 | NA | NA | NA | NA | NA | NA |
| Urea | HbA1c | IVW | 66 | -4.1E-02 | 3.0E-02 | 1.8E-01 | 6.8E+01 | 3.0E+01 | 8.7E+01 | 3.4E-02 | 2.3E+01 | NA | NA | NA |
| Urea | HbA1c | MR Egger | 66 | 9.8E-02 | 9.3E-02 | 3.0E-01 | 6.8E+01 | 3.0E+01 | NA | NA | NA | -2.8E-03 | 1.7E-03 | 1.2E-01 |
| Urea | HbA1c | WMed | 66 | -1.6E-02 | 3.9E-02 | 6.9E-01 | 6.8E+01 | 3.0E+01 | NA | NA | NA | NA | NA | NA |
| Urea | HbA1c | WMode | 66 | -2.4E-02 | 6.7E-02 | 7.2E-01 | 6.8E+01 | 3.0E+01 | NA | NA | NA | NA | NA | NA |
| Urea | HDL | IVW | 71 | 1.5E-02 | 6.9E-02 | 8.3E-01 | 7.6E+01 | 3.0E+01 | 3.8E+02 | 1.5E-44 | 8.2E+01 | NA | NA | NA |
| Urea | HDL | MR Egger | 71 | 2.5E-03 | 2.0E-01 | 9.9E-01 | 7.6E+01 | 3.0E+01 | NA | NA | NA | 2.7E-04 | 4.0E-03 | 9.5E-01 |
| Urea | HDL | WMed | 71 | 1.5E-02 | 5.5E-02 | 7.8E-01 | 7.6E+01 | 3.0E+01 | NA | NA | NA | NA | NA | NA |
| Urea | HDL | WMode | 71 | 2.7E-02 | 8.6E-02 | 7.5E-01 | 7.6E+01 | 3.0E+01 | NA | NA | NA | NA | NA | NA |
| Urea | LDL | IVW | 71 | 1.1E-01 | 5.7E-02 | 5.1E-02 | 7.6E+01 | 3.0E+01 | 2.2E+02 | 7.5E-18 | 6.8E+01 | NA | NA | NA |
| Urea | LDL | MR Egger | 71 | 3.7E-02 | 1.7E-01 | 8.2E-01 | 7.6E+01 | 3.0E+01 | NA | NA | NA | 1.6E-03 | 3.3E-03 | 6.3E-01 |
| Urea | LDL | WMed | 71 | 3.3E-02 | 5.7E-02 | 5.6E-01 | 7.6E+01 | 3.0E+01 | NA | NA | NA | NA | NA | NA |
| Urea | LDL | WMode | 71 | -2.1E-02 | 9.4E-02 | 8.2E-01 | 7.6E+01 | 3.0E+01 | NA | NA | NA | NA | NA | NA |
| Urea | MetS | IVW | 176 | 1.3E-01 | 5.5E-02 | 1.5E-02 | 7.6E+01 | 3.0E+01 | 7.5E+02 | 9.3E-72 | 7.6E+01 | NA | NA | NA |
| Urea | MetS | MR Egger | 176 | -2.6E-02 | 1.4E-01 | 8.5E-01 | 7.6E+01 | 3.0E+01 | NA | NA | NA | 3.7E-03 | 3.1E-03 | 2.3E-01 |
| Urea | MetS | WMed | 176 | 1.3E-01 | 4.6E-02 | 4.8E-03 | 7.6E+01 | 3.0E+01 | NA | NA | NA | NA | NA | NA |
| Urea | MetS | WMode | 176 | 1.2E-01 | 7.4E-02 | 1.1E-01 | 7.6E+01 | 3.0E+01 | NA | NA | NA | NA | NA | NA |
| Urea | SBP | IVW | 134 | -1.0E+00 | 5.8E-01 | 7.9E-02 | 7.5E+01 | 3.0E+01 | 2.2E+03 | 0.0E+00 | 9.4E+01 | NA | NA | NA |

| | | | | | | | | | | | | | |
|---|---|---|---|---|---|---|---|---|---|---|---|---|---|
| Urea | SBP | MR Egger | 134 | -1.4E+00 | 1.5E+00 | 3.4E-01 | 7.5E+01 | 3.0E+01 | NA | NA | NA | 8.9E-03 | 3.1E-02 | 7.7E-01 |
| Urea | SBP | WMed | 134 | -7.6E-01 | 3.3E-01 | 2.4E-02 | 7.5E+01 | 3.0E+01 | NA | NA | NA | NA | NA | NA |
| Urea | SBP | WMode | 134 | -2.7E-01 | 6.2E-01 | 6.7E-01 | 7.5E+01 | 3.0E+01 | NA | NA | NA | NA | NA | NA |
| Urea | T2D | IVW | 113 | 1.9E-02 | 9.1E-02 | 8.3E-01 | 7.8E+01 | 3.0E+01 | 6.0E+02 | 8.2E-68 | 8.1E+01 | NA | NA | NA |
| Urea | T2D | MR Egger | 113 | 5.3E-01 | 2.5E-01 | 3.7E-02 | 7.8E+01 | 3.0E+01 | NA | NA | NA | -1.1E-02 | 5.2E-03 | 3.1E-02 |
| Urea | T2D | WMed | 113 | 8.6E-02 | 6.8E-02 | 2.1E-01 | 7.8E+01 | 3.0E+01 | NA | NA | NA | NA | NA | NA |
| Urea | T2D | WMode | 113 | 1.2E-01 | 1.1E-01 | 2.6E-01 | 7.8E+01 | 3.0E+01 | NA | NA | NA | NA | NA | NA |
| Urea | Triglycerides | IVW | 71 | -2.3E-01 | 1.2E-01 | 5.7E-02 | 7.6E+01 | 3.0E+01 | 1.3E+03 | 1.6E-218 | 9.4E+01 | NA | NA | NA |
| Urea | Triglycerides | MR Egger | 71 | -4.5E-01 | 3.6E-01 | 2.1E-01 | 7.6E+01 | 3.0E+01 | NA | NA | NA | 4.6E-03 | 7.0E-03 | 5.1E-01 |
| Urea | Triglycerides | WMed | 71 | -5.0E-03 | 5.5E-02 | 9.3E-01 | 7.6E+01 | 3.0E+01 | NA | NA | NA | NA | NA | NA |
| Urea | Triglycerides | WMode | 71 | -7.9E-02 | 1.0E-01 | 4.5E-01 | 7.6E+01 | 3.0E+01 | NA | NA | NA | NA | NA | NA |
| Urinary Albumin Excretion | BMI | IVW | 31 | 1.4E-01 | 9.0E-02 | 1.2E-01 | 5.4E+01 | 3.0E+01 | 2.9E+02 | 1.2E-43 | 8.8E+01 | NA | NA | NA |
| Urinary Albumin Excretion | BMI | MR Egger | 31 | -2.3E-01 | 2.1E-01 | 3.0E-01 | 5.4E+01 | 3.0E+01 | NA | NA | NA | 5.8E-03 | 3.1E-03 | 7.1E-02 |
| Urinary Albumin Excretion | BMI | WMed | 31 | -1.4E-02 | 5.3E-02 | 7.9E-01 | 5.4E+01 | 3.0E+01 | NA | NA | NA | NA | NA | NA |
| Urinary Albumin Excretion | BMI | WMode | 31 | 1.1E-02 | 5.7E-02 | 8.5E-01 | 5.4E+01 | 3.0E+01 | NA | NA | NA | NA | NA | NA |
| Urinary Albumin Excretion | DBP | IVW | 27 | -2.2E+00 | 1.0E+00 | 2.9E-02 | 5.3E+01 | 3.0E+01 | 3.5E+02 | 7.1E-58 | 9.0E+01 | NA | NA | NA |
| Urinary Albumin Excretion | DBP | MR Egger | 27 | 4.4E+00 | 2.4E+00 | 7.7E-02 | 5.3E+01 | 3.0E+01 | NA | NA | NA | -1.0E-01 | 3.4E-02 | 6.2E-03 |

| | | | | | | | | | | | | | | |
|---|---|---|---|---|---|---|---|---|---|---|---|---|---|---|
| Urinary Albumin Excretion | DBP | WMed | 27 | -1.2E+00 | 6.0E-01 | 3.7E-02 | 5.3E+01 | 3.0E+01 | NA | NA | NA | NA | NA | NA |
| Urinary Albumin Excretion | DBP | WMode | 27 | -1.1E+00 | 6.5E-01 | 9.8E-02 | 5.3E+01 | 3.0E+01 | NA | NA | NA | NA | NA | NA |
| Urinary Albumin Excretion | HbA1c | IVW | 15 | -6.6E-02 | 8.2E-02 | 4.2E-01 | 6.5E+01 | 3.0E+01 | 1.9E+01 | 1.7E-01 | 2.0E+01 | NA | NA | NA |
| Urinary Albumin Excretion | HbA1c | MR Egger | 15 | 1.2E-01 | 2.1E-01 | 5.7E-01 | 6.5E+01 | 3.0E+01 | NA | NA | NA | -3.1E-03 | 3.1E-03 | 3.4E-01 |
| Urinary Albumin Excretion | HbA1c | WMed | 15 | -4.8E-02 | 1.0E-01 | 6.3E-01 | 6.5E+01 | 3.0E+01 | NA | NA | NA | NA | NA | NA |
| Urinary Albumin Excretion | HbA1c | WMode | 15 | -2.3E-02 | 1.1E-01 | 8.4E-01 | 6.5E+01 | 3.0E+01 | NA | NA | NA | NA | NA | NA |
| Urinary Albumin Excretion | HDL | IVW | 18 | -3.9E-02 | 2.0E-01 | 8.4E-01 | 6.0E+01 | 3.0E+01 | 9.3E+01 | 1.6E-12 | 8.2E+01 | NA | NA | NA |
| Urinary Albumin Excretion | HDL | MR Egger | 18 | -3.3E-02 | 5.6E-01 | 9.5E-01 | 6.0E+01 | 3.0E+01 | NA | NA | NA | -1.0E-04 | 8.0E-03 | 9.9E-01 |
| Urinary Albumin Excretion | HDL | WMed | 18 | 3.3E-02 | 1.5E-01 | 8.2E-01 | 6.0E+01 | 3.0E+01 | NA | NA | NA | NA | NA | NA |
| Urinary Albumin Excretion | HDL | WMode | 18 | -2.9E-02 | 1.7E-01 | 8.7E-01 | 6.0E+01 | 3.0E+01 | NA | NA | NA | NA | NA | NA |
| Urinary Albumin Excretion | LDL | IVW | 18 | -7.9E-02 | 1.0E-01 | 4.4E-01 | 6.0E+01 | 3.0E+01 | 2.1E+01 | 2.4E-01 | 1.8E+01 | NA | NA | NA |
| Urinary Albumin Excretion | LDL | MR Egger | 18 | 4.7E-03 | 2.8E-01 | 9.9E-01 | 6.0E+01 | 3.0E+01 | NA | NA | NA | -1.3E-03 | 4.1E-03 | 7.6E-01 |
| Urinary Albumin Excretion | LDL | WMed | 18 | -8.2E-03 | 1.3E-01 | 9.5E-01 | 6.0E+01 | 3.0E+01 | NA | NA | NA | NA | NA | NA |
| Urinary Albumin Excretion | LDL | WMode | 18 | -1.6E-01 | 1.8E-01 | 3.9E-01 | 6.0E+01 | 3.0E+01 | NA | NA | NA | NA | NA | NA |
| Urinary Albumin Excretion | MetS | IVW | 31 | 1.2E-01 | 2.4E-01 | 6.1E-01 | 5.4E+01 | 3.0E+01 | 1.8E+02 | 1.1E-23 | 8.3E+01 | NA | NA | NA |

| | | | | | | | | | | | | | | |
|---|---|---|---|---|---|---|---|---|---|---|---|---|---|---|
| Urinary Albumin Excretion | MetS | MR Egger | 31 | -1.8E-02 | 5.9E-01 | 9.8E-01 | 5.4E+01 | 3.0E+01 | NA | NA | NA | 2.2E-03 | 8.6E-03 | 8.0E-01 |
| Urinary Albumin Excretion | MetS | WMed | 31 | -1.4E-01 | 1.7E-01 | 4.2E-01 | 5.4E+01 | 3.0E+01 | NA | NA | NA | NA | NA | NA |
| Urinary Albumin Excretion | MetS | WMode | 31 | -4.0E-01 | 2.9E-01 | 1.8E-01 | 5.4E+01 | 3.0E+01 | NA | NA | NA | NA | NA | NA |
| Urinary Albumin Excretion | SBP | IVW | 26 | -4.8E+00 | 1.7E+00 | 5.4E-03 | 5.4E+01 | 3.0E+01 | 3.1E+02 | 9.4E-51 | 8.8E+01 | NA | NA | NA |
| Urinary Albumin Excretion | SBP | MR Egger | 26 | 7.8E+00 | 4.0E+00 | 6.1E-02 | 5.4E+01 | 3.0E+01 | NA | NA | NA | -1.9E-01 | 5.6E-02 | 2.4E-03 |
| Urinary Albumin Excretion | SBP | WMed | 26 | -3.1E+00 | 1.1E+00 | 4.4E-03 | 5.4E+01 | 3.0E+01 | NA | NA | NA | NA | NA | NA |
| Urinary Albumin Excretion | SBP | WMode | 26 | -1.6E+00 | 1.2E+00 | 1.8E-01 | 5.4E+01 | 3.0E+01 | NA | NA | NA | NA | NA | NA |
| Urinary Albumin Excretion | T2D | IVW | 23 | -3.7E-01 | 2.7E-01 | 1.6E-01 | 5.3E+01 | 3.0E+01 | 8.5E+01 | 2.6E-09 | 7.1E+01 | NA | NA | NA |
| Urinary Albumin Excretion | T2D | MR Egger | 23 | 6.7E-01 | 6.7E-01 | 3.3E-01 | 5.3E+01 | 3.0E+01 | NA | NA | NA | -1.6E-02 | 9.5E-03 | 1.1E-01 |
| Urinary Albumin Excretion | T2D | WMed | 23 | -3.6E-02 | 2.2E-01 | 8.7E-01 | 5.3E+01 | 3.0E+01 | NA | NA | NA | NA | NA | NA |
| Urinary Albumin Excretion | T2D | WMode | 23 | 8.0E-02 | 2.5E-01 | 7.5E-01 | 5.3E+01 | 3.0E+01 | NA | NA | NA | NA | NA | NA |
| Urinary Albumin Excretion | Triglycerides | IVW | 18 | -5.3E-02 | 1.0E-01 | 6.1E-01 | 6.0E+01 | 3.0E+01 | 2.6E+01 | 8.3E-02 | 3.3E+01 | NA | NA | NA |
| Urinary Albumin Excretion | Triglycerides | MR Egger | 18 | -7.3E-02 | 2.8E-01 | 8.0E-01 | 6.0E+01 | 3.0E+01 | NA | NA | NA | 3.1E-04 | 4.0E-03 | 9.4E-01 |
| Urinary Albumin Excretion | Triglycerides | WMed | 18 | 0.0E+00 | 1.2E-01 | 1.0E+00 | 6.0E+01 | 3.0E+01 | NA | NA | NA | NA | NA | NA |
| Urinary Albumin Excretion | Triglycerides | WMode | 18 | -4.5E-02 | 1.7E-01 | 8.0E-01 | 6.0E+01 | 3.0E+01 | NA | NA | NA | NA | NA | NA |

| | | | | | | | | | | | | | | |
|---|---|---|---|---|---|---|---|---|---|---|---|---|---|---|
| Water Consumption | BMI | IVW | 41 | -7.8E-02 | 8.2E-02 | 3.4E-01 | 4.2E+01 | 3.0E+01 | 2.0E+03 | 0.0E+00 | 9.8E+01 | NA | NA | NA |
| Water Consumption | BMI | MR Egger | 41 | 4.6E-01 | 4.1E-01 | 2.8E-01 | 4.2E+01 | 3.0E+01 | NA | NA | NA | -1.6E-02 | 1.2E-02 | 1.9E-01 |
| Water Consumption | BMI | WMed | 41 | -2.6E-02 | 2.7E-02 | 3.4E-01 | 4.2E+01 | 3.0E+01 | NA | NA | NA | NA | NA | NA |
| Water Consumption | BMI | WMode | 41 | -4.7E-02 | 6.6E-02 | 4.8E-01 | 4.2E+01 | 3.0E+01 | NA | NA | NA | NA | NA | NA |
| Water Consumption | DBP | IVW | 38 | 1.6E-01 | 3.0E-01 | 6.0E-01 | 4.2E+01 | 3.0E+01 | 3.0E+02 | 3.2E-42 | 8.7E+01 | NA | NA | NA |
| Water Consumption | DBP | MR Egger | 38 | -2.3E-01 | 1.6E+00 | 8.8E-01 | 4.2E+01 | 3.0E+01 | NA | NA | NA | 1.2E-02 | 4.6E-02 | 8.0E-01 |
| Water Consumption | DBP | WMed | 38 | 5.2E-01 | 1.8E-01 | 4.9E-03 | 4.2E+01 | 3.0E+01 | NA | NA | NA | NA | NA | NA |
| Water Consumption | DBP | WMode | 38 | 5.1E-01 | 2.7E-01 | 6.6E-02 | 4.2E+01 | 3.0E+01 | NA | NA | NA | NA | NA | NA |
| Water Consumption | HbA1c | IVW | 21 | 2.3E-02 | 3.6E-02 | 5.3E-01 | 4.7E+01 | 3.1E+01 | 3.4E+01 | 3.0E-02 | 3.8E+01 | NA | NA | NA |
| Water Consumption | HbA1c | MR Egger | 21 | 1.6E-01 | 1.8E-01 | 3.9E-01 | 4.7E+01 | 3.1E+01 | NA | NA | NA | -4.2E-03 | 5.5E-03 | 4.5E-01 |
| Water Consumption | HbA1c | WMed | 21 | 3.9E-02 | 4.3E-02 | 3.7E-01 | 4.7E+01 | 3.1E+01 | NA | NA | NA | NA | NA | NA |
| Water Consumption | HbA1c | WMode | 21 | 6.2E-02 | 7.2E-02 | 4.0E-01 | 4.7E+01 | 3.1E+01 | NA | NA | NA | NA | NA | NA |
| Water Consumption | HDL | IVW | 22 | 1.4E-01 | 1.1E-01 | 1.8E-01 | 4.7E+01 | 3.1E+01 | 2.0E+02 | 1.3E-31 | 9.0E+01 | NA | NA | NA |
| Water Consumption | HDL | MR Egger | 22 | 9.2E-02 | 5.1E-01 | 8.6E-01 | 4.7E+01 | 3.1E+01 | NA | NA | NA | 1.6E-03 | 1.6E-02 | 9.2E-01 |
| Water Consumption | HDL | WMed | 22 | 7.3E-02 | 6.1E-02 | 2.3E-01 | 4.7E+01 | 3.1E+01 | NA | NA | NA | NA | NA | NA |

| | | | | | | | | | | | | | | |
|---|---|---|---|---|---|---|---|---|---|---|---|---|---|---|
| Water Consumption | HDL | WMode | 22 | 1.0E-01 | 8.7E-02 | 2.6E-01 | 4.7E+01 | 3.1E+01 | NA | NA | NA | NA | NA | NA |
| Water Consumption | LDL | IVW | 22 | -1.3E-01 | 2.4E-01 | 6.0E-01 | 4.7E+01 | 3.1E+01 | 8.7E+02 | 7.5E-171 | 9.8E+01 | NA | NA | NA |
| Water Consumption | LDL | MR Egger | 22 | -3.5E-01 | 1.1E+00 | 7.6E-01 | 4.7E+01 | 3.1E+01 | NA | NA | NA | 7.2E-03 | 3.6E-02 | 8.4E-01 |
| Water Consumption | LDL | WMed | 22 | 6.2E-02 | 5.5E-02 | 2.6E-01 | 4.7E+01 | 3.1E+01 | NA | NA | NA | NA | NA | NA |
| Water Consumption | LDL | WMode | 22 | 7.4E-02 | 7.5E-02 | 3.4E-01 | 4.7E+01 | 3.1E+01 | NA | NA | NA | NA | NA | NA |
| Water Consumption | MetS | IVW | 43 | 1.7E-01 | 1.1E-01 | 1.4E-01 | 4.2E+01 | 3.0E+01 | 3.9E+02 | 2.0E-58 | 8.9E+01 | NA | NA | NA |
| Water Consumption | MetS | MR Egger | 43 | 1.9E-01 | 6.1E-01 | 7.5E-01 | 4.2E+01 | 3.0E+01 | NA | NA | NA | -7.3E-04 | 1.8E-02 | 9.7E-01 |
| Water Consumption | MetS | WMed | 43 | 1.2E-01 | 6.7E-02 | 7.3E-02 | 4.2E+01 | 3.0E+01 | NA | NA | NA | NA | NA | NA |
| Water Consumption | MetS | WMode | 43 | 2.3E-01 | 1.3E-01 | 9.9E-02 | 4.2E+01 | 3.0E+01 | NA | NA | NA | NA | NA | NA |
| Water Consumption | SBP | IVW | 38 | 1.9E-01 | 4.7E-01 | 6.9E-01 | 4.2E+01 | 3.0E+01 | 2.4E+02 | 2.4E-31 | 8.4E+01 | NA | NA | NA |
| Water Consumption | SBP | MR Egger | 38 | -1.1E+00 | 2.4E+00 | 6.6E-01 | 4.2E+01 | 3.0E+01 | NA | NA | NA | 3.8E-02 | 7.2E-02 | 6.0E-01 |
| Water Consumption | SBP | WMed | 38 | 7.6E-01 | 3.5E-01 | 3.2E-02 | 4.2E+01 | 3.0E+01 | NA | NA | NA | NA | NA | NA |
| Water Consumption | SBP | WMode | 38 | 1.2E+00 | 5.2E-01 | 2.2E-02 | 4.2E+01 | 3.0E+01 | NA | NA | NA | NA | NA | NA |
| Water Consumption | T2D | IVW | 33 | -1.5E-01 | 1.6E-01 | 3.6E-01 | 4.0E+01 | 3.0E+01 | 3.1E+02 | 7.3E-48 | 9.0E+01 | NA | NA | NA |
| Water Consumption | T2D | MR Egger | 33 | -3.1E-01 | 9.7E-01 | 7.5E-01 | 4.0E+01 | 3.0E+01 | NA | NA | NA | 4.6E-03 | 2.8E-02 | 8.7E-01 |

| | | | | | | | | | | | | | | |
|---|---|---|---|---|---|---|---|---|---|---|---|---|---|---|
| Water Consumption | T2D | WMed | 33 | -6.0E-02 | 8.9E-02 | 5.0E-01 | 4.0E+01 | 3.0E+01 | NA | NA | NA | NA | NA | NA |
| Water Consumption | T2D | WMode | 33 | -3.5E-02 | 1.7E-01 | 8.3E-01 | 4.0E+01 | 3.0E+01 | NA | NA | NA | NA | NA | NA |
| Water Consumption | Triglycerides | IVW | 22 | -2.3E-01 | 1.7E-01 | 1.8E-01 | 4.7E+01 | 3.1E+01 | 5.6E+02 | 3.9E-106 | 9.6E+01 | NA | NA | NA |
| Water Consumption | Triglycerides | MR Egger | 22 | -8.0E-02 | 8.1E-01 | 9.2E-01 | 4.7E+01 | 3.1E+01 | NA | NA | NA | -4.9E-03 | 2.6E-02 | 8.5E-01 |
| Water Consumption | Triglycerides | WMed | 22 | -7.7E-02 | 6.5E-02 | 2.4E-01 | 4.7E+01 | 3.1E+01 | NA | NA | NA | NA | NA | NA |
| Water Consumption | Triglycerides | WMode | 22 | -9.6E-02 | 1.1E-01 | 4.1E-01 | 4.7E+01 | 3.1E+01 | NA | NA | NA | NA | NA | NA |
| x_11440 | BMI | IVW | 3 | -2.4E-02 | 1.6E-02 | 1.3E-01 | 2.4E+02 | 3.2E+01 | 1.4E+00 | 5.0E-01 | 0.0E+00 | NA | NA | NA |
| x_11440 | BMI | MR Egger | 3 | -4.5E-02 | 2.4E-02 | 3.1E-01 | 2.4E+02 | 3.2E+01 | NA | NA | NA | 2.7E-03 | 2.3E-03 | 4.5E-01 |
| x_11440 | BMI | WMed | 3 | -2.5E-02 | 1.6E-02 | 1.1E-01 | 2.4E+02 | 3.2E+01 | NA | NA | NA | NA | NA | NA |
| x_11440 | BMI | WMode | 3 | -2.8E-02 | 1.5E-02 | 2.1E-01 | 2.4E+02 | 3.2E+01 | NA | NA | NA | NA | NA | NA |
| x_11440 | DBP | IVW | 3 | -2.6E-01 | 1.4E-01 | 5.9E-02 | 2.4E+02 | 3.2E+01 | 4.7E-01 | 7.9E-01 | 0.0E+00 | NA | NA | NA |
| x_11440 | DBP | MR Egger | 3 | -3.7E-01 | 2.1E-01 | 3.3E-01 | 2.4E+02 | 3.2E+01 | NA | NA | NA | 1.4E-02 | 2.0E-02 | 6.2E-01 |
| x_11440 | DBP | WMed | 3 | -2.7E-01 | 1.4E-01 | 5.9E-02 | 2.4E+02 | 3.2E+01 | NA | NA | NA | NA | NA | NA |
| x_11440 | DBP | WMode | 3 | -2.8E-01 | 1.5E-01 | 2.0E-01 | 2.4E+02 | 3.2E+01 | NA | NA | NA | NA | NA | NA |
| x_11440 | HbA1c | IVW | 3 | 3.8E-02 | 2.8E-02 | 1.7E-01 | 2.4E+02 | 3.2E+01 | 7.2E-01 | 7.0E-01 | 0.0E+00 | NA | NA | NA |
| x_11440 | HbA1c | MR Egger | 3 | 4.5E-02 | 4.3E-02 | 4.9E-01 | 2.4E+02 | 3.2E+01 | NA | NA | NA | -7.9E-04 | 4.0E-03 | 8.8E-01 |
| x_11440 | HbA1c | WMed | 3 | 4.2E-02 | 2.8E-02 | 1.3E-01 | 2.4E+02 | 3.2E+01 | NA | NA | NA | NA | NA | NA |
| x_11440 | HbA1c | WMode | 3 | 4.3E-02 | 2.9E-02 | 2.7E-01 | 2.4E+02 | 3.2E+01 | NA | NA | NA | NA | NA | NA |
| x_11440 | HDL | IVW | 3 | -6.6E-02 | 9.1E-02 | 4.7E-01 | 2.4E+02 | 3.2E+01 | 1.1E+01 | 3.7E-03 | 0.0E+00 | NA | NA | NA |
| x_11440 | HDL | MR Egger | 3 | 6.9E-02 | 5.6E-02 | 4.3E-01 | 2.4E+02 | 3.2E+01 | NA | NA | NA | -1.4E-02 | 4.2E-03 | 1.9E-01 |
| x_11440 | HDL | WMed | 3 | -3.1E-02 | 4.1E-02 | 4.5E-01 | 2.4E+02 | 3.2E+01 | NA | NA | NA | NA | NA | NA |

| | | | | | | | | | | | | | | |
|---|---|---|---|---|---|---|---|---|---|---|---|---|---|---|
| x_11440 | HDL | WMode | 3 | -2.2E-02 | 4.0E-02 | 6.4E-01 | 2.4E+02 | 3.2E+01 | NA | NA | NA | NA | NA | NA |
| x_11440 | LDL | IVW | 3 | -4.1E-02 | 7.9E-02 | 6.0E-01 | 2.4E+02 | 3.2E+01 | 7.2E+00 | 2.8E-02 | 0.0E+00 | NA | NA | NA |
| x_11440 | LDL | MR Egger | 3 | -1.6E-01 | 6.1E-02 | 2.3E-01 | 2.4E+02 | 3.2E+01 | NA | NA | NA | 1.2E-02 | 4.6E-03 | 2.3E-01 |
| x_11440 | LDL | WMed | 3 | -6.6E-02 | 4.2E-02 | 1.2E-01 | 2.4E+02 | 3.2E+01 | NA | NA | NA | NA | NA | NA |
| x_11440 | LDL | WMode | 3 | -7.8E-02 | 4.7E-02 | 2.4E-01 | 2.4E+02 | 3.2E+01 | NA | NA | NA | NA | NA | NA |
| x_11440 | MetS | IVW | 3 | -3.8E-02 | 8.1E-02 | 6.4E-01 | 2.4E+02 | 3.2E+01 | 5.0E+00 | 8.2E-02 | 4.2E+00 | NA | NA | NA |
| x_11440 | MetS | MR Egger | 3 | 6.7E-02 | 1.2E-01 | 6.6E-01 | 2.4E+02 | 3.2E+01 | NA | NA | NA | -1.3E-02 | 1.1E-02 | 4.5E-01 |
| x_11440 | MetS | WMed | 3 | -2.7E-02 | 5.2E-02 | 6.1E-01 | 2.4E+02 | 3.2E+01 | NA | NA | NA | NA | NA | NA |
| x_11440 | MetS | WMode | 3 | -4.5E-03 | 5.5E-02 | 9.4E-01 | 2.4E+02 | 3.2E+01 | NA | NA | NA | NA | NA | NA |
| x_11440 | SBP | IVW | 3 | -3.2E-01 | 2.4E-01 | 1.8E-01 | 2.4E+02 | 3.2E+01 | 1.5E+00 | 4.7E-01 | 0.0E+00 | NA | NA | NA |
| x_11440 | SBP | MR Egger | 3 | -5.1E-01 | 3.8E-01 | 4.1E-01 | 2.4E+02 | 3.2E+01 | NA | NA | NA | 2.3E-02 | 3.6E-02 | 6.4E-01 |
| x_11440 | SBP | WMed | 3 | -3.4E-01 | 2.4E-01 | 1.6E-01 | 2.4E+02 | 3.2E+01 | NA | NA | NA | NA | NA | NA |
| x_11440 | SBP | WMode | 3 | -3.5E-01 | 2.5E-01 | 3.0E-01 | 2.4E+02 | 3.2E+01 | NA | NA | NA | NA | NA | NA |
| x_11440 | T2D | IVW | 3 | 2.3E-02 | 1.0E-01 | 8.2E-01 | 2.4E+02 | 3.2E+01 | 5.4E+00 | 6.8E-02 | 0.0E+00 | NA | NA | NA |
| x_11440 | T2D | MR Egger | 3 | 1.9E-01 | 9.4E-02 | 3.0E-01 | 2.4E+02 | 3.2E+01 | NA | NA | NA | -1.9E-02 | 8.4E-03 | 2.6E-01 |
| x_11440 | T2D | WMed | 3 | 4.8E-02 | 6.6E-02 | 4.7E-01 | 2.4E+02 | 3.2E+01 | NA | NA | NA | NA | NA | NA |
| x_11440 | T2D | WMode | 3 | 5.8E-02 | 6.9E-02 | 4.9E-01 | 2.4E+02 | 3.2E+01 | NA | NA | NA | NA | NA | NA |
| x_11440 | Triglycerides | IVW | 3 | -3.6E-02 | 1.3E-01 | 7.9E-01 | 2.4E+02 | 3.2E+01 | 2.6E+01 | 2.5E-06 | 0.0E+00 | NA | NA | NA |
| x_11440 | Triglycerides | MR Egger | 3 | -2.4E-01 | 5.4E-02 | 1.4E-01 | 2.4E+02 | 3.2E+01 | NA | NA | NA | 2.1E-02 | 4.1E-03 | 1.2E-01 |
| x_11440 | Triglycerides | WMed | 3 | -7.9E-02 | 4.1E-02 | 5.5E-02 | 2.4E+02 | 3.2E+01 | NA | NA | NA | NA | NA | NA |
| x_11440 | Triglycerides | WMode | 3 | -9.9E-02 | 3.9E-02 | 1.3E-01 | 2.4E+02 | 3.2E+01 | NA | NA | NA | NA | NA | NA |

### Supplementary Table 6 - Cardiometabolic Traits' Effects on Candidate Biomarkers (MR)

| Exposure | Outcome | Method | nSNP | Beta | SE | P-Value | F Stat (Mean) | F Stat (Min) | Q Stat | Q Stat P-Value | I^2 | Egger Intercept | Egger Intercept SE | Egger Intercept P-Value |
|---|---|---|---|---|---|---|---|---|---|---|---|---|---|---|
| BMI | 25 Hydroxyvitamin D | Inverse variance weighted | 99 | -0.058810611 | 0.016518855 | 0.000370557 | 150.8 | 85.3 | 403.53 | 0 | 75.7 | NA | NA | NA |
| BMI | 25 Hydroxyvitamin D | MR Egger | 99 | -0.043291973 | 0.045594131 | 0.344722251 | 150.8 | 85.3 | NA | NA | NA | -0.000465 | 0.001272 | 0.7156 |
| BMI | 25 Hydroxyvitamin D | Weighted median | 99 | -0.02703231 | 0.01438759 | 0.060263203 | 150.8 | 85.3 | NA | NA | NA | NA | NA | NA |
| BMI | 25 Hydroxyvitamin D | Weighted mode | 99 | -0.005681454 | 0.04207629 | 0.892867127 | 150.8 | 85.3 | NA | NA | NA | NA | NA | NA |
| DBP | 25 Hydroxyvitamin D | Inverse variance weighted | 99 | 7.62E-05 | 0.001502489 | 0.959577191 | 210.2 | 114 | 364.34 | 0 | 73.1 | NA | NA | NA |
| DBP | 25 Hydroxyvitamin D | MR Egger | 99 | 0.000619026 | 0.005187572 | 0.905261711 | 210.2 | 114 | NA | NA | NA | -0.000168 | 0.00154 | 0.9131 |
| DBP | 25 Hydroxyvitamin D | Weighted median | 99 | 4.41E-05 | 0.001331468 | 0.97356655 | 210.2 | 114 | NA | NA | NA | NA | NA | NA |
| DBP | 25 Hydroxyvitamin D | Weighted mode | 99 | 0.001341533 | 0.002891526 | 0.643710148 | 210.2 | 114 | NA | NA | NA | NA | NA | NA |
| HDL | 25 Hydroxyvitamin D | Inverse variance weighted | 95 | 0.056973559 | 0.016601219 | 0.000599391 | 193.1 | 59.2 | 1627.21 | 0 | 94.2 | NA | NA | NA |
| HDL | 25 Hydroxyvitamin D | MR Egger | 95 | 0.072559186 | 0.032394632 | 0.027482985 | 193.1 | 59.2 | NA | NA | NA | -0.001149 | 0.002048 | 0.5761 |
| HDL | 25 Hydroxyvitamin D | Weighted median | 95 | 0.076266248 | 0.007769148 | 9.56E-23 | 193.1 | 59.2 | NA | NA | NA | NA | NA | NA |
| HDL | 25 Hydroxyvitamin D | Weighted mode | 95 | 0.077166402 | 0.00804071 | 1.32E-15 | 193.1 | 59.2 | NA | NA | NA | NA | NA | NA |

| | | | | | | | | | | | | | | |
|---|---|---|---|---|---|---|---|---|---|---|---|---|---|---|
| HbA1c | 25 Hydroxyvitamin D | Inverse variance weighted | 16 | -0.031941696 | 0.035003273 | 0.3614874 | 64.7 | 30.1 | 63.66 | 0 | 76.4 | NA | NA | NA |
| HbA1c | 25 Hydroxyvitamin D | MR Egger | 16 | -0.126730926 | 0.081045772 | 0.140206022 | 64.7 | 30.1 | NA | NA | NA | 0.003638 | 0.002819 | 0.2178 |
| HbA1c | 25 Hydroxyvitamin D | Weighted median | 16 | -0.033331164 | 0.027154214 | 0.219643485 | 64.7 | 30.1 | NA | NA | NA | NA | NA | NA |
| HbA1c | 25 Hydroxyvitamin D | Weighted mode | 16 | -0.045428061 | 0.035517935 | 0.220324154 | 64.7 | 30.1 | NA | NA | NA | NA | NA | NA |
| LDL | 25 Hydroxyvitamin D | Inverse variance weighted | 96 | 0.087265302 | 0.011221703 | 7.46E-15 | 213.2 | 56.4 | 1004.64 | 0 | 90.5 | NA | NA | NA |
| LDL | 25 Hydroxyvitamin D | MR Egger | 96 | 0.093981353 | 0.019548506 | 5.79E-06 | 213.2 | 56.4 | NA | NA | NA | -0.000598 | 0.001423 | 0.6751 |
| LDL | 25 Hydroxyvitamin D | Weighted median | 96 | 0.087169918 | 0.006765113 | 5.45E-38 | 213.2 | 56.4 | NA | NA | NA | NA | NA | NA |
| LDL | 25 Hydroxyvitamin D | Weighted mode | 96 | 0.087875901 | 0.006142064 | 1.96E-25 | 213.2 | 56.4 | NA | NA | NA | NA | NA | NA |
| MetS | 25 Hydroxyvitamin D | Inverse variance weighted | 89 | -0.020710068 | 0.013319191 | 0.119968849 | 101.5 | 41 | 1539.28 | 0 | 94.3 | NA | NA | NA |
| MetS | 25 Hydroxyvitamin D | MR Egger | 89 | -0.015063033 | 0.030448034 | 0.62204987 | 101.5 | 41 | NA | NA | NA | -0.000534 | 0.002586 | 0.8369 |
| MetS | 25 Hydroxyvitamin D | Weighted median | 89 | -0.020766065 | 0.006780626 | 0.002194535 | 101.5 | 41 | NA | NA | NA | NA | NA | NA |
| MetS | 25 Hydroxyvitamin D | Weighted mode | 89 | -0.018947501 | 0.010079668 | 0.063447722 | 101.5 | 41 | NA | NA | NA | NA | NA | NA |

| | | | | | | | | | | | | | | |
|---|---|---|---|---|---|---|---|---|---|---|---|---|---|---|
| SBP | 25 Hydroxyvitamin D | Inverse variance weighted | 100 | -0.00100149 | 0.000955913 | 0.294786474 | 189.9 | 103.5 | 409.76 | 0 | 75.8 | NA | NA | NA |
| SBP | 25 Hydroxyvitamin D | MR Egger | 100 | -0.002502885 | 0.00316856 | 0.431486102 | 189.9 | 103.5 | NA | NA | NA | 0.000758 | 0.001525 | 0.6202 |
| SBP | 25 Hydroxyvitamin D | Weighted median | 100 | -0.000696695 | 0.000803147 | 0.385692251 | 189.9 | 103.5 | NA | NA | NA | NA | NA | NA |
| SBP | 25 Hydroxyvitamin D | Weighted mode | 100 | -0.000404689 | 0.001847251 | 0.827041589 | 189.9 | 103.5 | NA | NA | NA | NA | NA | NA |
| T2D | 25 Hydroxyvitamin D | Inverse variance weighted | 100 | 0.015634362 | 0.007090706 | 0.027460464 | 89.6 | 38.5 | 609.9 | 0 | 83.8 | NA | NA | NA |
| T2D | 25 Hydroxyvitamin D | MR Egger | 100 | 0.008656798 | 0.01697168 | 0.61114711 | 89.6 | 38.5 | NA | NA | NA | 0.000618 | 0.001365 | 0.6516 |
| T2D | 25 Hydroxyvitamin D | Weighted median | 100 | 0.012667587 | 0.005616247 | 0.024100389 | 89.6 | 38.5 | NA | NA | NA | NA | NA | NA |
| T2D | 25 Hydroxyvitamin D | Weighted mode | 100 | 0.008362589 | 0.007004477 | 0.235373105 | 89.6 | 38.5 | NA | NA | NA | NA | NA | NA |
| TG | 25 Hydroxyvitamin D | Inverse variance weighted | 91 | 0.141499832 | 0.017113781 | 1.36E-16 | 149.6 | 43 | 1125.55 | 0 | 92 | NA | NA | NA |
| TG | 25 Hydroxyvitamin D | MR Egger | 91 | 0.128597378 | 0.03069004 | 6.55E-05 | 149.6 | 43 | NA | NA | NA | 0.000849 | 0.001674 | 0.6131 |
| TG | 25 Hydroxyvitamin D | Weighted median | 91 | 0.146409374 | 0.013218692 | 1.64E-28 | 149.6 | 43 | NA | NA | NA | NA | NA | NA |
| TG | 25 Hydroxyvitamin D | Weighted mode | 91 | 0.177950323 | 0.017103915 | 4.15E-17 | 149.6 | 43 | NA | NA | NA | NA | NA | NA |
| BMI | Bilirubin | Inverse variance weighted | 75 | 0.037821343 | 0.023424402 | 0.106394543 | 156.7 | 85.3 | 82.38 | 0.2362 | 10.2 | NA | NA | NA |

| | | | | | | | | | | | | | | |
|---|---|---|---|---|---|---|---|---|---|---|---|---|---|---|
| BMI | Bilirubin | MR Egger | 75 | 0.030933314 | 0.067835388 | 0.649739685 | 156.7 | 85.3 | NA | NA | NA | 0.000207 | 0.001908 | 0.9141 |
| BMI | Bilirubin | Weighted median | 75 | 0.0339413 | 0.036097588 | 0.347081621 | 156.7 | 85.3 | NA | NA | NA | NA | NA | NA |
| BMI | Bilirubin | Weighted mode | 75 | 0.052759059 | 0.055618703 | 0.34591942 | 156.7 | 85.3 | NA | NA | NA | NA | NA | NA |
| DBP | Bilirubin | Inverse variance weighted | 79 | -0.00106105 | 0.002568558 | 0.679539359 | 197 | 114 | 131.47 | 1.00E-04 | 40.7 | NA | NA | NA |
| DBP | Bilirubin | MR Egger | 79 | 0.007001936 | 0.0090908 | 0.443524805 | 197 | 114 | NA | NA | NA | -0.002478 | 0.00268 | 0.358 |
| DBP | Bilirubin | Weighted median | 79 | -0.003016048 | 0.00321685 | 0.348461289 | 197 | 114 | NA | NA | NA | NA | NA | NA |
| DBP | Bilirubin | Weighted mode | 79 | -0.00565341 | 0.006197229 | 0.364449078 | 197 | 114 | NA | NA | NA | NA | NA | NA |
| HbA1c | Bilirubin | Inverse variance weighted | 10 | 0.176628505 | 0.14517958 | 0.223748502 | 54.8 | 30.1 | 59.12 | 0 | 84.8 | NA | NA | NA |
| HbA1c | Bilirubin | MR Egger | 10 | -0.517358161 | 0.49808143 | 0.329315648 | 54.8 | 30.1 | NA | NA | NA | 0.020418 | 0.014088 | 0.1853 |
| HbA1c | Bilirubin | Weighted median | 10 | -0.120188618 | 0.088299965 | 0.173469439 | 54.8 | 30.1 | NA | NA | NA | NA | NA | NA |
| HbA1c | Bilirubin | Weighted mode | 10 | -0.116301266 | 0.090385015 | 0.230297132 | 54.8 | 30.1 | NA | NA | NA | NA | NA | NA |
| HDL | Bilirubin | Inverse variance weighted | 87 | -0.020936179 | 0.012882999 | 0.104140957 | 173.5 | 59.2 | 164.55 | 0 | 47.7 | NA | NA | NA |

| | | | | | | | | | | | | | | |
|---|---|---|---|---|---|---|---|---|---|---|---|---|---|---|
| HDL | Bilirubin | MR Egger | 87 | -0.054668042 | 0.027646251 | 0.051235677 | 173.5 | 59.2 | NA | NA | NA | 0.002332 | 0.001693 | 0.1721 |
| HDL | Bilirubin | Weighted median | 87 | -0.021823344 | 0.014751407 | 0.13903141 | 173.5 | 59.2 | NA | NA | NA | NA | NA | NA |
| HDL | Bilirubin | Weighted mode | 87 | -0.029946085 | 0.020376841 | 0.145315311 | 173.5 | 59.2 | NA | NA | NA | NA | NA | NA |
| LDL | Bilirubin | Inverse variance weighted | 77 | -0.005908362 | 0.010320083 | 0.566975746 | 209.8 | 56.8 | 111.77 | 0.0048 | 32 | NA | NA | NA |
| LDL | Bilirubin | MR Egger | 77 | -0.009013228 | 0.017687721 | 0.611845104 | 209.8 | 56.8 | NA | NA | NA | 0.000277 | 0.001276 | 0.8289 |
| LDL | Bilirubin | Weighted median | 77 | 0.002147642 | 0.012996188 | 0.868745865 | 209.8 | 56.8 | NA | NA | NA | NA | NA | NA |
| LDL | Bilirubin | Weighted mode | 77 | -0.003527557 | 0.011872335 | 0.767182363 | 209.8 | 56.8 | NA | NA | NA | NA | NA | NA |
| MetS | Bilirubin | Inverse variance weighted | 66 | -0.002996318 | 0.013596331 | 0.825577571 | 98.6 | 41 | 191.58 | 0 | 66.1 | NA | NA | NA |
| MetS | Bilirubin | MR Egger | 66 | 0.047383475 | 0.032197995 | 0.146021697 | 98.6 | 41 | NA | NA | NA | -0.004616 | 0.002683 | 0.0901 |
| MetS | Bilirubin | Weighted median | 66 | 0.017068869 | 0.012767799 | 0.181265552 | 98.6 | 41 | NA | NA | NA | NA | NA | NA |
| MetS | Bilirubin | Weighted mode | 66 | 0.022185627 | 0.014202617 | 0.123125074 | 98.6 | 41 | NA | NA | NA | NA | NA | NA |
| SBP | Bilirubin | Inverse variance weighted | 85 | -0.000588444 | 0.001525836 | 0.699753359 | 182.5 | 103.5 | 162.85 | 0 | 48.4 | NA | NA | NA |

| | | | | | | | | | | | | | | |
|---|---|---|---|---|---|---|---|---|---|---|---|---|---|---|
| SBP | Bilirubin | MR Egger | 85 | 0.007060611 | 0.004448051 | 0.116236033 | 182.5 | 103.5 | NA | NA | NA | -0.004055 | 0.002219 | 0.0712 |
| SBP | Bilirubin | Weighted median | 85 | -0.000383283 | 0.001817311 | 0.832960094 | 182.5 | 103.5 | NA | NA | NA | NA | NA | NA |
| SBP | Bilirubin | Weighted mode | 85 | 0.002381337 | 0.002864198 | 0.408096426 | 182.5 | 103.5 | NA | NA | NA | NA | NA | NA |
| T2D | Bilirubin | Inverse variance weighted | 86 | -0.011487805 | 0.008176017 | 0.160003041 | 92.5 | 38.6 | 98.44 | 0.1511 | 13.7 | NA | NA | NA |
| T2D | Bilirubin | MR Egger | 86 | -0.004339391 | 0.023623383 | 0.854698965 | 92.5 | 38.6 | NA | NA | NA | -0.000569 | 0.001763 | 0.7477 |
| T2D | Bilirubin | Weighted median | 86 | -0.019065289 | 0.011754818 | 0.104822024 | 92.5 | 38.6 | NA | NA | NA | NA | NA | NA |
| T2D | Bilirubin | Weighted mode | 86 | -0.018662903 | 0.018067794 | 0.304563135 | 92.5 | 38.6 | NA | NA | NA | NA | NA | NA |
| TG | Bilirubin | Inverse variance weighted | 76 | -0.00667934 | 0.021293969 | 0.753769505 | 159.8 | 43 | 299.34 | 0 | 74.9 | NA | NA | NA |
| TG | Bilirubin | MR Egger | 76 | -0.058955697 | 0.036098476 | 0.106676934 | 159.8 | 43 | NA | NA | NA | 0.003689 | 0.002072 | 0.0792 |
| TG | Bilirubin | Weighted median | 76 | -0.036101789 | 0.016529743 | 0.028958555 | 159.8 | 43 | NA | NA | NA | NA | NA | NA |
| TG | Bilirubin | Weighted mode | 76 | -0.029874132 | 0.016076682 | 0.067060912 | 159.8 | 43 | NA | NA | NA | NA | NA | NA |

| | | | | | | | | | | | | | | |
|---|---|---|---|---|---|---|---|---|---|---|---|---|---|---|
| BMI | Blood Urea Nitrogen | Inverse variance weighted | 75 | -0.132230565 | 0.035576083 | 0.000201731 | 156.7 | 85.3 | 275.46 | 0 | 73.1 | NA | NA | NA |
| BMI | Blood Urea Nitrogen | MR Egger | 75 | -0.428095703 | 0.096213024 | 3.03E-05 | 156.7 | 85.3 | NA | NA | NA | 0.008872 | 0.002705 | 0.0016 |
| BMI | Blood Urea Nitrogen | Weighted median | 75 | -0.080596913 | 0.035416522 | 0.022864759 | 156.7 | 85.3 | NA | NA | NA | NA | NA | NA |
| BMI | Blood Urea Nitrogen | Weighted mode | 75 | -0.052566576 | 0.076842994 | 0.496062853 | 156.7 | 85.3 | NA | NA | NA | NA | NA | NA |
| DBP | Blood Urea Nitrogen | Inverse variance weighted | 79 | -0.001020143 | 0.003488256 | 0.769942137 | 197 | 114 | 351.63 | 0 | 77.8 | NA | NA | NA |
| DBP | Blood Urea Nitrogen | MR Egger | 79 | 0.003518411 | 0.012403766 | 0.777434776 | 197 | 114 | NA | NA | NA | -0.001395 | 0.003657 | 0.7039 |
| DBP | Blood Urea Nitrogen | Weighted median | 79 | -0.005663796 | 0.002851403 | 0.046997966 | 197 | 114 | NA | NA | NA | NA | NA | NA |
| DBP | Blood Urea Nitrogen | Weighted mode | 79 | -0.009070322 | 0.004345423 | 0.04012396 | 197 | 114 | NA | NA | NA | NA | NA | NA |
| HbA1c | Blood Urea Nitrogen | Inverse variance weighted | 10 | 0.012615861 | 0.049761811 | 0.799862574 | 54.8 | 30.1 | 10.09 | 0.3433 | 10.8 | NA | NA | NA |
| HbA1c | Blood Urea Nitrogen | MR Egger | 10 | 0.030617531 | 0.191444405 | 0.876901215 | 54.8 | 30.1 | NA | NA | NA | -0.00053 | 0.005417 | 0.9245 |
| HbA1c | Blood Urea Nitrogen | Weighted median | 10 | -0.009709426 | 0.061971331 | 0.875500127 | 54.8 | 30.1 | NA | NA | NA | NA | NA | NA |
| HbA1c | Blood Urea Nitrogen | Weighted mode | 10 | -0.010674585 | 0.082031804 | 0.899327665 | 54.8 | 30.1 | NA | NA | NA | NA | NA | NA |

| | | | | | | | | | | | | | | |
|---|---|---|---|---|---|---|---|---|---|---|---|---|---|---|
| HDL | Blood Urea Nitrogen | Inverse variance weighted | 87 | -0.002482399 | 0.009546535 | 0.794839511 | 173.5 | 59.2 | 131.01 | 0.0013 | 34.4 | NA | NA | NA |
| HDL | Blood Urea Nitrogen | MR Egger | 87 | -0.041423301 | 0.020158288 | 0.042959047 | 173.5 | 59.2 | NA | NA | NA | 0.002692 | 0.001235 | 0.032 |
| HDL | Blood Urea Nitrogen | Weighted median | 87 | -0.009884072 | 0.012604101 | 0.432925767 | 173.5 | 59.2 | NA | NA | NA | NA | NA | NA |
| HDL | Blood Urea Nitrogen | Weighted mode | 87 | -0.015983739 | 0.013051035 | 0.224028423 | 173.5 | 59.2 | NA | NA | NA | NA | NA | NA |
| LDL | Blood Urea Nitrogen | Inverse variance weighted | 77 | 0.001686189 | 0.010505381 | 0.872481607 | 209.8 | 56.8 | 168.01 | 0 | 54.8 | NA | NA | NA |
| LDL | Blood Urea Nitrogen | MR Egger | 77 | 0.040618865 | 0.01713679 | 0.020346271 | 209.8 | 56.8 | NA | NA | NA | -0.00347 | 0.001236 | 0.0064 |
| LDL | Blood Urea Nitrogen | Weighted median | 77 | 0.018969522 | 0.011195212 | 0.090183292 | 209.8 | 56.8 | NA | NA | NA | NA | NA | NA |
| LDL | Blood Urea Nitrogen | Weighted mode | 77 | 0.014463582 | 0.010871726 | 0.187369754 | 209.8 | 56.8 | NA | NA | NA | NA | NA | NA |
| MetS | Blood Urea Nitrogen | Inverse variance weighted | 66 | 0.001776548 | 0.010234865 | 0.862197047 | 98.6 | 41 | 157.46 | 0 | 58.7 | NA | NA | NA |
| MetS | Blood Urea Nitrogen | MR Egger | 66 | -0.015108462 | 0.024677451 | 0.54254977 | 98.6 | 41 | NA | NA | NA | 0.001547 | 0.002057 | 0.4545 |
| MetS | Blood Urea Nitrogen | Weighted median | 66 | -0.000597272 | 0.010791344 | 0.955861742 | 98.6 | 41 | NA | NA | NA | NA | NA | NA |
| MetS | Blood Urea Nitrogen | Weighted mode | 66 | -0.010404699 | 0.012534046 | 0.409512345 | 98.6 | 41 | NA | NA | NA | NA | NA | NA |

| | | | | | | | | | | | | | | |
|---|---|---|---|---|---|---|---|---|---|---|---|---|---|---|
| SBP | Blood Urea Nitrogen | Inverse variance weighted | 85 | -0.001774552 | 0.001808126 | 0.326379932 | 182.5 | 103.5 | 331.57 | 0 | 74.7 | NA | NA | NA |
| SBP | Blood Urea Nitrogen | MR Egger | 85 | 0.000502361 | 0.005369598 | 0.925686846 | 182.5 | 103.5 | NA | NA | NA | -0.001207 | 0.002679 | 0.6534 |
| SBP | Blood Urea Nitrogen | Weighted median | 85 | -0.003599253 | 0.001596009 | 0.024123415 | 182.5 | 103.5 | NA | NA | NA | NA | NA | NA |
| SBP | Blood Urea Nitrogen | Weighted mode | 85 | -0.005344546 | 0.002303784 | 0.022772859 | 182.5 | 103.5 | NA | NA | NA | NA | NA | NA |
| T2D | Blood Urea Nitrogen | Inverse variance weighted | 86 | -0.026922641 | 0.011182211 | 0.016056384 | 92.5 | 38.6 | 267.11 | 0 | 68.2 | NA | NA | NA |
| T2D | Blood Urea Nitrogen | MR Egger | 86 | -0.031876048 | 0.032315056 | 0.326762443 | 92.5 | 38.6 | NA | NA | NA | 0.000394 | 0.002412 | 0.8705 |
| T2D | Blood Urea Nitrogen | Weighted median | 86 | -0.021899982 | 0.0100 8208 | 0.029842908 | 92.5 | 38.6 | NA | NA | NA | NA | NA | NA |
| T2D | Blood Urea Nitrogen | Weighted mode | 86 | -0.013965146 | 0.013804642 | 0.314587587 | 92.5 | 38.6 | NA | NA | NA | NA | NA | NA |
| TG | Blood Urea Nitrogen | Inverse variance weighted | 76 | -0.015046846 | 0.012873817 | 0.24248647 | 159.8 | 43 | 158.78 | 0 | 52.8 | NA | NA | NA |
| TG | Blood Urea Nitrogen | MR Egger | 76 | 0.003880537 | 0.022117762 | 0.861206133 | 159.8 | 43 | NA | NA | NA | -0.001336 | 0.00127 | 0.2962 |
| TG | Blood Urea Nitrogen | Weighted median | 76 | 0.004316783 | 0.014346216 | 0.763490426 | 159.8 | 43 | NA | NA | NA | NA | NA | NA |
| TG | Blood Urea Nitrogen | Weighted mode | 76 | 0.001158409 | 0.013654768 | 0.932618201 | 159.8 | 43 | NA | NA | NA | NA | NA | NA |

| | | | | | | | | | | | | | | |
|---|---|---|---|---|---|---|---|---|---|---|---|---|---|---|
| BMI | Coffee Consumption | Inverse variance weighted | 100 | -0.138708555 | 0.014816641 | 7.85E-21 | 150.7 | 85.3 | 490.96 | 0 | 79.8 | NA | NA | NA |
| BMI | Coffee Consumption | MR Egger | 100 | -0.291352211 | 0.037667821 | 9.29E-12 | 150.7 | 85.3 | NA | NA | NA | 0.004564 | 0.00105 | 0 |
| BMI | Coffee Consumption | Weighted median | 100 | -0.152087886 | 0.013679532 | 1.03E-28 | 150.7 | 85.3 | NA | NA | NA | NA | NA | NA |
| BMI | Coffee Consumption | Weighted mode | 100 | -0.23383983 | 0.023743074 | 2.34E-16 | 150.7 | 85.3 | NA | NA | NA | NA | NA | NA |
| DBP | Coffee Consumption | Inverse variance weighted | 100 | 0.006219905 | 0.001109493 | 2.07E-08 | 209.5 | 114 | 299.92 | 0 | 67 | NA | NA | NA |
| DBP | Coffee Consumption | MR Egger | 100 | 0.004610683 | 0.003829531 | 0.231496835 | 209.5 | 114 | NA | NA | NA | 0.000498 | 0.001133 | 0.6615 |
| DBP | Coffee Consumption | Weighted median | 100 | 0.004276186 | 0.001089058 | 8.62E-05 | 209.5 | 114 | NA | NA | NA | NA | NA | NA |
| DBP | Coffee Consumption | Weighted mode | 100 | 0.004218239 | 0.001967555 | 0.034489983 | 209.5 | 114 | NA | NA | NA | NA | NA | NA |
| HbA1c | Coffee Consumption | Inverse variance weighted | 16 | 0.010492886 | 0.014658756 | 0.474108591 | 64.7 | 30.1 | 16.48 | 0.3512 | 9 | NA | NA | NA |
| HbA1c | Coffee Consumption | MR Egger | 16 | -0.038285732 | 0.0331533 | 0.267499704 | 64.7 | 30.1 | NA | NA | NA | 0.001869 | 0.001152 | 0.1269 |
| HbA1c | Coffee Consumption | Weighted median | 16 | -0.011760244 | 0.020113528 | 0.558754005 | 64.7 | 30.1 | NA | NA | NA | NA | NA | NA |
| HbA1c | Coffee Consumption | Weighted mode | 16 | -0.012591067 | 0.023262046 | 0.596269004 | 64.7 | 30.1 | NA | NA | NA | NA | NA | NA |

| | | | | | | | | | | | | | | |
|---|---|---|---|---|---|---|---|---|---|---|---|---|---|---|
| HDL | Coffee Consumption | Inverse variance weighted | 100 | -0.006403244 | 0.004628976 | 0.16657411 | 193.1 | 59.2 | 208.4 | 0 | 52.5 | NA | NA | NA |
| HDL | Coffee Consumption | MR Egger | 100 | 0.003229417 | 0.009110651 | 0.723751182 | 193.1 | 59.2 | NA | NA | NA | -0.000704 | 0.000574 | 0.223 |
| HDL | Coffee Consumption | Weighted median | 100 | 8.02E-05 | 0.005107904 | 0.987479893 | 193.1 | 59.2 | NA | NA | NA | NA | NA | NA |
| HDL | Coffee Consumption | Weighted mode | 100 | 0.00087576 | 0.00576795 | 0.879628323 | 193.1 | 59.2 | NA | NA | NA | NA | NA | NA |
| LDL | Coffee Consumption | Inverse variance weighted | 100 | 0.003947272 | 0.003393274 | 0.24472253 | 207.8 | 56.4 | 144.03 | 0.0021 | 31.3 | NA | NA | NA |
| LDL | Coffee Consumption | MR Egger | 100 | 0.010496076 | 0.005904431 | 0.078561469 | 207.8 | 56.4 | NA | NA | NA | -0.000571 | 0.000422 | 0.1793 |
| LDL | Coffee Consumption | Weighted median | 100 | 0.002119906 | 0.00461747 | 0.64615837 | 207.8 | 56.4 | NA | NA | NA | NA | NA | NA |
| LDL | Coffee Consumption | Weighted mode | 100 | 0.003110328 | 0.005901292 | 0.599331845 | 207.8 | 56.4 | NA | NA | NA | NA | NA | NA |
| MetS | Coffee Consumption | Inverse variance weighted | 96 | -0.003369162 | 0.006513871 | 0.604996371 | 97.8 | 41 | 616.29 | 0 | 84.6 | NA | NA | NA |
| MetS | Coffee Consumption | MR Egger | 96 | -0.018770925 | 0.0151516 | 0.218476753 | 97.8 | 41 | NA | NA | NA | 0.001429 | 0.00127 | 0.2632 |
| MetS | Coffee Consumption | Weighted median | 96 | -0.007925792 | 0.004192387 | 0.058688449 | 97.8 | 41 | NA | NA | NA | NA | NA | NA |
| MetS | Coffee Consumption | Weighted mode | 96 | -0.006978466 | 0.006477442 | 0.284052452 | 97.8 | 41 | NA | NA | NA | NA | NA | NA |
| SBP | Coffee Consumption | Inverse variance weighted | 100 | 0.001829723 | 0.000719249 | 0.010961099 | 189.9 | 103.5 | 344.08 | 0 | 71.2 | NA | NA | NA |

| | | | | | | | | | | | | | | |
|---|---|---|---|---|---|---|---|---|---|---|---|---|---|---|
| SBP | Coffee Consumption | MR Egger | 100 | 0.005264659 | 0.002361213 | 0.028054048 | 189.9 | 103.5 | NA | NA | NA | -0.001735 | 0.001136 | 0.1302 |
| SBP | Coffee Consumption | Weighted median | 100 | 0.001882592 | 0.000626919 | 0.002673961 | 189.9 | 103.5 | NA | NA | NA | NA | NA | NA |
| SBP | Coffee Consumption | Weighted mode | 100 | 0.002320442 | 0.001070394 | 0.03256794 | 189.9 | 103.5 | NA | NA | NA | NA | NA | NA |
| T2D | Coffee Consumption | Inverse variance weighted | 100 | 0.002213344 | 0.005410583 | 0.682483365 | 89.6 | 38.5 | 525.17 | 0 | 81.1 | NA | NA | NA |
| T2D | Coffee Consumption | MR Egger | 100 | 0.010717182 | 0.01293496 | 0.409375888 | 89.6 | 38.5 | NA | NA | NA | -0.000753 | 0.00104 | 0.4707 |
| T2D | Coffee Consumption | Weighted median | 100 | 0.007880539 | 0.004830126 | 0.102776636 | 89.6 | 38.5 | NA | NA | NA | NA | NA | NA |
| T2D | Coffee Consumption | Weighted mode | 100 | 0.007641655 | 0.00444145 | 0.088459985 | 89.6 | 38.5 | NA | NA | NA | NA | NA | NA |
| TG | Coffee Consumption | Inverse variance weighted | 100 | 0.038498486 | 0.007238134 | 1.04E-07 | 149.4 | 43 | 360.43 | 0 | 72.5 | NA | NA | NA |
| TG | Coffee Consumption | MR Egger | 100 | 0.03362736 | 0.013262266 | 0.012807659 | 149.4 | 43 | NA | NA | NA | 0.000313 | 0.000712 | 0.6616 |
| TG | Coffee Consumption | Weighted median | 100 | 0.024764451 | 0.00696442 | 0.000376755 | 149.4 | 43 | NA | NA | NA | NA | NA | NA |
| TG | Coffee Consumption | Weighted mode | 100 | 0.015249118 | 0.007220921 | 0.037221205 | 149.4 | 43 | NA | NA | NA | NA | NA | NA |
| BMI | Cotinine | Inverse variance weighted | 100 | -0.052355469 | 0.008974231 | 5.41E-09 | 150.7 | 85.3 | 362.24 | 0 | 72.7 | NA | NA | NA |
| BMI | Cotinine | MR Egger | 100 | -0.042621199 | 0.024890472 | 0.089994755 | 150.7 | 85.3 | NA | NA | NA | -0.000291 | 0.000694 | 0.6757 |
| BMI | Cotinine | Weighted median | 100 | -0.057652769 | 0.008996813 | 1.47E-10 | 150.7 | 85.3 | NA | NA | NA | NA | NA | NA |

| | | | | | | | | | | | | | | |
|---|---|---|---|---|---|---|---|---|---|---|---|---|---|---|
| BMI | Cotinine | Weighted mode | 100 | -0.092179009 | 0.03967038 | 0.0221907 | 150.7 | 85.3 | NA | NA | NA | NA | NA | NA |
| DBP | Cotinine | Inverse variance weighted | 100 | 0.001157554 | 0.000705964 | 0.101072065 | 209.5 | 114 | 244.15 | 0 | 59.5 | NA | NA | NA |
| DBP | Cotinine | MR Egger | 100 | -0.001923117 | 0.002417275 | 0.428203486 | 209.5 | 114 | NA | NA | NA | 0.000953 | 0.000715 | 0.1859 |
| DBP | Cotinine | Weighted median | 100 | 0.001056986 | 0.000760915 | 0.164802772 | 209.5 | 114 | NA | NA | NA | NA | NA | NA |
| DBP | Cotinine | Weighted mode | 100 | 0.001182491 | 0.00145525 | 0.41841508 | 209.5 | 114 | NA | NA | NA | NA | NA | NA |
| HbA1c | Cotinine | Inverse variance weighted | 16 | -0.003012007 | 0.009863493 | 0.760084936 | 64.7 | 30.1 | 11.64 | 0.7057 | 0 | NA | NA | NA |
| HbA1c | Cotinine | MR Egger | 16 | 0.029780158 | 0.02337747 | 0.223447445 | 64.7 | 30.1 | NA | NA | NA | -0.001256 | 0.000812 | 0.1441 |
| HbA1c | Cotinine | Weighted median | 16 | -0.000166392 | 0.013901888 | 0.990450353 | 64.7 | 30.1 | NA | NA | NA | NA | NA | NA |
| HbA1c | Cotinine | Weighted mode | 16 | 0.006526655 | 0.017815636 | 0.719224032 | 64.7 | 30.1 | NA | NA | NA | NA | NA | NA |
| HDL | Cotinine | Inverse variance weighted | 100 | 0.002682679 | 0.003116862 | 0.389403942 | 193.1 | 59.2 | 189.91 | 0 | 47.9 | NA | NA | NA |
| HDL | Cotinine | MR Egger | 100 | 0.016106348 | 0.005977838 | 0.008299113 | 193.1 | 59.2 | NA | NA | NA | -0.000982 | 0.000377 | 0.0106 |
| HDL | Cotinine | Weighted median | 100 | 0.002134699 | 0.003769945 | 0.571229716 | 193.1 | 59.2 | NA | NA | NA | NA | NA | NA |
| HDL | Cotinine | Weighted mode | 100 | 0.004363224 | 0.004007249 | 0.278871093 | 193.1 | 59.2 | NA | NA | NA | NA | NA | NA |

| | | | | | | | | | | | | | | |
|---|---|---|---|---|---|---|---|---|---|---|---|---|---|---|
| LDL | Cotinine | Inverse variance weighted | 100 | 0.010756816 | 0.002233197 | 1.46E-06 | 207.8 | 56.4 | 125.46 | 0.0374 | 21.1 | NA | NA | NA |
| LDL | Cotinine | MR Egger | 100 | 0.008374274 | 0.003910819 | 0.034728872 | 207.8 | 56.4 | NA | NA | NA | 0.000208 | 0.00028 | 0.4593 |
| LDL | Cotinine | Weighted median | 100 | 0.011241931 | 0.003236134 | 0.000512995 | 207.8 | 56.4 | NA | NA | NA | NA | NA | NA |
| LDL | Cotinine | Weighted mode | 100 | 0.001028293 | 0.005495721 | 0.851958851 | 207.8 | 56.4 | NA | NA | NA | NA | NA | NA |
| MetS | Cotinine | Inverse variance weighted | 96 | 0.006374746 | 0.00272129 | 0.019152686 | 97.8 | 41 | 216.25 | 0 | 56.1 | NA | NA | NA |
| MetS | Cotinine | MR Egger | 96 | -0.003576372 | 0.006269142 | 0.569718573 | 97.8 | 41 | NA | NA | NA | 0.000924 | 0.000526 | 0.0821 |
| MetS | Cotinine | Weighted median | 96 | 0.00315731 | 0.003013271 | 0.29473007 | 97.8 | 41 | NA | NA | NA | NA | NA | NA |
| MetS | Cotinine | Weighted mode | 96 | 0.001358708 | 0.006498634 | 0.83483665 | 97.8 | 41 | NA | NA | NA | NA | NA | NA |
| SBP | Cotinine | Inverse variance weighted | 100 | 0.000616234 | 0.000420338 | 0.14263633 | 189.9 | 103.5 | 236.31 | 0 | 58.1 | NA | NA | NA |
| SBP | Cotinine | MR Egger | 100 | 0.001118744 | 0.00139514 | 0.424558943 | 189.9 | 103.5 | NA | NA | NA | -0.000254 | 0.000672 | 0.7063 |
| SBP | Cotinine | Weighted median | 100 | 0.000696691 | 0.000427726 | 0.103349695 | 189.9 | 103.5 | NA | NA | NA | NA | NA | NA |
| SBP | Cotinine | Weighted mode | 100 | 0.000847675 | 0.000842451 | 0.316772166 | 189.9 | 103.5 | NA | NA | NA | NA | NA | NA |
| T2D | Cotinine | Inverse variance weighted | 100 | -0.000291635 | 0.002335998 | 0.900647158 | 89.6 | 38.5 | 196.85 | 0 | 49.7 | NA | NA | NA |
| T2D | Cotinine | MR Egger | 100 | 0.008280094 | 0.005517416 | 0.136643163 | 89.6 | 38.5 | NA | NA | NA | -0.000759 | 0.000444 | 0.0902 |

| | | | | | | | | | | | | | | |
|---|---|---|---|---|---|---|---|---|---|---|---|---|---|---|
| T2D | Cotinine | Weighted median | 100 | 0.000610389 | 0.003303384 | 0.853404065 | 89.6 | 38.5 | NA | NA | NA | NA | NA | NA |
| T2D | Cotinine | Weighted mode | 100 | 0.002421798 | 0.003343821 | 0.470613871 | 89.6 | 38.5 | NA | NA | NA | NA | NA | NA |
| TG | Cotinine | Inverse variance weighted | 100 | -0.004484762 | 0.003054057 | 0.141979227 | 149.4 | 43 | 129.03 | 0.0229 | 23.3 | NA | NA | NA |
| TG | Cotinine | MR Egger | 100 | 0.004306875 | 0.005499297 | 0.435416941 | 149.4 | 43 | NA | NA | NA | -0.000564 | 0.000295 | 0.0589 |
| TG | Cotinine | Weighted median | 100 | -0.001884519 | 0.004240419 | 0.656740417 | 149.4 | 43 | NA | NA | NA | NA | NA | NA |
| TG | Cotinine | Weighted mode | 100 | 0.001744281 | 0.006246925 | 0.780656743 | 149.4 | 43 | NA | NA | NA | NA | NA | NA |
| BMI | Creatinine | Inverse variance weighted | 99 | 0.091137106 | 0.020791868 | 1.17E-05 | 150.8 | 85.3 | 467.14 | 0 | 79 | NA | NA | NA |
| BMI | Creatinine | MR Egger | 99 | 0.005884015 | 0.056677562 | 0.917530033 | 150.8 | 85.3 | NA | NA | NA | 0.002554 | 0.001581 | 0.1096 |
| BMI | Creatinine | Weighted median | 99 | 0.063633931 | 0.018732216 | 0.000681211 | 150.8 | 85.3 | NA | NA | NA | NA | NA | NA |
| BMI | Creatinine | Weighted mode | 99 | -0.025718503 | 0.076243079 | 0.736595233 | 150.8 | 85.3 | NA | NA | NA | NA | NA | NA |
| DBP | Creatinine | Inverse variance weighted | 100 | 6.56E-05 | 0.001613754 | 0.967588633 | 209.5 | 114 | 312.3 | 0 | 68.3 | NA | NA | NA |
| DBP | Creatinine | MR Egger | 100 | -0.013083491 | 0.005398993 | 0.017214438 | 209.5 | 114 | NA | NA | NA | 0.004069 | 0.001598 | 0.0125 |
| DBP | Creatinine | Weighted median | 100 | 0.000324853 | 0.001588839 | 0.837994724 | 209.5 | 114 | NA | NA | NA | NA | NA | NA |

| | | | | | | | | | | | | | | |
|---|---|---|---|---|---|---|---|---|---|---|---|---|---|---|
| DBP | Creatinine | Weighted mode | 100 | 0.00067668 | 0.00401861 | 0.866622653 | 209.5 | 114 | NA | NA | NA | NA | NA | NA |
| HbA1c | Creatinine | Inverse variance weighted | 16 | 0.062396755 | 0.02036305 | 0.002182423 | 64.7 | 30.1 | 15.68 | 0.4038 | 4.3 | NA | NA | NA |
| HbA1c | Creatinine | MR Egger | 16 | 0.140168066 | 0.047148138 | 0.010077823 | 64.7 | 30.1 | NA | NA | NA | -0.002983 | 0.001639 | 0.0902 |
| HbA1c | Creatinine | Weighted median | 16 | 0.091026007 | 0.027300699 | 0.000855449 | 64.7 | 30.1 | NA | NA | NA | NA | NA | NA |
| HbA1c | Creatinine | Weighted mode | 16 | 0.088020356 | 0.032918465 | 0.017342648 | 64.7 | 30.1 | NA | NA | NA | NA | NA | NA |
| HDL | Creatinine | Inverse variance weighted | 100 | -0.012036432 | 0.005779799 | 0.037296816 | 193.1 | 59.2 | 160.13 | 1.00E-04 | 38.2 | NA | NA | NA |
| HDL | Creatinine | MR Egger | 100 | -0.015721483 | 0.011442131 | 0.172576312 | 193.1 | 59.2 | NA | NA | NA | 0.00027 | 0.000722 | 0.7094 |
| HDL | Creatinine | Weighted median | 100 | -0.017703901 | 0.007590478 | 0.019680566 | 193.1 | 59.2 | NA | NA | NA | NA | NA | NA |
| HDL | Creatinine | Weighted mode | 100 | -0.015777776 | 0.008475193 | 0.065620125 | 193.1 | 59.2 | NA | NA | NA | NA | NA | NA |
| LDL | Creatinine | Inverse variance weighted | 99 | -0.018041737 | 0.005752469 | 0.001710668 | 208.9 | 56.4 | 200.74 | 0 | 51.2 | NA | NA | NA |
| LDL | Creatinine | MR Egger | 99 | -0.019759055 | 0.010086775 | 0.052995113 | 208.9 | 56.4 | NA | NA | NA | 0.00015 | 0.000723 | 0.8359 |
| LDL | Creatinine | Weighted median | 99 | -0.019750266 | 0.006525558 | 0.002473196 | 208.9 | 56.4 | NA | NA | NA | NA | NA | NA |

| | | | | | | | | | | | | | | |
|---|---|---|---|---|---|---|---|---|---|---|---|---|---|---|
| LDL | Creatinine | Weighted mode | 99 | -0.016208562 | 0.006794202 | 0.018971307 | 208.9 | 56.4 | NA | NA | NA | NA | NA | NA |
| MetS | Creatinine | Inverse variance weighted | 96 | -0.013280867 | 0.00624666 | 0.033497037 | 97.8 | 41 | 279.17 | 0 | 66 | NA | NA | NA |
| MetS | Creatinine | MR Egger | 96 | -0.000102977 | 0.014531143 | 0.994360764 | 97.8 | 41 | NA | NA | NA | -0.001224 | 0.001219 | 0.3178 |
| MetS | Creatinine | Weighted median | 96 | -0.010323052 | 0.006159927 | 0.093769577 | 97.8 | 41 | NA | NA | NA | NA | NA | NA |
| MetS | Creatinine | Weighted mode | 96 | -0.004380246 | 0.011056113 | 0.692858331 | 97.8 | 41 | NA | NA | NA | NA | NA | NA |
| SBP | Creatinine | Inverse variance weighted | 100 | -0.001661861 | 0.000963601 | 0.084592952 | 189.9 | 103.5 | 303.92 | 0 | 67.4 | NA | NA | NA |
| SBP | Creatinine | MR Egger | 100 | -0.003208526 | 0.003196812 | 0.318012148 | 189.9 | 103.5 | NA | NA | NA | 0.000781 | 0.001539 | 0.6129 |
| SBP | Creatinine | Weighted median | 100 | -0.000471991 | 0.00096363 | 0.624271927 | 189.9 | 103.5 | NA | NA | NA | NA | NA | NA |
| SBP | Creatinine | Weighted mode | 100 | -6.03E-05 | 0.002040744 | 0.976490674 | 189.9 | 103.5 | NA | NA | NA | NA | NA | NA |
| T2D | Creatinine | Inverse variance weighted | 100 | -0.001862121 | 0.004810263 | 0.698671748 | 89.6 | 38.5 | 204.37 | 0 | 51.6 | NA | NA | NA |
| T2D | Creatinine | MR Egger | 100 | 0.001253857 | 0.011523349 | 0.913575581 | 89.6 | 38.5 | NA | NA | NA | -0.000276 | 0.000927 | 0.7664 |

| | | | | | | | | | | | | | | |
|---|---|---|---|---|---|---|---|---|---|---|---|---|---|---|
| T2D | Creatinine | Weighted median | 100 | -0.010148409 | 0.006490225 | 0.117900898 | 89.6 | 38.5 | NA | NA | NA | NA | NA | NA |
| T2D | Creatinine | Weighted mode | 100 | -0.013932182 | 0.007022667 | 0.050037298 | 89.6 | 38.5 | NA | NA | NA | NA | NA | NA |
| TG | Creatinine | Inverse variance weighted | 99 | 0.001084408 | 0.010457061 | 0.917406622 | 150.5 | 43 | 364.98 | 0 | 73.1 | NA | NA | NA |
| TG | Creatinine | MR Egger | 99 | -0.020049084 | 0.019054442 | 0.295321949 | 150.5 | 43 | NA | NA | NA | 0.001364 | 0.001029 | 0.1884 |
| TG | Creatinine | Weighted median | 99 | -0.000973299 | 0.010898337 | 0.928837832 | 150.5 | 43 | NA | NA | NA | NA | NA | NA |
| TG | Creatinine | Weighted mode | 99 | -0.011249569 | 0.012519366 | 0.371082392 | 150.5 | 43 | NA | NA | NA | NA | NA | NA |
| BMI | CRP | Inverse variance weighted | 41 | -0.479013168 | 0.134896849 | 0.00038383 | 146.4 | 85.3 | 1249.42 | 0 | 96.8 | NA | NA | NA |
| BMI | CRP | MR Egger | 41 | -0.303789219 | 0.430534446 | 0.48462459 | 146.4 | 85.3 | NA | NA | NA | -0.00491 | 0.011443 | 0.6702 |
| BMI | CRP | Weighted median | 41 | -0.396925473 | 0.039772961 | 1.87E-23 | 146.4 | 85.3 | NA | NA | NA | NA | NA | NA |
| BMI | CRP | Weighted mode | 41 | -0.385223052 | 0.05508384 | 1.92E-08 | 146.4 | 85.3 | NA | NA | NA | NA | NA | NA |
| DBP | CRP | Inverse variance weighted | 43 | -0.009272398 | 0.004244345 | 0.028914675 | 196.1 | 114 | 141.75 | 0 | 70.4 | NA | NA | NA |

| | | | | | | | | | | | | | | |
|---|---|---|---|---|---|---|---|---|---|---|---|---|---|---|
| DBP | CRP | MR Egger | 43 | -0.006147805 | 0.016384056 | 0.70942295 | 196.1 | 114 | NA | NA | NA | -9.00E-04 | 0.004552 | 0.8443 |
| DBP | CRP | Weighted median | 43 | -0.004430006 | 0.003730513 | 0.235028071 | 196.1 | 114 | NA | NA | NA | NA | NA | NA |
| DBP | CRP | Weighted mode | 43 | -7.92E-06 | 0.006922735 | 0.999093035 | 196.1 | 114 | NA | NA | NA | NA | NA | NA |
| HbA1c | CRP | Inverse variance weighted | 15 | 0.053573958 | 0.058131649 | 0.356738785 | 66.7 | 30.1 | 36.85 | 8.00E-04 | 62 | NA | NA | NA |
| HbA1c | CRP | MR Egger | 15 | 0.111007178 | 0.149976805 | 0.472358061 | 66.7 | 30.1 | NA | NA | NA | -0.00209 | 0.005004 | 0.683 |
| HbA1c | CRP | Weighted median | 15 | -0.025244497 | 0.053458693 | 0.636766652 | 66.7 | 30.1 | NA | NA | NA | NA | NA | NA |
| HbA1c | CRP | Weighted mode | 15 | -0.028969106 | 0.06467057 | 0.661043869 | 66.7 | 30.1 | NA | NA | NA | NA | NA | NA |
| HDL | CRP | Inverse variance weighted | 86 | -0.028107852 | 0.037522576 | 0.453801913 | 202.7 | 59.2 | 1744.41 | 0 | 95.1 | NA | NA | NA |
| HDL | CRP | MR Egger | 86 | -0.073689937 | 0.076459753 | 0.337925534 | 202.7 | 59.2 | NA | NA | NA | 0.003178 | 0.00464 | 0.4953 |
| HDL | CRP | Weighted median | 86 | -0.004965581 | 0.015559302 | 0.749621058 | 202.7 | 59.2 | NA | NA | NA | NA | NA | NA |
| HDL | CRP | Weighted mode | 86 | -0.015375997 | 0.017944266 | 0.39392323 | 202.7 | 59.2 | NA | NA | NA | NA | NA | NA |
| LDL | CRP | Inverse variance weighted | 90 | 0.109196645 | 0.043164075 | 0.011412629 | 199.1 | 56.8 | 2946.52 | 0 | 97 | NA | NA | NA |

| | | | | | | | | | | | | | | |
|---|---|---|---|---|---|---|---|---|---|---|---|---|---|---|
| LDL | CRP | MR Egger | 90 | 0.167176756 | 0.080428673 | 0.040568492 | 199.1 | 56.8 | NA | NA | NA | -0.004576 | 0.005353 | 0.3949 |
| LDL | CRP | Weighted median | 90 | -0.013386645 | 0.017065012 | 0.432776229 | 199.1 | 56.8 | NA | NA | NA | NA | NA | NA |
| LDL | CRP | Weighted mode | 90 | -0.026530704 | 0.021098725 | 0.211878423 | 199.1 | 56.8 | NA | NA | NA | NA | NA | NA |
| MetS | CRP | Inverse variance weighted | 40 | 0.024207255 | 0.043257758 | 0.575748941 | 110.9 | 42.1 | 966.36 | 0 | 96 | NA | NA | NA |
| MetS | CRP | MR Egger | 40 | -0.022374415 | 0.111630595 | 0.842211115 | 110.9 | 42.1 | NA | NA | NA | 0.004085 | 0.009007 | 0.6528 |
| MetS | CRP | Weighted median | 40 | 0.005834856 | 0.014240112 | 0.681990922 | 110.9 | 42.1 | NA | NA | NA | NA | NA | NA |
| MetS | CRP | Weighted mode | 40 | 0.007712308 | 0.016835584 | 0.649426045 | 110.9 | 42.1 | NA | NA | NA | NA | NA | NA |
| SBP | CRP | Inverse variance weighted | 43 | -0.003710222 | 0.002496407 | 0.137219651 | 188.3 | 103.5 | 145.59 | 0 | 71.2 | NA | NA | NA |
| SBP | CRP | MR Egger | 43 | 0.012607049 | 0.007606554 | 0.10507362 | 188.3 | 103.5 | NA | NA | NA | -0.008134 | 0.003601 | 0.0293 |
| SBP | CRP | Weighted median | 43 | -0.001491963 | 0.002123756 | 0.482360112 | 188.3 | 103.5 | NA | NA | NA | NA | NA | NA |
| SBP | CRP | Weighted mode | 43 | -0.001481352 | 0.003580064 | 0.681142104 | 188.3 | 103.5 | NA | NA | NA | NA | NA | NA |
| T2D | CRP | Inverse variance weighted | 80 | -0.036401294 | 0.012349417 | 0.003202382 | 93.8 | 38.6 | 349.12 | 0 | 77.4 | NA | NA | NA |

| | | | | | | | | | | | | | | |
|---|---|---|---|---|---|---|---|---|---|---|---|---|---|---|
| T2D | CRP | MR Egger | 80 | 0.010719194 | 0.027317152 | 0.695834442 | 93.8 | 38.6 | NA | NA | NA | -0.004301 | 0.002234 | 0.0578 |
| T2D | CRP | Weighted median | 80 | -0.005555439 | 0.010400514 | 0.593237475 | 93.8 | 38.6 | NA | NA | NA | NA | NA | NA |
| T2D | CRP | Weighted mode | 80 | -0.008456534 | 0.012107412 | 0.48694104 | 93.8 | 38.6 | NA | NA | NA | NA | NA | NA |
| TG | CRP | Inverse variance weighted | 88 | -0.115898601 | 0.051030185 | 0.023136243 | 148 | 43 | 2503.81 | 0 | 96.5 | NA | NA | NA |
| TG | CRP | MR Egger | 88 | -0.203394019 | 0.099225369 | 0.043428293 | 148 | 43 | NA | NA | NA | 0.005282 | 0.005138 | 0.3068 |
| TG | CRP | Weighted median | 88 | -0.076008995 | 0.019786748 | 0.00012233 | 148 | 43 | NA | NA | NA | NA | NA | NA |
| TG | CRP | Weighted mode | 88 | -0.066130533 | 0.01956986 | 0.001089753 | 148 | 43 | NA | NA | NA | NA | NA | NA |
| BMI | Cystatin C | Inverse variance weighted | 42 | 0.068469304 | 0.014633519 | 2.88E-06 | 142.8 | 85.3 | 56.98 | 0.0497 | 28 | NA | NA | NA |
| BMI | Cystatin C | MR Egger | 42 | 0.059011034 | 0.049685665 | 0.241961525 | 142.8 | 85.3 | NA | NA | NA | 0.000256 | 0.001281 | 0.8429 |
| BMI | Cystatin C | Weighted median | 42 | 0.04696998 | 0.019368749 | 0.015306722 | 142.8 | 85.3 | NA | NA | NA | NA | NA | NA |
| BMI | Cystatin C | Weighted mode | 42 | 0.045938127 | 0.030979322 | 0.145757094 | 142.8 | 85.3 | NA | NA | NA | NA | NA | NA |
| DBP | Cystatin C | Inverse variance weighted | 42 | 0.000355974 | 0.001820352 | 0.844960552 | 196.7 | 114 | 96.41 | 0 | 57.5 | NA | NA | NA |

| | | | | | | | | | | | | | | |
|---|---|---|---|---|---|---|---|---|---|---|---|---|---|---|
| DBP | Cystatin C | MR Egger | 42 | -0.001696922 | 0.006909953 | 0.807265786 | 196.7 | 114 | NA | NA | NA | 0.000594 | 0.001926 | 0.7595 |
| DBP | Cystatin C | Weighted median | 42 | 0.000815087 | 0.001868235 | 0.662628363 | 196.7 | 114 | NA | NA | NA | NA | NA | NA |
| DBP | Cystatin C | Weighted mode | 42 | 0.001984192 | 0.003616077 | 0.586177641 | 196.7 | 114 | NA | NA | NA | NA | NA | NA |
| HbA1c | Cystatin C | Inverse variance weighted | 14 | 0.011471088 | 0.019272374 | 0.551703865 | 69.1 | 30.1 | 11.48 | 0.5707 | 0 | NA | NA | NA |
| HbA1c | Cystatin C | MR Egger | 14 | -0.003627189 | 0.051528099 | 0.9450408 | 69.1 | 30.1 | NA | NA | NA | 0.000525 | 0.001662 | 0.7575 |
| HbA1c | Cystatin C | Weighted median | 14 | 0.006511815 | 0.025995179 | 0.80220003 | 69.1 | 30.1 | NA | NA | NA | NA | NA | NA |
| HbA1c | Cystatin C | Weighted mode | 14 | -0.005233039 | 0.036181311 | 0.887218276 | 69.1 | 30.1 | NA | NA | NA | NA | NA | NA |
| HDL | Cystatin C | Inverse variance weighted | 86 | -0.001952365 | 0.005003836 | 0.696408098 | 188.5 | 59.2 | 108.46 | 0.044 | 21.6 | NA | NA | NA |
| HDL | Cystatin C | MR Egger | 86 | -0.001985192 | 0.011205716 | 0.85981009 | 188.5 | 59.2 | NA | NA | NA | 2.00E-06 | 0.000627 | 0.9974 |
| HDL | Cystatin C | Weighted median | 86 | 0.001166328 | 0.00683113 | 0.864430357 | 188.5 | 59.2 | NA | NA | NA | NA | NA | NA |
| HDL | Cystatin C | Weighted mode | 86 | 0.002785666 | 0.009322428 | 0.765811816 | 188.5 | 59.2 | NA | NA | NA | NA | NA | NA |
| LDL | Cystatin C | Inverse variance weighted | 90 | 0.004671555 | 0.004666741 | 0.316811492 | 194 | 56.4 | 120.71 | 0.0142 | 26.3 | NA | NA | NA |
| LDL | Cystatin C | MR Egger | 90 | 0.003193478 | 0.009637467 | 0.741159873 | 194 | 56.4 | NA | NA | NA | 0.000106 | 0.000603 | 0.861 |

| | | | | | | | | | | | | | | |
|---|---|---|---|---|---|---|---|---|---|---|---|---|---|---|
| LDL | Cystatin C | Weighted median | 90 | 0.010939527 | 0.006501816 | 0.092465335 | 194 | 56.4 | NA | NA | NA | NA | NA | NA |
| LDL | Cystatin C | Weighted mode | 90 | 0.013730151 | 0.009608948 | 0.156535624 | 194 | 56.4 | NA | NA | NA | NA | NA | NA |
| MetS | Cystatin C | Inverse variance weighted | 38 | -0.004078749 | 0.004925074 | 0.407579924 | 112.1 | 42.1 | 43.03 | 0.229 | 14 | NA | NA | NA |
| MetS | Cystatin C | MR Egger | 38 | -0.008000272 | 0.012549309 | 0.527828715 | 112.1 | 42.1 | NA | NA | NA | 0.000341 | 0.001003 | 0.7355 |
| MetS | Cystatin C | Weighted median | 38 | 8.72E-05 | 0.007129812 | 0.990237787 | 112.1 | 42.1 | NA | NA | NA | NA | NA | NA |
| MetS | Cystatin C | Weighted mode | 38 | 0.00452218 | 0.011000093 | 0.683367862 | 112.1 | 42.1 | NA | NA | NA | NA | NA | NA |
| SBP | Cystatin C | Inverse variance weighted | 43 | 0.000856907 | 0.000863318 | 0.320917525 | 188.3 | 103.5 | 65.29 | 0.0122 | 35.7 | NA | NA | NA |
| SBP | Cystatin C | MR Egger | 43 | -0.005605731 | 0.002581356 | 0.035723198 | 188.3 | 103.5 | NA | NA | NA | 0.003203 | 0.001215 | 0.0118 |
| SBP | Cystatin C | Weighted median | 43 | 0.000567965 | 0.001019142 | 0.577324384 | 188.3 | 103.5 | NA | NA | NA | NA | NA | NA |
| SBP | Cystatin C | Weighted mode | 43 | -0.000222975 | 0.001923479 | 0.908266177 | 188.3 | 103.5 | NA | NA | NA | NA | NA | NA |
| T2D | Cystatin C | Inverse variance weighted | 83 | 0.00016823 | 0.003691904 | 0.963655051 | 95.9 | 38.6 | 133.24 | 3.00E-04 | 38.5 | NA | NA | NA |
| T2D | Cystatin C | MR Egger | 83 | 0.008101112 | 0.008541343 | 0.345717144 | 95.9 | 38.6 | NA | NA | NA | -0.000711 | 0.00069 | 0.3061 |
| T2D | Cystatin C | Weighted median | 83 | 0.000650131 | 0.005400929 | 0.904186893 | 95.9 | 38.6 | NA | NA | NA | NA | NA | NA |

| | | | | | | | | | | | | | | |
|---|---|---|---|---|---|---|---|---|---|---|---|---|---|---|
| T2D | Cystatin C | Weighted mode | 83 | -0.001040338 | 0.005284355 | 0.844415314 | 95.9 | 38.6 | NA | NA | NA | NA | NA | NA |
| TG | Cystatin C | Inverse variance weighted | 85 | -0.003201865 | 0.006346848 | 0.613923282 | 160.5 | 43 | 145.17 | 0 | 42.1 | NA | NA | NA |
| TG | Cystatin C | MR Egger | 85 | -0.006971453 | 0.011552261 | 0.547841182 | 160.5 | 43 | NA | NA | NA | 0.000235 | 6.00E-04 | 0.6965 |
| TG | Cystatin C | Weighted median | 85 | 0.002303027 | 0.008781122 | 0.793113083 | 160.5 | 43 | NA | NA | NA | NA | NA | NA |
| TG | Cystatin C | Weighted mode | 85 | 0.011508361 | 0.012747417 | 0.369213937 | 160.5 | 43 | NA | NA | NA | NA | NA | NA |
| BMI | Fresh Fruit Consumption | Inverse variance weighted | 91 | -0.209187535 | 0.040346062 | 2.16E-07 | 154 | 85.3 | 379.34 | 0 | 76.3 | NA | NA | NA |
| BMI | Fresh Fruit Consumption | MR Egger | 91 | -0.435638315 | 0.105960364 | 8.74E-05 | 154 | 85.3 | NA | NA | NA | 0.006927 | 0.003008 | 0.0236 |
| BMI | Fresh Fruit Consumption | Weighted median | 91 | -0.202908914 | 0.038062009 | 9.77E-08 | 154 | 85.3 | NA | NA | NA | NA | NA | NA |
| BMI | Fresh Fruit Consumption | Weighted mode | 91 | -0.198443059 | 0.070721064 | 0.006147543 | 154 | 85.3 | NA | NA | NA | NA | NA | NA |
| DBP | Fresh Fruit Consumption | Inverse variance weighted | 100 | 0.000368628 | 0.002484916 | 0.882069447 | 209.5 | 114 | 185.47 | 0 | 46.6 | NA | NA | NA |
| DBP | Fresh Fruit Consumption | MR Egger | 100 | -0.006190531 | 0.008556202 | 0.471087654 | 209.5 | 114 | NA | NA | NA | 0.002029 | 0.002533 | 0.4249 |
| DBP | Fresh Fruit Consumption | Weighted median | 100 | 0.000238816 | 0.002861226 | 0.933480762 | 209.5 | 114 | NA | NA | NA | NA | NA | NA |

| | | | | | | | | | | | | | | |
|---|---|---|---|---|---|---|---|---|---|---|---|---|---|---|
| DBP | Fresh Fruit Consumption | Weighted mode | 100 | 0.000293943 | 0.005673014 | 0.958781113 | 209.5 | 114 | NA | NA | NA | NA | NA | NA |
| HbA1c | Fresh Fruit Consumption | Inverse variance weighted | 16 | 0.018551886 | 0.052635593 | 0.724494092 | 64.7 | 30.1 | 26.19 | 0.0361 | 42.7 | NA | NA | NA |
| HbA1c | Fresh Fruit Consumption | MR Egger | 16 | 0.042101027 | 0.129016277 | 0.749006683 | 64.7 | 30.1 | NA | NA | NA | -0.000902 | 0.004482 | 0.8434 |
| HbA1c | Fresh Fruit Consumption | Weighted median | 16 | 0.016908397 | 0.061860174 | 0.784597625 | 64.7 | 30.1 | NA | NA | NA | NA | NA | NA |
| HbA1c | Fresh Fruit Consumption | Weighted mode | 16 | 0.011869905 | 0.067701647 | 0.863167751 | 64.7 | 30.1 | NA | NA | NA | NA | NA | NA |
| HDL | Fresh Fruit Consumption | Inverse variance weighted | 92 | 0.007678107 | 0.010606163 | 0.469109481 | 201.7 | 59.2 | 118.99 | 0.0261 | 23.5 | NA | NA | NA |
| HDL | Fresh Fruit Consumption | MR Egger | 92 | -0.012309238 | 0.020725756 | 0.554062051 | 201.7 | 59.2 | NA | NA | NA | 0.00151 | 0.001346 | 0.2649 |
| HDL | Fresh Fruit Consumption | Weighted median | 92 | 0.00404241 | 0.014567908 | 0.781405912 | 201.7 | 59.2 | NA | NA | NA | NA | NA | NA |
| HDL | Fresh Fruit Consumption | Weighted mode | 92 | 0.004532095 | 0.016776591 | 0.787661191 | 201.7 | 59.2 | NA | NA | NA | NA | NA | NA |
| LDL | Fresh Fruit Consumption | Inverse variance weighted | 93 | -0.044750225 | 0.010688925 | 2.83E-05 | 203 | 56.8 | 149.11 | 2.00E-04 | 38.3 | NA | NA | NA |
| LDL | Fresh Fruit Consumption | MR Egger | 93 | -0.048474295 | 0.018624053 | 0.010796158 | 203 | 56.8 | NA | NA | NA | 0.000325 | 0.001328 | 0.8072 |
| LDL | Fresh Fruit Consumption | Weighted median | 93 | -0.027464634 | 0.013207115 | 0.037568372 | 203 | 56.8 | NA | NA | NA | NA | NA | NA |
| LDL | Fresh Fruit Consumption | Weighted mode | 93 | -0.033778149 | 0.016549784 | 0.044115518 | 203 | 56.8 | NA | NA | NA | NA | NA | NA |

| | | | | | | | | | | | | | | |
|---|---|---|---|---|---|---|---|---|---|---|---|---|---|---|
| MetS | Fresh Fruit Consumption | Inverse variance weighted | 91 | 0.014453212 | 0.011650872 | 0.214780815 | 100.4 | 41 | 223.88 | 0 | 59.8 | NA | NA | NA |
| MetS | Fresh Fruit Consumption | MR Egger | 91 | -0.01527571 | 0.026712616 | 0.568862598 | 100.4 | 41 | NA | NA | NA | 0.002798 | 0.002264 | 0.2197 |
| MetS | Fresh Fruit Consumption | Weighted median | 91 | -0.007096173 | 0.011430873 | 0.534737739 | 100.4 | 41 | NA | NA | NA | NA | NA | NA |
| MetS | Fresh Fruit Consumption | Weighted mode | 91 | -0.019271745 | 0.020769126 | 0.355939466 | 100.4 | 41 | NA | NA | NA | NA | NA | NA |
| SBP | Fresh Fruit Consumption | Inverse variance weighted | 100 | -0.00216331 | 0.001661949 | 0.19302889 | 189.9 | 103.5 | 226.51 | 0 | 56.3 | NA | NA | NA |
| SBP | Fresh Fruit Consumption | MR Egger | 100 | 0.003922457 | 0.005480544 | 0.475874932 | 189.9 | 103.5 | NA | NA | NA | -0.003074 | 0.002638 | 0.2468 |
| SBP | Fresh Fruit Consumption | Weighted median | 100 | -0.004077349 | 0.001693276 | 0.016041768 | 189.9 | 103.5 | NA | NA | NA | NA | NA | NA |
| SBP | Fresh Fruit Consumption | Weighted mode | 100 | -0.00520334 | 0.003341352 | 0.122600849 | 189.9 | 103.5 | NA | NA | NA | NA | NA | NA |
| T2D | Fresh Fruit Consumption | Inverse variance weighted | 91 | -0.015494276 | 0.011345142 | 0.172027166 | 88.4 | 38.5 | 232.04 | 0 | 61.2 | NA | NA | NA |
| T2D | Fresh Fruit Consumption | MR Egger | 91 | 0.003200949 | 0.026222521 | 0.903119807 | 88.4 | 38.5 | NA | NA | NA | -0.001674 | 0.002116 | 0.4309 |
| T2D | Fresh Fruit Consumption | Weighted median | 91 | -0.002019493 | 0.013556335 | 0.881576952 | 88.4 | 38.5 | NA | NA | NA | NA | NA | NA |
| T2D | Fresh Fruit Consumption | Weighted mode | 91 | -0.002884128 | 0.013824755 | 0.835215886 | 88.4 | 38.5 | NA | NA | NA | NA | NA | NA |

| | | | | | | | | | | | | | | |
|---|---|---|---|---|---|---|---|---|---|---|---|---|---|---|
| TG | Fresh Fruit Consumption | Inverse variance weighted | 93 | 0.027090931 | 0.01620141 | 0.094497871 | 143.3 | 43 | 180.67 | 0 | 49.1 | NA | NA | NA |
| TG | Fresh Fruit Consumption | MR Egger | 93 | 0.025076441 | 0.031814881 | 0.432629061 | 143.3 | 43 | NA | NA | NA | 0.000122 | 0.001654 | 0.9414 |
| TG | Fresh Fruit Consumption | Weighted median | 93 | 0.015255802 | 0.019716617 | 0.439076552 | 143.3 | 43 | NA | NA | NA | NA | NA | NA |
| TG | Fresh Fruit Consumption | Weighted mode | 93 | 0.013556918 | 0.02708318 | 0.617872031 | 143.3 | 43 | NA | NA | NA | NA | NA | NA |
| BMI | Gamma Glutamyl Transferase | Inverse variance weighted | 75 | -0.066955756 | 0.025580918 | 0.008859978 | 156.7 | 85.3 | 107.33 | 0.0069 | 31.1 | NA | NA | NA |
| BMI | Gamma Glutamyl Transferase | MR Egger | 75 | 0.137214556 | 0.069547718 | 0.052287441 | 156.7 | 85.3 | NA | NA | NA | -0.006124 | 0.001956 | 0.0025 |
| BMI | Gamma Glutamyl Transferase | Weighted median | 75 | -0.049508475 | 0.035977482 | 0.168791865 | 156.7 | 85.3 | NA | NA | NA | NA | NA | NA |
| BMI | Gamma Glutamyl Transferase | Weighted mode | 75 | 0.036311954 | 0.053247868 | 0.497404609 | 156.7 | 85.3 | NA | NA | NA | NA | NA | NA |
| DBP | Gamma Glutamyl Transferase | Inverse variance weighted | 79 | -0.001560613 | 0.003378411 | 0.644126524 | 197 | 114 | 248.31 | 0 | 68.6 | NA | NA | NA |
| DBP | Gamma Glutamyl Transferase | MR Egger | 79 | 0.002083989 | 0.012016669 | 0.8627726 | 197 | 114 | NA | NA | NA | -0.00112 | 0.003543 | 0.7527 |
| DBP | Gamma Glutamyl Transferase | Weighted median | 79 | 0.001839054 | 0.00292707 | 0.529812803 | 197 | 114 | NA | NA | NA | NA | NA | NA |
| DBP | Gamma Glutamyl Transferase | Weighted mode | 79 | 0.002199062 | 0.004361808 | 0.61556805 | 197 | 114 | NA | NA | NA | NA | NA | NA |
| HbA1c | Gamma Glutamyl Transferase | Inverse variance weighted | 10 | -0.02861621 | 0.054160984 | 0.597253184 | 54.8 | 30.1 | 7.88 | 0.5468 | 0 | NA | NA | NA |

| | | | | | | | | | | | | | | |
|---|---|---|---|---|---|---|---|---|---|---|---|---|---|---|
| HbA1c | Gamma Glutamyl Transferase | MR Egger | 10 | -0.206272084 | 0.196485429 | 0.324482268 | 54.8 | 30.1 | NA | NA | NA | 0.00523 | 0.00556 | 0.3744 |
| HbA1c | Gamma Glutamyl Transferase | Weighted median | 10 | -0.022374556 | 0.069285073 | 0.746744678 | 54.8 | 30.1 | NA | NA | NA | NA | NA | NA |
| HbA1c | Gamma Glutamyl Transferase | Weighted mode | 10 | -0.017609988 | 0.092181741 | 0.85273778 | 54.8 | 30.1 | NA | NA | NA | NA | NA | NA |
| HDL | Gamma Glutamyl Transferase | Inverse variance weighted | 87 | -0.018501858 | 0.016354864 | 0.257939125 | 173.5 | 59.2 | 289.44 | 0 | 70.3 | NA | NA | NA |
| HDL | Gamma Glutamyl Transferase | MR Egger | 87 | -0.026445173 | 0.035478503 | 0.458094743 | 173.5 | 59.2 | NA | NA | NA | 0.000549 | 0.002173 | 0.8011 |
| HDL | Gamma Glutamyl Transferase | Weighted median | 87 | -0.006231789 | 0.016097807 | 0.698667107 | 173.5 | 59.2 | NA | NA | NA | NA | NA | NA |
| HDL | Gamma Glutamyl Transferase | Weighted mode | 87 | -0.005805403 | 0.022161706 | 0.793981399 | 173.5 | 59.2 | NA | NA | NA | NA | NA | NA |
| LDL | Gamma Glutamyl Transferase | Inverse variance weighted | 77 | 0.0054958 | 0.025211522 | 0.827438746 | 209.8 | 56.8 | 728.77 | 0 | 89.6 | NA | NA | NA |
| LDL | Gamma Glutamyl Transferase | MR Egger | 77 | 0.004959568 | 0.043234609 | 0.908979234 | 209.8 | 56.8 | NA | NA | NA | 4.80E-05 | 0.003119 | 0.9878 |
| LDL | Gamma Glutamyl Transferase | Weighted median | 77 | 0.012478256 | 0.014149098 | 0.377824527 | 209.8 | 56.8 | NA | NA | NA | NA | NA | NA |
| LDL | Gamma Glutamyl Transferase | Weighted mode | 77 | 0.0137815 | 0.013173443 | 0.298804119 | 209.8 | 56.8 | NA | NA | NA | NA | NA | NA |
| MetS | Gamma Glutamyl Transferase | Inverse variance weighted | 66 | 0.021308511 | 0.019756933 | 0.280795823 | 98.6 | 41 | 442.04 | 0 | 85.3 | NA | NA | NA |

| | | | | | | | | | | | | | | |
|---|---|---|---|---|---|---|---|---|---|---|---|---|---|---|
| MetS | Gamma Glutamyl Transferase | MR Egger | 66 | 0.011164708 | 0.047835563 | 0.816197546 | 98.6 | 41 | NA | NA | NA | 0.00093 | 0.003986 | 0.8164 |
| MetS | Gamma Glutamyl Transferase | Weighted median | 66 | 0.019173194 | 0.012567256 | 0.127097853 | 98.6 | 41 | NA | NA | NA | NA | NA | NA |
| MetS | Gamma Glutamyl Transferase | Weighted mode | 66 | 0.01763144 | 0.013880021 | 0.208515025 | 98.6 | 41 | NA | NA | NA | NA | NA | NA |
| SBP | Gamma Glutamyl Transferase | Inverse variance weighted | 85 | -0.000638363 | 0.001530457 | 0.67660092 | 182.5 | 103.5 | 178.85 | 0 | 53 | NA | NA | NA |
| SBP | Gamma Glutamyl Transferase | MR Egger | 85 | 0.000167435 | 0.004549946 | 0.970733361 | 182.5 | 103.5 | NA | NA | NA | -0.000427 | 0.00227 | 0.8512 |
| SBP | Gamma Glutamyl Transferase | Weighted median | 85 | 0.002040505 | 0.001686153 | 0.226219729 | 182.5 | 103.5 | NA | NA | NA | NA | NA | NA |
| SBP | Gamma Glutamyl Transferase | Weighted mode | 85 | 0.001805122 | 0.002530159 | 0.477549627 | 182.5 | 103.5 | NA | NA | NA | NA | NA | NA |
| T2D | Gamma Glutamyl Transferase | Inverse variance weighted | 86 | 0.038290055 | 0.01584684 | 0.015680944 | 92.5 | 38.6 | 404.24 | 0 | 79 | NA | NA | NA |
| T2D | Gamma Glutamyl Transferase | MR Egger | 86 | 0.043603444 | 0.045767465 | 0.343466935 | 92.5 | 38.6 | NA | NA | NA | -0.000423 | 0.003417 | 0.9017 |
| T2D | Gamma Glutamyl Transferase | Weighted median | 86 | 0.032686257 | 0.012647028 | 0.009752004 | 92.5 | 38.6 | NA | NA | NA | NA | NA | NA |
| T2D | Gamma Glutamyl Transferase | Weighted mode | 86 | 0.039157719 | 0.023796135 | 0.103549647 | 92.5 | 38.6 | NA | NA | NA | NA | NA | NA |
| TG | Gamma Glutamyl Transferase | Inverse variance weighted | 76 | -0.099560322 | 0.026653739 | 0.000187475 | 159.8 | 43 | 512.86 | 0 | 85.4 | NA | NA | NA |
| TG | Gamma Glutamyl Transferase | MR Egger | 76 | -0.109168336 | 0.046113306 | 0.020529479 | 159.8 | 43 | NA | NA | NA | 0.000678 | 0.002648 | 0.7986 |

| | | | | | | | | | | | | | | |
|---|---|---|---|---|---|---|---|---|---|---|---|---|---|---|
| TG | Gamma Glutamyl Transferase | Weighted median | 76 | -0.02147355 | 0.018224034 | 0.238673312 | 159.8 | 43 | NA | NA | NA | NA | NA | NA |
| TG | Gamma Glutamyl Transferase | Weighted mode | 76 | -0.01886829 | 0.015843394 | 0.237438638 | 159.8 | 43 | NA | NA | NA | NA | NA | NA |
| BMI | Impedance of Arm (Left) | Inverse variance weighted | 99 | -0.433750069 | 0.018691934 | 4.03E-119 | 150.8 | 85.3 | 679.27 | 0 | 85.6 | NA | NA | NA |
| BMI | Impedance of Arm (Left) | MR Egger | 99 | -0.504779717 | 0.051046393 | 2.33E-16 | 150.8 | 85.3 | NA | NA | NA | 0.002128 | 0.001424 | 0.1384 |
| BMI | Impedance of Arm (Left) | Weighted median | 99 | -0.432370774 | 0.015370393 | 4.20E-174 | 150.8 | 85.3 | NA | NA | NA | NA | NA | NA |
| BMI | Impedance of Arm (Left) | Weighted mode | 99 | -0.458160366 | 0.023980825 | 8.16E-35 | 150.8 | 85.3 | NA | NA | NA | NA | NA | NA |
| DBP | Impedance of Arm (Left) | Inverse variance weighted | 100 | 0.006404265 | 0.002260063 | 0.004601729 | 209.5 | 114 | 1101.78 | 0 | 91 | NA | NA | NA |
| DBP | Impedance of Arm (Left) | MR Egger | 100 | 0.015084773 | 0.00775337 | 0.054570801 | 209.5 | 114 | NA | NA | NA | -0.002686 | 0.002295 | 0.2448 |
| DBP | Impedance of Arm (Left) | Weighted median | 100 | 0.005810998 | 0.001409966 | 3.77E-05 | 209.5 | 114 | NA | NA | NA | NA | NA | NA |
| DBP | Impedance of Arm (Left) | Weighted mode | 100 | 0.005297217 | 0.002639993 | 0.047527314 | 209.5 | 114 | NA | NA | NA | NA | NA | NA |
| HbA1c | Impedance of Arm (Left) | Inverse variance weighted | 16 | -0.075681354 | 0.029840978 | 0.011207704 | 64.7 | 30.1 | 60.55 | 0 | 75.2 | NA | NA | NA |
| HbA1c | Impedance of Arm (Left) | MR Egger | 16 | -0.173200108 | 0.067231039 | 0.021971354 | 64.7 | 30.1 | NA | NA | NA | 0.00374 | 0.002337 | 0.1319 |

| | | | | | | | | | | | | | | |
|---|---|---|---|---|---|---|---|---|---|---|---|---|---|---|
| HbA1c | Impedance of Arm (Left) | Weighted median | 16 | -0.090875117 | 0.023936325 | 0.000146732 | 64.7 | 30.1 | NA | NA | NA | NA | NA | NA |
| HbA1c | Impedance of Arm (Left) | Weighted mode | 16 | -0.092759101 | 0.025677285 | 0.002559234 | 64.7 | 30.1 | NA | NA | NA | NA | NA | NA |
| HDL | Impedance of Arm (Left) | Inverse variance weighted | 100 | 0.015093721 | 0.008753516 | 0.084652996 | 193.1 | 59.2 | 660.66 | 0 | 85 | NA | NA | NA |
| HDL | Impedance of Arm (Left) | MR Egger | 100 | -0.000442326 | 0.017247173 | 0.979591631 | 193.1 | 59.2 | NA | NA | NA | 0.001137 | 0.001088 | 0.2985 |
| HDL | Impedance of Arm (Left) | Weighted median | 100 | 0.010970845 | 0.005878292 | 0.061994915 | 193.1 | 59.2 | NA | NA | NA | NA | NA | NA |
| HDL | Impedance of Arm (Left) | Weighted mode | 100 | 0.0138742 | 0.006587196 | 0.037712701 | 193.1 | 59.2 | NA | NA | NA | NA | NA | NA |
| LDL | Impedance of Arm (Left) | Inverse variance weighted | 99 | 0.017004145 | 0.005804139 | 0.003393345 | 208.9 | 56.4 | 367.69 | 0 | 73.3 | NA | NA | NA |
| LDL | Impedance of Arm (Left) | MR Egger | 99 | 0.018807482 | 0.010175701 | 0.067609386 | 208.9 | 56.4 | NA | NA | NA | -0.000158 | 0.00073 | 0.8292 |
| LDL | Impedance of Arm (Left) | Weighted median | 99 | 0.014738717 | 0.005034299 | 0.003415232 | 208.9 | 56.4 | NA | NA | NA | NA | NA | NA |
| LDL | Impedance of Arm (Left) | Weighted mode | 99 | 0.015071403 | 0.005262159 | 0.005115878 | 208.9 | 56.4 | NA | NA | NA | NA | NA | NA |
| MetS | Impedance of Arm (Left) | Inverse variance weighted | 96 | 0.049504176 | 0.012033807 | 3.89E-05 | 97.8 | 41 | 1863.74 | 0 | 94.9 | NA | NA | NA |
| MetS | Impedance of Arm (Left) | MR Egger | 96 | -0.027996314 | 0.026718837 | 0.297412021 | 97.8 | 41 | NA | NA | NA | 0.007199 | 0.002241 | 0.0018 |
| MetS | Impedance of Arm (Left) | Weighted median | 96 | 0.018719334 | 0.004842613 | 0.000110842 | 97.8 | 41 | NA | NA | NA | NA | NA | NA |

| | | | | | | | | | | | | | | |
|---|---|---|---|---|---|---|---|---|---|---|---|---|---|---|
| MetS | Impedance of Arm (Left) | Weighted mode | 96 | 0.017649497 | 0.006122081 | 0.004872772 | 97.8 | 41 | NA | NA | NA | NA | NA | NA |
| SBP | Impedance of Arm (Left) | Inverse variance weighted | 100 | 0.002023139 | 0.001499375 | 0.177233723 | 189.9 | 103.5 | 1323.51 | 0 | 92.5 | NA | NA | NA |
| SBP | Impedance of Arm (Left) | MR Egger | 100 | 0.007319906 | 0.004949433 | 0.142363986 | 189.9 | 103.5 | NA | NA | NA | -0.002675 | 0.002382 | 0.2643 |
| SBP | Impedance of Arm (Left) | Weighted median | 100 | 0.00197435 | 0.000877187 | 0.024399808 | 189.9 | 103.5 | NA | NA | NA | NA | NA | NA |
| SBP | Impedance of Arm (Left) | Weighted mode | 100 | 0.002075668 | 0.001395729 | 0.140151479 | 189.9 | 103.5 | NA | NA | NA | NA | NA | NA |
| T2D | Impedance of Arm (Left) | Inverse variance weighted | 100 | -0.029164463 | 0.011969546 | 0.014827897 | 89.6 | 38.5 | 2276.93 | 0 | 95.7 | NA | NA | NA |
| T2D | Impedance of Arm (Left) | MR Egger | 100 | 0.033077574 | 0.027835686 | 0.237580971 | 89.6 | 38.5 | NA | NA | NA | -0.005514 | 0.002239 | 0.0155 |
| T2D | Impedance of Arm (Left) | Weighted median | 100 | 0.018982141 | 0.004889681 | 0.000103566 | 89.6 | 38.5 | NA | NA | NA | NA | NA | NA |
| T2D | Impedance of Arm (Left) | Weighted mode | 100 | 0.019134865 | 0.004886766 | 0.000165774 | 89.6 | 38.5 | NA | NA | NA | NA | NA | NA |
| TG | Impedance of Arm (Left) | Inverse variance weighted | 99 | 0.010902521 | 0.015175075 | 0.472480341 | 150.5 | 43 | 1382.82 | 0 | 92.9 | NA | NA | NA |
| TG | Impedance of Arm (Left) | MR Egger | 99 | 0.078178593 | 0.026680535 | 0.004223908 | 150.5 | 43 | NA | NA | NA | -0.004341 | 0.001441 | 0.0033 |
| TG | Impedance of Arm (Left) | Weighted median | 99 | -0.003070263 | 0.008062212 | 0.703335754 | 150.5 | 43 | NA | NA | NA | NA | NA | NA |
| TG | Impedance of Arm (Left) | Weighted mode | 99 | -0.010590584 | 0.006891272 | 0.127562027 | 150.5 | 43 | NA | NA | NA | NA | NA | NA |

| | | | | | | | | | | | | | | |
|---|---|---|---|---|---|---|---|---|---|---|---|---|---|---|
| BMI | Impedance of Arm (Right) | Inverse variance weighted | 99 | -0.425164351 | 0.018173197 | 4.79E-121 | 150.8 | 85.3 | 647.55 | 0 | 84.9 | NA | NA | NA |
| BMI | Impedance of Arm (Right) | MR Egger | 99 | -0.482587195 | 0.049805763 | 6.28E-16 | 150.8 | 85.3 | NA | NA | NA | 0.00172 | 0.00139 | 0.2188 |
| BMI | Impedance of Arm (Right) | Weighted median | 99 | -0.415905446 | 0.015400845 | 1.28E-160 | 150.8 | 85.3 | NA | NA | NA | NA | NA | NA |
| BMI | Impedance of Arm (Right) | Weighted mode | 99 | -0.438299481 | 0.02724721 | 3.00E-29 | 150.8 | 85.3 | NA | NA | NA | NA | NA | NA |
| DBP | Impedance of Arm (Right) | Inverse variance weighted | 100 | 0.006745615 | 0.002233753 | 0.002528943 | 209.5 | 114 | 1085.43 | 0 | 90.9 | NA | NA | NA |
| DBP | Impedance of Arm (Right) | MR Egger | 100 | 0.014772311 | 0.007669787 | 0.056997406 | 209.5 | 114 | NA | NA | NA | -0.002484 | 0.00227 | 0.2767 |
| DBP | Impedance of Arm (Right) | Weighted median | 100 | 0.005987438 | 0.001290469 | 3.49E-06 | 209.5 | 114 | NA | NA | NA | NA | NA | NA |
| DBP | Impedance of Arm (Right) | Weighted mode | 100 | 0.006471727 | 0.002518782 | 0.011679171 | 209.5 | 114 | NA | NA | NA | NA | NA | NA |
| HbA1c | Impedance of Arm (Right) | Inverse variance weighted | 16 | -0.072485813 | 0.027187329 | 0.00767228 | 64.7 | 30.1 | 50.69 | 0 | 70.4 | NA | NA | NA |
| HbA1c | Impedance of Arm (Right) | MR Egger | 16 | -0.148467032 | 0.062739478 | 0.032917035 | 64.7 | 30.1 | NA | NA | NA | 0.002914 | 0.002181 | 0.2028 |
| HbA1c | Impedance of Arm (Right) | Weighted median | 16 | -0.09375161 | 0.02309848 | 4.93E-05 | 64.7 | 30.1 | NA | NA | NA | NA | NA | NA |
| HbA1c | Impedance of Arm (Right) | Weighted mode | 16 | -0.093034112 | 0.026435911 | 0.003098798 | 64.7 | 30.1 | NA | NA | NA | NA | NA | NA |

| | | | | | | | | | | | | | | |
|---|---|---|---|---|---|---|---|---|---|---|---|---|---|---|
| HDL | Impedance of Arm (Right) | Inverse variance weighted | 100 | 0.017167851 | 0.008876191 | 0.053095154 | 193.1 | 59.2 | 685.09 | 0 | 85.5 | NA | NA | NA |
| HDL | Impedance of Arm (Right) | MR Egger | 100 | -0.003053554 | 0.017425748 | 0.861258649 | 193.1 | 59.2 | NA | NA | NA | 0.00148 | 0.001099 | 0.1812 |
| HDL | Impedance of Arm (Right) | Weighted median | 100 | 0.013373957 | 0.005933899 | 0.024207304 | 193.1 | 59.2 | NA | NA | NA | NA | NA | NA |
| HDL | Impedance of Arm (Right) | Weighted mode | 100 | 0.014119587 | 0.006762518 | 0.039372207 | 193.1 | 59.2 | NA | NA | NA | NA | NA | NA |
| LDL | Impedance of Arm (Right) | Inverse variance weighted | 99 | 0.016383274 | 0.005736011 | 0.004287264 | 208.9 | 56.4 | 362.16 | 0 | 72.9 | NA | NA | NA |
| LDL | Impedance of Arm (Right) | MR Egger | 99 | 0.018785865 | 0.010054364 | 0.064720327 | 208.9 | 56.4 | NA | NA | NA | -0.00021 | 0.000721 | 0.7712 |
| LDL | Impedance of Arm (Right) | Weighted median | 99 | 0.011961988 | 0.005207653 | 0.021618679 | 208.9 | 56.4 | NA | NA | NA | NA | NA | NA |
| LDL | Impedance of Arm (Right) | Weighted mode | 99 | 0.012582122 | 0.005050465 | 0.014408444 | 208.9 | 56.4 | NA | NA | NA | NA | NA | NA |
| MetS | Impedance of Arm (Right) | Inverse variance weighted | 96 | 0.050585052 | 0.01208421 | 2.84E-05 | 97.8 | 41 | 1895.39 | 0 | 95 | NA | NA | NA |
| MetS | Impedance of Arm (Right) | MR Egger | 96 | -0.028275094 | 0.026791313 | 0.293955242 | 97.8 | 41 | NA | NA | NA | 0.007326 | 0.002247 | 0.0016 |
| MetS | Impedance of Arm (Right) | Weighted median | 96 | 0.018976737 | 0.004590696 | 3.57E-05 | 97.8 | 41 | NA | NA | NA | NA | NA | NA |
| MetS | Impedance of Arm (Right) | Weighted mode | 96 | 0.017281022 | 0.005588999 | 0.002610019 | 97.8 | 41 | NA | NA | NA | NA | NA | NA |
| SBP | Impedance of Arm (Right) | Inverse variance weighted | 100 | 0.002179683 | 0.001501936 | 0.146710603 | 189.9 | 103.5 | 1339.33 | 0 | 92.6 | NA | NA | NA |
| SBP | Impedance of Arm (Right) | MR Egger | 100 | 0.006861482 | 0.004964955 | 0.170119058 | 189.9 | 103.5 | NA | NA | NA | -0.002364 | 0.00239 | 0.3249 |

| | | | | | | | | | | | | | | |
|---|---|---|---|---|---|---|---|---|---|---|---|---|---|---|
| SBP | Impedance of Arm (Right) | Weighted median | 100 | 0.00160014 | 0.000828594 | 0.053464381 | 189.9 | 103.5 | NA | NA | NA | NA | NA | NA |
| SBP | Impedance of Arm (Right) | Weighted mode | 100 | 0.002100561 | 0.001509079 | 0.167057173 | 189.9 | 103.5 | NA | NA | NA | NA | NA | NA |
| T2D | Impedance of Arm (Right) | Inverse variance weighted | 100 | -0.030279734 | 0.011865516 | 0.010713403 | 89.6 | 38.5 | 2256.56 | 0 | 95.6 | NA | NA | NA |
| T2D | Impedance of Arm (Right) | MR Egger | 100 | 0.03308732 | 0.027546983 | 0.232598356 | 89.6 | 38.5 | NA | NA | NA | -0.005614 | 0.002215 | 0.0129 |
| T2D | Impedance of Arm (Right) | Weighted median | 100 | 0.019695452 | 0.004674174 | 2.51E-05 | 89.6 | 38.5 | NA | NA | NA | NA | NA | NA |
| T2D | Impedance of Arm (Right) | Weighted mode | 100 | 0.020279289 | 0.004689981 | 3.66E-05 | 89.6 | 38.5 | NA | NA | NA | NA | NA | NA |
| TG | Impedance of Arm (Right) | Inverse variance weighted | 99 | 0.008570391 | 0.01548206 | 0.579873716 | 150.5 | 43 | 1451.6 | 0 | 93.2 | NA | NA | NA |
| TG | Impedance of Arm (Right) | MR Egger | 99 | 0.077509362 | 0.027209044 | 0.005360991 | 150.5 | 43 | NA | NA | NA | -0.004448 | 0.00147 | 0.0032 |
| TG | Impedance of Arm (Right) | Weighted median | 99 | 0.00153194 | 0.008181093 | 0.851461757 | 150.5 | 43 | NA | NA | NA | NA | NA | NA |
| TG | Impedance of Arm (Right) | Weighted mode | 99 | -0.011439024 | 0.007537884 | 0.132350324 | 150.5 | 43 | NA | NA | NA | NA | NA | NA |
| BMI | Overall Healthy Diet | Inverse variance weighted | 100 | -0.002488779 | 0.009430559 | 0.791852515 | 150.7 | 85.3 | 112.89 | 0.1607 | 12.3 | NA | NA | NA |
| BMI | Overall Healthy Diet | MR Egger | 100 | 0.013759416 | 0.026077204 | 0.598940773 | 150.7 | 85.3 | NA | NA | NA | -0.000486 | 0.000727 | 0.5053 |
| BMI | Overall Healthy Diet | Weighted median | 100 | -0.008193611 | 0.013960104 | 0.557250359 | 150.7 | 85.3 | NA | NA | NA | NA | NA | NA |

| | | | | | | | | | | | | | | |
|---|---|---|---|---|---|---|---|---|---|---|---|---|---|---|
| BMI | Overall Healthy Diet | Weighted mode | 100 | -0.014671954 | 0.021951951 | 0.505454883 | 150.7 | 85.3 | NA | NA | NA | NA | NA | NA |
| DBP | Overall Healthy Diet | Inverse variance weighted | 100 | -0.000209311 | 0.000846181 | 0.804630045 | 209.5 | 114 | 95.19 | 0.5896 | 0 | NA | NA | NA |
| DBP | Overall Healthy Diet | MR Egger | 100 | -0.00080527 | 0.002908522 | 0.782466219 | 209.5 | 114 | NA | NA | NA | 0.000184 | 0.000861 | 0.8309 |
| DBP | Overall Healthy Diet | Weighted median | 100 | -0.000607238 | 0.001252418 | 0.62778083 | 209.5 | 114 | NA | NA | NA | NA | NA | NA |
| DBP | Overall Healthy Diet | Weighted mode | 100 | -0.002491767 | 0.002320833 | 0.285588632 | 209.5 | 114 | NA | NA | NA | NA | NA | NA |
| HbA1c | Overall Healthy Diet | Inverse variance weighted | 16 | -0.038783904 | 0.018601554 | 0.037070936 | 64.7 | 30.1 | 11.36 | 0.727 | 0 | NA | NA | NA |
| HbA1c | Overall Healthy Diet | MR Egger | 16 | -0.07790102 | 0.044098329 | 0.099091833 | 64.7 | 30.1 | NA | NA | NA | 0.001498 | 0.001531 | 0.3445 |
| HbA1c | Overall Healthy Diet | Weighted median | 16 | -0.050510033 | 0.025569805 | 0.048225256 | 64.7 | 30.1 | NA | NA | NA | NA | NA | NA |
| HbA1c | Overall Healthy Diet | Weighted mode | 16 | -0.057172213 | 0.029668055 | 0.073130597 | 64.7 | 30.1 | NA | NA | NA | NA | NA | NA |
| HDL | Overall Healthy Diet | Inverse variance weighted | 100 | 0.015621378 | 0.004240882 | 0.000230034 | 193.1 | 59.2 | 97.96 | 0.5106 | 0 | NA | NA | NA |
| HDL | Overall Healthy Diet | MR Egger | 100 | 0.00776627 | 0.00837677 | 0.356141978 | 193.1 | 59.2 | NA | NA | NA | 0.000574 | 0.000528 | 0.2795 |
| HDL | Overall Healthy Diet | Weighted median | 100 | 0.019364642 | 0.006861905 | 0.004771769 | 193.1 | 59.2 | NA | NA | NA | NA | NA | NA |

| | | | | | | | | | | | | | | |
|---|---|---|---|---|---|---|---|---|---|---|---|---|---|---|
| HDL | Overall Healthy Diet | Weighted mode | 100 | 0.015853421 | 0.007698227 | 0.042084708 | 193.1 | 59.2 | NA | NA | NA | NA | NA | NA |
| LDL | Overall Healthy Diet | Inverse variance weighted | 100 | 0.006755321 | 0.0039586 | 0.087916413 | 207.8 | 56.4 | 111.3 | 0.1874 | 11.1 | NA | NA | NA |
| LDL | Overall Healthy Diet | MR Egger | 100 | 0.002335354 | 0.006920065 | 0.73648006 | 207.8 | 56.4 | NA | NA | NA | 0.000386 | 0.000496 | 0.4376 |
| LDL | Overall Healthy Diet | Weighted median | 100 | 0.006865918 | 0.006234817 | 0.270800017 | 207.8 | 56.4 | NA | NA | NA | NA | NA | NA |
| LDL | Overall Healthy Diet | Weighted mode | 100 | 0.002307753 | 0.007401152 | 0.755840702 | 207.8 | 56.4 | NA | NA | NA | NA | NA | NA |
| MetS | Overall Healthy Diet | Inverse variance weighted | 95 | 0.01113522 | 0.003405379 | 0.001075889 | 98.4 | 41 | 75.39 | 0.9207 | 0 | NA | NA | NA |
| MetS | Overall Healthy Diet | MR Egger | 95 | 0.021673874 | 0.007928403 | 0.007497482 | 98.4 | 41 | NA | NA | NA | -0.00098 | 0.000666 | 0.1444 |
| MetS | Overall Healthy Diet | Weighted median | 95 | 0.016018496 | 0.004901221 | 0.001082084 | 98.4 | 41 | NA | NA | NA | NA | NA | NA |
| MetS | Overall Healthy Diet | Weighted mode | 95 | 0.020550093 | 0.009611981 | 0.035113235 | 98.4 | 41 | NA | NA | NA | NA | NA | NA |
| SBP | Overall Healthy Diet | Inverse variance weighted | 100 | -0.000607223 | 0.000512035 | 0.235660811 | 189.9 | 103.5 | 74.04 | 0.9714 | 0 | NA | NA | NA |
| SBP | Overall Healthy Diet | MR Egger | 100 | -0.00034693 | 0.0016912 | 0.837889684 | 189.9 | 103.5 | NA | NA | NA | -0.000132 | 0.000814 | 0.872 |
| SBP | Overall Healthy Diet | Weighted median | 100 | -0.000827603 | 0.000720004 | 0.250373782 | 189.9 | 103.5 | NA | NA | NA | NA | NA | NA |
| SBP | Overall Healthy Diet | Weighted mode | 100 | -0.001558706 | 0.00170657 | 0.363274288 | 189.9 | 103.5 | NA | NA | NA | NA | NA | NA |

| | | | | | | | | | | | | | | |
|---|---|---|---|---|---|---|---|---|---|---|---|---|---|---|
| T2D | Overall Healthy Diet | Inverse variance weighted | 100 | -0.000733684 | 0.003453023 | 0.831735924 | 89.6 | 38.5 | 121.32 | 0.0633 | 18.4 | NA | NA | NA |
| T2D | Overall Healthy Diet | MR Egger | 100 | -0.000758858 | 0.008281335 | 0.927175316 | 89.6 | 38.5 | NA | NA | NA | 2.00E-06 | 0.000666 | 0.9973 |
| T2D | Overall Healthy Diet | Weighted median | 100 | -0.004240889 | 0.005141383 | 0.409454621 | 89.6 | 38.5 | NA | NA | NA | NA | NA | NA |
| T2D | Overall Healthy Diet | Weighted mode | 100 | -0.005599251 | 0.006807627 | 0.412770885 | 89.6 | 38.5 | NA | NA | NA | NA | NA | NA |
| TG | Overall Healthy Diet | Inverse variance weighted | 100 | -0.003023613 | 0.005522279 | 0.584014711 | 149.4 | 43 | 119.04 | 0.0831 | 16.8 | NA | NA | NA |
| TG | Overall Healthy Diet | MR Egger | 100 | -0.006981694 | 0.010114433 | 0.49165566 | 149.4 | 43 | NA | NA | NA | 0.000254 | 0.000543 | 0.6409 |
| TG | Overall Healthy Diet | Weighted median | 100 | -0.006675316 | 0.008476904 | 0.431006175 | 149.4 | 43 | NA | NA | NA | NA | NA | NA |
| TG | Overall Healthy Diet | Weighted mode | 100 | -0.013110458 | 0.011246254 | 0.246511521 | 149.4 | 43 | NA | NA | NA | NA | NA | NA |
| BMI | Peak Expiratory Flow | Inverse variance weighted | 99 | -0.013100893 | 0.017060649 | 0.442545873 | 150.8 | 85.3 | 356.2 | 0 | 72.5 | NA | NA | NA |
| BMI | Peak Expiratory Flow | MR Egger | 99 | 0.092682707 | 0.045701331 | 0.045301509 | 150.8 | 85.3 | NA | NA | NA | -0.003168 | 0.001275 | 0.0147 |
| BMI | Peak Expiratory Flow | Weighted median | 99 | 0.026761166 | 0.016641733 | 0.107818597 | 150.8 | 85.3 | NA | NA | NA | NA | NA | NA |
| BMI | Peak Expiratory Flow | Weighted mode | 99 | 0.051251935 | 0.023582756 | 0.03216883 | 150.8 | 85.3 | NA | NA | NA | NA | NA | NA |

| | | | | | | | | | | | | | | |
|---|---|---|---|---|---|---|---|---|---|---|---|---|---|---|
| DBP | Peak Expiratory Flow | Inverse variance weighted | 100 | 0.00232266 | 0.001728696 | 0.179080638 | 209.5 | 114 | 405.89 | 0 | 75.6 | NA | NA | NA |
| DBP | Peak Expiratory Flow | MR Egger | 100 | -0.007704666 | 0.005877716 | 0.192979833 | 209.5 | 114 | NA | NA | NA | 0.003103 | 0.00174 | 0.0777 |
| DBP | Peak Expiratory Flow | Weighted median | 100 | 0.001942322 | 0.001503098 | 0.196283538 | 209.5 | 114 | NA | NA | NA | NA | NA | NA |
| DBP | Peak Expiratory Flow | Weighted mode | 100 | 0.002737663 | 0.003060848 | 0.373270105 | 209.5 | 114 | NA | NA | NA | NA | NA | NA |
| HbA1c | Peak Expiratory Flow | Inverse variance weighted | 16 | -0.003835245 | 0.027540355 | 0.889245398 | 64.7 | 30.1 | 32.47 | 0.0055 | 53.8 | NA | NA | NA |
| HbA1c | Peak Expiratory Flow | MR Egger | 16 | -0.009357326 | 0.067465186 | 0.891663657 | 64.7 | 30.1 | NA | NA | NA | 0.000212 | 0.002345 | 0.9293 |
| HbA1c | Peak Expiratory Flow | Weighted median | 16 | 0.019106395 | 0.027914249 | 0.493680018 | 64.7 | 30.1 | NA | NA | NA | NA | NA | NA |
| HbA1c | Peak Expiratory Flow | Weighted mode | 16 | 0.017700718 | 0.035838308 | 0.628527188 | 64.7 | 30.1 | NA | NA | NA | NA | NA | NA |
| HDL | Peak Expiratory Flow | Inverse variance weighted | 100 | 0.009548232 | 0.005369096 | 0.075343321 | 193.1 | 59.2 | 156.47 | 2.00E-04 | 36.7 | NA | NA | NA |
| HDL | Peak Expiratory Flow | MR Egger | 100 | 0.013485124 | 0.010629101 | 0.207553885 | 193.1 | 59.2 | NA | NA | NA | -0.000288 | 0.00067 | 0.6683 |
| HDL | Peak Expiratory Flow | Weighted median | 100 | 0.014535432 | 0.006574353 | 0.027040717 | 193.1 | 59.2 | NA | NA | NA | NA | NA | NA |
| HDL | Peak Expiratory Flow | Weighted mode | 100 | 0.014085886 | 0.006905696 | 0.044039071 | 193.1 | 59.2 | NA | NA | NA | NA | NA | NA |
| LDL | Peak Expiratory Flow | Inverse variance weighted | 99 | -0.0051614 | 0.005051944 | 0.306938972 | 208.9 | 56.4 | 175.35 | 0 | 44.1 | NA | NA | NA |

| | | | | | | | | | | | | | | |
|---|---|---|---|---|---|---|---|---|---|---|---|---|---|---|
| LDL | Peak Expiratory Flow | MR Egger | 99 | 0.006232565 | 0.008747996 | 0.477892516 | 208.9 | 56.4 | NA | NA | NA | -0.000997 | 0.000627 | 0.1152 |
| LDL | Peak Expiratory Flow | Weighted median | 99 | -0.002799652 | 0.006472886 | 0.665363625 | 208.9 | 56.4 | NA | NA | NA | NA | NA | NA |
| LDL | Peak Expiratory Flow | Weighted mode | 99 | 0.002185768 | 0.00794881 | 0.783910098 | 208.9 | 56.4 | NA | NA | NA | NA | NA | NA |
| MetS | Peak Expiratory Flow | Inverse variance weighted | 96 | 0.000621368 | 0.005430256 | 0.908899304 | 97.8 | 41 | 238.92 | 0 | 60.2 | NA | NA | NA |
| MetS | Peak Expiratory Flow | MR Egger | 96 | 0.000570762 | 0.012701702 | 0.964253753 | 97.8 | 41 | NA | NA | NA | 5.00E-06 | 0.001065 | 0.9965 |
| MetS | Peak Expiratory Flow | Weighted median | 96 | 0.011171649 | 0.005485305 | 0.041685085 | 97.8 | 41 | NA | NA | NA | NA | NA | NA |
| MetS | Peak Expiratory Flow | Weighted mode | 96 | 0.018506032 | 0.008108849 | 0.02470876 | 97.8 | 41 | NA | NA | NA | NA | NA | NA |
| SBP | Peak Expiratory Flow | Inverse variance weighted | 100 | -0.002569742 | 0.000907254 | 0.004619447 | 189.9 | 103.5 | 305.12 | 0 | 67.6 | NA | NA | NA |
| SBP | Peak Expiratory Flow | MR Egger | 100 | -0.003213267 | 0.003012955 | 0.28882555 | 189.9 | 103.5 | NA | NA | NA | 0.000325 | 0.00145 | 0.8232 |
| SBP | Peak Expiratory Flow | Weighted median | 100 | -0.002429845 | 0.00087949 | 0.005730992 | 189.9 | 103.5 | NA | NA | NA | NA | NA | NA |
| SBP | Peak Expiratory Flow | Weighted mode | 100 | -0.004150873 | 0.001900464 | 0.031313338 | 189.9 | 103.5 | NA | NA | NA | NA | NA | NA |
| T2D | Peak Expiratory Flow | Inverse variance weighted | 100 | -0.012394524 | 0.006130216 | 0.043189396 | 89.6 | 38.5 | 375.9 | 0 | 73.7 | NA | NA | NA |

| | | | | | | | | | | | | | | |
|---|---|---|---|---|---|---|---|---|---|---|---|---|---|---|
| T2D | Peak Expiratory Flow | MR Egger | 100 | -0.018226267 | 0.014681265 | 0.217397526 | 89.6 | 38.5 | NA | NA | NA | 0.000517 | 0.001181 | 0.6627 |
| T2D | Peak Expiratory Flow | Weighted median | 100 | -0.003081687 | 0.006360311 | 0.628018048 | 89.6 | 38.5 | NA | NA | NA | NA | NA | NA |
| T2D | Peak Expiratory Flow | Weighted mode | 100 | -0.005110866 | 0.006640361 | 0.443330048 | 89.6 | 38.5 | NA | NA | NA | NA | NA | NA |
| TG | Peak Expiratory Flow | Inverse variance weighted | 99 | 0.009668599 | 0.007690895 | 0.208699799 | 150.5 | 43 | 223.56 | 0 | 56.2 | NA | NA | NA |
| TG | Peak Expiratory Flow | MR Egger | 99 | 0.004233162 | 0.014126844 | 0.765082418 | 150.5 | 43 | NA | NA | NA | 0.000351 | 0.000763 | 0.6469 |
| TG | Peak Expiratory Flow | Weighted median | 99 | 0.009112471 | 0.008992432 | 0.310893534 | 150.5 | 43 | NA | NA | NA | NA | NA | NA |
| TG | Peak Expiratory Flow | Weighted mode | 99 | -0.015818659 | 0.011079387 | 0.156541117 | 150.5 | 43 | NA | NA | NA | NA | NA | NA |
| BMI | Potassium in Urine | Inverse variance weighted | 99 | 0.049384674 | 0.01668073 | 0.00307058 | 150.8 | 85.3 | 267.42 | 0 | 63.4 | NA | NA | NA |
| BMI | Potassium in Urine | MR Egger | 99 | 0.051398854 | 0.046076797 | 0.267390438 | 150.8 | 85.3 | NA | NA | NA | -6.00E-05 | 0.001286 | 0.9627 |
| BMI | Potassium in Urine | Weighted median | 99 | 0.048736687 | 0.017748925 | 0.006034603 | 150.8 | 85.3 | NA | NA | NA | NA | NA | NA |
| BMI | Potassium in Urine | Weighted mode | 99 | 0.077154698 | 0.029758078 | 0.010978304 | 150.8 | 85.3 | NA | NA | NA | NA | NA | NA |
| DBP | Potassium in Urine | Inverse variance weighted | 100 | 0.002893242 | 0.001681734 | 0.085361222 | 209.5 | 114 | 301.66 | 0 | 67.2 | NA | NA | NA |
| DBP | Potassium in Urine | MR Egger | 100 | -0.00482154 | 0.005752096 | 0.40394511 | 209.5 | 114 | NA | NA | NA | 0.002387 | 0.001703 | 0.1641 |

| | | | | | | | | | | | | | | |
|---|---|---|---|---|---|---|---|---|---|---|---|---|---|---|
| DBP | Potassium in Urine | Weighted median | 100 | 0.000946051 | 0.001649717 | 0.566331422 | 209.5 | 114 | NA | NA | NA | NA | NA | NA |
| DBP | Potassium in Urine | Weighted mode | 100 | 0.00083494 | 0.003950175 | 0.833034713 | 209.5 | 114 | NA | NA | NA | NA | NA | NA |
| HbA1c | Potassium in Urine | Inverse variance weighted | 16 | 0.046523909 | 0.024248434 | 0.055030465 | 64.7 | 30.1 | 19.77 | 0.1808 | 24.1 | NA | NA | NA |
| HbA1c | Potassium in Urine | MR Egger | 16 | 0.171095231 | 0.049991482 | 0.00412423 | 64.7 | 30.1 | NA | NA | NA | -0.004778 | 0.001738 | 0.0157 |
| HbA1c | Potassium in Urine | Weighted median | 16 | 0.069196719 | 0.032190319 | 0.031585864 | 64.7 | 30.1 | NA | NA | NA | NA | NA | NA |
| HbA1c | Potassium in Urine | Weighted mode | 16 | 0.087618696 | 0.036156266 | 0.028493713 | 64.7 | 30.1 | NA | NA | NA | NA | NA | NA |
| HDL | Potassium in Urine | Inverse variance weighted | 100 | -0.000997533 | 0.005613525 | 0.858957292 | 193.1 | 59.2 | 134.35 | 0.0105 | 26.3 | NA | NA | NA |
| HDL | Potassium in Urine | MR Egger | 100 | 0.004732423 | 0.01110058 | 0.67080811 | 193.1 | 59.2 | NA | NA | NA | -0.000419 | 7.00E-04 | 0.5506 |
| HDL | Potassium in Urine | Weighted median | 100 | -0.002807412 | 0.007609454 | 0.712174426 | 193.1 | 59.2 | NA | NA | NA | NA | NA | NA |
| HDL | Potassium in Urine | Weighted mode | 100 | 0.001099124 | 0.008190157 | 0.8935164 | 193.1 | 59.2 | NA | NA | NA | NA | NA | NA |
| LDL | Potassium in Urine | Inverse variance weighted | 99 | -0.003885039 | 0.004949468 | 0.432488415 | 208.9 | 56.4 | 132.18 | 0.0122 | 25.9 | NA | NA | NA |
| LDL | Potassium in Urine | MR Egger | 99 | -0.00156462 | 0.008675836 | 0.857260125 | 208.9 | 56.4 | NA | NA | NA | -0.000203 | 0.000622 | 0.7448 |
| LDL | Potassium in Urine | Weighted median | 99 | 0.003476309 | 0.006783858 | 0.608344289 | 208.9 | 56.4 | NA | NA | NA | NA | NA | NA |

| | | | | | | | | | | | | | | |
|---|---|---|---|---|---|---|---|---|---|---|---|---|---|---|
| LDL | Potassium in Urine | Weighted mode | 99 | 0.003116937 | 0.007905837 | 0.694247428 | 208.9 | 56.4 | NA | NA | NA | NA | NA | NA |
| MetS | Potassium in Urine | Inverse variance weighted | 96 | 0.001209995 | 0.005516093 | 0.826371693 | 97.8 | 41 | 193.62 | 0 | 50.9 | NA | NA | NA |
| MetS | Potassium in Urine | MR Egger | 96 | 0.028327059 | 0.012522932 | 0.026000366 | 97.8 | 41 | NA | NA | NA | -0.002519 | 0.00105 | 0.0184 |
| MetS | Potassium in Urine | Weighted median | 96 | 0.007197408 | 0.006689963 | 0.281993613 | 97.8 | 41 | NA | NA | NA | NA | NA | NA |
| MetS | Potassium in Urine | Weighted mode | 96 | 0.009801667 | 0.010584736 | 0.356782564 | 97.8 | 41 | NA | NA | NA | NA | NA | NA |
| SBP | Potassium in Urine | Inverse variance weighted | 100 | -0.000681927 | 0.00094384 | 0.469985456 | 189.9 | 103.5 | 259.34 | 0 | 61.8 | NA | NA | NA |
| SBP | Potassium in Urine | MR Egger | 100 | 0.002450145 | 0.003117769 | 0.433842691 | 189.9 | 103.5 | NA | NA | NA | -0.001582 | 0.001501 | 0.2945 |
| SBP | Potassium in Urine | Weighted median | 100 | 0.00011509 | 0.00099687 | 0.90808716 | 189.9 | 103.5 | NA | NA | NA | NA | NA | NA |
| SBP | Potassium in Urine | Weighted mode | 100 | 0.001365506 | 0.001868313 | 0.466580849 | 189.9 | 103.5 | NA | NA | NA | NA | NA | NA |
| T2D | Potassium in Urine | Inverse variance weighted | 100 | -0.005059539 | 0.0048 1312 | 0.293168025 | 89.6 | 38.5 | 181.98 | 0 | 45.6 | NA | NA | NA |
| T2D | Potassium in Urine | MR Egger | 100 | 0.009793431 | 0.011416357 | 0.393072843 | 89.6 | 38.5 | NA | NA | NA | -0.001316 | 0.000918 | 0.155 |
| T2D | Potassium in Urine | Weighted median | 100 | -0.005249795 | 0.006864488 | 0.444405022 | 89.6 | 38.5 | NA | NA | NA | NA | NA | NA |
| T2D | Potassium in Urine | Weighted mode | 100 | -0.008084851 | 0.007082118 | 0.256379665 | 89.6 | 38.5 | NA | NA | NA | NA | NA | NA |

| | | | | | | | | | | | | | | |
|---|---|---|---|---|---|---|---|---|---|---|---|---|---|---|
| TG | Potassium in Urine | Inverse variance weighted | 99 | -0.006507043 | 0.009258356 | 0.482162142 | 150.5 | 43 | 254.46 | 0 | 61.5 | NA | NA | NA |
| TG | Potassium in Urine | MR Egger | 99 | -0.020113107 | 0.016941903 | 0.238055406 | 150.5 | 43 | NA | NA | NA | 0.000878 | 0.000915 | 0.3399 |
| TG | Potassium in Urine | Weighted median | 99 | -0.017402354 | 0.009538437 | 0.068084811 | 150.5 | 43 | NA | NA | NA | NA | NA | NA |
| TG | Potassium in Urine | Weighted mode | 99 | -0.024017158 | 0.011889919 | 0.046117179 | 150.5 | 43 | NA | NA | NA | NA | NA | NA |
| BMI | Red Blood Cell Distribution | Inverse variance weighted | 99 | -0.116681806 | 0.036134018 | 0.001241631 | 151.3 | 85.3 | 599.5 | 0 | 83.7 | NA | NA | NA |
| BMI | Red Blood Cell Distribution | MR Egger | 99 | 0.014921945 | 0.099351528 | 0.880923896 | 151.3 | 85.3 | NA | NA | NA | -0.003948 | 0.002778 | 0.1586 |
| BMI | Red Blood Cell Distribution | Weighted median | 99 | -0.070190795 | 0.024781047 | 0.004619444 | 151.3 | 85.3 | NA | NA | NA | NA | NA | NA |
| BMI | Red Blood Cell Distribution | Weighted mode | 99 | -0.099517552 | 0.039092617 | 0.012465083 | 151.3 | 85.3 | NA | NA | NA | NA | NA | NA |
| DBP | Red Blood Cell Distribution | Inverse variance weighted | 99 | 0.017021575 | 0.0064331 | 0.008146509 | 210 | 114 | 2076.29 | 0 | 95.3 | NA | NA | NA |
| DBP | Red Blood Cell Distribution | MR Egger | 99 | 0.015188417 | 0.022014703 | 0.491890903 | 210 | 114 | NA | NA | NA | 0.000569 | 0.006536 | 0.9308 |
| DBP | Red Blood Cell Distribution | Weighted median | 99 | 0.005602622 | 0.002515263 | 0.025917224 | 210 | 114 | NA | NA | NA | NA | NA | NA |
| DBP | Red Blood Cell Distribution | Weighted mode | 99 | 0.008644687 | 0.006437309 | 0.18240514 | 210 | 114 | NA | NA | NA | NA | NA | NA |

| | | | | | | | | | | | | | | |
|---|---|---|---|---|---|---|---|---|---|---|---|---|---|---|
| HbA1c | Red Blood Cell Distribution | Inverse variance weighted | 16 | -0.617366748 | 0.385304318 | 0.109092934 | 64.7 | 30.1 | 2389.36 | 0 | 99.4 | NA | NA | NA |
| HbA1c | Red Blood Cell Distribution | MR Egger | 16 | 0.062896768 | 0.920290943 | 0.946477986 | 64.7 | 30.1 | NA | NA | NA | -0.026181 | 0.032087 | 0.4282 |
| HbA1c | Red Blood Cell Distribution | Weighted median | 16 | 0.31184682 | 0.053785862 | 6.71E-09 | 64.7 | 30.1 | NA | NA | NA | NA | NA | NA |
| HbA1c | Red Blood Cell Distribution | Weighted mode | 16 | 0.263890106 | 0.041498465 | 1.28E-05 | 64.7 | 30.1 | NA | NA | NA | NA | NA | NA |
| HDL | Red Blood Cell Distribution | Inverse variance weighted | 99 | -0.081852655 | 0.024168861 | 0.00070739 | 193.7 | 59.2 | 1160.86 | 0 | 91.6 | NA | NA | NA |
| HDL | Red Blood Cell Distribution | MR Egger | 99 | -0.008576803 | 0.047032018 | 0.855680065 | 193.7 | 59.2 | NA | NA | NA | -0.005373 | 0.002971 | 0.0736 |
| HDL | Red Blood Cell Distribution | Weighted median | 99 | -0.02324279 | 0.013359579 | 0.081896847 | 193.7 | 59.2 | NA | NA | NA | NA | NA | NA |
| HDL | Red Blood Cell Distribution | Weighted mode | 99 | 0.000210358 | 0.011695516 | 0.985686445 | 193.7 | 59.2 | NA | NA | NA | NA | NA | NA |
| LDL | Red Blood Cell Distribution | Inverse variance weighted | 99 | 0.067169949 | 0.02726507 | 0.013755342 | 207.7 | 56.4 | 1917.29 | 0 | 94.9 | NA | NA | NA |
| LDL | Red Blood Cell Distribution | MR Egger | 99 | 0.113894908 | 0.047344494 | 0.018038664 | 207.7 | 56.4 | NA | NA | NA | -0.004092 | 0.003394 | 0.2308 |
| LDL | Red Blood Cell Distribution | Weighted median | 99 | 0.096499195 | 0.015129697 | 1.79E-10 | 207.7 | 56.4 | NA | NA | NA | NA | NA | NA |
| LDL | Red Blood Cell Distribution | Weighted mode | 99 | 0.121509823 | 0.045725338 | 0.009195965 | 207.7 | 56.4 | NA | NA | NA | NA | NA | NA |
| MetS | Red Blood Cell Distribution | Inverse variance weighted | 100 | -0.089072919 | 0.016046135 | 2.84E-08 | 97.4 | 41 | 841.47 | 0 | 88.2 | NA | NA | NA |

| | | | | | | | | | | | | | | |
|---|---|---|---|---|---|---|---|---|---|---|---|---|---|---|
| MetS | Red Blood Cell Distribution | MR Egger | 100 | -0.100985806 | 0.036812704 | 0.007234867 | 97.4 | 41 | NA | NA | NA | 0.001108 | 0.003079 | 0.7197 |
| MetS | Red Blood Cell Distribution | Weighted median | 100 | -0.100421653 | 0.01152905 | 3.03E-18 | 97.4 | 41 | NA | NA | NA | NA | NA | NA |
| MetS | Red Blood Cell Distribution | Weighted mode | 100 | -0.133173068 | 0.025341684 | 8.51E-07 | 97.4 | 41 | NA | NA | NA | NA | NA | NA |
| SBP | Red Blood Cell Distribution | Inverse variance weighted | 100 | 0.002467242 | 0.002974515 | 0.40684402 | 189.9 | 103.5 | 1240.58 | 0 | 92 | NA | NA | NA |
| SBP | Red Blood Cell Distribution | MR Egger | 100 | 0.003782931 | 0.009688576 | 0.697049226 | 189.9 | 103.5 | NA | NA | NA | -0.00067 | 0.00469 | 0.8868 |
| SBP | Red Blood Cell Distribution | Weighted median | 100 | 0.000620555 | 0.00153198 | 0.685428285 | 189.9 | 103.5 | NA | NA | NA | NA | NA | NA |
| SBP | Red Blood Cell Distribution | Weighted mode | 100 | -2.34E-05 | 0.003670995 | 0.994918675 | 189.9 | 103.5 | NA | NA | NA | NA | NA | NA |
| T2D | Red Blood Cell Distribution | Inverse variance weighted | 100 | 0.012137764 | 0.014012632 | 0.386379636 | 89.6 | 38.5 | 733.04 | 0 | 86.5 | NA | NA | NA |
| T2D | Red Blood Cell Distribution | MR Egger | 100 | 0.057114335 | 0.033221673 | 0.088737101 | 89.6 | 38.5 | NA | NA | NA | -0.003983 | 0.002671 | 0.1391 |
| T2D | Red Blood Cell Distribution | Weighted median | 100 | 0.036386337 | 0.010227851 | 0.000374296 | 89.6 | 38.5 | NA | NA | NA | NA | NA | NA |
| T2D | Red Blood Cell Distribution | Weighted mode | 100 | 0.034299634 | 0.010551422 | 0.001573766 | 89.6 | 38.5 | NA | NA | NA | NA | NA | NA |
| TG | Red Blood Cell Distribution | Inverse variance weighted | 100 | 0.175378242 | 0.026619451 | 4.45E-11 | 149.4 | 43 | 1012.21 | 0 | 90.2 | NA | NA | NA |
| TG | Red Blood Cell Distribution | MR Egger | 100 | 0.133322647 | 0.048628159 | 0.007266742 | 149.4 | 43 | NA | NA | NA | 0.002695 | 0.002608 | 0.304 |
| TG | Red Blood Cell Distribution | Weighted median | 100 | 0.20572459 | 0.016479277 | 9.15E-36 | 149.4 | 43 | NA | NA | NA | NA | NA | NA |

| | | | | | | | | | | | | | | |
|---|---|---|---|---|---|---|---|---|---|---|---|---|---|---|
| TG | Red Blood Cell Distribution | Weighted mode | 100 | 0.200355897 | 0.015982479 | 3.71E-22 | 149.4 | 43 | NA | NA | NA | NA | NA | NA |
| BMI | Serum Phosphate | Inverse variance weighted | 45 | 0.00467338 | 0.010692911 | 0.662072164 | 141.3 | 85.3 | 54.95 | 0.1247 | 19.9 | NA | NA | NA |
| BMI | Serum Phosphate | MR Egger | 45 | 0.066985234 | 0.031709856 | 0.040494152 | 141.3 | 85.3 | NA | NA | NA | -0.001768 | 0.000851 | 0.0437 |
| BMI | Serum Phosphate | Weighted median | 45 | 0.02284851 | 0.014365711 | 0.111724554 | 141.3 | 85.3 | NA | NA | NA | NA | NA | NA |
| BMI | Serum Phosphate | Weighted mode | 45 | 0.031274474 | 0.019396993 | 0.114039545 | 141.3 | 85.3 | NA | NA | NA | NA | NA | NA |
| DBP | Serum Phosphate | Inverse variance weighted | 49 | -9.25E-05 | 0.000931508 | 0.920940406 | 191.3 | 114 | 35.67 | 0.906 | 0 | NA | NA | NA |
| DBP | Serum Phosphate | MR Egger | 49 | 0.001209841 | 0.003444746 | 0.726997758 | 191.3 | 114 | NA | NA | NA | -0.000379 | 0.000966 | 0.6963 |
| DBP | Serum Phosphate | Weighted median | 49 | 0.00033881 | 0.001347748 | 0.801512687 | 191.3 | 114 | NA | NA | NA | NA | NA | NA |
| DBP | Serum Phosphate | Weighted mode | 49 | 0.000575927 | 0.002767623 | 0.836035631 | 191.3 | 114 | NA | NA | NA | NA | NA | NA |
| HbA1c | Serum Phosphate | Inverse variance weighted | 16 | 0.003529733 | 0.013900378 | 0.799549103 | 64.7 | 30.1 | 11.28 | 0.7322 | 0 | NA | NA | NA |
| HbA1c | Serum Phosphate | MR Egger | 16 | -0.008163136 | 0.033343443 | 0.810147897 | 64.7 | 30.1 | NA | NA | NA | 0.000449 | 0.001163 | 0.7054 |
| HbA1c | Serum Phosphate | Weighted median | 16 | 0.0171016 | 0.018825907 | 0.363662817 | 64.7 | 30.1 | NA | NA | NA | NA | NA | NA |
| HbA1c | Serum Phosphate | Weighted mode | 16 | 0.022162378 | 0.024607657 | 0.382017988 | 64.7 | 30.1 | NA | NA | NA | NA | NA | NA |
| HDL | Serum Phosphate | Inverse variance weighted | 94 | -0.020986448 | 0.003666891 | 1.05E-08 | 193.5 | 59.2 | 110.73 | 0.1014 | 16 | NA | NA | NA |

| | | | | | | | | | | | | | | |
|---|---|---|---|---|---|---|---|---|---|---|---|---|---|---|
| HDL | Serum Phosphate | MR Egger | 94 | -0.020551506 | 0.007754555 | 0.009469024 | 193.5 | 59.2 | NA | NA | NA | -2.90E-05 | 0.000456 | 0.9493 |
| HDL | Serum Phosphate | Weighted median | 94 | -0.020110588 | 0.005289388 | 0.000143496 | 193.5 | 59.2 | NA | NA | NA | NA | NA | NA |
| HDL | Serum Phosphate | Weighted mode | 94 | -0.021512964 | 0.006707481 | 0.001837863 | 193.5 | 59.2 | NA | NA | NA | NA | NA | NA |
| LDL | Serum Phosphate | Inverse variance weighted | 96 | -0.019818146 | 0.00368256 | 7.38E-08 | 205.5 | 56.4 | 145.55 | 7.00E-04 | 34.7 | NA | NA | NA |
| LDL | Serum Phosphate | MR Egger | 96 | -0.026356451 | 0.006848628 | 0.000216604 | 205.5 | 56.4 | NA | NA | NA | 0.000523 | 0.000462 | 0.2607 |
| LDL | Serum Phosphate | Weighted median | 96 | -0.020289446 | 0.005273622 | 0.000119404 | 205.5 | 56.4 | NA | NA | NA | NA | NA | NA |
| LDL | Serum Phosphate | Weighted mode | 96 | -0.012248266 | 0.007562583 | 0.108635981 | 205.5 | 56.4 | NA | NA | NA | NA | NA | NA |
| MetS | Serum Phosphate | Inverse variance weighted | 41 | -0.01638736 | 0.003957979 | 3.47E-05 | 110.4 | 42.1 | 47.39 | 0.1966 | 15.6 | NA | NA | NA |
| MetS | Serum Phosphate | MR Egger | 41 | -0.000781301 | 0.0099883 1 | 0.93805178 | 110.4 | 42.1 | NA | NA | NA | -0.001359 | 0.000802 | 0.0981 |
| MetS | Serum Phosphate | Weighted median | 41 | -0.012933585 | 0.005643564 | 0.021920607 | 110.4 | 42.1 | NA | NA | NA | NA | NA | NA |
| MetS | Serum Phosphate | Weighted mode | 41 | -0.009841349 | 0.009396725 | 0.301240879 | 110.4 | 42.1 | NA | NA | NA | NA | NA | NA |

| | | | | | | | | | | | | | | |
|---|---|---|---|---|---|---|---|---|---|---|---|---|---|---|
| SBP | Serum Phosphate | Inverse variance weighted | 45 | -0.000433759 | 0.000558342 | 0.437236204 | 186.3 | 103.5 | 32.37 | 0.9026 | 0 | NA | NA | NA |
| SBP | Serum Phosphate | MR Egger | 45 | 0.000879142 | 0.001769214 | 0.621783214 | 186.3 | 103.5 | NA | NA | NA | -0.00066 | 0.000844 | 0.4385 |
| SBP | Serum Phosphate | Weighted median | 45 | 0.000253315 | 0.00083635 | 0.761979628 | 186.3 | 103.5 | NA | NA | NA | NA | NA | NA |
| SBP | Serum Phosphate | Weighted mode | 45 | 0.000670253 | 0.001456022 | 0.64754496 | 186.3 | 103.5 | NA | NA | NA | NA | NA | NA |
| T2D | Serum Phosphate | Inverse variance weighted | 90 | -0.001023993 | 0.002380264 | 0.667049206 | 94.1 | 38.6 | 96.37 | 0.2785 | 7.6 | NA | NA | NA |
| T2D | Serum Phosphate | MR Egger | 90 | -0.00244406 | 0.005538075 | 0.660064212 | 94.1 | 38.6 | NA | NA | NA | 0.000129 | 0.000454 | 0.7768 |
| T2D | Serum Phosphate | Weighted median | 90 | -0.004809505 | 0.003862698 | 0.213089294 | 94.1 | 38.6 | NA | NA | NA | NA | NA | NA |
| T2D | Serum Phosphate | Weighted mode | 90 | -0.005515642 | 0.004471245 | 0.220607009 | 94.1 | 38.6 | NA | NA | NA | NA | NA | NA |
| TG | Serum Phosphate | Inverse variance weighted | 94 | 0.008140981 | 0.004769558 | 0.087847566 | 155.1 | 43 | 148.91 | 2.00E-04 | 37.5 | NA | NA | NA |
| TG | Serum Phosphate | MR Egger | 94 | -0.008840976 | 0.008546431 | 0.303630835 | 155.1 | 43 | NA | NA | NA | 0.001063 | 0.000449 | 0.0199 |
| TG | Serum Phosphate | Weighted median | 94 | 0.002585468 | 0.006307957 | 0.681898347 | 155.1 | 43 | NA | NA | NA | NA | NA | NA |
| TG | Serum Phosphate | Weighted mode | 94 | 7.00E-05 | 0.007000157 | 0.992046648 | 155.1 | 43 | NA | NA | NA | NA | NA | NA |
| BMI | Sodium in Urine | Inverse variance weighted | 99 | 0.107685136 | 0.021142547 | 3.52E-07 | 150.8 | 85.3 | 457.01 | 0 | 78.6 | NA | NA | NA |

| | | | | | | | | | | | | | | |
|---|---|---|---|---|---|---|---|---|---|---|---|---|---|---|
| BMI | Sodium in Urine | MR Egger | 99 | 0.028356723 | 0.057758115 | 0.624566133 | 150.8 | 85.3 | NA | NA | NA | 0.002376 | 0.001611 | 0.1436 |
| BMI | Sodium in Urine | Weighted median | 99 | 0.129709055 | 0.019311201 | 1.86E-11 | 150.8 | 85.3 | NA | NA | NA | NA | NA | NA |
| BMI | Sodium in Urine | Weighted mode | 99 | 0.219473393 | 0.039373214 | 2.19E-07 | 150.8 | 85.3 | NA | NA | NA | NA | NA | NA |
| DBP | Sodium in Urine | Inverse variance weighted | 100 | -0.00299023 | 0.001618281 | 0.064633953 | 209.5 | 114 | 297.12 | 0 | 66.7 | NA | NA | NA |
| DBP | Sodium in Urine | MR Egger | 100 | -0.015659491 | 0.005427847 | 0.004813082 | 209.5 | 114 | NA | NA | NA | 0.00392 | 0.001607 | 0.0165 |
| DBP | Sodium in Urine | Weighted median | 100 | -0.003866485 | 0.001637991 | 0.018250132 | 209.5 | 114 | NA | NA | NA | NA | NA | NA |
| DBP | Sodium in Urine | Weighted mode | 100 | -0.007288571 | 0.004069315 | 0.076332282 | 209.5 | 114 | NA | NA | NA | NA | NA | NA |
| HbA1c | Sodium in Urine | Inverse variance weighted | 16 | 0.053946455 | 0.022848485 | 0.018223188 | 64.7 | 30.1 | 18.67 | 0.2289 | 19.7 | NA | NA | NA |
| HbA1c | Sodium in Urine | MR Egger | 16 | 0.135028231 | 0.050625036 | 0.018399538 | 64.7 | 30.1 | NA | NA | NA | -0.003109 | 0.00176 | 0.099 |
| HbA1c | Sodium in Urine | Weighted median | 16 | 0.091173718 | 0.029040531 | 0.001692171 | 64.7 | 30.1 | NA | NA | NA | NA | NA | NA |
| HbA1c | Sodium in Urine | Weighted mode | 16 | 0.088928943 | 0.031641893 | 0.013178005 | 64.7 | 30.1 | NA | NA | NA | NA | NA | NA |
| HDL | Sodium in Urine | Inverse variance weighted | 100 | -0.004055798 | 0.006058485 | 0.50321429 | 193.1 | 59.2 | 166.45 | 0 | 40.5 | NA | NA | NA |
| HDL | Sodium in Urine | MR Egger | 100 | 0.018507158 | 0.011707531 | 0.117148144 | 193.1 | 59.2 | NA | NA | NA | -0.001652 | 0.000739 | 0.0276 |

| | | | | | | | | | | | | | | |
|---|---|---|---|---|---|---|---|---|---|---|---|---|---|---|
| HDL | Sodium in Urine | Weighted median | 100 | 0.007763754 | 0.007709364 | 0.313908372 | 193.1 | 59.2 | NA | NA | NA | NA | NA | NA |
| HDL | Sodium in Urine | Weighted mode | 100 | 0.005364154 | 0.009089629 | 0.556441419 | 193.1 | 59.2 | NA | NA | NA | NA | NA | NA |
| LDL | Sodium in Urine | Inverse variance weighted | 99 | -0.012592484 | 0.00586787 | 0.031872493 | 208.9 | 56.4 | 197.62 | 0 | 50.4 | NA | NA | NA |
| LDL | Sodium in Urine | MR Egger | 99 | -0.017489459 | 0.010273487 | 0.091885423 | 208.9 | 56.4 | NA | NA | NA | 0.000429 | 0.000737 | 0.5621 |
| LDL | Sodium in Urine | Weighted median | 99 | -0.012299667 | 0.006916329 | 0.075346091 | 208.9 | 56.4 | NA | NA | NA | NA | NA | NA |
| LDL | Sodium in Urine | Weighted mode | 99 | -0.011980862 | 0.008119719 | 0.143276766 | 208.9 | 56.4 | NA | NA | NA | NA | NA | NA |
| MetS | Sodium in Urine | Inverse variance weighted | 96 | -0.009762049 | 0.007061889 | 0.166862257 | 97.8 | 41 | 337.56 | 0 | 71.9 | NA | NA | NA |
| MetS | Sodium in Urine | MR Egger | 96 | 0.027724583 | 0.015950517 | 0.085456846 | 97.8 | 41 | NA | NA | NA | -0.003483 | 0.001338 | 0.0107 |
| MetS | Sodium in Urine | Weighted median | 96 | 0.001641185 | 0.005976547 | 0.783620349 | 97.8 | 41 | NA | NA | NA | NA | NA | NA |
| MetS | Sodium in Urine | Weighted mode | 96 | 0.003618474 | 0.01033695 | 0.7270743 | 97.8 | 41 | NA | NA | NA | NA | NA | NA |
| SBP | Sodium in Urine | Inverse variance weighted | 100 | -0.003478329 | 0.000946968 | 0.000239606 | 189.9 | 103.5 | 277.69 | 0 | 64.3 | NA | NA | NA |
| SBP | Sodium in Urine | MR Egger | 100 | -0.004766046 | 0.003142807 | 0.132613844 | 189.9 | 103.5 | NA | NA | NA | 0.00065 | 0.001513 | 0.6682 |

| | | | | | | | | | | | | | | |
|---|---|---|---|---|---|---|---|---|---|---|---|---|---|---|
| SBP | Sodium in Urine | Weighted median | 100 | -0.004092914 | 0.001001853 | 4.40E-05 | 189.9 | 103.5 | NA | NA | NA | NA | NA | NA |
| SBP | Sodium in Urine | Weighted mode | 100 | -0.005642877 | 0.002612036 | 0.033159966 | 189.9 | 103.5 | NA | NA | NA | NA | NA | NA |
| T2D | Sodium in Urine | Inverse variance weighted | 100 | 0.007541065 | 0.005704701 | 0.186200251 | 89.6 | 38.5 | 271.95 | 0 | 63.6 | NA | NA | NA |
| T2D | Sodium in Urine | MR Egger | 100 | 0.002508628 | 0.013660739 | 0.854677017 | 89.6 | 38.5 | NA | NA | NA | 0.000446 | 0.001099 | 0.6858 |
| T2D | Sodium in Urine | Weighted median | 100 | 0.001311077 | 0.005791358 | 0.820901939 | 89.6 | 38.5 | NA | NA | NA | NA | NA | NA |
| T2D | Sodium in Urine | Weighted mode | 100 | -0.006581283 | 0.006932161 | 0.344737177 | 89.6 | 38.5 | NA | NA | NA | NA | NA | NA |
| TG | Sodium in Urine | Inverse variance weighted | 99 | 0.044438245 | 0.010499664 | 2.31E-05 | 150.5 | 43 | 348.14 | 0 | 71.9 | NA | NA | NA |
| TG | Sodium in Urine | MR Egger | 99 | 0.029892598 | 0.019223144 | 0.123195314 | 150.5 | 43 | NA | NA | NA | 0.000939 | 0.001039 | 0.3684 |
| TG | Sodium in Urine | Weighted median | 99 | 0.033864173 | 0.011169712 | 0.002431118 | 150.5 | 43 | NA | NA | NA | NA | NA | NA |
| TG | Sodium in Urine | Weighted mode | 99 | -0.003951831 | 0.013361723 | 0.768040991 | 150.5 | 43 | NA | NA | NA | NA | NA | NA |
| BMI | Tea Consumption | Inverse variance weighted | 100 | 1.02027163 | 4.275073791 | 0.811372407 | 150.7 | 85.3 | 126.79 | 0.0313 | 21.9 | NA | NA | NA |
| BMI | Tea Consumption | MR Egger | 100 | 13.06121869 | 11.77637839 | 0.270100001 | 150.7 | 85.3 | NA | NA | NA | -0.360313 | 0.328407 | 0.2753 |
| BMI | Tea Consumption | Weighted median | 100 | 4.132111803 | 6.084599978 | 0.497068259 | 150.7 | 85.3 | NA | NA | NA | NA | NA | NA |

| | | | | | | | | | | | | | | |
|---|---|---|---|---|---|---|---|---|---|---|---|---|---|---|
| BMI | Tea Consumption | Weighted mode | 100 | -1.003777864 | 19.50062345 | 0.959051423 | 150.7 | 85.3 | NA | NA | NA | NA | NA | NA |
| DBP | Tea Consumption | Inverse variance weighted | 100 | 0.383494973 | 0.412782491 | 0.352863929 | 209.5 | 114 | 128.76 | 0.0238 | 23.1 | NA | NA | NA |
| DBP | Tea Consumption | MR Egger | 100 | 1.012199179 | 1.424505721 | 0.479043502 | 209.5 | 114 | NA | NA | NA | -0.194548 | 0.421736 | 0.6456 |
| DBP | Tea Consumption | Weighted median | 100 | 0.405456325 | 0.556346605 | 0.466133998 | 209.5 | 114 | NA | NA | NA | NA | NA | NA |
| DBP | Tea Consumption | Weighted mode | 100 | -0.384487103 | 1.274721445 | 0.763571067 | 209.5 | 114 | NA | NA | NA | NA | NA | NA |
| HbA1c | Tea Consumption | Inverse variance weighted | 16 | 7.925729748 | 7.956944056 | 0.319212686 | 64.7 | 30.1 | 5.19 | 0.9904 | 0 | NA | NA | NA |
| HbA1c | Tea Consumption | MR Egger | 16 | 26.65172242 | 18.86307537 | 0.17952894 | 64.7 | 30.1 | NA | NA | NA | -0.717001 | 0.654847 | 0.292 |
| HbA1c | Tea Consumption | Weighted median | 16 | 16.10991962 | 10.71536614 | 0.132725388 | 64.7 | 30.1 | NA | NA | NA | NA | NA | NA |
| HbA1c | Tea Consumption | Weighted mode | 16 | 16.74148351 | 14.01707627 | 0.250881187 | 64.7 | 30.1 | NA | NA | NA | NA | NA | NA |
| HDL | Tea Consumption | Inverse variance weighted | 100 | -2.689029899 | 1.814106078 | 0.138263353 | 193.1 | 59.2 | 98.77 | 0.4876 | 0 | NA | NA | NA |
| HDL | Tea Consumption | MR Egger | 100 | 2.285297723 | 3.583317454 | 0.525117049 | 193.1 | 59.2 | NA | NA | NA | -0.363306 | 0.225695 | 0.1107 |
| HDL | Tea Consumption | Weighted median | 100 | 0.379498411 | 2.784046135 | 0.891574776 | 193.1 | 59.2 | NA | NA | NA | NA | NA | NA |
| HDL | Tea Consumption | Weighted mode | 100 | -0.865406186 | 3.351197078 | 0.796759379 | 193.1 | 59.2 | NA | NA | NA | NA | NA | NA |

| | | | | | | | | | | | | | | |
|---|---|---|---|---|---|---|---|---|---|---|---|---|---|---|
| LDL | Tea Consumption | Inverse variance weighted | 100 | -0.025146195 | 1.805583286 | 0.988888293 | 207.8 | 56.4 | 126.56 | 0.0323 | 21.8 | NA | NA | NA |
| LDL | Tea Consumption | MR Egger | 100 | -0.217158096 | 3.166040814 | 0.945455896 | 207.8 | 56.4 | NA | NA | NA | 0.016783 | 0.226756 | 0.9412 |
| LDL | Tea Consumption | Weighted median | 100 | -0.185317631 | 2.65667437 | 0.944388272 | 207.8 | 56.4 | NA | NA | NA | NA | NA | NA |
| LDL | Tea Consumption | Weighted mode | 100 | -0.902142571 | 3.417997409 | 0.792375614 | 207.8 | 56.4 | NA | NA | NA | NA | NA | NA |
| MetS | Tea Consumption | Inverse variance weighted | 96 | -2.077424983 | 1.661151775 | 0.211082962 | 97.8 | 41 | 124.1 | 0.0242 | 23.4 | NA | NA | NA |
| MetS | Tea Consumption | MR Egger | 96 | 4.613007755 | 3.816416443 | 0.229799276 | 97.8 | 41 | NA | NA | NA | -0.620694 | 0.319815 | 0.0553 |
| MetS | Tea Consumption | Weighted median | 96 | -1.886992892 | 2.218871368 | 0.395086535 | 97.8 | 41 | NA | NA | NA | NA | NA | NA |
| MetS | Tea Consumption | Weighted mode | 96 | -0.776129066 | 3.764001188 | 0.837077604 | 97.8 | 41 | NA | NA | NA | NA | NA | NA |
| SBP | Tea Consumption | Inverse variance weighted | 100 | -0.109320407 | 0.256746464 | 0.670259936 | 189.9 | 103.5 | 136.04 | 0.008 | 27.2 | NA | NA | NA |
| SBP | Tea Consumption | MR Egger | 100 | -1.034848012 | 0.846657431 | 0.224535368 | 189.9 | 103.5 | NA | NA | NA | 0.467579 | 0.407658 | 0.2542 |
| SBP | Tea Consumption | Weighted median | 100 | -0.534585855 | 0.329797571 | 0.105028049 | 189.9 | 103.5 | NA | NA | NA | NA | NA | NA |

| | | | | | | | | | | | | | | |
|---|---|---|---|---|---|---|---|---|---|---|---|---|---|---|
| SBP | Tea Consumption | Weighted mode | 100 | -0.841448639 | 0.78007444 | 0.283352735 | 189.9 | 103.5 | NA | NA | NA | NA | NA | NA |
| T2D | Tea Consumption | Inverse variance weighted | 100 | 0.844891355 | 1.380948017 | 0.54065698 | 89.6 | 38.5 | 106.05 | 0.2957 | 6.7 | NA | NA | NA |
| T2D | Tea Consumption | MR Egger | 100 | 4.6585545 | 3.284617941 | 0.159277101 | 89.6 | 38.5 | NA | NA | NA | -0.337656 | 0.264044 | 0.204 |
| T2D | Tea Consumption | Weighted median | 100 | 2.297069194 | 2.335239055 | 0.325285253 | 89.6 | 38.5 | NA | NA | NA | NA | NA | NA |
| T2D | Tea Consumption | Weighted mode | 100 | 2.382727821 | 2.696525493 | 0.379037386 | 89.6 | 38.5 | NA | NA | NA | NA | NA | NA |
| TG | Tea Consumption | Inverse variance weighted | 100 | 0.063423734 | 2.230986318 | 0.977320344 | 149.4 | 43 | 106.18 | 0.2926 | 6.8 | NA | NA | NA |
| TG | Tea Consumption | MR Egger | 100 | -1.033028611 | 4.088622107 | 0.801060655 | 149.4 | 43 | NA | NA | NA | 0.070394 | 0.219546 | 0.7492 |
| TG | Tea Consumption | Weighted median | 100 | 0.453190213 | 3.493583106 | 0.896787367 | 149.4 | 43 | NA | NA | NA | NA | NA | NA |
| TG | Tea Consumption | Weighted mode | 100 | 2.491301557 | 5.329040569 | 0.641172943 | 149.4 | 43 | NA | NA | NA | NA | NA | NA |
| BMI | Testosterone | Inverse variance weighted | 91 | -0.065465261 | 0.022351405 | 0.003401525 | 154 | 85.3 | 248.79 | 0 | 63.8 | NA | NA | NA |
| BMI | Testosterone | MR Egger | 91 | 0.036644137 | 0.059305009 | 0.538223938 | 154 | 85.3 | NA | NA | NA | -0.003122 | 0.001683 | 0.0669 |
| BMI | Testosterone | Weighted median | 91 | -0.026359395 | 0.024152068 | 0.275100053 | 154 | 85.3 | NA | NA | NA | NA | NA | NA |
| BMI | Testosterone | Weighted mode | 91 | 0.010927689 | 0.032755437 | 0.739446507 | 154 | 85.3 | NA | NA | NA | NA | NA | NA |

| | | | | | | | | | | | | | | |
|---|---|---|---|---|---|---|---|---|---|---|---|---|---|---|
| DBP | Testosterone | Inverse variance weighted | 100 | 0.002309642 | 0.002876231 | 0.421969096 | 209.5 | 114 | 531.29 | 0 | 81.4 | NA | NA | NA |
| DBP | Testosterone | MR Egger | 100 | 0.012237491 | 0.009879817 | 0.21843922 | 209.5 | 114 | NA | NA | NA | -0.003072 | 0.002925 | 0.2962 |
| DBP | Testosterone | Weighted median | 100 | 0.001198798 | 0.002260154 | 0.59583095 | 209.5 | 114 | NA | NA | NA | NA | NA | NA |
| DBP | Testosterone | Weighted mode | 100 | -0.000430365 | 0.005807284 | 0.941074115 | 209.5 | 114 | NA | NA | NA | NA | NA | NA |
| HbA1c | Testosterone | Inverse variance weighted | 16 | -0.043424419 | 0.037847061 | 0.251230519 | 64.7 | 30.1 | 28.94 | 0.0164 | 48.2 | NA | NA | NA |
| HbA1c | Testosterone | MR Egger | 16 | -0.168557638 | 0.085232374 | 0.068001173 | 64.7 | 30.1 | NA | NA | NA | 0.004793 | 0.00296 | 0.1277 |
| HbA1c | Testosterone | Weighted median | 16 | -0.09691875 | 0.042039237 | 0.021142192 | 64.7 | 30.1 | NA | NA | NA | NA | NA | NA |
| HbA1c | Testosterone | Weighted mode | 16 | -0.10698054 | 0.049321007 | 0.046559223 | 64.7 | 30.1 | NA | NA | NA | NA | NA | NA |
| HDL | Testosterone | Inverse variance weighted | 92 | -0.024242907 | 0.010950161 | 0.026833491 | 201.7 | 59.2 | 270.84 | 0 | 66.4 | NA | NA | NA |
| HDL | Testosterone | MR Egger | 92 | 0.003881414 | 0.021284451 | 0.855711015 | 201.7 | 59.2 | NA | NA | NA | -0.002123 | 0.001381 | 0.1278 |
| HDL | Testosterone | Weighted median | 92 | -0.016637566 | 0.010477173 | 0.112290313 | 201.7 | 59.2 | NA | NA | NA | NA | NA | NA |
| HDL | Testosterone | Weighted mode | 92 | -0.015918259 | 0.011314485 | 0.162865592 | 201.7 | 59.2 | NA | NA | NA | NA | NA | NA |

| | | | | | | | | | | | | | | |
|---|---|---|---|---|---|---|---|---|---|---|---|---|---|---|
| LDL | Testosterone | Inverse variance weighted | 93 | 0.004363087 | 0.009250639 | 0.637174865 | 203 | 56.8 | 238.91 | 0 | 61.5 | NA | NA | NA |
| LDL | Testosterone | MR Egger | 93 | -0.001158523 | 0.01610644 | 0.942816115 | 203 | 56.8 | NA | NA | NA | 0.000482 | 0.001148 | 0.6757 |
| LDL | Testosterone | Weighted median | 93 | 0.010960819 | 0.009799781 | 0.2633638 | 203 | 56.8 | NA | NA | NA | NA | NA | NA |
| LDL | Testosterone | Weighted mode | 93 | 0.005132418 | 0.011496381 | 0.656330153 | 203 | 56.8 | NA | NA | NA | NA | NA | NA |
| MetS | Testosterone | Inverse variance weighted | 91 | -0.043974541 | 0.010479746 | 2.71E-05 | 100.4 | 41 | 386.82 | 0 | 76.7 | NA | NA | NA |
| MetS | Testosterone | MR Egger | 91 | 0.003004705 | 0.023619877 | 0.899060472 | 100.4 | 41 | NA | NA | NA | -0.00442 | 0.002001 | 0.0298 |
| MetS | Testosterone | Weighted median | 91 | -0.034325468 | 0.007686925 | 7.99E-06 | 100.4 | 41 | NA | NA | NA | NA | NA | NA |
| MetS | Testosterone | Weighted mode | 91 | -0.032833813 | 0.010758585 | 0.002989041 | 100.4 | 41 | NA | NA | NA | NA | NA | NA |
| SBP | Testosterone | Inverse variance weighted | 100 | 0.000593966 | 0.001829089 | 0.745383076 | 189.9 | 103.5 | 586.83 | 0 | 83.1 | NA | NA | NA |
| SBP | Testosterone | MR Egger | 100 | 0.001427933 | 0.006071628 | 0.814558798 | 189.9 | 103.5 | NA | NA | NA | -0.000421 | 0.002923 | 0.8857 |
| SBP | Testosterone | Weighted median | 100 | 6.01E-05 | 0.001324058 | 0.96377356 | 189.9 | 103.5 | NA | NA | NA | NA | NA | NA |
| SBP | Testosterone | Weighted mode | 100 | -0.000155379 | 0.002573822 | 0.951983616 | 189.9 | 103.5 | NA | NA | NA | NA | NA | NA |
| T2D | Testosterone | Inverse variance weighted | 91 | 0.009574112 | 0.008971488 | 0.285894579 | 88.4 | 38.5 | 310.19 | 0 | 71 | NA | NA | NA |

| | | | | | | | | | | | | | | |
|---|---|---|---|---|---|---|---|---|---|---|---|---|---|---|
| T2D | Testosterone | MR Egger | 91 | 0.007408659 | 0.020805811 | 0.72261883 | 88.4 | 38.5 | NA | NA | NA | 0.000194 | 0.001679 | 0.9083 |
| T2D | Testosterone | Weighted median | 91 | 0.001023838 | 0.009995002 | 0.918411406 | 88.4 | 38.5 | NA | NA | NA | NA | NA | NA |
| T2D | Testosterone | Weighted mode | 91 | -0.007316986 | 0.010055152 | 0.468695141 | 88.4 | 38.5 | NA | NA | NA | NA | NA | NA |
| TG | Testosterone | Inverse variance weighted | 93 | 0.10684884 | 0.016791764 | 1.98E-10 | 143.3 | 43 | 414.94 | 0 | 77.8 | NA | NA | NA |
| TG | Testosterone | MR Egger | 93 | 0.119475815 | 0.03293331 | 0.000471398 | 143.3 | 43 | NA | NA | NA | -0.000764 | 0.001712 | 0.6564 |
| TG | Testosterone | Weighted median | 93 | 0.063317516 | 0.014493695 | 1.25E-05 | 143.3 | 43 | NA | NA | NA | NA | NA | NA |
| TG | Testosterone | Weighted mode | 93 | 0.05856908 | 0.016991235 | 0.000856345 | 143.3 | 43 | NA | NA | NA | NA | NA | NA |
| BMI | Urea | Inverse variance weighted | 96 | -0.157824518 | 0.069250534 | 0.022664877 | 149 | 85.3 | 73.96 | 0.946 | 0 | NA | NA | NA |
| BMI | Urea | MR Egger | 96 | -0.626296143 | 0.198125959 | 0.002115689 | 149 | 85.3 | NA | NA | NA | 0.013743 | 0.005446 | 0.0133 |
| BMI | Urea | Weighted median | 96 | -0.293538005 | 0.107840808 | 0.006489665 | 149 | 85.3 | NA | NA | NA | NA | NA | NA |
| BMI | Urea | Weighted mode | 96 | -0.358106124 | 0.158729168 | 0.026358835 | 149 | 85.3 | NA | NA | NA | NA | NA | NA |
| DBP | Urea | Inverse variance weighted | 96 | -0.004774247 | 0.007823363 | 0.541692873 | 209.9 | 114 | 131.36 | 0.008 | 27.7 | NA | NA | NA |

| | | | | | | | | | | | | | | |
|---|---|---|---|---|---|---|---|---|---|---|---|---|---|---|
| DBP | Urea | MR Egger | 96 | -0.018407704 | 0.028396563 | 0.518412003 | 209.9 | 114 | NA | NA | NA | 0.004148 | 0.008302 | 0.6185 |
| DBP | Urea | Weighted median | 96 | -0.001008821 | 0.01002288 | 0.919826827 | 209.9 | 114 | NA | NA | NA | NA | NA | NA |
| DBP | Urea | Weighted mode | 96 | -0.010700885 | 0.01904883 | 0.575601969 | 209.9 | 114 | NA | NA | NA | NA | NA | NA |
| HbA1c | Urea | Inverse variance weighted | 15 | 0.494135089 | 0.158948107 | 0.0018786 | 63.4 | 30.1 | 12.42 | 0.5725 | 0 | NA | NA | NA |
| HbA1c | Urea | MR Egger | 15 | 0.024033646 | 0.422737696 | 0.955527326 | 63.4 | 30.1 | NA | NA | NA | 0.0164 | 0.013665 | 0.2515 |
| HbA1c | Urea | Weighted median | 15 | 0.500185699 | 0.218770681 | 0.022233964 | 63.4 | 30.1 | NA | NA | NA | NA | NA | NA |
| HbA1c | Urea | Weighted mode | 15 | 0.428378504 | 0.320747833 | 0.203000961 | 63.4 | 30.1 | NA | NA | NA | NA | NA | NA |
| HDL | Urea | Inverse variance weighted | 97 | -0.053228651 | 0.030643116 | 0.082378198 | 196.9 | 59.2 | 82.6 | 0.8333 | 0 | NA | NA | NA |
| HDL | Urea | MR Egger | 97 | -0.080479949 | 0.059528266 | 0.179597492 | 196.9 | 59.2 | NA | NA | NA | 0.00206 | 0.003858 | 0.5946 |
| HDL | Urea | Weighted median | 97 | -0.034325202 | 0.045027475 | 0.44587093 | 196.9 | 59.2 | NA | NA | NA | NA | NA | NA |
| HDL | Urea | Weighted mode | 97 | -0.035054848 | 0.047500391 | 0.462320298 | 196.9 | 59.2 | NA | NA | NA | NA | NA | NA |
| LDL | Urea | Inverse variance weighted | 96 | 0.036786239 | 0.032713708 | 0.260805187 | 206 | 56.4 | 128.23 | 0.0131 | 25.9 | NA | NA | NA |

| | | | | | | | | | | | | | | |
|---|---|---|---|---|---|---|---|---|---|---|---|---|---|---|
| LDL | Urea | MR Egger | 96 | 0.067761739 | 0.058080606 | 0.246289147 | 206 | 56.4 | NA | NA | NA | -0.002686 | 0.004155 | 0.5196 |
| LDL | Urea | Weighted median | 96 | 0.037671055 | 0.0445891 | 0.398195175 | 206 | 56.4 | NA | NA | NA | NA | NA | NA |
| LDL | Urea | Weighted mode | 96 | 0.02279391 | 0.055107803 | 0.680081756 | 206 | 56.4 | NA | NA | NA | NA | NA | NA |
| MetS | Urea | Inverse variance weighted | 92 | -0.025007055 | 0.025719924 | 0.330909581 | 97.5 | 41 | 92.06 | 0.4493 | 1.1 | NA | NA | NA |
| MetS | Urea | MR Egger | 92 | -0.005292876 | 0.060159176 | 0.930087097 | 97.5 | 41 | NA | NA | NA | -0.001812 | 0.004993 | 0.7175 |
| MetS | Urea | Weighted median | 92 | -0.006583247 | 0.038779192 | 0.865197038 | 97.5 | 41 | NA | NA | NA | NA | NA | NA |
| MetS | Urea | Weighted mode | 92 | -0.000792482 | 0.057034043 | 0.988944253 | 97.5 | 41 | NA | NA | NA | NA | NA | NA |
| SBP | Urea | Inverse variance weighted | 99 | -0.00734979 | 0.004657598 | 0.114560676 | 189.4 | 103.5 | 150.82 | 5.00E-04 | 35 | NA | NA | NA |
| SBP | Urea | MR Egger | 99 | 0.010710499 | 0.014364794 | 0.457707501 | 189.4 | 103.5 | NA | NA | NA | -0.009364 | 0.007049 | 0.1871 |
| SBP | Urea | Weighted median | 99 | 0.002373344 | 0.005855137 | 0.685224808 | 189.4 | 103.5 | NA | NA | NA | NA | NA | NA |
| SBP | Urea | Weighted mode | 99 | 0.003393179 | 0.009937184 | 0.733486258 | 189.4 | 103.5 | NA | NA | NA | NA | NA | NA |
| T2D | Urea | Inverse variance weighted | 99 | -0.055226263 | 0.024936577 | 0.026782796 | 90.1 | 38.5 | 111.3 | 0.1692 | 12 | NA | NA | NA |

| | | | | | | | | | | | | | | |
|---|---|---|---|---|---|---|---|---|---|---|---|---|---|---|
| T2D | Urea | MR Egger | 99 | -0.079724148 | 0.059368752 | 0.182449954 | 90.1 | 38.5 | NA | NA | NA | 0.002162 | 0.004751 | 0.6501 |
| T2D | Urea | Weighted median | 99 | -0.113653679 | 0.03923289 | 0.003768723 | 90.1 | 38.5 | NA | NA | NA | NA | NA | NA |
| T2D | Urea | Weighted mode | 99 | -0.112867711 | 0.049033976 | 0.023461689 | 90.1 | 38.5 | NA | NA | NA | NA | NA | NA |
| TG | Urea | Inverse variance weighted | 93 | -0.000386078 | 0.038948213 | 0.99209103 | 155.9 | 43 | 101.69 | 0.2298 | 9.5 | NA | NA | NA |
| TG | Urea | MR Egger | 93 | -0.005550345 | 0.06834891 | 0.935456422 | 155.9 | 43 | NA | NA | NA | 0.000352 | 0.003818 | 0.9268 |
| TG | Urea | Weighted median | 93 | 0.041240886 | 0.062890987 | 0.511983753 | 155.9 | 43 | NA | NA | NA | NA | NA | NA |
| TG | Urea | Weighted mode | 93 | 0.040877586 | 0.068407648 | 0.551602425 | 155.9 | 43 | NA | NA | NA | NA | NA | NA |
| BMI | Uric Acid | Inverse variance weighted | 75 | -0.115728072 | 0.035295487 | 0.001042366 | 156.7 | 85.3 | 191.72 | 0 | 61.4 | NA | NA | NA |
| BMI | Uric Acid | MR Egger | 75 | -0.065665437 | 0.102005366 | 0.521758225 | 156.7 | 85.3 | NA | NA | NA | -0.001501 | 0.002868 | 0.6022 |
| BMI | Uric Acid | Weighted median | 75 | -0.127904175 | 0.03725807 | 0.000597106 | 156.7 | 85.3 | NA | NA | NA | NA | NA | NA |
| BMI | Uric Acid | Weighted mode | 75 | -0.073684222 | 0.054767651 | 0.182605402 | 156.7 | 85.3 | NA | NA | NA | NA | NA | NA |

| | | | | | | | | | | | | | | |
|---|---|---|---|---|---|---|---|---|---|---|---|---|---|---|
| DBP | Uric Acid | Inverse variance weighted | 79 | -0.006707225 | 0.004389591 | 0.126516522 | 197 | 114 | 393.53 | 0 | 80.2 | NA | NA | NA |
| DBP | Uric Acid | MR Egger | 79 | 0.018837038 | 0.015320132 | 0.22260537 | 197 | 114 | NA | NA | NA | -0.007853 | 0.004518 | 0.0861 |
| DBP | Uric Acid | Weighted median | 79 | -0.007077426 | 0.003380726 | 0.036307829 | 197 | 114 | NA | NA | NA | NA | NA | NA |
| DBP | Uric Acid | Weighted mode | 79 | -0.010113927 | 0.007592928 | 0.186732902 | 197 | 114 | NA | NA | NA | NA | NA | NA |
| HbA1c | Uric Acid | Inverse variance weighted | 10 | 0.014344603 | 0.05740063 | 0.802662186 | 54.8 | 30.1 | 9.49 | 0.3938 | 5.1 | NA | NA | NA |
| HbA1c | Uric Acid | MR Egger | 10 | 0.195758832 | 0.210685583 | 0.379992343 | 54.8 | 30.1 | NA | NA | NA | -0.005339 | 0.00596 | 0.3965 |
| HbA1c | Uric Acid | Weighted median | 10 | -0.018548389 | 0.072224924 | 0.797322176 | 54.8 | 30.1 | NA | NA | NA | NA | NA | NA |
| HbA1c | Uric Acid | Weighted mode | 10 | -0.044225562 | 0.095872782 | 0.655533109 | 54.8 | 30.1 | NA | NA | NA | NA | NA | NA |
| HDL | Uric Acid | Inverse variance weighted | 87 | 0.029280658 | 0.012012201 | 0.01478609 | 173.5 | 59.2 | 146.65 | 1.00E-04 | 41.4 | NA | NA | NA |
| HDL | Uric Acid | MR Egger | 87 | 0.005522648 | 0.025898721 | 0.83165024 | 173.5 | 59.2 | NA | NA | NA | 0.001642 | 0.001586 | 0.3034 |
| HDL | Uric Acid | Weighted median | 87 | 0.024306563 | 0.015016514 | 0.105521399 | 173.5 | 59.2 | NA | NA | NA | NA | NA | NA |
| HDL | Uric Acid | Weighted mode | 87 | 0.017380455 | 0.01937813 | 0.372270413 | 173.5 | 59.2 | NA | NA | NA | NA | NA | NA |
| LDL | Uric Acid | Inverse variance weighted | 77 | 0.005488579 | 0.011549441 | 0.63462684 | 209.8 | 56.8 | 143.25 | 0 | 46.9 | NA | NA | NA |

| | | | | | | | | | | | | | | |
|---|---|---|---|---|---|---|---|---|---|---|---|---|---|---|
| LDL | Uric Acid | MR Egger | 77 | 0.006708751 | 0.019830145 | 0.73607427 | 209.8 | 56.8 | NA | NA | NA | -0.000109 | 0.001429 | 0.9397 |
| LDL | Uric Acid | Weighted median | 77 | 0.017997775 | 0.015053631 | 0.231861666 | 209.8 | 56.8 | NA | NA | NA | NA | NA | NA |
| LDL | Uric Acid | Weighted mode | 77 | 0.010291118 | 0.016005392 | 0.522173459 | 209.8 | 56.8 | NA | NA | NA | NA | NA | NA |
| MetS | Uric Acid | Inverse variance weighted | 66 | 0.037238327 | 0.014089406 | 0.008217459 | 98.6 | 41 | 210.93 | 0 | 69.2 | NA | NA | NA |
| MetS | Uric Acid | MR Egger | 66 | -0.033801191 | 0.032698521 | 0.305156435 | 98.6 | 41 | NA | NA | NA | 0.006511 | 0.002725 | 0.0198 |
| MetS | Uric Acid | Weighted median | 66 | 0.014520813 | 0.013333293 | 0.276125536 | 98.6 | 41 | NA | NA | NA | NA | NA | NA |
| MetS | Uric Acid | Weighted mode | 66 | 0.002719416 | 0.015218505 | 0.858735795 | 98.6 | 41 | NA | NA | NA | NA | NA | NA |
| SBP | Uric Acid | Inverse variance weighted | 85 | -0.004594299 | 0.002554599 | 0.072106933 | 182.5 | 103.5 | 467.85 | 0 | 82 | NA | NA | NA |
| SBP | Uric Acid | MR Egger | 85 | 0.000739518 | 0.007566952 | 0.922382368 | 182.5 | 103.5 | NA | NA | NA | -0.002829 | 0.003776 | 0.4559 |
| SBP | Uric Acid | Weighted median | 85 | -0.005274108 | 0.001983519 | 0.007838111 | 182.5 | 103.5 | NA | NA | NA | NA | NA | NA |
| SBP | Uric Acid | Weighted mode | 85 | -0.008393874 | 0.002930212 | 0.005273066 | 182.5 | 103.5 | NA | NA | NA | NA | NA | NA |
| T2D | Uric Acid | Inverse variance weighted | 86 | 0.042257723 | 0.012715751 | 0.000889725 | 92.5 | 38.6 | 244.29 | 0 | 65.2 | NA | NA | NA |
| T2D | Uric Acid | MR Egger | 86 | 0.060321554 | 0.036655505 | 0.103576068 | 92.5 | 38.6 | NA | NA | NA | -0.001439 | 0.002736 | 0.6005 |

| | | | | | | | | | | | | | | |
|---|---|---|---|---|---|---|---|---|---|---|---|---|---|---|
| T2D | Uric Acid | Weighted median | 86 | 0.030690321 | 0.013177589 | 0.019860203 | 92.5 | 38.6 | NA | NA | NA | NA | NA | NA |
| T2D | Uric Acid | Weighted mode | 86 | 0.032551661 | 0.019205562 | 0.093753619 | 92.5 | 38.6 | NA | NA | NA | NA | NA | NA |
| TG | Uric Acid | Inverse variance weighted | 76 | -0.040656142 | 0.018934332 | 0.031775913 | 159.8 | 43 | 242.98 | 0 | 69.1 | NA | NA | NA |
| TG | Uric Acid | MR Egger | 76 | -0.000341237 | 0.032252149 | 0.99158679 | 159.8 | 43 | NA | NA | NA | -0.002847 | 0.001853 | 0.1286 |
| TG | Uric Acid | Weighted median | 76 | -0.000399035 | 0.017945295 | 0.98225956 | 159.8 | 43 | NA | NA | NA | NA | NA | NA |
| TG | Uric Acid | Weighted mode | 76 | 0.006345159 | 0.018364955 | 0.730682964 | 159.8 | 43 | NA | NA | NA | NA | NA | NA |
| BMI | Urinary Albumin Excretion | Inverse variance weighted | 98 | -0.015899866 | 0.012229816 | 0.193569983 | 151.4 | 85.3 | 302.98 | 0 | 68 | NA | NA | NA |
| BMI | Urinary Albumin Excretion | MR Egger | 98 | -0.073040869 | 0.03319079 | 0.030159658 | 151.4 | 85.3 | NA | NA | NA | 0.001717 | 0.000929 | 0.0676 |
| BMI | Urinary Albumin Excretion | Weighted median | 98 | -0.004949505 | 0.012490525 | 0.691912653 | 151.4 | 85.3 | NA | NA | NA | NA | NA | NA |
| BMI | Urinary Albumin Excretion | Weighted mode | 98 | -0.000247319 | 0.028964175 | 0.993204642 | 151.4 | 85.3 | NA | NA | NA | NA | NA | NA |
| DBP | Urinary Albumin Excretion | Inverse variance weighted | 99 | -0.005736305 | 0.001300804 | 1.03E-05 | 210 | 114 | 380.02 | 0 | 74.2 | NA | NA | NA |
| DBP | Urinary Albumin Excretion | MR Egger | 99 | -0.007261606 | 0.004474264 | 0.107841 | 210 | 114 | NA | NA | NA | 0.000472 | 0.001325 | 0.7223 |

| | | | | | | | | | | | | | | |
|---|---|---|---|---|---|---|---|---|---|---|---|---|---|---|
| DBP | Urinary Albumin Excretion | Weighted median | 99 | -0.006419247 | 0.001122515 | 1.07E-08 | 210 | 114 | NA | NA | NA | NA | NA | NA |
| DBP | Urinary Albumin Excretion | Weighted mode | 99 | -0.007818229 | 0.002038308 | 0.000221615 | 210 | 114 | NA | NA | NA | NA | NA | NA |
| HbA1c | Urinary Albumin Excretion | Inverse variance weighted | 16 | 0.020353546 | 0.01442131 | 0.158140802 | 64.7 | 30.1 | 12.75 | 0.6219 | 0 | NA | NA | NA |
| HbA1c | Urinary Albumin Excretion | MR Egger | 16 | 0.023522798 | 0.034131523 | 0.501972567 | 64.7 | 30.1 | NA | NA | NA | -0.000122 | 0.001187 | 0.9199 |
| HbA1c | Urinary Albumin Excretion | Weighted median | 16 | 0.020445468 | 0.021102 | 0.332601215 | 64.7 | 30.1 | NA | NA | NA | NA | NA | NA |
| HbA1c | Urinary Albumin Excretion | Weighted mode | 16 | 0.02111981 | 0.024121121 | 0.395066828 | 64.7 | 30.1 | NA | NA | NA | NA | NA | NA |
| HDL | Urinary Albumin Excretion | Inverse variance weighted | 99 | 0.007754416 | 0.004908005 | 0.114117664 | 193.7 | 59.2 | 216.3 | 0 | 54.7 | NA | NA | NA |
| HDL | Urinary Albumin Excretion | MR Egger | 99 | 0.003403495 | 0.009673301 | 0.725718318 | 193.7 | 59.2 | NA | NA | NA | 0.00032 | 0.000611 | 0.6024 |
| HDL | Urinary Albumin Excretion | Weighted median | 99 | 0.006313628 | 0.005998111 | 0.292523063 | 193.7 | 59.2 | NA | NA | NA | NA | NA | NA |
| HDL | Urinary Albumin Excretion | Weighted mode | 99 | 0.002289377 | 0.00605782 | 0.706306684 | 193.7 | 59.2 | NA | NA | NA | NA | NA | NA |
| LDL | Urinary Albumin Excretion | Inverse variance weighted | 99 | -0.017358429 | 0.003999845 | 1.43E-05 | 207.7 | 56.4 | 184.21 | 0 | 46.8 | NA | NA | NA |
| LDL | Urinary Albumin Excretion | MR Egger | 99 | -0.018986505 | 0.006996243 | 0.007872196 | 207.7 | 56.4 | NA | NA | NA | 0.000142 | 0.000501 | 0.7768 |
| LDL | Urinary Albumin Excretion | Weighted median | 99 | -0.020203964 | 0.004876624 | 3.43E-05 | 207.7 | 56.4 | NA | NA | NA | NA | NA | NA |

| | | | | | | | | | | | | | | |
|---|---|---|---|---|---|---|---|---|---|---|---|---|---|---|
| LDL | Urinary Albumin Excretion | Weighted mode | 99 | -0.021065333 | 0.005194633 | 0.000100577 | 207.7 | 56.4 | NA | NA | NA | NA | NA | NA |
| MetS | Urinary Albumin Excretion | Inverse variance weighted | 96 | 0.014223408 | 0.004223372 | 0.000757747 | 97.8 | 41 | 243.24 | 0 | 60.9 | NA | NA | NA |
| MetS | Urinary Albumin Excretion | MR Egger | 96 | 0.009559829 | 0.009864632 | 0.334981501 | 97.8 | 41 | NA | NA | NA | 0.000433 | 0.000827 | 0.6018 |
| MetS | Urinary Albumin Excretion | Weighted median | 96 | 0.012207157 | 0.004606338 | 0.008047311 | 97.8 | 41 | NA | NA | NA | NA | NA | NA |
| MetS | Urinary Albumin Excretion | Weighted mode | 96 | 0.012681698 | 0.009223891 | 0.17240424 | 97.8 | 41 | NA | NA | NA | NA | NA | NA |
| SBP | Urinary Albumin Excretion | Inverse variance weighted | 100 | -0.004707544 | 0.000726229 | 9.04E-11 | 189.9 | 103.5 | 329.06 | 0 | 69.9 | NA | NA | NA |
| SBP | Urinary Albumin Excretion | MR Egger | 100 | -0.003122138 | 0.002406633 | 0.197571781 | 189.9 | 103.5 | NA | NA | NA | -0.000801 | 0.001158 | 0.4911 |
| SBP | Urinary Albumin Excretion | Weighted median | 100 | -0.005158872 | 0.000685208 | 5.12E-14 | 189.9 | 103.5 | NA | NA | NA | NA | NA | NA |
| SBP | Urinary Albumin Excretion | Weighted mode | 100 | -0.005905265 | 0.001338312 | 2.61E-05 | 189.9 | 103.5 | NA | NA | NA | NA | NA | NA |
| T2D | Urinary Albumin Excretion | Inverse variance weighted | 100 | -0.01749938 | 0.004043304 | 1.50E-05 | 89.6 | 38.5 | 275.22 | 0 | 64 | NA | NA | NA |
| T2D | Urinary Albumin Excretion | MR Egger | 100 | -0.020757134 | 0.009685777 | 0.034584847 | 89.6 | 38.5 | NA | NA | NA | 0.000289 | 0.000779 | 0.7118 |
| T2D | Urinary Albumin Excretion | Weighted median | 100 | -0.022909327 | 0.004064921 | 1.74E-08 | 89.6 | 38.5 | NA | NA | NA | NA | NA | NA |

| | | | | | | | | | | | | | | |
|---|---|---|---|---|---|---|---|---|---|---|---|---|---|---|
| T2D | Urinary Albumin Excretion | Weighted mode | 100 | -0.023527274 | 0.005194062 | 1.65E-05 | 89.6 | 38.5 | NA | NA | NA | NA | NA | NA |
| TG | Urinary Albumin Excretion | Inverse variance weighted | 100 | -0.032048104 | 0.00713657 | 7.10E-06 | 149.4 | 43 | 328.22 | 0 | 69.8 | NA | NA | NA |
| TG | Urinary Albumin Excretion | MR Egger | 100 | -0.030000094 | 0.013093002 | 0.02408673 | 149.4 | 43 | NA | NA | NA | -0.000131 | 0.000703 | 0.8521 |
| TG | Urinary Albumin Excretion | Weighted median | 100 | -0.033375699 | 0.007601584 | 1.13E-05 | 149.4 | 43 | NA | NA | NA | NA | NA | NA |
| TG | Urinary Albumin Excretion | Weighted mode | 100 | -0.033990123 | 0.009710408 | 0.000698178 | 149.4 | 43 | NA | NA | NA | NA | NA | NA |
| BMI | Water Consumption | Inverse variance weighted | 100 | -0.026843178 | 0.015525394 | 0.08381173 | 150.7 | 85.3 | 427.87 | 0 | 76.9 | NA | NA | NA |
| BMI | Water Consumption | MR Egger | 100 | -0.116356557 | 0.041985919 | 0.006681158 | 150.7 | 85.3 | NA | NA | NA | 0.002677 | 0.00117 | 0.0243 |
| BMI | Water Consumption | Weighted median | 100 | -0.039527658 | 0.013922079 | 0.004522587 | 150.7 | 85.3 | NA | NA | NA | NA | NA | NA |
| BMI | Water Consumption | Weighted mode | 100 | -0.046382826 | 0.040924207 | 0.259790432 | 150.7 | 85.3 | NA | NA | NA | NA | NA | NA |
| DBP | Water Consumption | Inverse variance weighted | 100 | -0.004874995 | 0.001200644 | 4.90E-05 | 209.5 | 114 | 278.67 | 0 | 64.5 | NA | NA | NA |
| DBP | Water Consumption | MR Egger | 100 | -0.009093111 | 0.004124168 | 0.02980465 | 209.5 | 114 | NA | NA | NA | 0.001305 | 0.001221 | 0.2877 |

| | | | | | | | | | | | | | | |
|---|---|---|---|---|---|---|---|---|---|---|---|---|---|---|
| DBP | Water Consumption | Weighted median | 100 | -0.005169742 | 0.001225698 | 2.47E-05 | 209.5 | 114 | NA | NA | NA | NA | NA | NA |
| DBP | Water Consumption | Weighted mode | 100 | -0.006459787 | 0.003822886 | 0.094219369 | 209.5 | 114 | NA | NA | NA | NA | NA | NA |
| HbA1c | Water Consumption | Inverse variance weighted | 16 | -0.004919202 | 0.022359736 | 0.825869048 | 64.7 | 30.1 | 30.42 | 0.0105 | 50.7 | NA | NA | NA |
| HbA1c | Water Consumption | MR Egger | 16 | 0.022641177 | 0.054255197 | 0.682781263 | 64.7 | 30.1 | NA | NA | NA | -0.001056 | 0.001884 | 0.5841 |
| HbA1c | Water Consumption | Weighted median | 16 | -0.020600901 | 0.021883369 | 0.346502329 | 64.7 | 30.1 | NA | NA | NA | NA | NA | NA |
| HbA1c | Water Consumption | Weighted mode | 16 | -0.01309075 | 0.025339852 | 0.612962626 | 64.7 | 30.1 | NA | NA | NA | NA | NA | NA |
| HDL | Water Consumption | Inverse variance weighted | 100 | 0.013278106 | 0.005936102 | 0.025296854 | 193.1 | 59.2 | 271.85 | 0 | 63.6 | NA | NA | NA |
| HDL | Water Consumption | MR Egger | 100 | 0.007312571 | 0.011750945 | 0.535192226 | 193.1 | 59.2 | NA | NA | NA | 0.000436 | 0.000741 | 0.5573 |
| HDL | Water Consumption | Weighted median | 100 | 0.007498897 | 0.005601406 | 0.180651261 | 193.1 | 59.2 | NA | NA | NA | NA | NA | NA |
| HDL | Water Consumption | Weighted mode | 100 | 0.00858713 | 0.005497264 | 0.121461807 | 193.1 | 59.2 | NA | NA | NA | NA | NA | NA |
| LDL | Water Consumption | Inverse variance weighted | 100 | -0.012761047 | 0.004911993 | 0.009378582 | 207.8 | 56.4 | 239.48 | 0 | 58.7 | NA | NA | NA |
| LDL | Water Consumption | MR Egger | 100 | -0.017679784 | 0.008606465 | 0.042614223 | 207.8 | 56.4 | NA | NA | NA | 0.000429 | 0.000616 | 0.4875 |

| | | | | | | | | | | | | | | |
|---|---|---|---|---|---|---|---|---|---|---|---|---|---|---|
| LDL | Water Consumption | Weighted median | 100 | -0.010598372 | 0.005714791 | 0.063660368 | 207.8 | 56.4 | NA | NA | NA | NA | NA | NA |
| LDL | Water Consumption | Weighted mode | 100 | -0.003607076 | 0.006179064 | 0.560711095 | 207.8 | 56.4 | NA | NA | NA | NA | NA | NA |
| MetS | Water Consumption | Inverse variance weighted | 96 | 0.017757287 | 0.005045301 | 0.000432248 | 97.8 | 41 | 293.33 | 0 | 67.6 | NA | NA | NA |
| MetS | Water Consumption | MR Egger | 96 | 0.007738008 | 0.011757777 | 0.512070732 | 97.8 | 41 | NA | NA | NA | 0.00093 | 0.000986 | 0.3478 |
| MetS | Water Consumption | Weighted median | 96 | 0.010144638 | 0.004765546 | 0.033275249 | 97.8 | 41 | NA | NA | NA | NA | NA | NA |
| MetS | Water Consumption | Weighted mode | 96 | 0.00632067 | 0.007171161 | 0.380325291 | 97.8 | 41 | NA | NA | NA | NA | NA | NA |
| SBP | Water Consumption | Inverse variance weighted | 100 | -0.001025561 | 0.000695003 | 0.140045579 | 189.9 | 103.5 | 254.95 | 0 | 61.2 | NA | NA | NA |
| SBP | Water Consumption | MR Egger | 100 | -0.00195486 | 0.002306441 | 0.398745134 | 189.9 | 103.5 | NA | NA | NA | 0.000469 | 0.00111 | 0.6734 |
| SBP | Water Consumption | Weighted median | 100 | -0.000794666 | 0.000715608 | 0.266793856 | 189.9 | 103.5 | NA | NA | NA | NA | NA | NA |
| SBP | Water Consumption | Weighted mode | 100 | -0.00311679 | 0.002207933 | 0.161193323 | 189.9 | 103.5 | NA | NA | NA | NA | NA | NA |
| T2D | Water Consumption | Inverse variance weighted | 100 | -0.009761887 | 0.004454911 | 0.028432664 | 89.6 | 38.5 | 282.52 | 0 | 65 | NA | NA | NA |
| T2D | Water Consumption | MR Egger | 100 | -0.011987952 | 0.010675451 | 0.26420407 | 89.6 | 38.5 | NA | NA | NA | 0.000197 | 0.000858 | 0.8188 |

| | | | | | | | | | | | | | | |
|---|---|---|---|---|---|---|---|---|---|---|---|---|---|---|
| T2D | Water Consumption | Weighted median | 100 | -0.006814747 | 0.005246002 | 0.193931479 | 89.6 | 38.5 | NA | NA | NA | NA | NA | NA |
| T2D | Water Consumption | Weighted mode | 100 | -0.005199639 | 0.005486535 | 0.345585152 | 89.6 | 38.5 | NA | NA | NA | NA | NA | NA |
| TG | Water Consumption | Inverse variance weighted | 100 | -0.029164515 | 0.007988902 | 0.000261599 | 149.4 | 43 | 348.41 | 0 | 71.6 | NA | NA | NA |
| TG | Water Consumption | MR Egger | 100 | -0.019188607 | 0.014601779 | 0.19187078 | 149.4 | 43 | NA | NA | NA | -0.00064 | 0.000784 | 0.416 |
| TG | Water Consumption | Weighted median | 100 | -0.016437374 | 0.007905776 | 0.037602636 | 149.4 | 43 | NA | NA | NA | NA | NA | NA |
| TG | Water Consumption | Weighted mode | 100 | -0.013469366 | 0.010881014 | 0.218689017 | 149.4 | 43 | NA | NA | NA | NA | NA | NA |
| BMI | X_11440 | Inverse variance weighted | 45 | -0.001131289 | 0.032419988 | 0.972163617 | 141.3 | 85.3 | 43.48 | 0.4936 | 0 | NA | NA | NA |
| BMI | X_11440 | MR Egger | 45 | 0.054915741 | 0.103148289 | 0.597192243 | 141.3 | 85.3 | NA | NA | NA | -0.001546 | 0.0027 | 0.57 |
| BMI | X_11440 | Weighted median | 45 | -0.0046143 | 0.048598044 | 0.924355923 | 141.3 | 85.3 | NA | NA | NA | NA | NA | NA |
| BMI | X_11440 | Weighted mode | 45 | 0.039707422 | 0.084898564 | 0.642304403 | 141.3 | 85.3 | NA | NA | NA | NA | NA | NA |
| DBP | X_11440 | Inverse variance weighted | 49 | -0.006393175 | 0.003097731 | 0.039034308 | 191.3 | 114 | 39.61 | 0.8003 | 0 | NA | NA | NA |

| | | | | | | | | | | | | | | |
|---|---|---|---|---|---|---|---|---|---|---|---|---|---|---|
| DBP | X_11440 | MR Egger | 49 | -0.014601488 | 0.011755854 | 0.220376259 | 191.3 | 114 | NA | NA | NA | 0.002353 | 0.003252 | 0.4728 |
| DBP | X_11440 | Weighted median | 49 | -0.005285615 | 0.004423616 | 0.232140572 | 191.3 | 114 | NA | NA | NA | NA | NA | NA |
| DBP | X_11440 | Weighted mode | 49 | -0.002569332 | 0.007875826 | 0.745668346 | 191.3 | 114 | NA | NA | NA | NA | NA | NA |
| HbA1c | X_11440 | Inverse variance weighted | 16 | -0.043090034 | 0.047690299 | 0.366240018 | 64.7 | 30.1 | 12.52 | 0.639 | 0 | NA | NA | NA |
| HbA1c | X_11440 | MR Egger | 16 | 0.04439886 | 0.117218413 | 0.710540628 | 64.7 | 30.1 | NA | NA | NA | -0.003237 | 0.003962 | 0.4276 |
| HbA1c | X_11440 | Weighted median | 16 | -0.019294663 | 0.06781541 | 0.776013915 | 64.7 | 30.1 | NA | NA | NA | NA | NA | NA |
| HbA1c | X_11440 | Weighted mode | 16 | 0.020965397 | 0.093439768 | 0.825494412 | 64.7 | 30.1 | NA | NA | NA | NA | NA | NA |
| HDL | X_11440 | Inverse variance weighted | 94 | -0.010641631 | 0.012545963 | 0.396320145 | 193.5 | 59.2 | 113.95 | 0.0692 | 18.4 | NA | NA | NA |
| HDL | X_11440 | MR Egger | 94 | -0.005782994 | 0.026091583 | 0.825083723 | 193.5 | 59.2 | NA | NA | NA | -0.000326 | 0.001531 | 0.832 |
| HDL | X_11440 | Weighted median | 94 | -0.024210902 | 0.018368625 | 0.187484445 | 193.5 | 59.2 | NA | NA | NA | NA | NA | NA |
| HDL | X_11440 | Weighted mode | 94 | -0.027333608 | 0.020756399 | 0.191115904 | 193.5 | 59.2 | NA | NA | NA | NA | NA | NA |
| LDL | X_11440 | Inverse variance weighted | 96 | 0.032550258 | 0.011503086 | 0.004659198 | 205.5 | 56.4 | 129.09 | 0.0115 | 26.4 | NA | NA | NA |

| | | | | | | | | | | | | | | |
|---|---|---|---|---|---|---|---|---|---|---|---|---|---|---|
| LDL | X_11440 | MR Egger | 96 | 0.020910939 | 0.021519624 | 0.333686243 | 205.5 | 56.4 | NA | NA | NA | 0.000929 | 0.00145 | 0.5232 |
| LDL | X_11440 | Weighted median | 96 | 0.022057596 | 0.015831222 | 0.163529969 | 205.5 | 56.4 | NA | NA | NA | NA | NA | NA |
| LDL | X_11440 | Weighted mode | 96 | 0.01758973 | 0.020011475 | 0.381628821 | 205.5 | 56.4 | NA | NA | NA | NA | NA | NA |
| MetS | X_11440 | Inverse variance weighted | 41 | -0.001238209 | 0.014584734 | 0.932342791 | 110.4 | 42.1 | 60.2 | 0.021 | 33.6 | NA | NA | NA |
| MetS | X_11440 | MR Egger | 41 | 0.029540821 | 0.037326911 | 0.433492898 | 110.4 | 42.1 | NA | NA | NA | -0.002685 | 0.002996 | 0.3756 |
| MetS | X_11440 | Weighted median | 41 | -0.008690566 | 0.018589563 | 0.64014432 | 110.4 | 42.1 | NA | NA | NA | NA | NA | NA |
| MetS | X_11440 | Weighted mode | 41 | -0.008007754 | 0.026094493 | 0.760530238 | 110.4 | 42.1 | NA | NA | NA | NA | NA | NA |
| SBP | X_11440 | Inverse variance weighted | 45 | -0.000749373 | 0.001961589 | 0.702444182 | 186.3 | 103.5 | 50.36 | 0.2363 | 12.6 | NA | NA | NA |
| SBP | X_11440 | MR Egger | 45 | -0.006435314 | 0.006257883 | 0.309533819 | 186.3 | 103.5 | NA | NA | NA | 0.002841 | 0.002969 | 0.344 |
| SBP | X_11440 | Weighted median | 45 | -0.003811593 | 0.002631496 | 0.147491015 | 186.3 | 103.5 | NA | NA | NA | NA | NA | NA |
| SBP | X_11440 | Weighted mode | 45 | -0.00456572 | 0.004400524 | 0.305152701 | 186.3 | 103.5 | NA | NA | NA | NA | NA | NA |
| T2D | X_11440 | Inverse variance weighted | 90 | 0.000766009 | 0.008873099 | 0.931204574 | 94.1 | 38.6 | 120.77 | 0.0141 | 26.3 | NA | NA | NA |

| | | | | | | | | | | | | | | |
|---|---|---|---|---|---|---|---|---|---|---|---|---|---|---|
| T2D | X_11440 | MR Egger | 90 | 0.018155455 | 0.021115181 | 0.392219418 | 94.1 | 38.6 | NA | NA | NA | -0.001547 | 0.001704 | 0.3665 |
| T2D | X_11440 | Weighted median | 90 | 0.003799405 | 0.014939818 | 0.799252978 | 94.1 | 38.6 | NA | NA | NA | NA | NA | NA |
| T2D | X_11440 | Weighted mode | 90 | 0.009837515 | 0.015528703 | 0.528028396 | 94.1 | 38.6 | NA | NA | NA | NA | NA | NA |
| TG | X_11440 | Inverse variance weighted | 94 | 0.005043125 | 0.013731587 | 0.713421716 | 155.1 | 43 | 113.64 | 0.0718 | 18.2 | NA | NA | NA |
| TG | X_11440 | MR Egger | 94 | -0.006782223 | 0.025014905 | 0.786900543 | 155.1 | 43 | NA | NA | NA | 0.000744 | 0.001313 | 0.5725 |
| TG | X_11440 | Weighted median | 94 | -0.009333703 | 0.020318333 | 0.645965978 | 155.1 | 43 | NA | NA | NA | NA | NA | NA |
| TG | X_11440 | Weighted mode | 94 | -0.018711606 | 0.022462305 | 0.40696626 | 155.1 | 43 | NA | NA | NA | NA | NA | NA |
| BMI | X_14473 | Inverse variance weighted | 45 | -0.029937518 | 0.030001024 | 0.318335993 | 141.3 | 85.3 | 42.52 | 0.5352 | 0 | NA | NA | NA |
| BMI | X_14473 | MR Egger | 45 | 0.043666718 | 0.095773093 | 0.650727998 | 141.3 | 85.3 | NA | NA | NA | -0.002025 | 0.002502 | 0.4228 |
| BMI | X_14473 | Weighted median | 45 | -0.021053667 | 0.044034754 | 0.632568504 | 141.3 | 85.3 | NA | NA | NA | NA | NA | NA |
| BMI | X_14473 | Weighted mode | 45 | -0.025273404 | 0.070589701 | 0.722030993 | 141.3 | 85.3 | NA | NA | NA | NA | NA | NA |
| DBP | X_14473 | Inverse variance weighted | 49 | -0.00065405 | 0.002853587 | 0.818711226 | 191.3 | 114 | 42.77 | 0.6865 | 0 | NA | NA | NA |

| | | | | | | | | | | | | | | |
|---|---|---|---|---|---|---|---|---|---|---|---|---|---|---|
| DBP | X_14473 | MR Egger | 49 | 0.00265517 | 0.010915028 | 0.808863994 | 191.3 | 114 | NA | NA | NA | -0.000946 | 0.003013 | 0.7548 |
| DBP | X_14473 | Weighted median | 49 | 0 | 0.0043702 | 1 | 191.3 | 114 | NA | NA | NA | NA | NA | NA |
| DBP | X_14473 | Weighted mode | 49 | -0.001451953 | 0.009122676 | 0.874211656 | 191.3 | 114 | NA | NA | NA | NA | NA | NA |
| HbA1c | X_14473 | Inverse variance weighted | 16 | -0.044966167 | 0.043918516 | 0.305904041 | 64.7 | 30.1 | 11.61 | 0.7084 | 0 | NA | NA | NA |
| HbA1c | X_14473 | MR Egger | 16 | 0.051184556 | 0.1093462 | 0.646919762 | 64.7 | 30.1 | NA | NA | NA | -0.003512 | 0.003658 | 0.3533 |
| HbA1c | X_14473 | Weighted median | 16 | -0.007198195 | 0.062967071 | 0.908986654 | 64.7 | 30.1 | NA | NA | NA | NA | NA | NA |
| HbA1c | X_14473 | Weighted mode | 16 | 0.007382263 | 0.087087886 | 0.933566908 | 64.7 | 30.1 | NA | NA | NA | NA | NA | NA |
| HDL | X_14473 | Inverse variance weighted | 94 | 0.025715813 | 0.010449251 | 0.013854269 | 193.5 | 59.2 | 81.98 | 0.786 | 0 | NA | NA | NA |
| HDL | X_14473 | MR Egger | 94 | 0.017403371 | 0.021705234 | 0.424731801 | 193.5 | 59.2 | NA | NA | NA | 0.000555 | 0.00127 | 0.6632 |
| HDL | X_14473 | Weighted median | 94 | -0.002048058 | 0.016254435 | 0.899731984 | 193.5 | 59.2 | NA | NA | NA | NA | NA | NA |
| HDL | X_14473 | Weighted mode | 94 | -0.006799903 | 0.018511829 | 0.714209355 | 193.5 | 59.2 | NA | NA | NA | NA | NA | NA |
| LDL | X_14473 | Inverse variance weighted | 96 | 0.004259686 | 0.009785149 | 0.663329028 | 205.5 | 56.4 | 107.49 | 0.1796 | 11.6 | NA | NA | NA |
| LDL | X_14473 | MR Egger | 96 | 0.017966745 | 0.018416498 | 0.331777199 | 205.5 | 56.4 | NA | NA | NA | -0.001081 | 0.00123 | 0.3817 |

| | | | | | | | | | | | | | | |
|---|---|---|---|---|---|---|---|---|---|---|---|---|---|---|
| LDL | X_14473 | Weighted median | 96 | 0.010379078 | 0.01508317 | 0.491375253 | 205.5 | 56.4 | NA | NA | NA | NA | NA | NA |
| LDL | X_14473 | Weighted mode | 96 | 0.014260162 | 0.018768569 | 0.44926178 | 205.5 | 56.4 | NA | NA | NA | NA | NA | NA |
| MetS | X_14473 | Inverse variance weighted | 41 | 0.005895679 | 0.012361205 | 0.633397615 | 110.4 | 42.1 | 50.81 | 0.1177 | 21.3 | NA | NA | NA |
| MetS | X_14473 | MR Egger | 41 | -0.021979896 | 0.031751958 | 0.492889408 | 110.4 | 42.1 | NA | NA | NA | 0.002431 | 0.00255 | 0.3463 |
| MetS | X_14473 | Weighted median | 41 | -0.010611901 | 0.015822623 | 0.502425041 | 110.4 | 42.1 | NA | NA | NA | NA | NA | NA |
| MetS | X_14473 | Weighted mode | 41 | -0.006057341 | 0.019306603 | 0.755345446 | 110.4 | 42.1 | NA | NA | NA | NA | NA | NA |
| SBP | X_14473 | Inverse variance weighted | 45 | 0.002390173 | 0.001840004 | 0.193942447 | 186.3 | 103.5 | 51.98 | 0.1911 | 15.3 | NA | NA | NA |
| SBP | X_14473 | MR Egger | 45 | 0.008654195 | 0.005873794 | 0.147938051 | 186.3 | 103.5 | NA | NA | NA | -0.003117 | 0.002776 | 0.2678 |
| SBP | X_14473 | Weighted median | 45 | 0.003456891 | 0.002617135 | 0.18654521 | 186.3 | 103.5 | NA | NA | NA | NA | NA | NA |
| SBP | X_14473 | Weighted mode | 45 | 0.00915192 | 0.005538287 | 0.105555656 | 186.3 | 103.5 | NA | NA | NA | NA | NA | NA |
| T2D | X_14473 | Inverse variance weighted | 90 | 0.000804496 | 0.007322307 | 0.912513066 | 94.1 | 38.6 | 96.12 | 0.2845 | 7.4 | NA | NA | NA |
| T2D | X_14473 | MR Egger | 90 | 0.026950596 | 0.017271407 | 0.122250652 | 94.1 | 38.6 | NA | NA | NA | -0.002325 | 0.001394 | 0.0989 |
| T2D | X_14473 | Weighted median | 90 | 0.018592313 | 0.011461419 | 0.104768014 | 94.1 | 38.6 | NA | NA | NA | NA | NA | NA |
| T2D | X_14473 | Weighted mode | 90 | 0.018797471 | 0.014207995 | 0.189217639 | 94.1 | 38.6 | NA | NA | NA | NA | NA | NA |

| | | | | | | | | | | | | | | |
|---|---|---|---|---|---|---|---|---|---|---|---|---|---|---|
| TG | X_14473 | Inverse variance weighted | 94 | -0.014915484 | 0.011492888 | 0.194355657 | 155.1 | 43 | 90.22 | 0.5623 | 0 | NA | NA | NA |
| TG | X_14473 | MR Egger | 94 | -0.025472179 | 0.020910053 | 0.226270965 | 155.1 | 43 | NA | NA | NA | 0.000666 | 0.001101 | 0.5471 |
| TG | X_14473 | Weighted median | 94 | -0.017128274 | 0.017794082 | 0.335757028 | 155.1 | 43 | NA | NA | NA | NA | NA | NA |
| TG | X_14473 | Weighted mode | 94 | -0.015945254 | 0.018140732 | 0.381680157 | 155.1 | 43 | NA | NA | NA | NA | NA | NA |

#### Supplementary Table 7 - Genetic Correlation Analysis Results (LDSC)

| Trait 1 | Trait 2 | rg | SE | P | nSNPs |
|---|---|---|---|---|---|
| Impedance of Left Arm | Impedance of Right Arm | 0.963752391 | 9.73E-05 | 6.19E-83 | 7521790 |
| Creatinine | Albumin | 0.594758013 | 0.000293725 | 8.23E-117 | 7490805 |
| Creatinine | Sodium in Urine | 0.555134801 | 0.000303199 | 0.023465436 | 7525592 |
| Sodium in Urine | Albumin | 0.367295082 | 0.000339835 | 0.034561347 | 7490805 |
| Impedance of Right Arm | Urate | 0.192619443 | 0.000359554 | 4.25E-05 | 7448195 |
| Impedance of Left Arm | Urate | 0.190486497 | 0.000359703 | 7.25E-161 | 7448377 |
| Creatinine | Impedance of Right Arm | -0.171358971 | 0.00035922 | 9.21E-78 | 7522019 |
| Creatinine | Impedance of Left Arm | -0.169005465 | 0.000359364 | 9.30E-119 | 7522218 |
| Creatinine | Urate | -0.155793442 | 0.00036186 | 7.68E-118 | 7451550 |
| Uric Acid | Urate | 0.153037055 | 0.000437205 | 0.086768066 | 5109022 |
| Impedance of Left Arm | Sodium in Urine | -0.145802052 | 0.000360712 | 0.161346521 | 7522220 |
| Impedance of Right Arm | Sodium in Urine | -0.145212484 | 0.000360749 | 7.22E-145 | 7522021 |
| GAMMA-GLUTAMYL TRANSFERASE | Uric Acid | 0.135569255 | 0.000436429 | 5.95E-126 | 5153659 |
| 25-Hydroxyvitamin D | Impedance of Right Arm | 0.123428146 | 0.000375788 | 6.23E-193 | 6973456 |
| 25-Hydroxyvitamin D | Impedance of Left Arm | 0.11994815 | 0.000375944 | 1.19E-79 | 6973633 |
| Cystatin C | Urate | -0.102948043 | 0.000682634 | 4.35E-137 | 2123227 |
| Sodium in Urine | Urate | -0.099370967 | 0.00036452 | 2.68E-16 | 7451551 |
| Coffee | Impedance of Right Arm | 0.079010977 | 0.000363638 | 7.25E-184 | 7515225 |
| Coffee | Impedance of Left Arm | 0.078885023 | 0.000363637 | 6.21E-75 | 7515425 |
| Testosterone | Urate | 0.068477761 | 0.000344656 | 9.33E-128 | 8378923 |
| Impedance of Right Arm | Albumin | -0.062587302 | 0.000364743 | 7.68E-16 | 7487232 |
| Urate | Albumin | -0.055772064 | 0.000364133 | 0.054675069 | 7518438 |

| | | | | | |
|---|---|---|---|---|---|
| Impedance of Left Arm | Albumin | -0.055426374 | 0.000364893 | 0.005089174 | 7487430 |
| GAMMA-GLUTAMYL TRANSFERASE | Urate | 0.048871731 | 0.000441883 | 1.24E-06 | 5109118 |
| Impedance of Right Arm | Testosterone | 0.038600427 | 0.000364534 | 1.24E-06 | 7514097 |
| Impedance of Left Arm | Testosterone | 0.038007017 | 0.000364538 | 1.39E-10 | 7514278 |
| Impedance of Right Arm | PEAK EXPIRATORY FLOW | -0.037195757 | 0.000364368 | 0.001756414 | 7521746 |
| 25-Hydroxyvitamin D | Albumin | -0.036311691 | 0.000378087 | 0.000989817 | 6986246 |
| Cystatin C | Albumin | 0.03527595 | 0.000688735 | 0.086768066 | 2105499 |
| Coffee | RED CELL DISTRIBUTION WIDTH | 0.034562873 | 0.000363416 | 9.99E-96 | 7562633 |
| PEAK EXPIRATORY FLOW | Albumin | 0.03353113 | 0.000365174 | 2.10E-21 | 7490529 |
| Cystatin C | Impedance of Left Arm | -0.033217995 | 0.000689667 | 0.016134652 | 2100106 |
| Cystatin C | Impedance of Right Arm | -0.033166274 | 0.000689681 | 0.054675069 | 2100031 |
| Cystatin C | Uric Acid | -0.032479434 | 0.000735575 | 0.081235452 | 1846240 |
| Impedance of Left Arm | PEAK EXPIRATORY FLOW | -0.032465394 | 0.000364423 | 7.35E-124 | 7521945 |
| Creatinine | Testosterone | -0.032218978 | 0.00036453 | 3.68E-18 | 7517640 |
| Coffee | Creatinine | 0.031877638 | 0.000364509 | 7.24E-194 | 7518695 |
| Coffee | Albumin | 0.031868098 | 0.000363736 | 2.58E-132 | 7550671 |
| Creatinine | Cystatin C | 0.029909401 | 0.000689504 | 0.002967123 | 2101538 |
| PEAK EXPIRATORY FLOW | Sodium in Urine | -0.026769476 | 0.000364403 | 3.55E-08 | 7525319 |
| Impedance of Right Arm | RED CELL DISTRIBUTION WIDTH | 0.02640069 | 0.000365658 | 4.57E-11 | 7473890 |
| Coffee | Sodium in Urine | 0.024501513 | 0.000364585 | 6.30E-144 | 7518695 |
| Impedance of Left Arm | RED CELL DISTRIBUTION WIDTH | 0.024127715 | 0.000365674 | 6.56E-132 | 7474084 |
| Sodium in Urine | Testosterone | -0.023205989 | 0.000364621 | 1.83E-106 | 7517642 |
| 25-Hydroxyvitamin D | PEAK EXPIRATORY FLOW | -0.021926327 | 0.000378518 | 1.10E-48 | 6976177 |
| Coffee | Urate | -0.021888582 | 0.000363907 | 3.22E-148 | 7547655 |

| | | | | | |
|---|---|---|---|---|---|
| PEAK EXPIRATORY FLOW | RED CELL DISTRIBUTION WIDTH | 0.02124133 | 0.000365623 | 5.12E-126 | 7477180 |
| Urea | Urate | 0.019487301 | 0.000646937 | 6.23E-193 | 2388421 |
| GAMMA-GLUTAMYL TRANSFERASE | Urea | -0.01909846 | 0.001055171 | 3.19E-79 | 897834 |
| Cystatin C | RED CELL DISTRIBUTION WIDTH | -0.01842678 | 0.000686856 | 3.53E-137 | 2118955 |
| PEAK EXPIRATORY FLOW | Urate | 0.017568948 | 0.000366284 | 8.68E-16 | 7451274 |
| GAMMA-GLUTAMYL TRANSFERASE | Sodium in Urine | -0.017386746 | 0.000452648 | 2.06E-31 | 4879187 |
| 25-Hydroxyvitamin D | RED CELL DISTRIBUTION WIDTH | 0.016730421 | 0.000365073 | 2.60E-73 | 7500979 |
| 25-Hydroxyvitamin D | Urate | 0.015572662 | 0.000364819 | 3.60E-76 | 7511747 |
| RED CELL DISTRIBUTION WIDTH | Phosphate | 0.015253695 | 0.000635868 | 3.64E-127 | 2472665 |
| 25-Hydroxyvitamin D | Sodium in Urine | -0.015144451 | 0.000378559 | 8.35E-06 | 6976445 |
| Cystatin C | Testosterone | -0.014974748 | 0.000685294 | 7.51E-106 | 2128866 |
| Creatinine | PEAK EXPIRATORY FLOW | 0.014591541 | 0.000364495 | 4.36E-08 | 7525317 |
| GAMMA-GLUTAMYL TRANSFERASE | Albumin | 0.013826144 | 0.0004526 | 5.96E-205 | 4880762 |
| Impedance of Left Arm | Urea | -0.013373857 | 0.000667853 | 3.32E-89 | 2241615 |
| Impedance of Right Arm | Uric Acid | 0.013323737 | 0.000452899 | 3.16E-190 | 4874385 |
| Cystatin C | Sodium in Urine | 0.012955041 | 0.000689755 | 1.06E-78 | 2101540 |
| 25-Hydroxyvitamin D | Cystatin C | -0.012641131 | 0.000697973 | 2.60E-73 | 2052360 |
| Impedance of Left Arm | Uric Acid | 0.012198582 | 0.000452896 | 8.64E-160 | 4874597 |
| Coffee | PEAK EXPIRATORY FLOW | -0.011789549 | 0.000364675 | 2.75E-229 | 7518418 |
| Impedance of Right Arm | Urea | -0.011573684 | 0.000667869 | 2.83E-67 | 2241607 |
| GAMMA-GLUTAMYL TRANSFERASE | RED CELL DISTRIBUTION WIDTH | -0.011278448 | 0.000441619 | 7.30E-144 | 5126829 |
| Uric Acid | Testosterone | 0.010698283 | 0.000440704 | 3.56E-130 | 5148210 |
| PEAK EXPIRATORY FLOW | Uric Acid | 0.009923038 | 0.000452777 | 1.83E-106 | 4877393 |
| 25-Hydroxyvitamin D | Phosphate | 0.009479879 | 0.000646473 | 1.10E-48 | 2392547 |

| | | | | | |
|---|---|---|---|---|---|
| RED CELL DISTRIBUTION WIDTH | Testosterone | 0.009134743 | 0.000345298 | 3.22E-154 | 8386419 |
| Coffee | Testosterone | 0.009075307 | 0.000362211 | 1.53E-138 | 7621496 |
| PEAK EXPIRATORY FLOW | Testosterone | -0.009012444 | 0.000364712 | 8.12E-135 | 7517365 |
| Urea | Sodium in Urine | -0.008398002 | 0.000667866 | 2.92E-36 | 2241769 |
| Coffee | Phosphate | 0.008260794 | 0.000635666 | 1.30E-38 | 2474645 |
| GAMMA-GLUTAMYL TRANSFERASE | Testosterone | 0.008047546 | 0.000440647 | 1.63E-74 | 5149809 |
| Creatinine | Urea | -0.007856851 | 0.000667869 | 5.98E-32 | 2241769 |
| Cystatin C | GAMMA-GLUTAMYL TRANSFERASE | -0.007383254 | 0.000735846 | 1.08E-23 | 1846725 |
| Cystatin C | Urea | 0.007003053 | 0.002043372 | 0.0609831 | 239488 |
| Cystatin C | Phosphate | -0.006722946 | 0.00068676 | 1.25E-22 | 2120169 |
| Phosphate | Urea | 0.006593483 | 0.001570467 | 2.69E-05 | 405437 |
| Coffee | Uric Acid | 0.006593177 | 0.000451273 | 2.42E-48 | 4910234 |
| GAMMA-GLUTAMYL TRANSFERASE | Impedance of Right Arm | 0.006559325 | 0.000452857 | 1.52E-47 | 4875943 |
| GAMMA-GLUTAMYL TRANSFERASE | PEAK EXPIRATORY FLOW | 0.006461413 | 0.00045272 | 3.25E-46 | 4878915 |
| Creatinine | Phosphate | -0.006312024 | 0.00063769 | 4.24E-23 | 2459027 |
| GAMMA-GLUTAMYL TRANSFERASE | Impedance of Left Arm | 0.006089379 | 0.000452849 | 3.21E-41 | 4876142 |
| Impedance of Right Arm | Phosphate | 0.006032922 | 0.000637896 | 3.15E-21 | 2457451 |
| RED CELL DISTRIBUTION WIDTH | Albumin | -0.005752272 | 0.000363653 | 2.34E-56 | 7561550 |
| RED CELL DISTRIBUTION WIDTH | Sodium in Urine | 0.005676472 | 0.000365693 | 2.44E-54 | 7477454 |
| Impedance of Left Arm | Phosphate | 0.005080032 | 0.000637888 | 1.67E-15 | 2457533 |
| RED CELL DISTRIBUTION WIDTH | Urea | 0.005010812 | 0.000640179 | 4.99E-15 | 2439982 |
| RED CELL DISTRIBUTION WIDTH | Uric Acid | -0.00485626 | 0.00044171 | 4.07E-28 | 5125247 |
| Urea | Testosterone | 0.004663309 | 0.00064461 | 4.68E-13 | 2406556 |
| Testosterone | Albumin | -0.004546697 | 0.000363914 | 8.06E-36 | 7550802 |

| | | | | | |
|---|---|---|---|---|---|
| Creatinine | GAMMA-GLUTAMYL TRANSFERASE | 0.004398553 | 0.000452712 | 2.58E-22 | 4879187 |
| Urea | Albumin | -0.00439331 | 0.000662283 | 3.28E-11 | 2279839 |
| Cystatin C | PEAK EXPIRATORY FLOW | -0.003934865 | 0.00068983 | 1.17E-08 | 2101399 |
| Creatinine | Uric Acid | 0.003855043 | 0.000452784 | 1.68E-17 | 4877665 |
| Uric Acid | Albumin | 0.003694565 | 0.000452712 | 3.32E-16 | 4879217 |
| Urea | Uric Acid | 0.003671381 | 0.001055541 | 0.0504798 | 897519 |
| PEAK EXPIRATORY FLOW | Phosphate | 0.00362509 | 0.000637717 | 1.31E-08 | 2458886 |
| Uric Acid | Sodium in Urine | 0.003601034 | 0.000452784 | 1.82E-15 | 4877665 |
| Phosphate | Urate | -0.003483476 | 0.000635391 | 4.20E-08 | 2476926 |
| Coffee | GAMMA-GLUTAMYL TRANSFERASE | -0.003340332 | 0.00045121 | 1.33E-13 | 4911763 |
| PEAK EXPIRATORY FLOW | Urea | -0.003322435 | 0.000667886 | 6.54E-07 | 2241769 |
| 25-Hydroxyvitamin D | GAMMA-GLUTAMYL TRANSFERASE | -0.002942951 | 0.00045087 | 6.70E-11 | 4919186 |
| 25-Hydroxyvitamin D | Testosterone | 0.002351178 | 0.000364539 | 1.12E-10 | 7525064 |
| 25-Hydroxyvitamin D | Coffee | 0.002299751 | 0.00037802 | 1.17E-09 | 6997889 |
| Phosphate | Testosterone | -0.002279267 | 0.00063372 | 0.000322344 | 2490022 |
| Creatinine | RED CELL DISTRIBUTION WIDTH | -0.001610809 | 0.000365698 | 1.06E-05 | 7477452 |
| Phosphate | Sodium in Urine | -0.001600081 | 0.000637702 | 0.01210267 | 2459029 |
| Phosphate | Uric Acid | 0.001596409 | 0.000691707 | 0.021003187 | 2090040 |
| 25-Hydroxyvitamin D | Uric Acid | 0.001339761 | 0.000450941 | 0.002967984 | 4917674 |
| 25-Hydroxyvitamin D | Creatinine | -0.001285099 | 0.000378602 | 0.00068797 | 6976443 |
| Phosphate | Albumin | 0.001225726 | 0.000637917 | 0.054675069 | 2457371 |
| 25-Hydroxyvitamin D | Urea | 0.001195373 | 0.00697947 | 0.008676807 | 2052837 |
| Coffee | Cystatin C | -0.000962985 | 0.00687573 | 0.016134652 | 2115251 |
| GAMMA-GLUTAMYL TRANSFERASE | Phosphate | -0.000874829 | 0.00691603 | 0.20589623 | 2090674 |

| | | | | | |
|---|---|---|---|---|---|
| Coffee | Urea | 0.000672694 | 0.066364 | 0.310753388 | 2270570 |
| RED CELL DISTRIBUTION WIDTH | Urate | 0.000668291 | 0.0345845 | 0.053317303 | 8360584 |

#### Supplementary Table 8 - LDSC $h^2$ Estimates (LDSC)

| Biomarker | h2SNP | SE | N SNPs | Cluster |
|---|---|---|---|---|
| Impedance of Left Arm | 0.493129 | 0.012133 | 7,474,084 | 1 |
| Impedance of Right Arm | 0.492093 | 0.01189 | 7,473,890 | 1 |
| Albumin (urinary excretion) | 0.078099 | 0.004813 | 7,487,430 | 2 |
| Creatinine | 0.354125 | 0.008514 | 7,477,452 | 2 |
| Sodium in Urine | 0.098134 | 0.005514 | 7,477,454 | 2 |
| 25-Hydroxyvitamin D | 0.134135 | 0.006813 | 6,976,177 | NA |
| Coffee consumption | 0.048135 | 0.003219 | 7,515,425 | NA |
| Cystatin C | 0.291398 | 0.044531 | 2,101,538 | NA |
| Gamma-glutamyl transferase | 0.403146 | 0.01135 | 4,879,187 | NA |
| Peak Expiratory Flow | 0.271039 | 0.007532 | 7,477,180 | NA |
| Phosphate | 0.348133 | 0.082139 | 2,457,533 | NA |
| Red Cell Distribution Width | 0.198135 | 0.006531 | 7,477,180 | NA |
| Testosterone | 0.25135 | 0.007239 | 7,517,365 | NA |
| Urate | 0.44201 | 0.009513 | 7,451,274 | NA |
| Urea | 0.317099 | 0.078315 | 2,241,769 | NA |
| Uric Acid | 0.438139 | 0.009409 | 4,877,665 | NA |

Supplementary Table 9 - Candidate Biomarkers' Independent Effects on Cardiometabolic Outcomes (MVMR)

| Exposure | Beta | SE | P Value | nSNPs | Outcome | F Stat |
|---|---|---|---|---|---|---|
| 25-Hydroxyvitamin D | 0.036741 | 0.005198 | 2.19E-12 | 1946 | BMI | 1.121879 |
| Coffee | 0.115134 | 0.026896 | 1.95E-05 | 1946 | BMI | 1.121879 |
| Creatinine | -0.26114 | 0.025996 | 3.53E-23 | 1946 | BMI | 1.121879 |
| Cystatin C | 0.032222 | 0.021746 | 0.138567 | 1946 | BMI | 1.121879 |
| GAMMA-GLUTAMYL TRANSFERASE | 0.030156 | 0.007179 | 2.79E-05 | 1946 | BMI | 1.121879 |
| Impedance of Left Arm | 0.365675 | 0.070945 | 2.81E-07 | 1946 | BMI | 1.121879 |
| Impedance of Right Arm | -0.68338 | 0.07375 | 4.99E-20 | 1946 | BMI | 1.121879 |
| PEAK EXPIRATORY FLOW | -0.11636 | 0.012566 | 5.27E-20 | 1946 | BMI | 1.121879 |
| RED CELL DISTRIBUTION WIDTH | 0.001471 | 0.004172 | 0.724474 | 1946 | BMI | 1.121879 |
| Phosphate | 0.090512 | 0.018696 | 1.39E-06 | 1946 | BMI | 1.121879 |
| Urea | 0.070092 | 0.007497 | 2.35E-20 | 1946 | BMI | 1.121879 |
| Uric Acid | -0.00507 | 0.002274 | 0.02599 | 1946 | BMI | 1.121879 |
| Sodium Urine | 0.411296 | 0.01425 | 1.37E-152 | 1946 | BMI | 1.121879 |
| Testosterone | 0.101369 | 0.003386 | 4.74E-162 | 1946 | BMI | 1.121879 |
| Urate | -0.05157 | 0.003318 | 2.06E-51 | 1946 | BMI | 1.121879 |
| Albumin | 0.40408 | 0.030097 | 2.34E-39 | 1946 | BMI | 1.121879 |
| 25-Hydroxyvitamin D | -0.40493 | 0.040443 | 5.28E-23 | 1823 | DBP | 1.149121 |
| Coffee | -4.12401 | 0.151367 | 3.18E-137 | 1823 | DBP | 1.149121 |
| Creatinine | -5.55084 | 0.214854 | 1.61E-125 | 1823 | DBP | 1.149121 |
| Cystatin C | -0.46198 | 0.084326 | 4.89E-08 | 1823 | DBP | 1.149121 |
| GAMMA-GLUTAMYL TRANSFERASE | 0.433549 | 0.059613 | 5.22E-13 | 1823 | DBP | 1.149121 |
| Impedance of Left Arm | -6.71355 | 0.651275 | 2.99E-24 | 1823 | DBP | 1.149121 |
| Impedance of Right Arm | 7.191368 | 0.650898 | 1.66E-27 | 1823 | DBP | 1.149121 |
| PEAK EXPIRATORY FLOW | 2.995662 | 0.116708 | 3.80E-124 | 1823 | DBP | 1.149121 |
| RED CELL DISTRIBUTION WIDTH | 0.50566 | 0.057222 | 2.28E-18 | 1823 | DBP | 1.149121 |
| Phosphate | 1.224969 | 0.324918 | 0.000168 | 1823 | DBP | 1.149121 |
| Urea | -0.44945 | 0.106808 | 2.70E-05 | 1823 | DBP | 1.149121 |
| Uric Acid | -0.65874 | 0.037504 | 6.92E-64 | 1823 | DBP | 1.149121 |
| Sodium Urine | 6.822594 | 0.177281 | 3.57E-237 | 1823 | DBP | 1.149121 |
| Testosterone | 0.04669 | 0.030034 | 0.120218 | 1823 | DBP | 1.149121 |
| Urate | 0.083701 | 0.062881 | 0.183323 | 1823 | DBP | 1.149121 |

| | | | | | | |
|---|---|---|---|---|---|---|
| Albumin | 0.270222 | 0.173136 | 0.118759 | 1823 | DBP | 1.149121 |
| 25-Hydroxyvitamin D | 0.08088 | 0.088836 | 0.362714 | 1803 | SBP | 1.130693 |
| Coffee | -4.95809 | 0.244188 | 1.22E-82 | 1803 | SBP | 1.130693 |
| Creatinine | -4.09543 | 0.328658 | 3.08E-34 | 1803 | SBP | 1.130693 |
| Cystatin C | -2.82541 | 0.33063 | 2.70E-17 | 1803 | SBP | 1.130693 |
| GAMMA-GLUTAMYL TRANSFERASE | 1.611787 | 0.096307 | 1.61E-58 | 1803 | SBP | 1.130693 |
| Impedance of Left Arm | -1.55385 | 0.959869 | 0.105664 | 1803 | SBP | 1.130693 |
| Impedance of Right Arm | 7.142803 | 0.984807 | 6.05E-13 | 1803 | SBP | 1.130693 |
| PEAK EXPIRATORY FLOW | -1.45584 | 0.209036 | 4.61E-12 | 1803 | SBP | 1.130693 |
| RED CELL DISTRIBUTION WIDTH | -0.235 | 0.076795 | 0.002245 | 1803 | SBP | 1.130693 |
| Phosphate | -2.50893 | 0.264304 | 6.93E-21 | 1803 | SBP | 1.130693 |
| Urea | 1.182672 | 0.133055 | 1.48E-18 | 1803 | SBP | 1.130693 |
| Uric Acid | 0.117342 | 0.027579 | 2.20E-05 | 1803 | SBP | 1.130693 |
| Sodium Urine | 6.558183 | 0.368931 | 3.19E-65 | 1803 | SBP | 1.130693 |
| Testosterone | -1.59084 | 0.155442 | 6.27E-24 | 1803 | SBP | 1.130693 |
| Urate | 0.135663 | 0.042237 | 0.001342 | 1803 | SBP | 1.130693 |
| Albumin | 2.94989 | 0.425951 | 6.04E-12 | 1803 | SBP | 1.130693 |
| 25-Hydroxyvitamin D | -0.17881 | 0.009781 | 1.21E-68 | 1825 | HbA1c | 1.151031 |
| Coffee | -0.0701 | 0.036671 | 0.056095 | 1825 | HbA1c | 1.151031 |
| Creatinine | 0.252717 | 0.047836 | 1.42E-07 | 1825 | HbA1c | 1.151031 |
| Cystatin C | 0.288326 | 0.023263 | 6.41E-34 | 1825 | HbA1c | 1.151031 |
| GAMMA-GLUTAMYL TRANSFERASE | -0.00666 | 0.003951 | 0.092084 | 1825 | HbA1c | 1.151031 |
| Impedance of Left Arm | 2.203457 | 0.092963 | 2.21E-108 | 1825 | HbA1c | 1.151031 |
| Impedance of Right Arm | -1.81762 | 0.087933 | 2.19E-85 | 1825 | HbA1c | 1.151031 |
| PEAK EXPIRATORY FLOW | 0.16186 | 0.022467 | 8.52E-13 | 1825 | HbA1c | 1.151031 |
| RED CELL DISTRIBUTION WIDTH | -0.09255 | 0.010802 | 2.22E-17 | 1825 | HbA1c | 1.151031 |
| Phosphate | 0.383521 | 0.064896 | 4.08E-09 | 1825 | HbA1c | 1.151031 |
| Urea | 0.07609 | 0.011102 | 9.83E-12 | 1825 | HbA1c | 1.151031 |
| Uric Acid | 0.052234 | 0.005285 | 1.79E-22 | 1825 | HbA1c | 1.151031 |
| Sodium Urine | -0.42012 | 0.024269 | 3.04E-62 | 1825 | HbA1c | 1.151031 |
| Testosterone | 0.215304 | 0.012891 | 2.35E-58 | 1825 | HbA1c | 1.151031 |
| Urate | -0.05449 | 0.00666 | 5.24E-16 | 1825 | HbA1c | 1.151031 |
| Albumin | -0.27388 | 0.051314 | 1.06E-07 | 1825 | HbA1c | 1.151031 |

| | | | | | | |
|---|---|---|---|---|---|---|
| 25-Hydroxyvitamin D | 0.08669 | 0.011083 | 8.58E-15 | 1906 | HDL | 1.107075 |
| Coffee | 0.334356 | 0.039983 | 1.18E-16 | 1906 | HDL | 1.107075 |
| Creatinine | -0.33075 | 0.041854 | 4.60E-15 | 1906 | HDL | 1.107075 |
| Cystatin C | -0.15589 | 0.040885 | 0.000142 | 1906 | HDL | 1.107075 |
| GAMMA-GLUTAMYL TRANSFERASE | 0.080502 | 0.009207 | 4.88E-18 | 1906 | HDL | 1.107075 |
| Impedance of Left Arm | 0.142476 | 0.142202 | 0.316504 | 1906 | HDL | 1.107075 |
| Impedance of Right Arm | -0.13072 | 0.152009 | 0.389943 | 1906 | HDL | 1.107075 |
| PEAK EXPIRATORY FLOW | -0.49303 | 0.019745 | 3.66E-119 | 1906 | HDL | 1.107075 |
| RED CELL DISTRIBUTION WIDTH | -0.11721 | 0.006067 | 4.81E-76 | 1906 | HDL | 1.107075 |
| Phosphate | 1.270655 | 0.066825 | 6.60E-74 | 1906 | HDL | 1.107075 |
| Urea | 0.097513 | 0.020775 | 2.88E-06 | 1906 | HDL | 1.107075 |
| Uric Acid | -0.21492 | 0.009151 | 3.07E-107 | 1906 | HDL | 1.107075 |
| Sodium Urine | 1.301575 | 0.034452 | 3.64E-233 | 1906 | HDL | 1.107075 |
| Testosterone | 0.110097 | 0.014753 | 1.28E-13 | 1906 | HDL | 1.107075 |
| Urate | 0.263986 | 0.010114 | 1.69E-128 | 1906 | HDL | 1.107075 |
| Albumin | -0.50434 | 0.052588 | 2.66E-21 | 1906 | HDL | 1.107075 |
| 25-Hydroxyvitamin D | 0.025377 | 0.018165 | 0.162568 | 1906 | LDL | 1.107075 |
| Coffee | 0.267917 | 0.064948 | 3.87E-05 | 1906 | LDL | 1.107075 |
| Creatinine | 0.261233 | 0.059098 | 1.04E-05 | 1906 | LDL | 1.107075 |
| Cystatin C | 0.356594 | 0.039362 | 3.19E-19 | 1906 | LDL | 1.107075 |
| GAMMA-GLUTAMYL TRANSFERASE | 0.103369 | 0.012187 | 4.40E-17 | 1906 | LDL | 1.107075 |
| Impedance of Left Arm | 0.92207 | 0.170168 | 6.78E-08 | 1906 | LDL | 1.107075 |
| Impedance of Right Arm | -0.85575 | 0.184077 | 3.57E-06 | 1906 | LDL | 1.107075 |
| PEAK EXPIRATORY FLOW | -0.79021 | 0.025276 | 2.69E-173 | 1906 | LDL | 1.107075 |
| RED CELL DISTRIBUTION WIDTH | -0.04045 | 0.01256 | 0.001301 | 1906 | LDL | 1.107075 |
| Phosphate | 1.378855 | 0.098384 | 1.57E-42 | 1906 | LDL | 1.107075 |
| Urea | 0.369447 | 0.019896 | 7.80E-71 | 1906 | LDL | 1.107075 |
| Uric Acid | -0.17223 | 0.00806 | 6.45E-91 | 1906 | LDL | 1.107075 |
| Sodium Urine | 0.423382 | 0.063064 | 2.50E-11 | 1906 | LDL | 1.107075 |
| Testosterone | 0.010613 | 0.01975 | 0.591055 | 1906 | LDL | 1.107075 |
| Urate | 0.153765 | 0.008034 | 9.06E-75 | 1906 | LDL | 1.107075 |
| Albumin | -0.54809 | 0.070375 | 1.11E-14 | 1906 | LDL | 1.107075 |
| 25-Hydroxyvitamin D | -0.53596 | 0.017874 | 6.29E-162 | 1906 | Triglycerides | 1.107075 |

| | | | | | | |
|---|---|---|---|---|---|---|
| Coffee | -1.42165 | 0.054055 | 3.58E-130 | 1906 | Triglycerides | 1.107075 |
| Creatinine | 0.756573 | 0.064932 | 2.39E-30 | 1906 | Triglycerides | 1.107075 |
| Cystatin C | 0.831801 | 0.052016 | 4.39E-54 | 1906 | Triglycerides | 1.107075 |
| GAMMA-GLUTAMYL TRANSFERASE | -0.01628 | 0.018868 | 0.388362 | 1906 | Triglycerides | 1.107075 |
| Impedance of Left Arm | -0.22671 | 0.124222 | 0.068159 | 1906 | Triglycerides | 1.107075 |
| Impedance of Right Arm | 0.829624 | 0.131996 | 4.05E-10 | 1906 | Triglycerides | 1.107075 |
| PEAK EXPIRATORY FLOW | 0.662247 | 0.054183 | 4.05E-33 | 1906 | Triglycerides | 1.107075 |
| RED CELL DISTRIBUTION WIDTH | -0.18702 | 0.006079 | 7.73E-169 | 1906 | Triglycerides | 1.107075 |
| Phosphate | -0.04533 | 0.059479 | 0.446038 | 1906 | Triglycerides | 1.107075 |
| Urea | 0.11396 | 0.020455 | 2.89E-08 | 1906 | Triglycerides | 1.107075 |
| Uric Acid | 0.041188 | 0.007878 | 1.90E-07 | 1906 | Triglycerides | 1.107075 |
| Sodium Urine | -0.59606 | 0.0481 | 5.78E-34 | 1906 | Triglycerides | 1.107075 |
| Testosterone | 0.373469 | 0.016209 | 9.84E-104 | 1906 | Triglycerides | 1.107075 |
| Urate | -0.00957 | 0.011147 | 0.390508 | 1906 | Triglycerides | 1.107075 |
| Albumin | -0.56437 | 0.078155 | 7.44E-13 | 1906 | Triglycerides | 1.107075 |
| 25-Hydroxyvitamin D | -1.59505 | 0.2272 | 3.06E-12 | 1937 | T2D | 1.123534 |
| Coffee | -1.99199 | 0.305544 | 8.99E-11 | 1937 | T2D | 1.123534 |
| Creatinine | 1.663269 | 0.455558 | 0.000268 | 1937 | T2D | 1.123534 |
| Cystatin C | -0.04113 | 0.569972 | 0.942485 | 1937 | T2D | 1.123534 |
| GAMMA-GLUTAMYL TRANSFERASE | 1.275898 | 0.153134 | 1.49E-16 | 1937 | T2D | 1.123534 |
| Impedance of Left Arm | 0.184556 | 1.594676 | 0.907877 | 1937 | T2D | 1.123534 |
| Impedance of Right Arm | 0.069045 | 1.562228 | 0.964752 | 1937 | T2D | 1.123534 |
| PEAK EXPIRATORY FLOW | 2.244235 | 0.476172 | 2.62E-06 | 1937 | T2D | 1.123534 |
| RED CELL DISTRIBUTION WIDTH | -0.1449 | 0.08972 | 0.106478 | 1937 | T2D | 1.123534 |
| Phosphate | 12.1544 | 1.027282 | 3.16E-31 | 1937 | T2D | 1.123534 |
| Urea | 0.180346 | 0.211187 | 0.393232 | 1937 | T2D | 1.123534 |
| Uric Acid | 0.395829 | 0.116465 | 0.000691 | 1937 | T2D | 1.123534 |
| Sodium Urine | 2.301616 | 0.483582 | 2.09E-06 | 1937 | T2D | 1.123534 |
| Testosterone | 0.824131 | 0.171599 | 1.69E-06 | 1937 | T2D | 1.123534 |
| Urate | 0.515043 | 0.1185 | 1.46E-05 | 1937 | T2D | 1.123534 |
| Albumin | 0.882247 | 0.580831 | 0.128942 | 1937 | T2D | 1.123534 |
| 25-Hydroxyvitamin D | -0.06268 | 0.009371 | 3.00E-11 | 1838 | MetS | 1.173883 |
| Coffee | 0.075297 | 0.027721 | 0.006666 | 1838 | MetS | 1.173883 |

| | | | | | | |
|---|---|---|---|---|---|---|
| Creatinine | -0.08162 | 0.029443 | 0.005628 | 1838 | MetS | 1.173883 |
| Cystatin C | -0.32062 | 0.02823 | 6.24E-29 | 1838 | MetS | 1.173883 |
| GAMMA-GLUTAMYL TRANSFERASE | 0.001214 | 0.006784 | 0.857979 | 1838 | MetS | 1.173883 |
| Impedance of Left Arm | 0.310972 | 0.092419 | 0.000782 | 1838 | MetS | 1.173883 |
| Impedance of Right Arm | -0.09167 | 0.091886 | 0.318605 | 1838 | MetS | 1.173883 |
| PEAK EXPIRATORY FLOW | 0.091494 | 0.016806 | 5.91E-08 | 1838 | MetS | 1.173883 |
| RED CELL DISTRIBUTION WIDTH | -0.01318 | 0.004906 | 0.007296 | 1838 | MetS | 1.173883 |
| Phosphate | -0.1598 | 0.044328 | 0.000321 | 1838 | MetS | 1.173883 |
| Urea | -0.06738 | 0.011453 | 4.78E-09 | 1838 | MetS | 1.173883 |
| Uric Acid | 0.017383 | 0.00545 | 0.001451 | 1838 | MetS | 1.173883 |
| Sodium Urine | -0.17884 | 0.026152 | 1.09E-11 | 1838 | MetS | 1.173883 |
| Testosterone | 0.024392 | 0.01101 | 0.026847 | 1838 | MetS | 1.173883 |
| Urate | 0.056425 | 0.007101 | 3.35E-15 | 1838 | MetS | 1.173883 |
| Albumin | 0.180384 | 0.031411 | 1.09E-08 | 1838 | MetS | 1.173883 |

### Supplementary Table 10 - Renal Cluster Independent Effects on Cardiometabolic Outcomes (MVMR)

| Exposure | Outcome | Beta | SE | P-Value | nSNPs | F Stat | Q Stat | Q P-Value |
|---|---|---|---|---|---|---|---|---|
| Creatinine | BMI | -0.0804671 | 0.026684048 | 0.003061737 | 139 | 9.718893 | 123.4125 | 0.01374515 |
| Sodium Urine | BMI | -0.026296435 | 0.028199483 | 0.352723098 | 139 | 12.59848 | 123.4125 | 0.01374515 |
| Albumin | BMI | -0.093932231 | 0.003029786 | 1.60E-63 | 139 | 11.96678 | 123.4125 | 0.01374515 |
| Creatinine | DBP | 2.340025361 | 0.27548723 | 5.22E-14 | 127 | 9.234259 | 47.1274 | 0.01498976 |
| Sodium Urine | DBP | -2.112041412 | 0.264444575 | 8.08E-13 | 127 | 12.95644 | 47.1274 | 0.01498976 |
| Albumin | DBP | -0.888572065 | 0.031013782 | 1.60E-56 | 127 | 11.6999 | 47.1274 | 0.01498976 |
| Creatinine | SBP | 2.92027086 | 0.505349831 | 5.73E-08 | 127 | 9.234259 | 170.37463 | 0.2087211 |
| Sodium Urine | SBP | -0.454522452 | 0.37611707 | 0.229169845 | 127 | 12.95644 | 170.37463 | 0.2087211 |
| Albumin | SBP | -1.052501447 | 0.039215985 | 1.73E-53 | 127 | 11.6999 | 170.37463 | 0.2087211 |
| Creatinine | MetS | 0.112246921 | 0.143574404 | 0.4381698 | 51 | 9.155787 | 234.84189 | 8.77E-02 |
| Sodium Urine | MetS | -0.297686156 | 0.136470182 | 0.034089175 | 51 | 11.65653 | 234.84189 | 8.77E-02 |
| Albumin | MetS | 0.12831458 | 0.1028238 | 0.218118463 | 51 | 10.25478 | 234.84189 | 8.77E-02 |
